# Disease progression Mendelian randomization of risk factors for kidney function decline in chronic kidney disease and the general population

**DOI:** 10.64898/2026.09.07.26362457

**Authors:** Aristomo Andries, Venexia Walker, Artemis Briasouli, Maria Sobczyk-Barad, Jie Zheng, Mathias Gorski, Iris M. Heid, Stein Ivar Hallan, Kate Tilling, Humaira Rasheed, Bjørn Olav Åsvold, Ben Michael Brumpton, Tom R Gaunt

## Abstract

**Introduction:** With limited treatment options, identifying risk factors for chronic kidney disease (CKD) disease progression is an important aspect of CKD management. Novel approaches for correcting case-only genome-wide association studies (GWAS) for index event bias and the availability of large GWAS conducted in people with CKD may allow us to identify such risk factors.

**Methods:** We applied Mendelian randomization (MR) using European ancestry data from CKDGen consortium to estimate the causal effect of 74 putative risk factors for CKD progression on annual decline in estimated glomerular filtration rate (eGFR) among people with CKD (N=26,653) and in the general population (N=343,339). Additionally, we examined the effect of the same risk factors on incident CKD (N=480,698). We accounted for index event bias using the Dudbridge et.al. and SlopeHunter methods in CKD case-only analyses.

**Results:** Type-2 diabetes (T2D) and high thyroid-stimulating hormone (TSH) increased annual decline in eGFR in CKD population of 0.102 (0.035,0.168) and 0.353 (0.108,0.598) ml/min/1.73 m^2^ per 1 log-odds higher exposure respectively. These effects were consistent after index event bias adjustment. In unadjusted analysis, pulse pressure (PP) (CKD population: 0.221 (0.041,0.402); general population: 0.059 (0.028,0.091) ml/min/1.73 m^2^ per SD increase); and serum uric acid level (CKD population: 0.252 (0.081,0.423); general population: 0.041 (0.007,0.075) ml/min/1.73 m^2^ per SD increase) also increased annual eGFR decline. Conversely, educational attainment decreased annual eGFR decline (CKD population: −0.051 (−0.102,-0.001); general population: −0.016 (−0.025,-0.007)).

**Conclusion:** We applied novel methodological approaches to correct for index event bias in CKD case-only GWAS, enabling investigation of putative risk factors for CKD progression.

Cardiometabolic traits showed suggestive effects on kidney function decline and there was evidence of a causal effect of T2D and high TSH on annual decline in eGFR. These findings highlight potential targets to slow CKD progression.

## Introduction

Chronic kidney disease (CKD) is characterized by a decrease in kidney function over time, lasting for at least three months or more^1–3^. Compared to the previous decade, efforts to manage CKD have gained more attention^4^ including promising drug repurposing strategies^5^, such as the use of anti-diabetic SGLT2 inhibitors, for improving kidney function while simultaneously benefitting the other cardiometabolic indices^6,7^.

Despite progress in treatment development, there is no cure for CKD and knowledge of factors which preserve kidney function is lacking^8^. Previous studies have recognised risk factors, such as hypertension, cardiovascular disease, and diabetes, are associated with kidney function deterioration^9,10^. Nevertheless, risk factors such as obesity and body mass index (BMI) have shown a more nuanced effect on CKD disease progression^11,12^. This seemingly counterintuitive result might have resulted from confounders or actual biological mechanism^13^, which requires more investigation. In this context, novel disease progression analysis methods for correcting genome-wide association studies (GWAS)^14,15^ and the recent availability of CKD case-only GWAS^16^ may allow us to better understand the effect of such factors. This strategy may direct us towards mechanisms which slow down or accelerate CKD progression and for which interventions may reduce the number of patients progressing to end stage kidney disease.

Mendelian randomisation (MR) is a causal inference approach that uses germline genetic variations as instruments to study disease risk factors^17^. The approach has some advantages compared to a traditional observational study in that it can give estimates which are not influenced by non-genetic confounders and reverse causation^17,18^. However, to study risk factors for disease progression, such as kidney function decline among CKD patients, is challenging because it requires access to large-scale genetic analysis of case-only designs. Additionally, case-only studies are potentially subject to index event bias, which can occur in traditional epidemiology and MR studies alike^17,19^. Index event bias can make a causal risk factor for incident disease falsely appear as a causal risk factor for disease progression, due to an induced association with another causal risk factor for incident disease^17,19^ (**Box 1**).

In this study, we applied Mendelian randomization to assess the causal effect of a range of risk factors for annual decline in estimated glomerular filtration rate (eGFR) in CKD and general populations, while accounting for the potential index event bias that arises from using a CKD population. Systematic assessment of risk factors for eGFR decline in both CKD and general populations will further our understanding of the impact of risk factors for CKD disease progression. These risk factors may suggest mechanisms specific to people with CKD, which can be used to guide CKD management.

## Methods

### Study design

We performed two-sample Mendelian randomization to estimate the causal effects of 74 putative risk factors for CKD progression on annual decline in eGFR in CKD population. Additionally, we tested the same risk factors against incident CKD and in the general population (**Figure 1**).

**Figure 1.**
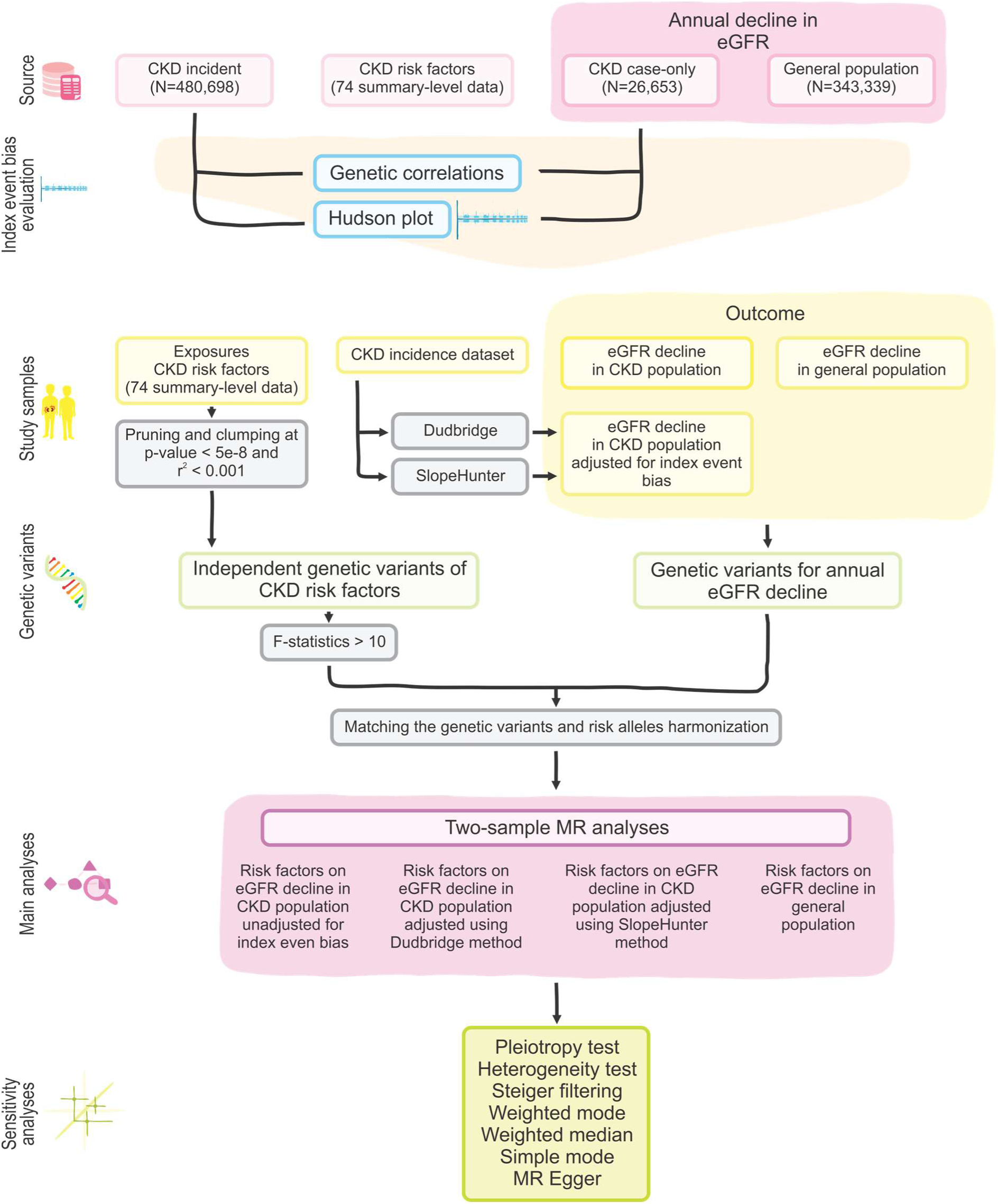
Study analyses flowchart

### Exposures

We identified putative risk factors for CKD progression from three sources. The first set of risk factors was derived from the Kidney Disease Improving Global Outcomes (KDIGO) 2024 guideline^3^. A second set was retrieved from the work of Zheng, et.al. which selected risk factors for CKD incidence from the PubMed database using MELODI-Presto^20^ . The final set were manually curated by clinicians to ensure relevant phenotypes were included.

Summary-level genome-wide association study (GWAS) data for the risk factors were obtained from the OpenGWAS database^21,22^. Additionally, we retrieved summary-level data from consortia for these risk factors when they were more recent, had larger sample size and/or the number of single nucleotide polymorphisms (SNPs) available was higher. We filtered to European population datasets (**Supplementary Table ST1**).

For each risk factor, we selected genome-wide significant (P < 5e-8) genetic variants to ensure they met the first MR assumption of relevance. The genetic variants were then clumped using a 10 Mb window and R^2^ linkage disequilibrium (LD) threshold of 0.001 against the 1000 genomes reference panel for the European super-population. The reference panel comprised bi-allelic variants and has minor allele frequency greater than 0.01. We transformed the continuous risk factors to the standard deviation (SD) scale and binary risk factors to log-odds to allow comparison between risk factors.

### Outcomes

We studied three outcomes: (1) annual eGFR decline in the CKD population; (2) annual eGFR decline in the general population; and (3) incident CKD. For each outcome, we used the latest summary-level data from the CKDGen consortium GWAS meta-analyses, which includes 62 longitudinal studies predominantly sourced from people of European ancestry. For annual decline in eGFR, the CKD case-only GWAS included 26,653 people, while the general population GWAS included 343,339 people^16,24^. For both GWAS, annual eGFR decline^16^ was measured in ml/min/1.73 m^2^ and defined as 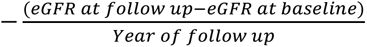. The incident CKD GWAS comprised of 41,395 CKD cases and 439,303 controls ^23^. We used this GWAS to assess and correct for index event bias in the case-only GWAS.

### Consideration of index event bias

#### Evaluation

We visually assessed whether index event bias was likely in our study using a Hudson plot of the GWAS for incident CKD^23^ and eGFR decline in CKD population^16^. The Hudson plots consist of two Miami plots aligned by chromosome position on the x-axis. If SNP associations with CKD incidence occur close to SNP associations with eGFR decline in CKD population then index event bias may be present. We further assessed whether index event bias was possible using the genetic correlation between these two phenotypes using ‘ldsc’ in python^25,26^.

*Correction using the Dudbridge et.al. method*

The Dudbridge et.al. method calculates an index event bias correction from the residuals of the regression of the case-only progression GWAS on the incident GWAS. This method assumes no correlation between genetic effects on incidence and on prognosis^14^. To account for regression dilution, we weighted using the simulation extrapolation (SIMEX) approach with lambda values between 0.25 to 5 with 0.25 increments. SIMEX was implemented with 10,000 simulations.

#### Correction using the SlopeHunter method

The SlopeHunter method models the bivariate distribution of the incidence and prognosis genetic effect sizes using model-based clustering to identify the set of variants likely affecting incidence only. These variants are used to estimate index event bias correction. Unlike the Dudbridge method, SlopeHunter does not assume no correlation between genetic effects on incidence and on prognosis^15^. We set the initial weight of the mixture components of SNPs affecting incidence only to 0.6 and the initial covariance between incidence and prognosis to 1e-5. Multiple p-value thresholds for SNP-incidence associations (1e-1, 1e-2, 1e-3, and 1e-4) were tested to assess sensitivity of the method. We performed bootstraps using 10,000 samples. We assessed plots of the SNP membership in the clusters as a visual aid to determine at which threshold the algorithm was able to distinguish clusters of SNPs affecting incidence only and both incidence and progression^15^. The ability to separate these two clusters indicates a more robust correction factor^15,27^.

### Mendelian randomization analyses

We applied two-sample MR to estimate the effect of putative risk factors for CKD progression on kidney function measured by the annual decline in eGFR^16,24^. The effects of risk factors on annual eGFR decline in CKD population, unadjusted for index event bias, were obtained using a two-sample MR approach^17,19,22^, presented as

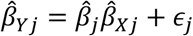

and in case-only group

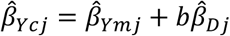

whereas *β̂_j_* is the effect estimates of risk factors on eGFR decline, *β̂_Xj_* is the association between SNPs and CKD risk factors, *β̂_Yj_* is the association between SNPs and the outcome eGFR decline, *β̂_Ycj_* is the *β̂_Yj_* in CKD population dataset, *β̂_Ymj_* is the *β̂_Yj_* without index event bias, *β̂_Dj_* is the genetic association with CKD incidence, b is the constant for index event bias. Then, we assessed the presence of index event bias resulting from conditioning on CKD population and subsequently applied adjustment factors (*b*) using Dudbridge^14^ and SlopeHunter^15^ methods. We compared MR with unadjusted (*β̂_cj_*) and adjusted (*β̂_cj,adj_*) estimates to infer the causal effects of CKD risk factors on CKD progression in the presence of potential index event bias. Additionally, we presented the unadjusted estimates of CKD risk factors on eGFR decline in both CKD (*β̂_cj_*) and general (*β̂_j_*) populations.

For our main analysis, we present both the unadjusted analysis (*β̂_cj_*), reflecting higher statistical power, and adjusted analysis (*β̂_cj,adj(Db)_* for Dudbridge and *β̂_cj,adj(SH)_* for SlopeHunter methods), representing lower index event bias. In both cases, we used inverse variance weighted (IVW) for phenotypes with multiple SNPs or Wald ratio when only one SNP was available^17^. If point estimates of the same risk factors were both positive or negative, we considered them to have same effect direction. If point estimates were in the opposite direction, we considered them as indeterminate.

### Sensitivity analyses

We applied MR using simple median, simple mode, weighted median, weighted mode and MR-Egger as sensitivity analyses to evaluate consistency of our results. Heterogeneity statistics were used to determine consistency of estimates for all instruments for each phenotype^28,29^. To assess pleiotropy, we inspected MR-Egger intercept and its P value^17,30^. The *I*^2^ statistics were calculated to assess the NO Measurement Error (NOME) assumption of MR-Egger as a measure of potential attenuation bias^31^. We repeated the analysis with Steiger filtering to further assess the validity of our instruments for the risk factors.

### Software and code availability

We used R version 4.4.1 and python version 3.12.12. Our code is publicly available: https://github.com/hunt-genes/CKDProgressionMR.

## Results

### Consideration of index event bias

Using the Hudson plot (**Figure 2**), we identified SNPs at the UMOD-PDILT loci on chromosome 16 that were associated with both incident CKD and eGFR decline in the CKD population. The estimated genetic correlation between the GWAS of incident CKD and eGFR decline in CKD population was 0.57 (SE=0.17) (**Supplementary Table ST2**). Using the Dudbridge, et.al. method, the index event bias adjustment factor was 0.60 (95% CI 0.60, 0.60). The SlopeHunter adjustment factors ranged from 0.33 (95% CI −0.10, 0.75; P-value threshold 1e-4) to 0.74 (95% CI 0.60, 0.87; P-value threshold 1e-1) (**Table 1**).

**Figure 2.**
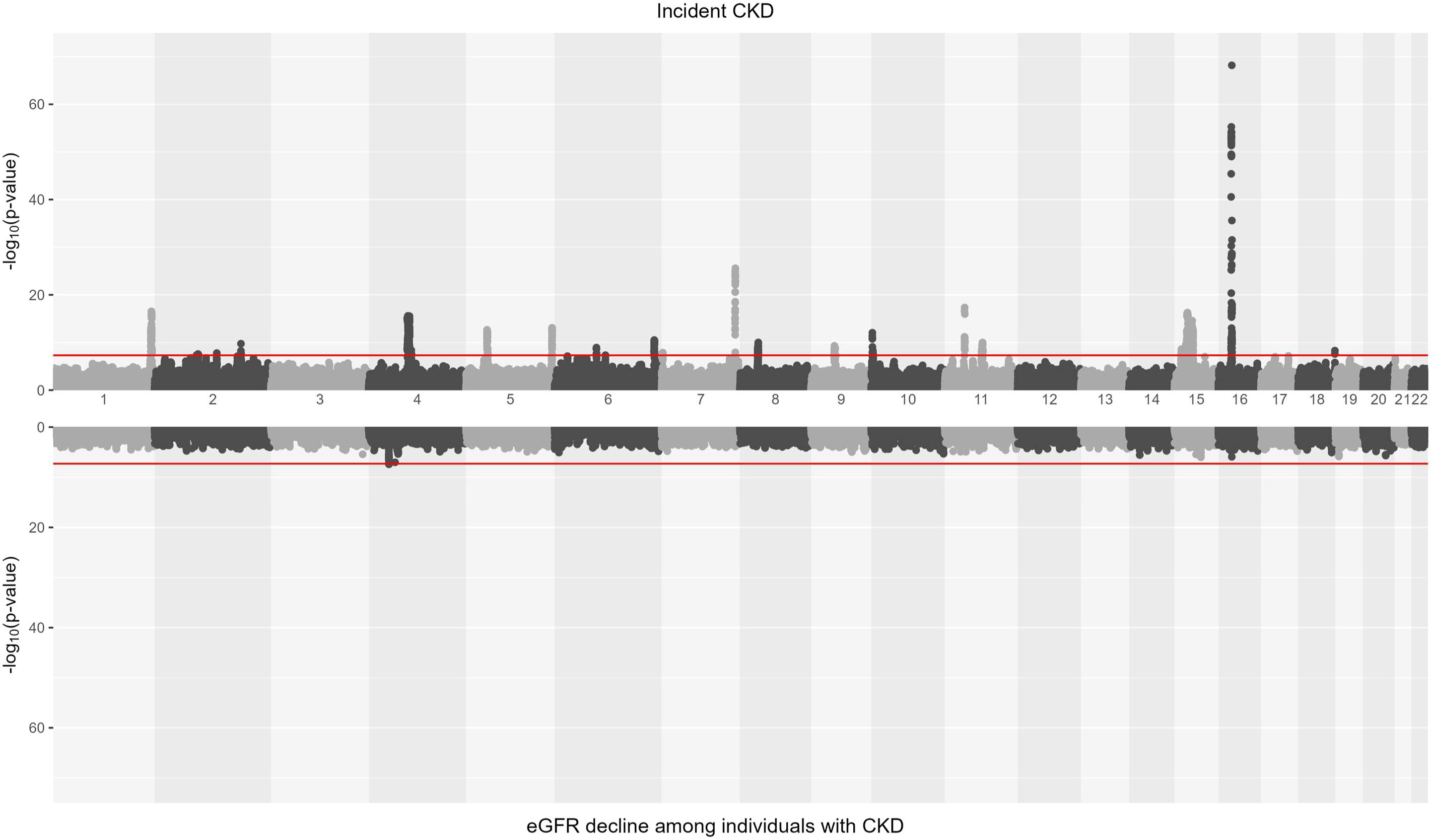
Hudson plot illustrates the incident CKD and eGFR decline in the CKD population

**Table 1.** Summary of adjustment factors for index event bias.

| Method | P-value thresholds for SNP-incidence associations | Adjustment factor estimates (95% CI) |
| --- | --- | --- |
| Dudbridge, et.al. | Not applicable | 0.60 (95% CI 0.60, 0.60) |
| SlopeHunter | 1e-1 | 0.74 (95% CI 0.60, 0.88) |
| SlopeHunter | 1e-2 | 0.58 (95% CI 0.35, 0.82) |
| SlopeHunter | 1e-3 | 0.54 (95% CI 0.16, 0.92) |
| SlopeHunter | 1e-4 | 0.33 (95% CI -0.10, 0.76) |

95% CI, 95% confidence interval

### Risk factors for disease progression of eGFR decline in the CKD population

Applying index event bias adjustment factors had minimal impact on the annual eGFR decline effect estimates in the CKD population. The risk factors type 2 diabetes (T2D) (0.102 (0.035, 0.168) ml/min/1.73 m^2^ per log-odds increase) and high thyroid-stimulating hormone (TSH) 0.353 (0.108, 0.598) ml/min/1.73 m^2^ per log-odds increase) increased annual eGFR decline in both unadjusted and adjusted analyses. Several other risk factors increased annual eGFR decline in CKD population, but effect estimates were attenuated after correcting for index event bias. These were body mass index (0.155 (0.004, 0.305) ml/min.1,73 m^2^ per SD increase), with *β̂_cj,adj(Db)_*=0.057 (−0.097, 0.212) and *β̂_cj,adj(SH)_*=0.102 (−0.051, 0.255); pulse pressure (PP) (0.221 (0.041, 0.402) ml/min/1.73 m^2^ per SD increase), with *β̂_cj,adj(Db)_*=0.117 (−0.068, 0.303) and *β̂_cj,adj(SH)_*=0.163 (−0.022, 0.347); and serum uric acid level (0.252 (0.081, 0.423) ml/min/1.73 m^2^ per SD increase), with *β̂_cj,adj(Db)_*=0.069 (−0.101, 0.239) and *β̂_cj,adj(SH)_*=0.151 (−0.020, 0.321). Conversely, educational attainment decreased the annual decline in eGFR (−0.051 (−0.102, −0.001) ml/min/1.73 m^2^ per SD increase), albeit the effect was attenuated with correction for index event bias (*β̂_cj,adj(Db)_*=-0.037 (−0.089, 0.014) and *β̂_cj,adj(SH)_*=-0.045 (−0.096, 0.007)) (**Figure 3**, **Supplementary Figure S1 and S2**, **Supplementary Table ST3**). Nine of the 74 risk factors had a different direction of effect (indeterminate) on annual eGFR decline in the unadjusted; and both Dudbridge and SlopeHunter adjusted analyses.

**Figure 3.**
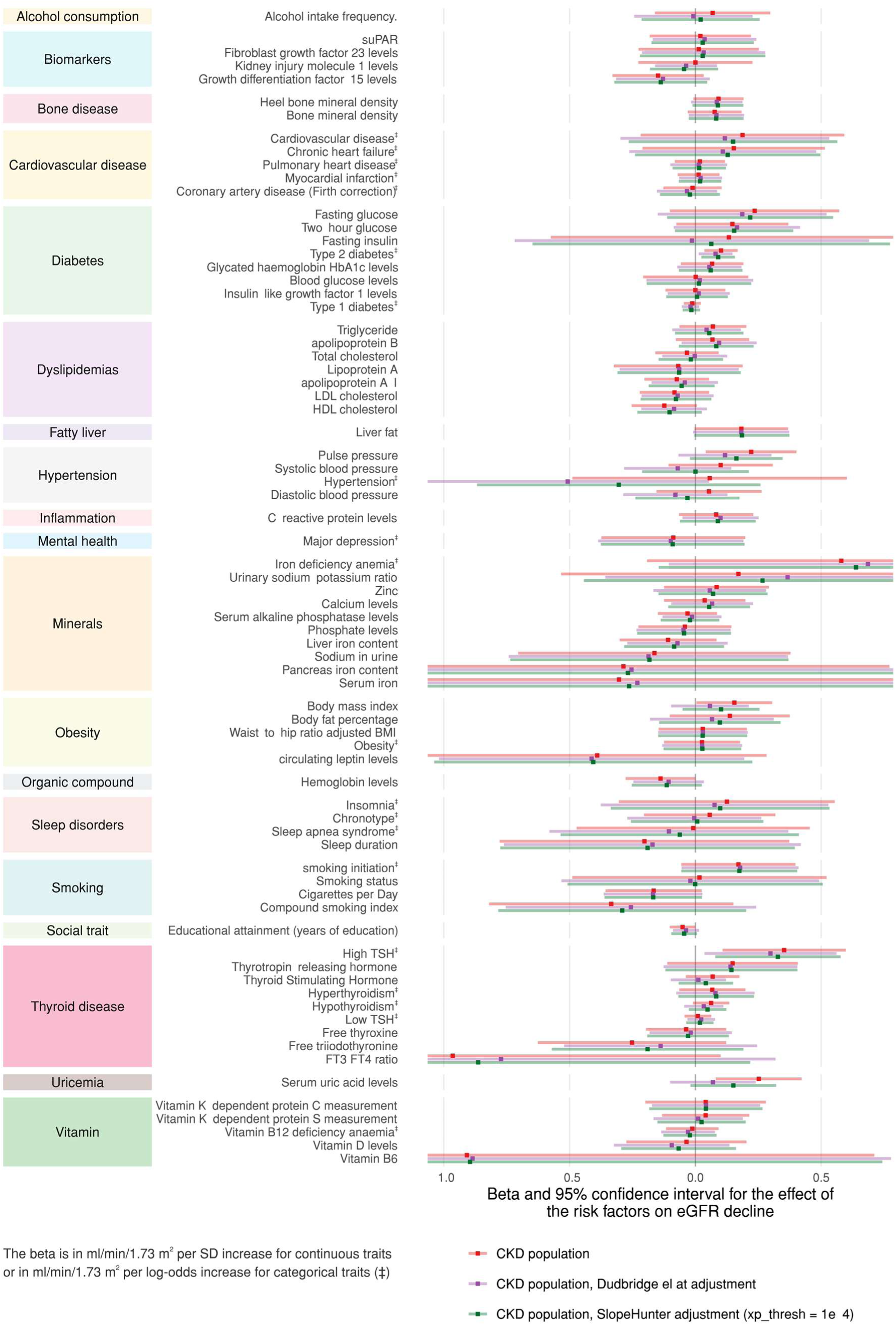
Mendelian randomisation estimates for the risk factors on the annual eGFR decline for CKD population, without and with adjusted estimates

### Risk factors for eGFR decline in the general population

Fifty of the 74 risk factors had the same effect direction of effect for eGFR decline in both the CKD and general populations, albeit with wider confidence interval in the CKD population. Risk factors including PP (0.059 (0.028, 0.091) ml/min/1.73 m^2^ per SD increase) and serum uric acid level (0.041 (0.007, 0.075) ml/min/1.73 m^2^ per SD increase) increased annual eGFR decline, while educational attainment (−0.016 (−0.025, −0.007) ml/min/1.73 m^2^ per SD increase) decreased annual eGFR decline. Additionally, several risk factors had evidence for an effect on annual eGFR decline in the general population but not in the CKD population. These included chronic heart failure (0.062 (0.001, 0.124) ml/min/1.73 m^2^ per log-odds increase), hypertension (0.165 (0.026, 0.305) ml/min/1.73 m^2^ per log-odds increase), systolic blood pressure (SBP) (0.063 (0.025, 0.100) ml/min/1.73 m^2^ per SD increase), triglycerides (TG)(0.028 (0.003, 0.052) ml/min/1.73 m^2^ per SD increase), body fat percentage (0.058 (0.016, 0.099) ml/min/1.73 m^2^ per SD increase), and low TSH (0.014 (0.003, 0.024) ml/min/1.73 m^2^ per log-odds increase). Insulin-like growth factor 1 level (−0.028 (−0.050, − 0.006) ml/min/1.73 m^2^ per SD increase), high density lipoprotein cholesterol (HDL-C) (−0.028 (−0.051, −0.006) ml/min/1.73 m^2^ per SD increase), apolipoprotein A-I (−0.038 (−0.061, −0.014) ml/min/1.73 m^2^ per SD increase), and haemoglobin level (−0.050 (−0.077, −0.024) ml/min/1.73 m^2^ per SD increase) reduced annual eGFR decline in the general population but had limited evidence of an effect in the CKD population (**Figure 4**, **Supplementary Figure S3**, **Supplementary Table ST3**).

**Figure 4.**
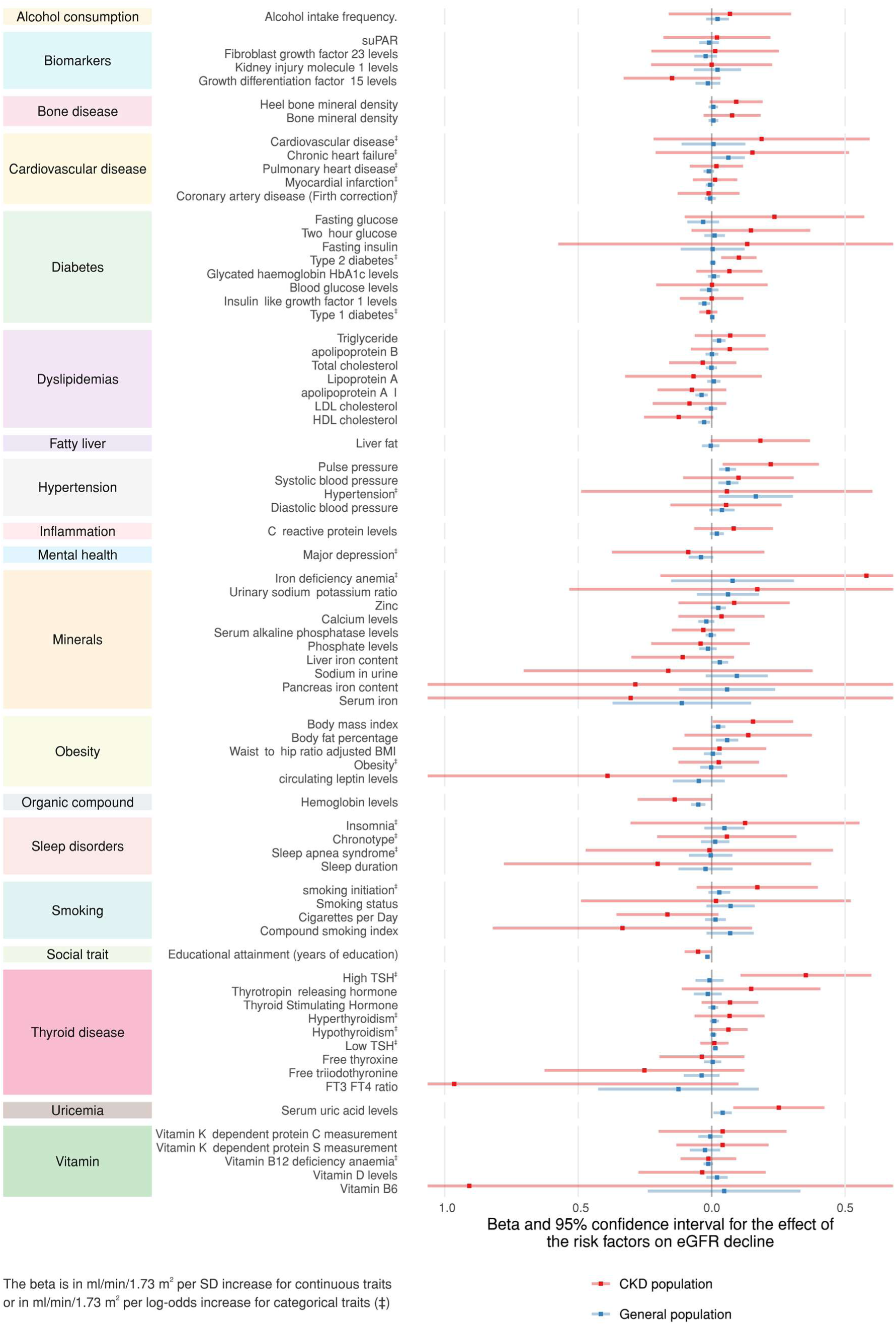
Mendelian randomisation estimates for the risk factors on the annual eGFR decline in the CKD and general populations

### Sensitivity analyses

The MR estimates using the simple mode, weighted median, weighted mode, and MR-Egger methods were consistent with the main analyses in both populations (**Supplementary Figure S4 and S5**, **Supplementary Table ST3**). The Q statistics suggested that heterogeneity may be present in 75 of 553 analyses (**Supplementary Table ST4**). Most of the analyses (510 of 528) had MR-Egger intercept p-value ≥ 0.05, suggesting little evidence of pleiotropy (**Supplementary Table ST5**). Most of the analyses (544 of 553) had *I*^2^ statistics over 0.9, which indicated these analyses had less than 10% bias toward the null (**Supplementary Table ST6**). Notably, chronic heart failure has low *I*^2^ statistics: 0.29 in the CKD population and 0.79 in incident CKD and the general population. Directionality tests suggested all risk factors have correct effect direction on annual eGFR decline, except for iron deficiency anaemia in the CKD population (**Supplementary Table ST7**). Steiger filtering removed iron deficiency anaemia risk factor in the annual eGFR decline for CKD population, due to only one SNP available. Several other risk factors were also attenuated after Steiger filtering (**Supplementary Figure S6**).

## Discussion

Using GWAS in the CKD and general populations from the CKDGen consortium, we performed two-sample MR to investigate the effect of putative risk factors for disease progression on kidney function decline. Using annual decline in eGFR as a measure of CKD progression, we found evidence that T2D and high TSH increase annual eGFR decline in the CKD population. The effect from these risk factors remained after adjusting for index event bias. Other risk factors identified in the CKD population, including BMI, PP and serum uric acid level, attenuated after applying index event bias correction.

In both CKD and general populations, PP and serum uric acid increased annual eGFR decline. Conversely, educational attainment, as measured by years spent in education, decreased annual eGFR decline in both populations. Some risk factors showed evidence of effects only in the general population but not in CKD population. For example, HDL-C on reduced annual eGFR decline in the general population only; while cardiovascular risk factors and blood pressure traits such as chronic heart failure, SBP, and hypertension increased annual decline in eGFR.

We found that T2D, a known CKD comorbidity, increased annual eGFR decline. A previous study shows that in the presence of T2D twice as many patients had eGFR less than 30 mL/min/1.73 m^2^ within 3 years of observation^32^, and a second study found that patients with T2D experienced eGFR decline twice as much as those without, leading to end-stage kidney disease and dialysis^33^. High TSH has also been reported as a risk factor for CKD in earlier studies^34–36^. In the current study, we found evidence for a causal role of this risk factor, with a possible mechanism through changes to kidney function and/or structure^35^.

We identified chronic heart failure, SBP, PP, and hypertension as risk factors for annual eGFR decline in the general population. These findings are in line with previous studies showing a detrimental effect of poor cardiovascular health and hypertension on the kidney^37–39^. An earlier MR study also reported the effect of SBP on decreased eGFR in the general population, consistent with our results^40,41^. While the effect from these risk factors (except PP) on eGFR remained inconclusive in our CKD population, the direction of effect matched the general population. Certain antihypertensives are known to benefit kidney function^9^. Our findings that increased blood pressure traits reduce kidney function serves as positive control, increasing the confidence in our analytical approach.

Finally, we report evidence to support a protective effect of HDL-C on eGFR decline in the general population that attenuated in the CKD population, especially after accounting for index event bias. The relationship between HDL-C and CKD is complex^42–44^. The present study indicates that HDL-C may play a role in development of the condition but not necessarily its progression.

Published MR studies on CKD have mainly focused on the effect of risk factors for incident disease and kidney function in the general population^43,45–47^. A strength of the current study is the consideration of risk factors on the annual eGFR decline in a CKD population, which is relevant to CKD progression and may inform CKD management. This was feasible due to the availability of a large-scale CKD incident and case-only GWAS from CKDGen. Moreover, advances in disease progression methods have provided tools to correct for index event bias in case-only GWAS^14,15^. Our approach is novel, as most earlier studies have instead used the associations identified for eGFR decline in CKD population to understand mechanisms and disease pathway for CKD and other chronic conditions^48–50^. We used the largest available GWAS summary statistics on risk factors from both OpenGWAS^21,22^ and international consortia, paired with the largest available GWAS to date on eGFR decline^16^, to ensure that our MR analyses were as well powered as possible.

Our study has several limitations. MR assumes the genetic instruments must 1) be associated with the exposure (relevance); 2) not be associated with unmeasured confounders between exposures and the outcome (independence); and 3) not affect the outcome other than via exposures (exclusion restriction). Additionally, assumption of homogeneity or monotonicity is required to obtain valid point estimates. The relevance assumption can be directly tested, and we have selected genome-wide significant SNPs and provided F-statistics to address this. However, the other assumptions are only falsifiable^17,19,51^. We have therefore provided several sensitivity analyses to support our inferences against these remaining assumptions^17^. The main challenge of MR in case-only settings is the potential for index event bias. To address this, we corrected the case-only GWAS to account for this type of bias prior to our MR analyses^14,15^. The magnitude of risk factor effects from MR reflects lifelong risk exposure to the risk factors which might be different than the effect estimates obtained from studies considering shorter timeframes of exposure^17^. Finally, CKD case-only GWAS has smaller sample size than eGFR decline GWAS in general population, which may be reflected on the effect differences between these populations.

In conclusion, we applied novel methods to correct for index event bias in case-only GWAS, leveraging recent large-scale GWAS of incident CKD and eGFR decline in CKD population, to estimate the causal effect of 74 risk factors on eGFR decline in CKD population using two-sample MR^16,23,24^. While several established cardiometabolic risk factors showed evidence consistent with kidney function decline, T2D and high TSH emerged as potential causal risk factors in CKD population after accounting for index event bias. These findings improve our understanding of CKD progression and may help priorities biological pathways and potential intervention targets to slow kidney function decline.

## Data availability statement

All data used in this study are publicly available. We accessed instruments for the risk factors from the OpenGWAS database, Open Access publications and GWAS consortia public data repositories (**Supplementary Table ST1**). The summary statistics for CKD incident and eGFR in the general and CKD populations were accessed from https://ckdgen.imbi.uni-freiburg.de/ and https://www.uni-regensburg.de/medizin/epidemiologie-praeventivmedizin/genetische-epidemiologie/gwas-summary-statistics/index.html.

## Data Availability

All data produced in the present study are available upon reasonable request to the authors

## Acknowledgements

We thank to Samarbeidsorganet, Helse Midt-Norge for research mobility grant to visit the Medical Research Council Integrative Epidemiology Unit at Bristol Medical School, University of Bristol.

## Contribution statement

AA and VW performed analysis and drafted the manuscript. All authors contributed to the design of the work, data interpretation, critical revision of the article, and final approval for publication.

## Ethical statement

This study used publicly available data and so did not require study-specific ethical approval.

### Box 1. Illustration of potential for index event bias in this study

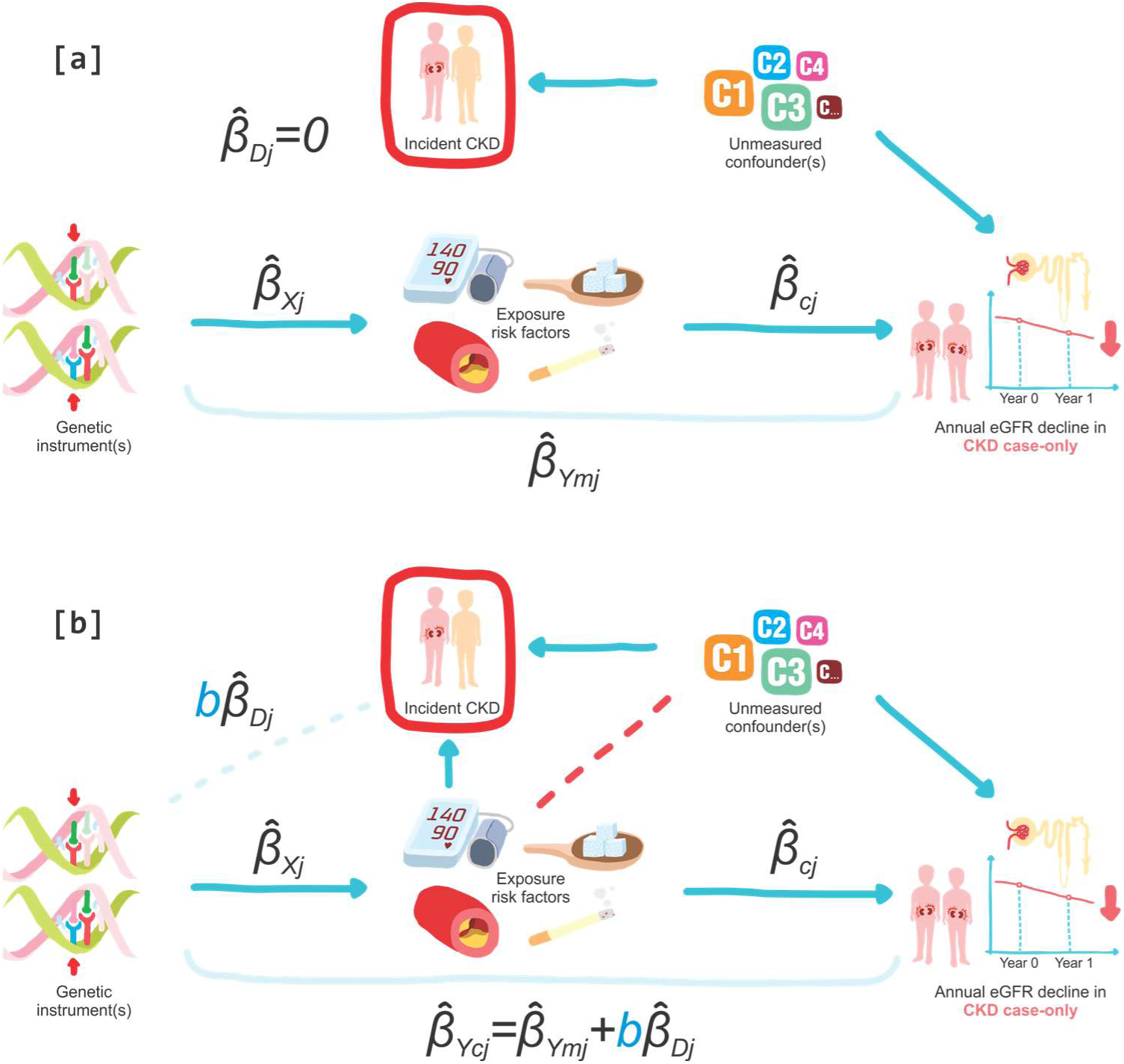

The solid blue lines indicate assume relationship, the faint blue line/dots represent association, the red-stripped line indicates induced association. The absent of red-stripped line shows that induced association is unlikely. The red boxes illustrate conditioned variable in CKD case-only analysis. eGFR, estimated glomerulus filtration rate; CKD, chronic kidney disease; C1, C2, C…, illustrates unmeasured confounder(s) relevant to each exposure risk factor. In **scenario [a]**, risk factors have no causal effect on the CKD incidence (*β̂_Dj_* = 0), thus index even bias is not expected. In this scenario, the exposure risk factors on the outcome of annual eGFR decline in CKD population is not bias (*β̂_Ymj_* = *β̂_cj_β̂_Xj_* + *ε*_j_). In **scenario [b]**, risk factors are likely to be the cause of CKD incidence when conditioning on CKD incidence (*bβ̂_Dj_* ≠ 0). Hence, there is a potential of index event bias (a collider bias) in this scenario and the genetic association with the outcome is *β̂_Ycj_* = *β̂_Ymj_* + *bβ̂_Dj_* . Such bias induces an effect of risk factors on the outcome (*β̂_cj_*) of annual eGFR decline in CKD population, where such effect should not have existed had there was no causal relationship between risk factors and CKD incidence. The Dudbridge^14^ and SlopeHunter^15^ methods estimate the constant *b* to correct for genetic association with the outcome. Hence, the association between risk factors and the outcome is adjusted for index event bias.

