## Supplementary Figures for "Disease progression Mendelian randomization of risk factors for kidney function decline in chronic kidney disease and the general population"

**Figure S1.** Mendelian randomisation estimates for the risk factors on the annual eGFR decline for CKD population, without and with adjusted estimates for continuous (a) and categorical (b) risk factors

(a)

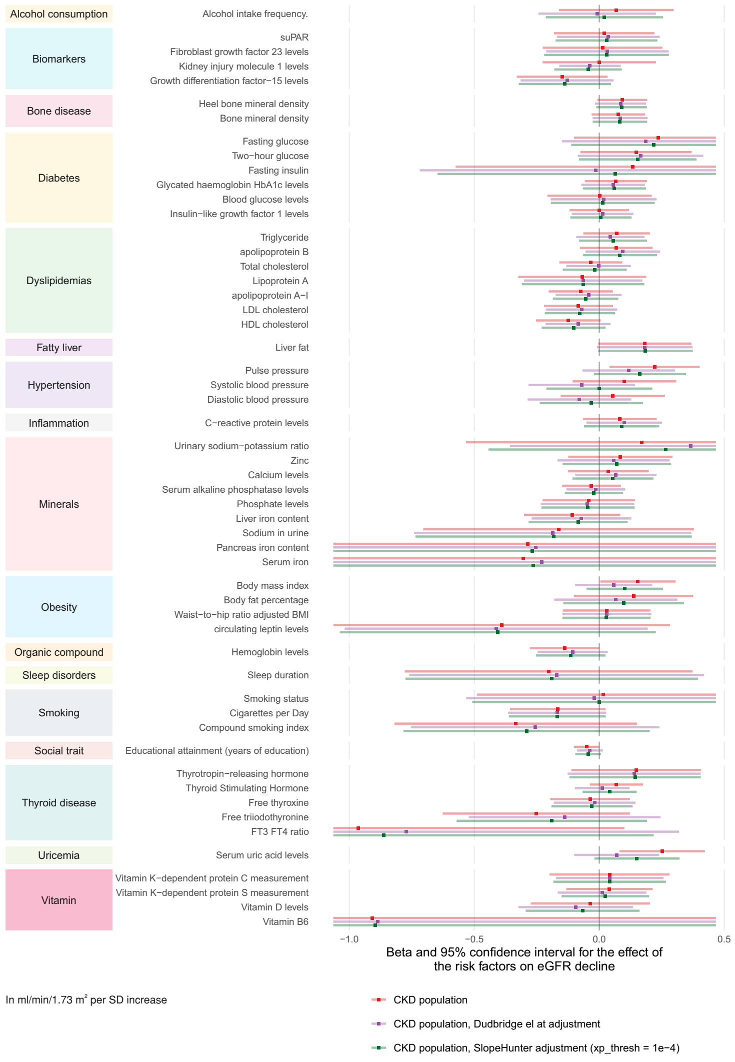


(b)

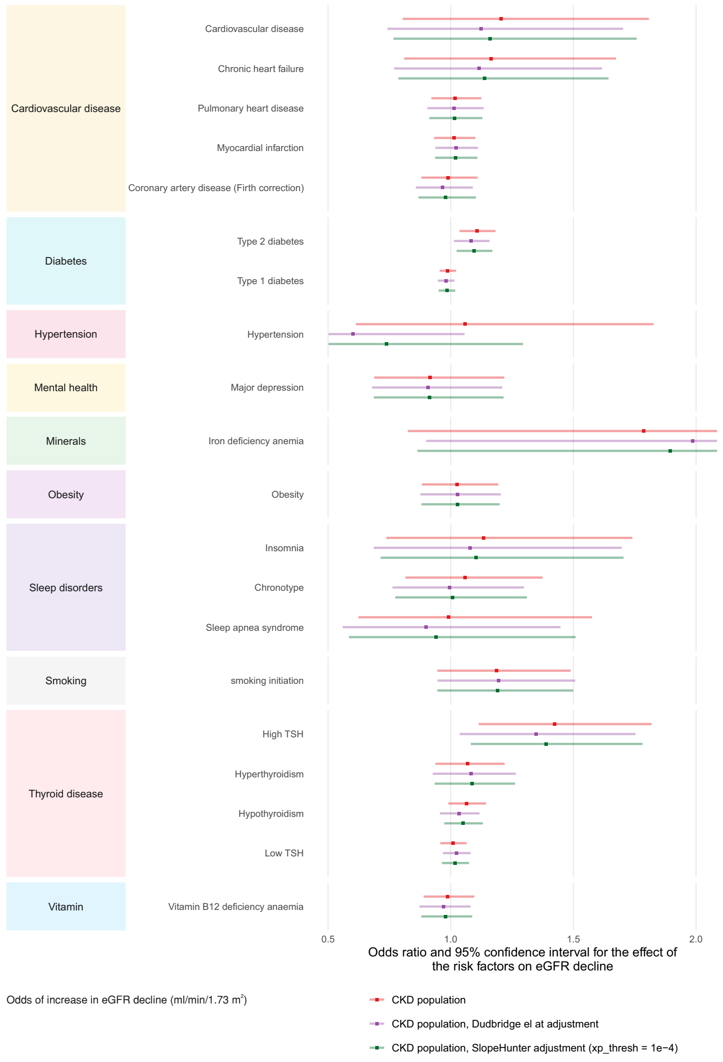

**Figure S2.** Mendelian randomisation estimates for the risk factors on the annual eGFR decline for CKD population, without adjustment and with adjustment using SlopeHunter method adjustment at various P-value thresholds for SNP-incidence associations

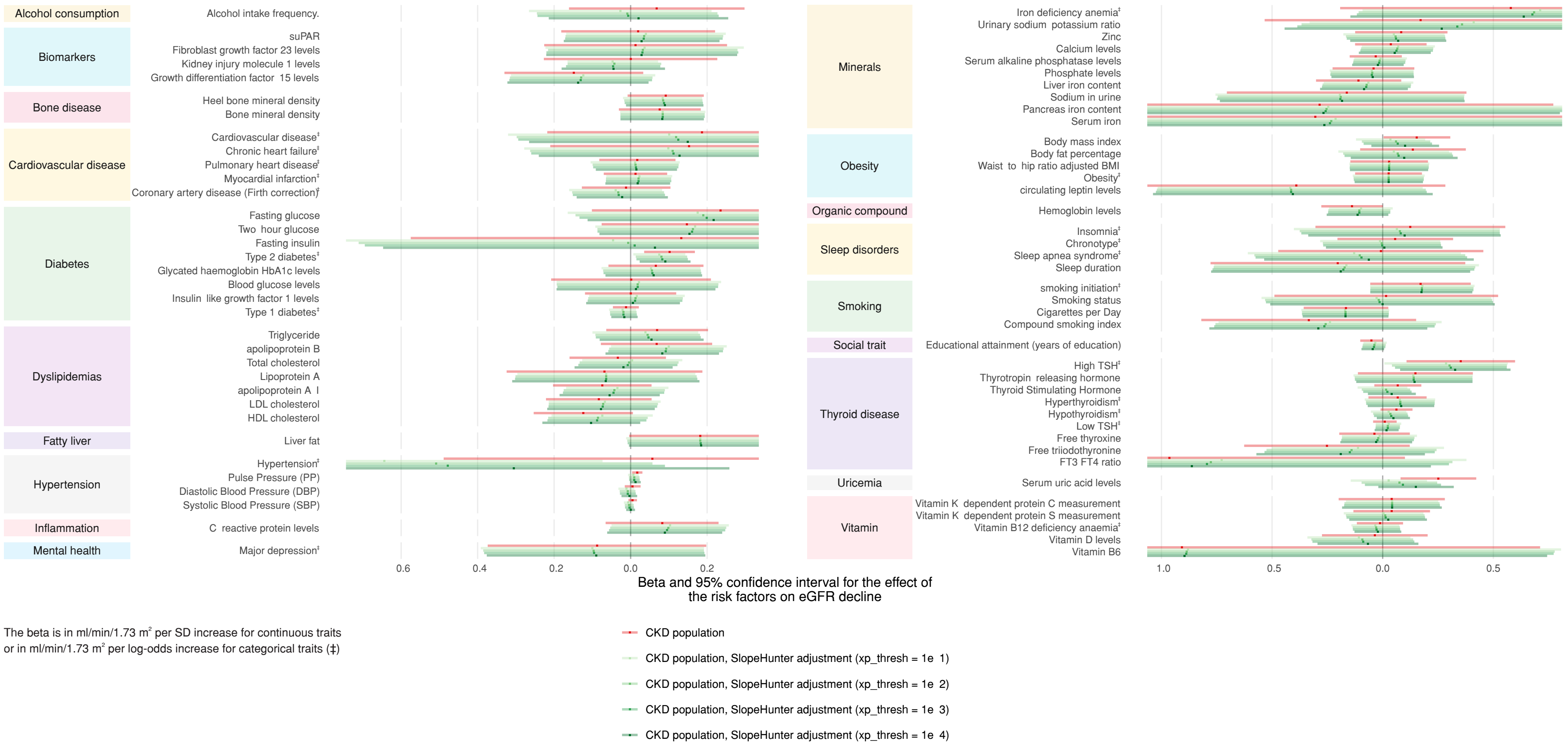

**Figure S3.** Mendelian randomisation estimates for the risk factors on the annual eGFR decline for CKD and general populations for continuous (a) and categorical (b) risk factors

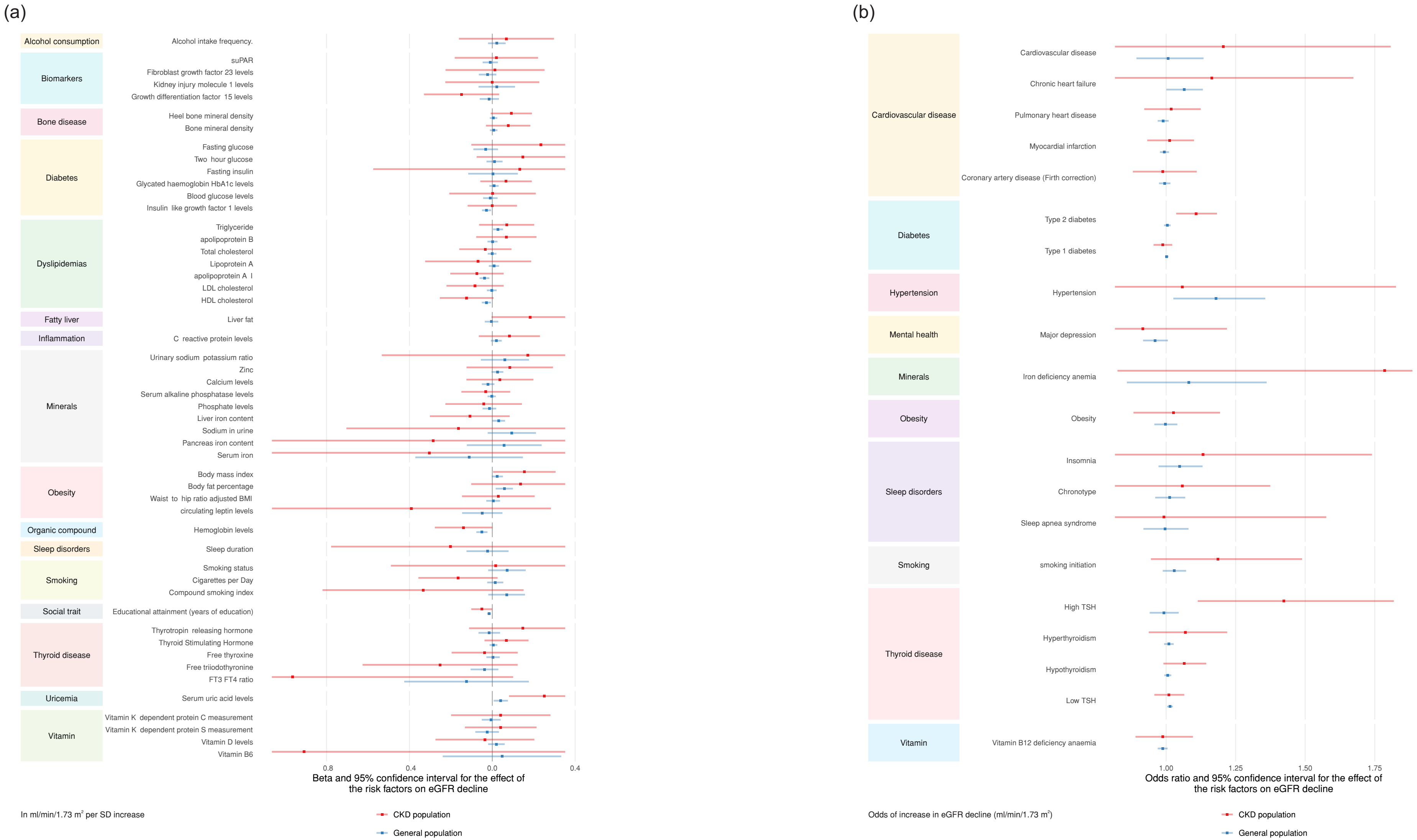

**Figure S4.** Comparison of estimates on annual eGFR decline in the CKD population from main analysis and sensitivity analyses (MR Egger, simple mode, weighted median, and weighted mode)

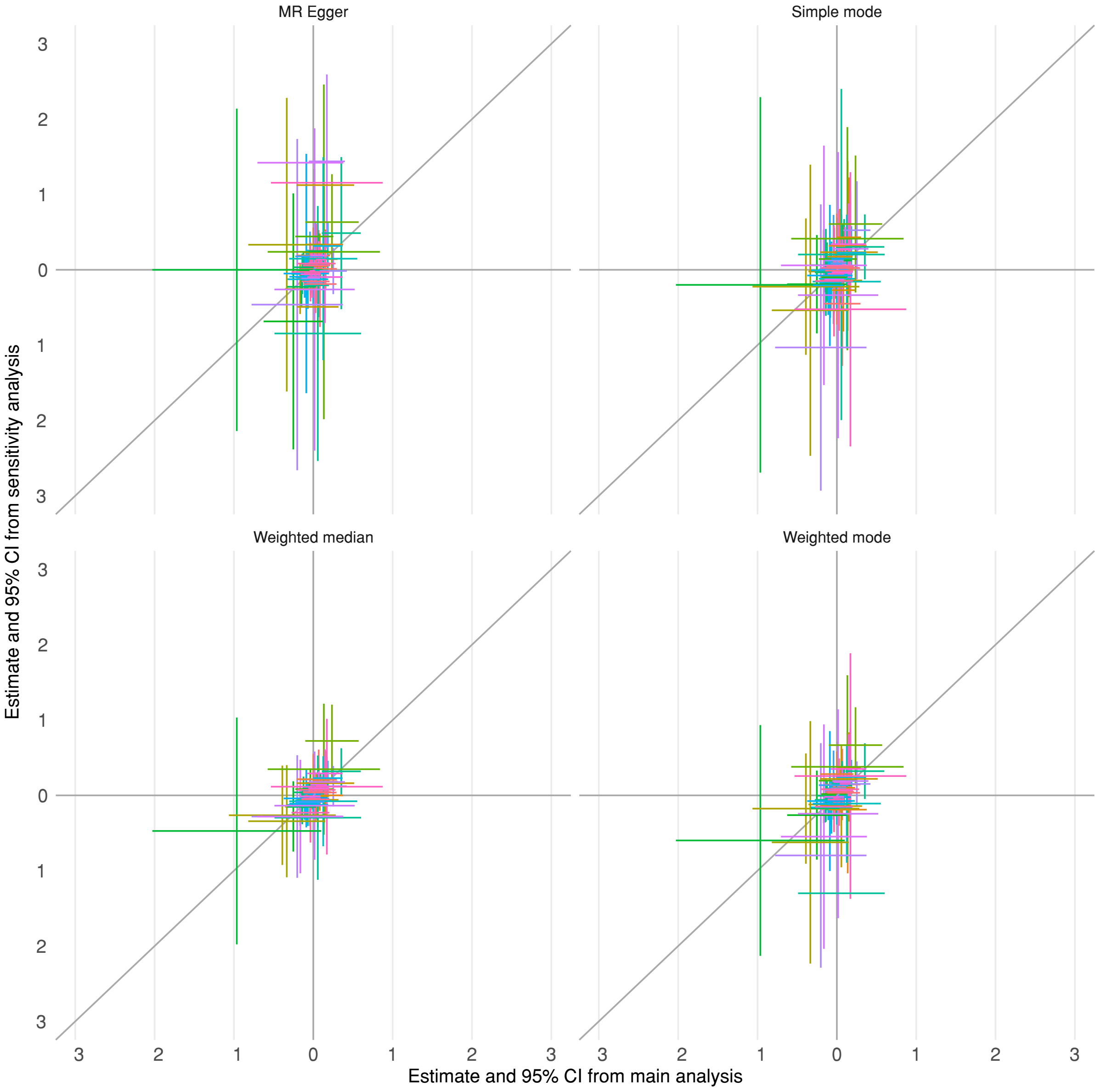

**Figure S5.** Comparison of estimates on annual eGFR decline in the general population from main analysis and sensitivity analyses (MR Egger, simple mode, weighted median, and weighted mode)

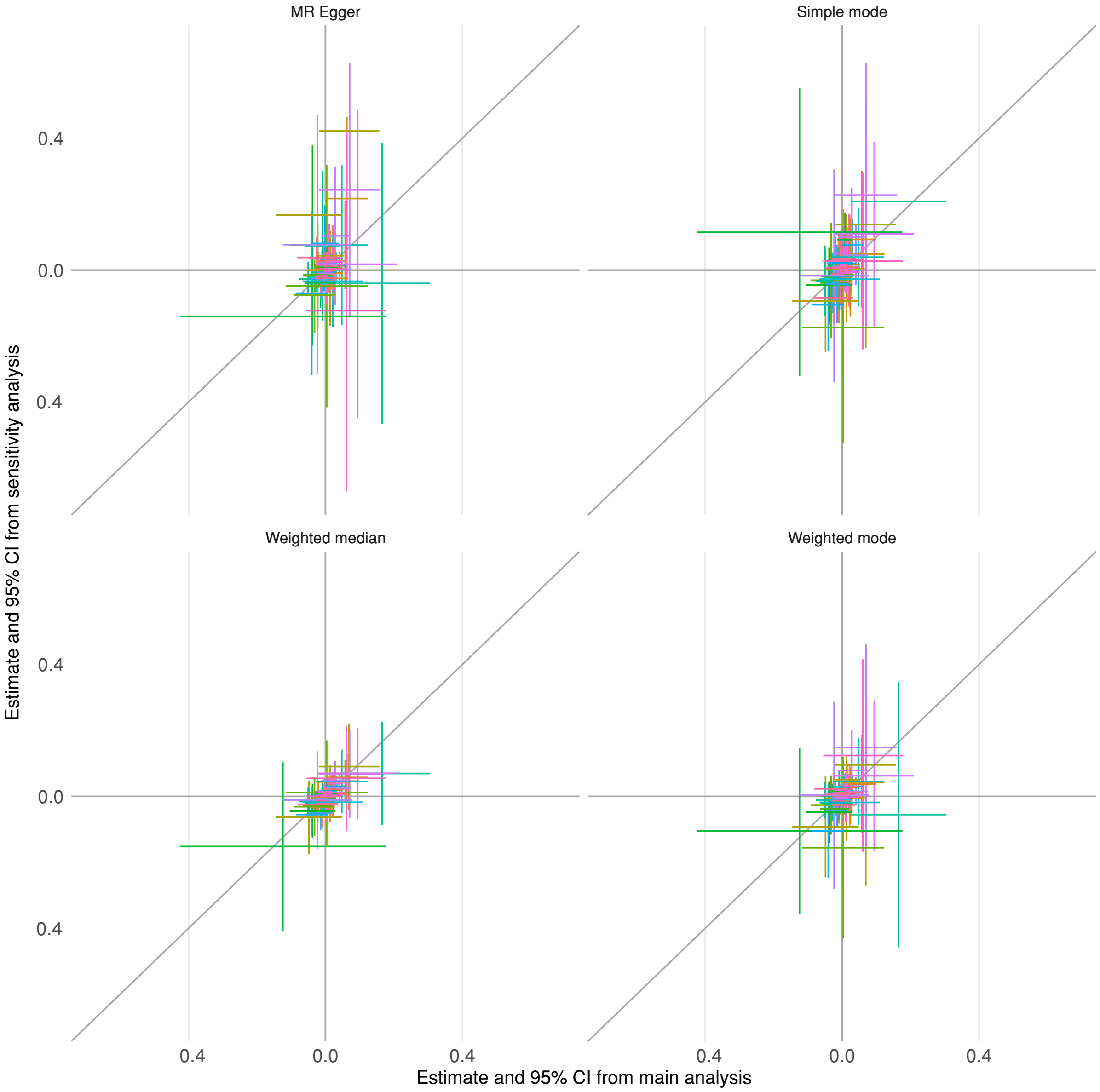

**Figure S6.** Mendelian randomisation estimates for the risk factors on the annual eGFR decline for CKD and general populations, after Steiger filtering

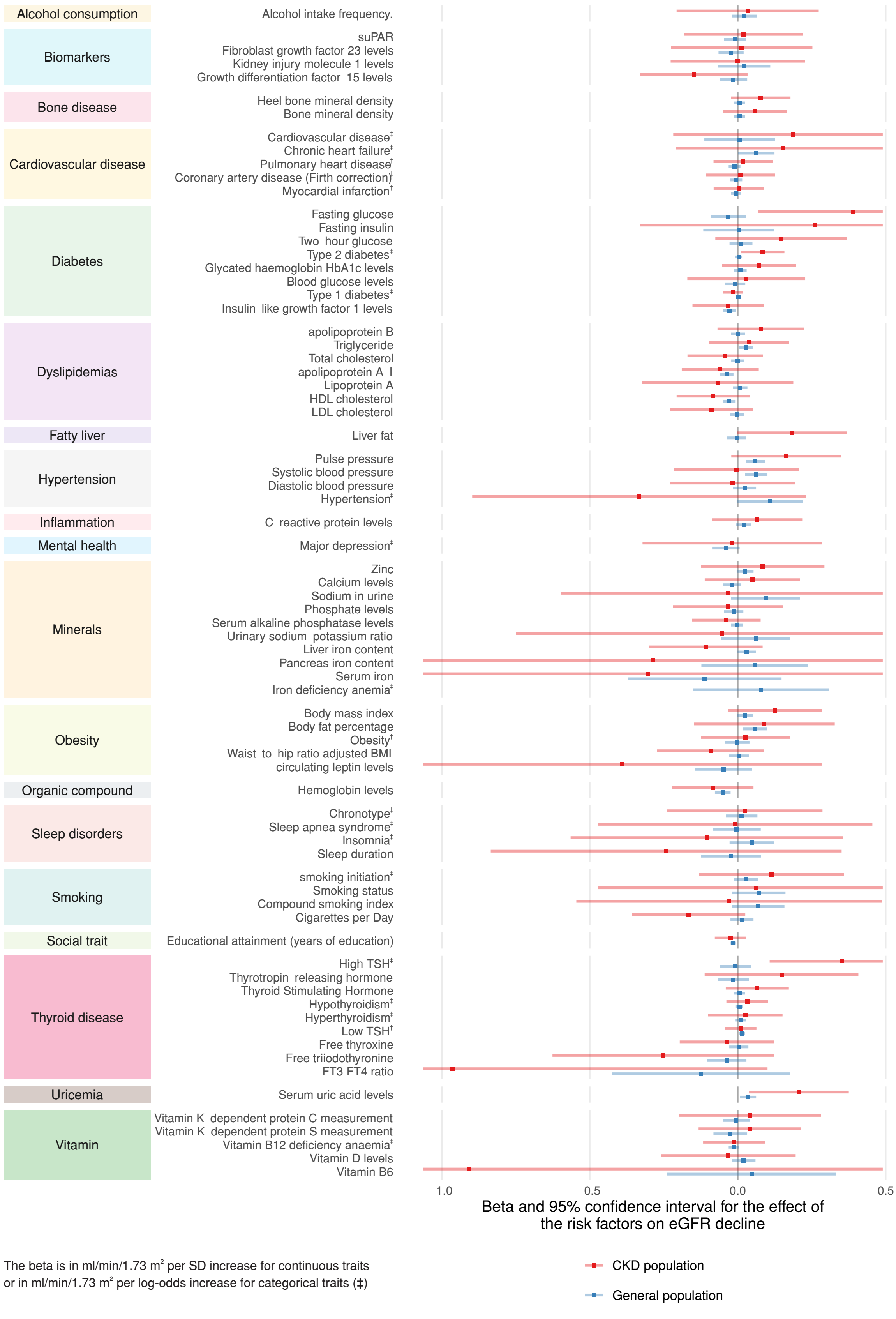


The beta is in ml/min/1.73 m² per SD increase for continuous traits or in ml/min/1.73 m² per log-odds increase for categorical traits (‡)
