## Supplementary Tables for "Disease progression Mendelian randomization of risk factors for kidney function decline in chronic kidney disease and the general population"

| ID | Trait | PhenotypeGroup | nSNP | N | Author | PMID | Link | Unit |
|---|---|---|---|---|---|---|---|---|
| ukb-b-5779 | Alcohol intake frequency. | Alcohol consumption | 9851867 | 462346 | Ben Elsworth | | | SD |
| ebi-a-GCST90012022 | Fibroblast growth factor 23 levels | Biomarkers | 13102275 | 21758 | Folkersen L | 33067605 | | SD |
| ebi-a-GCST90011998 | Growth differentiation factor-15 levels | Biomarkers | 13138240 | 21758 | Folkersen L | 33067605 | | SD |
| ebi-a-GCST90012041 | Kidney injury molecule 1 levels | Biomarkers | 13103458 | 21758 | Folkersen L | 33067605 | | SD |
| prot-c-2652_15_1 | suPAR | Biomarkers | 501428 | 997 | Suhre K | 28240269 | | SD |
| ebi-a-GCST90014022 | Bone mineral density | Bone disease | 10783906 | 365403 | Mbatchou J | 34017140 | | SD |
| ebi-a-GCST006979-consort | Heel bone mineral density | Bone disease | 13705641 | 426824 | | | | SD |
| ebi-a-GCST90086057 | Cardiovascular disease | Cardiovascular disease | 14485079 | 56637 | Guindo-Mart<U+00ED>nez M | 33893285 | | Log-odds |
| ebi-a-GCST90018806 | Chronic heart failure | Cardiovascular disease | 24178220 | 486160 | Sakaue S | 34594039 | | Log-odds |
| ebi-a-GCST90013864 | Coronary artery disease (Firth correction) | Cardiovascular disease | 11027870 | 352063 | Mbatchou J | 34017140 | | Log-odds |
| ebi-a-GCST90018877 | Myocardial infarction | Cardiovascular disease | 24172914 | 461823 | Sakaue S | 34594039 | | Log-odds |
| finn-b-FG_PULMHEART | Pulmonary heart disease | Cardiovascular disease | 16380466 | 218792 | | | | Log-odds |
| ebi-a-GCST90025986 | Blood glucose levels | Diabetes | 4218897 | 400458 | Barton AR | 34226706 | | SD |
| ebi-a-GCST90002232 | Fasting glucose | Diabetes | 31008728 | 200622 | Chen J | 34059833 | | SD |
| ebi-a-GCST90002238 | Fasting insulin | Diabetes | 29664438 | 151013 | Chen J | 34059833 | | SD |
| ebi-a-GCST90014006 | Glycated haemoglobin HbA1c levels | Diabetes | 10783722 | 389889 | Mbatchou J | 34017140 | | SD |
| ebi-a-GCST90025989 | Insulin-like growth factor 1 levels | Diabetes | 4231359 | 435516 | Barton AR | 34226706 | | SD |
| ebi-a-GCST90002227 | Two-hour glucose | Diabetes | 27330879 | 63396 | Chen J | 34059833 | | SD |
| ebi-a-GCST90014023 | Type 1 diabetes | Diabetes | 59999551 | 520580 | Chiou J | 34012112 | | Log-odds |
| T2DGGI | Type 2 diabetes | Diabetes | 19305792 | 1812013 | | | https://dia | Log-odds |
| HDL | HDL cholesterol | Dyslipidemias | 35200666 | 888227 | | | https://cs | SD |
| LDL | LDL cholesterol | Dyslipidemias | 34553200 | 842660 | | | https://cs | SD |
| ukb-d-30790_irnt | Lipoprotein A | Dyslipidemias | 13583854 | 273896 | Neale lab | | | SD |
| TC | Total cholesterol | Dyslipidemias | 35472026 | 930672 | | | https://cs | SD |
| TG | Triglyceride | Dyslipidemias | 34673395 | 864240 | | | https://cs | SD |
| ieu-b-107 | apolipoprotein A-I | Dyslipidemias | 12321875 | 393193 | Richardson, Tom | 32203549 | | SD |
| ieu-b-108 | apolipoprotein B | Dyslipidemias | 12321875 | 439214 | Richardson, Tom | 32203549 | | SD |
| ebi-a-GCST90029073 | Liver fat | Fatty liver | 9499333 | 32974 | Haas ME | 34957434 | | SD |
| DBPTang | Diastolic Blood Pressure (DBP) | Hypertension | 12321875 | 436083 | Tang H | | | SD |
| ieu-b-5144 | Hypertension | Hypertension | 12321875 | 462826 | Tang H | | | Log-odds |
| PPTang | Pulse Pressure (PP) | Hypertension | 12321875 | 435478 | Tang H | | | SD |
| SBPTang | Systolic Blood Pressure (SBP) | Hypertension | 12321875 | 435859 | Tang H | | | SD |
| ebi-a-GCST90029070 | C-reactive protein levels | Inflammation | 10713245 | 575531 | Said S | 35459240 | | SD |
| ieu-b-102 | Major depression | Mental health | | 500199 | Howard DM | 30718901 | | Log-odds |
| ebi-a-GCST90025990 | Calcium levels | Minerals | 4218949 | 400792 | Barton AR | 34226706 | | SD |
| ebi-a-GCST90018872 | Iron deficiency anemia | Minerals | 24180477 | 480941 | Sakaue S | 34594039 | | Log-odds |
| ebi-a-GCST90016674 | Liver iron content | Minerals | 9275407 | 32858 | Liu Y | 34128465 | | SD |
| ebi-a-GCST90016676 | Pancreas iron content | Minerals | 9275407 | 25617 | Liu Y | 34128465 | | SD |
| ebi-a-GCST90014010 | Phosphate levels | Minerals | 10783661 | 357302 | Mbatchou J | 34017140 | | SD |
| ebi-a-GCST90025947 | Serum alkaline phosphatase levels | Minerals | 4232004 | 437896 | Barton AR | 34226706 | | SD |
| Fe | Serum iron | Minerals | 21703644 | 236612 | | | | SD |
| ebi-a-GCST90013989 | Sodium in urine | Minerals | 10783698 | 396020 | Mbatchou J | 34017140 | | SD |
| ieu-b-72 | Urinary sodium-potassium ratio | Minerals | 19420026 | 326938 | Zanetti | 32008434 | | SD |
| Zn | Zinc | Minerals | 13354620 | 2819 | | | | SD |
| ebi-a-GCST90013975 | Body fat percentage | Obesity | 10783701 | 401772 | Mbatchou J | 34017140 | | SD |
| BMI | Body mass index | Obesity | 2529253 | 806834 | | | https://gia | SD |
| finn-b-E4_OBESITY | Obesity | Obesity | 16380465 | 218735 | | | | Log-odds |
| WHR | Waist-to-hip ratio adjusted BMI | Obesity | 2516890 | 694649 | | | https://gia | SD |
| ebi-a-GCST90007310 | circulating leptin levels | Obesity | 203423 | 49909 | Yaghootkar H | 32917775 | | SD |
| ebi-a-GCST90025969 | Hemoglobin levels | Organic compound | 4234826 | 445373 | Barton AR | 34226706 | | SD |
| ieu-b-4861 | Chronotype | Sleep disorders | 11979082 | 244207 | Samuel Jones | | | Log-odds |
| Insom | Insomnia | Sleep disorders | 12655095 | 386988 | | | https://vu | Log-odds |
| ebi-a-GCST90018916 | Sleep apnea syndrome | Sleep disorders | 24183940 | 476853 | Sakaue S | 34594039 | | Log-odds |
| ukb-b-4424 | Sleep duration | Sleep disorders | 9851867 | 460099 | Ben Elsworth | | | SD |
| ieu-b-25 | Cigarettes per Day | Smoking | 11913712 | 337334 | Liu, M | 30643251 | | SD |
| SIndex | Compound smoking index | Smoking | 7679983 | 462690 | | | https://da | SD |
| ebi-a-GCST90029014 | Smoking status | Smoking | 11973425 | 468170 | Loh PR | 29892013 | | SD |
| ieu-b-4877 | smoking initiation | Smoking | 11802365 | 607291 | Liu M | 30643251 | | Log-odds |
| ebi-a-GCST90029013 | Educational attainment (years of education) | Social trait | 11972619 | 461457 | Loh PR | 29892013 | | SD |
| FT3FT4 | FT3 FT4 ratio | Thyroid disease | 7771185 | 51095 | | | https://ge | SD |
| FT4 | Free thyroxine | Thyroid disease | 7815340 | 119120 | | | https://ge | SD |
| FT3 | Free triiodothyronine | Thyroid disease | 7905034 | 59061 | | | https://ge | SD |
| HiTSH | High TSH | Thyroid disease | 8101931 | 153241 | | | https://ge | Log-odds |
| ebi-a-GCST90018860 | Hyperthyroidism | Thyroid disease | 24189279 | 460499 | Sakaue S | 34594039 | | Log-odds |
| ebi-a-GCST90018862 | Hypothyroidism | Thyroid disease | 24138872 | 410141 | Sakaue S | 34594039 | | Log-odds |
| LoTSH | Low TSH | Thyroid disease | 8178699 | 141549 | | | https://ge | Log-odds |
| TSH | Thyroid Stimulating Hormone | Thyroid disease | 7987300 | 271040 | | | https://ge | SD |
| prot-a-3102 | Thyrotropin-releasing hormone | Thyroid disease | 10534735 | 3301 | Sun BB | 29875488 | | SD |
| ebi-a-GCST90018977 | Serum uric acid levels | Uricemia | 19041286 | 343836 | Sakaue S | 34594039 | | SD |
| finn-b-D3_ANAEMIA_B12_DEF | Vitamin B12 deficiency anaemia | Vitamin | 16380452 | 212822 | | | | Log-odds |
| ukb-b-7864 | Vitamin B6 | Vitamin | 9851867 | 64979 | Ben Elsworth | | | SD |
| ebi-a-GCST90025967 | Vitamin D levels | Vitamin | 4225238 | 418691 | Barton AR | 34226706 | | SD |
| ebi-a-GCST90019423 | Vitamin K-dependent protein C measurement | Vitamin | 15567760 | 10708 | Pietzner M | 33328453 | | SD |
| ebi-a-GCST90019417 | Vitamin K-dependent protein S measurement | Vitamin | 15567420 | 10708 | Pietzner M | 33328453 | | SD |

Summary of Genetic Correlation Results

| p1 | p2 | rg | se | z | p | h2_obs | h2_obs_se | h2_int | h2_int_se | gcov_int | gcov_int_se |
|---|---|---|---|---|---|---|---|---|---|---|---|
| incidence.sumstats.gz | ckdprognosis.sumstats.gz | 0,5721 | 0,1707 | 3,3511 | 0,0008 | 0,0381 | 0,0157 | 0,9395 | 0,0061 | 0,0174 | 0,0051 |
| incidence.sumstats.gz | genprognosis.sumstats.gz | 1,6616 | 0,5312 | 3,1278 | 0,0018 | 0,0158 | 0,01 | 0,9873 | 0,0068 | 0,0874 | 0,0059 |
| genprognosis.sumstats.gz | ckdprognosis.sumstats.gz | 1,6612 | 0,6452 | 2,5749 | 0,01 | 0,0396 | 0,0164 | 0,939 | 0,0063 | 0,0555 | 0,0043 |

| id | outcome | exposure | method | nsnp | b | se | pval | PhenotypeGroup | MeanFstat |
|---|---|---|---|---|---|---|---|---|---|
| BMI | Incident CKD | Body mass index | MR Egger | 506 | 0,110491 | 0,088677 | 0,213343 | Obesity | 74,21321 |
| BMI | Incident CKD | Body mass index | Weighted median | 506 | 0,194893 | 0,054404 | 0,000341 | Obesity | 74,21321 |
| BMI | Incident CKD | Body mass index | Inverse variance weighted | 506 | 0,157605 | 0,0341 | 3,80E-06 | Obesity | 74,21321 |
| BMI | Incident CKD | Body mass index | Simple mode | 506 | 0,151464 | 0,177315 | 0,393393 | Obesity | 74,21321 |
| BMI | Incident CKD | Body mass index | Weighted mode | 506 | 0,167259 | 0,092688 | 0,071741 | Obesity | 74,21321 |
| DBPTang | Incident CKD | Diastolic blood pressure | MR Egger | 259 | 0,070876 | 0,216825 | 0,744026 | Hypertension | 55,78276 |
| DBPTang | Incident CKD | Diastolic blood pressure | Weighted median | 259 | 0,142261 | 0,064694 | 0,02788 | Hypertension | 55,78276 |
| DBPTang | Incident CKD | Diastolic blood pressure | Inverse variance weighted | 259 | 0,225367 | 0,070141 | 0,001313 | Hypertension | 55,78276 |
| DBPTang | Incident CKD | Diastolic blood pressure | Simple mode | 259 | 0,017625 | 0,207472 | 0,932366 | Hypertension | 55,78276 |
| DBPTang | Incident CKD | Diastolic blood pressure | Weighted mode | 259 | 0,045971 | 0,173168 | 0,790861 | Hypertension | 55,78276 |
| FT3 | Incident CKD | Free triiodothyronine | MR Egger | 6 | -0,40934 | 0,325642 | 0,277136 | Thyroid disease | 82,22035 |
| FT3 | Incident CKD | Free triiodothyronine | Weighted median | 6 | -0,20513 | 0,091737 | 0,02535 | Thyroid disease | 82,22035 |
| FT3 | Incident CKD | Free triiodothyronine | Inverse variance weighted | 6 | -0,19064 | 0,071753 | 0,007887 | Thyroid disease | 82,22035 |
| FT3 | Incident CKD | Free triiodothyronine | Simple mode | 6 | -0,31759 | 0,140326 | 0,073056 | Thyroid disease | 82,22035 |
| FT3 | Incident CKD | Free triiodothyronine | Weighted mode | 6 | -0,23821 | 0,112624 | 0,088049 | Thyroid disease | 82,22035 |
| FT3FT4 | Incident CKD | FT3 FT4 ratio | MR Egger | 13 | -0,62047 | 0,409604 | 0,158016 | Thyroid disease | 132,3008 |
| FT3FT4 | Incident CKD | FT3 FT4 ratio | Weighted median | 13 | -0,44014 | 0,27279 | 0,10664 | Thyroid disease | 132,3008 |
| FT3FT4 | Incident CKD | FT3 FT4 ratio | Inverse variance weighted | 13 | -0,32057 | 0,204716 | 0,117361 | Thyroid disease | 132,3008 |
| FT3FT4 | Incident CKD | FT3 FT4 ratio | Simple mode | 13 | -0,5366 | 0,455467 | 0,261583 | Thyroid disease | 132,3008 |
| FT3FT4 | Incident CKD | FT3 FT4 ratio | Weighted mode | 13 | -0,47208 | 0,284072 | 0,122428 | Thyroid disease | 132,3008 |
| FT4 | Incident CKD | Free thyroxine | MR Egger | 63 | 0,103965 | 0,094563 | 0,2759 | Thyroid disease | 83,6751 |
| FT4 | Incident CKD | Free thyroxine | Weighted median | 63 | 0,087293 | 0,04887 | 0,074065 | Thyroid disease | 83,6751 |
| FT4 | Incident CKD | Free thyroxine | Inverse variance weighted | 63 | -0,03186 | 0,046033 | 0,488825 | Thyroid disease | 83,6751 |
| FT4 | Incident CKD | Free thyroxine | Simple mode | 63 | 0,08351 | 0,121273 | 0,493633 | Thyroid disease | 83,6751 |
| FT4 | Incident CKD | Free thyroxine | Weighted mode | 63 | 0,091417 | 0,057209 | 0,115139 | Thyroid disease | 83,6751 |
| Fe | Incident CKD | Serum iron | Inverse variance weighted | 2 | -0,12178 | 0,226406 | 0,590645 | Minerals | 291,9629 |
| HDL | Incident CKD | HDL cholesterol | MR Egger | 249 | 0,026328 | 0,053042 | 0,620075 | Dyslipidemias | 269,7662 |
| HDL | Incident CKD | HDL cholesterol | Weighted median | 249 | -0,01979 | 0,041784 | 0,635748 | Dyslipidemias | 269,7662 |
| HDL | Incident CKD | HDL cholesterol | Inverse variance weighted | 249 | -0,06359 | 0,035137 | 0,070316 | Dyslipidemias | 269,7662 |
| HDL | Incident CKD | HDL cholesterol | Simple mode | 249 | -0,01442 | 0,090318 | 0,873321 | Dyslipidemias | 269,7662 |
| HDL | Incident CKD | HDL cholesterol | Weighted mode | 249 | 0,017079 | 0,036842 | 0,643363 | Dyslipidemias | 269,7662 |
| HiTSH | Incident CKD | High TSH | MR Egger | 6 | -0,27327 | 0,277642 | 0,380708 | Thyroid disease | 49,53497 |
| HiTSH | Incident CKD | High TSH | Weighted median | 6 | 0,042501 | 0,060626 | 0,48328 | Thyroid disease | 49,53497 |
| HiTSH | Incident CKD | High TSH | Inverse variance weighted | 6 | 0,091217 | 0,072811 | 0,21028 | Thyroid disease | 49,53497 |
| HiTSH | Incident CKD | High TSH | Simple mode | 6 | 0,055978 | 0,076098 | 0,49501 | Thyroid disease | 49,53497 |
| HiTSH | Incident CKD | High TSH | Weighted mode | 6 | 0,035735 | 0,068443 | 0,623888 | Thyroid disease | 49,53497 |
| Insom | Incident CKD | Insomnia | MR Egger | 13 | 0,218862 | 0,252778 | 0,405075 | Sleep disorders | 44,05724 |
| Insom | Incident CKD | Insomnia | Weighted median | 13 | 0,096508 | 0,113004 | 0,393091 | Sleep disorders | 44,05724 |
| Insom | Incident CKD | Insomnia | Inverse variance weighted | 13 | 0,08519 | 0,081757 | 0,297415 | Sleep disorders | 44,05724 |
| Insom | Incident CKD | Insomnia | Simple mode | 13 | 0,069335 | 0,194188 | 0,727252 | Sleep disorders | 44,05724 |
| Insom | Incident CKD | Insomnia | Weighted mode | 13 | 0,140625 | 0,153918 | 0,378902 | Sleep disorders | 44,05724 |
| LDL | Incident CKD | LDL cholesterol | MR Egger | 228 | 0,004239 | 0,053436 | 0,936839 | Dyslipidemias | 240,9604 |
| LDL | Incident CKD | LDL cholesterol | Weighted median | 228 | -0,06606 | 0,042869 | 0,123297 | Dyslipidemias | 240,9604 |
| LDL | Incident CKD | LDL cholesterol | Inverse variance weighted | 228 | -0,02909 | 0,035263 | 0,409473 | Dyslipidemias | 240,9604 |
| LDL | Incident CKD | LDL cholesterol | Simple mode | 228 | -0,02275 | 0,100694 | 0,821431 | Dyslipidemias | 240,9604 |
| LDL | Incident CKD | LDL cholesterol | Weighted mode | 228 | -0,06468 | 0,04319 | 0,135623 | Dyslipidemias | 240,9604 |
| LoTSH | Incident CKD | Low TSH | MR Egger | 28 | -0,05271 | 0,058831 | 0,378453 | Thyroid disease | 72,70639 |
| LoTSH | Incident CKD | Low TSH | Weighted median | 28 | -0,02606 | 0,015417 | 0,090998 | Thyroid disease | 72,70639 |
| LoTSH | Incident CKD | Low TSH | Inverse variance weighted | 28 | -0,02036 | 0,022219 | 0,359507 | Thyroid disease | 72,70639 |
| LoTSH | Incident CKD | Low TSH | Simple mode | 28 | -0,02896 | 0,024499 | 0,247471 | Thyroid disease | 72,70639 |
| LoTSH | Incident CKD | Low TSH | Weighted mode | 28 | -0,02676 | 0,018479 | 0,159022 | Thyroid disease | 72,70639 |
| PPTang | Incident CKD | Pulse pressure | MR Egger | 308 | 0,323557 | 0,134666 | 0,016872 | Hypertension | 68,29648 |
| PPTang | Incident CKD | Pulse pressure | Weighted median | 308 | 0,221795 | 0,057488 | 0,000114 | Hypertension | 68,29648 |
| PPTang | Incident CKD | Pulse pressure | Inverse variance weighted | 308 | 0,168646 | 0,051619 | 0,001086 | Hypertension | 68,29648 |
| PPTang | Incident CKD | Pulse pressure | Simple mode | 308 | -0,10596 | 0,196003 | 0,589163 | Hypertension | 68,29648 |
| PPTang | Incident CKD | Pulse pressure | Weighted mode | 308 | 0,397703 | 0,139813 | 0,004746 | Hypertension | 68,29648 |
| SBPTang | Incident CKD | Systolic blood pressure | MR Egger | 257 | 0,108971 | 0,185436 | 0,557288 | Hypertension | 56,83473 |
| SBPTang | Incident CKD | Systolic blood pressure | Weighted median | 257 | 0,231645 | 0,066746 | 0,000519 | Hypertension | 56,83473 |
| SBPTang | Incident CKD | Systolic blood pressure | Inverse variance weighted | 257 | 0,267095 | 0,061345 | 1,34E-05 | Hypertension | 56,83473 |
| SBPTang | Incident CKD | Systolic blood pressure | Simple mode | 257 | 0,237897 | 0,212154 | 0,263192 | Hypertension | 56,83473 |
| SBPTang | Incident CKD | Systolic blood pressure | Weighted mode | 257 | 0,237897 | 0,180192 | 0,187934 | Hypertension | 56,83473 |
| SIndex | Incident CKD | Compound smoking index | MR Egger | 121 | 0,096416 | 0,393954 | 0,80708 | Smoking | 44,08236 |
| SIndex | Incident CKD | Compound smoking index | Weighted median | 121 | -0,16949 | 0,137827 | 0,218798 | Smoking | 44,08236 |
| SIndex | Incident CKD | Compound smoking index | Inverse variance weighted | 121 | -0,137 | 0,098841 | 0,165736 | Smoking | 44,08236 |
| SIndex | Incident CKD | Compound smoking index | Simple mode | 121 | -0,15504 | 0,378747 | 0,683021 | Smoking | 44,08236 |
| SIndex | Incident CKD | Compound smoking index | Weighted mode | 121 | -0,22477 | 0,347885 | 0,519439 | Smoking | 44,08236 |
| T2DGGI | Incident CKD | Type 2 diabetes | MR Egger | 450 | 0,029642 | 0,030053 | 0,3245 | Diabetes | 94,66069 |
| T2DGGI | Incident CKD | Type 2 diabetes | Weighted median | 450 | 0,022047 | 0,025301 | 0,383537 | Diabetes | 94,66069 |
| T2DGGI | Incident CKD | Type 2 diabetes | Inverse variance weighted | 450 | 0,03769 | 0,014851 | 0,011152 | Diabetes | 94,66069 |
| T2DGGI | Incident CKD | Type 2 diabetes | Simple mode | 450 | 0,03691 | 0,057017 | 0,517737 | Diabetes | 94,66069 |
| T2DGGI | Incident CKD | Type 2 diabetes | Weighted mode | 450 | 0,045343 | 0,030177 | 0,133647 | Diabetes | 94,66069 |
| TC | Incident CKD | Total cholesterol | MR Egger | 276 | 0,002292 | 0,050086 | 0,963531 | Dyslipidemias | 255,0395 |
| TC | Incident CKD | Total cholesterol | Weighted median | 276 | -0,07445 | 0,041192 | 0,070688 | Dyslipidemias | 255,0395 |
| TC | Incident CKD | Total cholesterol | Inverse variance weighted | 276 | -0,04866 | 0,032967 | 0,139966 | Dyslipidemias | 255,0395 |
| TC | Incident CKD | Total cholesterol | Simple mode | 276 | 0,006512 | 0,094303 | 0,944997 | Dyslipidemias | 255,0395 |
| TC | Incident CKD | Total cholesterol | Weighted mode | 276 | -0,04773 | 0,040224 | 0,236459 | Dyslipidemias | 255,0395 |
| TG | Incident CKD | Triglyceride | MR Egger | 226 | -0,06903 | 0,053278 | 0,196438 | Dyslipidemias | 237,206 |
| TG | Incident CKD | Triglyceride | Weighted median | 226 | 0,008711 | 0,044817 | 0,845895 | Dyslipidemias | 237,206 |
| TG | Incident CKD | Triglyceride | Inverse variance weighted | 226 | 0,038234 | 0,035712 | 0,284329 | Dyslipidemias | 237,206 |
| TG | Incident CKD | Triglyceride | Simple mode | 226 | 0,11346 | 0,118562 | 0,339613 | Dyslipidemias | 237,206 |
| TG | Incident CKD | Triglyceride | Weighted mode | 226 | -0,01083 | 0,041985 | 0,796758 | Dyslipidemias | 237,206 |
| TSH | Incident CKD | Thyroid Stimulating Hormone | MR Egger | 164 | 0,098909 | 0,04473 | 0,028419 | Thyroid disease | 153,509 |
| TSH | Incident CKD | Thyroid Stimulating Hormone | Weighted median | 164 | 0,056105 | 0,034783 | 0,106744 | Thyroid disease | 153,509 |
| TSH | Incident CKD | Thyroid Stimulating Hormone | Inverse variance weighted | 164 | 0,092128 | 0,024885 | 0,000214 | Thyroid disease | 153,509 |
| TSH | Incident CKD | Thyroid Stimulating Hormone | Simple mode | 164 | -0,00225 | 0,075545 | 0,976293 | Thyroid disease | 153,509 |
| TSH | Incident CKD | Thyroid Stimulating Hormone | Weighted mode | 164 | 0,042877 | 0,037293 | 0,251939 | Thyroid disease | 153,509 |
| WHR | Incident CKD | Waist-to-hip ratio adjusted BMI | MR Egger | 297 | 0,047949 | 0,109073 | 0,660545 | Obesity | 85,78362 |
| WHR | Incident CKD | Waist-to-hip ratio adjusted BMI | Weighted median | 297 | -0,01409 | 0,05906 | 0,811475 | Obesity | 85,78362 |
| WHR | Incident CKD | Waist-to-hip ratio adjusted BMI | Inverse variance weighted | 297 | 0,000222 | 0,045531 | 0,996116 | Obesity | 85,78362 |
| WHR | Incident CKD | Waist-to-hip ratio adjusted BMI | Simple mode | 297 | -0,02503 | 0,160362 | 0,876091 | Obesity | 85,78362 |
| WHR | Incident CKD | Waist-to-hip ratio adjusted BMI | Weighted mode | 297 | 0,005766 | 0,094688 | 0,951487 | Obesity | 85,78362 |
| Zn | Incident CKD | Zinc | MR Egger | 3 | -0,00548 | 0,115675 | 0,969854 | Minerals | 99,95282 |
| Zn | Incident CKD | Zinc | Weighted median | 3 | 0,018423 | 0,034064 | 0,588615 | Minerals | 99,95282 |
| Zn | Incident CKD | Zinc | Inverse variance weighted | 3 | 0,045326 | 0,032945 | 0,168873 | Minerals | 99,95282 |
| Zn | Incident CKD | Zinc | Simple mode | 3 | 0,018037 | 0,060853 | 0,79487 | Minerals | 99,95282 |
| Zn | Incident CKD | Zinc | Weighted mode | 3 | 0,01504 | 0,039499 | 0,74001 | Minerals | 99,95282 |
| ebi-a-GCST006979-consort | Incident CKD | Heel bone mineral density | MR Egger | 498 | -0,0686 | 0,043251 | 0,113359 | Bone disease | 152,6556 |
| ebi-a-GCST006979-consort | Incident CKD | Heel bone mineral density | Weighted median | 498 | -0,02668 | 0,031675 | 0,399664 | Bone disease | 152,6556 |
| ebi-a-GCST006979-consort | Incident CKD | Heel bone mineral density | Inverse variance weighted | 498 | 0,00589 | 0,022975 | 0,797673 | Bone disease | 152,6556 |
| ebi-a-GCST006979-consort | Incident CKD | Heel bone mineral density | Simple mode | 498 | -0,04185 | 0,079736 | 0,599952 | Bone disease | 152,6556 |
| ebi-a-GCST006979-consort | Incident CKD | Heel bone mineral density | Weighted mode | 498 | -0,05451 | 0,047168 | 0,248343 | Bone disease | 152,6556 |
| ebi-a-GCST90002227 | Incident CKD | Two-hour glucose | MR Egger | 14 | 0,111944 | 0,188184 | 0,562982 | Diabetes | 56,47851 |
| ebi-a-GCST90002227 | Incident CKD | Two-hour glucose | Weighted median | 14 | -0,00756 | 0,060618 | 0,900685 | Diabetes | 56,47851 |
| ebi-a-GCST90002227 | Incident CKD | Two-hour glucose | Inverse variance weighted | 14 | -0,03124 | 0,069265 | 0,651992 | Diabetes | 56,47851 |
| ebi-a-GCST90002227 | Incident CKD | Two-hour glucose | Simple mode | 14 | -0,08858 | 0,093034 | 0,35838 | Diabetes | 56,47851 |
| ebi-a-GCST90002227 | Incident CKD | Two-hour glucose | Weighted mode | 14 | -0,03463 | 0,074459 | 0,64953 | Diabetes | 56,47851 |
| ebi-a-GCST90002232 | Incident CKD | Fasting glucose | MR Egger | 69 | 0,131853 | 0,177807 | 0,460951 | Diabetes | 122,8115 |
| ebi-a-GCST90002232 | Incident CKD | Fasting glucose | Weighted median | 69 | -0,02557 | 0,112165 | 0,819645 | Diabetes | 122,8115 |
| ebi-a-GCST90002232 | Incident CKD | Fasting glucose | Inverse variance weighted | 69 | 0,081148 | 0,092314 | 0,379378 | Diabetes | 122,8115 |
| ebi-a-GCST90002232 | Incident CKD | Fasting glucose | Simple mode | 69 | -0,03746 | 0,210809 | 0,859489 | Diabetes | 122,8115 |
| ebi-a-GCST90002232 | Incident CKD | Fasting glucose | Weighted mode | 69 | -0,02037 | 0,113 | 0,857515 | Diabetes | 122,8115 |
| ebi-a-GCST90002238 | Incident CKD | Fasting insulin | MR Egger | 37 | -0,37928 | 0,521519 | 0,4719 | Diabetes | 52,15552 |

| | | | | | | | | | |
|---|---|---|---|---|---|---|---|---|---|
| ebi-a-GCST90002238 | Incident CKD | Fasting insulin | Weighted median | 37 | 0,108295 | 0,185014 | 0,558322 | Diabetes | 52,15552 |
| ebi-a-GCST90002238 | Incident CKD | Fasting insulin | Inverse variance weighted | 37 | 0,242265 | 0,164365 | 0,140497 | Diabetes | 52,15552 |
| ebi-a-GCST90002238 | Incident CKD | Fasting insulin | Simple mode | 37 | 0,045884 | 0,419865 | 0,913585 | Diabetes | 52,15552 |
| ebi-a-GCST90002238 | Incident CKD | Fasting insulin | Weighted mode | 37 | 0,018975 | 0,347193 | 0,956718 | Diabetes | 52,15552 |
| ebi-a-GCST90007310 | Incident CKD | circulating leptin levels | MR Egger | 3 | 0,208663 | 1,834228 | 0,927888 | Obesity | 56,18818 |
| ebi-a-GCST90007310 | Incident CKD | circulating leptin levels | Weighted median | 3 | -0,00672 | 0,147495 | 0,963675 | Obesity | 56,18818 |
| ebi-a-GCST90007310 | Incident CKD | circulating leptin levels | Inverse variance weighted | 3 | 0,033523 | 0,164289 | 0,838318 | Obesity | 56,18818 |
| ebi-a-GCST90007310 | Incident CKD | circulating leptin levels | Simple mode | 3 | -0,06544 | 0,225861 | 0,799296 | Obesity | 56,18818 |
| ebi-a-GCST90007310 | Incident CKD | circulating leptin levels | Weighted mode | 3 | -0,10809 | 0,242902 | 0,699857 | Obesity | 56,18818 |
| ebi-a-GCST90011998 | Incident CKD | Growth differentiation factor-15 levels | MR Egger | 3 | -0,08737 | 0,125182 | 0,612092 | Biomarkers | 294,0397 |
| ebi-a-GCST90011998 | Incident CKD | Growth differentiation factor-15 levels | Weighted median | 3 | -0,04786 | 0,036969 | 0,195493 | Biomarkers | 294,0397 |
| ebi-a-GCST90011998 | Incident CKD | Growth differentiation factor-15 levels | Inverse variance weighted | 3 | -0,03259 | 0,070907 | 0,645833 | Biomarkers | 294,0397 |
| ebi-a-GCST90011998 | Incident CKD | Growth differentiation factor-15 levels | Simple mode | 3 | -0,10857 | 0,064268 | 0,233222 | Biomarkers | 294,0397 |
| ebi-a-GCST90011998 | Incident CKD | Growth differentiation factor-15 levels | Weighted mode | 3 | -0,05112 | 0,035955 | 0,291034 | Biomarkers | 294,0397 |
| ebi-a-GCST90012022 | Incident CKD | Fibroblast growth factor 23 levels | MR Egger | 8 | 0,830553 | 0,568071 | 0,194041 | Biomarkers | 43,42098 |
| ebi-a-GCST90012022 | Incident CKD | Fibroblast growth factor 23 levels | Weighted median | 8 | 0,138472 | 0,075976 | 0,068367 | Biomarkers | 43,42098 |
| ebi-a-GCST90012022 | Incident CKD | Fibroblast growth factor 23 levels | Inverse variance weighted | 8 | -0,03372 | 0,167095 | 0,840079 | Biomarkers | 43,42098 |
| ebi-a-GCST90012022 | Incident CKD | Fibroblast growth factor 23 levels | Simple mode | 8 | 0,154053 | 0,083115 | 0,10622 | Biomarkers | 43,42098 |
| ebi-a-GCST90012022 | Incident CKD | Fibroblast growth factor 23 levels | Weighted mode | 8 | 0,206481 | 0,072416 | 0,024641 | Biomarkers | 43,42098 |
| ebi-a-GCST90012041 | Incident CKD | Kidney injury molecule 1 levels | MR Egger | 9 | -0,1299 | 0,211778 | 0,559027 | Biomarkers | 190,9807 |
| ebi-a-GCST90012041 | Incident CKD | Kidney injury molecule 1 levels | Weighted median | 9 | -0,04891 | 0,024243 | 0,043658 | Biomarkers | 190,9807 |
| ebi-a-GCST90012041 | Incident CKD | Kidney injury molecule 1 levels | Inverse variance weighted | 9 | 0,065134 | 0,137071 | 0,634658 | Biomarkers | 190,9807 |
| ebi-a-GCST90012041 | Incident CKD | Kidney injury molecule 1 levels | Simple mode | 9 | 0,013648 | 0,055495 | 0,811921 | Biomarkers | 190,9807 |
| ebi-a-GCST90012041 | Incident CKD | Kidney injury molecule 1 levels | Weighted mode | 9 | -0,05213 | 0,024591 | 0,066818 | Biomarkers | 190,9807 |
| ebi-a-GCST90013864 | Incident CKD | Coronary artery disease (Firth correction) | MR Egger | 39 | 0,071285 | 0,072265 | 0,330322 | Cardiovascular disease | 59,15571 |
| ebi-a-GCST90013864 | Incident CKD | Coronary artery disease (Firth correction) | Weighted median | 39 | 0,046964 | 0,039583 | 0,235439 | Cardiovascular disease | 59,15571 |
| ebi-a-GCST90013864 | Incident CKD | Coronary artery disease (Firth correction) | Inverse variance weighted | 39 | 0,040108 | 0,033033 | 0,224677 | Cardiovascular disease | 59,15571 |
| ebi-a-GCST90013864 | Incident CKD | Coronary artery disease (Firth correction) | Simple mode | 39 | 0,105889 | 0,075124 | 0,166811 | Cardiovascular disease | 59,15571 |
| ebi-a-GCST90013864 | Incident CKD | Coronary artery disease (Firth correction) | Weighted mode | 39 | 0,098912 | 0,049237 | 0,051695 | Cardiovascular disease | 59,15571 |
| ebi-a-GCST90013975 | Incident CKD | Body fat percentage | MR Egger | 318 | -0,05758 | 0,174764 | 0,742006 | Obesity | 56,09903 |
| ebi-a-GCST90013975 | Incident CKD | Body fat percentage | Weighted median | 318 | 0,174024 | 0,073938 | 0,01859 | Obesity | 56,09903 |
| ebi-a-GCST90013975 | Incident CKD | Body fat percentage | Inverse variance weighted | 318 | 0,120737 | 0,054794 | 0,027561 | Obesity | 56,09903 |
| ebi-a-GCST90013975 | Incident CKD | Body fat percentage | Simple mode | 318 | 0,261772 | 0,252714 | 0,301066 | Obesity | 56,09903 |
| ebi-a-GCST90013975 | Incident CKD | Body fat percentage | Weighted mode | 318 | 0,329886 | 0,168471 | 0,051094 | Obesity | 56,09903 |
| ebi-a-GCST90013989 | Incident CKD | Sodium in urine | MR Egger | 41 | 0,65113 | 0,633448 | 0,310324 | Minerals | 43,81211 |
| ebi-a-GCST90013989 | Incident CKD | Sodium in urine | Weighted median | 41 | 0,029694 | 0,173173 | 0,863855 | Minerals | 43,81211 |
| ebi-a-GCST90013989 | Incident CKD | Sodium in urine | Inverse variance weighted | 41 | 0,031964 | 0,161903 | 0,843495 | Minerals | 43,81211 |
| ebi-a-GCST90013989 | Incident CKD | Sodium in urine | Simple mode | 41 | -0,26166 | 0,443115 | 0,558169 | Minerals | 43,81211 |
| ebi-a-GCST90013989 | Incident CKD | Sodium in urine | Weighted mode | 41 | 0,659891 | 0,488312 | 0,18417 | Minerals | 43,81211 |
| ebi-a-GCST90014006 | Incident CKD | Glycated haemoglobin HbA1c levels | MR Egger | 316 | -0,00111 | 0,056801 | 0,984376 | Diabetes | 129,8664 |
| ebi-a-GCST90014006 | Incident CKD | Glycated haemoglobin HbA1c levels | Weighted median | 316 | -0,00564 | 0,039999 | 0,887848 | Diabetes | 129,8664 |
| ebi-a-GCST90014006 | Incident CKD | Glycated haemoglobin HbA1c levels | Inverse variance weighted | 316 | 0,014894 | 0,030704 | 0,62762 | Diabetes | 129,8664 |
| ebi-a-GCST90014006 | Incident CKD | Glycated haemoglobin HbA1c levels | Simple mode | 316 | 0,065323 | 0,095607 | 0,494951 | Diabetes | 129,8664 |
| ebi-a-GCST90014006 | Incident CKD | Glycated haemoglobin HbA1c levels | Weighted mode | 316 | 0,003936 | 0,048132 | 0,934874 | Diabetes | 129,8664 |
| ebi-a-GCST90014010 | Incident CKD | Phosphate levels | MR Egger | 162 | 0,054189 | 0,105523 | 0,608294 | Minerals | 88,26243 |
| ebi-a-GCST90014010 | Incident CKD | Phosphate levels | Weighted median | 162 | -0,04758 | 0,066579 | 0,474843 | Minerals | 88,26243 |
| ebi-a-GCST90014010 | Incident CKD | Phosphate levels | Inverse variance weighted | 162 | 0,022449 | 0,059097 | 0,70405 | Minerals | 88,26243 |
| ebi-a-GCST90014010 | Incident CKD | Phosphate levels | Simple mode | 162 | 0,036692 | 0,156586 | 0,815032 | Minerals | 88,26243 |
| ebi-a-GCST90014010 | Incident CKD | Phosphate levels | Weighted mode | 162 | -0,0493 | 0,080856 | 0,54289 | Minerals | 88,26243 |
| ebi-a-GCST90014022 | Incident CKD | Bone mineral density | MR Egger | 424 | -0,03431 | 0,048454 | 0,479225 | Bone disease | 121,6064 |
| ebi-a-GCST90014022 | Incident CKD | Bone mineral density | Weighted median | 424 | -0,00981 | 0,034088 | 0,773429 | Bone disease | 121,6064 |
| ebi-a-GCST90014022 | Incident CKD | Bone mineral density | Inverse variance weighted | 424 | -0,01175 | 0,02511 | 0,639868 | Bone disease | 121,6064 |
| ebi-a-GCST90014022 | Incident CKD | Bone mineral density | Simple mode | 424 | 0,039101 | 0,095366 | 0,682005 | Bone disease | 121,6064 |
| ebi-a-GCST90014022 | Incident CKD | Bone mineral density | Weighted mode | 424 | -0,01121 | 0,051923 | 0,829249 | Bone disease | 121,6064 |
| ebi-a-GCST90014023 | Incident CKD | Type 1 diabetes | MR Egger | 77 | 0,017281 | 0,012437 | 0,168802 | Diabetes | 130,8043 |
| ebi-a-GCST90014023 | Incident CKD | Type 1 diabetes | Weighted median | 77 | 0,025005 | 0,011198 | 0,025545 | Diabetes | 130,8043 |
| ebi-a-GCST90014023 | Incident CKD | Type 1 diabetes | Inverse variance weighted | 77 | 0,012297 | 0,007593 | 0,105351 | Diabetes | 130,8043 |
| ebi-a-GCST90014023 | Incident CKD | Type 1 diabetes | Simple mode | 77 | 0,016095 | 0,021404 | 0,454399 | Diabetes | 130,8043 |
| ebi-a-GCST90014023 | Incident CKD | Type 1 diabetes | Weighted mode | 77 | 0,022739 | 0,01107 | 0,043395 | Diabetes | 130,8043 |
| ebi-a-GCST90016674 | Incident CKD | Liver iron content | MR Egger | 5 | -0,09582 | 0,058795 | 0,201662 | Minerals | 257,3642 |
| ebi-a-GCST90016674 | Incident CKD | Liver iron content | Weighted median | 5 | -0,06002 | 0,039034 | 0,124128 | Minerals | 257,3642 |
| ebi-a-GCST90016674 | Incident CKD | Liver iron content | Inverse variance weighted | 5 | -0,05097 | 0,037421 | 0,173178 | Minerals | 257,3642 |
| ebi-a-GCST90016674 | Incident CKD | Liver iron content | Simple mode | 5 | -0,03805 | 0,072213 | 0,626134 | Minerals | 257,3642 |
| ebi-a-GCST90016674 | Incident CKD | Liver iron content | Weighted mode | 5 | -0,06602 | 0,043638 | 0,204847 | Minerals | 257,3642 |
| ebi-a-GCST90016676 | Incident CKD | Pancreas iron content | Wald ratio | 1 | -0,05375 | 0,203472 | 0,791664 | Minerals | 30,56027 |
| ebi-a-GCST90018806 | Incident CKD | Chronic heart failure | MR Egger | 6 | 0,104927 | 0,285258 | 0,731635 | Cardiovascular disease | 40,14382 |
| ebi-a-GCST90018806 | Incident CKD | Chronic heart failure | Weighted median | 6 | 0,143147 | 0,0819 | 0,080494 | Cardiovascular disease | 40,14382 |
| ebi-a-GCST90018806 | Incident CKD | Chronic heart failure | Inverse variance weighted | 6 | 0,062796 | 0,078284 | 0,422465 | Cardiovascular disease | 40,14382 |
| ebi-a-GCST90018806 | Incident CKD | Chronic heart failure | Simple mode | 6 | 0,151416 | 0,099592 | 0,188898 | Cardiovascular disease | 40,14382 |
| ebi-a-GCST90018806 | Incident CKD | Chronic heart failure | Weighted mode | 6 | 0,155171 | 0,102191 | 0,189364 | Cardiovascular disease | 40,14382 |
| ebi-a-GCST90018860 | Incident CKD | Hyperthyroidism | MR Egger | 8 | 0,061096 | 0,107265 | 0,589635 | Thyroid disease | 53,27573 |
| ebi-a-GCST90018860 | Incident CKD | Hyperthyroidism | Weighted median | 8 | 0,029863 | 0,027291 | 0,273839 | Thyroid disease | 53,27573 |
| ebi-a-GCST90018860 | Incident CKD | Hyperthyroidism | Inverse variance weighted | 8 | 0,011306 | 0,029123 | 0,697859 | Thyroid disease | 53,27573 |
| ebi-a-GCST90018860 | Incident CKD | Hyperthyroidism | Simple mode | 8 | 0,039189 | 0,03598 | 0,312137 | Thyroid disease | 53,27573 |
| ebi-a-GCST90018860 | Incident CKD | Hyperthyroidism | Weighted mode | 8 | 0,035753 | 0,029799 | 0,269245 | Thyroid disease | 53,27573 |
| ebi-a-GCST90018862 | Incident CKD | Hypothyroidism | MR Egger | 68 | 0,058383 | 0,037567 | 0,12494 | Thyroid disease | 79,29446 |
| ebi-a-GCST90018862 | Incident CKD | Hypothyroidism | Weighted median | 68 | 0,041075 | 0,019643 | 0,036524 | Thyroid disease | 79,29446 |
| ebi-a-GCST90018862 | Incident CKD | Hypothyroidism | Inverse variance weighted | 68 | 0,048001 | 0,016359 | 0,003343 | Thyroid disease | 79,29446 |
| ebi-a-GCST90018862 | Incident CKD | Hypothyroidism | Simple mode | 68 | 0,072795 | 0,038043 | 0,05996 | Thyroid disease | 79,29446 |
| ebi-a-GCST90018862 | Incident CKD | Hypothyroidism | Weighted mode | 68 | 0,035576 | 0,023129 | 0,128714 | Thyroid disease | 79,29446 |
| ebi-a-GCST90018872 | Incident CKD | Iron deficiency anemia | Wald ratio | 1 | -0,17801 | 0,149215 | 0,232876 | Minerals | 34,53822 |
| ebi-a-GCST90018877 | Incident CKD | Myocardial infarction | MR Egger | 75 | 0,008641 | 0,056328 | 0,878503 | Cardiovascular disease | 66,10062 |
| ebi-a-GCST90018877 | Incident CKD | Myocardial infarction | Weighted median | 75 | 0,035939 | 0,029254 | 0,219256 | Cardiovascular disease | 66,10062 |
| ebi-a-GCST90018877 | Incident CKD | Myocardial infarction | Inverse variance weighted | 75 | -0,01256 | 0,025436 | 0,621364 | Cardiovascular disease | 66,10062 |
| ebi-a-GCST90018877 | Incident CKD | Myocardial infarction | Simple mode | 75 | 0,015496 | 0,069835 | 0,825005 | Cardiovascular disease | 66,10062 |
| ebi-a-GCST90018877 | Incident CKD | Myocardial infarction | Weighted mode | 75 | 0,045564 | 0,046543 | 0,330788 | Cardiovascular disease | 66,10062 |
| ebi-a-GCST90018916 | Incident CKD | Sleep apnea syndrome | Wald ratio | 1 | 0,162162 | 0,088803 | 0,067837 | Sleep disorders | 63,50864 |
| ebi-a-GCST90018977 | Incident CKD | Serum uric acid levels | MR Egger | 254 | -0,03196 | 0,101622 | 0,7534 | Uricemia | 122,4585 |
| ebi-a-GCST90018977 | Incident CKD | Serum uric acid levels | Weighted median | 254 | 0,062953 | 0,057197 | 0,271058 | Uricemia | 122,4585 |
| ebi-a-GCST90018977 | Incident CKD | Serum uric acid levels | Inverse variance weighted | 254 | 0,244439 | 0,066325 | 0,000228 | Uricemia | 122,4585 |
| ebi-a-GCST90018977 | Incident CKD | Serum uric acid levels | Simple mode | 254 | 0,132221 | 0,140858 | 0,348789 | Uricemia | 122,4585 |
| ebi-a-GCST90018977 | Incident CKD | Serum uric acid levels | Weighted mode | 254 | 0,007957 | 0,047448 | 0,86695 | Uricemia | 122,4585 |
| ebi-a-GCST90019417 | Incident CKD | Vitamin K-dependent protein S measurement | MR Egger | 4 | 0,013057 | 0,058952 | 0,845267 | Vitamin | 118,3171 |
| ebi-a-GCST90019417 | Incident CKD | Vitamin K-dependent protein S measurement | Weighted median | 4 | 0,043738 | 0,037136 | 0,238888 | Vitamin | 118,3171 |
| ebi-a-GCST90019417 | Incident CKD | Vitamin K-dependent protein S measurement | Inverse variance weighted | 4 | 0,049762 | 0,033774 | 0,140642 | Vitamin | 118,3171 |
| ebi-a-GCST90019417 | Incident CKD | Vitamin K-dependent protein S measurement | Simple mode | 4 | 0,105401 | 0,065576 | 0,206349 | Vitamin | 118,3171 |
| ebi-a-GCST90019417 | Incident CKD | Vitamin K-dependent protein S measurement | Weighted mode | 4 | 0,012659 | 0,043253 | 0,788852 | Vitamin | 118,3171 |
| ebi-a-GCST90019423 | Incident CKD | Vitamin K-dependent protein C measurement | MR Egger | 4 | 0,006688 | 0,05073 | 0,907184 | Vitamin | 174,2568 |
| ebi-a-GCST90019423 | Incident CKD | Vitamin K-dependent protein C measurement | Weighted median | 4 | 0,005026 | 0,026904 | 0,851815 | Vitamin | 174,2568 |
| ebi-a-GCST90019423 | Incident CKD | Vitamin K-dependent protein C measurement | Inverse variance weighted | 4 | 0,007586 | 0,025531 | 0,76637 | Vitamin | 174,2568 |
| ebi-a-GCST90019423 | Incident CKD | Vitamin K-dependent protein C measurement | Simple mode | 4 | -0,00312 | 0,051999 | 0,955887 | Vitamin | 174,2568 |
| ebi-a-GCST90019423 | Incident CKD | Vitamin K-dependent protein C measurement | Weighted mode | 4 | 0,003972 | 0,027002 | 0,892383 | Vitamin | 174,2568 |
| ebi-a-GCST90025947 | Incident CKD | Serum alkaline phosphatase levels | MR Egger | 344 | -0,08632 | 0,049004 | 0,079063 | Minerals | 165,7959 |
| ebi-a-GCST90025947 | Incident CKD | Serum alkaline phosphatase levels | Weighted median | 344 | -0,04111 | 0,038519 | 0,285796 | Minerals | 165,7959 |
| ebi-a-GCST90025947 | Incident CKD | Serum alkaline phosphatase levels | Inverse variance weighted | 344 | -0,03345 | 0,029006 | 0,248849 | Minerals | 165,7959 |
| ebi-a-GCST90025947 | Incident CKD | Serum alkaline phosphatase levels | Simple mode | 344 | 0,048836 | 0,08116 | 0,547759 | Minerals | 165,7959 |
| ebi-a-GCST90025947 | Incident CKD | Serum alkaline phosphatase levels | Weighted mode | 344 | -0,00822 | 0,039964 | 0,837153 | Minerals | 165,7959 |
| ebi-a-GCST90025967 | Incident CKD | Vitamin D levels | MR Egger | 84 | 0,041336 | 0,108958 | 0,705387 | Vitamin | 131,2146 |
| ebi-a-GCST90025967 | Incident CKD | Vitamin D levels | Weighted median | 84 | 0,005519 | 0,072926 | 0,939672 | Vitamin | 131,2146 |
| ebi-a-GCST90025967 | Incident CKD | Vitamin D levels | Inverse variance weighted | 84 | 0,092957 | 0,066471 | 0,161975 | Vitamin | 131,2146 |
| ebi-a-GCST90025967 | Incident CKD | Vitamin D levels | Simple mode | 84 | -0,03731 | 0,140385 | 0,791058 | Vitamin | 131,2146 |
| ebi-a-GCST90025967 | Incident CKD | Vitamin D levels | Weighted mode | 84 | 0,019894 | 0,065446 | 0,761913 | Vitamin | 131,2146 |
| ebi-a-GCST90025969 | Incident CKD | Hemoglobin levels | MR Egger | 330 | -0,08536 | 0,089451 | 0,340632 | Organic compound | 105,1179 |

| | | | | | | | | | |
|---|---|---|---|---|---|---|---|---|---|
| ebi-a-GCST90025969 | Incident CKD | Hemoglobin levels | Weighted median | 330 | -0,04153 | 0,051818 | 0,422816 | Organic compound | 105,1179 |
| ebi-a-GCST90025969 | Incident CKD | Hemoglobin levels | Inverse variance weighted | 330 | -0,05759 | 0,047111 | 0,221567 | Organic compound | 105,1179 |
| ebi-a-GCST90025969 | Incident CKD | Hemoglobin levels | Simple mode | 330 | 0,085586 | 0,140065 | 0,541591 | Organic compound | 105,1179 |
| ebi-a-GCST90025969 | Incident CKD | Hemoglobin levels | Weighted mode | 330 | -0,07925 | 0,070551 | 0,262114 | Organic compound | 105,1179 |
| ebi-a-GCST90025986 | Incident CKD | Blood glucose levels | MR Egger | 110 | 0,013197 | 0,092769 | 0,887141 | Diabetes | 118,1557 |
| ebi-a-GCST90025986 | Incident CKD | Blood glucose levels | Weighted median | 110 | 0,010321 | 0,063363 | 0,870606 | Diabetes | 118,1557 |
| ebi-a-GCST90025986 | Incident CKD | Blood glucose levels | Inverse variance weighted | 110 | -0,02593 | 0,051477 | 0,614411 | Diabetes | 118,1557 |
| ebi-a-GCST90025986 | Incident CKD | Blood glucose levels | Simple mode | 110 | -0,08042 | 0,129348 | 0,535395 | Diabetes | 118,1557 |
| ebi-a-GCST90025986 | Incident CKD | Blood glucose levels | Weighted mode | 110 | 0,006243 | 0,058327 | 0,914964 | Diabetes | 118,1557 |
| ebi-a-GCST90025989 | Incident CKD | Insulin-like growth factor 1 levels | MR Egger | 382 | 0,005895 | 0,05834 | 0,91957 | Diabetes | 115,6066 |
| ebi-a-GCST90025989 | Incident CKD | Insulin-like growth factor 1 levels | Weighted median | 382 | 0,010359 | 0,040969 | 0,800385 | Diabetes | 115,6066 |
| ebi-a-GCST90025989 | Incident CKD | Insulin-like growth factor 1 levels | Inverse variance weighted | 382 | -0,01884 | 0,030562 | 0,537518 | Diabetes | 115,6066 |
| ebi-a-GCST90025989 | Incident CKD | Insulin-like growth factor 1 levels | Simple mode | 382 | 0,054236 | 0,100384 | 0,589315 | Diabetes | 115,6066 |
| ebi-a-GCST90025989 | Incident CKD | Insulin-like growth factor 1 levels | Weighted mode | 382 | 0,069173 | 0,059302 | 0,244159 | Diabetes | 115,6066 |
| ebi-a-GCST90025990 | Incident CKD | Calcium levels | MR Egger | 214 | -0,11081 | 0,090539 | 0,222342 | Minerals | 102,3373 |
| ebi-a-GCST90025990 | Incident CKD | Calcium levels | Weighted median | 214 | -0,18676 | 0,052296 | 0,000355 | Minerals | 102,3373 |
| ebi-a-GCST90025990 | Incident CKD | Calcium levels | Inverse variance weighted | 214 | -0,02625 | 0,0503 | 0,601726 | Minerals | 102,3373 |
| ebi-a-GCST90025990 | Incident CKD | Calcium levels | Simple mode | 214 | -0,15114 | 0,145016 | 0,2985 | Minerals | 102,3373 |
| ebi-a-GCST90025990 | Incident CKD | Calcium levels | Weighted mode | 214 | -0,17415 | 0,068948 | 0,012272 | Minerals | 102,3373 |
| ebi-a-GCST90029013 | Incident CKD | Educational attainment (years of education) | MR Egger | 210 | -0,04601 | 0,050544 | 0,363748 | Social trait | 45,00571 |
| ebi-a-GCST90029013 | Incident CKD | Educational attainment (years of education) | Weighted median | 210 | -0,01901 | 0,014624 | 0,193592 | Social trait | 45,00571 |
| ebi-a-GCST90029013 | Incident CKD | Educational attainment (years of education) | Inverse variance weighted | 210 | -0,02271 | 0,011697 | 0,052217 | Social trait | 45,00571 |
| ebi-a-GCST90029013 | Incident CKD | Educational attainment (years of education) | Simple mode | 210 | -0,03731 | 0,046421 | 0,422515 | Social trait | 45,00571 |
| ebi-a-GCST90029013 | Incident CKD | Educational attainment (years of education) | Weighted mode | 210 | -0,03401 | 0,045444 | 0,455058 | Social trait | 45,00571 |
| ebi-a-GCST90029014 | Incident CKD | Smoking status | MR Egger | 117 | 0,21187 | 0,423881 | 0,618147 | Smoking | 42,91926 |
| ebi-a-GCST90029014 | Incident CKD | Smoking status | Weighted median | 117 | 0,01442 | 0,143406 | 0,919907 | Smoking | 42,91926 |
| ebi-a-GCST90029014 | Incident CKD | Smoking status | Inverse variance weighted | 117 | 0,058042 | 0,099983 | 0,561566 | Smoking | 42,91926 |
| ebi-a-GCST90029014 | Incident CKD | Smoking status | Simple mode | 117 | -0,19552 | 0,420068 | 0,642486 | Smoking | 42,91926 |
| ebi-a-GCST90029014 | Incident CKD | Smoking status | Weighted mode | 117 | -0,22532 | 0,361624 | 0,534464 | Smoking | 42,91926 |
| ebi-a-GCST90029070 | Incident CKD | C-reactive protein levels | MR Egger | 228 | -0,07947 | 0,058347 | 0,174535 | Inflammation | 121,6981 |
| ebi-a-GCST90029070 | Incident CKD | C-reactive protein levels | Weighted median | 228 | -0,05454 | 0,051116 | 0,285964 | Inflammation | 121,6981 |
| ebi-a-GCST90029070 | Incident CKD | C-reactive protein levels | Inverse variance weighted | 228 | -0,03124 | 0,035037 | 0,372645 | Inflammation | 121,6981 |
| ebi-a-GCST90029070 | Incident CKD | C-reactive protein levels | Simple mode | 228 | 0,017641 | 0,122234 | 0,885377 | Inflammation | 121,6981 |
| ebi-a-GCST90029070 | Incident CKD | C-reactive protein levels | Weighted mode | 228 | -0,04505 | 0,061026 | 0,461106 | Inflammation | 121,6981 |
| ebi-a-GCST90029073 | Incident CKD | Liver fat | MR Egger | 8 | 0,047458 | 0,136756 | 0,740419 | Fatty liver | 140,8023 |
| ebi-a-GCST90029073 | Incident CKD | Liver fat | Weighted median | 8 | 0,027474 | 0,043847 | 0,53093 | Fatty liver | 140,8023 |
| ebi-a-GCST90029073 | Incident CKD | Liver fat | Inverse variance weighted | 8 | 0,000498 | 0,078099 | 0,994909 | Fatty liver | 140,8023 |
| ebi-a-GCST90029073 | Incident CKD | Liver fat | Simple mode | 8 | 0,056133 | 0,06924 | 0,444226 | Fatty liver | 140,8023 |
| ebi-a-GCST90029073 | Incident CKD | Liver fat | Weighted mode | 8 | 0,031011 | 0,041395 | 0,478175 | Fatty liver | 140,8023 |
| ebi-a-GCST90086057 | Incident CKD | Cardiovascular disease | Inverse variance weighted | 2 | 0,12156 | 0,185464 | 0,512186 | Cardiovascular disease | 32,018 |
| finn-b-D3_ANAEMIA_B12_DEF | Incident CKD | Vitamin B12 deficiency anaemia | MR Egger | 4 | 0,040247 | 0,056754 | 0,551754 | Vitamin | 52,61875 |
| finn-b-D3_ANAEMIA_B12_DEF | Incident CKD | Vitamin B12 deficiency anaemia | Weighted median | 4 | 0,026382 | 0,021892 | 0,228166 | Vitamin | 52,61875 |
| finn-b-D3_ANAEMIA_B12_DEF | Incident CKD | Vitamin B12 deficiency anaemia | Inverse variance weighted | 4 | 0,02804 | 0,019276 | 0,145772 | Vitamin | 52,61875 |
| finn-b-D3_ANAEMIA_B12_DEF | Incident CKD | Vitamin B12 deficiency anaemia | Simple mode | 4 | 0,020188 | 0,028261 | 0,526572 | Vitamin | 52,61875 |
| finn-b-D3_ANAEMIA_B12_DEF | Incident CKD | Vitamin B12 deficiency anaemia | Weighted mode | 4 | 0,026937 | 0,027989 | 0,406843 | Vitamin | 52,61875 |
| finn-b-E4_OBESITY | Incident CKD | Obesity | MR Egger | 8 | 0,169685 | 0,085507 | 0,09444 | Obesity | 55,97912 |
| finn-b-E4_OBESITY | Incident CKD | Obesity | Weighted median | 8 | 0,024613 | 0,037086 | 0,506901 | Obesity | 55,97912 |
| finn-b-E4_OBESITY | Incident CKD | Obesity | Inverse variance weighted | 8 | -0,00445 | 0,034315 | 0,896831 | Obesity | 55,97912 |
| finn-b-E4_OBESITY | Incident CKD | Obesity | Simple mode | 8 | -0,04926 | 0,076986 | 0,542607 | Obesity | 55,97912 |
| finn-b-E4_OBESITY | Incident CKD | Obesity | Weighted mode | 8 | 0,050433 | 0,03889 | 0,235799 | Obesity | 55,97912 |
| finn-b-FG_PULMHEART | Incident CKD | Pulmonary heart disease | MR Egger | 6 | 0,132815 | 0,061177 | 0,095703 | Cardiovascular disease | 80,37648 |
| finn-b-FG_PULMHEART | Incident CKD | Pulmonary heart disease | Weighted median | 6 | 0,026344 | 0,023445 | 0,261157 | Cardiovascular disease | 80,37648 |
| finn-b-FG_PULMHEART | Incident CKD | Pulmonary heart disease | Inverse variance weighted | 6 | 0,019661 | 0,027376 | 0,472632 | Cardiovascular disease | 80,37648 |
| finn-b-FG_PULMHEART | Incident CKD | Pulmonary heart disease | Simple mode | 6 | 0,022529 | 0,03066 | 0,495452 | Cardiovascular disease | 80,37648 |
| finn-b-FG_PULMHEART | Incident CKD | Pulmonary heart disease | Weighted mode | 6 | 0,025682 | 0,023911 | 0,33187 | Cardiovascular disease | 80,37648 |
| ieu-b-102 | Incident CKD | Major depression | MR Egger | 48 | -0,5835 | 0,315555 | 0,070871 | Mental health | 38,29386 |
| ieu-b-102 | Incident CKD | Major depression | Weighted median | 48 | 0,058899 | 0,076089 | 0,438883 | Mental health | 38,29386 |
| ieu-b-102 | Incident CKD | Major depression | Inverse variance weighted | 48 | 0,019967 | 0,059301 | 0,736334 | Mental health | 38,29386 |
| ieu-b-102 | Incident CKD | Major depression | Simple mode | 48 | 0,026985 | 0,176167 | 0,878912 | Mental health | 38,29386 |
| ieu-b-102 | Incident CKD | Major depression | Weighted mode | 48 | 0,061326 | 0,160388 | 0,703917 | Mental health | 38,29386 |
| ieu-b-107 | Incident CKD | apolipoprotein A-I | MR Egger | 285 | 0,020617 | 0,057798 | 0,721577 | Dyslipidemias | 140,1499 |
| ieu-b-107 | Incident CKD | apolipoprotein A-I | Weighted median | 285 | -0,03115 | 0,042951 | 0,468282 | Dyslipidemias | 140,1499 |
| ieu-b-107 | Incident CKD | apolipoprotein A-I | Inverse variance weighted | 285 | -0,03928 | 0,034938 | 0,260915 | Dyslipidemias | 140,1499 |
| ieu-b-107 | Incident CKD | apolipoprotein A-I | Simple mode | 285 | 0,012773 | 0,09355 | 0,891491 | Dyslipidemias | 140,1499 |
| ieu-b-107 | Incident CKD | apolipoprotein A-I | Weighted mode | 285 | 0,012773 | 0,048664 | 0,793142 | Dyslipidemias | 140,1499 |
| ieu-b-108 | Incident CKD | apolipoprotein B | MR Egger | 183 | -0,01145 | 0,063187 | 0,856417 | Dyslipidemias | 167,6864 |
| ieu-b-108 | Incident CKD | apolipoprotein B | Weighted median | 183 | -0,03503 | 0,05011 | 0,484469 | Dyslipidemias | 167,6864 |
| ieu-b-108 | Incident CKD | apolipoprotein B | Inverse variance weighted | 183 | -0,04104 | 0,040302 | 0,308481 | Dyslipidemias | 167,6864 |
| ieu-b-108 | Incident CKD | apolipoprotein B | Simple mode | 183 | 0,141953 | 0,123637 | 0,252419 | Dyslipidemias | 167,6864 |
| ieu-b-108 | Incident CKD | apolipoprotein B | Weighted mode | 183 | -0,02098 | 0,051235 | 0,682732 | Dyslipidemias | 167,6864 |
| ieu-b-25 | Incident CKD | Cigarettes per Day | MR Egger | 22 | -2,07E-05 | 0,067173 | 0,999758 | Smoking | 99,80012 |
| ieu-b-25 | Incident CKD | Cigarettes per Day | Weighted median | 22 | 0,047719 | 0,050675 | 0,346369 | Smoking | 99,80012 |
| ieu-b-25 | Incident CKD | Cigarettes per Day | Inverse variance weighted | 22 | 0,003761 | 0,037646 | 0,920424 | Smoking | 99,80012 |
| ieu-b-25 | Incident CKD | Cigarettes per Day | Simple mode | 22 | 0,0902 | 0,102547 | 0,38903 | Smoking | 99,80012 |
| ieu-b-25 | Incident CKD | Cigarettes per Day | Weighted mode | 22 | 0,028629 | 0,046824 | 0,547483 | Smoking | 99,80012 |
| ieu-b-4861 | Incident CKD | Chronotype | MR Egger | 62 | -0,02453 | 0,172688 | 0,887503 | Sleep disorders | 44,20616 |
| ieu-b-4861 | Incident CKD | Chronotype | Weighted median | 62 | 0,045337 | 0,075222 | 0,5467 | Sleep disorders | 44,20616 |
| ieu-b-4861 | Incident CKD | Chronotype | Inverse variance weighted | 62 | 0,035427 | 0,062998 | 0,573879 | Sleep disorders | 44,20616 |
| ieu-b-4861 | Incident CKD | Chronotype | Simple mode | 62 | -0,08187 | 0,191184 | 0,669983 | Sleep disorders | 44,20616 |
| ieu-b-4861 | Incident CKD | Chronotype | Weighted mode | 62 | -0,07361 | 0,1657 | 0,658457 | Sleep disorders | 44,20616 |
| ieu-b-4877 | Incident CKD | smoking initiation | MR Egger | 85 | -0,19382 | 0,242588 | 0,426582 | Smoking | 42,2587 |
| ieu-b-4877 | Incident CKD | smoking initiation | Weighted median | 85 | -0,03298 | 0,06545 | 0,614291 | Smoking | 42,2587 |
| ieu-b-4877 | Incident CKD | smoking initiation | Inverse variance weighted | 85 | -0,01261 | 0,0476 | 0,791081 | Smoking | 42,2587 |
| ieu-b-4877 | Incident CKD | smoking initiation | Simple mode | 85 | 0,044958 | 0,158348 | 0,77717 | Smoking | 42,2587 |
| ieu-b-4877 | Incident CKD | smoking initiation | Weighted mode | 85 | -0,08947 | 0,150275 | 0,553214 | Smoking | 42,2587 |
| ieu-b-5144 | Incident CKD | Hypertension | MR Egger | 192 | 0,988741 | 0,656227 | 0,133547 | Hypertension | 53,57455 |
| ieu-b-5144 | Incident CKD | Hypertension | Weighted median | 192 | 0,656367 | 0,181311 | 0,000294 | Hypertension | 53,57455 |
| ieu-b-5144 | Incident CKD | Hypertension | Inverse variance weighted | 192 | 0,948142 | 0,213283 | 8,77E-06 | Hypertension | 53,57455 |
| ieu-b-5144 | Incident CKD | Hypertension | Simple mode | 192 | 0,584594 | 0,524767 | 0,266677 | Hypertension | 53,57455 |
| ieu-b-5144 | Incident CKD | Hypertension | Weighted mode | 192 | 0,642902 | 0,442002 | 0,147443 | Hypertension | 53,57455 |
| ieu-b-72 | Incident CKD | Urinary sodium-potassium ratio | MR Egger | 24 | 0,424141 | 0,904667 | 0,643799 | Minerals | 39,88328 |
| ieu-b-72 | Incident CKD | Urinary sodium-potassium ratio | Weighted median | 24 | -0,18811 | 0,213244 | 0,377693 | Minerals | 39,88328 |
| ieu-b-72 | Incident CKD | Urinary sodium-potassium ratio | Inverse variance weighted | 24 | -0,33491 | 0,209346 | 0,109644 | Minerals | 39,88328 |
| ieu-b-72 | Incident CKD | Urinary sodium-potassium ratio | Simple mode | 24 | -0,08327 | 0,407791 | 0,839991 | Minerals | 39,88328 |
| ieu-b-72 | Incident CKD | Urinary sodium-potassium ratio | Weighted mode | 24 | -0,03087 | 0,41 | 0,940638 | Minerals | 39,88328 |
| prot-a-3102 | Incident CKD | Thyrotropin-releasing hormone | Inverse variance weighted | 2 | 0,01372 | 0,049475 | 0,781533 | Thyroid disease | 36,35446 |
| prot-c-2652_15_1 | Incident CKD | suPAR | Wald ratio | 1 | -0,02773 | 0,038213 | 0,467957 | Biomarkers | 33,5125 |
| ukb-b-4424 | Incident CKD | Sleep duration | MR Egger | 66 | -0,79052 | 0,576465 | 0,175063 | Sleep disorders | 42,80283 |
| ukb-b-4424 | Incident CKD | Sleep duration | Weighted median | 66 | -0,14618 | 0,176233 | 0,406849 | Sleep disorders | 42,80283 |
| ukb-b-4424 | Incident CKD | Sleep duration | Inverse variance weighted | 66 | -0,06883 | 0,15155 | 0,649709 | Sleep disorders | 42,80283 |
| ukb-b-4424 | Incident CKD | Sleep duration | Simple mode | 66 | 0,060985 | 0,448975 | 0,892375 | Sleep disorders | 42,80283 |
| ukb-b-4424 | Incident CKD | Sleep duration | Weighted mode | 66 | -0,18199 | 0,326191 | 0,578804 | Sleep disorders | 42,80283 |
| ukb-b-5779 | Incident CKD | Alcohol intake frequency. | MR Egger | 95 | 0,037485 | 0,138666 | 0,787508 | Alcohol consumption | 52,90557 |
| ukb-b-5779 | Incident CKD | Alcohol intake frequency. | Weighted median | 95 | 0,266293 | 0,081087 | 0,001023 | Alcohol consumption | 52,90557 |
| ukb-b-5779 | Incident CKD | Alcohol intake frequency. | Inverse variance weighted | 95 | 0,122854 | 0,060612 | 0,042672 | Alcohol consumption | 52,90557 |
| ukb-b-5779 | Incident CKD | Alcohol intake frequency. | Simple mode | 95 | 0,329168 | 0,180914 | 0,072022 | Alcohol consumption | 52,90557 |
| ukb-b-5779 | Incident CKD | Alcohol intake frequency. | Weighted mode | 95 | 0,283727 | 0,097772 | 0,00462 | Alcohol consumption | 52,90557 |
| ukb-b-7864 | Incident CKD | Vitamin B6 | Wald ratio | 1 | -0,03795 | 0,324056 | 0,906767 | Vitamin | 32,75897 |
| ukb-d-30790_irnt | Incident CKD | Lipoprotein A | MR Egger | 19 | -0,04209 | 0,042386 | 0,334636 | Dyslipidemias | 920,8256 |
| ukb-d-30790_irnt | Incident CKD | Lipoprotein A | Weighted median | 19 | -0,00579 | 0,036562 | 0,874194 | Dyslipidemias | 920,8256 |
| ukb-d-30790_irnt | Incident CKD | Lipoprotein A | Inverse variance weighted | 19 | -0,00123 | 0,026778 | 0,963463 | Dyslipidemias | 920,8256 |
| ukb-d-30790_irnt | Incident CKD | Lipoprotein A | Simple mode | 19 | -0,01668 | 0,05601 | 0,76927 | Dyslipidemias | 920,8256 |
| ukb-d-30790_irnt | Incident CKD | Lipoprotein A | Weighted mode | 19 | 0,014043 | 0,047386 | 0,770361 | Dyslipidemias | 920,8256 |

| | | | | | | | | | |
|---|---|---|---|---|---|---|---|---|---|
| BMI | eGFR decline (General population) | Body mass index | MR Egger | 505 | -0,00921 | 0,035656 | 0,796324 | Obesity | 74,21321 |
| BMI | eGFR decline (General population) | Body mass index | Weighted median | 505 | 0,021232 | 0,025603 | 0,406942 | Obesity | 74,21321 |
| BMI | eGFR decline (General population) | Body mass index | Inverse variance weighted | 505 | 0,024372 | 0,013704 | 0,075333 | Obesity | 74,21321 |
| BMI | eGFR decline (General population) | Body mass index | Simple mode | 505 | 0,007567 | 0,077526 | 0,922281 | Obesity | 74,21321 |
| BMI | eGFR decline (General population) | Body mass index | Weighted mode | 505 | 0,001351 | 0,039068 | 0,972434 | Obesity | 74,21321 |
| DBPTang | eGFR decline (General population) | Diastolic blood pressure | MR Egger | 268 | -0,05956 | 0,073765 | 0,420137 | Hypertension | 55,53771 |
| DBPTang | eGFR decline (General population) | Diastolic blood pressure | Weighted median | 268 | 0,009218 | 0,027314 | 0,735761 | Hypertension | 55,53771 |
| DBPTang | eGFR decline (General population) | Diastolic blood pressure | Inverse variance weighted | 268 | 0,038357 | 0,02419 | 0,112818 | Hypertension | 55,53771 |
| DBPTang | eGFR decline (General population) | Diastolic blood pressure | Simple mode | 268 | -0,01731 | 0,0954 | 0,856157 | Hypertension | 55,53771 |
| DBPTang | eGFR decline (General population) | Diastolic blood pressure | Weighted mode | 268 | -0,00753 | 0,080353 | 0,925433 | Hypertension | 55,53771 |
| FT3 | eGFR decline (General population) | Free triiodothyronine | MR Egger | 6 | 0,075536 | 0,155285 | 0,652107 | Thyroid disease | 82,22035 |
| FT3 | eGFR decline (General population) | Free triiodothyronine | Weighted median | 6 | -0,0449 | 0,043044 | 0,29686 | Thyroid disease | 82,22035 |
| FT3 | eGFR decline (General population) | Free triiodothyronine | Inverse variance weighted | 6 | -0,03774 | 0,034169 | 0,269363 | Thyroid disease | 82,22035 |
| FT3 | eGFR decline (General population) | Free triiodothyronine | Simple mode | 6 | -0,04515 | 0,056017 | 0,456819 | Thyroid disease | 82,22035 |
| FT3 | eGFR decline (General population) | Free triiodothyronine | Weighted mode | 6 | -0,04811 | 0,052157 | 0,398666 | Thyroid disease | 82,22035 |
| FT3FT4 | eGFR decline (General population) | FT3 FT4 ratio | MR Egger | 13 | -0,14014 | 0,323945 | 0,673651 | Thyroid disease | 132,3008 |
| FT3FT4 | eGFR decline (General population) | FT3 FT4 ratio | Weighted median | 13 | -0,15196 | 0,129247 | 0,239695 | Thyroid disease | 132,3008 |
| FT3FT4 | eGFR decline (General population) | FT3 FT4 ratio | Inverse variance weighted | 13 | -0,12452 | 0,153519 | 0,417317 | Thyroid disease | 132,3008 |
| FT3FT4 | eGFR decline (General population) | FT3 FT4 ratio | Simple mode | 13 | 0,114862 | 0,242278 | 0,643944 | Thyroid disease | 132,3008 |
| FT3FT4 | eGFR decline (General population) | FT3 FT4 ratio | Weighted mode | 13 | -0,10494 | 0,123259 | 0,411222 | Thyroid disease | 132,3008 |
| FT4 | eGFR decline (General population) | Free thyroxine | MR Egger | 63 | 0,008245 | 0,034999 | 0,814549 | Thyroid disease | 83,6751 |
| FT4 | eGFR decline (General population) | Free thyroxine | Weighted median | 63 | 0,006455 | 0,023407 | 0,782725 | Thyroid disease | 83,6751 |
| FT4 | eGFR decline (General population) | Free thyroxine | Inverse variance weighted | 63 | 0,003624 | 0,016474 | 0,825859 | Thyroid disease | 83,6751 |
| FT4 | eGFR decline (General population) | Free thyroxine | Simple mode | 63 | -0,01336 | 0,043531 | 0,759984 | Thyroid disease | 83,6751 |
| FT4 | eGFR decline (General population) | Free thyroxine | Weighted mode | 63 | 0,015923 | 0,023969 | 0,508961 | Thyroid disease | 83,6751 |
| Fe | eGFR decline (General population) | Serum iron | Inverse variance weighted | 2 | -0,11194 | 0,132531 | 0,398297 | Minerals | 291,9629 |
| HDL | eGFR decline (General population) | HDL cholesterol | MR Egger | 251 | -0,02134 | 0,017099 | 0,213129 | Dyslipidemias | 271,1249 |
| HDL | eGFR decline (General population) | HDL cholesterol | Weighted median | 251 | -0,01768 | 0,017851 | 0,322092 | Dyslipidemias | 271,1249 |
| HDL | eGFR decline (General population) | HDL cholesterol | Inverse variance weighted | 251 | -0,02849 | 0,011264 | 0,011435 | Dyslipidemias | 271,1249 |
| HDL | eGFR decline (General population) | HDL cholesterol | Simple mode | 251 | -0,02329 | 0,040468 | 0,56554 | Dyslipidemias | 271,1249 |
| HDL | eGFR decline (General population) | HDL cholesterol | Weighted mode | 251 | 0,001855 | 0,01829 | 0,919319 | Dyslipidemias | 271,1249 |
| HiTSH | eGFR decline (General population) | High TSH | MR Egger | 6 | 0,074285 | 0,116059 | 0,556956 | Thyroid disease | 49,53497 |
| HiTSH | eGFR decline (General population) | High TSH | Weighted median | 6 | -0,01246 | 0,029434 | 0,671971 | Thyroid disease | 49,53497 |
| HiTSH | eGFR decline (General population) | High TSH | Inverse variance weighted | 6 | -0,00833 | 0,02684 | 0,756188 | Thyroid disease | 49,53497 |
| HiTSH | eGFR decline (General population) | High TSH | Simple mode | 6 | -0,02568 | 0,047828 | 0,614404 | Thyroid disease | 49,53497 |
| HiTSH | eGFR decline (General population) | High TSH | Weighted mode | 6 | 0,002279 | 0,03331 | 0,948106 | Thyroid disease | 49,53497 |
| Insom | eGFR decline (General population) | Insomnia | MR Egger | 13 | 0,075693 | 0,123843 | 0,553486 | Sleep disorders | 44,05724 |
| Insom | eGFR decline (General population) | Insomnia | Weighted median | 13 | 0,045434 | 0,050239 | 0,365809 | Sleep disorders | 44,05724 |
| Insom | eGFR decline (General population) | Insomnia | Inverse variance weighted | 13 | 0,047691 | 0,038521 | 0,215698 | Sleep disorders | 44,05724 |
| Insom | eGFR decline (General population) | Insomnia | Simple mode | 13 | 0,039381 | 0,070672 | 0,58761 | Sleep disorders | 44,05724 |
| Insom | eGFR decline (General population) | Insomnia | Weighted mode | 13 | 0,045376 | 0,070495 | 0,531896 | Sleep disorders | 44,05724 |
| LDL | eGFR decline (General population) | LDL cholesterol | MR Egger | 231 | 0,034661 | 0,018143 | 0,057332 | Dyslipidemias | 239,3726 |
| LDL | eGFR decline (General population) | LDL cholesterol | Weighted median | 231 | 0,015653 | 0,018882 | 0,407111 | Dyslipidemias | 239,3726 |
| LDL | eGFR decline (General population) | LDL cholesterol | Inverse variance weighted | 231 | -0,00258 | 0,012081 | 0,830982 | Dyslipidemias | 239,3726 |
| LDL | eGFR decline (General population) | LDL cholesterol | Simple mode | 231 | -0,04396 | 0,036786 | 0,233259 | Dyslipidemias | 239,3726 |
| LDL | eGFR decline (General population) | LDL cholesterol | Weighted mode | 231 | 0,028658 | 0,019477 | 0,142563 | Dyslipidemias | 239,3726 |
| LoTSH | eGFR decline (General population) | Low TSH | MR Egger | 27 | 0,021605 | 0,013558 | 0,123621 | Thyroid disease | 72,92792 |
| LoTSH | eGFR decline (General population) | Low TSH | Weighted median | 27 | 0,015977 | 0,006579 | 0,01516 | Thyroid disease | 72,92792 |
| LoTSH | eGFR decline (General population) | Low TSH | Inverse variance weighted | 27 | 0,013631 | 0,005195 | 0,008687 | Thyroid disease | 72,92792 |
| LoTSH | eGFR decline (General population) | Low TSH | Simple mode | 27 | 0,018032 | 0,012114 | 0,148642 | Thyroid disease | 72,92792 |
| LoTSH | eGFR decline (General population) | Low TSH | Weighted mode | 27 | 0,019776 | 0,008846 | 0,034186 | Thyroid disease | 72,92792 |
| PPTang | eGFR decline (General population) | Pulse pressure | MR Egger | 315 | 0,023658 | 0,041463 | 0,568686 | Hypertension | 69,42147 |
| PPTang | eGFR decline (General population) | Pulse pressure | Weighted median | 315 | 0,059234 | 0,02491 | 0,01741 | Hypertension | 69,42147 |
| PPTang | eGFR decline (General population) | Pulse pressure | Inverse variance weighted | 315 | 0,059374 | 0,016214 | 0,00025 | Hypertension | 69,42147 |
| PPTang | eGFR decline (General population) | Pulse pressure | Simple mode | 315 | 0,043261 | 0,082511 | 0,600441 | Hypertension | 69,42147 |
| PPTang | eGFR decline (General population) | Pulse pressure | Weighted mode | 315 | 0,037014 | 0,075508 | 0,624332 | Hypertension | 69,42147 |
| SBPTang | eGFR decline (General population) | Systolic blood pressure | MR Egger | 266 | -0,03115 | 0,057524 | 0,588662 | Hypertension | 57,08057 |
| SBPTang | eGFR decline (General population) | Systolic blood pressure | Weighted median | 266 | 0,063078 | 0,028022 | 0,024387 | Hypertension | 57,08057 |
| SBPTang | eGFR decline (General population) | Systolic blood pressure | Inverse variance weighted | 266 | 0,062551 | 0,019236 | 0,001147 | Hypertension | 57,08057 |
| SBPTang | eGFR decline (General population) | Systolic blood pressure | Simple mode | 266 | 0,094178 | 0,090899 | 0,301116 | Hypertension | 57,08057 |
| SBPTang | eGFR decline (General population) | Systolic blood pressure | Weighted mode | 266 | 0,07844 | 0,080139 | 0,32857 | Hypertension | 57,08057 |
| SIndex | eGFR decline (General population) | Compound smoking index | MR Egger | 120 | 0,421804 | 0,177373 | 0,01901 | Smoking | 44,08236 |
| SIndex | eGFR decline (General population) | Compound smoking index | Weighted median | 120 | 0,090435 | 0,065959 | 0,17035 | Smoking | 44,08236 |
| SIndex | eGFR decline (General population) | Compound smoking index | Inverse variance weighted | 120 | 0,06916 | 0,045165 | 0,125704 | Smoking | 44,08236 |
| SIndex | eGFR decline (General population) | Compound smoking index | Simple mode | 120 | 0,13809 | 0,17914 | 0,442323 | Smoking | 44,08236 |
| SIndex | eGFR decline (General population) | Compound smoking index | Weighted mode | 120 | 0,095462 | 0,182867 | 0,60262 | Smoking | 44,08236 |
| T2DGGI | eGFR decline (General population) | Type 2 diabetes | MR Egger | 451 | -0,00369 | 0,012146 | 0,761694 | Diabetes | 94,60617 |
| T2DGGI | eGFR decline (General population) | Type 2 diabetes | Weighted median | 451 | 0,008475 | 0,010075 | 0,400248 | Diabetes | 94,60617 |
| T2DGGI | eGFR decline (General population) | Type 2 diabetes | Inverse variance weighted | 451 | 0,004176 | 0,005985 | 0,48525 | Diabetes | 94,60617 |
| T2DGGI | eGFR decline (General population) | Type 2 diabetes | Simple mode | 451 | 0,015721 | 0,027253 | 0,564329 | Diabetes | 94,60617 |
| T2DGGI | eGFR decline (General population) | Type 2 diabetes | Weighted mode | 451 | 0,019513 | 0,013499 | 0,149009 | Diabetes | 94,60617 |
| TC | eGFR decline (General population) | Total cholesterol | MR Egger | 279 | 0,018335 | 0,016264 | 0,260559 | Dyslipidemias | 252,8943 |
| TC | eGFR decline (General population) | Total cholesterol | Weighted median | 279 | 0,01346 | 0,017607 | 0,44459 | Dyslipidemias | 252,8943 |
| TC | eGFR decline (General population) | Total cholesterol | Inverse variance weighted | 279 | -0,00109 | 0,010731 | 0,919073 | Dyslipidemias | 252,8943 |
| TC | eGFR decline (General population) | Total cholesterol | Simple mode | 279 | 0,051294 | 0,036048 | 0,155877 | Dyslipidemias | 252,8943 |
| TC | eGFR decline (General population) | Total cholesterol | Weighted mode | 279 | 0,018498 | 0,018878 | 0,328012 | Dyslipidemias | 252,8943 |
| TG | eGFR decline (General population) | Triglyceride | MR Egger | 231 | 0,024261 | 0,018654 | 0,194715 | Dyslipidemias | 236,2982 |
| TG | eGFR decline (General population) | Triglyceride | Weighted median | 231 | 0,039224 | 0,018207 | 0,031216 | Dyslipidemias | 236,2982 |
| TG | eGFR decline (General population) | Triglyceride | Inverse variance weighted | 231 | 0,027597 | 0,012335 | 0,02527 | Dyslipidemias | 236,2982 |
| TG | eGFR decline (General population) | Triglyceride | Simple mode | 231 | -0,0064 | 0,038468 | 0,867914 | Dyslipidemias | 236,2982 |
| TG | eGFR decline (General population) | Triglyceride | Weighted mode | 231 | 0,030337 | 0,017145 | 0,078148 | Dyslipidemias | 236,2982 |
| TSH | eGFR decline (General population) | Thyroid Stimulating Hormone | MR Egger | 164 | -0,01027 | 0,01707 | 0,548178 | Thyroid disease | 152,7855 |
| TSH | eGFR decline (General population) | Thyroid Stimulating Hormone | Weighted median | 164 | 7,90E-06 | 0,015411 | 0,999591 | Thyroid disease | 152,7855 |
| TSH | eGFR decline (General population) | Thyroid Stimulating Hormone | Inverse variance weighted | 164 | 0,005455 | 0,00958 | 0,569054 | Thyroid disease | 152,7855 |
| TSH | eGFR decline (General population) | Thyroid Stimulating Hormone | Simple mode | 164 | 0,004624 | 0,033902 | 0,891687 | Thyroid disease | 152,7855 |
| TSH | eGFR decline (General population) | Thyroid Stimulating Hormone | Weighted mode | 164 | -0,00526 | 0,01815 | 0,772515 | Thyroid disease | 152,7855 |
| WHR | eGFR decline (General population) | Waist-to-hip ratio adjusted BMI | MR Egger | 296 | -0,02378 | 0,040115 | 0,553691 | Obesity | 85,78362 |
| WHR | eGFR decline (General population) | Waist-to-hip ratio adjusted BMI | Weighted median | 296 | -0,02221 | 0,028246 | 0,431626 | Obesity | 85,78362 |
| WHR | eGFR decline (General population) | Waist-to-hip ratio adjusted BMI | Inverse variance weighted | 296 | 0,004115 | 0,016998 | 0,808692 | Obesity | 85,78362 |
| WHR | eGFR decline (General population) | Waist-to-hip ratio adjusted BMI | Simple mode | 296 | 0,034385 | 0,072741 | 0,636773 | Obesity | 85,78362 |
| WHR | eGFR decline (General population) | Waist-to-hip ratio adjusted BMI | Weighted mode | 296 | -0,02597 | 0,044686 | 0,561531 | Obesity | 85,78362 |
| Zn | eGFR decline (General population) | Zinc | MR Egger | 3 | 0,011382 | 0,053131 | 0,865651 | Minerals | 99,95282 |
| Zn | eGFR decline (General population) | Zinc | Weighted median | 3 | 0,021501 | 0,015723 | 0,171487 | Minerals | 99,95282 |
| Zn | eGFR decline (General population) | Zinc | Inverse variance weighted | 3 | 0,02468 | 0,014548 | 0,089812 | Minerals | 99,95282 |
| Zn | eGFR decline (General population) | Zinc | Simple mode | 3 | 0,008839 | 0,02434 | 0,751274 | Minerals | 99,95282 |
| Zn | eGFR decline (General population) | Zinc | Weighted mode | 3 | 0,012135 | 0,019108 | 0,590352 | Minerals | 99,95282 |
| ebi-a-GCST006979-consort | eGFR decline (General population) | Heel bone mineral density | MR Egger | 499 | 0,010328 | 0,017195 | 0,54837 | Bone disease | 152,4254 |
| ebi-a-GCST006979-consort | eGFR decline (General population) | Heel bone mineral density | Weighted median | 499 | 0,014227 | 0,013479 | 0,291175 | Bone disease | 152,4254 |
| ebi-a-GCST006979-consort | eGFR decline (General population) | Heel bone mineral density | Inverse variance weighted | 499 | 0,005894 | 0,009134 | 0,518739 | Bone disease | 152,4254 |
| ebi-a-GCST006979-consort | eGFR decline (General population) | Heel bone mineral density | Simple mode | 499 | 0,031261 | 0,035586 | 0,380122 | Bone disease | 152,4254 |
| ebi-a-GCST006979-consort | eGFR decline (General population) | Heel bone mineral density | Weighted mode | 499 | 0,006267 | 0,019902 | 0,752966 | Bone disease | 152,4254 |
| ebi-a-GCST90002227 | eGFR decline (General population) | Two-hour glucose | MR Egger | 14 | 0,035974 | 0,052991 | 0,510106 | Diabetes | 56,47851 |
| ebi-a-GCST90002227 | eGFR decline (General population) | Two-hour glucose | Weighted median | 14 | 0,003069 | 0,026474 | 0,907698 | Diabetes | 56,47851 |
| ebi-a-GCST90002227 | eGFR decline (General population) | Two-hour glucose | Inverse variance weighted | 14 | 0,010804 | 0,019766 | 0,584658 | Diabetes | 56,47851 |
| ebi-a-GCST90002227 | eGFR decline (General population) | Two-hour glucose | Simple mode | 14 | 0,008815 | 0,040989 | 0,833055 | Diabetes | 56,47851 |
| ebi-a-GCST90002227 | eGFR decline (General population) | Two-hour glucose | Weighted mode | 14 | -0,00254 | 0,036886 | 0,946152 | Diabetes | 56,47851 |
| ebi-a-GCST90002232 | eGFR decline (General population) | Fasting glucose | MR Egger | 69 | -0,07606 | 0,058036 | 0,194473 | Diabetes | 122,8115 |
| ebi-a-GCST90002232 | eGFR decline (General population) | Fasting glucose | Weighted median | 69 | -0,03129 | 0,044791 | 0,484839 | Diabetes | 122,8115 |
| ebi-a-GCST90002232 | eGFR decline (General population) | Fasting glucose | Inverse variance weighted | 69 | -0,03201 | 0,030414 | 0,292585 | Diabetes | 122,8115 |
| ebi-a-GCST90002232 | eGFR decline (General population) | Fasting glucose | Simple mode | 69 | -0,03069 | 0,087743 | 0,727604 | Diabetes | 122,8115 |
| ebi-a-GCST90002232 | eGFR decline (General population) | Fasting glucose | Weighted mode | 69 | -0,02609 | 0,04583 | 0,571052 | Diabetes | 122,8115 |
| ebi-a-GCST90002238 | eGFR decline (General population) | Fasting insulin | MR Egger | 38 | -0,04831 | 0,187689 | 0,79833 | Diabetes | 51,80185 |
| ebi-a-GCST90002238 | eGFR decline (General population) | Fasting insulin | Weighted median | 38 | 0,011395 | 0,080091 | 0,886863 | Diabetes | 51,80185 |

| | | | | | | | | | |
|---|---|---|---|---|---|---|---|---|---|
| ebi-a-GCST90002238 | eGFR decline (General population) | Fasting insulin | Inverse variance weighted | 38 | 0,003459 | 0,06118 | 0,954909 | Diabetes | 51,80185 |
| ebi-a-GCST90002238 | eGFR decline (General population) | Fasting insulin | Simple mode | 38 | -0,17396 | 0,172327 | 0,319294 | Diabetes | 51,80185 |
| ebi-a-GCST90002238 | eGFR decline (General population) | Fasting insulin | Weighted mode | 38 | -0,15567 | 0,146443 | 0,294681 | Diabetes | 51,80185 |
| ebi-a-GCST90007310 | eGFR decline (General population) | circulating leptin levels | MR Egger | 3 | 0,167442 | 0,399361 | 0,747255 | Obesity | 56,18818 |
| ebi-a-GCST90007310 | eGFR decline (General population) | circulating leptin levels | Weighted median | 3 | -0,06341 | 0,058106 | 0,275124 | Obesity | 56,18818 |
| ebi-a-GCST90007310 | eGFR decline (General population) | circulating leptin levels | Inverse variance weighted | 3 | -0,0484 | 0,049622 | 0,329389 | Obesity | 56,18818 |
| ebi-a-GCST90007310 | eGFR decline (General population) | circulating leptin levels | Simple mode | 3 | -0,09426 | 0,075997 | 0,340634 | Obesity | 56,18818 |
| ebi-a-GCST90007310 | eGFR decline (General population) | circulating leptin levels | Weighted mode | 3 | -0,09251 | 0,079139 | 0,362912 | Obesity | 56,18818 |
| ebi-a-GCST90011998 | eGFR decline (General population) | Growth differentiation factor-15 levels | MR Egger | 3 | -0,03889 | 0,038633 | 0,497864 | Biomarkers | 294,0397 |
| ebi-a-GCST90011998 | eGFR decline (General population) | Growth differentiation factor-15 levels | Weighted median | 3 | -0,01951 | 0,017538 | 0,266047 | Biomarkers | 294,0397 |
| ebi-a-GCST90011998 | eGFR decline (General population) | Growth differentiation factor-15 levels | Inverse variance weighted | 3 | -0,01449 | 0,023713 | 0,541296 | Biomarkers | 294,0397 |
| ebi-a-GCST90011998 | eGFR decline (General population) | Growth differentiation factor-15 levels | Simple mode | 3 | -0,01799 | 0,034364 | 0,652823 | Biomarkers | 294,0397 |
| ebi-a-GCST90011998 | eGFR decline (General population) | Growth differentiation factor-15 levels | Weighted mode | 3 | -0,01945 | 0,017846 | 0,389555 | Biomarkers | 294,0397 |
| ebi-a-GCST90012022 | eGFR decline (General population) | Fibroblast growth factor 23 levels | MR Egger | 8 | -0,01406 | 0,082233 | 0,869883 | Biomarkers | 43,42098 |
| ebi-a-GCST90012022 | eGFR decline (General population) | Fibroblast growth factor 23 levels | Weighted median | 8 | -0,02812 | 0,028514 | 0,324132 | Biomarkers | 43,42098 |
| ebi-a-GCST90012022 | eGFR decline (General population) | Fibroblast growth factor 23 levels | Inverse variance weighted | 8 | -0,02275 | 0,021621 | 0,292722 | Biomarkers | 43,42098 |
| ebi-a-GCST90012022 | eGFR decline (General population) | Fibroblast growth factor 23 levels | Simple mode | 8 | -0,0386 | 0,045552 | 0,42485 | Biomarkers | 43,42098 |
| ebi-a-GCST90012022 | eGFR decline (General population) | Fibroblast growth factor 23 levels | Weighted mode | 8 | -0,0379 | 0,037709 | 0,348294 | Biomarkers | 43,42098 |
| ebi-a-GCST90012041 | eGFR decline (General population) | Kidney injury molecule 1 levels | MR Egger | 10 | -0,03382 | 0,070102 | 0,642388 | Biomarkers | 190,9807 |
| ebi-a-GCST90012041 | eGFR decline (General population) | Kidney injury molecule 1 levels | Weighted median | 10 | -0,01783 | 0,011408 | 0,118118 | Biomarkers | 190,9807 |
| ebi-a-GCST90012041 | eGFR decline (General population) | Kidney injury molecule 1 levels | Inverse variance weighted | 10 | 0,02161 | 0,045046 | 0,631411 | Biomarkers | 190,9807 |
| ebi-a-GCST90012041 | eGFR decline (General population) | Kidney injury molecule 1 levels | Simple mode | 10 | -0,0277 | 0,024054 | 0,279159 | Biomarkers | 190,9807 |
| ebi-a-GCST90012041 | eGFR decline (General population) | Kidney injury molecule 1 levels | Weighted mode | 10 | -0,01831 | 0,011005 | 0,130522 | Biomarkers | 190,9807 |
| ebi-a-GCST90013864 | eGFR decline (General population) | Coronary artery disease (Firth correction) | MR Egger | 40 | -0,00348 | 0,023569 | 0,883506 | Cardiovascular disease | 59,88817 |
| ebi-a-GCST90013864 | eGFR decline (General population) | Coronary artery disease (Firth correction) | Weighted median | 40 | -0,02069 | 0,015859 | 0,191982 | Cardiovascular disease | 59,88817 |
| ebi-a-GCST90013864 | eGFR decline (General population) | Coronary artery disease (Firth correction) | Inverse variance weighted | 40 | -0,0055 | 0,010533 | 0,60167 | Cardiovascular disease | 59,88817 |
| ebi-a-GCST90013864 | eGFR decline (General population) | Coronary artery disease (Firth correction) | Simple mode | 40 | -0,01672 | 0,029495 | 0,57404 | Cardiovascular disease | 59,88817 |
| ebi-a-GCST90013864 | eGFR decline (General population) | Coronary artery disease (Firth correction) | Weighted mode | 40 | -0,01793 | 0,019164 | 0,355153 | Cardiovascular disease | 59,88817 |
| ebi-a-GCST90013975 | eGFR decline (General population) | Body fat percentage | MR Egger | 328 | 0,076405 | 0,0681 | 0,262706 | Obesity | 56,01289 |
| ebi-a-GCST90013975 | eGFR decline (General population) | Body fat percentage | Weighted median | 328 | 0,046133 | 0,033507 | 0,168572 | Obesity | 56,01289 |
| ebi-a-GCST90013975 | eGFR decline (General population) | Body fat percentage | Inverse variance weighted | 328 | 0,057814 | 0,021269 | 0,006563 | Obesity | 56,01289 |
| ebi-a-GCST90013975 | eGFR decline (General population) | Body fat percentage | Simple mode | 328 | 0,093143 | 0,107316 | 0,38607 | Obesity | 56,01289 |
| ebi-a-GCST90013975 | eGFR decline (General population) | Body fat percentage | Weighted mode | 328 | 0,037636 | 0,075472 | 0,618343 | Obesity | 56,01289 |
| ebi-a-GCST90013989 | eGFR decline (General population) | Sodium in urine | MR Egger | 41 | 0,017772 | 0,238088 | 0,940878 | Minerals | 43,81211 |
| ebi-a-GCST90013989 | eGFR decline (General population) | Sodium in urine | Weighted median | 41 | 0,069914 | 0,074937 | 0,350834 | Minerals | 43,81211 |
| ebi-a-GCST90013989 | eGFR decline (General population) | Sodium in urine | Inverse variance weighted | 41 | 0,094113 | 0,059509 | 0,11377 | Minerals | 43,81211 |
| ebi-a-GCST90013989 | eGFR decline (General population) | Sodium in urine | Simple mode | 41 | 0,109457 | 0,139493 | 0,437266 | Minerals | 43,81211 |
| ebi-a-GCST90013989 | eGFR decline (General population) | Sodium in urine | Weighted mode | 41 | 0,06253 | 0,117932 | 0,598893 | Minerals | 43,81211 |
| ebi-a-GCST90014006 | eGFR decline (General population) | Glycated haemoglobin HbA1c levels | MR Egger | 325 | -0,005 | 0,020855 | 0,810607 | Diabetes | 128,5009 |
| ebi-a-GCST90014006 | eGFR decline (General population) | Glycated haemoglobin HbA1c levels | Weighted median | 325 | -0,008 | 0,018038 | 0,657588 | Diabetes | 128,5009 |
| ebi-a-GCST90014006 | eGFR decline (General population) | Glycated haemoglobin HbA1c levels | Inverse variance weighted | 325 | 0,00819 | 0,011124 | 0,473421 | Diabetes | 128,5009 |
| ebi-a-GCST90014006 | eGFR decline (General population) | Glycated haemoglobin HbA1c levels | Simple mode | 325 | 0,09257 | 0,045562 | 0,042997 | Diabetes | 128,5009 |
| ebi-a-GCST90014006 | eGFR decline (General population) | Glycated haemoglobin HbA1c levels | Weighted mode | 325 | -0,00597 | 0,022701 | 0,792782 | Diabetes | 128,5009 |
| ebi-a-GCST90014010 | eGFR decline (General population) | Phosphate levels | MR Egger | 167 | -0,02663 | 0,029739 | 0,371917 | Minerals | 88,79967 |
| ebi-a-GCST90014010 | eGFR decline (General population) | Phosphate levels | Weighted median | 167 | -0,05023 | 0,025231 | 0,046522 | Minerals | 88,79967 |
| ebi-a-GCST90014010 | eGFR decline (General population) | Phosphate levels | Inverse variance weighted | 167 | -0,01413 | 0,016851 | 0,401632 | Minerals | 88,79967 |
| ebi-a-GCST90014010 | eGFR decline (General population) | Phosphate levels | Simple mode | 167 | -0,04208 | 0,053829 | 0,435467 | Minerals | 88,79967 |
| ebi-a-GCST90014010 | eGFR decline (General population) | Phosphate levels | Weighted mode | 167 | -0,04208 | 0,028671 | 0,144064 | Minerals | 88,79967 |
| ebi-a-GCST90014022 | eGFR decline (General population) | Bone mineral density | MR Egger | 437 | 0,00936 | 0,017849 | 0,600266 | Bone disease | 120,5445 |
| ebi-a-GCST90014022 | eGFR decline (General population) | Bone mineral density | Weighted median | 437 | 0,00509 | 0,014775 | 0,73048 | Bone disease | 120,5445 |
| ebi-a-GCST90014022 | eGFR decline (General population) | Bone mineral density | Inverse variance weighted | 437 | 0,006499 | 0,00932 | 0,485632 | Bone disease | 120,5445 |
| ebi-a-GCST90014022 | eGFR decline (General population) | Bone mineral density | Simple mode | 437 | 0,019582 | 0,034572 | 0,571409 | Bone disease | 120,5445 |
| ebi-a-GCST90014022 | eGFR decline (General population) | Bone mineral density | Weighted mode | 437 | 0,001008 | 0,020213 | 0,960263 | Bone disease | 120,5445 |
| ebi-a-GCST90014023 | eGFR decline (General population) | Type 1 diabetes | MR Egger | 78 | 0,003074 | 0,005103 | 0,548727 | Diabetes | 161,9002 |
| ebi-a-GCST90014023 | eGFR decline (General population) | Type 1 diabetes | Weighted median | 78 | 0,004094 | 0,004887 | 0,402183 | Diabetes | 161,9002 |
| ebi-a-GCST90014023 | eGFR decline (General population) | Type 1 diabetes | Inverse variance weighted | 78 | 0,002141 | 0,003199 | 0,503378 | Diabetes | 161,9002 |
| ebi-a-GCST90014023 | eGFR decline (General population) | Type 1 diabetes | Simple mode | 78 | 0,000607 | 0,009678 | 0,950161 | Diabetes | 161,9002 |
| ebi-a-GCST90014023 | eGFR decline (General population) | Type 1 diabetes | Weighted mode | 78 | 0,004058 | 0,004966 | 0,4163 | Diabetes | 161,9002 |
| ebi-a-GCST90016674 | eGFR decline (General population) | Liver iron content | MR Egger | 6 | 0,011946 | 0,022564 | 0,624547 | Minerals | 221,7646 |
| ebi-a-GCST90016674 | eGFR decline (General population) | Liver iron content | Weighted median | 6 | 0,030657 | 0,017477 | 0,079413 | Minerals | 221,7646 |
| ebi-a-GCST90016674 | eGFR decline (General population) | Liver iron content | Inverse variance weighted | 6 | 0,030362 | 0,015925 | 0,056583 | Minerals | 221,7646 |
| ebi-a-GCST90016674 | eGFR decline (General population) | Liver iron content | Simple mode | 6 | 0,07712 | 0,037965 | 0,09794 | Minerals | 221,7646 |
| ebi-a-GCST90016674 | eGFR decline (General population) | Liver iron content | Weighted mode | 6 | 0,029652 | 0,017611 | 0,153051 | Minerals | 221,7646 |
| ebi-a-GCST90016676 | eGFR decline (General population) | Pancreas iron content | Wald ratio | 1 | 0,057586 | 0,092138 | 0,531971 | Minerals | 30,56027 |
| ebi-a-GCST90018806 | eGFR decline (General population) | Chronic heart failure | MR Egger | 6 | 0,217198 | 0,12584 | 0,159428 | Cardiovascular disease | 40,14382 |
| ebi-a-GCST90018806 | eGFR decline (General population) | Chronic heart failure | Weighted median | 6 | 0,057434 | 0,037685 | 0,127498 | Cardiovascular disease | 40,14382 |
| ebi-a-GCST90018806 | eGFR decline (General population) | Chronic heart failure | Inverse variance weighted | 6 | 0,062401 | 0,031453 | 0,047261 | Cardiovascular disease | 40,14382 |
| ebi-a-GCST90018806 | eGFR decline (General population) | Chronic heart failure | Simple mode | 6 | 0,048679 | 0,055261 | 0,418699 | Cardiovascular disease | 40,14382 |
| ebi-a-GCST90018806 | eGFR decline (General population) | Chronic heart failure | Weighted mode | 6 | 0,043392 | 0,050038 | 0,425493 | Cardiovascular disease | 40,14382 |
| ebi-a-GCST90018860 | eGFR decline (General population) | Hyperthyroidism | MR Egger | 11 | -0,01926 | 0,022958 | 0,423177 | Thyroid disease | 63,8325 |
| ebi-a-GCST90018860 | eGFR decline (General population) | Hyperthyroidism | Weighted median | 11 | 0,006823 | 0,011651 | 0,55814 | Thyroid disease | 63,8325 |
| ebi-a-GCST90018860 | eGFR decline (General population) | Hyperthyroidism | Inverse variance weighted | 11 | 0,00968 | 0,008711 | 0,266429 | Thyroid disease | 63,8325 |
| ebi-a-GCST90018860 | eGFR decline (General population) | Hyperthyroidism | Simple mode | 11 | 0,019922 | 0,017396 | 0,278791 | Thyroid disease | 63,8325 |
| ebi-a-GCST90018860 | eGFR decline (General population) | Hyperthyroidism | Weighted mode | 11 | 0,013625 | 0,013389 | 0,332854 | Thyroid disease | 63,8325 |
| ebi-a-GCST90018862 | eGFR decline (General population) | Hypothyroidism | MR Egger | 68 | 0,001496 | 0,014952 | 0,920603 | Thyroid disease | 79,29446 |
| ebi-a-GCST90018862 | eGFR decline (General population) | Hypothyroidism | Weighted median | 68 | 0,010174 | 0,009859 | 0,302088 | Thyroid disease | 79,29446 |
| ebi-a-GCST90018862 | eGFR decline (General population) | Hypothyroidism | Inverse variance weighted | 68 | 0,005775 | 0,006352 | 0,363265 | Thyroid disease | 79,29446 |
| ebi-a-GCST90018862 | eGFR decline (General population) | Hypothyroidism | Simple mode | 68 | 0,016636 | 0,020728 | 0,42505 | Thyroid disease | 79,29446 |
| ebi-a-GCST90018862 | eGFR decline (General population) | Hypothyroidism | Weighted mode | 68 | 0,008847 | 0,012705 | 0,488634 | Thyroid disease | 79,29446 |
| ebi-a-GCST90018872 | eGFR decline (General population) | Iron deficiency anemia | Inverse variance weighted | 2 | 0,078169 | 0,117557 | 0,506086 | Minerals | 36,20402 |
| ebi-a-GCST90018877 | eGFR decline (General population) | Myocardial infarction | MR Egger | 77 | -0,00812 | 0,018825 | 0,667434 | Cardiovascular disease | 65,28777 |
| ebi-a-GCST90018877 | eGFR decline (General population) | Myocardial infarction | Weighted median | 77 | -0,01919 | 0,012298 | 0,118656 | Cardiovascular disease | 65,28777 |
| ebi-a-GCST90018877 | eGFR decline (General population) | Myocardial infarction | Inverse variance weighted | 77 | -0,00616 | 0,008326 | 0,459691 | Cardiovascular disease | 65,28777 |
| ebi-a-GCST90018877 | eGFR decline (General population) | Myocardial infarction | Simple mode | 77 | -0,0198 | 0,023521 | 0,402519 | Cardiovascular disease | 65,28777 |
| ebi-a-GCST90018877 | eGFR decline (General population) | Myocardial infarction | Weighted mode | 77 | -0,0198 | 0,014846 | 0,186251 | Cardiovascular disease | 65,28777 |
| ebi-a-GCST90018916 | eGFR decline (General population) | Sleep apnea syndrome | Wald ratio | 1 | -0,00386 | 0,041506 | 0,925885 | Sleep disorders | 63,50864 |
| ebi-a-GCST90018977 | eGFR decline (General population) | Serum uric acid levels | MR Egger | 260 | 0,002746 | 0,027469 | 0,920454 | Uricemia | 121,4825 |
| ebi-a-GCST90018977 | eGFR decline (General population) | Serum uric acid levels | Weighted median | 260 | 0,02428 | 0,021961 | 0,268913 | Uricemia | 121,4825 |
| ebi-a-GCST90018977 | eGFR decline (General population) | Serum uric acid levels | Inverse variance weighted | 260 | 0,040927 | 0,01749 | 0,019285 | Uricemia | 121,4825 |
| ebi-a-GCST90018977 | eGFR decline (General population) | Serum uric acid levels | Simple mode | 260 | 0,045955 | 0,044635 | 0,304171 | Uricemia | 121,4825 |
| ebi-a-GCST90018977 | eGFR decline (General population) | Serum uric acid levels | Weighted mode | 260 | 0,015142 | 0,020437 | 0,459411 | Uricemia | 121,4825 |
| ebi-a-GCST90019417 | eGFR decline (General population) | Vitamin K-dependent protein S measurement | MR Egger | 4 | 0,038434 | 0,032155 | 0,354485 | Vitamin | 118,3171 |
| ebi-a-GCST90019417 | eGFR decline (General population) | Vitamin K-dependent protein S measurement | Weighted median | 4 | -0,02522 | 0,022186 | 0,25562 | Vitamin | 118,3171 |
| ebi-a-GCST90019417 | eGFR decline (General population) | Vitamin K-dependent protein S measurement | Inverse variance weighted | 4 | -0,02517 | 0,029095 | 0,387057 | Vitamin | 118,3171 |
| ebi-a-GCST90019417 | eGFR decline (General population) | Vitamin K-dependent protein S measurement | Simple mode | 4 | -0,08342 | 0,042345 | 0,143431 | Vitamin | 118,3171 |
| ebi-a-GCST90019417 | eGFR decline (General population) | Vitamin K-dependent protein S measurement | Weighted mode | 4 | 0,022868 | 0,023114 | 0,39543 | Vitamin | 118,3171 |
| ebi-a-GCST90019423 | eGFR decline (General population) | Vitamin K-dependent protein C measurement | MR Egger | 4 | 0,038219 | 0,025702 | 0,275377 | Vitamin | 174,2568 |
| ebi-a-GCST90019423 | eGFR decline (General population) | Vitamin K-dependent protein C measurement | Weighted median | 4 | 0,000137 | 0,012686 | 0,991381 | Vitamin | 174,2568 |
| ebi-a-GCST90019423 | eGFR decline (General population) | Vitamin K-dependent protein C measurement | Inverse variance weighted | 4 | -0,00518 | 0,023261 | 0,823632 | Vitamin | 174,2568 |
| ebi-a-GCST90019423 | eGFR decline (General population) | Vitamin K-dependent protein C measurement | Simple mode | 4 | 0,006417 | 0,056599 | 0,916889 | Vitamin | 174,2568 |
| ebi-a-GCST90019423 | eGFR decline (General population) | Vitamin K-dependent protein C measurement | Weighted mode | 4 | 0,006417 | 0,012152 | 0,634016 | Vitamin | 174,2568 |
| ebi-a-GCST90025947 | eGFR decline (General population) | Serum alkaline phosphatase levels | MR Egger | 351 | -0,00973 | 0,01706 | 0,568694 | Minerals | 164,3204 |
| ebi-a-GCST90025947 | eGFR decline (General population) | Serum alkaline phosphatase levels | Weighted median | 351 | 0,000863 | 0,018751 | 0,9633 | Minerals | 164,3204 |
| ebi-a-GCST90025947 | eGFR decline (General population) | Serum alkaline phosphatase levels | Inverse variance weighted | 351 | -0,00316 | 0,010106 | 0,754336 | Minerals | 164,3204 |
| ebi-a-GCST90025947 | eGFR decline (General population) | Serum alkaline phosphatase levels | Simple mode | 351 | 0,03014 | 0,042634 | 0,480076 | Minerals | 164,3204 |
| ebi-a-GCST90025947 | eGFR decline (General population) | Serum alkaline phosphatase levels | Weighted mode | 351 | -0,00412 | 0,02118 | 0,846054 | Minerals | 164,3204 |
| ebi-a-GCST90025967 | eGFR decline (General population) | Vitamin D levels | MR Egger | 84 | 0,004653 | 0,033399 | 0,889546 | Vitamin | 131,2146 |
| ebi-a-GCST90025967 | eGFR decline (General population) | Vitamin D levels | Weighted median | 84 | 0,011667 | 0,030363 | 0,700793 | Vitamin | 131,2146 |
| ebi-a-GCST90025967 | eGFR decline (General population) | Vitamin D levels | Inverse variance weighted | 84 | 0,019894 | 0,020387 | 0,329154 | Vitamin | 131,2146 |
| ebi-a-GCST90025967 | eGFR decline (General population) | Vitamin D levels | Simple mode | 84 | 0,030716 | 0,06564 | 0,641055 | Vitamin | 131,2146 |
| ebi-a-GCST90025967 | eGFR decline (General population) | Vitamin D levels | Weighted mode | 84 | 0,010436 | 0,028913 | 0,71905 | Vitamin | 131,2146 |
| ebi-a-GCST90025969 | eGFR decline (General population) | Hemoglobin levels | MR Egger | 331 | -0,02683 | 0,025988 | 0,302565 | Organic compound | 105,1851 |
| ebi-a-GCST90025969 | eGFR decline (General population) | Hemoglobin levels | Weighted median | 331 | -0,01537 | 0,021274 | 0,470111 | Organic compound | 105,1851 |

| | | | | | | | | | |
|---|---|---|---|---|---|---|---|---|---|
| ebi-a-GCST90025969 | eGFR decline (General population) | Hemoglobin levels | Inverse variance weighted | 331 | -0,05031 | 0,013676 | 0,000234 | Organic compound | 105,1851 |
| ebi-a-GCST90025969 | eGFR decline (General population) | Hemoglobin levels | Simple mode | 331 | -0,03222 | 0,056398 | 0,568208 | Organic compound | 105,1851 |
| ebi-a-GCST90025969 | eGFR decline (General population) | Hemoglobin levels | Weighted mode | 331 | -0,01186 | 0,028586 | 0,678451 | Organic compound | 105,1851 |
| ebi-a-GCST90025986 | eGFR decline (General population) | Blood glucose levels | MR Egger | 113 | -0,00136 | 0,03193 | 0,966188 | Diabetes | 116,2197 |
| ebi-a-GCST90025986 | eGFR decline (General population) | Blood glucose levels | Weighted median | 113 | -0,03347 | 0,030143 | 0,266828 | Diabetes | 116,2197 |
| ebi-a-GCST90025986 | eGFR decline (General population) | Blood glucose levels | Inverse variance weighted | 113 | -0,00952 | 0,017815 | 0,592919 | Diabetes | 116,2197 |
| ebi-a-GCST90025986 | eGFR decline (General population) | Blood glucose levels | Simple mode | 113 | -0,02247 | 0,075035 | 0,765195 | Diabetes | 116,2197 |
| ebi-a-GCST90025986 | eGFR decline (General population) | Blood glucose levels | Weighted mode | 113 | -0,01921 | 0,032341 | 0,55373 | Diabetes | 116,2197 |
| ebi-a-GCST90025989 | eGFR decline (General population) | Insulin-like growth factor 1 levels | MR Egger | 384 | -0,0231 | 0,021432 | 0,281749 | Diabetes | 118,3851 |
| ebi-a-GCST90025989 | eGFR decline (General population) | Insulin-like growth factor 1 levels | Weighted median | 384 | -0,02114 | 0,018577 | 0,255065 | Diabetes | 118,3851 |
| ebi-a-GCST90025989 | eGFR decline (General population) | Insulin-like growth factor 1 levels | Inverse variance weighted | 384 | -0,02775 | 0,011327 | 0,014291 | Diabetes | 118,3851 |
| ebi-a-GCST90025989 | eGFR decline (General population) | Insulin-like growth factor 1 levels | Simple mode | 384 | -0,03959 | 0,04886 | 0,418289 | Diabetes | 118,3851 |
| ebi-a-GCST90025989 | eGFR decline (General population) | Insulin-like growth factor 1 levels | Weighted mode | 384 | -0,02425 | 0,027991 | 0,386937 | Diabetes | 118,3851 |
| ebi-a-GCST90025990 | eGFR decline (General population) | Calcium levels | MR Egger | 217 | 0,000112 | 0,028312 | 0,996849 | Minerals | 101,6909 |
| ebi-a-GCST90025990 | eGFR decline (General population) | Calcium levels | Weighted median | 217 | -0,00057 | 0,022744 | 0,980164 | Minerals | 101,6909 |
| ebi-a-GCST90025990 | eGFR decline (General population) | Calcium levels | Inverse variance weighted | 217 | -0,02018 | 0,015504 | 0,193109 | Minerals | 101,6909 |
| ebi-a-GCST90025990 | eGFR decline (General population) | Calcium levels | Simple mode | 217 | 0,002251 | 0,052507 | 0,965849 | Minerals | 101,6909 |
| ebi-a-GCST90025990 | eGFR decline (General population) | Calcium levels | Weighted mode | 217 | -0,0075 | 0,027846 | 0,787975 | Minerals | 101,6909 |
| ebi-a-GCST90029013 | eGFR decline (General population) | Educational attainment (years of education) | MR Egger | 211 | -0,01411 | 0,019422 | 0,468371 | Social trait | 44,93868 |
| ebi-a-GCST90029013 | eGFR decline (General population) | Educational attainment (years of education) | Weighted median | 211 | -0,01413 | 0,006635 | 0,033258 | Social trait | 44,93868 |
| ebi-a-GCST90029013 | eGFR decline (General population) | Educational attainment (years of education) | Inverse variance weighted | 211 | -0,01601 | 0,00451 | 0,000386 | Social trait | 44,93868 |
| ebi-a-GCST90029013 | eGFR decline (General population) | Educational attainment (years of education) | Simple mode | 211 | -0,01214 | 0,02019 | 0,548413 | Social trait | 44,93868 |
| ebi-a-GCST90029013 | eGFR decline (General population) | Educational attainment (years of education) | Weighted mode | 211 | -0,01342 | 0,019052 | 0,482002 | Social trait | 44,93868 |
| ebi-a-GCST90029014 | eGFR decline (General population) | Smoking status | MR Egger | 117 | 0,243353 | 0,19567 | 0,216144 | Smoking | 42,91926 |
| ebi-a-GCST90029014 | eGFR decline (General population) | Smoking status | Weighted median | 117 | 0,068459 | 0,071969 | 0,341488 | Smoking | 42,91926 |
| ebi-a-GCST90029014 | eGFR decline (General population) | Smoking status | Inverse variance weighted | 117 | 0,070669 | 0,046176 | 0,125908 | Smoking | 42,91926 |
| ebi-a-GCST90029014 | eGFR decline (General population) | Smoking status | Simple mode | 117 | 0,227624 | 0,187378 | 0,226914 | Smoking | 42,91926 |
| ebi-a-GCST90029014 | eGFR decline (General population) | Smoking status | Weighted mode | 117 | 0,14829 | 0,152633 | 0,333301 | Smoking | 42,91926 |
| ebi-a-GCST90029070 | eGFR decline (General population) | C-reactive protein levels | MR Egger | 231 | 0,010348 | 0,021962 | 0,637943 | Inflammation | 120,6737 |
| ebi-a-GCST90029070 | eGFR decline (General population) | C-reactive protein levels | Weighted median | 231 | 0,019266 | 0,021283 | 0,36534 | Inflammation | 120,6737 |
| ebi-a-GCST90029070 | eGFR decline (General population) | C-reactive protein levels | Inverse variance weighted | 231 | 0,019145 | 0,013444 | 0,154438 | Inflammation | 120,6737 |
| ebi-a-GCST90029070 | eGFR decline (General population) | C-reactive protein levels | Simple mode | 231 | 0,009419 | 0,042875 | 0,82631 | Inflammation | 120,6737 |
| ebi-a-GCST90029070 | eGFR decline (General population) | C-reactive protein levels | Weighted mode | 231 | 0,027924 | 0,020889 | 0,182621 | Inflammation | 120,6737 |
| ebi-a-GCST90029073 | eGFR decline (General population) | Liver fat | MR Egger | 9 | -0,00696 | 0,029414 | 0,819636 | Fatty liver | 131,2797 |
| ebi-a-GCST90029073 | eGFR decline (General population) | Liver fat | Weighted median | 9 | -0,01181 | 0,020306 | 0,560701 | Fatty liver | 131,2797 |
| ebi-a-GCST90029073 | eGFR decline (General population) | Liver fat | Inverse variance weighted | 9 | -0,00373 | 0,016613 | 0,822426 | Fatty liver | 131,2797 |
| ebi-a-GCST90029073 | eGFR decline (General population) | Liver fat | Simple mode | 9 | 0,022868 | 0,035476 | 0,53723 | Fatty liver | 131,2797 |
| ebi-a-GCST90029073 | eGFR decline (General population) | Liver fat | Weighted mode | 9 | -0,01004 | 0,019786 | 0,625381 | Fatty liver | 131,2797 |
| ebi-a-GCST90086057 | eGFR decline (General population) | Cardiovascular disease | Inverse variance weighted | 2 | 0,006591 | 0,061003 | 0,913956 | Cardiovascular disease | 32,018 |
| finn-b-D3_ANAEMIA_B12_DEF | eGFR decline (General population) | Vitamin B12 deficiency anaemia | MR Egger | 4 | -0,01777 | 0,027417 | 0,583443 | Vitamin | 52,61875 |
| finn-b-D3_ANAEMIA_B12_DEF | eGFR decline (General population) | Vitamin B12 deficiency anaemia | Weighted median | 4 | -0,01278 | 0,010859 | 0,239357 | Vitamin | 52,61875 |
| finn-b-D3_ANAEMIA_B12_DEF | eGFR decline (General population) | Vitamin B12 deficiency anaemia | Inverse variance weighted | 4 | -0,01281 | 0,009213 | 0,164368 | Vitamin | 52,61875 |
| finn-b-D3_ANAEMIA_B12_DEF | eGFR decline (General population) | Vitamin B12 deficiency anaemia | Simple mode | 4 | -0,01221 | 0,01393 | 0,445258 | Vitamin | 52,61875 |
| finn-b-D3_ANAEMIA_B12_DEF | eGFR decline (General population) | Vitamin B12 deficiency anaemia | Weighted mode | 4 | -0,01269 | 0,013169 | 0,406309 | Vitamin | 52,61875 |
| finn-b-E4_OBESITY | eGFR decline (General population) | Obesity | MR Egger | 8 | 0,080729 | 0,058409 | 0,216181 | Obesity | 55,97912 |
| finn-b-E4_OBESITY | eGFR decline (General population) | Obesity | Weighted median | 8 | 0,003048 | 0,01595 | 0,848441 | Obesity | 55,97912 |
| finn-b-E4_OBESITY | eGFR decline (General population) | Obesity | Inverse variance weighted | 8 | -0,00192 | 0,021155 | 0,927559 | Obesity | 55,97912 |
| finn-b-E4_OBESITY | eGFR decline (General population) | Obesity | Simple mode | 8 | 0,019308 | 0,024057 | 0,448589 | Obesity | 55,97912 |
| finn-b-E4_OBESITY | eGFR decline (General population) | Obesity | Weighted mode | 8 | 0,009417 | 0,017156 | 0,600155 | Obesity | 55,97912 |
| finn-b-FG_PULMHEART | eGFR decline (General population) | Pulmonary heart disease | MR Egger | 6 | -0,0266 | 0,029146 | 0,413 | Cardiovascular disease | 80,37648 |
| finn-b-FG_PULMHEART | eGFR decline (General population) | Pulmonary heart disease | Weighted median | 6 | -0,00795 | 0,011273 | 0,480407 | Cardiovascular disease | 80,37648 |
| finn-b-FG_PULMHEART | eGFR decline (General population) | Pulmonary heart disease | Inverse variance weighted | 6 | -0,01095 | 0,010188 | 0,282679 | Cardiovascular disease | 80,37648 |
| finn-b-FG_PULMHEART | eGFR decline (General population) | Pulmonary heart disease | Simple mode | 6 | 0,005689 | 0,018409 | 0,769745 | Cardiovascular disease | 80,37648 |
| finn-b-FG_PULMHEART | eGFR decline (General population) | Pulmonary heart disease | Weighted mode | 6 | -0,00516 | 0,015146 | 0,747152 | Cardiovascular disease | 80,37648 |
| ieu-b-102 | eGFR decline (General population) | Major depression | MR Egger | 48 | -0,07018 | 0,126628 | 0,582123 | Mental health | 38,29386 |
| ieu-b-102 | eGFR decline (General population) | Major depression | Weighted median | 48 | -0,05498 | 0,032448 | 0,090167 | Mental health | 38,29386 |
| ieu-b-102 | eGFR decline (General population) | Major depression | Inverse variance weighted | 48 | -0,04018 | 0,023664 | 0,089482 | Mental health | 38,29386 |
| ieu-b-102 | eGFR decline (General population) | Major depression | Simple mode | 48 | -0,10507 | 0,073711 | 0,160653 | Mental health | 38,29386 |
| ieu-b-102 | eGFR decline (General population) | Major depression | Weighted mode | 48 | -0,10507 | 0,071226 | 0,146848 | Mental health | 38,29386 |
| ieu-b-107 | eGFR decline (General population) | apolipoprotein A-I | MR Egger | 288 | -0,01748 | 0,019633 | 0,374157 | Dyslipidemias | 139,9829 |
| ieu-b-107 | eGFR decline (General population) | apolipoprotein A-I | Weighted median | 288 | -0,0085 | 0,01893 | 0,653313 | Dyslipidemias | 139,9829 |
| ieu-b-107 | eGFR decline (General population) | apolipoprotein A-I | Inverse variance weighted | 288 | -0,03794 | 0,011963 | 0,001516 | Dyslipidemias | 139,9829 |
| ieu-b-107 | eGFR decline (General population) | apolipoprotein A-I | Simple mode | 288 | -0,08369 | 0,043344 | 0,054494 | Dyslipidemias | 139,9829 |
| ieu-b-107 | eGFR decline (General population) | apolipoprotein A-I | Weighted mode | 288 | 0,001513 | 0,021125 | 0,942936 | Dyslipidemias | 139,9829 |
| ieu-b-108 | eGFR decline (General population) | apolipoprotein B | MR Egger | 189 | -0,00783 | 0,019061 | 0,681616 | Dyslipidemias | 168,3252 |
| ieu-b-108 | eGFR decline (General population) | apolipoprotein B | Weighted median | 189 | -0,01661 | 0,020224 | 0,41157 | Dyslipidemias | 168,3252 |
| ieu-b-108 | eGFR decline (General population) | apolipoprotein B | Inverse variance weighted | 189 | 0,000824 | 0,012294 | 0,946571 | Dyslipidemias | 168,3252 |
| ieu-b-108 | eGFR decline (General population) | apolipoprotein B | Simple mode | 189 | 0,005334 | 0,04301 | 0,90143 | Dyslipidemias | 168,3252 |
| ieu-b-108 | eGFR decline (General population) | apolipoprotein B | Weighted mode | 189 | -0,01408 | 0,022554 | 0,533317 | Dyslipidemias | 168,3252 |
| ieu-b-25 | eGFR decline (General population) | Cigarettes per Day | MR Egger | 22 | 0,044038 | 0,034613 | 0,217866 | Smoking | 99,80012 |
| ieu-b-25 | eGFR decline (General population) | Cigarettes per Day | Weighted median | 22 | 0,043955 | 0,025734 | 0,087624 | Smoking | 99,80012 |
| ieu-b-25 | eGFR decline (General population) | Cigarettes per Day | Inverse variance weighted | 22 | 0,014191 | 0,019837 | 0,474372 | Smoking | 99,80012 |
| ieu-b-25 | eGFR decline (General population) | Cigarettes per Day | Simple mode | 22 | 0,016619 | 0,061642 | 0,790102 | Smoking | 99,80012 |
| ieu-b-25 | eGFR decline (General population) | Cigarettes per Day | Weighted mode | 22 | 0,050634 | 0,026692 | 0,071665 | Smoking | 99,80012 |
| ieu-b-4861 | eGFR decline (General population) | Chronotype | MR Egger | 62 | -0,02517 | 0,073767 | 0,734157 | Sleep disorders | 44,20616 |
| ieu-b-4861 | eGFR decline (General population) | Chronotype | Weighted median | 62 | -0,00961 | 0,033393 | 0,773581 | Sleep disorders | 44,20616 |
| ieu-b-4861 | eGFR decline (General population) | Chronotype | Inverse variance weighted | 62 | 0,013113 | 0,027168 | 0,629328 | Sleep disorders | 44,20616 |
| ieu-b-4861 | eGFR decline (General population) | Chronotype | Simple mode | 62 | 0,00483 | 0,075057 | 0,948906 | Sleep disorders | 44,20616 |
| ieu-b-4861 | eGFR decline (General population) | Chronotype | Weighted mode | 62 | -0,00416 | 0,070526 | 0,953176 | Sleep disorders | 44,20616 |
| ieu-b-4877 | eGFR decline (General population) | smoking initiation | MR Egger | 85 | 0,104083 | 0,106256 | 0,330156 | Smoking | 42,2587 |
| ieu-b-4877 | eGFR decline (General population) | smoking initiation | Weighted median | 85 | 0,048514 | 0,030405 | 0,11058 | Smoking | 42,2587 |
| ieu-b-4877 | eGFR decline (General population) | smoking initiation | Inverse variance weighted | 85 | 0,028518 | 0,020844 | 0,171273 | Smoking | 42,2587 |
| ieu-b-4877 | eGFR decline (General population) | smoking initiation | Simple mode | 85 | 0,100414 | 0,074191 | 0,17954 | Smoking | 42,2587 |
| ieu-b-4877 | eGFR decline (General population) | smoking initiation | Weighted mode | 85 | 0,078516 | 0,063341 | 0,21858 | Smoking | 42,2587 |
| ieu-b-5144 | eGFR decline (General population) | Hypertension | MR Egger | 203 | -0,04031 | 0,21744 | 0,853122 | Hypertension | 53,29304 |
| ieu-b-5144 | eGFR decline (General population) | Hypertension | Weighted median | 203 | 0,069435 | 0,078692 | 0,37758 | Hypertension | 53,29304 |
| ieu-b-5144 | eGFR decline (General population) | Hypertension | Inverse variance weighted | 203 | 0,165209 | 0,071261 | 0,020429 | Hypertension | 53,29304 |
| ieu-b-5144 | eGFR decline (General population) | Hypertension | Simple mode | 203 | 0,208377 | 0,23945 | 0,385208 | Hypertension | 53,29304 |
| ieu-b-5144 | eGFR decline (General population) | Hypertension | Weighted mode | 203 | -0,0552 | 0,212767 | 0,795573 | Hypertension | 53,29304 |
| ieu-b-72 | eGFR decline (General population) | Urinary sodium-potassium ratio | MR Egger | 24 | -0,12307 | 0,278713 | 0,663117 | Minerals | 39,88328 |
| ieu-b-72 | eGFR decline (General population) | Urinary sodium-potassium ratio | Weighted median | 24 | 0,054697 | 0,082514 | 0,507408 | Minerals | 39,88328 |
| ieu-b-72 | eGFR decline (General population) | Urinary sodium-potassium ratio | Inverse variance weighted | 24 | 0,061012 | 0,059325 | 0,303746 | Minerals | 39,88328 |
| ieu-b-72 | eGFR decline (General population) | Urinary sodium-potassium ratio | Simple mode | 24 | 0,027418 | 0,129272 | 0,833898 | Minerals | 39,88328 |
| ieu-b-72 | eGFR decline (General population) | Urinary sodium-potassium ratio | Weighted mode | 24 | 0,123983 | 0,140509 | 0,3867 | Minerals | 39,88328 |
| prot-a-3102 | eGFR decline (General population) | Thyrotropin-releasing hormone | Inverse variance weighted | 2 | -0,01469 | 0,026598 | 0,580761 | Thyroid disease | 36,35446 |
| prot-c-2652_15_1 | eGFR decline (General population) | suPAR | Wald ratio | 1 | -0,00986 | 0,018798 | 0,599868 | Biomarkers | 33,5125 |
| ukb-b-4424 | eGFR decline (General population) | Sleep duration | MR Egger | 67 | 0,077291 | 0,200027 | 0,70046 | Sleep disorders | 42,62064 |
| ukb-b-4424 | eGFR decline (General population) | Sleep duration | Weighted median | 67 | -0,01078 | 0,0739 | 0,884049 | Sleep disorders | 42,62064 |
| ukb-b-4424 | eGFR decline (General population) | Sleep duration | Inverse variance weighted | 67 | -0,0232 | 0,051804 | 0,654335 | Sleep disorders | 42,62064 |
| ukb-b-4424 | eGFR decline (General population) | Sleep duration | Simple mode | 67 | -0,01713 | 0,166924 | 0,918596 | Sleep disorders | 42,62064 |
| ukb-b-4424 | eGFR decline (General population) | Sleep duration | Weighted mode | 67 | 0,003393 | 0,141733 | 0,980975 | Sleep disorders | 42,62064 |
| ukb-b-5779 | eGFR decline (General population) | Alcohol intake frequency. | MR Egger | 95 | 0,027074 | 0,055452 | 0,626525 | Alcohol consumption | 52,90557 |
| ukb-b-5779 | eGFR decline (General population) | Alcohol intake frequency. | Weighted median | 95 | 0,00715 | 0,03173 | 0,821725 | Alcohol consumption | 52,90557 |
| ukb-b-5779 | eGFR decline (General population) | Alcohol intake frequency. | Inverse variance weighted | 95 | 0,021885 | 0,021727 | 0,313803 | Alcohol consumption | 52,90557 |
| ukb-b-5779 | eGFR decline (General population) | Alcohol intake frequency. | Simple mode | 95 | 0,02733 | 0,075064 | 0,716611 | Alcohol consumption | 52,90557 |
| ukb-b-5779 | eGFR decline (General population) | Alcohol intake frequency. | Weighted mode | 95 | -0,00078 | 0,048737 | 0,987199 | Alcohol consumption | 52,90557 |
| ukb-b-7864 | eGFR decline (General population) | Vitamin B6 | Wald ratio | 1 | 0,046711 | 0,145971 | 0,748968 | Vitamin | 32,75897 |
| ukb-d-30790_irnt | eGFR decline (General population) | Lipoprotein A | MR Egger | 20 | 0,020137 | 0,019884 | 0,324613 | Dyslipidemias | 950,9105 |
| ukb-d-30790_irnt | eGFR decline (General population) | Lipoprotein A | Weighted median | 20 | 0,015928 | 0,018193 | 0,381284 | Dyslipidemias | 950,9105 |
| ukb-d-30790_irnt | eGFR decline (General population) | Lipoprotein A | Inverse variance weighted | 20 | 0,00788 | 0,012605 | 0,531851 | Dyslipidemias | 950,9105 |
| ukb-d-30790_irnt | eGFR decline (General population) | Lipoprotein A | Simple mode | 20 | 0,022451 | 0,026156 | 0,401395 | Dyslipidemias | 950,9105 |
| ukb-d-30790_irnt | eGFR decline (General population) | Lipoprotein A | Weighted mode | 20 | 0,020393 | 0,023291 | 0,392203 | Dyslipidemias | 950,9105 |
| BMI | eGFR decline (CKD population) | Body mass index | MR Egger | 504 | 0,142106 | 0,200017 | 0,477743 | Obesity | 74,21321 |

| | | | | | | | | | |
|---|---|---|---|---|---|---|---|---|---|
| BMI | eGFR decline (CKD population) | Body mass index | Weighted median | 504 | 0,183581 | 0,137889 | 0,183067 | Obesity | 74,21321 |
| BMI | eGFR decline (CKD population) | Body mass index | Inverse variance weighted | 504 | 0,154688 | 0,076931 | 0,044355 | Obesity | 74,21321 |
| BMI | eGFR decline (CKD population) | Body mass index | Simple mode | 504 | 0,430975 | 0,409857 | 0,293523 | Obesity | 74,21321 |
| BMI | eGFR decline (CKD population) | Body mass index | Weighted mode | 504 | 0,369672 | 0,253049 | 0,144675 | Obesity | 74,21321 |
| DBPTang | eGFR decline (CKD population) | Diastolic blood pressure | MR Egger | 256 | -0,05325 | 0,34333 | 0,876856 | Hypertension | 55,60753 |
| DBPTang | eGFR decline (CKD population) | Diastolic blood pressure | Weighted median | 256 | -0,03285 | 0,158637 | 0,835972 | Hypertension | 55,60753 |
| DBPTang | eGFR decline (CKD population) | Diastolic blood pressure | Inverse variance weighted | 256 | 0,053932 | 0,106509 | 0,612606 | Hypertension | 55,60753 |
| DBPTang | eGFR decline (CKD population) | Diastolic blood pressure | Simple mode | 256 | 0,181665 | 0,454456 | 0,689681 | Hypertension | 55,60753 |
| DBPTang | eGFR decline (CKD population) | Diastolic blood pressure | Weighted mode | 256 | -0,14069 | 0,333415 | 0,673408 | Hypertension | 55,60753 |
| FT3 | eGFR decline (CKD population) | Free triiodothyronine | MR Egger | 6 | -0,68526 | 0,866911 | 0,473486 | Thyroid disease | 82,22035 |
| FT3 | eGFR decline (CKD population) | Free triiodothyronine | Weighted median | 6 | -0,27931 | 0,232715 | 0,230044 | Thyroid disease | 82,22035 |
| FT3 | eGFR decline (CKD population) | Free triiodothyronine | Inverse variance weighted | 6 | -0,25194 | 0,190959 | 0,187051 | Thyroid disease | 82,22035 |
| FT3 | eGFR decline (CKD population) | Free triiodothyronine | Simple mode | 6 | -0,19122 | 0,328614 | 0,585887 | Thyroid disease | 82,22035 |
| FT3 | eGFR decline (CKD population) | Free triiodothyronine | Weighted mode | 6 | -0,26207 | 0,300928 | 0,423661 | Thyroid disease | 82,22035 |
| FT3FT4 | eGFR decline (CKD population) | FT3 FT4 ratio | MR Egger | 13 | -0,00057 | 1,091739 | 0,999592 | Thyroid disease | 132,3008 |
| FT3FT4 | eGFR decline (CKD population) | FT3 FT4 ratio | Weighted median | 13 | -0,47243 | 0,724714 | 0,51448 | Thyroid disease | 132,3008 |
| FT3FT4 | eGFR decline (CKD population) | FT3 FT4 ratio | Inverse variance weighted | 13 | -0,96421 | 0,543215 | 0,075897 | Thyroid disease | 132,3008 |
| FT3FT4 | eGFR decline (CKD population) | FT3 FT4 ratio | Simple mode | 13 | -0,20019 | 1,32015 | 0,881987 | Thyroid disease | 132,3008 |
| FT3FT4 | eGFR decline (CKD population) | FT3 FT4 ratio | Weighted mode | 13 | -0,59836 | 0,810418 | 0,474503 | Thyroid disease | 132,3008 |
| FT4 | eGFR decline (CKD population) | Free thyroxine | MR Egger | 62 | -0,00394 | 0,170287 | 0,981622 | Thyroid disease | 84,3797 |
| FT4 | eGFR decline (CKD population) | Free thyroxine | Weighted median | 62 | 0,082117 | 0,131285 | 0,53165 | Thyroid disease | 84,3797 |
| FT4 | eGFR decline (CKD population) | Free thyroxine | Inverse variance weighted | 62 | -0,03687 | 0,081356 | 0,650431 | Thyroid disease | 84,3797 |
| FT4 | eGFR decline (CKD population) | Free thyroxine | Simple mode | 62 | -0,09876 | 0,263799 | 0,709421 | Thyroid disease | 84,3797 |
| FT4 | eGFR decline (CKD population) | Free thyroxine | Weighted mode | 62 | 0,017818 | 0,148996 | 0,905202 | Thyroid disease | 84,3797 |
| Fe | eGFR decline (CKD population) | Serum iron | Inverse variance weighted | 2 | -0,30365 | 0,606072 | 0,616367 | Minerals | 291,9629 |
| HDL | eGFR decline (CKD population) | HDL cholesterol | MR Egger | 234 | 0,0332 | 0,101892 | 0,744841 | Dyslipidemias | 270,2118 |
| HDL | eGFR decline (CKD population) | HDL cholesterol | Weighted median | 234 | 0,040149 | 0,108182 | 0,710542 | Dyslipidemias | 270,2118 |
| HDL | eGFR decline (CKD population) | HDL cholesterol | Inverse variance weighted | 234 | -0,12372 | 0,066179 | 0,061562 | Dyslipidemias | 270,2118 |
| HDL | eGFR decline (CKD population) | HDL cholesterol | Simple mode | 234 | -0,03028 | 0,205212 | 0,882807 | Dyslipidemias | 270,2118 |
| HDL | eGFR decline (CKD population) | HDL cholesterol | Weighted mode | 234 | -0,00436 | 0,099597 | 0,965109 | Dyslipidemias | 270,2118 |
| HiTSH | eGFR decline (CKD population) | High TSH | MR Egger | 6 | 0,486707 | 0,515095 | 0,398218 | Thyroid disease | 49,53497 |
| HiTSH | eGFR decline (CKD population) | High TSH | Weighted median | 6 | 0,318734 | 0,154009 | 0,038491 | Thyroid disease | 49,53497 |
| HiTSH | eGFR decline (CKD population) | High TSH | Inverse variance weighted | 6 | 0,352939 | 0,125156 | 0,004802 | Thyroid disease | 49,53497 |
| HiTSH | eGFR decline (CKD population) | High TSH | Simple mode | 6 | 0,303913 | 0,226252 | 0,236934 | Thyroid disease | 49,53497 |
| HiTSH | eGFR decline (CKD population) | High TSH | Weighted mode | 6 | 0,321681 | 0,189322 | 0,150047 | Thyroid disease | 49,53497 |
| Insom | eGFR decline (CKD population) | Insomnia | MR Egger | 13 | 0,147319 | 0,687165 | 0,834169 | Sleep disorders | 44,05724 |
| Insom | eGFR decline (CKD population) | Insomnia | Weighted median | 13 | -0,07771 | 0,280972 | 0,782099 | Sleep disorders | 44,05724 |
| Insom | eGFR decline (CKD population) | Insomnia | Inverse variance weighted | 13 | 0,124966 | 0,21892 | 0,568117 | Sleep disorders | 44,05724 |
| Insom | eGFR decline (CKD population) | Insomnia | Simple mode | 13 | -0,15681 | 0,420559 | 0,715752 | Sleep disorders | 44,05724 |
| Insom | eGFR decline (CKD population) | Insomnia | Weighted mode | 13 | -0,10863 | 0,417261 | 0,799016 | Sleep disorders | 44,05724 |
| LDL | eGFR decline (CKD population) | LDL cholesterol | MR Egger | 218 | -0,05623 | 0,112804 | 0,618661 | Dyslipidemias | 221,0222 |
| LDL | eGFR decline (CKD population) | LDL cholesterol | Weighted median | 218 | -0,09623 | 0,108612 | 0,375614 | Dyslipidemias | 221,0222 |
| LDL | eGFR decline (CKD population) | LDL cholesterol | Inverse variance weighted | 218 | -0,08321 | 0,070454 | 0,23759 | Dyslipidemias | 221,0222 |
| LDL | eGFR decline (CKD population) | LDL cholesterol | Simple mode | 218 | -0,16776 | 0,22454 | 0,455797 | Dyslipidemias | 221,0222 |
| LDL | eGFR decline (CKD population) | LDL cholesterol | Weighted mode | 218 | -0,11462 | 0,124411 | 0,357916 | Dyslipidemias | 221,0222 |
| LoTSH | eGFR decline (CKD population) | Low TSH | MR Egger | 26 | -0,04357 | 0,078626 | 0,58458 | Thyroid disease | 70,97091 |
| LoTSH | eGFR decline (CKD population) | Low TSH | Weighted median | 26 | 0,03229 | 0,041913 | 0,441069 | Thyroid disease | 70,97091 |
| LoTSH | eGFR decline (CKD population) | Low TSH | Inverse variance weighted | 26 | 0,009852 | 0,027141 | 0,716609 | Thyroid disease | 70,97091 |
| LoTSH | eGFR decline (CKD population) | Low TSH | Simple mode | 26 | 0,06839 | 0,077584 | 0,386443 | Thyroid disease | 70,97091 |
| LoTSH | eGFR decline (CKD population) | Low TSH | Weighted mode | 26 | 0,060078 | 0,058836 | 0,316985 | Thyroid disease | 70,97091 |
| PPTang | eGFR decline (CKD population) | Pulse pressure | MR Egger | 302 | 0,323901 | 0,247494 | 0,191631 | Hypertension | 68,42683 |
| PPTang | eGFR decline (CKD population) | Pulse pressure | Weighted median | 302 | 0,214614 | 0,148324 | 0,147918 | Hypertension | 68,42683 |
| PPTang | eGFR decline (CKD population) | Pulse pressure | Inverse variance weighted | 302 | 0,221405 | 0,09214 | 0,016265 | Hypertension | 68,42683 |
| PPTang | eGFR decline (CKD population) | Pulse pressure | Simple mode | 302 | -0,08906 | 0,415197 | 0,8303 | Hypertension | 68,42683 |
| PPTang | eGFR decline (CKD population) | Pulse pressure | Weighted mode | 302 | 0,232096 | 0,281009 | 0,409495 | Hypertension | 68,42683 |
| SBPTang | eGFR decline (CKD population) | Systolic blood pressure | MR Egger | 254 | -0,21636 | 0,328419 | 0,51063 | Hypertension | 56,77375 |
| SBPTang | eGFR decline (CKD population) | Systolic blood pressure | Weighted median | 254 | 0,002894 | 0,154025 | 0,985008 | Hypertension | 56,77375 |
| SBPTang | eGFR decline (CKD population) | Systolic blood pressure | Inverse variance weighted | 254 | 0,100379 | 0,105815 | 0,342811 | Hypertension | 56,77375 |
| SBPTang | eGFR decline (CKD population) | Systolic blood pressure | Simple mode | 254 | 0,165614 | 0,446376 | 0,710935 | Hypertension | 56,77375 |
| SBPTang | eGFR decline (CKD population) | Systolic blood pressure | Weighted mode | 254 | -0,00924 | 0,324393 | 0,977288 | Hypertension | 56,77375 |
| SIndex | eGFR decline (CKD population) | Compound smoking index | MR Egger | 121 | 0,332774 | 0,994456 | 0,738494 | Smoking | 44,08236 |
| SIndex | eGFR decline (CKD population) | Compound smoking index | Weighted median | 121 | -0,34268 | 0,376425 | 0,362632 | Smoking | 44,08236 |
| SIndex | eGFR decline (CKD population) | Compound smoking index | Inverse variance weighted | 121 | -0,33433 | 0,247706 | 0,17711 | Smoking | 44,08236 |
| SIndex | eGFR decline (CKD population) | Compound smoking index | Simple mode | 121 | -0,53749 | 0,996831 | 0,590749 | Smoking | 44,08236 |
| SIndex | eGFR decline (CKD population) | Compound smoking index | Weighted mode | 121 | -0,62315 | 0,856023 | 0,468055 | Smoking | 44,08236 |
| T2DGGI | eGFR decline (CKD population) | Type 2 diabetes | MR Egger | 441 | 0,133837 | 0,070124 | 0,056967 | Diabetes | 93,04757 |
| T2DGGI | eGFR decline (CKD population) | Type 2 diabetes | Weighted median | 441 | 0,124112 | 0,069617 | 0,074621 | Diabetes | 93,04757 |
| T2DGGI | eGFR decline (CKD population) | Type 2 diabetes | Inverse variance weighted | 441 | 0,101682 | 0,033823 | 0,002644 | Diabetes | 93,04757 |
| T2DGGI | eGFR decline (CKD population) | Type 2 diabetes | Simple mode | 441 | 0,271971 | 0,142184 | 0,056422 | Diabetes | 93,04757 |
| T2DGGI | eGFR decline (CKD population) | Type 2 diabetes | Weighted mode | 441 | 0,152818 | 0,075906 | 0,044696 | Diabetes | 93,04757 |
| TC | eGFR decline (CKD population) | Total cholesterol | MR Egger | 264 | -0,05998 | 0,101087 | 0,553449 | Dyslipidemias | 246,1189 |
| TC | eGFR decline (CKD population) | Total cholesterol | Weighted median | 264 | -0,01305 | 0,106651 | 0,902619 | Dyslipidemias | 246,1189 |
| TC | eGFR decline (CKD population) | Total cholesterol | Inverse variance weighted | 264 | -0,03371 | 0,064304 | 0,600155 | Dyslipidemias | 246,1189 |
| TC | eGFR decline (CKD population) | Total cholesterol | Simple mode | 264 | -0,13018 | 0,212148 | 0,539991 | Dyslipidemias | 246,1189 |
| TC | eGFR decline (CKD population) | Total cholesterol | Weighted mode | 264 | -0,02001 | 0,112355 | 0,858818 | Dyslipidemias | 246,1189 |
| TG | eGFR decline (CKD population) | Triglyceride | MR Egger | 220 | 0,089701 | 0,105154 | 0,394573 | Dyslipidemias | 231,1265 |
| TG | eGFR decline (CKD population) | Triglyceride | Weighted median | 220 | 0,091252 | 0,112355 | 0,41669 | Dyslipidemias | 231,1265 |
| TG | eGFR decline (CKD population) | Triglyceride | Inverse variance weighted | 220 | 0,069304 | 0,067873 | 0,30721 | Dyslipidemias | 231,1265 |
| TG | eGFR decline (CKD population) | Triglyceride | Simple mode | 220 | -0,03129 | 0,225787 | 0,88992 | Dyslipidemias | 231,1265 |
| TG | eGFR decline (CKD population) | Triglyceride | Weighted mode | 220 | 0,06487 | 0,109718 | 0,554967 | Dyslipidemias | 231,1265 |
| TSH | eGFR decline (CKD population) | Thyroid Stimulating Hormone | MR Egger | 159 | 0,189704 | 0,097719 | 0,054011 | Thyroid disease | 154,4324 |
| TSH | eGFR decline (CKD population) | Thyroid Stimulating Hormone | Weighted median | 159 | 0,14131 | 0,085936 | 0,1001 | Thyroid disease | 154,4324 |
| TSH | eGFR decline (CKD population) | Thyroid Stimulating Hormone | Inverse variance weighted | 159 | 0,068453 | 0,054213 | 0,206706 | Thyroid disease | 154,4324 |
| TSH | eGFR decline (CKD population) | Thyroid Stimulating Hormone | Simple mode | 159 | -0,04252 | 0,182605 | 0,816173 | Thyroid disease | 154,4324 |
| TSH | eGFR decline (CKD population) | Thyroid Stimulating Hormone | Weighted mode | 159 | 0,176019 | 0,100571 | 0,082024 | Thyroid disease | 154,4324 |
| WHR | eGFR decline (CKD population) | Waist-to-hip ratio adjusted BMI | MR Egger | 296 | -0,15489 | 0,214147 | 0,470069 | Obesity | 85,86852 |
| WHR | eGFR decline (CKD population) | Waist-to-hip ratio adjusted BMI | Weighted median | 296 | -0,0531 | 0,1558 | 0,733228 | Obesity | 85,86852 |
| WHR | eGFR decline (CKD population) | Waist-to-hip ratio adjusted BMI | Inverse variance weighted | 296 | 0,029089 | 0,089529 | 0,745247 | Obesity | 85,86852 |
| WHR | eGFR decline (CKD population) | Waist-to-hip ratio adjusted BMI | Simple mode | 296 | -0,01178 | 0,393818 | 0,976148 | Obesity | 85,86852 |
| WHR | eGFR decline (CKD population) | Waist-to-hip ratio adjusted BMI | Weighted mode | 296 | 0,043332 | 0,251226 | 0,863175 | Obesity | 85,86852 |
| Zn | eGFR decline (CKD population) | Zinc | MR Egger | 3 | -0,1895 | 0,290457 | 0,631984 | Minerals | 99,95282 |
| Zn | eGFR decline (CKD population) | Zinc | Weighted median | 3 | 0,03756 | 0,087564 | 0,667962 | Minerals | 99,95282 |
| Zn | eGFR decline (CKD population) | Zinc | Inverse variance weighted | 3 | 0,084148 | 0,106455 | 0,429262 | Minerals | 99,95282 |
| Zn | eGFR decline (CKD population) | Zinc | Simple mode | 3 | 0,022658 | 0,112009 | 0,858404 | Minerals | 99,95282 |
| Zn | eGFR decline (CKD population) | Zinc | Weighted mode | 3 | 0,034374 | 0,09393 | 0,749481 | Minerals | 99,95282 |
| ebi-a-GCST006979-consort | eGFR decline (CKD population) | Heel bone mineral density | MR Egger | 483 | -0,00819 | 0,098437 | 0,933697 | Bone disease | 151,4375 |
| ebi-a-GCST006979-consort | eGFR decline (CKD population) | Heel bone mineral density | Weighted median | 483 | 0,131793 | 0,080496 | 0,101576 | Bone disease | 151,4375 |
| ebi-a-GCST006979-consort | eGFR decline (CKD population) | Heel bone mineral density | Inverse variance weighted | 483 | 0,091729 | 0,050833 | 0,071149 | Bone disease | 151,4375 |
| ebi-a-GCST006979-consort | eGFR decline (CKD population) | Heel bone mineral density | Simple mode | 483 | 0,292804 | 0,203897 | 0,15164 | Bone disease | 151,4375 |
| ebi-a-GCST006979-consort | eGFR decline (CKD population) | Heel bone mineral density | Weighted mode | 483 | 0,134444 | 0,128942 | 0,297625 | Bone disease | 151,4375 |
| ebi-a-GCST90002227 | eGFR decline (CKD population) | Two-hour glucose | MR Egger | 14 | -0,09628 | 0,312215 | 0,763089 | Diabetes | 56,47851 |
| ebi-a-GCST90002227 | eGFR decline (CKD population) | Two-hour glucose | Weighted median | 14 | 0,29134 | 0,153948 | 0,05843 | Diabetes | 56,47851 |
| ebi-a-GCST90002227 | eGFR decline (CKD population) | Two-hour glucose | Inverse variance weighted | 14 | 0,146871 | 0,113614 | 0,196109 | Diabetes | 56,47851 |
| ebi-a-GCST90002227 | eGFR decline (CKD population) | Two-hour glucose | Simple mode | 14 | 0,339715 | 0,271612 | 0,233075 | Diabetes | 56,47851 |
| ebi-a-GCST90002227 | eGFR decline (CKD population) | Two-hour glucose | Weighted mode | 14 | 0,350868 | 0,241229 | 0,169525 | Diabetes | 56,47851 |
| ebi-a-GCST90002232 | eGFR decline (CKD population) | Fasting glucose | MR Egger | 68 | 0,632936 | 0,324503 | 0,055368 | Diabetes | 124,2211 |
| ebi-a-GCST90002232 | eGFR decline (CKD population) | Fasting glucose | Weighted median | 68 | 0,722952 | 0,255898 | 0,004726 | Diabetes | 124,2211 |
| ebi-a-GCST90002232 | eGFR decline (CKD population) | Fasting glucose | Inverse variance weighted | 68 | 0,235297 | 0,171721 | 0,170615 | Diabetes | 124,2211 |
| ebi-a-GCST90002232 | eGFR decline (CKD population) | Fasting glucose | Simple mode | 68 | 0,607866 | 0,460985 | 0,191784 | Diabetes | 124,2211 |
| ebi-a-GCST90002232 | eGFR decline (CKD population) | Fasting glucose | Weighted mode | 68 | 0,667156 | 0,251066 | 0,009839 | Diabetes | 124,2211 |
| ebi-a-GCST90002238 | eGFR decline (CKD population) | Fasting insulin | MR Egger | 38 | 0,238289 | 1,133657 | 0,834701 | Diabetes | 51,80185 |
| ebi-a-GCST90002238 | eGFR decline (CKD population) | Fasting insulin | Weighted median | 38 | 0,347606 | 0,426675 | 0,415253 | Diabetes | 51,80185 |
| ebi-a-GCST90002238 | eGFR decline (CKD population) | Fasting insulin | Inverse variance weighted | 38 | 0,132902 | 0,361123 | 0,712854 | Diabetes | 51,80185 |

| | | | | | | | | | |
|---|---|---|---|---|---|---|---|---|---|
| ebi-a-GCST90002238 | eGFR decline (CKD population) | Fasting insulin | Simple mode | 38 | 0,413171 | 0,768524 | 0,594061 | Diabetes | 51,80185 |
| ebi-a-GCST90002238 | eGFR decline (CKD population) | Fasting insulin | Weighted mode | 38 | 0,379219 | 0,639349 | 0,5567 | Diabetes | 51,80185 |
| ebi-a-GCST90007310 | eGFR decline (CKD population) | circulating leptin levels | MR Egger | 3 | 3,205083 | 2,27809 | 0,39338 | Obesity | 56,18818 |
| ebi-a-GCST90007310 | eGFR decline (CKD population) | circulating leptin levels | Weighted median | 3 | -0,26374 | 0,331959 | 0,426913 | Obesity | 56,18818 |
| ebi-a-GCST90007310 | eGFR decline (CKD population) | circulating leptin levels | Inverse variance weighted | 3 | -0,39028 | 0,343599 | 0,25602 | Obesity | 56,18818 |
| ebi-a-GCST90007310 | eGFR decline (CKD population) | circulating leptin levels | Simple mode | 3 | -0,22368 | 0,466673 | 0,679016 | Obesity | 56,18818 |
| ebi-a-GCST90007310 | eGFR decline (CKD population) | circulating leptin levels | Weighted mode | 3 | -0,17572 | 0,399212 | 0,702816 | Obesity | 56,18818 |
| ebi-a-GCST90011998 | eGFR decline (CKD population) | Growth differentiation factor-15 levels | MR Egger | 3 | -0,22572 | 0,135176 | 0,34351 | Biomarkers | 294,0397 |
| ebi-a-GCST90011998 | eGFR decline (CKD population) | Growth differentiation factor-15 levels | Weighted median | 3 | -0,15286 | 0,094967 | 0,107471 | Biomarkers | 294,0397 |
| ebi-a-GCST90011998 | eGFR decline (CKD population) | Growth differentiation factor-15 levels | Inverse variance weighted | 3 | -0,14848 | 0,092598 | 0,108819 | Biomarkers | 294,0397 |
| ebi-a-GCST90011998 | eGFR decline (CKD population) | Growth differentiation factor-15 levels | Simple mode | 3 | 0,06095 | 0,20227 | 0,791605 | Biomarkers | 294,0397 |
| ebi-a-GCST90011998 | eGFR decline (CKD population) | Growth differentiation factor-15 levels | Weighted mode | 3 | -0,1705 | 0,098532 | 0,225701 | Biomarkers | 294,0397 |
| ebi-a-GCST90012022 | eGFR decline (CKD population) | Fibroblast growth factor 23 levels | MR Egger | 8 | 0,442206 | 0,455365 | 0,369001 | Biomarkers | 43,42098 |
| ebi-a-GCST90012022 | eGFR decline (CKD population) | Fibroblast growth factor 23 levels | Weighted median | 8 | 0,083296 | 0,16543 | 0,614604 | Biomarkers | 43,42098 |
| ebi-a-GCST90012022 | eGFR decline (CKD population) | Fibroblast growth factor 23 levels | Inverse variance weighted | 8 | 0,013106 | 0,121968 | 0,91443 | Biomarkers | 43,42098 |
| ebi-a-GCST90012022 | eGFR decline (CKD population) | Fibroblast growth factor 23 levels | Simple mode | 8 | 0,142976 | 0,266284 | 0,60795 | Biomarkers | 43,42098 |
| ebi-a-GCST90012022 | eGFR decline (CKD population) | Fibroblast growth factor 23 levels | Weighted mode | 8 | 0,195612 | 0,247738 | 0,455674 | Biomarkers | 43,42098 |
| ebi-a-GCST90012041 | eGFR decline (CKD population) | Kidney injury molecule 1 levels | MR Egger | 9 | -0,09729 | 0,189965 | 0,624322 | Biomarkers | 190,9807 |
| ebi-a-GCST90012041 | eGFR decline (CKD population) | Kidney injury molecule 1 levels | Weighted median | 9 | -0,06749 | 0,06302 | 0,28419 | Biomarkers | 190,9807 |
| ebi-a-GCST90012041 | eGFR decline (CKD population) | Kidney injury molecule 1 levels | Inverse variance weighted | 9 | -0,00012 | 0,115729 | 0,999187 | Biomarkers | 190,9807 |
| ebi-a-GCST90012041 | eGFR decline (CKD population) | Kidney injury molecule 1 levels | Simple mode | 9 | -0,11907 | 0,139255 | 0,417384 | Biomarkers | 190,9807 |
| ebi-a-GCST90012041 | eGFR decline (CKD population) | Kidney injury molecule 1 levels | Weighted mode | 9 | -0,07376 | 0,059823 | 0,252598 | Biomarkers | 190,9807 |
| ebi-a-GCST90013864 | eGFR decline (CKD population) | Coronary artery disease (Firth correction) | MR Egger | 38 | -0,05509 | 0,132584 | 0,680223 | Cardiovascular disease | 60,15128 |
| ebi-a-GCST90013864 | eGFR decline (CKD population) | Coronary artery disease (Firth correction) | Weighted median | 38 | -0,02084 | 0,086518 | 0,809671 | Cardiovascular disease | 60,15128 |
| ebi-a-GCST90013864 | eGFR decline (CKD population) | Coronary artery disease (Firth correction) | Inverse variance weighted | 38 | -0,01164 | 0,059099 | 0,843905 | Cardiovascular disease | 60,15128 |
| ebi-a-GCST90013864 | eGFR decline (CKD population) | Coronary artery disease (Firth correction) | Simple mode | 38 | -0,06476 | 0,155564 | 0,679592 | Cardiovascular disease | 60,15128 |
| ebi-a-GCST90013864 | eGFR decline (CKD population) | Coronary artery disease (Firth correction) | Weighted mode | 38 | -0,07425 | 0,117131 | 0,530048 | Cardiovascular disease | 60,15128 |
| ebi-a-GCST90013975 | eGFR decline (CKD population) | Body fat percentage | MR Egger | 321 | 0,335649 | 0,392405 | 0,392992 | Obesity | 56,13441 |
| ebi-a-GCST90013975 | eGFR decline (CKD population) | Body fat percentage | Weighted median | 321 | 0 | 0,186371 | 1 | Obesity | 56,13441 |
| ebi-a-GCST90013975 | eGFR decline (CKD population) | Body fat percentage | Inverse variance weighted | 321 | 0,136886 | 0,121862 | 0,261316 | Obesity | 56,13441 |
| ebi-a-GCST90013975 | eGFR decline (CKD population) | Body fat percentage | Simple mode | 321 | 0,318523 | 0,585249 | 0,586647 | Obesity | 56,13441 |
| ebi-a-GCST90013975 | eGFR decline (CKD population) | Body fat percentage | Weighted mode | 321 | -0,18924 | 0,431454 | 0,661242 | Obesity | 56,13441 |
| ebi-a-GCST90013989 | eGFR decline (CKD population) | Sodium in urine | MR Egger | 41 | 1,421704 | 1,074606 | 0,19354 | Minerals | 43,81211 |
| ebi-a-GCST90013989 | eGFR decline (CKD population) | Sodium in urine | Weighted median | 41 | -0,28094 | 0,394632 | 0,476531 | Minerals | 43,81211 |
| ebi-a-GCST90013989 | eGFR decline (CKD population) | Sodium in urine | Inverse variance weighted | 41 | -0,1629 | 0,276249 | 0,555401 | Minerals | 43,81211 |
| ebi-a-GCST90013989 | eGFR decline (CKD population) | Sodium in urine | Simple mode | 41 | 0,058941 | 0,849996 | 0,945062 | Minerals | 43,81211 |
| ebi-a-GCST90013989 | eGFR decline (CKD population) | Sodium in urine | Weighted mode | 41 | -0,54762 | 0,764276 | 0,47783 | Minerals | 43,81211 |
| ebi-a-GCST90014006 | eGFR decline (CKD population) | Glycated haemoglobin HbA1c levels | MR Egger | 304 | 0,247712 | 0,120468 | 0,040617 | Diabetes | 128,2125 |
| ebi-a-GCST90014006 | eGFR decline (CKD population) | Glycated haemoglobin HbA1c levels | Weighted median | 304 | 0,140388 | 0,103883 | 0,176565 | Diabetes | 128,2125 |
| ebi-a-GCST90014006 | eGFR decline (CKD population) | Glycated haemoglobin HbA1c levels | Inverse variance weighted | 304 | 0,066236 | 0,063392 | 0,296088 | Diabetes | 128,2125 |
| ebi-a-GCST90014006 | eGFR decline (CKD population) | Glycated haemoglobin HbA1c levels | Simple mode | 304 | 0,122407 | 0,236324 | 0,604862 | Diabetes | 128,2125 |
| ebi-a-GCST90014006 | eGFR decline (CKD population) | Glycated haemoglobin HbA1c levels | Weighted mode | 304 | 0,147713 | 0,14055 | 0,294113 | Diabetes | 128,2125 |
| ebi-a-GCST90014010 | eGFR decline (CKD population) | Phosphate levels | MR Egger | 161 | 0,178765 | 0,167384 | 0,287143 | Minerals | 87,46484 |
| ebi-a-GCST90014010 | eGFR decline (CKD population) | Phosphate levels | Weighted median | 161 | -0,08682 | 0,161229 | 0,590236 | Minerals | 87,46484 |
| ebi-a-GCST90014010 | eGFR decline (CKD population) | Phosphate levels | Inverse variance weighted | 161 | -0,04181 | 0,094267 | 0,657365 | Minerals | 87,46484 |
| ebi-a-GCST90014010 | eGFR decline (CKD population) | Phosphate levels | Simple mode | 161 | 0,004565 | 0,357365 | 0,989824 | Minerals | 87,46484 |
| ebi-a-GCST90014010 | eGFR decline (CKD population) | Phosphate levels | Weighted mode | 161 | 0,135802 | 0,206774 | 0,512273 | Minerals | 87,46484 |
| ebi-a-GCST90014022 | eGFR decline (CKD population) | Bone mineral density | MR Egger | 418 | 0,048085 | 0,108443 | 0,657698 | Bone disease | 120,0164 |
| ebi-a-GCST90014022 | eGFR decline (CKD population) | Bone mineral density | Weighted median | 418 | 0,087093 | 0,085881 | 0,31053 | Bone disease | 120,0164 |
| ebi-a-GCST90014022 | eGFR decline (CKD population) | Bone mineral density | Inverse variance weighted | 418 | 0,076375 | 0,054645 | 0,162217 | Bone disease | 120,0164 |
| ebi-a-GCST90014022 | eGFR decline (CKD population) | Bone mineral density | Simple mode | 418 | 0,219537 | 0,211216 | 0,299223 | Bone disease | 120,0164 |
| ebi-a-GCST90014022 | eGFR decline (CKD population) | Bone mineral density | Weighted mode | 418 | 0,099633 | 0,116625 | 0,39343 | Bone disease | 120,0164 |
| ebi-a-GCST90014023 | eGFR decline (CKD population) | Type 1 diabetes | MR Egger | 76 | -0,01484 | 0,028382 | 0,602537 | Diabetes | 131,7111 |
| ebi-a-GCST90014023 | eGFR decline (CKD population) | Type 1 diabetes | Weighted median | 76 | -0,03538 | 0,027753 | 0,202424 | Diabetes | 131,7111 |
| ebi-a-GCST90014023 | eGFR decline (CKD population) | Type 1 diabetes | Inverse variance weighted | 76 | -0,0124 | 0,017299 | 0,473623 | Diabetes | 131,7111 |
| ebi-a-GCST90014023 | eGFR decline (CKD population) | Type 1 diabetes | Simple mode | 76 | -0,05179 | 0,051599 | 0,318758 | Diabetes | 131,7111 |
| ebi-a-GCST90014023 | eGFR decline (CKD population) | Type 1 diabetes | Weighted mode | 76 | -0,02645 | 0,026406 | 0,319809 | Diabetes | 131,7111 |
| ebi-a-GCST90016674 | eGFR decline (CKD population) | Liver iron content | MR Egger | 6 | -0,09209 | 0,142285 | 0,552746 | Minerals | 221,7646 |
| ebi-a-GCST90016674 | eGFR decline (CKD population) | Liver iron content | Weighted median | 6 | -0,10916 | 0,106537 | 0,305538 | Minerals | 221,7646 |
| ebi-a-GCST90016674 | eGFR decline (CKD population) | Liver iron content | Inverse variance weighted | 6 | -0,10868 | 0,098263 | 0,268712 | Minerals | 221,7646 |
| ebi-a-GCST90016674 | eGFR decline (CKD population) | Liver iron content | Simple mode | 6 | -0,2061 | 0,196115 | 0,341423 | Minerals | 221,7646 |
| ebi-a-GCST90016674 | eGFR decline (CKD population) | Liver iron content | Weighted mode | 6 | -0,11218 | 0,114364 | 0,371699 | Minerals | 221,7646 |
| ebi-a-GCST90016676 | eGFR decline (CKD population) | Pancreas iron content | Wald ratio | 1 | -0,28601 | 0,539392 | 0,595939 | Minerals | 30,56027 |
| ebi-a-GCST90018806 | eGFR decline (CKD population) | Chronic heart failure | MR Egger | 5 | 1,1243 | 1,282342 | 0,445162 | Cardiovascular disease | 41,81649 |
| ebi-a-GCST90018806 | eGFR decline (CKD population) | Chronic heart failure | Weighted median | 5 | 0,161694 | 0,230004 | 0,482052 | Cardiovascular disease | 41,81649 |
| ebi-a-GCST90018806 | eGFR decline (CKD population) | Chronic heart failure | Inverse variance weighted | 5 | 0,15242 | 0,184986 | 0,409965 | Cardiovascular disease | 41,81649 |
| ebi-a-GCST90018806 | eGFR decline (CKD population) | Chronic heart failure | Simple mode | 5 | 0,233287 | 0,306381 | 0,48882 | Cardiovascular disease | 41,81649 |
| ebi-a-GCST90018806 | eGFR decline (CKD population) | Chronic heart failure | Weighted mode | 5 | 0,219318 | 0,305215 | 0,512144 | Cardiovascular disease | 41,81649 |
| ebi-a-GCST90018860 | eGFR decline (CKD population) | Hyperthyroidism | MR Egger | 10 | -0,20596 | 0,215951 | 0,36814 | Thyroid disease | 51,92777 |
| ebi-a-GCST90018860 | eGFR decline (CKD population) | Hyperthyroidism | Weighted median | 10 | 0,116403 | 0,078202 | 0,136622 | Thyroid disease | 51,92777 |
| ebi-a-GCST90018860 | eGFR decline (CKD population) | Hyperthyroidism | Inverse variance weighted | 10 | 0,067028 | 0,067084 | 0,317708 | Thyroid disease | 51,92777 |
| ebi-a-GCST90018860 | eGFR decline (CKD population) | Hyperthyroidism | Simple mode | 10 | 0,149887 | 0,147638 | 0,33651 | Thyroid disease | 51,92777 |
| ebi-a-GCST90018860 | eGFR decline (CKD population) | Hyperthyroidism | Weighted mode | 10 | 0,158342 | 0,147685 | 0,311555 | Thyroid disease | 51,92777 |
| ebi-a-GCST90018862 | eGFR decline (CKD population) | Hypothyroidism | MR Egger | 64 | -0,01569 | 0,08557 | 0,85512 | Thyroid disease | 82,13671 |
| ebi-a-GCST90018862 | eGFR decline (CKD population) | Hypothyroidism | Weighted median | 64 | 0,023343 | 0,059166 | 0,693188 | Thyroid disease | 82,13671 |
| ebi-a-GCST90018862 | eGFR decline (CKD population) | Hypothyroidism | Inverse variance weighted | 64 | 0,06243 | 0,036869 | 0,090396 | Thyroid disease | 82,13671 |
| ebi-a-GCST90018862 | eGFR decline (CKD population) | Hypothyroidism | Simple mode | 64 | 0,029247 | 0,115894 | 0,801587 | Thyroid disease | 82,13671 |
| ebi-a-GCST90018862 | eGFR decline (CKD population) | Hypothyroidism | Weighted mode | 64 | -0,00079 | 0,076955 | 0,991815 | Thyroid disease | 82,13671 |
| ebi-a-GCST90018872 | eGFR decline (CKD population) | Iron deficiency anemia | Wald ratio | 1 | 0,579843 | 0,393979 | 0,141085 | Minerals | 34,53822 |
| ebi-a-GCST90018877 | eGFR decline (CKD population) | Myocardial infarction | MR Egger | 70 | -0,11397 | 0,094674 | 0,232852 | Cardiovascular disease | 66,68595 |
| ebi-a-GCST90018877 | eGFR decline (CKD population) | Myocardial infarction | Weighted median | 70 | 0,013962 | 0,063504 | 0,825977 | Cardiovascular disease | 66,68595 |
| ebi-a-GCST90018877 | eGFR decline (CKD population) | Myocardial infarction | Inverse variance weighted | 70 | 0,012661 | 0,042315 | 0,764772 | Cardiovascular disease | 66,68595 |
| ebi-a-GCST90018877 | eGFR decline (CKD population) | Myocardial infarction | Simple mode | 70 | 0,146033 | 0,143168 | 0,311286 | Cardiovascular disease | 66,68595 |
| ebi-a-GCST90018877 | eGFR decline (CKD population) | Myocardial infarction | Weighted mode | 70 | -0,02538 | 0,0836 | 0,762372 | Cardiovascular disease | 66,68595 |
| ebi-a-GCST90018916 | eGFR decline (CKD population) | Sleep apnea syndrome | Wald ratio | 1 | -0,00869 | 0,236486 | 0,970697 | Sleep disorders | 63,50864 |
| ebi-a-GCST90018977 | eGFR decline (CKD population) | Serum uric acid levels | MR Egger | 238 | -0,01591 | 0,156562 | 0,919147 | Uricemia | 109,8482 |
| ebi-a-GCST90018977 | eGFR decline (CKD population) | Serum uric acid levels | Weighted median | 238 | 0,122066 | 0,141936 | 0,389785 | Uricemia | 109,8482 |
| ebi-a-GCST90018977 | eGFR decline (CKD population) | Serum uric acid levels | Inverse variance weighted | 238 | 0,25169 | 0,08729 | 0,003934 | Uricemia | 109,8482 |
| ebi-a-GCST90018977 | eGFR decline (CKD population) | Serum uric acid levels | Simple mode | 238 | 0,525093 | 0,320498 | 0,102672 | Uricemia | 109,8482 |
| ebi-a-GCST90018977 | eGFR decline (CKD population) | Serum uric acid levels | Weighted mode | 238 | 0,152311 | 0,158963 | 0,338961 | Uricemia | 109,8482 |
| ebi-a-GCST90019417 | eGFR decline (CKD population) | Vitamin K-dependent protein S measurement | MR Egger | 4 | 0,057004 | 0,154245 | 0,747165 | Vitamin | 118,3171 |
| ebi-a-GCST90019417 | eGFR decline (CKD population) | Vitamin K-dependent protein S measurement | Weighted median | 4 | 0,045654 | 0,096125 | 0,634826 | Vitamin | 118,3171 |
| ebi-a-GCST90019417 | eGFR decline (CKD population) | Vitamin K-dependent protein S measurement | Inverse variance weighted | 4 | 0,040674 | 0,088346 | 0,645232 | Vitamin | 118,3171 |
| ebi-a-GCST90019417 | eGFR decline (CKD population) | Vitamin K-dependent protein S measurement | Simple mode | 4 | 0,048446 | 0,140779 | 0,753457 | Vitamin | 118,3171 |
| ebi-a-GCST90019417 | eGFR decline (CKD population) | Vitamin K-dependent protein S measurement | Weighted mode | 4 | 0,048929 | 0,116147 | 0,701919 | Vitamin | 118,3171 |
| ebi-a-GCST90019423 | eGFR decline (CKD population) | Vitamin K-dependent protein C measurement | MR Egger | 3 | 0,082776 | 0,270246 | 0,81078 | Vitamin | 207,8561 |
| ebi-a-GCST90019423 | eGFR decline (CKD population) | Vitamin K-dependent protein C measurement | Weighted median | 3 | 0,073514 | 0,069627 | 0,291046 | Vitamin | 207,8561 |
| ebi-a-GCST90019423 | eGFR decline (CKD population) | Vitamin K-dependent protein C measurement | Inverse variance weighted | 3 | 0,040872 | 0,122425 | 0,738492 | Vitamin | 207,8561 |
| ebi-a-GCST90019423 | eGFR decline (CKD population) | Vitamin K-dependent protein C measurement | Simple mode | 3 | 0,026077 | 0,171693 | 0,893216 | Vitamin | 207,8561 |
| ebi-a-GCST90019423 | eGFR decline (CKD population) | Vitamin K-dependent protein C measurement | Weighted mode | 3 | 0,081488 | 0,072413 | 0,377346 | Vitamin | 207,8561 |
| ebi-a-GCST90025947 | eGFR decline (CKD population) | Serum alkaline phosphatase levels | MR Egger | 335 | -0,11488 | 0,102637 | 0,263821 | Minerals | 166,037 |
| ebi-a-GCST90025947 | eGFR decline (CKD population) | Serum alkaline phosphatase levels | Weighted median | 335 | -0,04276 | 0,100281 | 0,669828 | Minerals | 166,037 |
| ebi-a-GCST90025947 | eGFR decline (CKD population) | Serum alkaline phosphatase levels | Inverse variance weighted | 335 | -0,03143 | 0,060107 | 0,601074 | Minerals | 166,037 |
| ebi-a-GCST90025947 | eGFR decline (CKD population) | Serum alkaline phosphatase levels | Simple mode | 335 | 0,122023 | 0,229892 | 0,595922 | Minerals | 166,037 |
| ebi-a-GCST90025947 | eGFR decline (CKD population) | Serum alkaline phosphatase levels | Weighted mode | 335 | 0,067408 | 0,123684 | 0,586114 | Minerals | 166,037 |
| ebi-a-GCST90025967 | eGFR decline (CKD population) | Vitamin D levels | MR Egger | 81 | -0,02836 | 0,199842 | 0,887494 | Vitamin | 134,1402 |
| ebi-a-GCST90025967 | eGFR decline (CKD population) | Vitamin D levels | Weighted median | 81 | -0,22933 | 0,203703 | 0,260252 | Vitamin | 134,1402 |
| ebi-a-GCST90025967 | eGFR decline (CKD population) | Vitamin D levels | Inverse variance weighted | 81 | -0,0357 | 0,121847 | 0,769538 | Vitamin | 134,1402 |
| ebi-a-GCST90025967 | eGFR decline (CKD population) | Vitamin D levels | Simple mode | 81 | -0,1496 | 0,407253 | 0,714341 | Vitamin | 134,1402 |
| ebi-a-GCST90025967 | eGFR decline (CKD population) | Vitamin D levels | Weighted mode | 81 | -0,1496 | 0,175088 | 0,395427 | Vitamin | 134,1402 |
| ebi-a-GCST90025969 | eGFR decline (CKD population) | Hemoglobin levels | MR Egger | 325 | -0,05348 | 0,137245 | 0,697055 | Organic compound | 104,3556 |
| ebi-a-GCST90025969 | eGFR decline (CKD population) | Hemoglobin levels | Weighted median | 325 | -0,14345 | 0,123667 | 0,246058 | Organic compound | 104,3556 |
| ebi-a-GCST90025969 | eGFR decline (CKD population) | Hemoglobin levels | Inverse variance weighted | 325 | -0,13865 | 0,070832 | 0,050298 | Organic compound | 104,3556 |

| | | | | | | | | | |
|---|---|---|---|---|---|---|---|---|---|
| ebi-a-GCST90025969 | eGFR decline (CKD population) | Hemoglobin levels | Simple mode | 325 | -0,03492 | 0,273176 | 0,898366 | Organic compound | 104,3556 |
| ebi-a-GCST90025969 | eGFR decline (CKD population) | Hemoglobin levels | Weighted mode | 325 | -0,05988 | 0,131368 | 0,64881 | Organic compound | 104,3556 |
| ebi-a-GCST90025986 | eGFR decline (CKD population) | Blood glucose levels | MR Egger | 109 | 0,189135 | 0,189626 | 0,320816 | Diabetes | 118,7554 |
| ebi-a-GCST90025986 | eGFR decline (CKD population) | Blood glucose levels | Weighted median | 109 | 0,214938 | 0,161288 | 0,182653 | Diabetes | 118,7554 |
| ebi-a-GCST90025986 | eGFR decline (CKD population) | Blood glucose levels | Inverse variance weighted | 109 | 0,001075 | 0,106587 | 0,991953 | Diabetes | 118,7554 |
| ebi-a-GCST90025986 | eGFR decline (CKD population) | Blood glucose levels | Simple mode | 109 | 0,009281 | 0,364986 | 0,97976 | Diabetes | 118,7554 |
| ebi-a-GCST90025986 | eGFR decline (CKD population) | Blood glucose levels | Weighted mode | 109 | 0,282754 | 0,153944 | 0,069 | Diabetes | 118,7554 |
| ebi-a-GCST90025989 | eGFR decline (CKD population) | Insulin-like growth factor 1 levels | MR Egger | 374 | -0,18559 | 0,115571 | 0,109163 | Diabetes | 119,1633 |
| ebi-a-GCST90025989 | eGFR decline (CKD population) | Insulin-like growth factor 1 levels | Weighted median | 374 | -0,07365 | 0,101367 | 0,467518 | Diabetes | 119,1633 |
| ebi-a-GCST90025989 | eGFR decline (CKD population) | Insulin-like growth factor 1 levels | Inverse variance weighted | 374 | -4,46E-05 | 0,060843 | 0,999415 | Diabetes | 119,1633 |
| ebi-a-GCST90025989 | eGFR decline (CKD population) | Insulin-like growth factor 1 levels | Simple mode | 374 | -0,05752 | 0,223596 | 0,797112 | Diabetes | 119,1633 |
| ebi-a-GCST90025989 | eGFR decline (CKD population) | Insulin-like growth factor 1 levels | Weighted mode | 374 | -0,11841 | 0,139725 | 0,39729 | Diabetes | 119,1633 |
| ebi-a-GCST90025990 | eGFR decline (CKD population) | Calcium levels | MR Egger | 205 | 0,231355 | 0,151865 | 0,12921 | Minerals | 101,3922 |
| ebi-a-GCST90025990 | eGFR decline (CKD population) | Calcium levels | Weighted median | 205 | -0,0102 | 0,156119 | 0,9479 | Minerals | 101,3922 |
| ebi-a-GCST90025990 | eGFR decline (CKD population) | Calcium levels | Inverse variance weighted | 205 | 0,036767 | 0,0825 | 0,655837 | Minerals | 101,3922 |
| ebi-a-GCST90025990 | eGFR decline (CKD population) | Calcium levels | Simple mode | 205 | 0,174689 | 0,326517 | 0,593227 | Minerals | 101,3922 |
| ebi-a-GCST90025990 | eGFR decline (CKD population) | Calcium levels | Weighted mode | 205 | 0,080092 | 0,159867 | 0,616919 | Minerals | 101,3922 |
| ebi-a-GCST90029013 | eGFR decline (CKD population) | Educational attainment (years of education) | MR Egger | 209 | 0,000996 | 0,11262 | 0,99295 | Social trait | 45,05626 |
| ebi-a-GCST90029013 | eGFR decline (CKD population) | Educational attainment (years of education) | Weighted median | 209 | -0,02665 | 0,038254 | 0,486047 | Social trait | 45,05626 |
| ebi-a-GCST90029013 | eGFR decline (CKD population) | Educational attainment (years of education) | Inverse variance weighted | 209 | -0,05105 | 0,025769 | 0,047606 | Social trait | 45,05626 |
| ebi-a-GCST90029013 | eGFR decline (CKD population) | Educational attainment (years of education) | Simple mode | 209 | -0,03354 | 0,117002 | 0,774677 | Social trait | 45,05626 |
| ebi-a-GCST90029013 | eGFR decline (CKD population) | Educational attainment (years of education) | Weighted mode | 209 | 0,013457 | 0,095452 | 0,888018 | Social trait | 45,05626 |
| ebi-a-GCST90029014 | eGFR decline (CKD population) | Smoking status | MR Egger | 117 | -0,26134 | 1,091491 | 0,811195 | Smoking | 42,91926 |
| ebi-a-GCST90029014 | eGFR decline (CKD population) | Smoking status | Weighted median | 117 | -0,13627 | 0,382354 | 0,721535 | Smoking | 42,91926 |
| ebi-a-GCST90029014 | eGFR decline (CKD population) | Smoking status | Inverse variance weighted | 117 | 0,016079 | 0,257981 | 0,950304 | Smoking | 42,91926 |
| ebi-a-GCST90029014 | eGFR decline (CKD population) | Smoking status | Simple mode | 117 | -0,33645 | 0,912003 | 0,712864 | Smoking | 42,91926 |
| ebi-a-GCST90029014 | eGFR decline (CKD population) | Smoking status | Weighted mode | 117 | -0,24385 | 0,72653 | 0,737749 | Smoking | 42,91926 |
| ebi-a-GCST90029070 | eGFR decline (CKD population) | C-reactive protein levels | MR Egger | 215 | 0,231424 | 0,127682 | 0,071315 | Inflammation | 122,3126 |
| ebi-a-GCST90029070 | eGFR decline (CKD population) | C-reactive protein levels | Weighted median | 215 | 0,042531 | 0,124257 | 0,732138 | Inflammation | 122,3126 |
| ebi-a-GCST90029070 | eGFR decline (CKD population) | C-reactive protein levels | Inverse variance weighted | 215 | 0,082267 | 0,075336 | 0,274833 | Inflammation | 122,3126 |
| ebi-a-GCST90029070 | eGFR decline (CKD population) | C-reactive protein levels | Simple mode | 215 | -0,27044 | 0,271534 | 0,320387 | Inflammation | 122,3126 |
| ebi-a-GCST90029070 | eGFR decline (CKD population) | C-reactive protein levels | Weighted mode | 215 | 0,100473 | 0,124494 | 0,420531 | Inflammation | 122,3126 |
| ebi-a-GCST90029073 | eGFR decline (CKD population) | Liver fat | MR Egger | 8 | 0,316085 | 0,156458 | 0,089869 | Fatty liver | 140,8023 |
| ebi-a-GCST90029073 | eGFR decline (CKD population) | Liver fat | Weighted median | 8 | 0,228512 | 0,121271 | 0,059523 | Fatty liver | 140,8023 |
| ebi-a-GCST90029073 | eGFR decline (CKD population) | Liver fat | Inverse variance weighted | 8 | 0,182498 | 0,09504 | 0,05483 | Fatty liver | 140,8023 |
| ebi-a-GCST90029073 | eGFR decline (CKD population) | Liver fat | Simple mode | 8 | 0,225939 | 0,183485 | 0,257943 | Fatty liver | 140,8023 |
| ebi-a-GCST90029073 | eGFR decline (CKD population) | Liver fat | Weighted mode | 8 | 0,243095 | 0,120114 | 0,082658 | Fatty liver | 140,8023 |
| ebi-a-GCST90086057 | eGFR decline (CKD population) | Cardiovascular disease | Inverse variance weighted | 2 | 0,187045 | 0,206618 | 0,365321 | Cardiovascular disease | 32,018 |
| finn-b-D3_ANAEMIA_B12_DEF | eGFR decline (CKD population) | Vitamin B12 deficiency anaemia | MR Egger | 4 | -0,0502 | 0,163783 | 0,788172 | Vitamin | 52,61875 |
| finn-b-D3_ANAEMIA_B12_DEF | eGFR decline (CKD population) | Vitamin B12 deficiency anaemia | Weighted median | 4 | -0,04114 | 0,060023 | 0,493114 | Vitamin | 52,61875 |
| finn-b-D3_ANAEMIA_B12_DEF | eGFR decline (CKD population) | Vitamin B12 deficiency anaemia | Inverse variance weighted | 4 | -0,01245 | 0,053282 | 0,815284 | Vitamin | 52,61875 |
| finn-b-D3_ANAEMIA_B12_DEF | eGFR decline (CKD population) | Vitamin B12 deficiency anaemia | Simple mode | 4 | -0,05316 | 0,087638 | 0,586942 | Vitamin | 52,61875 |
| finn-b-D3_ANAEMIA_B12_DEF | eGFR decline (CKD population) | Vitamin B12 deficiency anaemia | Weighted mode | 4 | -0,05528 | 0,072788 | 0,502784 | Vitamin | 52,61875 |
| finn-b-E4_OBESITY | eGFR decline (CKD population) | Obesity | MR Egger | 7 | 0,081693 | 0,280762 | 0,782758 | Obesity | 59,53804 |
| finn-b-E4_OBESITY | eGFR decline (CKD population) | Obesity | Weighted median | 7 | -0,03329 | 0,090791 | 0,713846 | Obesity | 59,53804 |
| finn-b-E4_OBESITY | eGFR decline (CKD population) | Obesity | Inverse variance weighted | 7 | 0,026255 | 0,077088 | 0,733418 | Obesity | 59,53804 |
| finn-b-E4_OBESITY | eGFR decline (CKD population) | Obesity | Simple mode | 7 | -0,03948 | 0,155766 | 0,808389 | Obesity | 59,53804 |
| finn-b-E4_OBESITY | eGFR decline (CKD population) | Obesity | Weighted mode | 7 | -0,02375 | 0,106855 | 0,831506 | Obesity | 59,53804 |
| finn-b-FG_PULMHEART | eGFR decline (CKD population) | Pulmonary heart disease | MR Egger | 5 | -0,17522 | 0,150399 | 0,328229 | Cardiovascular disease | 89,00197 |
| finn-b-FG_PULMHEART | eGFR decline (CKD population) | Pulmonary heart disease | Weighted median | 5 | 0,043495 | 0,063687 | 0,494642 | Cardiovascular disease | 89,00197 |
| finn-b-FG_PULMHEART | eGFR decline (CKD population) | Pulmonary heart disease | Inverse variance weighted | 5 | 0,017643 | 0,05085 | 0,728615 | Cardiovascular disease | 89,00197 |
| finn-b-FG_PULMHEART | eGFR decline (CKD population) | Pulmonary heart disease | Simple mode | 5 | 0,087626 | 0,107526 | 0,460849 | Cardiovascular disease | 89,00197 |
| finn-b-FG_PULMHEART | eGFR decline (CKD population) | Pulmonary heart disease | Weighted mode | 5 | 0,095639 | 0,102284 | 0,402709 | Cardiovascular disease | 89,00197 |
| ieu-b-102 | eGFR decline (CKD population) | Major depression | MR Egger | 48 | -0,04976 | 0,810448 | 0,951313 | Mental health | 38,29386 |
| ieu-b-102 | eGFR decline (CKD population) | Major depression | Weighted median | 48 | -0,04009 | 0,199064 | 0,840382 | Mental health | 38,29386 |
| ieu-b-102 | eGFR decline (CKD population) | Major depression | Inverse variance weighted | 48 | -0,08796 | 0,1458 | 0,546334 | Mental health | 38,29386 |
| ieu-b-102 | eGFR decline (CKD population) | Major depression | Simple mode | 48 | -0,07561 | 0,437939 | 0,863669 | Mental health | 38,29386 |
| ieu-b-102 | eGFR decline (CKD population) | Major depression | Weighted mode | 48 | -0,07561 | 0,451892 | 0,867837 | Mental health | 38,29386 |
| ieu-b-107 | eGFR decline (CKD population) | apolipoprotein A-I | MR Egger | 269 | -0,1167 | 0,11121 | 0,294968 | Dyslipidemias | 139,7111 |
| ieu-b-107 | eGFR decline (CKD population) | apolipoprotein A-I | Weighted median | 269 | -0,11897 | 0,109287 | 0,276321 | Dyslipidemias | 139,7111 |
| ieu-b-107 | eGFR decline (CKD population) | apolipoprotein A-I | Inverse variance weighted | 269 | -0,07409 | 0,065723 | 0,259641 | Dyslipidemias | 139,7111 |
| ieu-b-107 | eGFR decline (CKD population) | apolipoprotein A-I | Simple mode | 269 | -0,09103 | 0,221347 | 0,681216 | Dyslipidemias | 139,7111 |
| ieu-b-107 | eGFR decline (CKD population) | apolipoprotein A-I | Weighted mode | 269 | -0,14591 | 0,135793 | 0,283546 | Dyslipidemias | 139,7111 |
| ieu-b-108 | eGFR decline (CKD population) | apolipoprotein B | MR Egger | 175 | 0,069487 | 0,120847 | 0,566039 | Dyslipidemias | 161,0895 |
| ieu-b-108 | eGFR decline (CKD population) | apolipoprotein B | Weighted median | 175 | 0,167183 | 0,120812 | 0,166411 | Dyslipidemias | 161,0895 |
| ieu-b-108 | eGFR decline (CKD population) | apolipoprotein B | Inverse variance weighted | 175 | 0,067709 | 0,074239 | 0,361753 | Dyslipidemias | 161,0895 |
| ieu-b-108 | eGFR decline (CKD population) | apolipoprotein B | Simple mode | 175 | 0,043564 | 0,234079 | 0,852575 | Dyslipidemias | 161,0895 |
| ieu-b-108 | eGFR decline (CKD population) | apolipoprotein B | Weighted mode | 175 | 0,193764 | 0,13591 | 0,155753 | Dyslipidemias | 161,0895 |
| ieu-b-25 | eGFR decline (CKD population) | Cigarettes per Day | MR Egger | 22 | -0,25429 | 0,169551 | 0,149285 | Smoking | 99,80012 |
| ieu-b-25 | eGFR decline (CKD population) | Cigarettes per Day | Weighted median | 22 | -0,15021 | 0,13735 | 0,274109 | Smoking | 99,80012 |
| ieu-b-25 | eGFR decline (CKD population) | Cigarettes per Day | Inverse variance weighted | 22 | -0,16586 | 0,097576 | 0,089159 | Smoking | 99,80012 |
| ieu-b-25 | eGFR decline (CKD population) | Cigarettes per Day | Simple mode | 22 | -0,02222 | 0,288871 | 0,939411 | Smoking | 99,80012 |
| ieu-b-25 | eGFR decline (CKD population) | Cigarettes per Day | Weighted mode | 22 | -0,12072 | 0,130711 | 0,366213 | Smoking | 99,80012 |
| ieu-b-4861 | eGFR decline (CKD population) | Chronotype | MR Egger | 59 | -0,49236 | 0,3671 | 0,185174 | Sleep disorders | 44,49296 |
| ieu-b-4861 | eGFR decline (CKD population) | Chronotype | Weighted median | 59 | -0,05905 | 0,199489 | 0,767211 | Sleep disorders | 44,49296 |
| ieu-b-4861 | eGFR decline (CKD population) | Chronotype | Inverse variance weighted | 59 | 0,0568 | 0,133343 | 0,67013 | Sleep disorders | 44,49296 |
| ieu-b-4861 | eGFR decline (CKD population) | Chronotype | Simple mode | 59 | -0,14152 | 0,450971 | 0,754792 | Sleep disorders | 44,49296 |
| ieu-b-4861 | eGFR decline (CKD population) | Chronotype | Weighted mode | 59 | -0,14152 | 0,411829 | 0,732363 | Sleep disorders | 44,49296 |
| ieu-b-4877 | eGFR decline (CKD population) | smoking initiation | MR Egger | 85 | 1,439223 | 0,588626 | 0,016598 | Smoking | 42,2587 |
| ieu-b-4877 | eGFR decline (CKD population) | smoking initiation | Weighted median | 85 | 0,17844 | 0,168881 | 0,290695 | Smoking | 42,2587 |
| ieu-b-4877 | eGFR decline (CKD population) | smoking initiation | Inverse variance weighted | 85 | 0,170845 | 0,115899 | 0,140458 | Smoking | 42,2587 |
| ieu-b-4877 | eGFR decline (CKD population) | smoking initiation | Simple mode | 85 | 0,278887 | 0,376844 | 0,461327 | Smoking | 42,2587 |
| ieu-b-4877 | eGFR decline (CKD population) | smoking initiation | Weighted mode | 85 | 0,190505 | 0,358675 | 0,596728 | Smoking | 42,2587 |
| ieu-b-5144 | eGFR decline (CKD population) | Hypertension | MR Egger | 192 | -0,84548 | 0,863468 | 0,328744 | Hypertension | 53,79452 |
| ieu-b-5144 | eGFR decline (CKD population) | Hypertension | Weighted median | 192 | -0,2956 | 0,414874 | 0,476152 | Hypertension | 53,79452 |
| ieu-b-5144 | eGFR decline (CKD population) | Hypertension | Inverse variance weighted | 192 | 0,056811 | 0,278408 | 0,83831 | Hypertension | 53,79452 |
| ieu-b-5144 | eGFR decline (CKD population) | Hypertension | Simple mode | 192 | 0,202939 | 1,112608 | 0,855463 | Hypertension | 53,79452 |
| ieu-b-5144 | eGFR decline (CKD population) | Hypertension | Weighted mode | 192 | -1,29952 | 0,869834 | 0,13683 | Hypertension | 53,79452 |
| ieu-b-72 | eGFR decline (CKD population) | Urinary sodium-potassium ratio | MR Egger | 24 | 1,15533 | 1,540232 | 0,461141 | Minerals | 39,88328 |
| ieu-b-72 | eGFR decline (CKD population) | Urinary sodium-potassium ratio | Weighted median | 24 | 0,115441 | 0,466382 | 0,804503 | Minerals | 39,88328 |
| ieu-b-72 | eGFR decline (CKD population) | Urinary sodium-potassium ratio | Inverse variance weighted | 24 | 0,170711 | 0,359346 | 0,634743 | Minerals | 39,88328 |
| ieu-b-72 | eGFR decline (CKD population) | Urinary sodium-potassium ratio | Simple mode | 24 | -0,52443 | 0,845593 | 0,541227 | Minerals | 39,88328 |
| ieu-b-72 | eGFR decline (CKD population) | Urinary sodium-potassium ratio | Weighted mode | 24 | 0,256827 | 0,842179 | 0,763143 | Minerals | 39,88328 |
| prot-a-3102 | eGFR decline (CKD population) | Thyrotropin-releasing hormone | Inverse variance weighted | 2 | 0,147736 | 0,132549 | 0,265029 | Thyroid disease | 36,35446 |
| prot-c-2652_15_1 | eGFR decline (CKD population) | suPAR | Wald ratio | 1 | 0,019723 | 0,102619 | 0,847592 | Biomarkers | 33,5125 |
| ukb-b-4424 | eGFR decline (CKD population) | Sleep duration | MR Egger | 66 | -0,46223 | 1,121994 | 0,681738 | Sleep disorders | 42,80283 |
| ukb-b-4424 | eGFR decline (CKD population) | Sleep duration | Weighted median | 66 | -0,28057 | 0,430239 | 0,514318 | Sleep disorders | 42,80283 |
| ukb-b-4424 | eGFR decline (CKD population) | Sleep duration | Inverse variance weighted | 66 | -0,20247 | 0,293737 | 0,490644 | Sleep disorders | 42,80283 |
| ukb-b-4424 | eGFR decline (CKD population) | Sleep duration | Simple mode | 66 | -1,03155 | 0,989681 | 0,301134 | Sleep disorders | 42,80283 |
| ukb-b-4424 | eGFR decline (CKD population) | Sleep duration | Weighted mode | 66 | -0,79729 | 0,746944 | 0,289741 | Sleep disorders | 42,80283 |
| ukb-b-5779 | eGFR decline (CKD population) | Alcohol intake frequency. | MR Egger | 94 | 0,012633 | 0,263147 | 0,961814 | Alcohol consumption | 53,13815 |
| ukb-b-5779 | eGFR decline (CKD population) | Alcohol intake frequency. | Weighted median | 94 | 0,210716 | 0,2015 | 0,295682 | Alcohol consumption | 53,13815 |
| ukb-b-5779 | eGFR decline (CKD population) | Alcohol intake frequency. | Inverse variance weighted | 94 | 0,068376 | 0,117084 | 0,559224 | Alcohol consumption | 53,13815 |
| ukb-b-5779 | eGFR decline (CKD population) | Alcohol intake frequency. | Simple mode | 94 | -0,45007 | 0,404511 | 0,268735 | Alcohol consumption | 53,13815 |
| ukb-b-5779 | eGFR decline (CKD population) | Alcohol intake frequency. | Weighted mode | 94 | 0,14438 | 0,242864 | 0,55363 | Alcohol consumption | 53,13815 |
| ukb-b-7864 | eGFR decline (CKD population) | Vitamin B6 | Wald ratio | 1 | -0,90794 | 0,826198 | 0,271794 | Vitamin | 32,75897 |
| ukb-d-30790_irnt | eGFR decline (CKD population) | Lipoprotein A | MR Egger | 11 | -0,12549 | 0,200334 | 0,546615 | Dyslipidemias | 985,6435 |
| ukb-d-30790_irnt | eGFR decline (CKD population) | Lipoprotein A | Weighted median | 11 | -0,12587 | 0,140374 | 0,369878 | Dyslipidemias | 985,6435 |
| ukb-d-30790_irnt | eGFR decline (CKD population) | Lipoprotein A | Inverse variance weighted | 11 | -0,06826 | 0,130592 | 0,601189 | Dyslipidemias | 985,6435 |
| ukb-d-30790_irnt | eGFR decline (CKD population) | Lipoprotein A | Simple mode | 11 | -0,00779 | 0,191663 | 0,96839 | Dyslipidemias | 985,6435 |
| ukb-d-30790_irnt | eGFR decline (CKD population) | Lipoprotein A | Weighted mode | 11 | -0,15915 | 0,169087 | 0,368754 | Dyslipidemias | 985,6435 |
| BMI | eGFR decline (CKD population) [DB01] | Body mass index | MR Egger | 506 | 0,075768 | 0,204761 | 0,711514 | Obesity | 74,21321 |
| BMI | eGFR decline (CKD population) [DB01] | Body mass index | Weighted median | 506 | 0,021583 | 0,140711 | 0,878092 | Obesity | 74,21321 |

| | | | | | | | | | |
|---|---|---|---|---|---|---|---|---|---|
| BMI | eGFR decline (CKD population) [DB01] | Body mass index | Inverse variance weighted | 506 | 0,057554 | 0,078697 | 0,464573 | Obesity | 74,21321 |
| BMI | eGFR decline (CKD population) [DB01] | Body mass index | Simple mode | 506 | -0,17422 | 0,434342 | 0,688506 | Obesity | 74,21321 |
| BMI | eGFR decline (CKD population) [DB01] | Body mass index | Weighted mode | 506 | 0,039386 | 0,256543 | 0,878043 | Obesity | 74,21321 |
| DBPTang | eGFR decline (CKD population) [DB01] | Diastolic blood pressure | MR Egger | 251 | -0,12238 | 0,339619 | 0,718884 | Hypertension | 55,91029 |
| DBPTang | eGFR decline (CKD population) [DB01] | Diastolic blood pressure | Weighted median | 251 | -0,08917 | 0,160235 | 0,57789 | Hypertension | 55,91029 |
| DBPTang | eGFR decline (CKD population) [DB01] | Diastolic blood pressure | Inverse variance weighted | 251 | -0,07965 | 0,1057 | 0,451148 | Hypertension | 55,91029 |
| DBPTang | eGFR decline (CKD population) [DB01] | Diastolic blood pressure | Simple mode | 251 | 0,149009 | 0,437722 | 0,733826 | Hypertension | 55,91029 |
| DBPTang | eGFR decline (CKD population) [DB01] | Diastolic blood pressure | Weighted mode | 251 | -0,09496 | 0,372155 | 0,798798 | Hypertension | 55,91029 |
| FT3 | eGFR decline (CKD population) [DB01] | Free triiodothyronine | MR Egger | 6 | -0,44108 | 0,888358 | 0,645577 | Thyroid disease | 82,22035 |
| FT3 | eGFR decline (CKD population) [DB01] | Free triiodothyronine | Weighted median | 6 | -0,18126 | 0,244348 | 0,458213 | Thyroid disease | 82,22035 |
| FT3 | eGFR decline (CKD population) [DB01] | Free triiodothyronine | Inverse variance weighted | 6 | -0,13834 | 0,195687 | 0,479585 | Thyroid disease | 82,22035 |
| FT3 | eGFR decline (CKD population) [DB01] | Free triiodothyronine | Simple mode | 6 | -0,17058 | 0,345975 | 0,642862 | Thyroid disease | 82,22035 |
| FT3 | eGFR decline (CKD population) [DB01] | Free triiodothyronine | Weighted mode | 6 | -0,13375 | 0,307836 | 0,682041 | Thyroid disease | 82,22035 |
| FT3FT4 | eGFR decline (CKD population) [DB01] | FT3 FT4 ratio | MR Egger | 13 | 0,366044 | 1,118687 | 0,749653 | Thyroid disease | 132,3008 |
| FT3FT4 | eGFR decline (CKD population) [DB01] | FT3 FT4 ratio | Weighted median | 13 | -0,22057 | 0,747849 | 0,768045 | Thyroid disease | 132,3008 |
| FT3FT4 | eGFR decline (CKD population) [DB01] | FT3 FT4 ratio | Inverse variance weighted | 13 | -0,77236 | 0,556747 | 0,165357 | Thyroid disease | 132,3008 |
| FT3FT4 | eGFR decline (CKD population) [DB01] | FT3 FT4 ratio | Simple mode | 13 | 0,142095 | 1,454479 | 0,923788 | Thyroid disease | 132,3008 |
| FT3FT4 | eGFR decline (CKD population) [DB01] | FT3 FT4 ratio | Weighted mode | 13 | -0,25019 | 0,795659 | 0,75858 | Thyroid disease | 132,3008 |
| FT4 | eGFR decline (CKD population) [DB01] | Free thyroxine | MR Egger | 62 | -0,0685 | 0,175403 | 0,697522 | Thyroid disease | 84,3797 |
| FT4 | eGFR decline (CKD population) [DB01] | Free thyroxine | Weighted median | 62 | 0,008263 | 0,138741 | 0,952506 | Thyroid disease | 84,3797 |
| FT4 | eGFR decline (CKD population) [DB01] | Free thyroxine | Inverse variance weighted | 62 | -0,01861 | 0,083386 | 0,823411 | Thyroid disease | 84,3797 |
| FT4 | eGFR decline (CKD population) [DB01] | Free thyroxine | Simple mode | 62 | -0,09317 | 0,263807 | 0,72517 | Thyroid disease | 84,3797 |
| FT4 | eGFR decline (CKD population) [DB01] | Free thyroxine | Weighted mode | 62 | -0,01587 | 0,14536 | 0,913412 | Thyroid disease | 84,3797 |
| Fe | eGFR decline (CKD population) [DB01] | Serum iron | Inverse variance weighted | 2 | -0,23071 | 0,620903 | 0,710216 | Minerals | 291,9629 |
| HDL | eGFR decline (CKD population) [DB01] | HDL cholesterol | MR Egger | 233 | 0,015375 | 0,103317 | 0,881831 | Dyslipidemias | 268,9835 |
| HDL | eGFR decline (CKD population) [DB01] | HDL cholesterol | Weighted median | 233 | -0,03941 | 0,105634 | 0,709087 | Dyslipidemias | 268,9835 |
| HDL | eGFR decline (CKD population) [DB01] | HDL cholesterol | Inverse variance weighted | 233 | -0,08483 | 0,066572 | 0,202554 | Dyslipidemias | 268,9835 |
| HDL | eGFR decline (CKD population) [DB01] | HDL cholesterol | Simple mode | 233 | -0,12164 | 0,214945 | 0,571991 | Dyslipidemias | 268,9835 |
| HDL | eGFR decline (CKD population) [DB01] | HDL cholesterol | Weighted mode | 233 | -0,02732 | 0,114077 | 0,810917 | Dyslipidemias | 268,9835 |
| HiTSH | eGFR decline (CKD population) [DB01] | High TSH | MR Egger | 6 | 0,650569 | 0,589538 | 0,331722 | Thyroid disease | 49,53497 |
| HiTSH | eGFR decline (CKD population) [DB01] | High TSH | Weighted median | 6 | 0,301742 | 0,161578 | 0,061837 | Thyroid disease | 49,53497 |
| HiTSH | eGFR decline (CKD population) [DB01] | High TSH | Inverse variance weighted | 6 | 0,298603 | 0,134047 | 0,025907 | Thyroid disease | 49,53497 |
| HiTSH | eGFR decline (CKD population) [DB01] | High TSH | Simple mode | 6 | 0,325815 | 0,240587 | 0,233643 | Thyroid disease | 49,53497 |
| HiTSH | eGFR decline (CKD population) [DB01] | High TSH | Weighted mode | 6 | 0,316438 | 0,189476 | 0,155773 | Thyroid disease | 49,53497 |
| Insom | eGFR decline (CKD population) [DB01] | Insomnia | MR Egger | 13 | 0,016357 | 0,756336 | 0,983133 | Sleep disorders | 44,05724 |
| Insom | eGFR decline (CKD population) [DB01] | Insomnia | Weighted median | 13 | -0,21801 | 0,31433 | 0,487956 | Sleep disorders | 44,05724 |
| Insom | eGFR decline (CKD population) [DB01] | Insomnia | Inverse variance weighted | 13 | 0,076061 | 0,230973 | 0,741922 | Sleep disorders | 44,05724 |
| Insom | eGFR decline (CKD population) [DB01] | Insomnia | Simple mode | 13 | -0,37189 | 0,471767 | 0,445819 | Sleep disorders | 44,05724 |
| Insom | eGFR decline (CKD population) [DB01] | Insomnia | Weighted mode | 13 | -0,28727 | 0,428025 | 0,514841 | Sleep disorders | 44,05724 |
| LDL | eGFR decline (CKD population) [DB01] | LDL cholesterol | MR Egger | 217 | -0,07143 | 0,117089 | 0,542452 | Dyslipidemias | 221,0222 |
| LDL | eGFR decline (CKD population) [DB01] | LDL cholesterol | Weighted median | 217 | -0,09343 | 0,115242 | 0,417517 | Dyslipidemias | 221,0222 |
| LDL | eGFR decline (CKD population) [DB01] | LDL cholesterol | Inverse variance weighted | 217 | -0,07072 | 0,073017 | 0,332747 | Dyslipidemias | 221,0222 |
| LDL | eGFR decline (CKD population) [DB01] | LDL cholesterol | Simple mode | 217 | -0,18403 | 0,237341 | 0,43896 | Dyslipidemias | 221,0222 |
| LDL | eGFR decline (CKD population) [DB01] | LDL cholesterol | Weighted mode | 217 | -0,13004 | 0,126737 | 0,306029 | Dyslipidemias | 221,0222 |
| LoTSH | eGFR decline (CKD population) [DB01] | Low TSH | MR Egger | 26 | 0,001665 | 0,082865 | 0,984134 | Thyroid disease | 70,97091 |
| LoTSH | eGFR decline (CKD population) [DB01] | Low TSH | Weighted median | 26 | 0,021031 | 0,040099 | 0,59995 | Thyroid disease | 70,97091 |
| LoTSH | eGFR decline (CKD population) [DB01] | Low TSH | Inverse variance weighted | 26 | 0,022971 | 0,028066 | 0,413099 | Thyroid disease | 70,97091 |
| LoTSH | eGFR decline (CKD population) [DB01] | Low TSH | Simple mode | 26 | 0,046559 | 0,081025 | 0,570677 | Thyroid disease | 70,97091 |
| LoTSH | eGFR decline (CKD population) [DB01] | Low TSH | Weighted mode | 26 | -0,01397 | 0,063927 | 0,828799 | Thyroid disease | 70,97091 |
| PPTang | eGFR decline (CKD population) [DB01] | Pulse pressure | MR Egger | 300 | 0,103035 | 0,253676 | 0,68491 | Hypertension | 68,57434 |
| PPTang | eGFR decline (CKD population) [DB01] | Pulse pressure | Weighted median | 300 | 0,032203 | 0,15606 | 0,836516 | Hypertension | 68,57434 |
| PPTang | eGFR decline (CKD population) [DB01] | Pulse pressure | Inverse variance weighted | 300 | 0,117554 | 0,094527 | 0,213645 | Hypertension | 68,57434 |
| PPTang | eGFR decline (CKD population) [DB01] | Pulse pressure | Simple mode | 300 | -0,24409 | 0,449368 | 0,587405 | Hypertension | 68,57434 |
| PPTang | eGFR decline (CKD population) [DB01] | Pulse pressure | Weighted mode | 300 | -0,04462 | 0,290988 | 0,878227 | Hypertension | 68,57434 |
| SBPTang | eGFR decline (CKD population) [DB01] | Systolic blood pressure | MR Egger | 251 | -0,25518 | 0,337506 | 0,450321 | Hypertension | 56,96232 |
| SBPTang | eGFR decline (CKD population) [DB01] | Systolic blood pressure | Weighted median | 251 | -0,06377 | 0,164471 | 0,698235 | Hypertension | 56,96232 |
| SBPTang | eGFR decline (CKD population) [DB01] | Systolic blood pressure | Inverse variance weighted | 251 | -0,07037 | 0,10867 | 0,51727 | Hypertension | 56,96232 |
| SBPTang | eGFR decline (CKD population) [DB01] | Systolic blood pressure | Simple mode | 251 | 0,023694 | 0,479114 | 0,960597 | Hypertension | 56,96232 |
| SBPTang | eGFR decline (CKD population) [DB01] | Systolic blood pressure | Weighted mode | 251 | -0,06621 | 0,32627 | 0,839344 | Hypertension | 56,96232 |
| SIndex | eGFR decline (CKD population) [DB01] | Compound smoking index | MR Egger | 121 | 0,276219 | 1,01852 | 0,78671 | Smoking | 44,08236 |
| SIndex | eGFR decline (CKD population) [DB01] | Compound smoking index | Weighted median | 121 | -0,05235 | 0,373381 | 0,888503 | Smoking | 44,08236 |
| SIndex | eGFR decline (CKD population) [DB01] | Compound smoking index | Inverse variance weighted | 121 | -0,25646 | 0,253825 | 0,312312 | Smoking | 44,08236 |
| SIndex | eGFR decline (CKD population) [DB01] | Compound smoking index | Simple mode | 121 | -0,35369 | 0,963366 | 0,71416 | Smoking | 44,08236 |
| SIndex | eGFR decline (CKD population) [DB01] | Compound smoking index | Weighted mode | 121 | -0,04742 | 0,913853 | 0,958701 | Smoking | 44,08236 |
| T2DGGI | eGFR decline (CKD population) [DB01] | Type 2 diabetes | MR Egger | 441 | 0,117887 | 0,070952 | 0,097327 | Diabetes | 93,04757 |
| T2DGGI | eGFR decline (CKD population) [DB01] | Type 2 diabetes | Weighted median | 441 | 0,106737 | 0,070192 | 0,12835 | Diabetes | 93,04757 |
| T2DGGI | eGFR decline (CKD population) [DB01] | Type 2 diabetes | Inverse variance weighted | 441 | 0,08006 | 0,034251 | 0,019416 | Diabetes | 93,04757 |
| T2DGGI | eGFR decline (CKD population) [DB01] | Type 2 diabetes | Simple mode | 441 | 0,116515 | 0,149593 | 0,436472 | Diabetes | 93,04757 |
| T2DGGI | eGFR decline (CKD population) [DB01] | Type 2 diabetes | Weighted mode | 441 | 0,102695 | 0,076988 | 0,182924 | Diabetes | 93,04757 |
| TC | eGFR decline (CKD population) [DB01] | Total cholesterol | MR Egger | 264 | -0,06068 | 0,103577 | 0,558473 | Dyslipidemias | 246,1189 |
| TC | eGFR decline (CKD population) [DB01] | Total cholesterol | Weighted median | 264 | 0,040656 | 0,107997 | 0,706575 | Dyslipidemias | 246,1189 |
| TC | eGFR decline (CKD population) [DB01] | Total cholesterol | Inverse variance weighted | 264 | -0,00276 | 0,065893 | 0,966564 | Dyslipidemias | 246,1189 |
| TC | eGFR decline (CKD population) [DB01] | Total cholesterol | Simple mode | 264 | -0,03999 | 0,24018 | 0,867886 | Dyslipidemias | 246,1189 |
| TC | eGFR decline (CKD population) [DB01] | Total cholesterol | Weighted mode | 264 | -0,01279 | 0,116658 | 0,91278 | Dyslipidemias | 246,1189 |
| TG | eGFR decline (CKD population) [DB01] | Triglyceride | MR Egger | 219 | 0,135118 | 0,107828 | 0,211522 | Dyslipidemias | 231,1265 |
| TG | eGFR decline (CKD population) [DB01] | Triglyceride | Weighted median | 219 | 0,08626 | 0,122632 | 0,481805 | Dyslipidemias | 231,1265 |
| TG | eGFR decline (CKD population) [DB01] | Triglyceride | Inverse variance weighted | 219 | 0,044259 | 0,069595 | 0,524807 | Dyslipidemias | 231,1265 |
| TG | eGFR decline (CKD population) [DB01] | Triglyceride | Simple mode | 219 | -0,05662 | 0,243342 | 0,81624 | Dyslipidemias | 231,1265 |
| TG | eGFR decline (CKD population) [DB01] | Triglyceride | Weighted mode | 219 | 0,099641 | 0,106527 | 0,350636 | Dyslipidemias | 231,1265 |
| TSH | eGFR decline (CKD population) [DB01] | Thyroid Stimulating Hormone | MR Egger | 159 | 0,126483 | 0,100588 | 0,210462 | Thyroid disease | 154,4324 |
| TSH | eGFR decline (CKD population) [DB01] | Thyroid Stimulating Hormone | Weighted median | 159 | 0,099272 | 0,091598 | 0,278464 | Thyroid disease | 154,4324 |
| TSH | eGFR decline (CKD population) [DB01] | Thyroid Stimulating Hormone | Inverse variance weighted | 159 | 0,011792 | 0,055956 | 0,833087 | Thyroid disease | 154,4324 |
| TSH | eGFR decline (CKD population) [DB01] | Thyroid Stimulating Hormone | Simple mode | 159 | 0,003655 | 0,193095 | 0,984924 | Thyroid disease | 154,4324 |
| TSH | eGFR decline (CKD population) [DB01] | Thyroid Stimulating Hormone | Weighted mode | 159 | 0,159234 | 0,100261 | 0,114241 | Thyroid disease | 154,4324 |
| WHR | eGFR decline (CKD population) [DB01] | Waist-to-hip ratio adjusted BMI | MR Egger | 296 | -0,17843 | 0,21734 | 0,412318 | Obesity | 85,86852 |
| WHR | eGFR decline (CKD population) [DB01] | Waist-to-hip ratio adjusted BMI | Weighted median | 296 | 0,001213 | 0,164626 | 0,994122 | Obesity | 85,86852 |
| WHR | eGFR decline (CKD population) [DB01] | Waist-to-hip ratio adjusted BMI | Inverse variance weighted | 296 | 0,030058 | 0,090872 | 0,74082 | Obesity | 85,86852 |
| WHR | eGFR decline (CKD population) [DB01] | Waist-to-hip ratio adjusted BMI | Simple mode | 296 | -0,21378 | 0,401321 | 0,594649 | Obesity | 85,86852 |
| WHR | eGFR decline (CKD population) [DB01] | Waist-to-hip ratio adjusted BMI | Weighted mode | 296 | -0,05155 | 0,223236 | 0,817544 | Obesity | 85,86852 |
| Zn | eGFR decline (CKD population) [DB01] | Zinc | MR Egger | 3 | -0,1862 | 0,359317 | 0,695625 | Minerals | 99,95282 |
| Zn | eGFR decline (CKD population) [DB01] | Zinc | Weighted median | 3 | 0,015998 | 0,090695 | 0,859985 | Minerals | 99,95282 |
| Zn | eGFR decline (CKD population) [DB01] | Zinc | Inverse variance weighted | 3 | 0,05719 | 0,114477 | 0,617377 | Minerals | 99,95282 |
| Zn | eGFR decline (CKD population) [DB01] | Zinc | Simple mode | 3 | -0,0161 | 0,107289 | 0,894493 | Minerals | 99,95282 |
| Zn | eGFR decline (CKD population) [DB01] | Zinc | Weighted mode | 3 | 0,006945 | 0,096264 | 0,949054 | Minerals | 99,95282 |
| ebi-a-GCST006979-consort | eGFR decline (CKD population) [DB01] | Heel bone mineral density | MR Egger | 483 | 0,028563 | 0,100887 | 0,777208 | Bone disease | 151,4375 |
| ebi-a-GCST006979-consort | eGFR decline (CKD population) [DB01] | Heel bone mineral density | Weighted median | 483 | 0,126914 | 0,082901 | 0,125791 | Bone disease | 151,4375 |
| ebi-a-GCST006979-consort | eGFR decline (CKD population) [DB01] | Heel bone mineral density | Inverse variance weighted | 483 | 0,084885 | 0,052101 | 0,103261 | Bone disease | 151,4375 |
| ebi-a-GCST006979-consort | eGFR decline (CKD population) [DB01] | Heel bone mineral density | Simple mode | 483 | 0,154474 | 0,216567 | 0,476013 | Bone disease | 151,4375 |
| ebi-a-GCST006979-consort | eGFR decline (CKD population) [DB01] | Heel bone mineral density | Weighted mode | 483 | 0,12625 | 0,135719 | 0,352715 | Bone disease | 151,4375 |
| ebi-a-GCST90002227 | eGFR decline (CKD population) [DB01] | Two-hour glucose | MR Egger | 14 | -0,16863 | 0,347048 | 0,635795 | Diabetes | 56,47851 |
| ebi-a-GCST90002227 | eGFR decline (CKD population) [DB01] | Two-hour glucose | Weighted median | 14 | 0,192891 | 0,166936 | 0,247896 | Diabetes | 56,47851 |
| ebi-a-GCST90002227 | eGFR decline (CKD population) [DB01] | Two-hour glucose | Inverse variance weighted | 14 | 0,165048 | 0,128186 | 0,197899 | Diabetes | 56,47851 |
| ebi-a-GCST90002227 | eGFR decline (CKD population) [DB01] | Two-hour glucose | Simple mode | 14 | -0,06615 | 0,305805 | 0,832107 | Diabetes | 56,47851 |
| ebi-a-GCST90002227 | eGFR decline (CKD population) [DB01] | Two-hour glucose | Weighted mode | 14 | 0,216309 | 0,249463 | 0,401609 | Diabetes | 56,47851 |
| ebi-a-GCST90002232 | eGFR decline (CKD population) [DB01] | Fasting glucose | MR Egger | 68 | 0,553198 | 0,324213 | 0,092658 | Diabetes | 124,2211 |
| ebi-a-GCST90002232 | eGFR decline (CKD population) [DB01] | Fasting glucose | Weighted median | 68 | 0,548002 | 0,25301 | 0,030317 | Diabetes | 124,2211 |
| ebi-a-GCST90002232 | eGFR decline (CKD population) [DB01] | Fasting glucose | Inverse variance weighted | 68 | 0,185952 | 0,171151 | 0,277267 | Diabetes | 124,2211 |
| ebi-a-GCST90002232 | eGFR decline (CKD population) [DB01] | Fasting glucose | Simple mode | 68 | 0,445719 | 0,472856 | 0,349267 | Diabetes | 124,2211 |
| ebi-a-GCST90002232 | eGFR decline (CKD population) [DB01] | Fasting glucose | Weighted mode | 68 | 0,656546 | 0,252779 | 0,01154 | Diabetes | 124,2211 |
| ebi-a-GCST90002238 | eGFR decline (CKD population) [DB01] | Fasting insulin | MR Egger | 37 | 0,445628 | 1,167969 | 0,705108 | Diabetes | 52,15552 |
| ebi-a-GCST90002238 | eGFR decline (CKD population) [DB01] | Fasting insulin | Weighted median | 37 | 0,397881 | 0,440524 | 0,36642 | Diabetes | 52,15552 |
| ebi-a-GCST90002238 | eGFR decline (CKD population) [DB01] | Fasting insulin | Inverse variance weighted | 37 | -0,01396 | 0,359311 | 0,969017 | Diabetes | 52,15552 |
| ebi-a-GCST90002238 | eGFR decline (CKD population) [DB01] | Fasting insulin | Simple mode | 37 | 0,439013 | 0,876478 | 0,619502 | Diabetes | 52,15552 |

| | | | | | | | | | |
|---|---|---|---|---|---|---|---|---|---|
| ebi-a-GCST90002238 | eGFR decline (CKD population) [DB01] | Fasting insulin | Weighted mode | 37 | 0,404699 | 0,681936 | 0,556586 | Diabetes | 52,15552 |
| ebi-a-GCST90007310 | eGFR decline (CKD population) [DB01] | circulating leptin levels | MR Egger | 3 | 3,075027 | 2,331823 | 0,413038 | Obesity | 56,18818 |
| ebi-a-GCST90007310 | eGFR decline (CKD population) [DB01] | circulating leptin levels | Weighted median | 3 | -0,21522 | 0,329727 | 0,513934 | Obesity | 56,18818 |
| ebi-a-GCST90007310 | eGFR decline (CKD population) [DB01] | circulating leptin levels | Inverse variance weighted | 3 | -0,4125 | 0,309332 | 0,182365 | Obesity | 56,18818 |
| ebi-a-GCST90007310 | eGFR decline (CKD population) [DB01] | circulating leptin levels | Simple mode | 3 | -0,18733 | 0,458523 | 0,72246 | Obesity | 56,18818 |
| ebi-a-GCST90007310 | eGFR decline (CKD population) [DB01] | circulating leptin levels | Weighted mode | 3 | -0,18365 | 0,398671 | 0,690281 | Obesity | 56,18818 |
| ebi-a-GCST90011998 | eGFR decline (CKD population) [DB01] | Growth differentiation factor-15 levels | MR Egger | 3 | -0,17367 | 0,138602 | 0,428816 | Biomarkers | 294,0397 |
| ebi-a-GCST90011998 | eGFR decline (CKD population) [DB01] | Growth differentiation factor-15 levels | Weighted median | 3 | -0,14044 | 0,095117 | 0,139823 | Biomarkers | 294,0397 |
| ebi-a-GCST90011998 | eGFR decline (CKD population) [DB01] | Growth differentiation factor-15 levels | Inverse variance weighted | 3 | -0,12888 | 0,094954 | 0,174696 | Biomarkers | 294,0397 |
| ebi-a-GCST90011998 | eGFR decline (CKD population) [DB01] | Growth differentiation factor-15 levels | Simple mode | 3 | -0,18808 | 0,171827 | 0,387926 | Biomarkers | 294,0397 |
| ebi-a-GCST90011998 | eGFR decline (CKD population) [DB01] | Growth differentiation factor-15 levels | Weighted mode | 3 | -0,14909 | 0,103323 | 0,28582 | Biomarkers | 294,0397 |
| ebi-a-GCST90012022 | eGFR decline (CKD population) [DB01] | Fibroblast growth factor 23 levels | MR Egger | 8 | -0,04796 | 0,482886 | 0,924115 | Biomarkers | 43,42098 |
| ebi-a-GCST90012022 | eGFR decline (CKD population) [DB01] | Fibroblast growth factor 23 levels | Weighted median | 8 | 0,14071 | 0,169965 | 0,407742 | Biomarkers | 43,42098 |
| ebi-a-GCST90012022 | eGFR decline (CKD population) [DB01] | Fibroblast growth factor 23 levels | Inverse variance weighted | 8 | 0,032581 | 0,125049 | 0,794444 | Biomarkers | 43,42098 |
| ebi-a-GCST90012022 | eGFR decline (CKD population) [DB01] | Fibroblast growth factor 23 levels | Simple mode | 8 | 0,149057 | 0,274394 | 0,603836 | Biomarkers | 43,42098 |
| ebi-a-GCST90012022 | eGFR decline (CKD population) [DB01] | Fibroblast growth factor 23 levels | Weighted mode | 8 | 0,181352 | 0,253664 | 0,497798 | Biomarkers | 43,42098 |
| ebi-a-GCST90012041 | eGFR decline (CKD population) [DB01] | Kidney injury molecule 1 levels | MR Egger | 9 | -0,01974 | 0,106036 | 0,857578 | Biomarkers | 190,9807 |
| ebi-a-GCST90012041 | eGFR decline (CKD population) [DB01] | Kidney injury molecule 1 levels | Weighted median | 9 | -0,03128 | 0,065053 | 0,630635 | Biomarkers | 190,9807 |
| ebi-a-GCST90012041 | eGFR decline (CKD population) [DB01] | Kidney injury molecule 1 levels | Inverse variance weighted | 9 | -0,03722 | 0,062877 | 0,553864 | Biomarkers | 190,9807 |
| ebi-a-GCST90012041 | eGFR decline (CKD population) [DB01] | Kidney injury molecule 1 levels | Simple mode | 9 | 0,015223 | 0,132882 | 0,911619 | Biomarkers | 190,9807 |
| ebi-a-GCST90012041 | eGFR decline (CKD population) [DB01] | Kidney injury molecule 1 levels | Weighted mode | 9 | -0,02257 | 0,066798 | 0,744142 | Biomarkers | 190,9807 |
| ebi-a-GCST90013864 | eGFR decline (CKD population) [DB01] | Coronary artery disease (Firth correction) | MR Egger | 37 | -0,10439 | 0,135867 | 0,44746 | Cardiovascular disease | 59,8949 |
| ebi-a-GCST90013864 | eGFR decline (CKD population) [DB01] | Coronary artery disease (Firth correction) | Weighted median | 37 | -0,12363 | 0,08903 | 0,164941 | Cardiovascular disease | 59,8949 |
| ebi-a-GCST90013864 | eGFR decline (CKD population) [DB01] | Coronary artery disease (Firth correction) | Inverse variance weighted | 37 | -0,0334 | 0,061162 | 0,585015 | Cardiovascular disease | 59,8949 |
| ebi-a-GCST90013864 | eGFR decline (CKD population) [DB01] | Coronary artery disease (Firth correction) | Simple mode | 37 | -0,14778 | 0,177465 | 0,410492 | Cardiovascular disease | 59,8949 |
| ebi-a-GCST90013864 | eGFR decline (CKD population) [DB01] | Coronary artery disease (Firth correction) | Weighted mode | 37 | -0,13818 | 0,111844 | 0,224659 | Cardiovascular disease | 59,8949 |
| ebi-a-GCST90013975 | eGFR decline (CKD population) [DB01] | Body fat percentage | MR Egger | 316 | 0,4465 | 0,404517 | 0,270531 | Obesity | 56,17886 |
| ebi-a-GCST90013975 | eGFR decline (CKD population) [DB01] | Body fat percentage | Weighted median | 316 | -0,08294 | 0,187938 | 0,659001 | Obesity | 56,17886 |
| ebi-a-GCST90013975 | eGFR decline (CKD population) [DB01] | Body fat percentage | Inverse variance weighted | 316 | 0,065913 | 0,125687 | 0,599985 | Obesity | 56,17886 |
| ebi-a-GCST90013975 | eGFR decline (CKD population) [DB01] | Body fat percentage | Simple mode | 316 | 0,177096 | 0,578332 | 0,75964 | Obesity | 56,17886 |
| ebi-a-GCST90013975 | eGFR decline (CKD population) [DB01] | Body fat percentage | Weighted mode | 316 | -0,23649 | 0,42237 | 0,575931 | Obesity | 56,17886 |
| ebi-a-GCST90013989 | eGFR decline (CKD population) [DB01] | Sodium in urine | MR Egger | 41 | 1,035035 | 1,102051 | 0,353414 | Minerals | 43,81211 |
| ebi-a-GCST90013989 | eGFR decline (CKD population) [DB01] | Sodium in urine | Weighted median | 41 | -0,3102 | 0,409909 | 0,449192 | Minerals | 43,81211 |
| ebi-a-GCST90013989 | eGFR decline (CKD population) [DB01] | Sodium in urine | Inverse variance weighted | 41 | -0,18647 | 0,283215 | 0,510275 | Minerals | 43,81211 |
| ebi-a-GCST90013989 | eGFR decline (CKD population) [DB01] | Sodium in urine | Simple mode | 41 | -0,25878 | 0,889152 | 0,772524 | Minerals | 43,81211 |
| ebi-a-GCST90013989 | eGFR decline (CKD population) [DB01] | Sodium in urine | Weighted mode | 41 | -0,5671 | 0,841808 | 0,504394 | Minerals | 43,81211 |
| ebi-a-GCST90014006 | eGFR decline (CKD population) [DB01] | Glycated haemoglobin HbA1c levels | MR Egger | 302 | 0,245933 | 0,123526 | 0,047394 | Diabetes | 128,2905 |
| ebi-a-GCST90014006 | eGFR decline (CKD population) [DB01] | Glycated haemoglobin HbA1c levels | Weighted median | 302 | 0,160936 | 0,101208 | 0,1118 | Diabetes | 128,2905 |
| ebi-a-GCST90014006 | eGFR decline (CKD population) [DB01] | Glycated haemoglobin HbA1c levels | Inverse variance weighted | 302 | 0,055304 | 0,065043 | 0,395174 | Diabetes | 128,2905 |
| ebi-a-GCST90014006 | eGFR decline (CKD population) [DB01] | Glycated haemoglobin HbA1c levels | Simple mode | 302 | -0,04754 | 0,255676 | 0,852618 | Diabetes | 128,2905 |
| ebi-a-GCST90014006 | eGFR decline (CKD population) [DB01] | Glycated haemoglobin HbA1c levels | Weighted mode | 302 | 0,103126 | 0,139521 | 0,460393 | Diabetes | 128,2905 |
| ebi-a-GCST90014010 | eGFR decline (CKD population) [DB01] | Phosphate levels | MR Egger | 160 | 0,154558 | 0,170265 | 0,365392 | Minerals | 87,71201 |
| ebi-a-GCST90014010 | eGFR decline (CKD population) [DB01] | Phosphate levels | Weighted median | 160 | -0,01397 | 0,161773 | 0,931188 | Minerals | 87,71201 |
| ebi-a-GCST90014010 | eGFR decline (CKD population) [DB01] | Phosphate levels | Inverse variance weighted | 160 | -0,0473 | 0,095443 | 0,620199 | Minerals | 87,71201 |
| ebi-a-GCST90014010 | eGFR decline (CKD population) [DB01] | Phosphate levels | Simple mode | 160 | -0,01518 | 0,350543 | 0,96552 | Minerals | 87,71201 |
| ebi-a-GCST90014010 | eGFR decline (CKD population) [DB01] | Phosphate levels | Weighted mode | 160 | 0,079048 | 0,200985 | 0,694623 | Minerals | 87,71201 |
| ebi-a-GCST90014022 | eGFR decline (CKD population) [DB01] | Bone mineral density | MR Egger | 415 | 0,060528 | 0,11127 | 0,586754 | Bone disease | 120,0409 |
| ebi-a-GCST90014022 | eGFR decline (CKD population) [DB01] | Bone mineral density | Weighted median | 415 | 0,098058 | 0,090365 | 0,277859 | Bone disease | 120,0409 |
| ebi-a-GCST90014022 | eGFR decline (CKD population) [DB01] | Bone mineral density | Inverse variance weighted | 415 | 0,083392 | 0,056105 | 0,137183 | Bone disease | 120,0409 |
| ebi-a-GCST90014022 | eGFR decline (CKD population) [DB01] | Bone mineral density | Simple mode | 415 | 0,021997 | 0,221198 | 0,920833 | Bone disease | 120,0409 |
| ebi-a-GCST90014022 | eGFR decline (CKD population) [DB01] | Bone mineral density | Weighted mode | 415 | 0,054163 | 0,134103 | 0,686499 | Bone disease | 120,0409 |
| ebi-a-GCST90014023 | eGFR decline (CKD population) [DB01] | Type 1 diabetes | MR Egger | 76 | -0,02482 | 0,029065 | 0,395818 | Diabetes | 131,7111 |
| ebi-a-GCST90014023 | eGFR decline (CKD population) [DB01] | Type 1 diabetes | Weighted median | 76 | -0,02625 | 0,026696 | 0,325489 | Diabetes | 131,7111 |
| ebi-a-GCST90014023 | eGFR decline (CKD population) [DB01] | Type 1 diabetes | Inverse variance weighted | 76 | -0,01951 | 0,017719 | 0,270833 | Diabetes | 131,7111 |
| ebi-a-GCST90014023 | eGFR decline (CKD population) [DB01] | Type 1 diabetes | Simple mode | 76 | -0,04942 | 0,052758 | 0,351869 | Diabetes | 131,7111 |
| ebi-a-GCST90014023 | eGFR decline (CKD population) [DB01] | Type 1 diabetes | Weighted mode | 76 | -0,0399 | 0,025496 | 0,121757 | Diabetes | 131,7111 |
| ebi-a-GCST90016674 | eGFR decline (CKD population) [DB01] | Liver iron content | MR Egger | 5 | -0,06562 | 0,178397 | 0,737407 | Minerals | 257,3642 |
| ebi-a-GCST90016674 | eGFR decline (CKD population) [DB01] | Liver iron content | Weighted median | 5 | -0,07574 | 0,108318 | 0,484407 | Minerals | 257,3642 |
| ebi-a-GCST90016674 | eGFR decline (CKD population) [DB01] | Liver iron content | Inverse variance weighted | 5 | -0,07138 | 0,101871 | 0,483482 | Minerals | 257,3642 |
| ebi-a-GCST90016674 | eGFR decline (CKD population) [DB01] | Liver iron content | Simple mode | 5 | -0,13207 | 0,201804 | 0,548532 | Minerals | 257,3642 |
| ebi-a-GCST90016674 | eGFR decline (CKD population) [DB01] | Liver iron content | Weighted mode | 5 | -0,07796 | 0,106094 | 0,503194 | Minerals | 257,3642 |
| ebi-a-GCST90016676 | eGFR decline (CKD population) [DB01] | Pancreas iron content | Wald ratio | 1 | -0,25399 | 0,552847 | 0,645933 | Minerals | 30,56027 |
| ebi-a-GCST90018806 | eGFR decline (CKD population) [DB01] | Chronic heart failure | MR Egger | 5 | 0,69511 | 1,314204 | 0,633496 | Cardiovascular disease | 41,81649 |
| ebi-a-GCST90018806 | eGFR decline (CKD population) [DB01] | Chronic heart failure | Weighted median | 5 | 0,245943 | 0,228991 | 0,282809 | Cardiovascular disease | 41,81649 |
| ebi-a-GCST90018806 | eGFR decline (CKD population) [DB01] | Chronic heart failure | Inverse variance weighted | 5 | 0,108753 | 0,189579 | 0,566201 | Cardiovascular disease | 41,81649 |
| ebi-a-GCST90018806 | eGFR decline (CKD population) [DB01] | Chronic heart failure | Simple mode | 5 | 0,299567 | 0,331452 | 0,41721 | Cardiovascular disease | 41,81649 |
| ebi-a-GCST90018806 | eGFR decline (CKD population) [DB01] | Chronic heart failure | Weighted mode | 5 | 0,303028 | 0,312919 | 0,387688 | Cardiovascular disease | 41,81649 |
| ebi-a-GCST90018860 | eGFR decline (CKD population) [DB01] | Hyperthyroidism | MR Egger | 8 | -0,15602 | 0,28101 | 0,598821 | Thyroid disease | 53,27573 |
| ebi-a-GCST90018860 | eGFR decline (CKD population) [DB01] | Hyperthyroidism | Weighted median | 8 | 0,160174 | 0,080081 | 0,045484 | Thyroid disease | 53,27573 |
| ebi-a-GCST90018860 | eGFR decline (CKD population) [DB01] | Hyperthyroidism | Inverse variance weighted | 8 | 0,079578 | 0,079452 | 0,316548 | Thyroid disease | 53,27573 |
| ebi-a-GCST90018860 | eGFR decline (CKD population) [DB01] | Hyperthyroidism | Simple mode | 8 | 0,250109 | 0,155753 | 0,152353 | Thyroid disease | 53,27573 |
| ebi-a-GCST90018860 | eGFR decline (CKD population) [DB01] | Hyperthyroidism | Weighted mode | 8 | 0,188529 | 0,148744 | 0,245521 | Thyroid disease | 53,27573 |
| ebi-a-GCST90018862 | eGFR decline (CKD population) [DB01] | Hypothyroidism | MR Egger | 64 | -0,04994 | 0,092397 | 0,590813 | Thyroid disease | 82,13671 |
| ebi-a-GCST90018862 | eGFR decline (CKD population) [DB01] | Hypothyroidism | Weighted median | 64 | 0,026367 | 0,060572 | 0,663343 | Thyroid disease | 82,13671 |
| ebi-a-GCST90018862 | eGFR decline (CKD population) [DB01] | Hypothyroidism | Inverse variance weighted | 64 | 0,033249 | 0,039823 | 0,40377 | Thyroid disease | 82,13671 |
| ebi-a-GCST90018862 | eGFR decline (CKD population) [DB01] | Hypothyroidism | Simple mode | 64 | 0,079802 | 0,122696 | 0,517796 | Thyroid disease | 82,13671 |
| ebi-a-GCST90018862 | eGFR decline (CKD population) [DB01] | Hypothyroidism | Weighted mode | 64 | 0,004507 | 0,083915 | 0,957339 | Thyroid disease | 82,13671 |
| ebi-a-GCST90018872 | eGFR decline (CKD population) [DB01] | Iron deficiency anemia | Wald ratio | 1 | 0,685903 | 0,403885 | 0,089458 | Minerals | 34,53822 |
| ebi-a-GCST90018877 | eGFR decline (CKD population) [DB01] | Myocardial infarction | MR Egger | 70 | -0,1219 | 0,097058 | 0,213421 | Cardiovascular disease | 66,68595 |
| ebi-a-GCST90018877 | eGFR decline (CKD population) [DB01] | Myocardial infarction | Weighted median | 70 | -0,0484 | 0,064781 | 0,455028 | Cardiovascular disease | 66,68595 |
| ebi-a-GCST90018877 | eGFR decline (CKD population) [DB01] | Myocardial infarction | Inverse variance weighted | 70 | 0,020544 | 0,043378 | 0,635777 | Cardiovascular disease | 66,68595 |
| ebi-a-GCST90018877 | eGFR decline (CKD population) [DB01] | Myocardial infarction | Simple mode | 70 | -0,0006 | 0,130661 | 0,996377 | Cardiovascular disease | 66,68595 |
| ebi-a-GCST90018877 | eGFR decline (CKD population) [DB01] | Myocardial infarction | Weighted mode | 70 | -0,06741 | 0,084934 | 0,430117 | Cardiovascular disease | 66,68595 |
| ebi-a-GCST90018916 | eGFR decline (CKD population) [DB01] | Sleep apnea syndrome | Wald ratio | 1 | -0,1053 | 0,242333 | 0,663893 | Sleep disorders | 63,50864 |
| ebi-a-GCST90018977 | eGFR decline (CKD population) [DB01] | Serum uric acid levels | MR Egger | 237 | -0,01262 | 0,156494 | 0,935814 | Uricemia | 110,1094 |
| ebi-a-GCST90018977 | eGFR decline (CKD population) [DB01] | Serum uric acid levels | Weighted median | 237 | 0,114161 | 0,142886 | 0,424311 | Uricemia | 110,1094 |
| ebi-a-GCST90018977 | eGFR decline (CKD population) [DB01] | Serum uric acid levels | Inverse variance weighted | 237 | 0,069549 | 0,086703 | 0,422462 | Uricemia | 110,1094 |
| ebi-a-GCST90018977 | eGFR decline (CKD population) [DB01] | Serum uric acid levels | Simple mode | 237 | 0,365022 | 0,358175 | 0,309191 | Uricemia | 110,1094 |
| ebi-a-GCST90018977 | eGFR decline (CKD population) [DB01] | Serum uric acid levels | Weighted mode | 237 | 0,110212 | 0,175982 | 0,531746 | Uricemia | 110,1094 |
| ebi-a-GCST90019417 | eGFR decline (CKD population) [DB01] | Vitamin K-dependent protein S measurement | MR Egger | 4 | 0,049357 | 0,158194 | 0,784563 | Vitamin | 118,3171 |
| ebi-a-GCST90019417 | eGFR decline (CKD population) [DB01] | Vitamin K-dependent protein S measurement | Weighted median | 4 | 0,019764 | 0,103471 | 0,84852 | Vitamin | 118,3171 |
| ebi-a-GCST90019417 | eGFR decline (CKD population) [DB01] | Vitamin K-dependent protein S measurement | Inverse variance weighted | 4 | 0,011039 | 0,090608 | 0,903036 | Vitamin | 118,3171 |
| ebi-a-GCST90019417 | eGFR decline (CKD population) [DB01] | Vitamin K-dependent protein S measurement | Simple mode | 4 | -0,03216 | 0,138096 | 0,830817 | Vitamin | 118,3171 |
| ebi-a-GCST90019417 | eGFR decline (CKD population) [DB01] | Vitamin K-dependent protein S measurement | Weighted mode | 4 | 0,034422 | 0,109241 | 0,77333 | Vitamin | 118,3171 |
| ebi-a-GCST90019423 | eGFR decline (CKD population) [DB01] | Vitamin K-dependent protein C measurement | MR Egger | 3 | 0,081855 | 0,241378 | 0,79186 | Vitamin | 207,8561 |
| ebi-a-GCST90019423 | eGFR decline (CKD population) [DB01] | Vitamin K-dependent protein C measurement | Weighted median | 3 | 0,070565 | 0,070535 | 0,317103 | Vitamin | 207,8561 |
| ebi-a-GCST90019423 | eGFR decline (CKD population) [DB01] | Vitamin K-dependent protein C measurement | Inverse variance weighted | 3 | 0,042021 | 0,109651 | 0,701552 | Vitamin | 207,8561 |
| ebi-a-GCST90019423 | eGFR decline (CKD population) [DB01] | Vitamin K-dependent protein C measurement | Simple mode | 3 | 0,024397 | 0,1881 | 0,90867 | Vitamin | 207,8561 |
| ebi-a-GCST90019423 | eGFR decline (CKD population) [DB01] | Vitamin K-dependent protein C measurement | Weighted mode | 3 | 0,078998 | 0,073281 | 0,393772 | Vitamin | 207,8561 |
| ebi-a-GCST90025947 | eGFR decline (CKD population) [DB01] | Serum alkaline phosphatase levels | MR Egger | 335 | -0,06945 | 0,102446 | 0,498299 | Minerals | 166,037 |
| ebi-a-GCST90025947 | eGFR decline (CKD population) [DB01] | Serum alkaline phosphatase levels | Weighted median | 335 | 0,121704 | 0,102102 | 0,233269 | Minerals | 166,037 |
| ebi-a-GCST90025947 | eGFR decline (CKD population) [DB01] | Serum alkaline phosphatase levels | Inverse variance weighted | 335 | -0,01368 | 0,059951 | 0,819561 | Minerals | 166,037 |
| ebi-a-GCST90025947 | eGFR decline (CKD population) [DB01] | Serum alkaline phosphatase levels | Simple mode | 335 | 0,162022 | 0,229923 | 0,481501 | Minerals | 166,037 |
| ebi-a-GCST90025947 | eGFR decline (CKD population) [DB01] | Serum alkaline phosphatase levels | Weighted mode | 335 | 0,10909 | 0,117215 | 0,352691 | Minerals | 166,037 |
| ebi-a-GCST90025967 | eGFR decline (CKD population) [DB01] | Vitamin D levels | MR Egger | 81 | -0,0599 | 0,190917 | 0,754544 | Vitamin | 134,1402 |
| ebi-a-GCST90025967 | eGFR decline (CKD population) [DB01] | Vitamin D levels | Weighted median | 81 | -0,21235 | 0,201263 | 0,291376 | Vitamin | 134,1402 |
| ebi-a-GCST90025967 | eGFR decline (CKD population) [DB01] | Vitamin D levels | Inverse variance weighted | 81 | -0,09409 | 0,11713 | 0,421783 | Vitamin | 134,1402 |
| ebi-a-GCST90025967 | eGFR decline (CKD population) [DB01] | Vitamin D levels | Simple mode | 81 | -0,11233 | 0,373562 | 0,764425 | Vitamin | 134,1402 |
| ebi-a-GCST90025967 | eGFR decline (CKD population) [DB01] | Vitamin D levels | Weighted mode | 81 | -0,1359 | 0,17236 | 0,432758 | Vitamin | 134,1402 |
| ebi-a-GCST90025969 | eGFR decline (CKD population) [DB01] | Hemoglobin levels | MR Egger | 325 | -0,00804 | 0,138654 | 0,953772 | Organic compound | 104,3556 |
| ebi-a-GCST90025969 | eGFR decline (CKD population) [DB01] | Hemoglobin levels | Weighted median | 325 | -0,07833 | 0,119235 | 0,511248 | Organic compound | 104,3556 |
| ebi-a-GCST90025969 | eGFR decline (CKD population) [DB01] | Hemoglobin levels | Inverse variance weighted | 325 | -0,10646 | 0,071593 | 0,137014 | Organic compound | 104,3556 |
| ebi-a-GCST90025969 | eGFR decline (CKD population) [DB01] | Hemoglobin levels | Simple mode | 325 | 0,193537 | 0,287185 | 0,500849 | Organic compound | 104,3556 |

| | | | | | | | | | |
|---|---|---|---|---|---|---|---|---|---|
| ebi-a-GCST90025969 | eGFR decline (CKD population) [DB01] | Hemoglobin levels | Weighted mode | 325 | -0,06948 | 0,137169 | 0,612843 | Organic compound | 104,3556 |
| ebi-a-GCST90025986 | eGFR decline (CKD population) [DB01] | Blood glucose levels | MR Egger | 109 | 0,186615 | 0,192815 | 0,335304 | Diabetes | 118,7554 |
| ebi-a-GCST90025986 | eGFR decline (CKD population) [DB01] | Blood glucose levels | Weighted median | 109 | 0,386502 | 0,185302 | 0,036997 | Diabetes | 118,7554 |
| ebi-a-GCST90025986 | eGFR decline (CKD population) [DB01] | Blood glucose levels | Inverse variance weighted | 109 | 0,017402 | 0,108191 | 0,872217 | Diabetes | 118,7554 |
| ebi-a-GCST90025986 | eGFR decline (CKD population) [DB01] | Blood glucose levels | Simple mode | 109 | 0,329492 | 0,382692 | 0,391154 | Diabetes | 118,7554 |
| ebi-a-GCST90025986 | eGFR decline (CKD population) [DB01] | Blood glucose levels | Weighted mode | 109 | 0,329492 | 0,170693 | 0,056189 | Diabetes | 118,7554 |
| ebi-a-GCST90025989 | eGFR decline (CKD population) [DB01] | Insulin-like growth factor 1 levels | MR Egger | 373 | -0,19418 | 0,121507 | 0,110873 | Diabetes | 115,9356 |
| ebi-a-GCST90025989 | eGFR decline (CKD population) [DB01] | Insulin-like growth factor 1 levels | Weighted median | 373 | -0,0122 | 0,10823 | 0,910247 | Diabetes | 115,9356 |
| ebi-a-GCST90025989 | eGFR decline (CKD population) [DB01] | Insulin-like growth factor 1 levels | Inverse variance weighted | 373 | 0,013257 | 0,06293 | 0,833156 | Diabetes | 115,9356 |
| ebi-a-GCST90025989 | eGFR decline (CKD population) [DB01] | Insulin-like growth factor 1 levels | Simple mode | 373 | -0,10295 | 0,263693 | 0,69644 | Diabetes | 115,9356 |
| ebi-a-GCST90025989 | eGFR decline (CKD population) [DB01] | Insulin-like growth factor 1 levels | Weighted mode | 373 | -0,1345 | 0,13494 | 0,31952 | Diabetes | 115,9356 |
| ebi-a-GCST90025990 | eGFR decline (CKD population) [DB01] | Calcium levels | MR Egger | 202 | 0,300841 | 0,153128 | 0,050841 | Minerals | 102,0722 |
| ebi-a-GCST90025990 | eGFR decline (CKD population) [DB01] | Calcium levels | Weighted median | 202 | 0,110435 | 0,173033 | 0,523323 | Minerals | 102,0722 |
| ebi-a-GCST90025990 | eGFR decline (CKD population) [DB01] | Calcium levels | Inverse variance weighted | 202 | 0,066268 | 0,083035 | 0,424827 | Minerals | 102,0722 |
| ebi-a-GCST90025990 | eGFR decline (CKD population) [DB01] | Calcium levels | Simple mode | 202 | -0,01985 | 0,326114 | 0,95153 | Minerals | 102,0722 |
| ebi-a-GCST90025990 | eGFR decline (CKD population) [DB01] | Calcium levels | Weighted mode | 202 | 0,139139 | 0,167892 | 0,408233 | Minerals | 102,0722 |
| ebi-a-GCST90029013 | eGFR decline (CKD population) [DB01] | Educational attainment (years of education) | MR Egger | 209 | 0,028723 | 0,115448 | 0,803763 | Social trait | 45,05626 |
| ebi-a-GCST90029013 | eGFR decline (CKD population) [DB01] | Educational attainment (years of education) | Weighted median | 209 | -0,00498 | 0,040459 | 0,902042 | Social trait | 45,05626 |
| ebi-a-GCST90029013 | eGFR decline (CKD population) [DB01] | Educational attainment (years of education) | Inverse variance weighted | 209 | -0,03734 | 0,026414 | 0,157424 | Social trait | 45,05626 |
| ebi-a-GCST90029013 | eGFR decline (CKD population) [DB01] | Educational attainment (years of education) | Simple mode | 209 | 0,013566 | 0,113295 | 0,904805 | Social trait | 45,05626 |
| ebi-a-GCST90029013 | eGFR decline (CKD population) [DB01] | Educational attainment (years of education) | Weighted mode | 209 | 0,050732 | 0,099672 | 0,611298 | Social trait | 45,05626 |
| ebi-a-GCST90029014 | eGFR decline (CKD population) [DB01] | Smoking status | MR Egger | 117 | -0,40495 | 1,101783 | 0,713888 | Smoking | 42,91926 |
| ebi-a-GCST90029014 | eGFR decline (CKD population) [DB01] | Smoking status | Weighted median | 117 | 0,033669 | 0,380844 | 0,929553 | Smoking | 42,91926 |
| ebi-a-GCST90029014 | eGFR decline (CKD population) [DB01] | Smoking status | Inverse variance weighted | 117 | -0,02011 | 0,261441 | 0,938698 | Smoking | 42,91926 |
| ebi-a-GCST90029014 | eGFR decline (CKD population) [DB01] | Smoking status | Simple mode | 117 | -0,4948 | 0,949971 | 0,603462 | Smoking | 42,91926 |
| ebi-a-GCST90029014 | eGFR decline (CKD population) [DB01] | Smoking status | Weighted mode | 117 | -0,30229 | 0,733348 | 0,68095 | Smoking | 42,91926 |
| ebi-a-GCST90029070 | eGFR decline (CKD population) [DB01] | C-reactive protein levels | MR Egger | 214 | 0,279263 | 0,130798 | 0,033902 | Inflammation | 122,6906 |
| ebi-a-GCST90029070 | eGFR decline (CKD population) [DB01] | C-reactive protein levels | Weighted median | 214 | 0,101449 | 0,129241 | 0,432474 | Inflammation | 122,6906 |
| ebi-a-GCST90029070 | eGFR decline (CKD population) [DB01] | C-reactive protein levels | Inverse variance weighted | 214 | 0,099774 | 0,077225 | 0,19636 | Inflammation | 122,6906 |
| ebi-a-GCST90029070 | eGFR decline (CKD population) [DB01] | C-reactive protein levels | Simple mode | 214 | -0,37402 | 0,291461 | 0,200794 | Inflammation | 122,6906 |
| ebi-a-GCST90029070 | eGFR decline (CKD population) [DB01] | C-reactive protein levels | Weighted mode | 214 | 0,199059 | 0,130852 | 0,129681 | Inflammation | 122,6906 |
| ebi-a-GCST90029073 | eGFR decline (CKD population) [DB01] | Liver fat | MR Egger | 8 | 0,289261 | 0,160518 | 0,121607 | Fatty liver | 140,8023 |
| ebi-a-GCST90029073 | eGFR decline (CKD population) [DB01] | Liver fat | Weighted median | 8 | 0,185404 | 0,114312 | 0,104821 | Fatty liver | 140,8023 |
| ebi-a-GCST90029073 | eGFR decline (CKD population) [DB01] | Liver fat | Inverse variance weighted | 8 | 0,181961 | 0,097508 | 0,062027 | Fatty liver | 140,8023 |
| ebi-a-GCST90029073 | eGFR decline (CKD population) [DB01] | Liver fat | Simple mode | 8 | 0,237487 | 0,173897 | 0,214299 | Fatty liver | 140,8023 |
| ebi-a-GCST90029073 | eGFR decline (CKD population) [DB01] | Liver fat | Weighted mode | 8 | 0,228735 | 0,12413 | 0,107916 | Fatty liver | 140,8023 |
| ebi-a-GCST90086057 | eGFR decline (CKD population) [DB01] | Cardiovascular disease | Inverse variance weighted | 2 | 0,117005 | 0,21181 | 0,580671 | Cardiovascular disease | 32,018 |
| finn-b-D3_ANAEMIA_B12_DEF | eGFR decline (CKD population) [DB01] | Vitamin B12 deficiency anaemia | MR Egger | 4 | -0,07429 | 0,156032 | 0,68091 | Vitamin | 52,61875 |
| finn-b-D3_ANAEMIA_B12_DEF | eGFR decline (CKD population) [DB01] | Vitamin B12 deficiency anaemia | Weighted median | 4 | -0,05699 | 0,063928 | 0,372651 | Vitamin | 52,61875 |
| finn-b-D3_ANAEMIA_B12_DEF | eGFR decline (CKD population) [DB01] | Vitamin B12 deficiency anaemia | Inverse variance weighted | 4 | -0,02937 | 0,054519 | 0,59005 | Vitamin | 52,61875 |
| finn-b-D3_ANAEMIA_B12_DEF | eGFR decline (CKD population) [DB01] | Vitamin B12 deficiency anaemia | Simple mode | 4 | -0,06971 | 0,089358 | 0,492234 | Vitamin | 52,61875 |
| finn-b-D3_ANAEMIA_B12_DEF | eGFR decline (CKD population) [DB01] | Vitamin B12 deficiency anaemia | Weighted mode | 4 | -0,07284 | 0,073876 | 0,396848 | Vitamin | 52,61875 |
| finn-b-E4_OBESITY | eGFR decline (CKD population) [DB01] | Obesity | MR Egger | 7 | -0,05857 | 0,293739 | 0,849813 | Obesity | 59,53804 |
| finn-b-E4_OBESITY | eGFR decline (CKD population) [DB01] | Obesity | Weighted median | 7 | -0,02488 | 0,099536 | 0,802596 | Obesity | 59,53804 |
| finn-b-E4_OBESITY | eGFR decline (CKD population) [DB01] | Obesity | Inverse variance weighted | 7 | 0,026791 | 0,081051 | 0,740993 | Obesity | 59,53804 |
| finn-b-E4_OBESITY | eGFR decline (CKD population) [DB01] | Obesity | Simple mode | 7 | 0,005295 | 0,161399 | 0,974895 | Obesity | 59,53804 |
| finn-b-E4_OBESITY | eGFR decline (CKD population) [DB01] | Obesity | Weighted mode | 7 | -0,06178 | 0,1114 | 0,599206 | Obesity | 59,53804 |
| finn-b-FG_PULMHEART | eGFR decline (CKD population) [DB01] | Pulmonary heart disease | MR Egger | 5 | -0,22911 | 0,15412 | 0,233826 | Cardiovascular disease | 89,00197 |
| finn-b-FG_PULMHEART | eGFR decline (CKD population) [DB01] | Pulmonary heart disease | Weighted median | 5 | 0,078679 | 0,063148 | 0,212782 | Cardiovascular disease | 89,00197 |
| finn-b-FG_PULMHEART | eGFR decline (CKD population) [DB01] | Pulmonary heart disease | Inverse variance weighted | 5 | 0,013069 | 0,057578 | 0,82044 | Cardiovascular disease | 89,00197 |
| finn-b-FG_PULMHEART | eGFR decline (CKD population) [DB01] | Pulmonary heart disease | Simple mode | 5 | 0,111366 | 0,109058 | 0,364911 | Cardiovascular disease | 89,00197 |
| finn-b-FG_PULMHEART | eGFR decline (CKD population) [DB01] | Pulmonary heart disease | Weighted mode | 5 | 0,111366 | 0,106327 | 0,354033 | Cardiovascular disease | 89,00197 |
| ieu-b-102 | eGFR decline (CKD population) [DB01] | Major depression | MR Egger | 48 | 0,282354 | 0,816547 | 0,731077 | Mental health | 38,29386 |
| ieu-b-102 | eGFR decline (CKD population) [DB01] | Major depression | Weighted median | 48 | -0,01348 | 0,207542 | 0,948224 | Mental health | 38,29386 |
| ieu-b-102 | eGFR decline (CKD population) [DB01] | Major depression | Inverse variance weighted | 48 | -0,09779 | 0,147286 | 0,506747 | Mental health | 38,29386 |
| ieu-b-102 | eGFR decline (CKD population) [DB01] | Major depression | Simple mode | 48 | 0,155192 | 0,493254 | 0,754436 | Mental health | 38,29386 |
| ieu-b-102 | eGFR decline (CKD population) [DB01] | Major depression | Weighted mode | 48 | 0,078086 | 0,438161 | 0,859322 | Mental health | 38,29386 |
| ieu-b-107 | eGFR decline (CKD population) [DB01] | apolipoprotein A-I | MR Egger | 269 | -0,11167 | 0,113998 | 0,328175 | Dyslipidemias | 139,7111 |
| ieu-b-107 | eGFR decline (CKD population) [DB01] | apolipoprotein A-I | Weighted median | 269 | -0,11305 | 0,114348 | 0,322834 | Dyslipidemias | 139,7111 |
| ieu-b-107 | eGFR decline (CKD population) [DB01] | apolipoprotein A-I | Inverse variance weighted | 269 | -0,04236 | 0,067368 | 0,529463 | Dyslipidemias | 139,7111 |
| ieu-b-107 | eGFR decline (CKD population) [DB01] | apolipoprotein A-I | Simple mode | 269 | -0,13593 | 0,232073 | 0,558549 | Dyslipidemias | 139,7111 |
| ieu-b-107 | eGFR decline (CKD population) [DB01] | apolipoprotein A-I | Weighted mode | 269 | -0,13593 | 0,135428 | 0,316416 | Dyslipidemias | 139,7111 |
| ieu-b-108 | eGFR decline (CKD population) [DB01] | apolipoprotein B | MR Egger | 173 | 0,072925 | 0,123825 | 0,556681 | Dyslipidemias | 162,3296 |
| ieu-b-108 | eGFR decline (CKD population) [DB01] | apolipoprotein B | Weighted median | 173 | 0,148851 | 0,119018 | 0,211059 | Dyslipidemias | 162,3296 |
| ieu-b-108 | eGFR decline (CKD population) [DB01] | apolipoprotein B | Inverse variance weighted | 173 | 0,094004 | 0,076158 | 0,217081 | Dyslipidemias | 162,3296 |
| ieu-b-108 | eGFR decline (CKD population) [DB01] | apolipoprotein B | Simple mode | 173 | 0,128062 | 0,243919 | 0,600248 | Dyslipidemias | 162,3296 |
| ieu-b-108 | eGFR decline (CKD population) [DB01] | apolipoprotein B | Weighted mode | 173 | 0,151343 | 0,121522 | 0,214678 | Dyslipidemias | 162,3296 |
| ieu-b-25 | eGFR decline (CKD population) [DB01] | Cigarettes per Day | MR Egger | 22 | -0,25477 | 0,173827 | 0,158285 | Smoking | 99,80012 |
| ieu-b-25 | eGFR decline (CKD population) [DB01] | Cigarettes per Day | Weighted median | 22 | -0,18798 | 0,139242 | 0,177003 | Smoking | 99,80012 |
| ieu-b-25 | eGFR decline (CKD population) [DB01] | Cigarettes per Day | Inverse variance weighted | 22 | -0,16854 | 0,100027 | 0,092007 | Smoking | 99,80012 |
| ieu-b-25 | eGFR decline (CKD population) [DB01] | Cigarettes per Day | Simple mode | 22 | -0,50739 | 0,281006 | 0,085335 | Smoking | 99,80012 |
| ieu-b-25 | eGFR decline (CKD population) [DB01] | Cigarettes per Day | Weighted mode | 22 | -0,16327 | 0,13644 | 0,24477 | Smoking | 99,80012 |
| ieu-b-4861 | eGFR decline (CKD population) [DB01] | Chronotype | MR Egger | 60 | -0,45389 | 0,375419 | 0,231555 | Sleep disorders | 44,49296 |
| ieu-b-4861 | eGFR decline (CKD population) [DB01] | Chronotype | Weighted median | 60 | -0,10305 | 0,205799 | 0,616559 | Sleep disorders | 44,49296 |
| ieu-b-4861 | eGFR decline (CKD population) [DB01] | Chronotype | Inverse variance weighted | 60 | -0,00466 | 0,135796 | 0,972632 | Sleep disorders | 44,49296 |
| ieu-b-4861 | eGFR decline (CKD population) [DB01] | Chronotype | Simple mode | 60 | -0,03181 | 0,483322 | 0,947749 | Sleep disorders | 44,49296 |
| ieu-b-4861 | eGFR decline (CKD population) [DB01] | Chronotype | Weighted mode | 60 | -0,12474 | 0,419762 | 0,767376 | Sleep disorders | 44,49296 |
| ieu-b-4877 | eGFR decline (CKD population) [DB01] | smoking initiation | MR Egger | 85 | 1,554247 | 0,60333 | 0,011763 | Smoking | 42,2587 |
| ieu-b-4877 | eGFR decline (CKD population) [DB01] | smoking initiation | Weighted median | 85 | 0,142495 | 0,173181 | 0,410618 | Smoking | 42,2587 |
| ieu-b-4877 | eGFR decline (CKD population) [DB01] | smoking initiation | Inverse variance weighted | 85 | 0,17756 | 0,118789 | 0,134979 | Smoking | 42,2587 |
| ieu-b-4877 | eGFR decline (CKD population) [DB01] | smoking initiation | Simple mode | 85 | 0,298237 | 0,411175 | 0,470267 | Smoking | 42,2587 |
| ieu-b-4877 | eGFR decline (CKD population) [DB01] | smoking initiation | Weighted mode | 85 | 0,227661 | 0,356965 | 0,52536 | Smoking | 42,2587 |
| ieu-b-5144 | eGFR decline (CKD population) [DB01] | Hypertension | MR Egger | 187 | -1,4626 | 0,887094 | 0,100894 | Hypertension | 54,118 |
| ieu-b-5144 | eGFR decline (CKD population) [DB01] | Hypertension | Weighted median | 187 | -0,65719 | 0,439332 | 0,134684 | Hypertension | 54,118 |
| ieu-b-5144 | eGFR decline (CKD population) [DB01] | Hypertension | Inverse variance weighted | 187 | -0,50741 | 0,286911 | 0,076973 | Hypertension | 54,118 |
| ieu-b-5144 | eGFR decline (CKD population) [DB01] | Hypertension | Simple mode | 187 | -1,2332 | 1,162907 | 0,290317 | Hypertension | 54,118 |
| ieu-b-5144 | eGFR decline (CKD population) [DB01] | Hypertension | Weighted mode | 187 | -1,39817 | 0,945445 | 0,140872 | Hypertension | 54,118 |
| ieu-b-72 | eGFR decline (CKD population) [DB01] | Urinary sodium-potassium ratio | MR Egger | 24 | 0,879873 | 1,597087 | 0,587238 | Minerals | 39,88328 |
| ieu-b-72 | eGFR decline (CKD population) [DB01] | Urinary sodium-potassium ratio | Weighted median | 24 | 0,222702 | 0,486591 | 0,647183 | Minerals | 39,88328 |
| ieu-b-72 | eGFR decline (CKD population) [DB01] | Urinary sodium-potassium ratio | Inverse variance weighted | 24 | 0,366708 | 0,369641 | 0,321166 | Minerals | 39,88328 |
| ieu-b-72 | eGFR decline (CKD population) [DB01] | Urinary sodium-potassium ratio | Simple mode | 24 | -0,214 | 0,972745 | 0,827812 | Minerals | 39,88328 |
| ieu-b-72 | eGFR decline (CKD population) [DB01] | Urinary sodium-potassium ratio | Weighted mode | 24 | -0,214 | 0,917723 | 0,817676 | Minerals | 39,88328 |
| prot-a-3102 | eGFR decline (CKD population) [DB01] | Thyrotropin-releasing hormone | Inverse variance weighted | 2 | 0,139421 | 0,135787 | 0,304531 | Thyroid disease | 36,35446 |
| prot-c-2652_15_1 | eGFR decline (CKD population) [DB01] | suPAR | Wald ratio | 1 | 0,036247 | 0,105115 | 0,730218 | Biomarkers | 33,5125 |
| ukb-b-4424 | eGFR decline (CKD population) [DB01] | Sleep duration | MR Egger | 66 | 0,008039 | 1,15011 | 0,994445 | Sleep disorders | 42,80283 |
| ukb-b-4424 | eGFR decline (CKD population) [DB01] | Sleep duration | Weighted median | 66 | -0,129 | 0,430677 | 0,764535 | Sleep disorders | 42,80283 |
| ukb-b-4424 | eGFR decline (CKD population) [DB01] | Sleep duration | Inverse variance weighted | 66 | -0,17059 | 0,301093 | 0,571009 | Sleep disorders | 42,80283 |
| ukb-b-4424 | eGFR decline (CKD population) [DB01] | Sleep duration | Simple mode | 66 | -0,38114 | 0,951974 | 0,6902 | Sleep disorders | 42,80283 |
| ukb-b-4424 | eGFR decline (CKD population) [DB01] | Sleep duration | Weighted mode | 66 | -0,28383 | 0,797944 | 0,723215 | Sleep disorders | 42,80283 |
| ukb-b-5779 | eGFR decline (CKD population) [DB01] | Alcohol intake frequency. | MR Egger | 94 | -0,02846 | 0,270031 | 0,916278 | Alcohol consumption | 53,13815 |
| ukb-b-5779 | eGFR decline (CKD population) [DB01] | Alcohol intake frequency. | Weighted median | 94 | 0,044961 | 0,218073 | 0,836656 | Alcohol consumption | 53,13815 |
| ukb-b-5779 | eGFR decline (CKD population) [DB01] | Alcohol intake frequency. | Inverse variance weighted | 94 | -0,00815 | 0,120036 | 0,945839 | Alcohol consumption | 53,13815 |
| ukb-b-5779 | eGFR decline (CKD population) [DB01] | Alcohol intake frequency. | Simple mode | 94 | -0,32747 | 0,436443 | 0,45496 | Alcohol consumption | 53,13815 |
| ukb-b-5779 | eGFR decline (CKD population) [DB01] | Alcohol intake frequency. | Weighted mode | 94 | -0,05007 | 0,253927 | 0,844121 | Alcohol consumption | 53,13815 |
| ukb-b-7864 | eGFR decline (CKD population) [DB01] | Vitamin B6 | Wald ratio | 1 | -0,88533 | 0,848458 | 0,296737 | Vitamin | 32,75897 |
| ukb-d-30790_irnt | eGFR decline (CKD population) [DB01] | Lipoprotein A | MR Egger | 11 | -0,08765 | 0,186545 | 0,649639 | Dyslipidemias | 985,6435 |
| ukb-d-30790_irnt | eGFR decline (CKD population) [DB01] | Lipoprotein A | Weighted median | 11 | -0,20562 | 0,133682 | 0,124025 | Dyslipidemias | 985,6435 |
| ukb-d-30790_irnt | eGFR decline (CKD population) [DB01] | Lipoprotein A | Inverse variance weighted | 11 | -0,06438 | 0,120688 | 0,593731 | Dyslipidemias | 985,6435 |
| ukb-d-30790_irnt | eGFR decline (CKD population) [DB01] | Lipoprotein A | Simple mode | 11 | -0,20313 | 0,193491 | 0,318521 | Dyslipidemias | 985,6435 |
| ukb-d-30790_irnt | eGFR decline (CKD population) [DB01] | Lipoprotein A | Weighted mode | 11 | -0,18722 | 0,172313 | 0,302744 | Dyslipidemias | 985,6435 |
| BMI | eGFR decline (CKD population) [SH01] | Body mass index | MR Egger | 506 | 0,060835 | 0,207538 | 0,769545 | Obesity | 74,21321 |
| BMI | eGFR decline (CKD population) [SH01] | Body mass index | Weighted median | 506 | 0,004206 | 0,143038 | 0,976543 | Obesity | 74,21321 |
| BMI | eGFR decline (CKD population) [SH01] | Body mass index | Inverse variance weighted | 506 | 0,03574 | 0,079758 | 0,654074 | Obesity | 74,21321 |

| | | | | | | | | | |
|---|---|---|---|---|---|---|---|---|---|
| BMI | eGFR decline (CKD population) [SH01] | Body mass index | Simple mode | 506 | -0,28854 | 0,382126 | 0,450544 | Obesity | 74,21321 |
| BMI | eGFR decline (CKD population) [SH01] | Body mass index | Weighted mode | 506 | -0,00409 | 0,251661 | 0,987045 | Obesity | 74,21321 |
| DBPTang | eGFR decline (CKD population) [SH01] | Diastolic blood pressure | MR Egger | 251 | -0,13362 | 0,344569 | 0,698512 | Hypertension | 55,91029 |
| DBPTang | eGFR decline (CKD population) [SH01] | Diastolic blood pressure | Weighted median | 251 | -0,04121 | 0,157962 | 0,794187 | Hypertension | 55,91029 |
| DBPTang | eGFR decline (CKD population) [SH01] | Diastolic blood pressure | Inverse variance weighted | 251 | -0,11127 | 0,107216 | 0,299338 | Hypertension | 55,91029 |
| DBPTang | eGFR decline (CKD population) [SH01] | Diastolic blood pressure | Simple mode | 251 | 0,114408 | 0,449524 | 0,799311 | Hypertension | 55,91029 |
| DBPTang | eGFR decline (CKD population) [SH01] | Diastolic blood pressure | Weighted mode | 251 | -0,02074 | 0,389966 | 0,957625 | Hypertension | 55,91029 |
| FT3 | eGFR decline (CKD population) [SH01] | Free triiodothyronine | MR Egger | 6 | -0,38342 | 0,909555 | 0,695021 | Thyroid disease | 82,22035 |
| FT3 | eGFR decline (CKD population) [SH01] | Free triiodothyronine | Weighted median | 6 | -0,13621 | 0,245133 | 0,578432 | Thyroid disease | 82,22035 |
| FT3 | eGFR decline (CKD population) [SH01] | Free triiodothyronine | Inverse variance weighted | 6 | -0,11163 | 0,198331 | 0,573552 | Thyroid disease | 82,22035 |
| FT3 | eGFR decline (CKD population) [SH01] | Free triiodothyronine | Simple mode | 6 | -0,14786 | 0,342189 | 0,683664 | Thyroid disease | 82,22035 |
| FT3 | eGFR decline (CKD population) [SH01] | Free triiodothyronine | Weighted mode | 6 | -0,10803 | 0,303657 | 0,736532 | Thyroid disease | 82,22035 |
| FT3FT4 | eGFR decline (CKD population) [SH01] | FT3 FT4 ratio | MR Egger | 13 | 0,452539 | 1,133626 | 0,69739 | Thyroid disease | 132,3008 |
| FT3FT4 | eGFR decline (CKD population) [SH01] | FT3 FT4 ratio | Weighted median | 13 | -0,24806 | 0,760152 | 0,744175 | Thyroid disease | 132,3008 |
| FT3FT4 | eGFR decline (CKD population) [SH01] | FT3 FT4 ratio | Inverse variance weighted | 13 | -0,72728 | 0,564202 | 0,197381 | Thyroid disease | 132,3008 |
| FT3FT4 | eGFR decline (CKD population) [SH01] | FT3 FT4 ratio | Simple mode | 13 | 0,194531 | 1,496335 | 0,898716 | Thyroid disease | 132,3008 |
| FT3FT4 | eGFR decline (CKD population) [SH01] | FT3 FT4 ratio | Weighted mode | 13 | -0,18677 | 0,82635 | 0,824987 | Thyroid disease | 132,3008 |
| FT4 | eGFR decline (CKD population) [SH01] | Free thyroxine | MR Egger | 62 | -0,08311 | 0,182375 | 0,65026 | Thyroid disease | 84,3797 |
| FT4 | eGFR decline (CKD population) [SH01] | Free thyroxine | Weighted median | 62 | -0,00907 | 0,138132 | 0,947648 | Thyroid disease | 84,3797 |
| FT4 | eGFR decline (CKD population) [SH01] | Free thyroxine | Inverse variance weighted | 62 | -0,01476 | 0,086557 | 0,864612 | Thyroid disease | 84,3797 |
| FT4 | eGFR decline (CKD population) [SH01] | Free thyroxine | Simple mode | 62 | -0,06669 | 0,258504 | 0,797278 | Thyroid disease | 84,3797 |
| FT4 | eGFR decline (CKD population) [SH01] | Free thyroxine | Weighted mode | 62 | -0,01032 | 0,144697 | 0,943389 | Thyroid disease | 84,3797 |
| Fe | eGFR decline (CKD population) [SH01] | Serum iron | Inverse variance weighted | 2 | -0,21349 | 0,628838 | 0,734232 | Minerals | 291,9629 |
| HDL | eGFR decline (CKD population) [SH01] | HDL cholesterol | MR Egger | 233 | 0,012929 | 0,104971 | 0,902078 | Dyslipidemias | 268,9835 |
| HDL | eGFR decline (CKD population) [SH01] | HDL cholesterol | Weighted median | 233 | -0,03344 | 0,107395 | 0,75555 | Dyslipidemias | 268,9835 |
| HDL | eGFR decline (CKD population) [SH01] | HDL cholesterol | Inverse variance weighted | 233 | -0,07449 | 0,067585 | 0,270372 | Dyslipidemias | 268,9835 |
| HDL | eGFR decline (CKD population) [SH01] | HDL cholesterol | Simple mode | 233 | -0,06695 | 0,219198 | 0,760301 | Dyslipidemias | 268,9835 |
| HDL | eGFR decline (CKD population) [SH01] | HDL cholesterol | Weighted mode | 233 | -0,01726 | 0,106128 | 0,870972 | Dyslipidemias | 268,9835 |
| HiTSH | eGFR decline (CKD population) [SH01] | High TSH | MR Egger | 6 | 0,687065 | 0,621733 | 0,331121 | Thyroid disease | 49,53497 |
| HiTSH | eGFR decline (CKD population) [SH01] | High TSH | Weighted median | 6 | 0,297983 | 0,172923 | 0,084849 | Thyroid disease | 49,53497 |
| HiTSH | eGFR decline (CKD population) [SH01] | High TSH | Inverse variance weighted | 6 | 0,286476 | 0,142393 | 0,044233 | Thyroid disease | 49,53497 |
| HiTSH | eGFR decline (CKD population) [SH01] | High TSH | Simple mode | 6 | 0,335259 | 0,230635 | 0,205787 | Thyroid disease | 49,53497 |
| HiTSH | eGFR decline (CKD population) [SH01] | High TSH | Weighted mode | 6 | 0,315432 | 0,194482 | 0,16575 | Thyroid disease | 49,53497 |
| Insom | eGFR decline (CKD population) [SH01] | Insomnia | MR Egger | 13 | -0,0147 | 0,776892 | 0,98524 | Sleep disorders | 44,05724 |
| Insom | eGFR decline (CKD population) [SH01] | Insomnia | Weighted median | 13 | -0,24475 | 0,299517 | 0,413841 | Sleep disorders | 44,05724 |
| Insom | eGFR decline (CKD population) [SH01] | Insomnia | Inverse variance weighted | 13 | 0,063942 | 0,237371 | 0,78764 | Sleep disorders | 44,05724 |
| Insom | eGFR decline (CKD population) [SH01] | Insomnia | Simple mode | 13 | -0,41414 | 0,479723 | 0,404905 | Sleep disorders | 44,05724 |
| Insom | eGFR decline (CKD population) [SH01] | Insomnia | Weighted mode | 13 | -0,32773 | 0,402971 | 0,431894 | Sleep disorders | 44,05724 |
| LDL | eGFR decline (CKD population) [SH01] | LDL cholesterol | MR Egger | 217 | -0,07521 | 0,119653 | 0,530289 | Dyslipidemias | 221,0222 |
| LDL | eGFR decline (CKD population) [SH01] | LDL cholesterol | Weighted median | 217 | -0,11401 | 0,117498 | 0,331885 | Dyslipidemias | 221,0222 |
| LDL | eGFR decline (CKD population) [SH01] | LDL cholesterol | Inverse variance weighted | 217 | -0,06775 | 0,074618 | 0,363876 | Dyslipidemias | 221,0222 |
| LDL | eGFR decline (CKD population) [SH01] | LDL cholesterol | Simple mode | 217 | -0,17518 | 0,228187 | 0,443491 | Dyslipidemias | 221,0222 |
| LDL | eGFR decline (CKD population) [SH01] | LDL cholesterol | Weighted mode | 217 | -0,11871 | 0,116457 | 0,309172 | Dyslipidemias | 221,0222 |
| LoTSH | eGFR decline (CKD population) [SH01] | Low TSH | MR Egger | 26 | 0,010305 | 0,08679 | 0,906474 | Thyroid disease | 70,97091 |
| LoTSH | eGFR decline (CKD population) [SH01] | Low TSH | Weighted median | 26 | 0,010152 | 0,041624 | 0,807315 | Thyroid disease | 70,97091 |
| LoTSH | eGFR decline (CKD population) [SH01] | Low TSH | Inverse variance weighted | 26 | 0,026273 | 0,029357 | 0,370817 | Thyroid disease | 70,97091 |
| LoTSH | eGFR decline (CKD population) [SH01] | Low TSH | Simple mode | 26 | 0,022398 | 0,07832 | 0,777252 | Thyroid disease | 70,97091 |
| LoTSH | eGFR decline (CKD population) [SH01] | Low TSH | Weighted mode | 26 | -0,01062 | 0,065036 | 0,87154 | Thyroid disease | 70,97091 |
| PPTang | eGFR decline (CKD population) [SH01] | Pulse pressure | MR Egger | 300 | 0,053365 | 0,257262 | 0,835811 | Hypertension | 68,57434 |
| PPTang | eGFR decline (CKD population) [SH01] | Pulse pressure | Weighted median | 300 | 0,024229 | 0,153604 | 0,874664 | Hypertension | 68,57434 |
| PPTang | eGFR decline (CKD population) [SH01] | Pulse pressure | Inverse variance weighted | 300 | 0,092385 | 0,095831 | 0,335027 | Hypertension | 68,57434 |
| PPTang | eGFR decline (CKD population) [SH01] | Pulse pressure | Simple mode | 300 | -0,26577 | 0,429651 | 0,536662 | Hypertension | 68,57434 |
| PPTang | eGFR decline (CKD population) [SH01] | Pulse pressure | Weighted mode | 300 | -0,06085 | 0,282089 | 0,82936 | Hypertension | 68,57434 |
| SBPTang | eGFR decline (CKD population) [SH01] | Systolic blood pressure | MR Egger | 251 | -0,26958 | 0,34238 | 0,431818 | Hypertension | 56,96232 |
| SBPTang | eGFR decline (CKD population) [SH01] | Systolic blood pressure | Weighted median | 251 | -0,0431 | 0,168265 | 0,797833 | Hypertension | 56,96232 |
| SBPTang | eGFR decline (CKD population) [SH01] | Systolic blood pressure | Inverse variance weighted | 251 | -0,10873 | 0,110197 | 0,323813 | Hypertension | 56,96232 |
| SBPTang | eGFR decline (CKD population) [SH01] | Systolic blood pressure | Simple mode | 251 | 0,030311 | 0,452686 | 0,946668 | Hypertension | 56,96232 |
| SBPTang | eGFR decline (CKD population) [SH01] | Systolic blood pressure | Weighted mode | 251 | -0,03031 | 0,324854 | 0,925741 | Hypertension | 56,96232 |
| SIndex | eGFR decline (CKD population) [SH01] | Compound smoking index | MR Egger | 121 | 0,263457 | 1,031671 | 0,798879 | Smoking | 44,08236 |
| SIndex | eGFR decline (CKD population) [SH01] | Compound smoking index | Weighted median | 121 | 0,020583 | 0,357056 | 0,954031 | Smoking | 44,08236 |
| SIndex | eGFR decline (CKD population) [SH01] | Compound smoking index | Inverse variance weighted | 121 | -0,23781 | 0,257178 | 0,355135 | Smoking | 44,08236 |
| SIndex | eGFR decline (CKD population) [SH01] | Compound smoking index | Simple mode | 121 | -0,24572 | 0,963098 | 0,799055 | Smoking | 44,08236 |
| SIndex | eGFR decline (CKD population) [SH01] | Compound smoking index | Weighted mode | 121 | 0,152609 | 0,886106 | 0,863551 | Smoking | 44,08236 |
| T2DGGI | eGFR decline (CKD population) [SH01] | Type 2 diabetes | MR Egger | 441 | 0,11436 | 0,071898 | 0,112421 | Diabetes | 93,04757 |
| T2DGGI | eGFR decline (CKD population) [SH01] | Type 2 diabetes | Weighted median | 441 | 0,103676 | 0,069574 | 0,136184 | Diabetes | 93,04757 |
| T2DGGI | eGFR decline (CKD population) [SH01] | Type 2 diabetes | Inverse variance weighted | 441 | 0,075011 | 0,03471 | 0,03069 | Diabetes | 93,04757 |
| T2DGGI | eGFR decline (CKD population) [SH01] | Type 2 diabetes | Simple mode | 441 | 0,059107 | 0,153526 | 0,700427 | Diabetes | 93,04757 |
| T2DGGI | eGFR decline (CKD population) [SH01] | Type 2 diabetes | Weighted mode | 441 | 0,086556 | 0,075742 | 0,253752 | Diabetes | 93,04757 |
| TC | eGFR decline (CKD population) [SH01] | Total cholesterol | MR Egger | 264 | -0,06125 | 0,104961 | 0,560004 | Dyslipidemias | 246,1189 |
| TC | eGFR decline (CKD population) [SH01] | Total cholesterol | Weighted median | 264 | 0,043707 | 0,105216 | 0,677845 | Dyslipidemias | 246,1189 |
| TC | eGFR decline (CKD population) [SH01] | Total cholesterol | Inverse variance weighted | 264 | 0,0043 | 0,066783 | 0,948657 | Dyslipidemias | 246,1189 |
| TC | eGFR decline (CKD population) [SH01] | Total cholesterol | Simple mode | 264 | -0,07318 | 0,221412 | 0,741261 | Dyslipidemias | 246,1189 |
| TC | eGFR decline (CKD population) [SH01] | Total cholesterol | Weighted mode | 264 | -0,01913 | 0,124196 | 0,877688 | Dyslipidemias | 246,1189 |
| TG | eGFR decline (CKD population) [SH01] | Triglyceride | MR Egger | 219 | 0,145159 | 0,10933 | 0,185667 | Dyslipidemias | 231,1265 |
| TG | eGFR decline (CKD population) [SH01] | Triglyceride | Weighted median | 219 | 0,089509 | 0,121534 | 0,461431 | Dyslipidemias | 231,1265 |
| TG | eGFR decline (CKD population) [SH01] | Triglyceride | Inverse variance weighted | 219 | 0,038508 | 0,070564 | 0,585263 | Dyslipidemias | 231,1265 |
| TG | eGFR decline (CKD population) [SH01] | Triglyceride | Simple mode | 219 | -0,03471 | 0,251017 | 0,890157 | Dyslipidemias | 231,1265 |
| TG | eGFR decline (CKD population) [SH01] | Triglyceride | Weighted mode | 219 | 0,097426 | 0,103885 | 0,349371 | Dyslipidemias | 231,1265 |
| TSH | eGFR decline (CKD population) [SH01] | Thyroid Stimulating Hormone | MR Egger | 159 | 0,112789 | 0,103301 | 0,276572 | Thyroid disease | 154,4324 |
| TSH | eGFR decline (CKD population) [SH01] | Thyroid Stimulating Hormone | Weighted median | 159 | 0,0881 | 0,090594 | 0,330817 | Thyroid disease | 154,4324 |
| TSH | eGFR decline (CKD population) [SH01] | Thyroid Stimulating Hormone | Inverse variance weighted | 159 | -0,00092 | 0,057433 | 0,987153 | Thyroid disease | 154,4324 |
| TSH | eGFR decline (CKD population) [SH01] | Thyroid Stimulating Hormone | Simple mode | 159 | 0,007156 | 0,204023 | 0,972066 | Thyroid disease | 154,4324 |
| TSH | eGFR decline (CKD population) [SH01] | Thyroid Stimulating Hormone | Weighted mode | 159 | 0,147262 | 0,10032 | 0,144113 | Thyroid disease | 154,4324 |
| WHR | eGFR decline (CKD population) [SH01] | Waist-to-hip ratio adjusted BMI | MR Egger | 296 | -0,18366 | 0,220271 | 0,405071 | Obesity | 85,86852 |
| WHR | eGFR decline (CKD population) [SH01] | Waist-to-hip ratio adjusted BMI | Weighted median | 296 | 0,005442 | 0,163112 | 0,973386 | Obesity | 85,86852 |
| WHR | eGFR decline (CKD population) [SH01] | Waist-to-hip ratio adjusted BMI | Inverse variance weighted | 296 | 0,030287 | 0,092107 | 0,742291 | Obesity | 85,86852 |
| WHR | eGFR decline (CKD population) [SH01] | Waist-to-hip ratio adjusted BMI | Simple mode | 296 | -0,34422 | 0,401816 | 0,392331 | Obesity | 85,86852 |
| WHR | eGFR decline (CKD population) [SH01] | Waist-to-hip ratio adjusted BMI | Weighted mode | 296 | -0,09752 | 0,236385 | 0,680244 | Obesity | 85,86852 |
| Zn | eGFR decline (CKD population) [SH01] | Zinc | MR Egger | 3 | -0,18551 | 0,375319 | 0,707759 | Minerals | 99,95282 |
| Zn | eGFR decline (CKD population) [SH01] | Zinc | Weighted median | 3 | 0,011053 | 0,091916 | 0,904288 | Minerals | 99,95282 |
| Zn | eGFR decline (CKD population) [SH01] | Zinc | Inverse variance weighted | 3 | 0,050923 | 0,116792 | 0,662826 | Minerals | 99,95282 |
| Zn | eGFR decline (CKD population) [SH01] | Zinc | Simple mode | 3 | -0,02765 | 0,120795 | 0,84022 | Minerals | 99,95282 |
| Zn | eGFR decline (CKD population) [SH01] | Zinc | Weighted mode | 3 | 0,00282 | 0,097597 | 0,979573 | Minerals | 99,95282 |
| ebi-a-GCST006979-consort | eGFR decline (CKD population) [SH01] | Heel bone mineral density | MR Egger | 483 | 0,03744 | 0,102242 | 0,714388 | Bone disease | 151,4375 |
| ebi-a-GCST006979-consort | eGFR decline (CKD population) [SH01] | Heel bone mineral density | Weighted median | 483 | 0,129923 | 0,08174 | 0,111954 | Bone disease | 151,4375 |
| ebi-a-GCST006979-consort | eGFR decline (CKD population) [SH01] | Heel bone mineral density | Inverse variance weighted | 483 | 0,083491 | 0,052803 | 0,113835 | Bone disease | 151,4375 |
| ebi-a-GCST006979-consort | eGFR decline (CKD population) [SH01] | Heel bone mineral density | Simple mode | 483 | 0,133381 | 0,208513 | 0,522687 | Bone disease | 151,4375 |
| ebi-a-GCST006979-consort | eGFR decline (CKD population) [SH01] | Heel bone mineral density | Weighted mode | 483 | 0,133381 | 0,124166 | 0,283264 | Bone disease | 151,4375 |
| ebi-a-GCST90002227 | eGFR decline (CKD population) [SH01] | Two-hour glucose | MR Egger | 14 | -0,18554 | 0,359943 | 0,615591 | Diabetes | 56,47851 |
| ebi-a-GCST90002227 | eGFR decline (CKD population) [SH01] | Two-hour glucose | Weighted median | 14 | 0,165001 | 0,169761 | 0,331072 | Diabetes | 56,47851 |
| ebi-a-GCST90002227 | eGFR decline (CKD population) [SH01] | Two-hour glucose | Inverse variance weighted | 14 | 0,16886 | 0,133276 | 0,205158 | Diabetes | 56,47851 |
| ebi-a-GCST90002227 | eGFR decline (CKD population) [SH01] | Two-hour glucose | Simple mode | 14 | -0,10755 | 0,309313 | 0,733623 | Diabetes | 56,47851 |
| ebi-a-GCST90002227 | eGFR decline (CKD population) [SH01] | Two-hour glucose | Weighted mode | 14 | 0,114874 | 0,257732 | 0,663139 | Diabetes | 56,47851 |
| ebi-a-GCST90002232 | eGFR decline (CKD population) [SH01] | Fasting glucose | MR Egger | 68 | 0,534847 | 0,328956 | 0,108739 | Diabetes | 124,2211 |
| ebi-a-GCST90002232 | eGFR decline (CKD population) [SH01] | Fasting glucose | Weighted median | 68 | 0,488808 | 0,265677 | 0,065789 | Diabetes | 124,2211 |
| ebi-a-GCST90002232 | eGFR decline (CKD population) [SH01] | Fasting glucose | Inverse variance weighted | 68 | 0,175493 | 0,173491 | 0,31176 | Diabetes | 124,2211 |
| ebi-a-GCST90002232 | eGFR decline (CKD population) [SH01] | Fasting glucose | Simple mode | 68 | 0,628656 | 0,524836 | 0,23521 | Diabetes | 124,2211 |
| ebi-a-GCST90002232 | eGFR decline (CKD population) [SH01] | Fasting glucose | Weighted mode | 68 | 0,659586 | 0,272219 | 0,018102 | Diabetes | 124,2211 |
| ebi-a-GCST90002238 | eGFR decline (CKD population) [SH01] | Fasting insulin | MR Egger | 37 | 0,501626 | 1,170618 | 0,670903 | Diabetes | 52,15552 |
| ebi-a-GCST90002238 | eGFR decline (CKD population) [SH01] | Fasting insulin | Weighted median | 37 | 0,376156 | 0,462686 | 0,416228 | Diabetes | 52,15552 |
| ebi-a-GCST90002238 | eGFR decline (CKD population) [SH01] | Fasting insulin | Inverse variance weighted | 37 | -0,04608 | 0,360422 | 0,898275 | Diabetes | 52,15552 |
| ebi-a-GCST90002238 | eGFR decline (CKD population) [SH01] | Fasting insulin | Simple mode | 37 | 0,423181 | 0,923703 | 0,649609 | Diabetes | 52,15552 |
| ebi-a-GCST90002238 | eGFR decline (CKD population) [SH01] | Fasting insulin | Weighted mode | 37 | 0,423181 | 0,756332 | 0,579273 | Diabetes | 52,15552 |

| | | | | | | | | | |
|---|---|---|---|---|---|---|---|---|---|
| ebi-a-GCST90007310 | eGFR decline (CKD population) [SH01] | circulating leptin levels | MR Egger | 3 | 3,04429 | 2,361106 | 0,419962 | Obesity | 56,18818 |
| ebi-a-GCST90007310 | eGFR decline (CKD population) [SH01] | circulating leptin levels | Weighted median | 3 | -0,20348 | 0,362256 | 0,574321 | Obesity | 56,18818 |
| ebi-a-GCST90007310 | eGFR decline (CKD population) [SH01] | circulating leptin levels | Inverse variance weighted | 3 | -0,41823 | 0,305611 | 0,171152 | Obesity | 56,18818 |
| ebi-a-GCST90007310 | eGFR decline (CKD population) [SH01] | circulating leptin levels | Simple mode | 3 | -0,19289 | 0,412923 | 0,686355 | Obesity | 56,18818 |
| ebi-a-GCST90007310 | eGFR decline (CKD population) [SH01] | circulating leptin levels | Weighted mode | 3 | -0,19289 | 0,370961 | 0,65491 | Obesity | 56,18818 |
| ebi-a-GCST90011998 | eGFR decline (CKD population) [SH01] | Growth differentiation factor-15 levels | MR Egger | 3 | -0,16146 | 0,140527 | 0,455951 | Biomarkers | 294,0397 |
| ebi-a-GCST90011998 | eGFR decline (CKD population) [SH01] | Growth differentiation factor-15 levels | Weighted median | 3 | -0,1337 | 0,101086 | 0,185948 | Biomarkers | 294,0397 |
| ebi-a-GCST90011998 | eGFR decline (CKD population) [SH01] | Growth differentiation factor-15 levels | Inverse variance weighted | 3 | -0,12421 | 0,096262 | 0,196922 | Biomarkers | 294,0397 |
| ebi-a-GCST90011998 | eGFR decline (CKD population) [SH01] | Growth differentiation factor-15 levels | Simple mode | 3 | -0,20926 | 0,171101 | 0,345869 | Biomarkers | 294,0397 |
| ebi-a-GCST90011998 | eGFR decline (CKD population) [SH01] | Growth differentiation factor-15 levels | Weighted mode | 3 | -0,1412 | 0,097346 | 0,284002 | Biomarkers | 294,0397 |
| ebi-a-GCST90012022 | eGFR decline (CKD population) [SH01] | Fibroblast growth factor 23 levels | MR Egger | 8 | -0,15906 | 0,526626 | 0,772821 | Biomarkers | 43,42098 |
| ebi-a-GCST90012022 | eGFR decline (CKD population) [SH01] | Fibroblast growth factor 23 levels | Weighted median | 8 | 0,13849 | 0,167753 | 0,409055 | Biomarkers | 43,42098 |
| ebi-a-GCST90012022 | eGFR decline (CKD population) [SH01] | Fibroblast growth factor 23 levels | Inverse variance weighted | 8 | 0,036635 | 0,132337 | 0,781908 | Biomarkers | 43,42098 |
| ebi-a-GCST90012022 | eGFR decline (CKD population) [SH01] | Fibroblast growth factor 23 levels | Simple mode | 8 | 0,128301 | 0,27477 | 0,654729 | Biomarkers | 43,42098 |
| ebi-a-GCST90012022 | eGFR decline (CKD population) [SH01] | Fibroblast growth factor 23 levels | Weighted mode | 8 | 0,151692 | 0,255179 | 0,570915 | Biomarkers | 43,42098 |
| ebi-a-GCST90012041 | eGFR decline (CKD population) [SH01] | Kidney injury molecule 1 levels | MR Egger | 9 | -0,00134 | 0,099406 | 0,98965 | Biomarkers | 190,9807 |
| ebi-a-GCST90012041 | eGFR decline (CKD population) [SH01] | Kidney injury molecule 1 levels | Weighted median | 9 | -0,02495 | 0,065401 | 0,70289 | Biomarkers | 190,9807 |
| ebi-a-GCST90012041 | eGFR decline (CKD population) [SH01] | Kidney injury molecule 1 levels | Inverse variance weighted | 9 | -0,04657 | 0,060392 | 0,440614 | Biomarkers | 190,9807 |
| ebi-a-GCST90012041 | eGFR decline (CKD population) [SH01] | Kidney injury molecule 1 levels | Simple mode | 9 | -0,02239 | 0,128368 | 0,865889 | Biomarkers | 190,9807 |
| ebi-a-GCST90012041 | eGFR decline (CKD population) [SH01] | Kidney injury molecule 1 levels | Weighted mode | 9 | -0,02239 | 0,061831 | 0,726684 | Biomarkers | 190,9807 |
| ebi-a-GCST90013864 | eGFR decline (CKD population) [SH01] | Coronary artery disease (Firth correction) | MR Egger | 37 | -0,11643 | 0,137723 | 0,403623 | Cardiovascular disease | 59,8949 |
| ebi-a-GCST90013864 | eGFR decline (CKD population) [SH01] | Coronary artery disease (Firth correction) | Weighted median | 37 | -0,13153 | 0,094209 | 0,162672 | Cardiovascular disease | 59,8949 |
| ebi-a-GCST90013864 | eGFR decline (CKD population) [SH01] | Coronary artery disease (Firth correction) | Inverse variance weighted | 37 | -0,0392 | 0,061998 | 0,527162 | Cardiovascular disease | 59,8949 |
| ebi-a-GCST90013864 | eGFR decline (CKD population) [SH01] | Coronary artery disease (Firth correction) | Simple mode | 37 | -0,16832 | 0,18033 | 0,35683 | Cardiovascular disease | 59,8949 |
| ebi-a-GCST90013864 | eGFR decline (CKD population) [SH01] | Coronary artery disease (Firth correction) | Weighted mode | 37 | -0,15377 | 0,104536 | 0,149986 | Cardiovascular disease | 59,8949 |
| ebi-a-GCST90013975 | eGFR decline (CKD population) [SH01] | Body fat percentage | MR Egger | 316 | 0,460102 | 0,410794 | 0,263557 | Obesity | 56,17886 |
| ebi-a-GCST90013975 | eGFR decline (CKD population) [SH01] | Body fat percentage | Weighted median | 316 | -0,08383 | 0,186755 | 0,65353 | Obesity | 56,17886 |
| ebi-a-GCST90013975 | eGFR decline (CKD population) [SH01] | Body fat percentage | Inverse variance weighted | 316 | 0,049727 | 0,127659 | 0,696883 | Obesity | 56,17886 |
| ebi-a-GCST90013975 | eGFR decline (CKD population) [SH01] | Body fat percentage | Simple mode | 316 | 0,077227 | 0,56388 | 0,891153 | Obesity | 56,17886 |
| ebi-a-GCST90013975 | eGFR decline (CKD population) [SH01] | Body fat percentage | Weighted mode | 316 | -0,17781 | 0,432028 | 0,680937 | Obesity | 56,17886 |
| ebi-a-GCST90013989 | eGFR decline (CKD population) [SH01] | Sodium in urine | MR Egger | 41 | 0,941057 | 1,117942 | 0,405045 | Minerals | 43,81211 |
| ebi-a-GCST90013989 | eGFR decline (CKD population) [SH01] | Sodium in urine | Weighted median | 41 | -0,20016 | 0,411773 | 0,626905 | Minerals | 43,81211 |
| ebi-a-GCST90013989 | eGFR decline (CKD population) [SH01] | Sodium in urine | Inverse variance weighted | 41 | -0,19289 | 0,28722 | 0,501845 | Minerals | 43,81211 |
| ebi-a-GCST90013989 | eGFR decline (CKD population) [SH01] | Sodium in urine | Simple mode | 41 | -0,23449 | 0,887344 | 0,792936 | Minerals | 43,81211 |
| ebi-a-GCST90013989 | eGFR decline (CKD population) [SH01] | Sodium in urine | Weighted mode | 41 | -0,45913 | 0,862956 | 0,59764 | Minerals | 43,81211 |
| ebi-a-GCST90014006 | eGFR decline (CKD population) [SH01] | Glycated haemoglobin HbA1c levels | MR Egger | 302 | 0,246004 | 0,125196 | 0,050343 | Diabetes | 128,2905 |
| ebi-a-GCST90014006 | eGFR decline (CKD population) [SH01] | Glycated haemoglobin HbA1c levels | Weighted median | 302 | 0,139854 | 0,104893 | 0,182433 | Diabetes | 128,2905 |
| ebi-a-GCST90014006 | eGFR decline (CKD population) [SH01] | Glycated haemoglobin HbA1c levels | Inverse variance weighted | 302 | 0,053109 | 0,06592 | 0,420444 | Diabetes | 128,2905 |
| ebi-a-GCST90014006 | eGFR decline (CKD population) [SH01] | Glycated haemoglobin HbA1c levels | Simple mode | 302 | -0,06417 | 0,231971 | 0,782257 | Diabetes | 128,2905 |
| ebi-a-GCST90014006 | eGFR decline (CKD population) [SH01] | Glycated haemoglobin HbA1c levels | Weighted mode | 302 | 0,087327 | 0,139658 | 0,532252 | Diabetes | 128,2905 |
| ebi-a-GCST90014010 | eGFR decline (CKD population) [SH01] | Phosphate levels | MR Egger | 160 | 0,150083 | 0,172702 | 0,386149 | Minerals | 87,71201 |
| ebi-a-GCST90014010 | eGFR decline (CKD population) [SH01] | Phosphate levels | Weighted median | 160 | 0,018705 | 0,166002 | 0,910286 | Minerals | 87,71201 |
| ebi-a-GCST90014010 | eGFR decline (CKD population) [SH01] | Phosphate levels | Inverse variance weighted | 160 | -0,04982 | 0,0968 | 0,606792 | Minerals | 87,71201 |
| ebi-a-GCST90014010 | eGFR decline (CKD population) [SH01] | Phosphate levels | Simple mode | 160 | 0,056506 | 0,365109 | 0,877202 | Minerals | 87,71201 |
| ebi-a-GCST90014010 | eGFR decline (CKD population) [SH01] | Phosphate levels | Weighted mode | 160 | 0,151757 | 0,20001 | 0,449126 | Minerals | 87,71201 |
| ebi-a-GCST90014022 | eGFR decline (CKD population) [SH01] | Bone mineral density | MR Egger | 415 | 0,06426 | 0,112765 | 0,569086 | Bone disease | 120,0409 |
| ebi-a-GCST90014022 | eGFR decline (CKD population) [SH01] | Bone mineral density | Weighted median | 415 | 0,067134 | 0,094705 | 0,478404 | Bone disease | 120,0409 |
| ebi-a-GCST90014022 | eGFR decline (CKD population) [SH01] | Bone mineral density | Inverse variance weighted | 415 | 0,084138 | 0,056861 | 0,138952 | Bone disease | 120,0409 |
| ebi-a-GCST90014022 | eGFR decline (CKD population) [SH01] | Bone mineral density | Simple mode | 415 | 0,028518 | 0,218679 | 0,896305 | Bone disease | 120,0409 |
| ebi-a-GCST90014022 | eGFR decline (CKD population) [SH01] | Bone mineral density | Weighted mode | 415 | 0,061312 | 0,141925 | 0,665966 | Bone disease | 120,0409 |
| ebi-a-GCST90014023 | eGFR decline (CKD population) [SH01] | Type 1 diabetes | MR Egger | 76 | -0,02712 | 0,02945 | 0,360101 | Diabetes | 131,7111 |
| ebi-a-GCST90014023 | eGFR decline (CKD population) [SH01] | Type 1 diabetes | Weighted median | 76 | -0,03073 | 0,0278 | 0,269049 | Diabetes | 131,7111 |
| ebi-a-GCST90014023 | eGFR decline (CKD population) [SH01] | Type 1 diabetes | Inverse variance weighted | 76 | -0,02114 | 0,017956 | 0,239089 | Diabetes | 131,7111 |
| ebi-a-GCST90014023 | eGFR decline (CKD population) [SH01] | Type 1 diabetes | Simple mode | 76 | -0,05421 | 0,053063 | 0,310272 | Diabetes | 131,7111 |
| ebi-a-GCST90014023 | eGFR decline (CKD population) [SH01] | Type 1 diabetes | Weighted mode | 76 | -0,0411 | 0,026555 | 0,125872 | Diabetes | 131,7111 |
| ebi-a-GCST90016674 | eGFR decline (CKD population) [SH01] | Liver iron content | MR Egger | 5 | -0,05218 | 0,181683 | 0,792633 | Minerals | 257,3642 |
| ebi-a-GCST90016674 | eGFR decline (CKD population) [SH01] | Liver iron content | Weighted median | 5 | -0,06804 | 0,105235 | 0,517897 | Minerals | 257,3642 |
| ebi-a-GCST90016674 | eGFR decline (CKD population) [SH01] | Liver iron content | Inverse variance weighted | 5 | -0,06421 | 0,10326 | 0,534048 | Minerals | 257,3642 |
| ebi-a-GCST90016674 | eGFR decline (CKD population) [SH01] | Liver iron content | Simple mode | 5 | -0,12579 | 0,198861 | 0,561391 | Minerals | 257,3642 |
| ebi-a-GCST90016674 | eGFR decline (CKD population) [SH01] | Liver iron content | Weighted mode | 5 | -0,06917 | 0,108064 | 0,556945 | Minerals | 257,3642 |
| ebi-a-GCST90016676 | eGFR decline (CKD population) [SH01] | Pancreas iron content | Wald ratio | 1 | -0,24641 | 0,560029 | 0,659935 | Minerals | 30,56027 |
| ebi-a-GCST90018806 | eGFR decline (CKD population) [SH01] | Chronic heart failure | MR Egger | 5 | 0,59422 | 1,331888 | 0,685726 | Cardiovascular disease | 41,81649 |
| ebi-a-GCST90018806 | eGFR decline (CKD population) [SH01] | Chronic heart failure | Weighted median | 5 | 0,247266 | 0,233694 | 0,290021 | Cardiovascular disease | 41,81649 |
| ebi-a-GCST90018806 | eGFR decline (CKD population) [SH01] | Chronic heart failure | Inverse variance weighted | 5 | 0,098323 | 0,192124 | 0,608813 | Cardiovascular disease | 41,81649 |
| ebi-a-GCST90018806 | eGFR decline (CKD population) [SH01] | Chronic heart failure | Simple mode | 5 | 0,302544 | 0,321075 | 0,399401 | Cardiovascular disease | 41,81649 |
| ebi-a-GCST90018806 | eGFR decline (CKD population) [SH01] | Chronic heart failure | Weighted mode | 5 | 0,302544 | 0,312485 | 0,387777 | Cardiovascular disease | 41,81649 |
| ebi-a-GCST90018860 | eGFR decline (CKD population) [SH01] | Hyperthyroidism | MR Egger | 8 | -0,16461 | 0,287151 | 0,587291 | Thyroid disease | 53,27573 |
| ebi-a-GCST90018860 | eGFR decline (CKD population) [SH01] | Hyperthyroidism | Weighted median | 8 | 0,170337 | 0,085558 | 0,046493 | Thyroid disease | 53,27573 |
| ebi-a-GCST90018860 | eGFR decline (CKD population) [SH01] | Hyperthyroidism | Inverse variance weighted | 8 | 0,077847 | 0,081245 | 0,337972 | Thyroid disease | 53,27573 |
| ebi-a-GCST90018860 | eGFR decline (CKD population) [SH01] | Hyperthyroidism | Simple mode | 8 | 0,272938 | 0,158378 | 0,128491 | Thyroid disease | 53,27573 |
| ebi-a-GCST90018860 | eGFR decline (CKD population) [SH01] | Hyperthyroidism | Weighted mode | 8 | 0,192069 | 0,1508 | 0,243438 | Thyroid disease | 53,27573 |
| ebi-a-GCST90018862 | eGFR decline (CKD population) [SH01] | Hypothyroidism | MR Egger | 64 | -0,05789 | 0,094707 | 0,543251 | Thyroid disease | 82,13671 |
| ebi-a-GCST90018862 | eGFR decline (CKD population) [SH01] | Hypothyroidism | Weighted median | 64 | 0,024643 | 0,059782 | 0,680183 | Thyroid disease | 82,13671 |
| ebi-a-GCST90018862 | eGFR decline (CKD population) [SH01] | Hypothyroidism | Inverse variance weighted | 64 | 0,02666 | 0,040822 | 0,513714 | Thyroid disease | 82,13671 |
| ebi-a-GCST90018862 | eGFR decline (CKD population) [SH01] | Hypothyroidism | Simple mode | 64 | 0,093244 | 0,131597 | 0,481211 | Thyroid disease | 82,13671 |
| ebi-a-GCST90018862 | eGFR decline (CKD population) [SH01] | Hypothyroidism | Weighted mode | 64 | 0,041792 | 0,083544 | 0,618651 | Thyroid disease | 82,13671 |
| ebi-a-GCST90018872 | eGFR decline (CKD population) [SH01] | Iron deficiency anemia | Wald ratio | 1 | 0,710991 | 0,409353 | 0,082411 | Minerals | 34,53822 |
| ebi-a-GCST90018877 | eGFR decline (CKD population) [SH01] | Myocardial infarction | MR Egger | 70 | -0,12373 | 0,098414 | 0,212976 | Cardiovascular disease | 66,68595 |
| ebi-a-GCST90018877 | eGFR decline (CKD population) [SH01] | Myocardial infarction | Weighted median | 70 | -0,06726 | 0,066144 | 0,309209 | Cardiovascular disease | 66,68595 |
| ebi-a-GCST90018877 | eGFR decline (CKD population) [SH01] | Myocardial infarction | Inverse variance weighted | 70 | 0,022526 | 0,043986 | 0,608561 | Cardiovascular disease | 66,68595 |
| ebi-a-GCST90018877 | eGFR decline (CKD population) [SH01] | Myocardial infarction | Simple mode | 70 | 0,020804 | 0,139052 | 0,881506 | Cardiovascular disease | 66,68595 |
| ebi-a-GCST90018877 | eGFR decline (CKD population) [SH01] | Myocardial infarction | Weighted mode | 70 | -0,06805 | 0,081809 | 0,40836 | Cardiovascular disease | 66,68595 |
| ebi-a-GCST90018916 | eGFR decline (CKD population) [SH01] | Sleep apnea syndrome | Wald ratio | 1 | -0,12816 | 0,245711 | 0,601961 | Sleep disorders | 63,50864 |
| ebi-a-GCST90018977 | eGFR decline (CKD population) [SH01] | Serum uric acid levels | MR Egger | 237 | -0,00999 | 0,158679 | 0,949863 | Uricemia | 110,1094 |
| ebi-a-GCST90018977 | eGFR decline (CKD population) [SH01] | Serum uric acid levels | Weighted median | 237 | 0,114994 | 0,149588 | 0,44205 | Uricemia | 110,1094 |
| ebi-a-GCST90018977 | eGFR decline (CKD population) [SH01] | Serum uric acid levels | Inverse variance weighted | 237 | 0,027876 | 0,08799 | 0,751389 | Uricemia | 110,1094 |
| ebi-a-GCST90018977 | eGFR decline (CKD population) [SH01] | Serum uric acid levels | Simple mode | 237 | 0,36055 | 0,350659 | 0,304905 | Uricemia | 110,1094 |
| ebi-a-GCST90018977 | eGFR decline (CKD population) [SH01] | Serum uric acid levels | Weighted mode | 237 | 0,105512 | 0,170727 | 0,537161 | Uricemia | 110,1094 |
| ebi-a-GCST90019417 | eGFR decline (CKD population) [SH01] | Vitamin K-dependent protein S measurement | MR Egger | 4 | 0,04757 | 0,160318 | 0,794658 | Vitamin | 118,3171 |
| ebi-a-GCST90019417 | eGFR decline (CKD population) [SH01] | Vitamin K-dependent protein S measurement | Weighted median | 4 | 0,009188 | 0,100173 | 0,926918 | Vitamin | 118,3171 |
| ebi-a-GCST90019417 | eGFR decline (CKD population) [SH01] | Vitamin K-dependent protein S measurement | Inverse variance weighted | 4 | 0,00404 | 0,091834 | 0,964908 | Vitamin | 118,3171 |
| ebi-a-GCST90019417 | eGFR decline (CKD population) [SH01] | Vitamin K-dependent protein S measurement | Simple mode | 4 | -0,05028 | 0,134318 | 0,733052 | Vitamin | 118,3171 |
| ebi-a-GCST90019417 | eGFR decline (CKD population) [SH01] | Vitamin K-dependent protein S measurement | Weighted mode | 4 | 0,035549 | 0,112902 | 0,773494 | Vitamin | 118,3171 |
| ebi-a-GCST90019423 | eGFR decline (CKD population) [SH01] | Vitamin K-dependent protein C measurement | MR Egger | 3 | 0,08169 | 0,234256 | 0,786392 | Vitamin | 207,8561 |
| ebi-a-GCST90019423 | eGFR decline (CKD population) [SH01] | Vitamin K-dependent protein C measurement | Weighted median | 3 | 0,069877 | 0,073086 | 0,339026 | Vitamin | 207,8561 |
| ebi-a-GCST90019423 | eGFR decline (CKD population) [SH01] | Vitamin K-dependent protein C measurement | Inverse variance weighted | 3 | 0,042372 | 0,10651 | 0,69076 | Vitamin | 207,8561 |
| ebi-a-GCST90019423 | eGFR decline (CKD population) [SH01] | Vitamin K-dependent protein C measurement | Simple mode | 3 | 0,022718 | 0,195754 | 0,918213 | Vitamin | 207,8561 |
| ebi-a-GCST90019423 | eGFR decline (CKD population) [SH01] | Vitamin K-dependent protein C measurement | Weighted mode | 3 | 0,0764 | 0,073662 | 0,408609 | Vitamin | 207,8561 |
| ebi-a-GCST90025947 | eGFR decline (CKD population) [SH01] | Serum alkaline phosphatase levels | MR Egger | 335 | -0,05882 | 0,103701 | 0,570955 | Minerals | 166,037 |
| ebi-a-GCST90025947 | eGFR decline (CKD population) [SH01] | Serum alkaline phosphatase levels | Weighted median | 335 | 0,130756 | 0,104363 | 0,210242 | Minerals | 166,037 |
| ebi-a-GCST90025947 | eGFR decline (CKD population) [SH01] | Serum alkaline phosphatase levels | Inverse variance weighted | 335 | -0,00962 | 0,060742 | 0,874139 | Minerals | 166,037 |
| ebi-a-GCST90025947 | eGFR decline (CKD population) [SH01] | Serum alkaline phosphatase levels | Simple mode | 335 | 0,162737 | 0,241094 | 0,500146 | Minerals | 166,037 |
| ebi-a-GCST90025947 | eGFR decline (CKD population) [SH01] | Serum alkaline phosphatase levels | Weighted mode | 335 | 0,109455 | 0,130523 | 0,402302 | Minerals | 166,037 |
| ebi-a-GCST90025967 | eGFR decline (CKD population) [SH01] | Vitamin D levels | MR Egger | 81 | -0,0682 | 0,193566 | 0,725527 | Vitamin | 134,1402 |
| ebi-a-GCST90025967 | eGFR decline (CKD population) [SH01] | Vitamin D levels | Weighted median | 81 | -0,23176 | 0,194493 | 0,233406 | Vitamin | 134,1402 |
| ebi-a-GCST90025967 | eGFR decline (CKD population) [SH01] | Vitamin D levels | Inverse variance weighted | 81 | -0,10762 | 0,118758 | 0,364833 | Vitamin | 134,1402 |
| ebi-a-GCST90025967 | eGFR decline (CKD population) [SH01] | Vitamin D levels | Simple mode | 81 | -0,1142 | 0,378641 | 0,763729 | Vitamin | 134,1402 |
| ebi-a-GCST90025967 | eGFR decline (CKD population) [SH01] | Vitamin D levels | Weighted mode | 81 | -0,13805 | 0,170901 | 0,421598 | Vitamin | 134,1402 |
| ebi-a-GCST90025969 | eGFR decline (CKD population) [SH01] | Hemoglobin levels | MR Egger | 325 | 0,004413 | 0,142314 | 0,975284 | Organic compound | 104,3556 |
| ebi-a-GCST90025969 | eGFR decline (CKD population) [SH01] | Hemoglobin levels | Weighted median | 325 | -0,09747 | 0,123165 | 0,42871 | Organic compound | 104,3556 |
| ebi-a-GCST90025969 | eGFR decline (CKD population) [SH01] | Hemoglobin levels | Inverse variance weighted | 325 | -0,09867 | 0,073452 | 0,179158 | Organic compound | 104,3556 |
| ebi-a-GCST90025969 | eGFR decline (CKD population) [SH01] | Hemoglobin levels | Simple mode | 325 | 0,092802 | 0,281141 | 0,741546 | Organic compound | 104,3556 |
| ebi-a-GCST90025969 | eGFR decline (CKD population) [SH01] | Hemoglobin levels | Weighted mode | 325 | -0,05376 | 0,15258 | 0,724815 | Organic compound | 104,3556 |

| | | | | | | | | | |
|---|---|---|---|---|---|---|---|---|---|
| ebi-a-GCST90025986 | eGFR decline (CKD population) [SH01] | Blood glucose levels | MR Egger | 109 | 0,186736 | 0,195588 | 0,341859 | Diabetes | 118,7554 |
| ebi-a-GCST90025986 | eGFR decline (CKD population) [SH01] | Blood glucose levels | Weighted median | 109 | 0,434939 | 0,173696 | 0,012279 | Diabetes | 118,7554 |
| ebi-a-GCST90025986 | eGFR decline (CKD population) [SH01] | Blood glucose levels | Inverse variance weighted | 109 | 0,021588 | 0,109719 | 0,844018 | Diabetes | 118,7554 |
| ebi-a-GCST90025986 | eGFR decline (CKD population) [SH01] | Blood glucose levels | Simple mode | 109 | 0,414807 | 0,401773 | 0,304172 | Diabetes | 118,7554 |
| ebi-a-GCST90025986 | eGFR decline (CKD population) [SH01] | Blood glucose levels | Weighted mode | 109 | 0,36746 | 0,168173 | 0,031049 | Diabetes | 118,7554 |
| ebi-a-GCST90025989 | eGFR decline (CKD population) [SH01] | Insulin-like growth factor 1 levels | MR Egger | 373 | -0,19295 | 0,123159 | 0,118051 | Diabetes | 115,9356 |
| ebi-a-GCST90025989 | eGFR decline (CKD population) [SH01] | Insulin-like growth factor 1 levels | Weighted median | 373 | -0,04361 | 0,110594 | 0,69332 | Diabetes | 115,9356 |
| ebi-a-GCST90025989 | eGFR decline (CKD population) [SH01] | Insulin-like growth factor 1 levels | Inverse variance weighted | 373 | 0,016615 | 0,063781 | 0,794475 | Diabetes | 115,9356 |
| ebi-a-GCST90025989 | eGFR decline (CKD population) [SH01] | Insulin-like growth factor 1 levels | Simple mode | 373 | -0,11826 | 0,244579 | 0,628999 | Diabetes | 115,9356 |
| ebi-a-GCST90025989 | eGFR decline (CKD population) [SH01] | Insulin-like growth factor 1 levels | Weighted mode | 373 | -0,15013 | 0,142643 | 0,293262 | Diabetes | 115,9356 |
| ebi-a-GCST90025990 | eGFR decline (CKD population) [SH01] | Calcium levels | MR Egger | 202 | 0,316142 | 0,155371 | 0,043196 | Minerals | 102,0722 |
| ebi-a-GCST90025990 | eGFR decline (CKD population) [SH01] | Calcium levels | Weighted median | 202 | 0,140813 | 0,177997 | 0,428888 | Minerals | 102,0722 |
| ebi-a-GCST90025990 | eGFR decline (CKD population) [SH01] | Calcium levels | Inverse variance weighted | 202 | 0,070865 | 0,084215 | 0,400083 | Minerals | 102,0722 |
| ebi-a-GCST90025990 | eGFR decline (CKD population) [SH01] | Calcium levels | Simple mode | 202 | 0,040378 | 0,345428 | 0,907062 | Minerals | 102,0722 |
| ebi-a-GCST90025990 | eGFR decline (CKD population) [SH01] | Calcium levels | Weighted mode | 202 | 0,148926 | 0,182714 | 0,415994 | Minerals | 102,0722 |
| ebi-a-GCST90029013 | eGFR decline (CKD population) [SH01] | Educational attainment (years of education) | MR Egger | 209 | 0,034884 | 0,117002 | 0,765889 | Social trait | 45,05626 |
| ebi-a-GCST90029013 | eGFR decline (CKD population) [SH01] | Educational attainment (years of education) | Weighted median | 209 | -0,00223 | 0,040451 | 0,955944 | Social trait | 45,05626 |
| ebi-a-GCST90029013 | eGFR decline (CKD population) [SH01] | Educational attainment (years of education) | Inverse variance weighted | 209 | -0,03435 | 0,026771 | 0,199523 | Social trait | 45,05626 |
| ebi-a-GCST90029013 | eGFR decline (CKD population) [SH01] | Educational attainment (years of education) | Simple mode | 209 | 0,042997 | 0,123934 | 0,728992 | Social trait | 45,05626 |
| ebi-a-GCST90029013 | eGFR decline (CKD population) [SH01] | Educational attainment (years of education) | Weighted mode | 209 | 0,059274 | 0,10133 | 0,559207 | Social trait | 45,05626 |
| ebi-a-GCST90029014 | eGFR decline (CKD population) [SH01] | Smoking status | MR Egger | 117 | -0,43839 | 1,116747 | 0,695372 | Smoking | 42,91926 |
| ebi-a-GCST90029014 | eGFR decline (CKD population) [SH01] | Smoking status | Weighted median | 117 | 0,037122 | 0,367983 | 0,919645 | Smoking | 42,91926 |
| ebi-a-GCST90029014 | eGFR decline (CKD population) [SH01] | Smoking status | Inverse variance weighted | 117 | -0,02846 | 0,264941 | 0,914463 | Smoking | 42,91926 |
| ebi-a-GCST90029014 | eGFR decline (CKD population) [SH01] | Smoking status | Simple mode | 117 | -0,58808 | 0,92202 | 0,524852 | Smoking | 42,91926 |
| ebi-a-GCST90029014 | eGFR decline (CKD population) [SH01] | Smoking status | Weighted mode | 117 | -0,2944 | 0,777708 | 0,705713 | Smoking | 42,91926 |
| ebi-a-GCST90029070 | eGFR decline (CKD population) [SH01] | C-reactive protein levels | MR Egger | 214 | 0,290229 | 0,132562 | 0,029661 | Inflammation | 122,6906 |
| ebi-a-GCST90029070 | eGFR decline (CKD population) [SH01] | C-reactive protein levels | Weighted median | 214 | 0,125493 | 0,127835 | 0,326256 | Inflammation | 122,6906 |
| ebi-a-GCST90029070 | eGFR decline (CKD population) [SH01] | C-reactive protein levels | Inverse variance weighted | 214 | 0,103448 | 0,078269 | 0,18627 | Inflammation | 122,6906 |
| ebi-a-GCST90029070 | eGFR decline (CKD population) [SH01] | C-reactive protein levels | Simple mode | 214 | -0,40705 | 0,304115 | 0,182167 | Inflammation | 122,6906 |
| ebi-a-GCST90029070 | eGFR decline (CKD population) [SH01] | C-reactive protein levels | Weighted mode | 214 | 0,227096 | 0,137773 | 0,100758 | Inflammation | 122,6906 |
| ebi-a-GCST90029073 | eGFR decline (CKD population) [SH01] | Liver fat | MR Egger | 8 | 0,28276 | 0,16279 | 0,133068 | Fatty liver | 140,8023 |
| ebi-a-GCST90029073 | eGFR decline (CKD population) [SH01] | Liver fat | Weighted median | 8 | 0,20489 | 0,121435 | 0,091556 | Fatty liver | 140,8023 |
| ebi-a-GCST90029073 | eGFR decline (CKD population) [SH01] | Liver fat | Inverse variance weighted | 8 | 0,182 | 0,098867 | 0,065641 | Fatty liver | 140,8023 |
| ebi-a-GCST90029073 | eGFR decline (CKD population) [SH01] | Liver fat | Simple mode | 8 | 0,231794 | 0,189745 | 0,261397 | Fatty liver | 140,8023 |
| ebi-a-GCST90029073 | eGFR decline (CKD population) [SH01] | Liver fat | Weighted mode | 8 | 0,223431 | 0,120781 | 0,106785 | Fatty liver | 140,8023 |
| ebi-a-GCST90086057 | eGFR decline (CKD population) [SH01] | Cardiovascular disease | Inverse variance weighted | 2 | 0,100644 | 0,214866 | 0,639496 | Cardiovascular disease | 32,018 |
| finn-b-D3_ANAEMIA_B12_DEF | eGFR decline (CKD population) [SH01] | Vitamin B12 deficiency anaemia | MR Egger | 4 | -0,08002 | 0,158056 | 0,662945 | Vitamin | 52,61875 |
| finn-b-D3_ANAEMIA_B12_DEF | eGFR decline (CKD population) [SH01] | Vitamin B12 deficiency anaemia | Weighted median | 4 | -0,06064 | 0,064329 | 0,345867 | Vitamin | 52,61875 |
| finn-b-D3_ANAEMIA_B12_DEF | eGFR decline (CKD population) [SH01] | Vitamin B12 deficiency anaemia | Inverse variance weighted | 4 | -0,03333 | 0,055191 | 0,545905 | Vitamin | 52,61875 |
| finn-b-D3_ANAEMIA_B12_DEF | eGFR decline (CKD population) [SH01] | Vitamin B12 deficiency anaemia | Simple mode | 4 | -0,07333 | 0,09507 | 0,496686 | Vitamin | 52,61875 |
| finn-b-D3_ANAEMIA_B12_DEF | eGFR decline (CKD population) [SH01] | Vitamin B12 deficiency anaemia | Weighted mode | 4 | -0,07645 | 0,071617 | 0,36401 | Vitamin | 52,61875 |
| finn-b-E4_OBESITY | eGFR decline (CKD population) [SH01] | Obesity | MR Egger | 7 | -0,09173 | 0,29805 | 0,77066 | Obesity | 59,53804 |
| finn-b-E4_OBESITY | eGFR decline (CKD population) [SH01] | Obesity | Weighted median | 7 | -0,02282 | 0,10046 | 0,820335 | Obesity | 59,53804 |
| finn-b-E4_OBESITY | eGFR decline (CKD population) [SH01] | Obesity | Inverse variance weighted | 7 | 0,027047 | 0,082887 | 0,744194 | Obesity | 59,53804 |
| finn-b-E4_OBESITY | eGFR decline (CKD population) [SH01] | Obesity | Simple mode | 7 | 0,058446 | 0,17554 | 0,750499 | Obesity | 59,53804 |
| finn-b-E4_OBESITY | eGFR decline (CKD population) [SH01] | Obesity | Weighted mode | 7 | -0,07929 | 0,10357 | 0,472966 | Obesity | 59,53804 |
| finn-b-FG_PULMHEART | eGFR decline (CKD population) [SH01] | Pulmonary heart disease | MR Egger | 5 | -0,24185 | 0,156145 | 0,219167 | Cardiovascular disease | 89,00197 |
| finn-b-FG_PULMHEART | eGFR decline (CKD population) [SH01] | Pulmonary heart disease | Weighted median | 5 | 0,034173 | 0,067442 | 0,612364 | Cardiovascular disease | 89,00197 |
| finn-b-FG_PULMHEART | eGFR decline (CKD population) [SH01] | Pulmonary heart disease | Inverse variance weighted | 5 | 0,011969 | 0,059472 | 0,840496 | Cardiovascular disease | 89,00197 |
| finn-b-FG_PULMHEART | eGFR decline (CKD population) [SH01] | Pulmonary heart disease | Simple mode | 5 | 0,115396 | 0,110659 | 0,355914 | Cardiovascular disease | 89,00197 |
| finn-b-FG_PULMHEART | eGFR decline (CKD population) [SH01] | Pulmonary heart disease | Weighted mode | 5 | 0,11266 | 0,113888 | 0,378554 | Cardiovascular disease | 89,00197 |
| ieu-b-102 | eGFR decline (CKD population) [SH01] | Major depression | MR Egger | 48 | 0,360597 | 0,825318 | 0,664214 | Mental health | 38,29386 |
| ieu-b-102 | eGFR decline (CKD population) [SH01] | Major depression | Weighted median | 48 | 0,011057 | 0,20253 | 0,956461 | Mental health | 38,29386 |
| ieu-b-102 | eGFR decline (CKD population) [SH01] | Major depression | Inverse variance weighted | 48 | -0,09998 | 0,148872 | 0,50187 | Mental health | 38,29386 |
| ieu-b-102 | eGFR decline (CKD population) [SH01] | Major depression | Simple mode | 48 | 0,252734 | 0,489814 | 0,608288 | Mental health | 38,29386 |
| ieu-b-102 | eGFR decline (CKD population) [SH01] | Major depression | Weighted mode | 48 | 0,15015 | 0,4304 | 0,728752 | Mental health | 38,29386 |
| ieu-b-107 | eGFR decline (CKD population) [SH01] | apolipoprotein A-I | MR Egger | 269 | -0,11042 | 0,115534 | 0,340053 | Dyslipidemias | 139,7111 |
| ieu-b-107 | eGFR decline (CKD population) [SH01] | apolipoprotein A-I | Weighted median | 269 | -0,08318 | 0,111167 | 0,454338 | Dyslipidemias | 139,7111 |
| ieu-b-107 | eGFR decline (CKD population) [SH01] | apolipoprotein A-I | Inverse variance weighted | 269 | -0,03493 | 0,068283 | 0,608921 | Dyslipidemias | 139,7111 |
| ieu-b-107 | eGFR decline (CKD population) [SH01] | apolipoprotein A-I | Simple mode | 269 | -0,16074 | 0,249388 | 0,519765 | Dyslipidemias | 139,7111 |
| ieu-b-107 | eGFR decline (CKD population) [SH01] | apolipoprotein A-I | Weighted mode | 269 | -0,10076 | 0,12843 | 0,433405 | Dyslipidemias | 139,7111 |
| ieu-b-108 | eGFR decline (CKD population) [SH01] | apolipoprotein B | MR Egger | 173 | 0,073617 | 0,125464 | 0,558142 | Dyslipidemias | 162,3296 |
| ieu-b-108 | eGFR decline (CKD population) [SH01] | apolipoprotein B | Weighted median | 173 | 0,134609 | 0,120473 | 0,263848 | Dyslipidemias | 162,3296 |
| ieu-b-108 | eGFR decline (CKD population) [SH01] | apolipoprotein B | Inverse variance weighted | 173 | 0,099999 | 0,077192 | 0,195161 | Dyslipidemias | 162,3296 |
| ieu-b-108 | eGFR decline (CKD population) [SH01] | apolipoprotein B | Simple mode | 173 | 0,137125 | 0,239355 | 0,567464 | Dyslipidemias | 162,3296 |
| ieu-b-108 | eGFR decline (CKD population) [SH01] | apolipoprotein B | Weighted mode | 173 | 0,161247 | 0,131855 | 0,223036 | Dyslipidemias | 162,3296 |
| ieu-b-25 | eGFR decline (CKD population) [SH01] | Cigarettes per Day | MR Egger | 22 | -0,25493 | 0,176156 | 0,163336 | Smoking | 99,80012 |
| ieu-b-25 | eGFR decline (CKD population) [SH01] | Cigarettes per Day | Weighted median | 22 | -0,19769 | 0,136564 | 0,147725 | Smoking | 99,80012 |
| ieu-b-25 | eGFR decline (CKD population) [SH01] | Cigarettes per Day | Inverse variance weighted | 22 | -0,16926 | 0,101361 | 0,094938 | Smoking | 99,80012 |
| ieu-b-25 | eGFR decline (CKD population) [SH01] | Cigarettes per Day | Simple mode | 22 | -0,52195 | 0,288851 | 0,085116 | Smoking | 99,80012 |
| ieu-b-25 | eGFR decline (CKD population) [SH01] | Cigarettes per Day | Weighted mode | 22 | -0,18353 | 0,142097 | 0,210535 | Smoking | 99,80012 |
| ieu-b-4861 | eGFR decline (CKD population) [SH01] | Chronotype | MR Egger | 60 | -0,46065 | 0,381176 | 0,231763 | Sleep disorders | 44,49296 |
| ieu-b-4861 | eGFR decline (CKD population) [SH01] | Chronotype | Weighted median | 60 | -0,08697 | 0,206097 | 0,673047 | Sleep disorders | 44,49296 |
| ieu-b-4861 | eGFR decline (CKD population) [SH01] | Chronotype | Inverse variance weighted | 60 | -0,01145 | 0,138413 | 0,93409 | Sleep disorders | 44,49296 |
| ieu-b-4861 | eGFR decline (CKD population) [SH01] | Chronotype | Simple mode | 60 | 0,022545 | 0,466133 | 0,961588 | Sleep disorders | 44,49296 |
| ieu-b-4861 | eGFR decline (CKD population) [SH01] | Chronotype | Weighted mode | 60 | -0,09898 | 0,452006 | 0,827427 | Sleep disorders | 44,49296 |
| ieu-b-4877 | eGFR decline (CKD population) [SH01] | smoking initiation | MR Egger | 85 | 1,581481 | 0,611491 | 0,011447 | Smoking | 42,2587 |
| ieu-b-4877 | eGFR decline (CKD population) [SH01] | smoking initiation | Weighted median | 85 | 0,171812 | 0,176137 | 0,329339 | Smoking | 42,2587 |
| ieu-b-4877 | eGFR decline (CKD population) [SH01] | smoking initiation | Inverse variance weighted | 85 | 0,17917 | 0,120377 | 0,136643 | Smoking | 42,2587 |
| ieu-b-4877 | eGFR decline (CKD population) [SH01] | smoking initiation | Simple mode | 85 | 0,291869 | 0,397498 | 0,464832 | Smoking | 42,2587 |
| ieu-b-4877 | eGFR decline (CKD population) [SH01] | smoking initiation | Weighted mode | 85 | 0,238576 | 0,37439 | 0,525702 | Smoking | 42,2587 |
| ieu-b-5144 | eGFR decline (CKD population) [SH01] | Hypertension | MR Egger | 187 | -1,60782 | 0,900963 | 0,075971 | Hypertension | 54,118 |
| ieu-b-5144 | eGFR decline (CKD population) [SH01] | Hypertension | Weighted median | 187 | -0,89257 | 0,435579 | 0,040446 | Hypertension | 54,118 |
| ieu-b-5144 | eGFR decline (CKD population) [SH01] | Hypertension | Inverse variance weighted | 187 | -0,64418 | 0,291192 | 0,02695 | Hypertension | 54,118 |
| ieu-b-5144 | eGFR decline (CKD population) [SH01] | Hypertension | Simple mode | 187 | -1,40799 | 1,209905 | 0,246029 | Hypertension | 54,118 |
| ieu-b-5144 | eGFR decline (CKD population) [SH01] | Hypertension | Weighted mode | 187 | -1,628 | 0,964035 | 0,092946 | Hypertension | 54,118 |
| ieu-b-72 | eGFR decline (CKD population) [SH01] | Urinary sodium-potassium ratio | MR Egger | 24 | 0,811983 | 1,637963 | 0,625003 | Minerals | 39,88328 |
| ieu-b-72 | eGFR decline (CKD population) [SH01] | Urinary sodium-potassium ratio | Weighted median | 24 | 0,190436 | 0,52103 | 0,714739 | Minerals | 39,88328 |
| ieu-b-72 | eGFR decline (CKD population) [SH01] | Urinary sodium-potassium ratio | Inverse variance weighted | 24 | 0,411221 | 0,378347 | 0,277086 | Minerals | 39,88328 |
| ieu-b-72 | eGFR decline (CKD population) [SH01] | Urinary sodium-potassium ratio | Simple mode | 24 | -0,19537 | 1,016826 | 0,849318 | Minerals | 39,88328 |
| ieu-b-72 | eGFR decline (CKD population) [SH01] | Urinary sodium-potassium ratio | Weighted mode | 24 | -0,22198 | 0,984203 | 0,823552 | Minerals | 39,88328 |
| prot-a-3102 | eGFR decline (CKD population) [SH01] | Thyrotropin-releasing hormone | Inverse variance weighted | 2 | 0,137458 | 0,13752 | 0,317528 | Thyroid disease | 36,35446 |
| prot-c-2652_15_1 | eGFR decline (CKD population) [SH01] | suPAR | Wald ratio | 1 | 0,040156 | 0,106463 | 0,706035 | Biomarkers | 33,5125 |
| ukb-b-4424 | eGFR decline (CKD population) [SH01] | Sleep duration | MR Egger | 66 | 0,118781 | 1,165573 | 0,919149 | Sleep disorders | 42,80283 |
| ukb-b-4424 | eGFR decline (CKD population) [SH01] | Sleep duration | Weighted median | 66 | -0,09165 | 0,462212 | 0,842822 | Sleep disorders | 42,80283 |
| ukb-b-4424 | eGFR decline (CKD population) [SH01] | Sleep duration | Inverse variance weighted | 66 | -0,16335 | 0,305189 | 0,592475 | Sleep disorders | 42,80283 |
| ukb-b-4424 | eGFR decline (CKD population) [SH01] | Sleep duration | Simple mode | 66 | -0,42897 | 0,969793 | 0,65972 | Sleep disorders | 42,80283 |
| ukb-b-4424 | eGFR decline (CKD population) [SH01] | Sleep duration | Weighted mode | 66 | -0,29094 | 0,79755 | 0,716451 | Sleep disorders | 42,80283 |
| ukb-b-5779 | eGFR decline (CKD population) [SH01] | Alcohol intake frequency. | MR Egger | 94 | -0,04049 | 0,274255 | 0,882956 | Alcohol consumption | 53,13815 |
| ukb-b-5779 | eGFR decline (CKD population) [SH01] | Alcohol intake frequency. | Weighted median | 94 | 0,011643 | 0,233589 | 0,960248 | Alcohol consumption | 53,13815 |
| ukb-b-5779 | eGFR decline (CKD population) [SH01] | Alcohol intake frequency. | Inverse variance weighted | 94 | -0,02711 | 0,121747 | 0,823799 | Alcohol consumption | 53,13815 |
| ukb-b-5779 | eGFR decline (CKD population) [SH01] | Alcohol intake frequency. | Simple mode | 94 | -0,40477 | 0,447725 | 0,368304 | Alcohol consumption | 53,13815 |
| ukb-b-5779 | eGFR decline (CKD population) [SH01] | Alcohol intake frequency. | Weighted mode | 94 | -0,08304 | 0,277222 | 0,765204 | Alcohol consumption | 53,13815 |
| ukb-b-7864 | eGFR decline (CKD population) [SH01] | Vitamin B6 | Wald ratio | 1 | -0,87998 | 0,860305 | 0,306369 | Vitamin | 32,75897 |
| ukb-d-30790_irnt | eGFR decline (CKD population) [SH01] | Lipoprotein A | MR Egger | 11 | -0,07878 | 0,183864 | 0,678366 | Dyslipidemias | 985,6435 |
| ukb-d-30790_irnt | eGFR decline (CKD population) [SH01] | Lipoprotein A | Weighted median | 11 | -0,17527 | 0,131606 | 0,18294 | Dyslipidemias | 985,6435 |
| ukb-d-30790_irnt | eGFR decline (CKD population) [SH01] | Lipoprotein A | Inverse variance weighted | 11 | -0,06345 | 0,118764 | 0,59319 | Dyslipidemias | 985,6435 |
| ukb-d-30790_irnt | eGFR decline (CKD population) [SH01] | Lipoprotein A | Simple mode | 11 | -0,25592 | 0,199919 | 0,229404 | Dyslipidemias | 985,6435 |
| ukb-d-30790_irnt | eGFR decline (CKD population) [SH01] | Lipoprotein A | Weighted mode | 11 | -0,1935 | 0,155538 | 0,241836 | Dyslipidemias | 985,6435 |
| BMI | eGFR decline (CKD population) [SH02] | Body mass index | MR Egger | 506 | 0,077118 | 0,205175 | 0,707174 | Obesity | 74,21321 |
| BMI | eGFR decline (CKD population) [SH02] | Body mass index | Weighted median | 506 | 0,028708 | 0,13178 | 0,827549 | Obesity | 74,21321 |
| BMI | eGFR decline (CKD population) [SH02] | Body mass index | Inverse variance weighted | 506 | 0,060272 | 0,07883 | 0,444526 | Obesity | 74,21321 |
| BMI | eGFR decline (CKD population) [SH02] | Body mass index | Simple mode | 506 | -0,17513 | 0,431449 | 0,684981 | Obesity | 74,21321 |

| | | | | | | | | | |
|---|---|---|---|---|---|---|---|---|---|
| BMI | eGFR decline (CKD population) [SH02] | Body mass index | Weighted mode | 506 | 0,068284 | 0,2441 | 0,779794 | Obesity | 74,21321 |
| DBPTang | eGFR decline (CKD population) [SH02] | Diastolic blood pressure | MR Egger | 251 | -0,11563 | 0,341223 | 0,734989 | Hypertension | 55,91029 |
| DBPTang | eGFR decline (CKD population) [SH02] | Diastolic blood pressure | Weighted median | 251 | -0,10768 | 0,159274 | 0,498995 | Hypertension | 55,91029 |
| DBPTang | eGFR decline (CKD population) [SH02] | Diastolic blood pressure | Inverse variance weighted | 251 | -0,07887 | 0,106131 | 0,45738 | Hypertension | 55,91029 |
| DBPTang | eGFR decline (CKD population) [SH02] | Diastolic blood pressure | Simple mode | 251 | 0,153876 | 0,467089 | 0,742101 | Hypertension | 55,91029 |
| DBPTang | eGFR decline (CKD population) [SH02] | Diastolic blood pressure | Weighted mode | 251 | -0,11723 | 0,359475 | 0,744608 | Hypertension | 55,91029 |
| FT3 | eGFR decline (CKD population) [SH02] | Free triiodothyronine | MR Egger | 6 | -0,44598 | 0,889122 | 0,642303 | Thyroid disease | 82,22035 |
| FT3 | eGFR decline (CKD population) [SH02] | Free triiodothyronine | Weighted median | 6 | -0,18523 | 0,249219 | 0,457331 | Thyroid disease | 82,22035 |
| FT3 | eGFR decline (CKD population) [SH02] | Free triiodothyronine | Inverse variance weighted | 6 | -0,14082 | 0,196082 | 0,47266 | Thyroid disease | 82,22035 |
| FT3 | eGFR decline (CKD population) [SH02] | Free triiodothyronine | Simple mode | 6 | -0,17581 | 0,342779 | 0,629873 | Thyroid disease | 82,22035 |
| FT3 | eGFR decline (CKD population) [SH02] | Free triiodothyronine | Weighted mode | 6 | -0,13399 | 0,31763 | 0,69067 | Thyroid disease | 82,22035 |
| FT3FT4 | eGFR decline (CKD population) [SH02] | FT3 FT4 ratio | MR Egger | 13 | 0,358204 | 1,120538 | 0,755207 | Thyroid disease | 132,3008 |
| FT3FT4 | eGFR decline (CKD population) [SH02] | FT3 FT4 ratio | Weighted median | 13 | -0,2188 | 0,735283 | 0,766026 | Thyroid disease | 132,3008 |
| FT3FT4 | eGFR decline (CKD population) [SH02] | FT3 FT4 ratio | Inverse variance weighted | 13 | -0,77748 | 0,55754 | 0,163174 | Thyroid disease | 132,3008 |
| FT3FT4 | eGFR decline (CKD population) [SH02] | FT3 FT4 ratio | Simple mode | 13 | 0,137842 | 1,312371 | 0,918085 | Thyroid disease | 132,3008 |
| FT3FT4 | eGFR decline (CKD population) [SH02] | FT3 FT4 ratio | Weighted mode | 13 | -0,25297 | 0,769103 | 0,747884 | Thyroid disease | 132,3008 |
| FT4 | eGFR decline (CKD population) [SH02] | Free thyroxine | MR Egger | 62 | -0,06551 | 0,174937 | 0,70939 | Thyroid disease | 84,3797 |
| FT4 | eGFR decline (CKD population) [SH02] | Free thyroxine | Weighted median | 62 | 0,010127 | 0,143041 | 0,943559 | Thyroid disease | 84,3797 |
| FT4 | eGFR decline (CKD population) [SH02] | Free thyroxine | Inverse variance weighted | 62 | -0,02018 | 0,083572 | 0,809219 | Thyroid disease | 84,3797 |
| FT4 | eGFR decline (CKD population) [SH02] | Free thyroxine | Simple mode | 62 | -0,08864 | 0,252072 | 0,726323 | Thyroid disease | 84,3797 |
| FT4 | eGFR decline (CKD population) [SH02] | Free thyroxine | Weighted mode | 62 | -0,0113 | 0,150361 | 0,940322 | Thyroid disease | 84,3797 |
| Fe | eGFR decline (CKD population) [SH02] | Serum iron | Inverse variance weighted | 2 | -0,2321 | 0,621053 | 0,708611 | Minerals | 291,9629 |
| HDL | eGFR decline (CKD population) [SH02] | HDL cholesterol | MR Egger | 233 | 0,015143 | 0,103008 | 0,883254 | Dyslipidemias | 268,9835 |
| HDL | eGFR decline (CKD population) [SH02] | HDL cholesterol | Weighted median | 233 | -0,03862 | 0,108653 | 0,722227 | Dyslipidemias | 268,9835 |
| HDL | eGFR decline (CKD population) [SH02] | HDL cholesterol | Inverse variance weighted | 233 | -0,08541 | 0,0664 | 0,198359 | Dyslipidemias | 268,9835 |
| HDL | eGFR decline (CKD population) [SH02] | HDL cholesterol | Simple mode | 233 | -0,13379 | 0,206313 | 0,517323 | Dyslipidemias | 268,9835 |
| HDL | eGFR decline (CKD population) [SH02] | HDL cholesterol | Weighted mode | 233 | -0,01643 | 0,106085 | 0,877043 | Dyslipidemias | 268,9835 |
| HiTSH | eGFR decline (CKD population) [SH02] | High TSH | MR Egger | 6 | 0,641541 | 0,583926 | 0,333599 | Thyroid disease | 49,53497 |
| HiTSH | eGFR decline (CKD population) [SH02] | High TSH | Weighted median | 6 | 0,30252 | 0,167669 | 0,071189 | Thyroid disease | 49,53497 |
| HiTSH | eGFR decline (CKD population) [SH02] | High TSH | Inverse variance weighted | 6 | 0,301583 | 0,132555 | 0,022897 | Thyroid disease | 49,53497 |
| HiTSH | eGFR decline (CKD population) [SH02] | High TSH | Simple mode | 6 | 0,328591 | 0,245225 | 0,237928 | Thyroid disease | 49,53497 |
| HiTSH | eGFR decline (CKD population) [SH02] | High TSH | Weighted mode | 6 | 0,314589 | 0,204326 | 0,184271 | Thyroid disease | 49,53497 |
| Insom | eGFR decline (CKD population) [SH02] | Insomnia | MR Egger | 13 | 0,019466 | 0,755305 | 0,9799 | Sleep disorders | 44,05724 |
| Insom | eGFR decline (CKD population) [SH02] | Insomnia | Weighted median | 13 | -0,21581 | 0,30258 | 0,475694 | Sleep disorders | 44,05724 |
| Insom | eGFR decline (CKD population) [SH02] | Insomnia | Inverse variance weighted | 13 | 0,077041 | 0,230564 | 0,738272 | Sleep disorders | 44,05724 |
| Insom | eGFR decline (CKD population) [SH02] | Insomnia | Simple mode | 13 | -0,37111 | 0,466583 | 0,441839 | Sleep disorders | 44,05724 |
| Insom | eGFR decline (CKD population) [SH02] | Insomnia | Weighted mode | 13 | -0,28663 | 0,416666 | 0,50459 | Sleep disorders | 44,05724 |
| LDL | eGFR decline (CKD population) [SH02] | LDL cholesterol | MR Egger | 217 | -0,07138 | 0,116801 | 0,541777 | Dyslipidemias | 221,0222 |
| LDL | eGFR decline (CKD population) [SH02] | LDL cholesterol | Weighted median | 217 | -0,0907 | 0,116845 | 0,437603 | Dyslipidemias | 221,0222 |
| LDL | eGFR decline (CKD population) [SH02] | LDL cholesterol | Inverse variance weighted | 217 | -0,07126 | 0,072852 | 0,327996 | Dyslipidemias | 221,0222 |
| LDL | eGFR decline (CKD population) [SH02] | LDL cholesterol | Simple mode | 217 | -0,20197 | 0,227341 | 0,375309 | Dyslipidemias | 221,0222 |
| LDL | eGFR decline (CKD population) [SH02] | LDL cholesterol | Weighted mode | 217 | -0,12116 | 0,110893 | 0,275789 | Dyslipidemias | 221,0222 |
| LoTSH | eGFR decline (CKD population) [SH02] | Low TSH | MR Egger | 26 | -0,00402 | 0,08185 | 0,961199 | Thyroid disease | 70,97091 |
| LoTSH | eGFR decline (CKD population) [SH02] | Low TSH | Weighted median | 26 | 0,021893 | 0,039384 | 0,578286 | Thyroid disease | 70,97091 |
| LoTSH | eGFR decline (CKD population) [SH02] | Low TSH | Inverse variance weighted | 26 | 0,023145 | 0,02792 | 0,407113 | Thyroid disease | 70,97091 |
| LoTSH | eGFR decline (CKD population) [SH02] | Low TSH | Simple mode | 26 | 0,051649 | 0,080607 | 0,527515 | Thyroid disease | 70,97091 |
| LoTSH | eGFR decline (CKD population) [SH02] | Low TSH | Weighted mode | 26 | -0,01483 | 0,067801 | 0,828651 | Thyroid disease | 70,97091 |
| PPTang | eGFR decline (CKD population) [SH02] | Pulse pressure | MR Egger | 300 | 0,111751 | 0,25466 | 0,661108 | Hypertension | 68,57434 |
| PPTang | eGFR decline (CKD population) [SH02] | Pulse pressure | Weighted median | 300 | 0,025359 | 0,153824 | 0,869056 | Hypertension | 68,57434 |
| PPTang | eGFR decline (CKD population) [SH02] | Pulse pressure | Inverse variance weighted | 300 | 0,118961 | 0,094782 | 0,209441 | Hypertension | 68,57434 |
| PPTang | eGFR decline (CKD population) [SH02] | Pulse pressure | Simple mode | 300 | -0,26811 | 0,425269 | 0,528886 | Hypertension | 68,57434 |
| PPTang | eGFR decline (CKD population) [SH02] | Pulse pressure | Weighted mode | 300 | -0,06936 | 0,294269 | 0,813822 | Hypertension | 68,57434 |
| SBPTang | eGFR decline (CKD population) [SH02] | Systolic blood pressure | MR Egger | 251 | -0,24958 | 0,339024 | 0,462322 | Hypertension | 56,96232 |
| SBPTang | eGFR decline (CKD population) [SH02] | Systolic blood pressure | Weighted median | 251 | -0,05981 | 0,16426 | 0,715785 | Hypertension | 56,96232 |
| SBPTang | eGFR decline (CKD population) [SH02] | Systolic blood pressure | Inverse variance weighted | 251 | -0,06791 | 0,109023 | 0,533352 | Hypertension | 56,96232 |
| SBPTang | eGFR decline (CKD population) [SH02] | Systolic blood pressure | Simple mode | 251 | 0,003914 | 0,45923 | 0,993207 | Hypertension | 56,96232 |
| SBPTang | eGFR decline (CKD population) [SH02] | Systolic blood pressure | Weighted mode | 251 | -0,05587 | 0,330289 | 0,865809 | Hypertension | 56,96232 |
| SIndex | eGFR decline (CKD population) [SH02] | Compound smoking index | MR Egger | 121 | 0,278247 | 1,019597 | 0,785404 | Smoking | 44,08236 |
| SIndex | eGFR decline (CKD population) [SH02] | Compound smoking index | Weighted median | 121 | -0,05681 | 0,37005 | 0,877996 | Smoking | 44,08236 |
| SIndex | eGFR decline (CKD population) [SH02] | Compound smoking index | Inverse variance weighted | 121 | -0,25786 | 0,254129 | 0,310265 | Smoking | 44,08236 |
| SIndex | eGFR decline (CKD population) [SH02] | Compound smoking index | Simple mode | 121 | -0,39173 | 0,978183 | 0,689525 | Smoking | 44,08236 |
| SIndex | eGFR decline (CKD population) [SH02] | Compound smoking index | Weighted mode | 121 | -0,08578 | 0,864835 | 0,921157 | Smoking | 44,08236 |
| T2DGGI | eGFR decline (CKD population) [SH02] | Type 2 diabetes | MR Egger | 441 | 0,118792 | 0,071043 | 0,095214 | Diabetes | 93,04757 |
| T2DGGI | eGFR decline (CKD population) [SH02] | Type 2 diabetes | Weighted median | 441 | 0,107496 | 0,067629 | 0,11195 | Diabetes | 93,04757 |
| T2DGGI | eGFR decline (CKD population) [SH02] | Type 2 diabetes | Inverse variance weighted | 441 | 0,080616 | 0,034303 | 0,018766 | Diabetes | 93,04757 |
| T2DGGI | eGFR decline (CKD population) [SH02] | Type 2 diabetes | Simple mode | 441 | 0,110093 | 0,166848 | 0,509702 | Diabetes | 93,04757 |
| T2DGGI | eGFR decline (CKD population) [SH02] | Type 2 diabetes | Weighted mode | 441 | 0,110093 | 0,08174 | 0,17872 | Diabetes | 93,04757 |
| TC | eGFR decline (CKD population) [SH02] | Total cholesterol | MR Egger | 264 | -0,06179 | 0,103759 | 0,552026 | Dyslipidemias | 246,1189 |
| TC | eGFR decline (CKD population) [SH02] | Total cholesterol | Weighted median | 264 | 0,040734 | 0,110704 | 0,71291 | Dyslipidemias | 246,1189 |
| TC | eGFR decline (CKD population) [SH02] | Total cholesterol | Inverse variance weighted | 264 | -0,004 | 0,06603 | 0,951721 | Dyslipidemias | 246,1189 |
| TC | eGFR decline (CKD population) [SH02] | Total cholesterol | Simple mode | 264 | -0,02091 | 0,232749 | 0,928482 | Dyslipidemias | 246,1189 |
| TC | eGFR decline (CKD population) [SH02] | Total cholesterol | Weighted mode | 264 | -0,02091 | 0,109001 | 0,848018 | Dyslipidemias | 246,1189 |
| TG | eGFR decline (CKD population) [SH02] | Triglyceride | MR Egger | 219 | 0,133228 | 0,108222 | 0,21963 | Dyslipidemias | 231,1265 |
| TG | eGFR decline (CKD population) [SH02] | Triglyceride | Weighted median | 219 | 0,083447 | 0,115439 | 0,469761 | Dyslipidemias | 231,1265 |
| TG | eGFR decline (CKD population) [SH02] | Triglyceride | Inverse variance weighted | 219 | 0,044648 | 0,069823 | 0,522537 | Dyslipidemias | 231,1265 |
| TG | eGFR decline (CKD population) [SH02] | Triglyceride | Simple mode | 219 | -0,07715 | 0,242069 | 0,750262 | Dyslipidemias | 231,1265 |
| TG | eGFR decline (CKD population) [SH02] | Triglyceride | Weighted mode | 219 | 0,104404 | 0,102645 | 0,310216 | Dyslipidemias | 231,1265 |
| TSH | eGFR decline (CKD population) [SH02] | Thyroid Stimulating Hormone | MR Egger | 159 | 0,130944 | 0,100471 | 0,194381 | Thyroid disease | 154,4324 |
| TSH | eGFR decline (CKD population) [SH02] | Thyroid Stimulating Hormone | Weighted median | 159 | 0,100038 | 0,089106 | 0,261567 | Thyroid disease | 154,4324 |
| TSH | eGFR decline (CKD population) [SH02] | Thyroid Stimulating Hormone | Inverse variance weighted | 159 | 0,014716 | 0,055853 | 0,792175 | Thyroid disease | 154,4324 |
| TSH | eGFR decline (CKD population) [SH02] | Thyroid Stimulating Hormone | Simple mode | 159 | -0,01249 | 0,19972 | 0,950231 | Thyroid disease | 154,4324 |
| TSH | eGFR decline (CKD population) [SH02] | Thyroid Stimulating Hormone | Weighted mode | 159 | 0,160566 | 0,095858 | 0,095902 | Thyroid disease | 154,4324 |
| WHR | eGFR decline (CKD population) [SH02] | Waist-to-hip ratio adjusted BMI | MR Egger | 296 | -0,17683 | 0,217654 | 0,417208 | Obesity | 85,86852 |
| WHR | eGFR decline (CKD population) [SH02] | Waist-to-hip ratio adjusted BMI | Weighted median | 296 | -0,00099 | 0,161352 | 0,995122 | Obesity | 85,86852 |
| WHR | eGFR decline (CKD population) [SH02] | Waist-to-hip ratio adjusted BMI | Inverse variance weighted | 296 | 0,029882 | 0,091041 | 0,742741 | Obesity | 85,86852 |
| WHR | eGFR decline (CKD population) [SH02] | Waist-to-hip ratio adjusted BMI | Simple mode | 296 | -0,21857 | 0,388967 | 0,574594 | Obesity | 85,86852 |
| WHR | eGFR decline (CKD population) [SH02] | Waist-to-hip ratio adjusted BMI | Weighted mode | 296 | -0,02955 | 0,258503 | 0,909072 | Obesity | 85,86852 |
| Zn | eGFR decline (CKD population) [SH02] | Zinc | MR Egger | 3 | -0,1865 | 0,357179 | 0,693649 | Minerals | 99,95282 |
| Zn | eGFR decline (CKD population) [SH02] | Zinc | Weighted median | 3 | 0,016636 | 0,092938 | 0,857938 | Minerals | 99,95282 |
| Zn | eGFR decline (CKD population) [SH02] | Zinc | Inverse variance weighted | 3 | 0,058006 | 0,114353 | 0,611978 | Minerals | 99,95282 |
| Zn | eGFR decline (CKD population) [SH02] | Zinc | Simple mode | 3 | -0,01642 | 0,116028 | 0,900439 | Minerals | 99,95282 |
| Zn | eGFR decline (CKD population) [SH02] | Zinc | Weighted mode | 3 | 0,00902 | 0,102257 | 0,937751 | Minerals | 99,95282 |
| ebi-a-GCST006979-consort | eGFR decline (CKD population) [SH02] | Heel bone mineral density | MR Egger | 483 | 0,028485 | 0,101045 | 0,778138 | Bone disease | 151,4375 |
| ebi-a-GCST006979-consort | eGFR decline (CKD population) [SH02] | Heel bone mineral density | Weighted median | 483 | 0,12415 | 0,083575 | 0,137414 | Bone disease | 151,4375 |
| ebi-a-GCST006979-consort | eGFR decline (CKD population) [SH02] | Heel bone mineral density | Inverse variance weighted | 483 | 0,085546 | 0,052185 | 0,101156 | Bone disease | 151,4375 |
| ebi-a-GCST006979-consort | eGFR decline (CKD population) [SH02] | Heel bone mineral density | Simple mode | 483 | 0,1333 | 0,223372 | 0,550948 | Bone disease | 151,4375 |
| ebi-a-GCST006979-consort | eGFR decline (CKD population) [SH02] | Heel bone mineral density | Weighted mode | 483 | 0,1333 | 0,130053 | 0,305893 | Bone disease | 151,4375 |
| ebi-a-GCST90002227 | eGFR decline (CKD population) [SH02] | Two-hour glucose | MR Egger | 14 | -0,16604 | 0,34576 | 0,639706 | Diabetes | 56,47851 |
| ebi-a-GCST90002227 | eGFR decline (CKD population) [SH02] | Two-hour glucose | Weighted median | 14 | 0,194876 | 0,159299 | 0,221203 | Diabetes | 56,47851 |
| ebi-a-GCST90002227 | eGFR decline (CKD population) [SH02] | Two-hour glucose | Inverse variance weighted | 14 | 0,163472 | 0,127675 | 0,200415 | Diabetes | 56,47851 |
| ebi-a-GCST90002227 | eGFR decline (CKD population) [SH02] | Two-hour glucose | Simple mode | 14 | -0,06621 | 0,307314 | 0,832767 | Diabetes | 56,47851 |
| ebi-a-GCST90002227 | eGFR decline (CKD population) [SH02] | Two-hour glucose | Weighted mode | 14 | 0,222235 | 0,256619 | 0,402185 | Diabetes | 56,47851 |
| ebi-a-GCST90002232 | eGFR decline (CKD population) [SH02] | Fasting glucose | MR Egger | 68 | 0,556153 | 0,323327 | 0,090102 | Diabetes | 124,2211 |
| ebi-a-GCST90002232 | eGFR decline (CKD population) [SH02] | Fasting glucose | Weighted median | 68 | 0,548739 | 0,258427 | 0,033722 | Diabetes | 124,2211 |
| ebi-a-GCST90002232 | eGFR decline (CKD population) [SH02] | Fasting glucose | Inverse variance weighted | 68 | 0,190449 | 0,170715 | 0,264595 | Diabetes | 124,2211 |
| ebi-a-GCST90002232 | eGFR decline (CKD population) [SH02] | Fasting glucose | Simple mode | 68 | 0,446221 | 0,48707 | 0,362883 | Diabetes | 124,2211 |
| ebi-a-GCST90002232 | eGFR decline (CKD population) [SH02] | Fasting glucose | Weighted mode | 68 | 0,656636 | 0,267064 | 0,016533 | Diabetes | 124,2211 |
| ebi-a-GCST90002238 | eGFR decline (CKD population) [SH02] | Fasting insulin | MR Egger | 37 | 0,45983 | 1,169713 | 0,69662 | Diabetes | 52,15552 |
| ebi-a-GCST90002238 | eGFR decline (CKD population) [SH02] | Fasting insulin | Weighted median | 37 | 0,399141 | 0,482084 | 0,4077 | Diabetes | 52,15552 |
| ebi-a-GCST90002238 | eGFR decline (CKD population) [SH02] | Fasting insulin | Inverse variance weighted | 37 | -0,00652 | 0,359522 | 0,985539 | Diabetes | 52,15552 |
| ebi-a-GCST90002238 | eGFR decline (CKD population) [SH02] | Fasting insulin | Simple mode | 37 | 0,444843 | 0,872566 | 0,613296 | Diabetes | 52,15552 |
| ebi-a-GCST90002238 | eGFR decline (CKD population) [SH02] | Fasting insulin | Weighted mode | 37 | 0,410539 | 0,688429 | 0,554679 | Diabetes | 52,15552 |
| ebi-a-GCST90007310 | eGFR decline (CKD population) [SH02] | circulating leptin levels | MR Egger | 3 | 3,077039 | 2,333926 | 0,413114 | Obesity | 56,18818 |

| | | | | | | | | | |
|---|---|---|---|---|---|---|---|---|---|
| ebi-a-GCST90007310 | eGFR decline (CKD population) [SH02] | circulating leptin levels | Weighted median | 3 | -0,21641 | 0,32664 | 0,507631 | Obesity | 56,18818 |
| ebi-a-GCST90007310 | eGFR decline (CKD population) [SH02] | circulating leptin levels | Inverse variance weighted | 3 | -0,41306 | 0,310123 | 0,182882 | Obesity | 56,18818 |
| ebi-a-GCST90007310 | eGFR decline (CKD population) [SH02] | circulating leptin levels | Simple mode | 3 | -0,186 | 0,404879 | 0,69105 | Obesity | 56,18818 |
| ebi-a-GCST90007310 | eGFR decline (CKD population) [SH02] | circulating leptin levels | Weighted mode | 3 | -0,186 | 0,379665 | 0,672669 | Obesity | 56,18818 |
| ebi-a-GCST90011998 | eGFR decline (CKD population) [SH02] | Growth differentiation factor-15 levels | MR Egger | 3 | -0,17491 | 0,138917 | 0,42731 | Biomarkers | 294,0397 |
| ebi-a-GCST90011998 | eGFR decline (CKD population) [SH02] | Growth differentiation factor-15 levels | Weighted median | 3 | -0,14081 | 0,099187 | 0,155705 | Biomarkers | 294,0397 |
| ebi-a-GCST90011998 | eGFR decline (CKD population) [SH02] | Growth differentiation factor-15 levels | Inverse variance weighted | 3 | -0,1292 | 0,095122 | 0,174385 | Biomarkers | 294,0397 |
| ebi-a-GCST90011998 | eGFR decline (CKD population) [SH02] | Growth differentiation factor-15 levels | Simple mode | 3 | -0,18454 | 0,179668 | 0,412348 | Biomarkers | 294,0397 |
| ebi-a-GCST90011998 | eGFR decline (CKD population) [SH02] | Growth differentiation factor-15 levels | Weighted mode | 3 | -0,14902 | 0,099332 | 0,272353 | Biomarkers | 294,0397 |
| ebi-a-GCST90012022 | eGFR decline (CKD population) [SH02] | Fibroblast growth factor 23 levels | MR Egger | 8 | -0,02755 | 0,477123 | 0,955821 | Biomarkers | 43,42098 |
| ebi-a-GCST90012022 | eGFR decline (CKD population) [SH02] | Fibroblast growth factor 23 levels | Weighted median | 8 | 0,143892 | 0,169425 | 0,395718 | Biomarkers | 43,42098 |
| ebi-a-GCST90012022 | eGFR decline (CKD population) [SH02] | Fibroblast growth factor 23 levels | Inverse variance weighted | 8 | 0,031775 | 0,126367 | 0,801465 | Biomarkers | 43,42098 |
| ebi-a-GCST90012022 | eGFR decline (CKD population) [SH02] | Fibroblast growth factor 23 levels | Simple mode | 8 | 0,154383 | 0,299355 | 0,621935 | Biomarkers | 43,42098 |
| ebi-a-GCST90012022 | eGFR decline (CKD population) [SH02] | Fibroblast growth factor 23 levels | Weighted mode | 8 | 0,186831 | 0,260606 | 0,496647 | Biomarkers | 43,42098 |
| ebi-a-GCST90012041 | eGFR decline (CKD population) [SH02] | Kidney injury molecule 1 levels | MR Egger | 9 | -0,02086 | 0,104418 | 0,847339 | Biomarkers | 190,9807 |
| ebi-a-GCST90012041 | eGFR decline (CKD population) [SH02] | Kidney injury molecule 1 levels | Weighted median | 9 | -0,03283 | 0,063436 | 0,604773 | Biomarkers | 190,9807 |
| ebi-a-GCST90012041 | eGFR decline (CKD population) [SH02] | Kidney injury molecule 1 levels | Inverse variance weighted | 9 | -0,04231 | 0,062482 | 0,498353 | Biomarkers | 190,9807 |
| ebi-a-GCST90012041 | eGFR decline (CKD population) [SH02] | Kidney injury molecule 1 levels | Simple mode | 9 | 0,016626 | 0,136393 | 0,905988 | Biomarkers | 190,9807 |
| ebi-a-GCST90012041 | eGFR decline (CKD population) [SH02] | Kidney injury molecule 1 levels | Weighted mode | 9 | -0,02673 | 0,067804 | 0,703667 | Biomarkers | 190,9807 |
| ebi-a-GCST90013864 | eGFR decline (CKD population) [SH02] | Coronary artery disease (Firth correction) | MR Egger | 37 | -0,10381 | 0,136232 | 0,451154 | Cardiovascular disease | 59,8949 |
| ebi-a-GCST90013864 | eGFR decline (CKD population) [SH02] | Coronary artery disease (Firth correction) | Weighted median | 37 | -0,1229 | 0,08972 | 0,170742 | Cardiovascular disease | 59,8949 |
| ebi-a-GCST90013864 | eGFR decline (CKD population) [SH02] | Coronary artery disease (Firth correction) | Inverse variance weighted | 37 | -0,03273 | 0,061318 | 0,593522 | Cardiovascular disease | 59,8949 |
| ebi-a-GCST90013864 | eGFR decline (CKD population) [SH02] | Coronary artery disease (Firth correction) | Simple mode | 37 | -0,14276 | 0,166445 | 0,396746 | Cardiovascular disease | 59,8949 |
| ebi-a-GCST90013864 | eGFR decline (CKD population) [SH02] | Coronary artery disease (Firth correction) | Weighted mode | 37 | -0,13317 | 0,115732 | 0,257435 | Cardiovascular disease | 59,8949 |
| ebi-a-GCST90013975 | eGFR decline (CKD population) [SH02] | Body fat percentage | MR Egger | 316 | 0,444484 | 0,404139 | 0,272249 | Obesity | 56,17886 |
| ebi-a-GCST90013975 | eGFR decline (CKD population) [SH02] | Body fat percentage | Weighted median | 316 | -0,07019 | 0,194354 | 0,717998 | Obesity | 56,17886 |
| ebi-a-GCST90013975 | eGFR decline (CKD population) [SH02] | Body fat percentage | Inverse variance weighted | 316 | 0,067395 | 0,125532 | 0,591356 | Obesity | 56,17886 |
| ebi-a-GCST90013975 | eGFR decline (CKD population) [SH02] | Body fat percentage | Simple mode | 316 | 0,181548 | 0,51993 | 0,727189 | Obesity | 56,17886 |
| ebi-a-GCST90013975 | eGFR decline (CKD population) [SH02] | Body fat percentage | Weighted mode | 316 | -0,2311 | 0,423501 | 0,585666 | Obesity | 56,17886 |
| ebi-a-GCST90013989 | eGFR decline (CKD population) [SH02] | Sodium in urine | MR Egger | 41 | 1,036612 | 1,106001 | 0,354392 | Minerals | 43,81211 |
| ebi-a-GCST90013989 | eGFR decline (CKD population) [SH02] | Sodium in urine | Weighted median | 41 | -0,32326 | 0,405294 | 0,425113 | Minerals | 43,81211 |
| ebi-a-GCST90013989 | eGFR decline (CKD population) [SH02] | Sodium in urine | Inverse variance weighted | 41 | -0,1888 | 0,284129 | 0,506387 | Minerals | 43,81211 |
| ebi-a-GCST90013989 | eGFR decline (CKD population) [SH02] | Sodium in urine | Simple mode | 41 | -0,24439 | 0,913954 | 0,790534 | Minerals | 43,81211 |
| ebi-a-GCST90013989 | eGFR decline (CKD population) [SH02] | Sodium in urine | Weighted mode | 41 | -0,57402 | 0,792198 | 0,472922 | Minerals | 43,81211 |
| ebi-a-GCST90014006 | eGFR decline (CKD population) [SH02] | Glycated haemoglobin HbA1c levels | MR Egger | 302 | 0,246521 | 0,123711 | 0,047198 | Diabetes | 128,2905 |
| ebi-a-GCST90014006 | eGFR decline (CKD population) [SH02] | Glycated haemoglobin HbA1c levels | Weighted median | 302 | 0,155415 | 0,106248 | 0,143535 | Diabetes | 128,2905 |
| ebi-a-GCST90014006 | eGFR decline (CKD population) [SH02] | Glycated haemoglobin HbA1c levels | Inverse variance weighted | 302 | 0,055793 | 0,065154 | 0,391816 | Diabetes | 128,2905 |
| ebi-a-GCST90014006 | eGFR decline (CKD population) [SH02] | Glycated haemoglobin HbA1c levels | Simple mode | 302 | -0,05305 | 0,257274 | 0,836783 | Diabetes | 128,2905 |
| ebi-a-GCST90014006 | eGFR decline (CKD population) [SH02] | Glycated haemoglobin HbA1c levels | Weighted mode | 302 | 0,097443 | 0,143533 | 0,497728 | Diabetes | 128,2905 |
| ebi-a-GCST90014010 | eGFR decline (CKD population) [SH02] | Phosphate levels | MR Egger | 160 | 0,157435 | 0,170886 | 0,358306 | Minerals | 87,71201 |
| ebi-a-GCST90014010 | eGFR decline (CKD population) [SH02] | Phosphate levels | Weighted median | 160 | -0,01875 | 0,165142 | 0,90959 | Minerals | 87,71201 |
| ebi-a-GCST90014010 | eGFR decline (CKD population) [SH02] | Phosphate levels | Inverse variance weighted | 160 | -0,04808 | 0,095794 | 0,61574 | Minerals | 87,71201 |
| ebi-a-GCST90014010 | eGFR decline (CKD population) [SH02] | Phosphate levels | Simple mode | 160 | -0,02079 | 0,351929 | 0,952978 | Minerals | 87,71201 |
| ebi-a-GCST90014010 | eGFR decline (CKD population) [SH02] | Phosphate levels | Weighted mode | 160 | 0,096823 | 0,212986 | 0,65002 | Minerals | 87,71201 |
| ebi-a-GCST90014022 | eGFR decline (CKD population) [SH02] | Bone mineral density | MR Egger | 415 | 0,060737 | 0,111441 | 0,586037 | Bone disease | 120,0409 |
| ebi-a-GCST90014022 | eGFR decline (CKD population) [SH02] | Bone mineral density | Weighted median | 415 | 0,099802 | 0,094415 | 0,290484 | Bone disease | 120,0409 |
| ebi-a-GCST90014022 | eGFR decline (CKD population) [SH02] | Bone mineral density | Inverse variance weighted | 415 | 0,083495 | 0,056199 | 0,137359 | Bone disease | 120,0409 |
| ebi-a-GCST90014022 | eGFR decline (CKD population) [SH02] | Bone mineral density | Simple mode | 415 | 0,04789 | 0,206908 | 0,817075 | Bone disease | 120,0409 |
| ebi-a-GCST90014022 | eGFR decline (CKD population) [SH02] | Bone mineral density | Weighted mode | 415 | 0,04789 | 0,128032 | 0,70856 | Bone disease | 120,0409 |
| ebi-a-GCST90014023 | eGFR decline (CKD population) [SH02] | Type 1 diabetes | MR Egger | 76 | -0,02453 | 0,029131 | 0,402551 | Diabetes | 131,7111 |
| ebi-a-GCST90014023 | eGFR decline (CKD population) [SH02] | Type 1 diabetes | Weighted median | 76 | -0,02905 | 0,027309 | 0,287505 | Diabetes | 131,7111 |
| ebi-a-GCST90014023 | eGFR decline (CKD population) [SH02] | Type 1 diabetes | Inverse variance weighted | 76 | -0,01926 | 0,01776 | 0,278181 | Diabetes | 131,7111 |
| ebi-a-GCST90014023 | eGFR decline (CKD population) [SH02] | Type 1 diabetes | Simple mode | 76 | -0,04861 | 0,053805 | 0,369133 | Diabetes | 131,7111 |
| ebi-a-GCST90014023 | eGFR decline (CKD population) [SH02] | Type 1 diabetes | Weighted mode | 76 | -0,03911 | 0,027497 | 0,159071 | Diabetes | 131,7111 |
| ebi-a-GCST90016674 | eGFR decline (CKD population) [SH02] | Liver iron content | MR Egger | 5 | -0,06668 | 0,178445 | 0,733485 | Minerals | 257,3642 |
| ebi-a-GCST90016674 | eGFR decline (CKD population) [SH02] | Liver iron content | Weighted median | 5 | -0,07638 | 0,108515 | 0,481533 | Minerals | 257,3642 |
| ebi-a-GCST90016674 | eGFR decline (CKD population) [SH02] | Liver iron content | Inverse variance weighted | 5 | -0,0719 | 0,102097 | 0,481267 | Minerals | 257,3642 |
| ebi-a-GCST90016674 | eGFR decline (CKD population) [SH02] | Liver iron content | Simple mode | 5 | -0,13503 | 0,206846 | 0,549502 | Minerals | 257,3642 |
| ebi-a-GCST90016674 | eGFR decline (CKD population) [SH02] | Liver iron content | Weighted mode | 5 | -0,08077 | 0,119867 | 0,537347 | Minerals | 257,3642 |
| ebi-a-GCST90016676 | eGFR decline (CKD population) [SH02] | Pancreas iron content | Wald ratio | 1 | -0,25459 | 0,552941 | 0,645212 | Minerals | 30,56027 |
| ebi-a-GCST90018806 | eGFR decline (CKD population) [SH02] | Chronic heart failure | MR Egger | 5 | 0,704018 | 1,316459 | 0,629898 | Cardiovascular disease | 41,81649 |
| ebi-a-GCST90018806 | eGFR decline (CKD population) [SH02] | Chronic heart failure | Weighted median | 5 | 0,243074 | 0,219096 | 0,26724 | Cardiovascular disease | 41,81649 |
| ebi-a-GCST90018806 | eGFR decline (CKD population) [SH02] | Chronic heart failure | Inverse variance weighted | 5 | 0,109492 | 0,189891 | 0,564207 | Cardiovascular disease | 41,81649 |
| ebi-a-GCST90018806 | eGFR decline (CKD population) [SH02] | Chronic heart failure | Simple mode | 5 | 0,300158 | 0,311731 | 0,390137 | Cardiovascular disease | 41,81649 |
| ebi-a-GCST90018806 | eGFR decline (CKD population) [SH02] | Chronic heart failure | Weighted mode | 5 | 0,303748 | 0,305874 | 0,376895 | Cardiovascular disease | 41,81649 |
| ebi-a-GCST90018860 | eGFR decline (CKD population) [SH02] | Hyperthyroidism | MR Egger | 8 | -0,15503 | 0,280834 | 0,60086 | Thyroid disease | 53,27573 |
| ebi-a-GCST90018860 | eGFR decline (CKD population) [SH02] | Hyperthyroidism | Weighted median | 8 | 0,159385 | 0,082238 | 0,052613 | Thyroid disease | 53,27573 |
| ebi-a-GCST90018860 | eGFR decline (CKD population) [SH02] | Hyperthyroidism | Inverse variance weighted | 8 | 0,079629 | 0,079341 | 0,315559 | Thyroid disease | 53,27573 |
| ebi-a-GCST90018860 | eGFR decline (CKD population) [SH02] | Hyperthyroidism | Simple mode | 8 | 0,247045 | 0,156103 | 0,157534 | Thyroid disease | 53,27573 |
| ebi-a-GCST90018860 | eGFR decline (CKD population) [SH02] | Hyperthyroidism | Weighted mode | 8 | 0,185564 | 0,135123 | 0,21203 | Thyroid disease | 53,27573 |
| ebi-a-GCST90018862 | eGFR decline (CKD population) [SH02] | Hypothyroidism | MR Egger | 64 | -0,04899 | 0,092146 | 0,596828 | Thyroid disease | 82,13671 |
| ebi-a-GCST90018862 | eGFR decline (CKD population) [SH02] | Hypothyroidism | Weighted median | 64 | 0,026224 | 0,05804 | 0,651399 | Thyroid disease | 82,13671 |
| ebi-a-GCST90018862 | eGFR decline (CKD population) [SH02] | Hypothyroidism | Inverse variance weighted | 64 | 0,03462 | 0,039714 | 0,383352 | Thyroid disease | 82,13671 |
| ebi-a-GCST90018862 | eGFR decline (CKD population) [SH02] | Hypothyroidism | Simple mode | 64 | 0,075388 | 0,125795 | 0,551127 | Thyroid disease | 82,13671 |
| ebi-a-GCST90018862 | eGFR decline (CKD population) [SH02] | Hypothyroidism | Weighted mode | 64 | 0,00516 | 0,0815 | 0,949713 | Thyroid disease | 82,13671 |
| ebi-a-GCST90018872 | eGFR decline (CKD population) [SH02] | Iron deficiency anemia | Wald ratio | 1 | 0,683922 | 0,404498 | 0,090876 | Minerals | 34,53822 |
| ebi-a-GCST90018877 | eGFR decline (CKD population) [SH02] | Myocardial infarction | MR Egger | 70 | -0,12179 | 0,097327 | 0,215104 | Cardiovascular disease | 66,68595 |
| ebi-a-GCST90018877 | eGFR decline (CKD population) [SH02] | Myocardial infarction | Weighted median | 70 | -0,0465 | 0,066351 | 0,483411 | Cardiovascular disease | 66,68595 |
| ebi-a-GCST90018877 | eGFR decline (CKD population) [SH02] | Myocardial infarction | Inverse variance weighted | 70 | 0,020807 | 0,043506 | 0,632476 | Cardiovascular disease | 66,68595 |
| ebi-a-GCST90018877 | eGFR decline (CKD population) [SH02] | Myocardial infarction | Simple mode | 70 | -0,00154 | 0,132968 | 0,99079 | Cardiovascular disease | 66,68595 |
| ebi-a-GCST90018877 | eGFR decline (CKD population) [SH02] | Myocardial infarction | Weighted mode | 70 | -0,06306 | 0,084606 | 0,458608 | Cardiovascular disease | 66,68595 |
| ebi-a-GCST90018916 | eGFR decline (CKD population) [SH02] | Sleep apnea syndrome | Wald ratio | 1 | -0,1035 | 0,243148 | 0,670349 | Sleep disorders | 63,50864 |
| ebi-a-GCST90018977 | eGFR decline (CKD population) [SH02] | Serum uric acid levels | MR Egger | 237 | -0,00832 | 0,157067 | 0,957823 | Uricemia | 110,1094 |
| ebi-a-GCST90018977 | eGFR decline (CKD population) [SH02] | Serum uric acid levels | Weighted median | 237 | 0,114502 | 0,144018 | 0,426582 | Uricemia | 110,1094 |
| ebi-a-GCST90018977 | eGFR decline (CKD population) [SH02] | Serum uric acid levels | Inverse variance weighted | 237 | 0,075424 | 0,087206 | 0,387098 | Uricemia | 110,1094 |
| ebi-a-GCST90018977 | eGFR decline (CKD population) [SH02] | Serum uric acid levels | Simple mode | 237 | 0,365613 | 0,34859 | 0,295328 | Uricemia | 110,1094 |
| ebi-a-GCST90018977 | eGFR decline (CKD population) [SH02] | Serum uric acid levels | Weighted mode | 237 | 0,110992 | 0,161622 | 0,492925 | Uricemia | 110,1094 |
| ebi-a-GCST90019417 | eGFR decline (CKD population) [SH02] | Vitamin K-dependent protein S measurement | MR Egger | 4 | 0,049563 | 0,158275 | 0,783809 | Vitamin | 118,3171 |
| ebi-a-GCST90019417 | eGFR decline (CKD population) [SH02] | Vitamin K-dependent protein S measurement | Weighted median | 4 | 0,020642 | 0,102693 | 0,840692 | Vitamin | 118,3171 |
| ebi-a-GCST90019417 | eGFR decline (CKD population) [SH02] | Vitamin K-dependent protein S measurement | Inverse variance weighted | 4 | 0,011625 | 0,090679 | 0,897989 | Vitamin | 118,3171 |
| ebi-a-GCST90019417 | eGFR decline (CKD population) [SH02] | Vitamin K-dependent protein S measurement | Simple mode | 4 | -0,03048 | 0,13689 | 0,838104 | Vitamin | 118,3171 |
| ebi-a-GCST90019417 | eGFR decline (CKD population) [SH02] | Vitamin K-dependent protein S measurement | Weighted mode | 4 | 0,034597 | 0,113708 | 0,780815 | Vitamin | 118,3171 |
| ebi-a-GCST90019423 | eGFR decline (CKD population) [SH02] | Vitamin K-dependent protein C measurement | MR Egger | 3 | 0,081928 | 0,241616 | 0,791878 | Vitamin | 207,8561 |
| ebi-a-GCST90019423 | eGFR decline (CKD population) [SH02] | Vitamin K-dependent protein C measurement | Weighted median | 3 | 0,07063 | 0,073433 | 0,336135 | Vitamin | 207,8561 |
| ebi-a-GCST90019423 | eGFR decline (CKD population) [SH02] | Vitamin K-dependent protein C measurement | Inverse variance weighted | 3 | 0,042082 | 0,109763 | 0,701426 | Vitamin | 207,8561 |
| ebi-a-GCST90019423 | eGFR decline (CKD population) [SH02] | Vitamin K-dependent protein C measurement | Simple mode | 3 | 0,024084 | 0,189223 | 0,910365 | Vitamin | 207,8561 |
| ebi-a-GCST90019423 | eGFR decline (CKD population) [SH02] | Vitamin K-dependent protein C measurement | Weighted mode | 3 | 0,078757 | 0,073001 | 0,393474 | Vitamin | 207,8561 |
| ebi-a-GCST90025947 | eGFR decline (CKD population) [SH02] | Serum alkaline phosphatase levels | MR Egger | 335 | -0,07111 | 0,102504 | 0,488333 | Minerals | 166,037 |
| ebi-a-GCST90025947 | eGFR decline (CKD population) [SH02] | Serum alkaline phosphatase levels | Weighted median | 335 | 0,119128 | 0,105518 | 0,258905 | Minerals | 166,037 |
| ebi-a-GCST90025947 | eGFR decline (CKD population) [SH02] | Serum alkaline phosphatase levels | Inverse variance weighted | 335 | -0,01428 | 0,060046 | 0,812024 | Minerals | 166,037 |
| ebi-a-GCST90025947 | eGFR decline (CKD population) [SH02] | Serum alkaline phosphatase levels | Simple mode | 335 | 0,15389 | 0,2277 | 0,499606 | Minerals | 166,037 |
| ebi-a-GCST90025947 | eGFR decline (CKD population) [SH02] | Serum alkaline phosphatase levels | Weighted mode | 335 | 0,100956 | 0,122526 | 0,41055 | Minerals | 166,037 |
| ebi-a-GCST90025967 | eGFR decline (CKD population) [SH02] | Vitamin D levels | MR Egger | 81 | -0,06217 | 0,191308 | 0,746077 | Vitamin | 134,1402 |
| ebi-a-GCST90025967 | eGFR decline (CKD population) [SH02] | Vitamin D levels | Weighted median | 81 | -0,21387 | 0,19095 | 0,26269 | Vitamin | 134,1402 |
| ebi-a-GCST90025967 | eGFR decline (CKD population) [SH02] | Vitamin D levels | Inverse variance weighted | 81 | -0,09226 | 0,117391 | 0,431933 | Vitamin | 134,1402 |
| ebi-a-GCST90025967 | eGFR decline (CKD population) [SH02] | Vitamin D levels | Simple mode | 81 | -0,11057 | 0,360234 | 0,759688 | Vitamin | 134,1402 |
| ebi-a-GCST90025967 | eGFR decline (CKD population) [SH02] | Vitamin D levels | Weighted mode | 81 | -0,13411 | 0,169934 | 0,43235 | Vitamin | 134,1402 |
| ebi-a-GCST90025969 | eGFR decline (CKD population) [SH02] | Hemoglobin levels | MR Egger | 325 | -0,00196 | 0,138872 | 0,988754 | Organic compound | 104,3556 |
| ebi-a-GCST90025969 | eGFR decline (CKD population) [SH02] | Hemoglobin levels | Weighted median | 325 | -0,06895 | 0,122028 | 0,572044 | Organic compound | 104,3556 |
| ebi-a-GCST90025969 | eGFR decline (CKD population) [SH02] | Hemoglobin levels | Inverse variance weighted | 325 | -0,10569 | 0,071601 | 0,139906 | Organic compound | 104,3556 |
| ebi-a-GCST90025969 | eGFR decline (CKD population) [SH02] | Hemoglobin levels | Simple mode | 325 | 0,193666 | 0,28312 | 0,494439 | Organic compound | 104,3556 |
| ebi-a-GCST90025969 | eGFR decline (CKD population) [SH02] | Hemoglobin levels | Weighted mode | 325 | -0,06925 | 0,144965 | 0,633196 | Organic compound | 104,3556 |
| ebi-a-GCST90025986 | eGFR decline (CKD population) [SH02] | Blood glucose levels | MR Egger | 109 | 0,188904 | 0,19205 | 0,32752 | Diabetes | 118,7554 |

| | | | | | | | | | |
|---|---|---|---|---|---|---|---|---|---|
| ebi-a-GCST90025986 | eGFR decline (CKD population) [SH02] | Blood glucose levels | Weighted median | 109 | 0,385016 | 0,168833 | 0,02258 | Diabetes | 118,7554 |
| ebi-a-GCST90025986 | eGFR decline (CKD population) [SH02] | Blood glucose levels | Inverse variance weighted | 109 | 0,018101 | 0,107867 | 0,866732 | Diabetes | 118,7554 |
| ebi-a-GCST90025986 | eGFR decline (CKD population) [SH02] | Blood glucose levels | Simple mode | 109 | 0,324951 | 0,421002 | 0,441887 | Diabetes | 118,7554 |
| ebi-a-GCST90025986 | eGFR decline (CKD population) [SH02] | Blood glucose levels | Weighted mode | 109 | 0,324951 | 0,170201 | 0,058887 | Diabetes | 118,7554 |
| ebi-a-GCST90025989 | eGFR decline (CKD population) [SH02] | Insulin-like growth factor 1 levels | MR Egger | 373 | -0,19411 | 0,121854 | 0,11201 | Diabetes | 115,9356 |
| ebi-a-GCST90025989 | eGFR decline (CKD population) [SH02] | Insulin-like growth factor 1 levels | Weighted median | 373 | -0,00564 | 0,11056 | 0,959289 | Diabetes | 115,9356 |
| ebi-a-GCST90025989 | eGFR decline (CKD population) [SH02] | Insulin-like growth factor 1 levels | Inverse variance weighted | 373 | 0,012837 | 0,063074 | 0,838732 | Diabetes | 115,9356 |
| ebi-a-GCST90025989 | eGFR decline (CKD population) [SH02] | Insulin-like growth factor 1 levels | Simple mode | 373 | -0,10954 | 0,256183 | 0,6692 | Diabetes | 115,9356 |
| ebi-a-GCST90025989 | eGFR decline (CKD population) [SH02] | Insulin-like growth factor 1 levels | Weighted mode | 373 | -0,14107 | 0,130767 | 0,28138 | Diabetes | 115,9356 |
| ebi-a-GCST90025990 | eGFR decline (CKD population) [SH02] | Calcium levels | MR Egger | 202 | 0,299167 | 0,153971 | 0,053419 | Minerals | 102,0722 |
| ebi-a-GCST90025990 | eGFR decline (CKD population) [SH02] | Calcium levels | Weighted median | 202 | 0,108352 | 0,171123 | 0,526613 | Minerals | 102,0722 |
| ebi-a-GCST90025990 | eGFR decline (CKD population) [SH02] | Calcium levels | Inverse variance weighted | 202 | 0,065179 | 0,083364 | 0,434295 | Minerals | 102,0722 |
| ebi-a-GCST90025990 | eGFR decline (CKD population) [SH02] | Calcium levels | Simple mode | 202 | -0,0168 | 0,322891 | 0,958557 | Minerals | 102,0722 |
| ebi-a-GCST90025990 | eGFR decline (CKD population) [SH02] | Calcium levels | Weighted mode | 202 | 0,116033 | 0,170824 | 0,497757 | Minerals | 102,0722 |
| ebi-a-GCST90029013 | eGFR decline (CKD population) [SH02] | Educational attainment (years of education) | MR Egger | 209 | 0,027181 | 0,115602 | 0,814343 | Social trait | 45,05626 |
| ebi-a-GCST90029013 | eGFR decline (CKD population) [SH02] | Educational attainment (years of education) | Weighted median | 209 | -0,00583 | 0,039702 | 0,883237 | Social trait | 45,05626 |
| ebi-a-GCST90029013 | eGFR decline (CKD population) [SH02] | Educational attainment (years of education) | Inverse variance weighted | 209 | -0,03819 | 0,026457 | 0,148844 | Social trait | 45,05626 |
| ebi-a-GCST90029013 | eGFR decline (CKD population) [SH02] | Educational attainment (years of education) | Simple mode | 209 | 0,010823 | 0,1143 | 0,924653 | Social trait | 45,05626 |
| ebi-a-GCST90029013 | eGFR decline (CKD population) [SH02] | Educational attainment (years of education) | Weighted mode | 209 | 0,047911 | 0,097923 | 0,625162 | Social trait | 45,05626 |
| ebi-a-GCST90029014 | eGFR decline (CKD population) [SH02] | Smoking status | MR Egger | 117 | -0,40299 | 1,103727 | 0,715695 | Smoking | 42,91926 |
| ebi-a-GCST90029014 | eGFR decline (CKD population) [SH02] | Smoking status | Weighted median | 117 | 0,031972 | 0,380237 | 0,932989 | Smoking | 42,91926 |
| ebi-a-GCST90029014 | eGFR decline (CKD population) [SH02] | Smoking status | Inverse variance weighted | 117 | -0,01855 | 0,261787 | 0,943508 | Smoking | 42,91926 |
| ebi-a-GCST90029014 | eGFR decline (CKD population) [SH02] | Smoking status | Simple mode | 117 | -0,49434 | 0,93278 | 0,597151 | Smoking | 42,91926 |
| ebi-a-GCST90029014 | eGFR decline (CKD population) [SH02] | Smoking status | Weighted mode | 117 | -0,30282 | 0,788362 | 0,701597 | Smoking | 42,91926 |
| ebi-a-GCST90029070 | eGFR decline (CKD population) [SH02] | C-reactive protein levels | MR Egger | 214 | 0,277442 | 0,131134 | 0,035537 | Inflammation | 122,6906 |
| ebi-a-GCST90029070 | eGFR decline (CKD population) [SH02] | C-reactive protein levels | Weighted median | 214 | 0,0914 | 0,123229 | 0,458265 | Inflammation | 122,6906 |
| ebi-a-GCST90029070 | eGFR decline (CKD population) [SH02] | C-reactive protein levels | Inverse variance weighted | 214 | 0,098334 | 0,077398 | 0,203908 | Inflammation | 122,6906 |
| ebi-a-GCST90029070 | eGFR decline (CKD population) [SH02] | C-reactive protein levels | Simple mode | 214 | -0,3915 | 0,272574 | 0,152387 | Inflammation | 122,6906 |
| ebi-a-GCST90029070 | eGFR decline (CKD population) [SH02] | C-reactive protein levels | Weighted mode | 214 | 0,205201 | 0,122974 | 0,096654 | Inflammation | 122,6906 |
| ebi-a-GCST90029073 | eGFR decline (CKD population) [SH02] | Liver fat | MR Egger | 8 | 0,289242 | 0,160865 | 0,122283 | Fatty liver | 140,8023 |
| ebi-a-GCST90029073 | eGFR decline (CKD population) [SH02] | Liver fat | Weighted median | 8 | 0,185086 | 0,116139 | 0,111012 | Fatty liver | 140,8023 |
| ebi-a-GCST90029073 | eGFR decline (CKD population) [SH02] | Liver fat | Inverse variance weighted | 8 | 0,182612 | 0,09765 | 0,061473 | Fatty liver | 140,8023 |
| ebi-a-GCST90029073 | eGFR decline (CKD population) [SH02] | Liver fat | Simple mode | 8 | 0,23967 | 0,191794 | 0,251598 | Fatty liver | 140,8023 |
| ebi-a-GCST90029073 | eGFR decline (CKD population) [SH02] | Liver fat | Weighted mode | 8 | 0,227952 | 0,124876 | 0,110685 | Fatty liver | 140,8023 |
| ebi-a-GCST90086057 | eGFR decline (CKD population) [SH02] | Cardiovascular disease | Inverse variance weighted | 2 | 0,118965 | 0,212701 | 0,575954 | Cardiovascular disease | 32,018 |
| finn-b-D3_ANAEMIA_B12_DEF | eGFR decline (CKD population) [SH02] | Vitamin B12 deficiency anaemia | MR Egger | 4 | -0,07375 | 0,156186 | 0,683294 | Vitamin | 52,61875 |
| finn-b-D3_ANAEMIA_B12_DEF | eGFR decline (CKD population) [SH02] | Vitamin B12 deficiency anaemia | Weighted median | 4 | -0,05668 | 0,063505 | 0,372076 | Vitamin | 52,61875 |
| finn-b-D3_ANAEMIA_B12_DEF | eGFR decline (CKD population) [SH02] | Vitamin B12 deficiency anaemia | Inverse variance weighted | 4 | -0,02904 | 0,054564 | 0,594576 | Vitamin | 52,61875 |
| finn-b-D3_ANAEMIA_B12_DEF | eGFR decline (CKD population) [SH02] | Vitamin B12 deficiency anaemia | Simple mode | 4 | -0,06934 | 0,089388 | 0,494466 | Vitamin | 52,61875 |
| finn-b-D3_ANAEMIA_B12_DEF | eGFR decline (CKD population) [SH02] | Vitamin B12 deficiency anaemia | Weighted mode | 4 | -0,07247 | 0,070463 | 0,379384 | Vitamin | 52,61875 |
| finn-b-E4_OBESITY | eGFR decline (CKD population) [SH02] | Obesity | MR Egger | 7 | -0,05561 | 0,293843 | 0,857345 | Obesity | 59,53804 |
| finn-b-E4_OBESITY | eGFR decline (CKD population) [SH02] | Obesity | Weighted median | 7 | -0,02483 | 0,100688 | 0,805233 | Obesity | 59,53804 |
| finn-b-E4_OBESITY | eGFR decline (CKD population) [SH02] | Obesity | Inverse variance weighted | 7 | 0,027062 | 0,080999 | 0,738307 | Obesity | 59,53804 |
| finn-b-E4_OBESITY | eGFR decline (CKD population) [SH02] | Obesity | Simple mode | 7 | 0,005263 | 0,161811 | 0,975109 | Obesity | 59,53804 |
| finn-b-E4_OBESITY | eGFR decline (CKD population) [SH02] | Obesity | Weighted mode | 7 | -0,05896 | 0,108714 | 0,607102 | Obesity | 59,53804 |
| finn-b-FG_PULMHEART | eGFR decline (CKD population) [SH02] | Pulmonary heart disease | MR Egger | 5 | -0,22808 | 0,154262 | 0,235796 | Cardiovascular disease | 89,00197 |
| finn-b-FG_PULMHEART | eGFR decline (CKD population) [SH02] | Pulmonary heart disease | Weighted median | 5 | 0,078044 | 0,067015 | 0,244189 | Cardiovascular disease | 89,00197 |
| finn-b-FG_PULMHEART | eGFR decline (CKD population) [SH02] | Pulmonary heart disease | Inverse variance weighted | 5 | 0,01315 | 0,057426 | 0,818873 | Cardiovascular disease | 89,00197 |
| finn-b-FG_PULMHEART | eGFR decline (CKD population) [SH02] | Pulmonary heart disease | Simple mode | 5 | 0,110032 | 0,111033 | 0,377787 | Cardiovascular disease | 89,00197 |
| finn-b-FG_PULMHEART | eGFR decline (CKD population) [SH02] | Pulmonary heart disease | Weighted mode | 5 | 0,110032 | 0,108206 | 0,366715 | Cardiovascular disease | 89,00197 |
| ieu-b-102 | eGFR decline (CKD population) [SH02] | Major depression | MR Egger | 48 | 0,27815 | 0,819154 | 0,735735 | Mental health | 38,29386 |
| ieu-b-102 | eGFR decline (CKD population) [SH02] | Major depression | Weighted median | 48 | -0,02068 | 0,203064 | 0,918902 | Mental health | 38,29386 |
| ieu-b-102 | eGFR decline (CKD population) [SH02] | Major depression | Inverse variance weighted | 48 | -0,09692 | 0,147236 | 0,510376 | Mental health | 38,29386 |
| ieu-b-102 | eGFR decline (CKD population) [SH02] | Major depression | Simple mode | 48 | 0,142766 | 0,461779 | 0,758562 | Mental health | 38,29386 |
| ieu-b-102 | eGFR decline (CKD population) [SH02] | Major depression | Weighted mode | 48 | 0,078599 | 0,441995 | 0,859622 | Mental health | 38,29386 |
| ieu-b-107 | eGFR decline (CKD population) [SH02] | apolipoprotein A-I | MR Egger | 269 | -0,11121 | 0,114161 | 0,330874 | Dyslipidemias | 139,7111 |
| ieu-b-107 | eGFR decline (CKD population) [SH02] | apolipoprotein A-I | Weighted median | 269 | -0,11546 | 0,113437 | 0,308755 | Dyslipidemias | 139,7111 |
| ieu-b-107 | eGFR decline (CKD population) [SH02] | apolipoprotein A-I | Inverse variance weighted | 269 | -0,04272 | 0,067494 | 0,526736 | Dyslipidemias | 139,7111 |
| ieu-b-107 | eGFR decline (CKD population) [SH02] | apolipoprotein A-I | Simple mode | 269 | -0,13386 | 0,229395 | 0,560033 | Dyslipidemias | 139,7111 |
| ieu-b-107 | eGFR decline (CKD population) [SH02] | apolipoprotein A-I | Weighted mode | 269 | -0,13386 | 0,126493 | 0,29091 | Dyslipidemias | 139,7111 |
| ieu-b-108 | eGFR decline (CKD population) [SH02] | apolipoprotein B | MR Egger | 173 | 0,072272 | 0,124069 | 0,560989 | Dyslipidemias | 162,3296 |
| ieu-b-108 | eGFR decline (CKD population) [SH02] | apolipoprotein B | Weighted median | 173 | 0,142546 | 0,12522 | 0,254967 | Dyslipidemias | 162,3296 |
| ieu-b-108 | eGFR decline (CKD population) [SH02] | apolipoprotein B | Inverse variance weighted | 173 | 0,09342 | 0,076345 | 0,22108 | Dyslipidemias | 162,3296 |
| ieu-b-108 | eGFR decline (CKD population) [SH02] | apolipoprotein B | Simple mode | 173 | 0,132605 | 0,240879 | 0,582689 | Dyslipidemias | 162,3296 |
| ieu-b-108 | eGFR decline (CKD population) [SH02] | apolipoprotein B | Weighted mode | 173 | 0,155761 | 0,133184 | 0,243812 | Dyslipidemias | 162,3296 |
| ieu-b-25 | eGFR decline (CKD population) [SH02] | Cigarettes per Day | MR Egger | 22 | -0,25476 | 0,173999 | 0,158695 | Smoking | 99,80012 |
| ieu-b-25 | eGFR decline (CKD population) [SH02] | Cigarettes per Day | Weighted median | 22 | -0,18724 | 0,135907 | 0,168282 | Smoking | 99,80012 |
| ieu-b-25 | eGFR decline (CKD population) [SH02] | Cigarettes per Day | Inverse variance weighted | 22 | -0,16863 | 0,100122 | 0,092143 | Smoking | 99,80012 |
| ieu-b-25 | eGFR decline (CKD population) [SH02] | Cigarettes per Day | Simple mode | 22 | -0,5038 | 0,298335 | 0,10607 | Smoking | 99,80012 |
| ieu-b-25 | eGFR decline (CKD population) [SH02] | Cigarettes per Day | Weighted mode | 22 | -0,16004 | 0,146846 | 0,288116 | Smoking | 99,80012 |
| ieu-b-4861 | eGFR decline (CKD population) [SH02] | Chronotype | MR Egger | 60 | -0,45381 | 0,375938 | 0,232274 | Sleep disorders | 44,49296 |
| ieu-b-4861 | eGFR decline (CKD population) [SH02] | Chronotype | Weighted median | 60 | -0,10205 | 0,198451 | 0,607084 | Sleep disorders | 44,49296 |
| ieu-b-4861 | eGFR decline (CKD population) [SH02] | Chronotype | Inverse variance weighted | 60 | -0,00433 | 0,135874 | 0,974559 | Sleep disorders | 44,49296 |
| ieu-b-4861 | eGFR decline (CKD population) [SH02] | Chronotype | Simple mode | 60 | -0,03611 | 0,474935 | 0,939645 | Sleep disorders | 44,49296 |
| ieu-b-4861 | eGFR decline (CKD population) [SH02] | Chronotype | Weighted mode | 60 | -0,12893 | 0,44113 | 0,771105 | Sleep disorders | 44,49296 |
| ieu-b-4877 | eGFR decline (CKD population) [SH02] | smoking initiation | MR Egger | 85 | 1,551365 | 0,604373 | 0,012056 | Smoking | 42,2587 |
| ieu-b-4877 | eGFR decline (CKD population) [SH02] | smoking initiation | Weighted median | 85 | 0,140178 | 0,18134 | 0,439513 | Smoking | 42,2587 |
| ieu-b-4877 | eGFR decline (CKD population) [SH02] | smoking initiation | Inverse variance weighted | 85 | 0,177448 | 0,118946 | 0,135741 | Smoking | 42,2587 |
| ieu-b-4877 | eGFR decline (CKD population) [SH02] | smoking initiation | Simple mode | 85 | 0,300718 | 0,429707 | 0,485973 | Smoking | 42,2587 |
| ieu-b-4877 | eGFR decline (CKD population) [SH02] | smoking initiation | Weighted mode | 85 | 0,230096 | 0,380381 | 0,54687 | Smoking | 42,2587 |
| ieu-b-5144 | eGFR decline (CKD population) [SH02] | Hypertension | MR Egger | 187 | -1,49178 | 0,893973 | 0,096866 | Hypertension | 54,118 |
| ieu-b-5144 | eGFR decline (CKD population) [SH02] | Hypertension | Weighted median | 187 | -0,63568 | 0,413236 | 0,123977 | Hypertension | 54,118 |
| ieu-b-5144 | eGFR decline (CKD population) [SH02] | Hypertension | Inverse variance weighted | 187 | -0,50839 | 0,288507 | 0,078044 | Hypertension | 54,118 |
| ieu-b-5144 | eGFR decline (CKD population) [SH02] | Hypertension | Simple mode | 187 | -1,16176 | 1,147515 | 0,312656 | Hypertension | 54,118 |
| ieu-b-5144 | eGFR decline (CKD population) [SH02] | Hypertension | Weighted mode | 187 | -1,38256 | 0,938091 | 0,142226 | Hypertension | 54,118 |
| ieu-b-72 | eGFR decline (CKD population) [SH02] | Urinary sodium-potassium ratio | MR Egger | 24 | 0,876749 | 1,59991 | 0,58921 | Minerals | 39,88328 |
| ieu-b-72 | eGFR decline (CKD population) [SH02] | Urinary sodium-potassium ratio | Weighted median | 24 | 0,223278 | 0,48343 | 0,64418 | Minerals | 39,88328 |
| ieu-b-72 | eGFR decline (CKD population) [SH02] | Urinary sodium-potassium ratio | Inverse variance weighted | 24 | 0,357651 | 0,369653 | 0,333279 | Minerals | 39,88328 |
| ieu-b-72 | eGFR decline (CKD population) [SH02] | Urinary sodium-potassium ratio | Simple mode | 24 | -0,21697 | 0,842747 | 0,799112 | Minerals | 39,88328 |
| ieu-b-72 | eGFR decline (CKD population) [SH02] | Urinary sodium-potassium ratio | Weighted mode | 24 | -0,1901 | 0,945706 | 0,842457 | Minerals | 39,88328 |
| prot-a-3102 | eGFR decline (CKD population) [SH02] | Thyrotropin-releasing hormone | Inverse variance weighted | 2 | 0,139578 | 0,135821 | 0,30411 | Thyroid disease | 36,35446 |
| prot-c-2652_15_1 | eGFR decline (CKD population) [SH02] | suPAR | Wald ratio | 1 | 0,035939 | 0,105178 | 0,732581 | Biomarkers | 33,5125 |
| ukb-b-4424 | eGFR decline (CKD population) [SH02] | Sleep duration | MR Egger | 66 | -0,00146 | 1,151687 | 0,998993 | Sleep disorders | 42,80283 |
| ukb-b-4424 | eGFR decline (CKD population) [SH02] | Sleep duration | Weighted median | 66 | -0,13854 | 0,444314 | 0,75519 | Sleep disorders | 42,80283 |
| ukb-b-4424 | eGFR decline (CKD population) [SH02] | Sleep duration | Inverse variance weighted | 66 | -0,17288 | 0,301653 | 0,566572 | Sleep disorders | 42,80283 |
| ukb-b-4424 | eGFR decline (CKD population) [SH02] | Sleep duration | Simple mode | 66 | -0,36106 | 0,972395 | 0,711617 | Sleep disorders | 42,80283 |
| ukb-b-4424 | eGFR decline (CKD population) [SH02] | Sleep duration | Weighted mode | 66 | -0,26403 | 0,804847 | 0,743926 | Sleep disorders | 42,80283 |
| ukb-b-5779 | eGFR decline (CKD population) [SH02] | Alcohol intake frequency. | MR Egger | 94 | -0,03393 | 0,271732 | 0,900899 | Alcohol consumption | 53,13815 |
| ukb-b-5779 | eGFR decline (CKD population) [SH02] | Alcohol intake frequency. | Weighted median | 94 | 0,044786 | 0,223183 | 0,840958 | Alcohol consumption | 53,13815 |
| ukb-b-5779 | eGFR decline (CKD population) [SH02] | Alcohol intake frequency. | Inverse variance weighted | 94 | -0,00903 | 0,120463 | 0,940263 | Alcohol consumption | 53,13815 |
| ukb-b-5779 | eGFR decline (CKD population) [SH02] | Alcohol intake frequency. | Simple mode | 94 | -0,32844 | 0,418076 | 0,434095 | Alcohol consumption | 53,13815 |
| ukb-b-5779 | eGFR decline (CKD population) [SH02] | Alcohol intake frequency. | Weighted mode | 94 | -0,05147 | 0,25684 | 0,84161 | Alcohol consumption | 53,13815 |
| ukb-b-7864 | eGFR decline (CKD population) [SH02] | Vitamin B6 | Wald ratio | 1 | -0,88575 | 0,848561 | 0,296565 | Vitamin | 32,75897 |
| ukb-d-30790_irnt | eGFR decline (CKD population) [SH02] | Lipoprotein A | MR Egger | 11 | -0,08818 | 0,187021 | 0,648511 | Dyslipidemias | 985,6435 |
| ukb-d-30790_irnt | eGFR decline (CKD population) [SH02] | Lipoprotein A | Weighted median | 11 | -0,20219 | 0,136626 | 0,138907 | Dyslipidemias | 985,6435 |
| ukb-d-30790_irnt | eGFR decline (CKD population) [SH02] | Lipoprotein A | Inverse variance weighted | 11 | -0,06412 | 0,120891 | 0,595828 | Dyslipidemias | 985,6435 |
| ukb-d-30790_irnt | eGFR decline (CKD population) [SH02] | Lipoprotein A | Simple mode | 11 | -0,20407 | 0,185548 | 0,297174 | Dyslipidemias | 985,6435 |
| ukb-d-30790_irnt | eGFR decline (CKD population) [SH02] | Lipoprotein A | Weighted mode | 11 | -0,19618 | 0,158128 | 0,24306 | Dyslipidemias | 985,6435 |
| BMI | eGFR decline (CKD population) [SH03] | Body mass index | MR Egger | 506 | 0,081932 | 0,205431 | 0,690188 | Obesity | 74,21321 |
| BMI | eGFR decline (CKD population) [SH03] | Body mass index | Weighted median | 506 | 0,059404 | 0,142317 | 0,676384 | Obesity | 74,21321 |
| BMI | eGFR decline (CKD population) [SH03] | Body mass index | Inverse variance weighted | 506 | 0,068329 | 0,078888 | 0,386407 | Obesity | 74,21321 |
| BMI | eGFR decline (CKD population) [SH03] | Body mass index | Simple mode | 506 | -0,15026 | 0,407952 | 0,712785 | Obesity | 74,21321 |
| BMI | eGFR decline (CKD population) [SH03] | Body mass index | Weighted mode | 506 | 0,091209 | 0,236784 | 0,700251 | Obesity | 74,21321 |

| | | | | | | | | | |
|---|---|---|---|---|---|---|---|---|---|
| DBPTang | eGFR decline (CKD population) [SH03] | Diastolic blood pressure | MR Egger | 251 | -0,10264 | 0,34279 | 0,764873 | Hypertension | 55,91029 |
| DBPTang | eGFR decline (CKD population) [SH03] | Diastolic blood pressure | Weighted median | 251 | -0,12205 | 0,157962 | 0,439709 | Hypertension | 55,91029 |
| DBPTang | eGFR decline (CKD population) [SH03] | Diastolic blood pressure | Inverse variance weighted | 251 | -0,07097 | 0,10648 | 0,50509 | Hypertension | 55,91029 |
| DBPTang | eGFR decline (CKD population) [SH03] | Diastolic blood pressure | Simple mode | 251 | 0,145371 | 0,456205 | 0,750256 | Hypertension | 55,91029 |
| DBPTang | eGFR decline (CKD population) [SH03] | Diastolic blood pressure | Weighted mode | 251 | -0,12665 | 0,372845 | 0,73437 | Hypertension | 55,91029 |
| FT3 | eGFR decline (CKD population) [SH03] | Free triiodothyronine | MR Egger | 6 | -0,46405 | 0,888654 | 0,629118 | Thyroid disease | 82,22035 |
| FT3 | eGFR decline (CKD population) [SH03] | Free triiodothyronine | Weighted median | 6 | -0,21845 | 0,250767 | 0,383682 | Thyroid disease | 82,22035 |
| FT3 | eGFR decline (CKD population) [SH03] | Free triiodothyronine | Inverse variance weighted | 6 | -0,1495 | 0,196329 | 0,446387 | Thyroid disease | 82,22035 |
| FT3 | eGFR decline (CKD population) [SH03] | Free triiodothyronine | Simple mode | 6 | -0,17737 | 0,345599 | 0,629665 | Thyroid disease | 82,22035 |
| FT3 | eGFR decline (CKD population) [SH03] | Free triiodothyronine | Weighted mode | 6 | -0,14619 | 0,326462 | 0,673025 | Thyroid disease | 82,22035 |
| FT3FT4 | eGFR decline (CKD population) [SH03] | FT3 FT4 ratio | MR Egger | 13 | 0,330273 | 1,12134 | 0,773834 | Thyroid disease | 132,3008 |
| FT3FT4 | eGFR decline (CKD population) [SH03] | FT3 FT4 ratio | Weighted median | 13 | -0,21078 | 0,766443 | 0,783312 | Thyroid disease | 132,3008 |
| FT3FT4 | eGFR decline (CKD population) [SH03] | FT3 FT4 ratio | Inverse variance weighted | 13 | -0,79365 | 0,55773 | 0,154734 | Thyroid disease | 132,3008 |
| FT3FT4 | eGFR decline (CKD population) [SH03] | FT3 FT4 ratio | Simple mode | 13 | 0,121469 | 1,340553 | 0,929297 | Thyroid disease | 132,3008 |
| FT3FT4 | eGFR decline (CKD population) [SH03] | FT3 FT4 ratio | Weighted mode | 13 | -0,26368 | 0,836225 | 0,757934 | Thyroid disease | 132,3008 |
| FT4 | eGFR decline (CKD population) [SH03] | Free thyroxine | MR Egger | 62 | -0,05851 | 0,175268 | 0,739689 | Thyroid disease | 84,3797 |
| FT4 | eGFR decline (CKD population) [SH03] | Free thyroxine | Weighted median | 62 | 0,027136 | 0,145668 | 0,852218 | Thyroid disease | 84,3797 |
| FT4 | eGFR decline (CKD population) [SH03] | Free thyroxine | Inverse variance weighted | 62 | -0,02298 | 0,083697 | 0,783665 | Thyroid disease | 84,3797 |
| FT4 | eGFR decline (CKD population) [SH03] | Free thyroxine | Simple mode | 62 | -0,1021 | 0,26969 | 0,706309 | Thyroid disease | 84,3797 |
| FT4 | eGFR decline (CKD population) [SH03] | Free thyroxine | Weighted mode | 62 | -0,01426 | 0,15409 | 0,92658 | Thyroid disease | 84,3797 |
| Fe | eGFR decline (CKD population) [SH03] | Serum iron | Inverse variance weighted | 2 | -0,23739 | 0,620139 | 0,701866 | Minerals | 291,9629 |
| HDL | eGFR decline (CKD population) [SH03] | HDL cholesterol | MR Egger | 233 | 0,015271 | 0,102321 | 0,881488 | Dyslipidemias | 268,9835 |
| HDL | eGFR decline (CKD population) [SH03] | HDL cholesterol | Weighted median | 233 | -0,03331 | 0,108173 | 0,758107 | Dyslipidemias | 268,9835 |
| HDL | eGFR decline (CKD population) [SH03] | HDL cholesterol | Inverse variance weighted | 233 | -0,0881 | 0,06601 | 0,181969 | Dyslipidemias | 268,9835 |
| HDL | eGFR decline (CKD population) [SH03] | HDL cholesterol | Simple mode | 233 | -0,13566 | 0,204949 | 0,508682 | Dyslipidemias | 268,9835 |
| HDL | eGFR decline (CKD population) [SH03] | HDL cholesterol | Weighted mode | 233 | -0,01999 | 0,103778 | 0,847412 | Dyslipidemias | 268,9835 |
| HiTSH | eGFR decline (CKD population) [SH03] | High TSH | MR Egger | 6 | 0,621222 | 0,569642 | 0,336763 | Thyroid disease | 49,53497 |
| HiTSH | eGFR decline (CKD population) [SH03] | High TSH | Weighted median | 6 | 0,304412 | 0,158435 | 0,054686 | Thyroid disease | 49,53497 |
| HiTSH | eGFR decline (CKD population) [SH03] | High TSH | Inverse variance weighted | 6 | 0,30826 | 0,128821 | 0,016715 | Thyroid disease | 49,53497 |
| HiTSH | eGFR decline (CKD population) [SH03] | High TSH | Simple mode | 6 | 0,325522 | 0,237344 | 0,228559 | Thyroid disease | 49,53497 |
| HiTSH | eGFR decline (CKD population) [SH03] | High TSH | Weighted mode | 6 | 0,316351 | 0,195096 | 0,165833 | Thyroid disease | 49,53497 |
| Insom | eGFR decline (CKD population) [SH03] | Insomnia | MR Egger | 13 | 0,029965 | 0,750129 | 0,968852 | Sleep disorders | 44,05724 |
| Insom | eGFR decline (CKD population) [SH03] | Insomnia | Weighted median | 13 | -0,20762 | 0,317472 | 0,513116 | Sleep disorders | 44,05724 |
| Insom | eGFR decline (CKD population) [SH03] | Insomnia | Inverse variance weighted | 13 | 0,080706 | 0,228815 | 0,724303 | Sleep disorders | 44,05724 |
| Insom | eGFR decline (CKD population) [SH03] | Insomnia | Simple mode | 13 | -0,35763 | 0,478796 | 0,469487 | Sleep disorders | 44,05724 |
| Insom | eGFR decline (CKD population) [SH03] | Insomnia | Weighted mode | 13 | -0,27369 | 0,443778 | 0,548943 | Sleep disorders | 44,05724 |
| LDL | eGFR decline (CKD population) [SH03] | LDL cholesterol | MR Egger | 217 | -0,07062 | 0,115998 | 0,543284 | Dyslipidemias | 221,0222 |
| LDL | eGFR decline (CKD population) [SH03] | LDL cholesterol | Weighted median | 217 | -0,08604 | 0,119332 | 0,470879 | Dyslipidemias | 221,0222 |
| LDL | eGFR decline (CKD population) [SH03] | LDL cholesterol | Inverse variance weighted | 217 | -0,07264 | 0,072494 | 0,316349 | Dyslipidemias | 221,0222 |
| LDL | eGFR decline (CKD population) [SH03] | LDL cholesterol | Simple mode | 217 | -0,19515 | 0,230529 | 0,398186 | Dyslipidemias | 221,0222 |
| LDL | eGFR decline (CKD population) [SH03] | LDL cholesterol | Weighted mode | 217 | -0,11539 | 0,114065 | 0,312852 | Dyslipidemias | 221,0222 |
| LoTSH | eGFR decline (CKD population) [SH03] | Low TSH | MR Egger | 26 | -0,01243 | 0,081316 | 0,879749 | Thyroid disease | 70,97091 |
| LoTSH | eGFR decline (CKD population) [SH03] | Low TSH | Weighted median | 26 | 0,024478 | 0,041688 | 0,557085 | Thyroid disease | 70,97091 |
| LoTSH | eGFR decline (CKD population) [SH03] | Low TSH | Inverse variance weighted | 26 | 0,022589 | 0,027995 | 0,419726 | Thyroid disease | 70,97091 |
| LoTSH | eGFR decline (CKD population) [SH03] | Low TSH | Simple mode | 26 | 0,065146 | 0,081379 | 0,430943 | Thyroid disease | 70,97091 |
| LoTSH | eGFR decline (CKD population) [SH03] | Low TSH | Weighted mode | 26 | -0,01601 | 0,062099 | 0,798661 | Thyroid disease | 70,97091 |
| PPTang | eGFR decline (CKD population) [SH03] | Pulse pressure | MR Egger | 300 | 0,133009 | 0,255646 | 0,603251 | Hypertension | 68,57434 |
| PPTang | eGFR decline (CKD population) [SH03] | Pulse pressure | Weighted median | 300 | 0,03953 | 0,153438 | 0,796694 | Hypertension | 68,57434 |
| PPTang | eGFR decline (CKD population) [SH03] | Pulse pressure | Inverse variance weighted | 300 | 0,126022 | 0,09498 | 0,184566 | Hypertension | 68,57434 |
| PPTang | eGFR decline (CKD population) [SH03] | Pulse pressure | Simple mode | 300 | -0,25248 | 0,409659 | 0,538161 | Hypertension | 68,57434 |
| PPTang | eGFR decline (CKD population) [SH03] | Pulse pressure | Weighted mode | 300 | -0,05532 | 0,293828 | 0,850793 | Hypertension | 68,57434 |
| SBPTang | eGFR decline (CKD population) [SH03] | Systolic blood pressure | MR Egger | 251 | -0,23937 | 0,340602 | 0,48284 | Hypertension | 56,96232 |
| SBPTang | eGFR decline (CKD population) [SH03] | Systolic blood pressure | Weighted median | 251 | -0,05148 | 0,161472 | 0,749871 | Hypertension | 56,96232 |
| SBPTang | eGFR decline (CKD population) [SH03] | Systolic blood pressure | Inverse variance weighted | 251 | -0,05641 | 0,109327 | 0,605846 | Hypertension | 56,96232 |
| SBPTang | eGFR decline (CKD population) [SH03] | Systolic blood pressure | Simple mode | 251 | 0,013795 | 0,434705 | 0,974709 | Hypertension | 56,96232 |
| SBPTang | eGFR decline (CKD population) [SH03] | Systolic blood pressure | Weighted mode | 251 | -0,07502 | 0,331261 | 0,821016 | Hypertension | 56,96232 |
| SIndex | eGFR decline (CKD population) [SH03] | Compound smoking index | MR Egger | 121 | 0,283727 | 1,019408 | 0,781246 | Smoking | 44,08236 |
| SIndex | eGFR decline (CKD population) [SH03] | Compound smoking index | Weighted median | 121 | -0,07408 | 0,366242 | 0,839701 | Smoking | 44,08236 |
| SIndex | eGFR decline (CKD population) [SH03] | Compound smoking index | Inverse variance weighted | 121 | -0,26335 | 0,254126 | 0,300072 | Smoking | 44,08236 |
| SIndex | eGFR decline (CKD population) [SH03] | Compound smoking index | Simple mode | 121 | -0,40434 | 0,940508 | 0,66803 | Smoking | 44,08236 |
| SIndex | eGFR decline (CKD population) [SH03] | Compound smoking index | Weighted mode | 121 | -0,09818 | 0,858508 | 0,909141 | Smoking | 44,08236 |
| T2DGGI | eGFR decline (CKD population) [SH03] | Type 2 diabetes | MR Egger | 441 | 0,12083 | 0,07105 | 0,08972 | Diabetes | 93,04757 |
| T2DGGI | eGFR decline (CKD population) [SH03] | Type 2 diabetes | Weighted median | 441 | 0,108424 | 0,067986 | 0,110755 | Diabetes | 93,04757 |
| T2DGGI | eGFR decline (CKD population) [SH03] | Type 2 diabetes | Inverse variance weighted | 441 | 0,082404 | 0,034317 | 0,01634 | Diabetes | 93,04757 |
| T2DGGI | eGFR decline (CKD population) [SH03] | Type 2 diabetes | Simple mode | 441 | 0,126323 | 0,146029 | 0,387478 | Diabetes | 93,04757 |
| T2DGGI | eGFR decline (CKD population) [SH03] | Type 2 diabetes | Weighted mode | 441 | 0,112431 | 0,079849 | 0,159825 | Diabetes | 93,04757 |
| TC | eGFR decline (CKD population) [SH03] | Total cholesterol | MR Egger | 264 | -0,06331 | 0,103846 | 0,542595 | Dyslipidemias | 246,1189 |
| TC | eGFR decline (CKD population) [SH03] | Total cholesterol | Weighted median | 264 | 0,054134 | 0,113778 | 0,634225 | Dyslipidemias | 246,1189 |
| TC | eGFR decline (CKD population) [SH03] | Total cholesterol | Inverse variance weighted | 264 | -0,00715 | 0,066116 | 0,913922 | Dyslipidemias | 246,1189 |
| TC | eGFR decline (CKD population) [SH03] | Total cholesterol | Simple mode | 264 | -0,02302 | 0,230968 | 0,920669 | Dyslipidemias | 246,1189 |
| TC | eGFR decline (CKD population) [SH03] | Total cholesterol | Weighted mode | 264 | -0,02302 | 0,113325 | 0,839157 | Dyslipidemias | 246,1189 |
| TG | eGFR decline (CKD population) [SH03] | Triglyceride | MR Egger | 219 | 0,128712 | 0,108611 | 0,237285 | Dyslipidemias | 231,1265 |
| TG | eGFR decline (CKD population) [SH03] | Triglyceride | Weighted median | 219 | 0,086006 | 0,123318 | 0,485531 | Dyslipidemias | 231,1265 |
| TG | eGFR decline (CKD population) [SH03] | Triglyceride | Inverse variance weighted | 219 | 0,046282 | 0,070037 | 0,508726 | Dyslipidemias | 231,1265 |
| TG | eGFR decline (CKD population) [SH03] | Triglyceride | Simple mode | 219 | -0,09147 | 0,218825 | 0,676367 | Dyslipidemias | 231,1265 |
| TG | eGFR decline (CKD population) [SH03] | Triglyceride | Weighted mode | 219 | 0,088926 | 0,11326 | 0,433215 | Dyslipidemias | 231,1265 |
| TSH | eGFR decline (CKD population) [SH03] | Thyroid Stimulating Hormone | MR Egger | 159 | 0,139666 | 0,100712 | 0,167474 | Thyroid disease | 154,4324 |
| TSH | eGFR decline (CKD population) [SH03] | Thyroid Stimulating Hormone | Weighted median | 159 | 0,102405 | 0,090803 | 0,259415 | Thyroid disease | 154,4324 |
| TSH | eGFR decline (CKD population) [SH03] | Thyroid Stimulating Hormone | Inverse variance weighted | 159 | 0,021215 | 0,055824 | 0,703919 | Thyroid disease | 154,4324 |
| TSH | eGFR decline (CKD population) [SH03] | Thyroid Stimulating Hormone | Simple mode | 159 | -0,02557 | 0,196944 | 0,896879 | Thyroid disease | 154,4324 |
| TSH | eGFR decline (CKD population) [SH03] | Thyroid Stimulating Hormone | Weighted mode | 159 | 0,165545 | 0,103397 | 0,111362 | Thyroid disease | 154,4324 |
| WHR | eGFR decline (CKD population) [SH03] | Waist-to-hip ratio adjusted BMI | MR Egger | 296 | -0,17361 | 0,217724 | 0,425861 | Obesity | 85,86852 |
| WHR | eGFR decline (CKD population) [SH03] | Waist-to-hip ratio adjusted BMI | Weighted median | 296 | -0,00877 | 0,157332 | 0,955552 | Obesity | 85,86852 |
| WHR | eGFR decline (CKD population) [SH03] | Waist-to-hip ratio adjusted BMI | Inverse variance weighted | 296 | 0,029414 | 0,091125 | 0,746855 | Obesity | 85,86852 |
| WHR | eGFR decline (CKD population) [SH03] | Waist-to-hip ratio adjusted BMI | Simple mode | 296 | -0,1835 | 0,369637 | 0,619965 | Obesity | 85,86852 |
| WHR | eGFR decline (CKD population) [SH03] | Waist-to-hip ratio adjusted BMI | Weighted mode | 296 | -0,02213 | 0,263077 | 0,933015 | Obesity | 85,86852 |
| Zn | eGFR decline (CKD population) [SH03] | Zinc | MR Egger | 3 | -0,1871 | 0,350934 | 0,688171 | Minerals | 99,95282 |
| Zn | eGFR decline (CKD population) [SH03] | Zinc | Weighted median | 3 | 0,018519 | 0,093345 | 0,842734 | Minerals | 99,95282 |
| Zn | eGFR decline (CKD population) [SH03] | Zinc | Inverse variance weighted | 3 | 0,060408 | 0,11377 | 0,59544 | Minerals | 99,95282 |
| Zn | eGFR decline (CKD population) [SH03] | Zinc | Simple mode | 3 | -0,01523 | 0,125673 | 0,914608 | Minerals | 99,95282 |
| Zn | eGFR decline (CKD population) [SH03] | Zinc | Weighted mode | 3 | 0,009568 | 0,092349 | 0,926933 | Minerals | 99,95282 |
| ebi-a-GCST006979-consort | eGFR decline (CKD population) [SH03] | Heel bone mineral density | MR Egger | 483 | 0,026734 | 0,101099 | 0,791559 | Bone disease | 151,4375 |
| ebi-a-GCST006979-consort | eGFR decline (CKD population) [SH03] | Heel bone mineral density | Weighted median | 483 | 0,132709 | 0,083973 | 0,11402 | Bone disease | 151,4375 |
| ebi-a-GCST006979-consort | eGFR decline (CKD population) [SH03] | Heel bone mineral density | Inverse variance weighted | 483 | 0,086771 | 0,052215 | 0,096549 | Bone disease | 151,4375 |
| ebi-a-GCST006979-consort | eGFR decline (CKD population) [SH03] | Heel bone mineral density | Simple mode | 483 | 0,160454 | 0,207752 | 0,440295 | Bone disease | 151,4375 |
| ebi-a-GCST006979-consort | eGFR decline (CKD population) [SH03] | Heel bone mineral density | Weighted mode | 483 | 0,13239 | 0,128035 | 0,30165 | Bone disease | 151,4375 |
| ebi-a-GCST90002227 | eGFR decline (CKD population) [SH03] | Two-hour glucose | MR Egger | 14 | -0,15939 | 0,341793 | 0,649337 | Diabetes | 56,47851 |
| ebi-a-GCST90002227 | eGFR decline (CKD population) [SH03] | Two-hour glucose | Weighted median | 14 | 0,203051 | 0,166613 | 0,22296 | Diabetes | 56,47851 |
| ebi-a-GCST90002227 | eGFR decline (CKD population) [SH03] | Two-hour glucose | Inverse variance weighted | 14 | 0,16046 | 0,126107 | 0,203226 | Diabetes | 56,47851 |
| ebi-a-GCST90002227 | eGFR decline (CKD population) [SH03] | Two-hour glucose | Simple mode | 14 | -0,03328 | 0,295397 | 0,912017 | Diabetes | 56,47851 |
| ebi-a-GCST90002227 | eGFR decline (CKD population) [SH03] | Two-hour glucose | Weighted mode | 14 | 0,239074 | 0,250787 | 0,357831 | Diabetes | 56,47851 |
| ebi-a-GCST90002232 | eGFR decline (CKD population) [SH03] | Fasting glucose | MR Egger | 68 | 0,563836 | 0,321413 | 0,084031 | Diabetes | 124,2211 |
| ebi-a-GCST90002232 | eGFR decline (CKD population) [SH03] | Fasting glucose | Weighted median | 68 | 0,549038 | 0,256812 | 0,032525 | Diabetes | 124,2211 |
| ebi-a-GCST90002232 | eGFR decline (CKD population) [SH03] | Fasting glucose | Inverse variance weighted | 68 | 0,199052 | 0,169776 | 0,241023 | Diabetes | 124,2211 |
| ebi-a-GCST90002232 | eGFR decline (CKD population) [SH03] | Fasting glucose | Simple mode | 68 | 0,463896 | 0,476406 | 0,333689 | Diabetes | 124,2211 |
| ebi-a-GCST90002232 | eGFR decline (CKD population) [SH03] | Fasting glucose | Weighted mode | 68 | 0,670922 | 0,251994 | 0,009704 | Diabetes | 124,2211 |
| ebi-a-GCST90002238 | eGFR decline (CKD population) [SH03] | Fasting insulin | MR Egger | 37 | 0,47131 | 1,172819 | 0,69023 | Diabetes | 52,15552 |
| ebi-a-GCST90002238 | eGFR decline (CKD population) [SH03] | Fasting insulin | Weighted median | 37 | 0,434546 | 0,469349 | 0,354525 | Diabetes | 52,15552 |
| ebi-a-GCST90002238 | eGFR decline (CKD population) [SH03] | Fasting insulin | Inverse variance weighted | 37 | 0,010458 | 0,35993 | 0,976821 | Diabetes | 52,15552 |
| ebi-a-GCST90002238 | eGFR decline (CKD population) [SH03] | Fasting insulin | Simple mode | 37 | 0,431982 | 0,855816 | 0,616803 | Diabetes | 52,15552 |
| ebi-a-GCST90002238 | eGFR decline (CKD population) [SH03] | Fasting insulin | Weighted mode | 37 | 0,431982 | 0,701972 | 0,542172 | Diabetes | 52,15552 |
| ebi-a-GCST90007310 | eGFR decline (CKD population) [SH03] | circulating leptin levels | MR Egger | 3 | 3,085538 | 2,333052 | 0,412154 | Obesity | 56,18818 |
| ebi-a-GCST90007310 | eGFR decline (CKD population) [SH03] | circulating leptin levels | Weighted median | 3 | -0,22049 | 0,352388 | 0,531518 | Obesity | 56,18818 |

| | | | | | | | | | |
|---|---|---|---|---|---|---|---|---|---|
| ebi-a-GCST90007310 | eGFR decline (CKD population) [SH03] | circulating leptin levels | Inverse variance weighted | 3 | -0,41302 | 0,312307 | 0,186009 | Obesity | 56,18818 |
| ebi-a-GCST90007310 | eGFR decline (CKD population) [SH03] | circulating leptin levels | Simple mode | 3 | -0,18316 | 0,435102 | 0,714713 | Obesity | 56,18818 |
| ebi-a-GCST90007310 | eGFR decline (CKD population) [SH03] | circulating leptin levels | Weighted mode | 3 | -0,18316 | 0,397969 | 0,690544 | Obesity | 56,18818 |
| ebi-a-GCST90011998 | eGFR decline (CKD population) [SH03] | Growth differentiation factor-15 levels | MR Egger | 3 | -0,17903 | 0,139141 | 0,4206 | Biomarkers | 294,0397 |
| ebi-a-GCST90011998 | eGFR decline (CKD population) [SH03] | Growth differentiation factor-15 levels | Weighted median | 3 | -0,14261 | 0,097722 | 0,144455 | Biomarkers | 294,0397 |
| ebi-a-GCST90011998 | eGFR decline (CKD population) [SH03] | Growth differentiation factor-15 levels | Inverse variance weighted | 3 | -0,13056 | 0,095202 | 0,170251 | Biomarkers | 294,0397 |
| ebi-a-GCST90011998 | eGFR decline (CKD population) [SH03] | Growth differentiation factor-15 levels | Simple mode | 3 | -0,17653 | 0,17632 | 0,422182 | Biomarkers | 294,0397 |
| ebi-a-GCST90011998 | eGFR decline (CKD population) [SH03] | Growth differentiation factor-15 levels | Weighted mode | 3 | -0,14996 | 0,102315 | 0,280363 | Biomarkers | 294,0397 |
| ebi-a-GCST90012022 | eGFR decline (CKD population) [SH03] | Fibroblast growth factor 23 levels | MR Egger | 8 | 0,021446 | 0,473853 | 0,96537 | Biomarkers | 43,42098 |
| ebi-a-GCST90012022 | eGFR decline (CKD population) [SH03] | Fibroblast growth factor 23 levels | Weighted median | 8 | 0,148498 | 0,167676 | 0,375821 | Biomarkers | 43,42098 |
| ebi-a-GCST90012022 | eGFR decline (CKD population) [SH03] | Fibroblast growth factor 23 levels | Inverse variance weighted | 8 | 0,030687 | 0,12803 | 0,810572 | Biomarkers | 43,42098 |
| ebi-a-GCST90012022 | eGFR decline (CKD population) [SH03] | Fibroblast growth factor 23 levels | Simple mode | 8 | 0,168582 | 0,298939 | 0,5904 | Biomarkers | 43,42098 |
| ebi-a-GCST90012022 | eGFR decline (CKD population) [SH03] | Fibroblast growth factor 23 levels | Weighted mode | 8 | 0,197489 | 0,254573 | 0,463287 | Biomarkers | 43,42098 |
| ebi-a-GCST90012041 | eGFR decline (CKD population) [SH03] | Kidney injury molecule 1 levels | MR Egger | 9 | -0,0257 | 0,104841 | 0,813402 | Biomarkers | 190,9807 |
| ebi-a-GCST90012041 | eGFR decline (CKD population) [SH03] | Kidney injury molecule 1 levels | Weighted median | 9 | -0,03598 | 0,068142 | 0,597523 | Biomarkers | 190,9807 |
| ebi-a-GCST90012041 | eGFR decline (CKD population) [SH03] | Kidney injury molecule 1 levels | Inverse variance weighted | 9 | -0,04564 | 0,062886 | 0,467992 | Biomarkers | 190,9807 |
| ebi-a-GCST90012041 | eGFR decline (CKD population) [SH03] | Kidney injury molecule 1 levels | Simple mode | 9 | 0,020861 | 0,146605 | 0,890364 | Biomarkers | 190,9807 |
| ebi-a-GCST90012041 | eGFR decline (CKD population) [SH03] | Kidney injury molecule 1 levels | Weighted mode | 9 | -0,03394 | 0,062534 | 0,602044 | Biomarkers | 190,9807 |
| ebi-a-GCST90013864 | eGFR decline (CKD population) [SH03] | Coronary artery disease (Firth correction) | MR Egger | 37 | -0,10077 | 0,136546 | 0,465435 | Cardiovascular disease | 59,8949 |
| ebi-a-GCST90013864 | eGFR decline (CKD population) [SH03] | Coronary artery disease (Firth correction) | Weighted median | 37 | -0,11266 | 0,089069 | 0,205923 | Cardiovascular disease | 59,8949 |
| ebi-a-GCST90013864 | eGFR decline (CKD population) [SH03] | Coronary artery disease (Firth correction) | Inverse variance weighted | 37 | -0,03066 | 0,061443 | 0,617802 | Cardiovascular disease | 59,8949 |
| ebi-a-GCST90013864 | eGFR decline (CKD population) [SH03] | Coronary artery disease (Firth correction) | Simple mode | 37 | -0,13241 | 0,180682 | 0,468417 | Cardiovascular disease | 59,8949 |
| ebi-a-GCST90013864 | eGFR decline (CKD population) [SH03] | Coronary artery disease (Firth correction) | Weighted mode | 37 | -0,13241 | 0,114011 | 0,253147 | Cardiovascular disease | 59,8949 |
| ebi-a-GCST90013975 | eGFR decline (CKD population) [SH03] | Body fat percentage | MR Egger | 316 | 0,439194 | 0,402678 | 0,276248 | Obesity | 56,17886 |
| ebi-a-GCST90013975 | eGFR decline (CKD population) [SH03] | Body fat percentage | Weighted median | 316 | -0,03793 | 0,18723 | 0,839475 | Obesity | 56,17886 |
| ebi-a-GCST90013975 | eGFR decline (CKD population) [SH03] | Body fat percentage | Inverse variance weighted | 316 | 0,072617 | 0,125015 | 0,561333 | Obesity | 56,17886 |
| ebi-a-GCST90013975 | eGFR decline (CKD population) [SH03] | Body fat percentage | Simple mode | 316 | 0,243704 | 0,574354 | 0,67163 | Obesity | 56,17886 |
| ebi-a-GCST90013975 | eGFR decline (CKD population) [SH03] | Body fat percentage | Weighted mode | 316 | -0,24941 | 0,40901 | 0,542434 | Obesity | 56,17886 |
| ebi-a-GCST90013989 | eGFR decline (CKD population) [SH03] | Sodium in urine | MR Egger | 41 | 1,057074 | 1,109953 | 0,34678 | Minerals | 43,81211 |
| ebi-a-GCST90013989 | eGFR decline (CKD population) [SH03] | Sodium in urine | Weighted median | 41 | -0,37881 | 0,393479 | 0,335693 | Minerals | 43,81211 |
| ebi-a-GCST90013989 | eGFR decline (CKD population) [SH03] | Sodium in urine | Inverse variance weighted | 41 | -0,19086 | 0,284976 | 0,503022 | Minerals | 43,81211 |
| ebi-a-GCST90013989 | eGFR decline (CKD population) [SH03] | Sodium in urine | Simple mode | 41 | -0,25192 | 0,88075 | 0,776331 | Minerals | 43,81211 |
| ebi-a-GCST90013989 | eGFR decline (CKD population) [SH03] | Sodium in urine | Weighted mode | 41 | -0,6224 | 0,810987 | 0,447315 | Minerals | 43,81211 |
| ebi-a-GCST90014006 | eGFR decline (CKD population) [SH03] | Glycated haemoglobin HbA1c levels | MR Egger | 302 | 0,247385 | 0,123761 | 0,046521 | Diabetes | 128,2905 |
| ebi-a-GCST90014006 | eGFR decline (CKD population) [SH03] | Glycated haemoglobin HbA1c levels | Weighted median | 302 | 0,161237 | 0,103073 | 0,117748 | Diabetes | 128,2905 |
| ebi-a-GCST90014006 | eGFR decline (CKD population) [SH03] | Glycated haemoglobin HbA1c levels | Inverse variance weighted | 302 | 0,056947 | 0,065201 | 0,382439 | Diabetes | 128,2905 |
| ebi-a-GCST90014006 | eGFR decline (CKD population) [SH03] | Glycated haemoglobin HbA1c levels | Simple mode | 302 | -0,0456 | 0,234472 | 0,845921 | Diabetes | 128,2905 |
| ebi-a-GCST90014006 | eGFR decline (CKD population) [SH03] | Glycated haemoglobin HbA1c levels | Weighted mode | 302 | 0,104683 | 0,139959 | 0,455072 | Diabetes | 128,2905 |
| ebi-a-GCST90014010 | eGFR decline (CKD population) [SH03] | Phosphate levels | MR Egger | 160 | 0,162736 | 0,171457 | 0,344002 | Minerals | 87,71201 |
| ebi-a-GCST90014010 | eGFR decline (CKD population) [SH03] | Phosphate levels | Weighted median | 160 | -0,02172 | 0,171907 | 0,899464 | Minerals | 87,71201 |
| ebi-a-GCST90014010 | eGFR decline (CKD population) [SH03] | Phosphate levels | Inverse variance weighted | 160 | -0,04839 | 0,096113 | 0,614599 | Minerals | 87,71201 |
| ebi-a-GCST90014010 | eGFR decline (CKD population) [SH03] | Phosphate levels | Simple mode | 160 | -0,04216 | 0,367728 | 0,908873 | Minerals | 87,71201 |
| ebi-a-GCST90014010 | eGFR decline (CKD population) [SH03] | Phosphate levels | Weighted mode | 160 | 0,097793 | 0,218853 | 0,655594 | Minerals | 87,71201 |
| ebi-a-GCST90014022 | eGFR decline (CKD population) [SH03] | Bone mineral density | MR Egger | 415 | 0,060393 | 0,111495 | 0,588345 | Bone disease | 120,0409 |
| ebi-a-GCST90014022 | eGFR decline (CKD population) [SH03] | Bone mineral density | Weighted median | 415 | 0,101009 | 0,090125 | 0,262387 | Bone disease | 120,0409 |
| ebi-a-GCST90014022 | eGFR decline (CKD population) [SH03] | Bone mineral density | Inverse variance weighted | 415 | 0,083536 | 0,056236 | 0,137418 | Bone disease | 120,0409 |
| ebi-a-GCST90014022 | eGFR decline (CKD population) [SH03] | Bone mineral density | Simple mode | 415 | 0,028482 | 0,227119 | 0,900265 | Bone disease | 120,0409 |
| ebi-a-GCST90014022 | eGFR decline (CKD population) [SH03] | Bone mineral density | Weighted mode | 415 | 0,06036 | 0,131903 | 0,647474 | Bone disease | 120,0409 |
| ebi-a-GCST90014023 | eGFR decline (CKD population) [SH03] | Type 1 diabetes | MR Egger | 76 | -0,02366 | 0,029181 | 0,420106 | Diabetes | 131,7111 |
| ebi-a-GCST90014023 | eGFR decline (CKD population) [SH03] | Type 1 diabetes | Weighted median | 76 | -0,02987 | 0,027024 | 0,268954 | Diabetes | 131,7111 |
| ebi-a-GCST90014023 | eGFR decline (CKD population) [SH03] | Type 1 diabetes | Inverse variance weighted | 76 | -0,01859 | 0,017792 | 0,29605 | Diabetes | 131,7111 |
| ebi-a-GCST90014023 | eGFR decline (CKD population) [SH03] | Type 1 diabetes | Simple mode | 76 | -0,04865 | 0,05161 | 0,348907 | Diabetes | 131,7111 |
| ebi-a-GCST90014023 | eGFR decline (CKD population) [SH03] | Type 1 diabetes | Weighted mode | 76 | -0,03605 | 0,027005 | 0,185882 | Diabetes | 131,7111 |
| ebi-a-GCST90016674 | eGFR decline (CKD population) [SH03] | Liver iron content | MR Egger | 5 | -0,07076 | 0,177969 | 0,717521 | Minerals | 257,3642 |
| ebi-a-GCST90016674 | eGFR decline (CKD population) [SH03] | Liver iron content | Weighted median | 5 | -0,07876 | 0,105322 | 0,454589 | Minerals | 257,3642 |
| ebi-a-GCST90016674 | eGFR decline (CKD population) [SH03] | Liver iron content | Inverse variance weighted | 5 | -0,074 | 0,102258 | 0,469272 | Minerals | 257,3642 |
| ebi-a-GCST90016674 | eGFR decline (CKD population) [SH03] | Liver iron content | Simple mode | 5 | -0,13361 | 0,197784 | 0,536368 | Minerals | 257,3642 |
| ebi-a-GCST90016674 | eGFR decline (CKD population) [SH03] | Liver iron content | Weighted mode | 5 | -0,07879 | 0,107175 | 0,503007 | Minerals | 257,3642 |
| ebi-a-GCST90016676 | eGFR decline (CKD population) [SH03] | Pancreas iron content | Wald ratio | 1 | -0,25689 | 0,552046 | 0,641688 | Minerals | 30,56027 |
| ebi-a-GCST90018806 | eGFR decline (CKD population) [SH03] | Chronic heart failure | MR Egger | 5 | 0,736026 | 1,317508 | 0,615388 | Cardiovascular disease | 41,81649 |
| ebi-a-GCST90018806 | eGFR decline (CKD population) [SH03] | Chronic heart failure | Weighted median | 5 | 0,231875 | 0,227078 | 0,307195 | Cardiovascular disease | 41,81649 |
| ebi-a-GCST90018806 | eGFR decline (CKD population) [SH03] | Chronic heart failure | Inverse variance weighted | 5 | 0,112557 | 0,190021 | 0,553622 | Cardiovascular disease | 41,81649 |
| ebi-a-GCST90018806 | eGFR decline (CKD population) [SH03] | Chronic heart failure | Simple mode | 5 | 0,300537 | 0,315098 | 0,394202 | Cardiovascular disease | 41,81649 |
| ebi-a-GCST90018806 | eGFR decline (CKD population) [SH03] | Chronic heart failure | Weighted mode | 5 | 0,304625 | 0,300234 | 0,367669 | Cardiovascular disease | 41,81649 |
| ebi-a-GCST90018860 | eGFR decline (CKD population) [SH03] | Hyperthyroidism | MR Egger | 8 | -0,15199 | 0,27958 | 0,606281 | Thyroid disease | 53,27573 |
| ebi-a-GCST90018860 | eGFR decline (CKD population) [SH03] | Hyperthyroidism | Weighted median | 8 | 0,15632 | 0,082813 | 0,059076 | Thyroid disease | 53,27573 |
| ebi-a-GCST90018860 | eGFR decline (CKD population) [SH03] | Hyperthyroidism | Inverse variance weighted | 8 | 0,080036 | 0,078879 | 0,310266 | Thyroid disease | 53,27573 |
| ebi-a-GCST90018860 | eGFR decline (CKD population) [SH03] | Hyperthyroidism | Simple mode | 8 | 0,240733 | 0,148774 | 0,149672 | Thyroid disease | 53,27573 |
| ebi-a-GCST90018860 | eGFR decline (CKD population) [SH03] | Hyperthyroidism | Weighted mode | 8 | 0,185445 | 0,148806 | 0,252771 | Thyroid disease | 53,27573 |
| ebi-a-GCST90018862 | eGFR decline (CKD population) [SH03] | Hypothyroidism | MR Egger | 64 | -0,046 | 0,091429 | 0,616648 | Thyroid disease | 82,13671 |
| ebi-a-GCST90018862 | eGFR decline (CKD population) [SH03] | Hypothyroidism | Weighted median | 64 | 0,024753 | 0,060232 | 0,681098 | Thyroid disease | 82,13671 |
| ebi-a-GCST90018862 | eGFR decline (CKD population) [SH03] | Hypothyroidism | Inverse variance weighted | 64 | 0,037804 | 0,039401 | 0,337327 | Thyroid disease | 82,13671 |
| ebi-a-GCST90018862 | eGFR decline (CKD population) [SH03] | Hypothyroidism | Simple mode | 64 | 0,072718 | 0,122644 | 0,555357 | Thyroid disease | 82,13671 |
| ebi-a-GCST90018862 | eGFR decline (CKD population) [SH03] | Hypothyroidism | Weighted mode | 64 | -0,00233 | 0,08581 | 0,978432 | Thyroid disease | 82,13671 |
| ebi-a-GCST90018872 | eGFR decline (CKD population) [SH03] | Iron deficiency anemia | Wald ratio | 1 | 0,676297 | 0,404689 | 0,094692 | Minerals | 34,53822 |
| ebi-a-GCST90018877 | eGFR decline (CKD population) [SH03] | Myocardial infarction | MR Egger | 70 | -0,12135 | 0,097554 | 0,217815 | Cardiovascular disease | 66,68595 |
| ebi-a-GCST90018877 | eGFR decline (CKD population) [SH03] | Myocardial infarction | Weighted median | 70 | -0,04386 | 0,065763 | 0,50479 | Cardiovascular disease | 66,68595 |
| ebi-a-GCST90018877 | eGFR decline (CKD population) [SH03] | Myocardial infarction | Inverse variance weighted | 70 | 0,020888 | 0,043618 | 0,63202 | Cardiovascular disease | 66,68595 |
| ebi-a-GCST90018877 | eGFR decline (CKD population) [SH03] | Myocardial infarction | Simple mode | 70 | -0,00089 | 0,131675 | 0,994609 | Cardiovascular disease | 66,68595 |
| ebi-a-GCST90018877 | eGFR decline (CKD population) [SH03] | Myocardial infarction | Weighted mode | 70 | -0,06618 | 0,079039 | 0,405289 | Cardiovascular disease | 66,68595 |
| ebi-a-GCST90018916 | eGFR decline (CKD population) [SH03] | Sleep apnea syndrome | Wald ratio | 1 | -0,09655 | 0,243965 | 0,692274 | Sleep disorders | 63,50864 |
| ebi-a-GCST90018977 | eGFR decline (CKD population) [SH03] | Serum uric acid levels | MR Egger | 237 | -0,0036 | 0,157613 | 0,981817 | Uricemia | 110,1094 |
| ebi-a-GCST90018977 | eGFR decline (CKD population) [SH03] | Serum uric acid levels | Weighted median | 237 | 0,116966 | 0,145711 | 0,422132 | Uricemia | 110,1094 |
| ebi-a-GCST90018977 | eGFR decline (CKD population) [SH03] | Serum uric acid levels | Inverse variance weighted | 237 | 0,091641 | 0,087737 | 0,296257 | Uricemia | 110,1094 |
| ebi-a-GCST90018977 | eGFR decline (CKD population) [SH03] | Serum uric acid levels | Simple mode | 237 | 0,395387 | 0,334522 | 0,238417 | Uricemia | 110,1094 |
| ebi-a-GCST90018977 | eGFR decline (CKD population) [SH03] | Serum uric acid levels | Weighted mode | 237 | 0,112666 | 0,167938 | 0,502955 | Uricemia | 110,1094 |
| ebi-a-GCST90019417 | eGFR decline (CKD population) [SH03] | Vitamin K-dependent protein S measurement | MR Egger | 4 | 0,050207 | 0,158096 | 0,7809 | Vitamin | 118,3171 |
| ebi-a-GCST90019417 | eGFR decline (CKD population) [SH03] | Vitamin K-dependent protein S measurement | Weighted median | 4 | 0,023915 | 0,09954 | 0,810132 | Vitamin | 118,3171 |
| ebi-a-GCST90019417 | eGFR decline (CKD population) [SH03] | Vitamin K-dependent protein S measurement | Inverse variance weighted | 4 | 0,0138 | 0,090614 | 0,878954 | Vitamin | 118,3171 |
| ebi-a-GCST90019417 | eGFR decline (CKD population) [SH03] | Vitamin K-dependent protein S measurement | Simple mode | 4 | -0,01813 | 0,139443 | 0,904788 | Vitamin | 118,3171 |
| ebi-a-GCST90019417 | eGFR decline (CKD population) [SH03] | Vitamin K-dependent protein S measurement | Weighted mode | 4 | 0,035299 | 0,112962 | 0,775132 | Vitamin | 118,3171 |
| ebi-a-GCST90019423 | eGFR decline (CKD population) [SH03] | Vitamin K-dependent protein C measurement | MR Egger | 3 | 0,082078 | 0,24324 | 0,792819 | Vitamin | 207,8561 |
| ebi-a-GCST90019423 | eGFR decline (CKD population) [SH03] | Vitamin K-dependent protein C measurement | Weighted median | 3 | 0,070857 | 0,072452 | 0,328087 | Vitamin | 207,8561 |
| ebi-a-GCST90019423 | eGFR decline (CKD population) [SH03] | Vitamin K-dependent protein C measurement | Inverse variance weighted | 3 | 0,042124 | 0,11049 | 0,70302 | Vitamin | 207,8561 |
| ebi-a-GCST90019423 | eGFR decline (CKD population) [SH03] | Vitamin K-dependent protein C measurement | Simple mode | 3 | 0,025769 | 0,187358 | 0,903203 | Vitamin | 207,8561 |
| ebi-a-GCST90019423 | eGFR decline (CKD population) [SH03] | Vitamin K-dependent protein C measurement | Weighted mode | 3 | 0,07783 | 0,070388 | 0,384057 | Vitamin | 207,8561 |
| ebi-a-GCST90025947 | eGFR decline (CKD population) [SH03] | Serum alkaline phosphatase levels | MR Egger | 335 | -0,07548 | 0,102567 | 0,462283 | Minerals | 166,037 |
| ebi-a-GCST90025947 | eGFR decline (CKD population) [SH03] | Serum alkaline phosphatase levels | Weighted median | 335 | 0,108673 | 0,103659 | 0,29447 | Minerals | 166,037 |
| ebi-a-GCST90025947 | eGFR decline (CKD population) [SH03] | Serum alkaline phosphatase levels | Inverse variance weighted | 335 | -0,01583 | 0,060099 | 0,792236 | Minerals | 166,037 |
| ebi-a-GCST90025947 | eGFR decline (CKD population) [SH03] | Serum alkaline phosphatase levels | Simple mode | 335 | 0,149922 | 0,229855 | 0,51469 | Minerals | 166,037 |
| ebi-a-GCST90025947 | eGFR decline (CKD population) [SH03] | Serum alkaline phosphatase levels | Weighted mode | 335 | 0,096905 | 0,125217 | 0,439538 | Minerals | 166,037 |
| ebi-a-GCST90025967 | eGFR decline (CKD population) [SH03] | Vitamin D levels | MR Egger | 81 | -0,06423 | 0,191536 | 0,738241 | Vitamin | 134,1402 |
| ebi-a-GCST90025967 | eGFR decline (CKD population) [SH03] | Vitamin D levels | Weighted median | 81 | -0,22566 | 0,19449 | 0,245946 | Vitamin | 134,1402 |
| ebi-a-GCST90025967 | eGFR decline (CKD population) [SH03] | Vitamin D levels | Inverse variance weighted | 81 | -0,0872 | 0,117562 | 0,458241 | Vitamin | 134,1402 |
| ebi-a-GCST90025967 | eGFR decline (CKD population) [SH03] | Vitamin D levels | Simple mode | 81 | -0,10144 | 0,339994 | 0,766203 | Vitamin | 134,1402 |
| ebi-a-GCST90025967 | eGFR decline (CKD population) [SH03] | Vitamin D levels | Weighted mode | 81 | -0,12495 | 0,185462 | 0,502432 | Vitamin | 134,1402 |
| ebi-a-GCST90025969 | eGFR decline (CKD population) [SH03] | Hemoglobin levels | MR Egger | 325 | 0,003815 | 0,139264 | 0,978162 | Organic compound | 104,3556 |
| ebi-a-GCST90025969 | eGFR decline (CKD population) [SH03] | Hemoglobin levels | Weighted median | 325 | -0,07951 | 0,124858 | 0,524254 | Organic compound | 104,3556 |
| ebi-a-GCST90025969 | eGFR decline (CKD population) [SH03] | Hemoglobin levels | Inverse variance weighted | 325 | -0,10629 | 0,071708 | 0,138255 | Organic compound | 104,3556 |
| ebi-a-GCST90025969 | eGFR decline (CKD population) [SH03] | Hemoglobin levels | Simple mode | 325 | 0,218469 | 0,304224 | 0,473202 | Organic compound | 104,3556 |
| ebi-a-GCST90025969 | eGFR decline (CKD population) [SH03] | Hemoglobin levels | Weighted mode | 325 | -0,0658 | 0,154827 | 0,671149 | Organic compound | 104,3556 |
| ebi-a-GCST90025986 | eGFR decline (CKD population) [SH03] | Blood glucose levels | MR Egger | 109 | 0,19226 | 0,190583 | 0,315347 | Diabetes | 118,7554 |
| ebi-a-GCST90025986 | eGFR decline (CKD population) [SH03] | Blood glucose levels | Weighted median | 109 | 0,381159 | 0,169276 | 0,024341 | Diabetes | 118,7554 |

| | | | | | | | | | |
|---|---|---|---|---|---|---|---|---|---|
| ebi-a-GCST90025986 | eGFR decline (CKD population) [SH03] | Blood glucose levels | Inverse variance weighted | 109 | 0,01863 | 0,107203 | 0,862038 | Diabetes | 118,7554 |
| ebi-a-GCST90025986 | eGFR decline (CKD population) [SH03] | Blood glucose levels | Simple mode | 109 | 0,281606 | 0,392471 | 0,474602 | Diabetes | 118,7554 |
| ebi-a-GCST90025986 | eGFR decline (CKD population) [SH03] | Blood glucose levels | Weighted mode | 109 | 0,329387 | 0,163432 | 0,046342 | Diabetes | 118,7554 |
| ebi-a-GCST90025989 | eGFR decline (CKD population) [SH03] | Insulin-like growth factor 1 levels | MR Egger | 373 | -0,19427 | 0,122156 | 0,112609 | Diabetes | 115,9356 |
| ebi-a-GCST90025989 | eGFR decline (CKD population) [SH03] | Insulin-like growth factor 1 levels | Weighted median | 373 | -0,00257 | 0,104455 | 0,980371 | Diabetes | 115,9356 |
| ebi-a-GCST90025989 | eGFR decline (CKD population) [SH03] | Insulin-like growth factor 1 levels | Inverse variance weighted | 373 | 0,011569 | 0,063177 | 0,854702 | Diabetes | 115,9356 |
| ebi-a-GCST90025989 | eGFR decline (CKD population) [SH03] | Insulin-like growth factor 1 levels | Simple mode | 373 | -0,10201 | 0,248525 | 0,681691 | Diabetes | 115,9356 |
| ebi-a-GCST90025989 | eGFR decline (CKD population) [SH03] | Insulin-like growth factor 1 levels | Weighted mode | 373 | -0,13344 | 0,139558 | 0,33962 | Diabetes | 115,9356 |
| ebi-a-GCST90025990 | eGFR decline (CKD population) [SH03] | Calcium levels | MR Egger | 202 | 0,29374 | 0,154972 | 0,059477 | Minerals | 102,0722 |
| ebi-a-GCST90025990 | eGFR decline (CKD population) [SH03] | Calcium levels | Weighted median | 202 | 0,100617 | 0,17098 | 0,556216 | Minerals | 102,0722 |
| ebi-a-GCST90025990 | eGFR decline (CKD population) [SH03] | Calcium levels | Inverse variance weighted | 202 | 0,062961 | 0,083696 | 0,451896 | Minerals | 102,0722 |
| ebi-a-GCST90025990 | eGFR decline (CKD population) [SH03] | Calcium levels | Simple mode | 202 | -0,01808 | 0,329784 | 0,956337 | Minerals | 102,0722 |
| ebi-a-GCST90025990 | eGFR decline (CKD population) [SH03] | Calcium levels | Weighted mode | 202 | 0,113873 | 0,172813 | 0,51069 | Minerals | 102,0722 |
| ebi-a-GCST90029013 | eGFR decline (CKD population) [SH03] | Educational attainment (years of education) | MR Egger | 209 | 0,023976 | 0,115619 | 0,835925 | Social trait | 45,05626 |
| ebi-a-GCST90029013 | eGFR decline (CKD population) [SH03] | Educational attainment (years of education) | Weighted median | 209 | -0,00907 | 0,039288 | 0,817356 | Social trait | 45,05626 |
| ebi-a-GCST90029013 | eGFR decline (CKD population) [SH03] | Educational attainment (years of education) | Inverse variance weighted | 209 | -0,03989 | 0,026471 | 0,131841 | Social trait | 45,05626 |
| ebi-a-GCST90029013 | eGFR decline (CKD population) [SH03] | Educational attainment (years of education) | Simple mode | 209 | 0,005516 | 0,113159 | 0,961167 | Social trait | 45,05626 |
| ebi-a-GCST90029013 | eGFR decline (CKD population) [SH03] | Educational attainment (years of education) | Weighted mode | 209 | 0,042309 | 0,097817 | 0,665805 | Social trait | 45,05626 |
| ebi-a-GCST90029014 | eGFR decline (CKD population) [SH03] | Smoking status | MR Egger | 117 | -0,39371 | 1,104631 | 0,722182 | Smoking | 42,91926 |
| ebi-a-GCST90029014 | eGFR decline (CKD population) [SH03] | Smoking status | Weighted median | 117 | 0,005551 | 0,38479 | 0,988491 | Smoking | 42,91926 |
| ebi-a-GCST90029014 | eGFR decline (CKD population) [SH03] | Smoking status | Inverse variance weighted | 117 | -0,01459 | 0,261826 | 0,955573 | Smoking | 42,91926 |
| ebi-a-GCST90029014 | eGFR decline (CKD population) [SH03] | Smoking status | Simple mode | 117 | -0,49582 | 0,88406 | 0,575987 | Smoking | 42,91926 |
| ebi-a-GCST90029014 | eGFR decline (CKD population) [SH03] | Smoking status | Weighted mode | 117 | -0,30523 | 0,73808 | 0,679973 | Smoking | 42,91926 |
| ebi-a-GCST90029070 | eGFR decline (CKD population) [SH03] | C-reactive protein levels | MR Egger | 214 | 0,272741 | 0,131404 | 0,039138 | Inflammation | 122,6906 |
| ebi-a-GCST90029070 | eGFR decline (CKD population) [SH03] | C-reactive protein levels | Weighted median | 214 | 0,077494 | 0,125432 | 0,536698 | Inflammation | 122,6906 |
| ebi-a-GCST90029070 | eGFR decline (CKD population) [SH03] | C-reactive protein levels | Inverse variance weighted | 214 | 0,095501 | 0,077518 | 0,217954 | Inflammation | 122,6906 |
| ebi-a-GCST90029070 | eGFR decline (CKD population) [SH03] | C-reactive protein levels | Simple mode | 214 | -0,36721 | 0,282086 | 0,194397 | Inflammation | 122,6906 |
| ebi-a-GCST90029070 | eGFR decline (CKD population) [SH03] | C-reactive protein levels | Weighted mode | 214 | 0,20219 | 0,136083 | 0,138814 | Inflammation | 122,6906 |
| ebi-a-GCST90029073 | eGFR decline (CKD population) [SH03] | Liver fat | MR Egger | 8 | 0,290276 | 0,161078 | 0,121601 | Fatty liver | 140,8023 |
| ebi-a-GCST90029073 | eGFR decline (CKD population) [SH03] | Liver fat | Weighted median | 8 | 0,181279 | 0,117086 | 0,121559 | Fatty liver | 140,8023 |
| ebi-a-GCST90029073 | eGFR decline (CKD population) [SH03] | Liver fat | Inverse variance weighted | 8 | 0,183685 | 0,097672 | 0,060023 | Fatty liver | 140,8023 |
| ebi-a-GCST90029073 | eGFR decline (CKD population) [SH03] | Liver fat | Simple mode | 8 | 0,241447 | 0,18426 | 0,23143 | Fatty liver | 140,8023 |
| ebi-a-GCST90029073 | eGFR decline (CKD population) [SH03] | Liver fat | Weighted mode | 8 | 0,229539 | 0,119228 | 0,095594 | Fatty liver | 140,8023 |
| ebi-a-GCST90086057 | eGFR decline (CKD population) [SH03] | Cardiovascular disease | Inverse variance weighted | 2 | 0,124873 | 0,213666 | 0,558931 | Cardiovascular disease | 32,018 |
| finn-b-D3_ANAEMIA_B12_DEF | eGFR decline (CKD population) [SH03] | Vitamin B12 deficiency anaemia | MR Egger | 4 | -0,07186 | 0,156427 | 0,691046 | Vitamin | 52,61875 |
| finn-b-D3_ANAEMIA_B12_DEF | eGFR decline (CKD population) [SH03] | Vitamin B12 deficiency anaemia | Weighted median | 4 | -0,05554 | 0,061359 | 0,365407 | Vitamin | 52,61875 |
| finn-b-D3_ANAEMIA_B12_DEF | eGFR decline (CKD population) [SH03] | Vitamin B12 deficiency anaemia | Inverse variance weighted | 4 | -0,0278 | 0,054539 | 0,610211 | Vitamin | 52,61875 |
| finn-b-D3_ANAEMIA_B12_DEF | eGFR decline (CKD population) [SH03] | Vitamin B12 deficiency anaemia | Simple mode | 4 | -0,06792 | 0,086637 | 0,490266 | Vitamin | 52,61875 |
| finn-b-D3_ANAEMIA_B12_DEF | eGFR decline (CKD population) [SH03] | Vitamin B12 deficiency anaemia | Weighted mode | 4 | -0,07106 | 0,073748 | 0,406366 | Vitamin | 52,61875 |
| finn-b-E4_OBESITY | eGFR decline (CKD population) [SH03] | Obesity | MR Egger | 7 | -0,045 | 0,293306 | 0,884069 | Obesity | 59,53804 |
| finn-b-E4_OBESITY | eGFR decline (CKD population) [SH03] | Obesity | Weighted median | 7 | -0,02513 | 0,103456 | 0,808064 | Obesity | 59,53804 |
| finn-b-E4_OBESITY | eGFR decline (CKD population) [SH03] | Obesity | Inverse variance weighted | 7 | 0,02746 | 0,08064 | 0,733459 | Obesity | 59,53804 |
| finn-b-E4_OBESITY | eGFR decline (CKD population) [SH03] | Obesity | Simple mode | 7 | -0,00414 | 0,15996 | 0,98018 | Obesity | 59,53804 |
| finn-b-E4_OBESITY | eGFR decline (CKD population) [SH03] | Obesity | Weighted mode | 7 | -0,05365 | 0,102807 | 0,620453 | Obesity | 59,53804 |
| finn-b-FG_PULMHEART | eGFR decline (CKD population) [SH03] | Pulmonary heart disease | MR Egger | 5 | -0,22417 | 0,154196 | 0,241965 | Cardiovascular disease | 89,00197 |
| finn-b-FG_PULMHEART | eGFR decline (CKD population) [SH03] | Pulmonary heart disease | Weighted median | 5 | 0,075549 | 0,066255 | 0,254166 | Cardiovascular disease | 89,00197 |
| finn-b-FG_PULMHEART | eGFR decline (CKD population) [SH03] | Pulmonary heart disease | Inverse variance weighted | 5 | 0,013476 | 0,056858 | 0,812641 | Cardiovascular disease | 89,00197 |
| finn-b-FG_PULMHEART | eGFR decline (CKD population) [SH03] | Pulmonary heart disease | Simple mode | 5 | 0,109956 | 0,106712 | 0,361046 | Cardiovascular disease | 89,00197 |
| finn-b-FG_PULMHEART | eGFR decline (CKD population) [SH03] | Pulmonary heart disease | Weighted mode | 5 | 0,109956 | 0,11077 | 0,377068 | Cardiovascular disease | 89,00197 |
| ieu-b-102 | eGFR decline (CKD population) [SH03] | Major depression | MR Egger | 48 | 0,258806 | 0,821908 | 0,754273 | Mental health | 38,29386 |
| ieu-b-102 | eGFR decline (CKD population) [SH03] | Major depression | Weighted median | 48 | -0,04893 | 0,20813 | 0,814143 | Mental health | 38,29386 |
| ieu-b-102 | eGFR decline (CKD population) [SH03] | Major depression | Inverse variance weighted | 48 | -0,09505 | 0,146942 | 0,517729 | Mental health | 38,29386 |
| ieu-b-102 | eGFR decline (CKD population) [SH03] | Major depression | Simple mode | 48 | 0,118913 | 0,474886 | 0,803366 | Mental health | 38,29386 |
| ieu-b-102 | eGFR decline (CKD population) [SH03] | Major depression | Weighted mode | 48 | 0,055081 | 0,44629 | 0,902301 | Mental health | 38,29386 |
| ieu-b-107 | eGFR decline (CKD population) [SH03] | apolipoprotein A-I | MR Egger | 269 | -0,1107 | 0,114196 | 0,333245 | Dyslipidemias | 139,7111 |
| ieu-b-107 | eGFR decline (CKD population) [SH03] | apolipoprotein A-I | Weighted median | 269 | -0,10107 | 0,11385 | 0,374678 | Dyslipidemias | 139,7111 |
| ieu-b-107 | eGFR decline (CKD population) [SH03] | apolipoprotein A-I | Inverse variance weighted | 269 | -0,04455 | 0,067558 | 0,509651 | Dyslipidemias | 139,7111 |
| ieu-b-107 | eGFR decline (CKD population) [SH03] | apolipoprotein A-I | Simple mode | 269 | -0,12626 | 0,240514 | 0,600052 | Dyslipidemias | 139,7111 |
| ieu-b-107 | eGFR decline (CKD population) [SH03] | apolipoprotein A-I | Weighted mode | 269 | -0,12626 | 0,123835 | 0,308854 | Dyslipidemias | 139,7111 |
| ieu-b-108 | eGFR decline (CKD population) [SH03] | apolipoprotein B | MR Egger | 173 | 0,071309 | 0,124215 | 0,566669 | Dyslipidemias | 162,3296 |
| ieu-b-108 | eGFR decline (CKD population) [SH03] | apolipoprotein B | Weighted median | 173 | 0,152093 | 0,116085 | 0,190132 | Dyslipidemias | 162,3296 |
| ieu-b-108 | eGFR decline (CKD population) [SH03] | apolipoprotein B | Inverse variance weighted | 173 | 0,091521 | 0,076488 | 0,231488 | Dyslipidemias | 162,3296 |
| ieu-b-108 | eGFR decline (CKD population) [SH03] | apolipoprotein B | Simple mode | 173 | 0,157636 | 0,246203 | 0,522851 | Dyslipidemias | 162,3296 |
| ieu-b-108 | eGFR decline (CKD population) [SH03] | apolipoprotein B | Weighted mode | 173 | 0,157636 | 0,128153 | 0,220356 | Dyslipidemias | 162,3296 |
| ieu-b-25 | eGFR decline (CKD population) [SH03] | Cigarettes per Day | MR Egger | 22 | -0,25472 | 0,173937 | 0,158608 | Smoking | 99,80012 |
| ieu-b-25 | eGFR decline (CKD population) [SH03] | Cigarettes per Day | Weighted median | 22 | -0,18434 | 0,14773 | 0,212094 | Smoking | 99,80012 |
| ieu-b-25 | eGFR decline (CKD population) [SH03] | Cigarettes per Day | Inverse variance weighted | 22 | -0,16864 | 0,100082 | 0,09199 | Smoking | 99,80012 |
| ieu-b-25 | eGFR decline (CKD population) [SH03] | Cigarettes per Day | Simple mode | 22 | -0,49665 | 0,302363 | 0,115361 | Smoking | 99,80012 |
| ieu-b-25 | eGFR decline (CKD population) [SH03] | Cigarettes per Day | Weighted mode | 22 | -0,16184 | 0,134722 | 0,243016 | Smoking | 99,80012 |
| ieu-b-4861 | eGFR decline (CKD population) [SH03] | Chronotype | MR Egger | 60 | -0,45249 | 0,376017 | 0,233721 | Sleep disorders | 44,49296 |
| ieu-b-4861 | eGFR decline (CKD population) [SH03] | Chronotype | Weighted median | 60 | -0,10658 | 0,20145 | 0,596759 | Sleep disorders | 44,49296 |
| ieu-b-4861 | eGFR decline (CKD population) [SH03] | Chronotype | Inverse variance weighted | 60 | -0,00264 | 0,135936 | 0,984515 | Sleep disorders | 44,49296 |
| ieu-b-4861 | eGFR decline (CKD population) [SH03] | Chronotype | Simple mode | 60 | -0,07317 | 0,516987 | 0,887939 | Sleep disorders | 44,49296 |
| ieu-b-4861 | eGFR decline (CKD population) [SH03] | Chronotype | Weighted mode | 60 | -0,13927 | 0,398876 | 0,72822 | Sleep disorders | 44,49296 |
| ieu-b-4877 | eGFR decline (CKD population) [SH03] | smoking initiation | MR Egger | 85 | 1,542099 | 0,604846 | 0,012624 | Smoking | 42,2587 |
| ieu-b-4877 | eGFR decline (CKD population) [SH03] | smoking initiation | Weighted median | 85 | 0,132912 | 0,17485 | 0,447166 | Smoking | 42,2587 |
| ieu-b-4877 | eGFR decline (CKD population) [SH03] | smoking initiation | Inverse variance weighted | 85 | 0,176953 | 0,118965 | 0,136898 | Smoking | 42,2587 |
| ieu-b-4877 | eGFR decline (CKD population) [SH03] | smoking initiation | Simple mode | 85 | 0,292216 | 0,412571 | 0,480732 | Smoking | 42,2587 |
| ieu-b-4877 | eGFR decline (CKD population) [SH03] | smoking initiation | Weighted mode | 85 | 0,239158 | 0,402712 | 0,554196 | Smoking | 42,2587 |
| ieu-b-5144 | eGFR decline (CKD population) [SH03] | Hypertension | MR Egger | 187 | -1,48555 | 0,901162 | 0,10095 | Hypertension | 54,118 |
| ieu-b-5144 | eGFR decline (CKD population) [SH03] | Hypertension | Weighted median | 187 | -0,54021 | 0,433392 | 0,212593 | Hypertension | 54,118 |
| ieu-b-5144 | eGFR decline (CKD population) [SH03] | Hypertension | Inverse variance weighted | 187 | -0,47851 | 0,289923 | 0,098848 | Hypertension | 54,118 |
| ieu-b-5144 | eGFR decline (CKD population) [SH03] | Hypertension | Simple mode | 187 | -1,14278 | 1,234154 | 0,355668 | Hypertension | 54,118 |
| ieu-b-5144 | eGFR decline (CKD population) [SH03] | Hypertension | Weighted mode | 187 | -1,41927 | 0,954554 | 0,138751 | Hypertension | 54,118 |
| ieu-b-72 | eGFR decline (CKD population) [SH03] | Urinary sodium-potassium ratio | MR Egger | 24 | 0,885593 | 1,598996 | 0,585272 | Minerals | 39,88328 |
| ieu-b-72 | eGFR decline (CKD population) [SH03] | Urinary sodium-potassium ratio | Weighted median | 24 | 0,225569 | 0,490764 | 0,645784 | Minerals | 39,88328 |
| ieu-b-72 | eGFR decline (CKD population) [SH03] | Urinary sodium-potassium ratio | Inverse variance weighted | 24 | 0,33625 | 0,368539 | 0,361566 | Minerals | 39,88328 |
| ieu-b-72 | eGFR decline (CKD population) [SH03] | Urinary sodium-potassium ratio | Simple mode | 24 | -0,22796 | 0,831602 | 0,786438 | Minerals | 39,88328 |
| ieu-b-72 | eGFR decline (CKD population) [SH03] | Urinary sodium-potassium ratio | Weighted mode | 24 | -0,2009 | 0,906404 | 0,826546 | Minerals | 39,88328 |
| prot-a-3102 | eGFR decline (CKD population) [SH03] | Thyrotropin-releasing hormone | Inverse variance weighted | 2 | 0,140176 | 0,135623 | 0,301337 | Thyroid disease | 36,35446 |
| prot-c-2652_15_1 | eGFR decline (CKD population) [SH03] | suPAR | Wald ratio | 1 | 0,034751 | 0,105085 | 0,740877 | Biomarkers | 33,5125 |
| ukb-b-4424 | eGFR decline (CKD population) [SH03] | Sleep duration | MR Egger | 66 | -0,03626 | 1,151961 | 0,974986 | Sleep disorders | 42,80283 |
| ukb-b-4424 | eGFR decline (CKD population) [SH03] | Sleep duration | Weighted median | 66 | -0,15141 | 0,462639 | 0,743463 | Sleep disorders | 42,80283 |
| ukb-b-4424 | eGFR decline (CKD population) [SH03] | Sleep duration | Inverse variance weighted | 66 | -0,17761 | 0,301941 | 0,556382 | Sleep disorders | 42,80283 |
| ukb-b-4424 | eGFR decline (CKD population) [SH03] | Sleep duration | Simple mode | 66 | -0,37664 | 0,960398 | 0,696215 | Sleep disorders | 42,80283 |
| ukb-b-4424 | eGFR decline (CKD population) [SH03] | Sleep duration | Weighted mode | 66 | -0,28071 | 0,775011 | 0,718377 | Sleep disorders | 42,80283 |
| ukb-b-5779 | eGFR decline (CKD population) [SH03] | Alcohol intake frequency. | MR Egger | 94 | -0,03954 | 0,273783 | 0,885472 | Alcohol consumption | 53,13815 |
| ukb-b-5779 | eGFR decline (CKD population) [SH03] | Alcohol intake frequency. | Weighted median | 94 | 0,038658 | 0,221809 | 0,861641 | Alcohol consumption | 53,13815 |
| ukb-b-5779 | eGFR decline (CKD population) [SH03] | Alcohol intake frequency. | Inverse variance weighted | 94 | -0,00663 | 0,120876 | 0,956244 | Alcohol consumption | 53,13815 |
| ukb-b-5779 | eGFR decline (CKD population) [SH03] | Alcohol intake frequency. | Simple mode | 94 | -0,31134 | 0,447346 | 0,488184 | Alcohol consumption | 53,13815 |
| ukb-b-5779 | eGFR decline (CKD population) [SH03] | Alcohol intake frequency. | Weighted mode | 94 | -0,03633 | 0,261703 | 0,889899 | Alcohol consumption | 53,13815 |
| ukb-b-7864 | eGFR decline (CKD population) [SH03] | Vitamin B6 | Wald ratio | 1 | -0,88738 | 0,847003 | 0,294792 | Vitamin | 32,75897 |
| ukb-d-30790_irnt | eGFR decline (CKD population) [SH03] | Lipoprotein A | MR Egger | 11 | -0,09058 | 0,188293 | 0,641939 | Dyslipidemias | 985,6435 |
| ukb-d-30790_irnt | eGFR decline (CKD population) [SH03] | Lipoprotein A | Weighted median | 11 | -0,18899 | 0,134482 | 0,159925 | Dyslipidemias | 985,6435 |
| ukb-d-30790_irnt | eGFR decline (CKD population) [SH03] | Lipoprotein A | Inverse variance weighted | 11 | -0,06387 | 0,12158 | 0,599374 | Dyslipidemias | 985,6435 |
| ukb-d-30790_irnt | eGFR decline (CKD population) [SH03] | Lipoprotein A | Simple mode | 11 | -0,19698 | 0,202261 | 0,353082 | Dyslipidemias | 985,6435 |
| ukb-d-30790_irnt | eGFR decline (CKD population) [SH03] | Lipoprotein A | Weighted mode | 11 | -0,22727 | 0,161126 | 0,188729 | Dyslipidemias | 985,6435 |
| BMI | eGFR decline (CKD population) [SH04] | Body mass index | MR Egger | 506 | 0,104562 | 0,203251 | 0,607164 | Obesity | 74,21321 |
| BMI | eGFR decline (CKD population) [SH04] | Body mass index | Weighted median | 506 | 0,039623 | 0,133719 | 0,766988 | Obesity | 74,21321 |
| BMI | eGFR decline (CKD population) [SH04] | Body mass index | Inverse variance weighted | 506 | 0,101696 | 0,078029 | 0,192471 | Obesity | 74,21321 |
| BMI | eGFR decline (CKD population) [SH04] | Body mass index | Simple mode | 506 | -0,27719 | 0,393651 | 0,481669 | Obesity | 74,21321 |
| BMI | eGFR decline (CKD population) [SH04] | Body mass index | Weighted mode | 506 | 0,170356 | 0,248002 | 0,492452 | Obesity | 74,21321 |
| DBPTang | eGFR decline (CKD population) [SH04] | Diastolic blood pressure | MR Egger | 251 | -0,08935 | 0,339591 | 0,792694 | Hypertension | 55,91029 |

| | | | | | | | | | |
|---|---|---|---|---|---|---|---|---|---|
| DBPTang | eGFR decline (CKD population) [SH04] | Diastolic blood pressure | Weighted median | 251 | -0,07313 | 0,159745 | 0,647096 | Hypertension | 55,91029 |
| DBPTang | eGFR decline (CKD population) [SH04] | Diastolic blood pressure | Inverse variance weighted | 251 | -0,03174 | 0,105426 | 0,763357 | Hypertension | 55,91029 |
| DBPTang | eGFR decline (CKD population) [SH04] | Diastolic blood pressure | Simple mode | 251 | 0,129149 | 0,449849 | 0,774278 | Hypertension | 55,91029 |
| DBPTang | eGFR decline (CKD population) [SH04] | Diastolic blood pressure | Weighted mode | 251 | -0,1985 | 0,352817 | 0,574211 | Hypertension | 55,91029 |
| FT3 | eGFR decline (CKD population) [SH04] | Free triiodothyronine | MR Egger | 6 | -0,55148 | 0,878549 | 0,564243 | Thyroid disease | 82,22035 |
| FT3 | eGFR decline (CKD population) [SH04] | Free triiodothyronine | Weighted median | 6 | -0,19516 | 0,245823 | 0,42725 | Thyroid disease | 82,22035 |
| FT3 | eGFR decline (CKD population) [SH04] | Free triiodothyronine | Inverse variance weighted | 6 | -0,19006 | 0,194256 | 0,327881 | Thyroid disease | 82,22035 |
| FT3 | eGFR decline (CKD population) [SH04] | Free triiodothyronine | Simple mode | 6 | -0,18774 | 0,327343 | 0,591123 | Thyroid disease | 82,22035 |
| FT3 | eGFR decline (CKD population) [SH04] | Free triiodothyronine | Weighted mode | 6 | -0,19288 | 0,307522 | 0,558046 | Thyroid disease | 82,22035 |
| FT3FT4 | eGFR decline (CKD population) [SH04] | FT3 FT4 ratio | MR Egger | 13 | 0,198516 | 1,10926 | 0,861221 | Thyroid disease | 132,3008 |
| FT3FT4 | eGFR decline (CKD population) [SH04] | FT3 FT4 ratio | Weighted median | 13 | -0,29869 | 0,710005 | 0,673984 | Thyroid disease | 132,3008 |
| FT3FT4 | eGFR decline (CKD population) [SH04] | FT3 FT4 ratio | Inverse variance weighted | 13 | -0,86322 | 0,551569 | 0,117576 | Thyroid disease | 132,3008 |
| FT3FT4 | eGFR decline (CKD population) [SH04] | FT3 FT4 ratio | Simple mode | 13 | 0,011912 | 1,362695 | 0,993169 | Thyroid disease | 132,3008 |
| FT3FT4 | eGFR decline (CKD population) [SH04] | FT3 FT4 ratio | Weighted mode | 13 | -0,39479 | 0,818215 | 0,638133 | Thyroid disease | 132,3008 |
| FT4 | eGFR decline (CKD population) [SH04] | Free thyroxine | MR Egger | 62 | -0,03528 | 0,173462 | 0,83951 | Thyroid disease | 84,3797 |
| FT4 | eGFR decline (CKD population) [SH04] | Free thyroxine | Weighted median | 62 | 0,027238 | 0,135747 | 0,840971 | Thyroid disease | 84,3797 |
| FT4 | eGFR decline (CKD population) [SH04] | Free thyroxine | Inverse variance weighted | 62 | -0,02919 | 0,082814 | 0,72445 | Thyroid disease | 84,3797 |
| FT4 | eGFR decline (CKD population) [SH04] | Free thyroxine | Simple mode | 62 | -0,14596 | 0,267474 | 0,587276 | Thyroid disease | 84,3797 |
| FT4 | eGFR decline (CKD population) [SH04] | Free thyroxine | Weighted mode | 62 | -0,00649 | 0,143006 | 0,963953 | Thyroid disease | 84,3797 |
| Fe | eGFR decline (CKD population) [SH04] | Serum iron | Inverse variance weighted | 2 | -0,26345 | 0,61289 | 0,667306 | Minerals | 291,9629 |
| HDL | eGFR decline (CKD population) [SH04] | HDL cholesterol | MR Egger | 233 | 0,018467 | 0,101031 | 0,855126 | Dyslipidemias | 268,9835 |
| HDL | eGFR decline (CKD population) [SH04] | HDL cholesterol | Weighted median | 233 | -0,01706 | 0,110287 | 0,877073 | Dyslipidemias | 268,9835 |
| HDL | eGFR decline (CKD population) [SH04] | HDL cholesterol | Inverse variance weighted | 233 | -0,10285 | 0,065294 | 0,1152 | Dyslipidemias | 268,9835 |
| HDL | eGFR decline (CKD population) [SH04] | HDL cholesterol | Simple mode | 233 | -0,10165 | 0,202465 | 0,616089 | Dyslipidemias | 268,9835 |
| HDL | eGFR decline (CKD population) [SH04] | HDL cholesterol | Weighted mode | 233 | -0,03011 | 0,101023 | 0,765907 | Dyslipidemias | 268,9835 |
| HiTSH | eGFR decline (CKD population) [SH04] | High TSH | MR Egger | 6 | 0,561066 | 0,522981 | 0,343763 | Thyroid disease | 49,53497 |
| HiTSH | eGFR decline (CKD population) [SH04] | High TSH | Weighted median | 6 | 0,310757 | 0,158409 | 0,049794 | Thyroid disease | 49,53497 |
| HiTSH | eGFR decline (CKD population) [SH04] | High TSH | Inverse variance weighted | 6 | 0,328095 | 0,127268 | 0,009938 | Thyroid disease | 49,53497 |
| HiTSH | eGFR decline (CKD population) [SH04] | High TSH | Simple mode | 6 | 0,31297 | 0,222359 | 0,218302 | Thyroid disease | 49,53497 |
| HiTSH | eGFR decline (CKD population) [SH04] | High TSH | Weighted mode | 6 | 0,317151 | 0,196788 | 0,167959 | Thyroid disease | 49,53497 |
| Insom | eGFR decline (CKD population) [SH04] | Insomnia | MR Egger | 13 | 0,077429 | 0,72155 | 0,916476 | Sleep disorders | 44,05724 |
| Insom | eGFR decline (CKD population) [SH04] | Insomnia | Weighted median | 13 | -0,16656 | 0,292963 | 0,569671 | Sleep disorders | 44,05724 |
| Insom | eGFR decline (CKD population) [SH04] | Insomnia | Inverse variance weighted | 13 | 0,098384 | 0,221878 | 0,657467 | Sleep disorders | 44,05724 |
| Insom | eGFR decline (CKD population) [SH04] | Insomnia | Simple mode | 13 | -0,27397 | 0,468617 | 0,56962 | Sleep disorders | 44,05724 |
| Insom | eGFR decline (CKD population) [SH04] | Insomnia | Weighted mode | 13 | -0,2129 | 0,423042 | 0,623898 | Sleep disorders | 44,05724 |
| LDL | eGFR decline (CKD population) [SH04] | LDL cholesterol | MR Egger | 217 | -0,06531 | 0,114661 | 0,569543 | Dyslipidemias | 221,0222 |
| LDL | eGFR decline (CKD population) [SH04] | LDL cholesterol | Weighted median | 217 | -0,07477 | 0,114211 | 0,512692 | Dyslipidemias | 221,0222 |
| LDL | eGFR decline (CKD population) [SH04] | LDL cholesterol | Inverse variance weighted | 217 | -0,0774 | 0,071719 | 0,280469 | Dyslipidemias | 221,0222 |
| LDL | eGFR decline (CKD population) [SH04] | LDL cholesterol | Simple mode | 217 | -0,18533 | 0,231408 | 0,424089 | Dyslipidemias | 221,0222 |
| LDL | eGFR decline (CKD population) [SH04] | LDL cholesterol | Weighted mode | 217 | -0,13355 | 0,122982 | 0,278712 | Dyslipidemias | 221,0222 |
| LoTSH | eGFR decline (CKD population) [SH04] | Low TSH | MR Egger | 26 | -0,02684 | 0,080502 | 0,741737 | Thyroid disease | 70,97091 |
| LoTSH | eGFR decline (CKD population) [SH04] | Low TSH | Weighted median | 26 | 0,034989 | 0,040996 | 0,393406 | Thyroid disease | 70,97091 |
| LoTSH | eGFR decline (CKD population) [SH04] | Low TSH | Inverse variance weighted | 26 | 0,017775 | 0,027706 | 0,521166 | Thyroid disease | 70,97091 |
| LoTSH | eGFR decline (CKD population) [SH04] | Low TSH | Simple mode | 26 | 0,085364 | 0,079666 | 0,294172 | Thyroid disease | 70,97091 |
| LoTSH | eGFR decline (CKD population) [SH04] | Low TSH | Weighted mode | 26 | 0,071092 | 0,062239 | 0,264171 | Thyroid disease | 70,97091 |
| PPTang | eGFR decline (CKD population) [SH04] | Pulse pressure | MR Egger | 300 | 0,213599 | 0,253233 | 0,399633 | Hypertension | 68,57434 |
| PPTang | eGFR decline (CKD population) [SH04] | Pulse pressure | Weighted median | 300 | 0,101772 | 0,157136 | 0,5172 | Hypertension | 68,57434 |
| PPTang | eGFR decline (CKD population) [SH04] | Pulse pressure | Inverse variance weighted | 300 | 0,162669 | 0,094004 | 0,083552 | Hypertension | 68,57434 |
| PPTang | eGFR decline (CKD population) [SH04] | Pulse pressure | Simple mode | 300 | -0,17859 | 0,457125 | 0,696305 | Hypertension | 68,57434 |
| PPTang | eGFR decline (CKD population) [SH04] | Pulse pressure | Weighted mode | 300 | 0,013462 | 0,299103 | 0,964132 | Hypertension | 68,57434 |
| SBPTang | eGFR decline (CKD population) [SH04] | Systolic blood pressure | MR Egger | 251 | -0,21491 | 0,337462 | 0,524823 | Hypertension | 56,96232 |
| SBPTang | eGFR decline (CKD population) [SH04] | Systolic blood pressure | Weighted median | 251 | -0,04741 | 0,16299 | 0,771141 | Hypertension | 56,96232 |
| SBPTang | eGFR decline (CKD population) [SH04] | Systolic blood pressure | Inverse variance weighted | 251 | 0,000135 | 0,108227 | 0,999002 | Hypertension | 56,96232 |
| SBPTang | eGFR decline (CKD population) [SH04] | Systolic blood pressure | Simple mode | 251 | 0,093673 | 0,467534 | 0,841366 | Hypertension | 56,96232 |
| SBPTang | eGFR decline (CKD population) [SH04] | Systolic blood pressure | Weighted mode | 251 | -0,05318 | 0,333391 | 0,873405 | Hypertension | 56,96232 |
| SIndex | eGFR decline (CKD population) [SH04] | Compound smoking index | MR Egger | 121 | 0,304289 | 1,008218 | 0,763325 | Smoking | 44,08236 |
| SIndex | eGFR decline (CKD population) [SH04] | Compound smoking index | Weighted median | 121 | -0,18608 | 0,366444 | 0,611586 | Smoking | 44,08236 |
| SIndex | eGFR decline (CKD population) [SH04] | Compound smoking index | Inverse variance weighted | 121 | -0,29092 | 0,251301 | 0,247008 | Smoking | 44,08236 |
| SIndex | eGFR decline (CKD population) [SH04] | Compound smoking index | Simple mode | 121 | -0,41153 | 0,954182 | 0,667028 | Smoking | 44,08236 |
| SIndex | eGFR decline (CKD population) [SH04] | Compound smoking index | Weighted mode | 121 | -0,28212 | 0,801179 | 0,725357 | Smoking | 44,08236 |
| T2DGGI | eGFR decline (CKD population) [SH04] | Type 2 diabetes | MR Egger | 441 | 0,126784 | 0,070258 | 0,071829 | Diabetes | 93,04757 |
| T2DGGI | eGFR decline (CKD population) [SH04] | Type 2 diabetes | Weighted median | 441 | 0,112844 | 0,067881 | 0,096434 | Diabetes | 93,04757 |
| T2DGGI | eGFR decline (CKD population) [SH04] | Type 2 diabetes | Inverse variance weighted | 441 | 0,090179 | 0,033939 | 0,007883 | Diabetes | 93,04757 |
| T2DGGI | eGFR decline (CKD population) [SH04] | Type 2 diabetes | Simple mode | 441 | 0,217099 | 0,147236 | 0,141063 | Diabetes | 93,04757 |
| T2DGGI | eGFR decline (CKD population) [SH04] | Type 2 diabetes | Weighted mode | 441 | 0,13207 | 0,079623 | 0,09789 | Diabetes | 93,04757 |
| TC | eGFR decline (CKD population) [SH04] | Total cholesterol | MR Egger | 264 | -0,06331 | 0,102734 | 0,53827 | Dyslipidemias | 246,1189 |
| TC | eGFR decline (CKD population) [SH04] | Total cholesterol | Weighted median | 264 | 0,009376 | 0,106842 | 0,930074 | Dyslipidemias | 246,1189 |
| TC | eGFR decline (CKD population) [SH04] | Total cholesterol | Inverse variance weighted | 264 | -0,01829 | 0,06542 | 0,779749 | Dyslipidemias | 246,1189 |
| TC | eGFR decline (CKD population) [SH04] | Total cholesterol | Simple mode | 264 | -0,08501 | 0,228183 | 0,709767 | Dyslipidemias | 246,1189 |
| TC | eGFR decline (CKD population) [SH04] | Total cholesterol | Weighted mode | 264 | -0,00392 | 0,112728 | 0,972298 | Dyslipidemias | 246,1189 |
| TG | eGFR decline (CKD population) [SH04] | Triglyceride | MR Egger | 219 | 0,113497 | 0,107586 | 0,292627 | Dyslipidemias | 231,1265 |
| TG | eGFR decline (CKD population) [SH04] | Triglyceride | Weighted median | 219 | 0,084772 | 0,119203 | 0,476991 | Dyslipidemias | 231,1265 |
| TG | eGFR decline (CKD population) [SH04] | Triglyceride | Inverse variance weighted | 219 | 0,055012 | 0,069353 | 0,427645 | Dyslipidemias | 231,1265 |
| TG | eGFR decline (CKD population) [SH04] | Triglyceride | Simple mode | 219 | -0,08347 | 0,261421 | 0,749815 | Dyslipidemias | 231,1265 |
| TG | eGFR decline (CKD population) [SH04] | Triglyceride | Weighted mode | 219 | 0,091142 | 0,110771 | 0,411526 | Dyslipidemias | 231,1265 |
| TSH | eGFR decline (CKD population) [SH04] | Thyroid Stimulating Hormone | MR Egger | 159 | 0,16217 | 0,099674 | 0,105743 | Thyroid disease | 154,4324 |
| TSH | eGFR decline (CKD population) [SH04] | Thyroid Stimulating Hormone | Weighted median | 159 | 0,110548 | 0,088885 | 0,2136 | Thyroid disease | 154,4324 |
| TSH | eGFR decline (CKD population) [SH04] | Thyroid Stimulating Hormone | Inverse variance weighted | 159 | 0,041407 | 0,05524 | 0,453505 | Thyroid disease | 154,4324 |
| TSH | eGFR decline (CKD population) [SH04] | Thyroid Stimulating Hormone | Simple mode | 159 | -0,02969 | 0,185302 | 0,87292 | Thyroid disease | 154,4324 |
| TSH | eGFR decline (CKD population) [SH04] | Thyroid Stimulating Hormone | Weighted mode | 159 | 0,174843 | 0,097622 | 0,075205 | Thyroid disease | 154,4324 |
| WHR | eGFR decline (CKD population) [SH04] | Waist-to-hip ratio adjusted BMI | MR Egger | 296 | -0,16489 | 0,215298 | 0,444371 | Obesity | 85,86852 |
| WHR | eGFR decline (CKD population) [SH04] | Waist-to-hip ratio adjusted BMI | Weighted median | 296 | -0,02068 | 0,158863 | 0,896437 | Obesity | 85,86852 |
| WHR | eGFR decline (CKD population) [SH04] | Waist-to-hip ratio adjusted BMI | Inverse variance weighted | 296 | 0,028661 | 0,090137 | 0,750506 | Obesity | 85,86852 |
| WHR | eGFR decline (CKD population) [SH04] | Waist-to-hip ratio adjusted BMI | Simple mode | 296 | -0,11352 | 0,396895 | 0,775055 | Obesity | 85,86852 |
| WHR | eGFR decline (CKD population) [SH04] | Waist-to-hip ratio adjusted BMI | Weighted mode | 296 | 0,021271 | 0,226732 | 0,925318 | Obesity | 85,86852 |
| Zn | eGFR decline (CKD population) [SH04] | Zinc | MR Egger | 3 | -0,1884 | 0,325912 | 0,666327 | Minerals | 99,95282 |
| Zn | eGFR decline (CKD population) [SH04] | Zinc | Weighted median | 3 | 0,026208 | 0,094422 | 0,781348 | Minerals | 99,95282 |
| Zn | eGFR decline (CKD population) [SH04] | Zinc | Inverse variance weighted | 3 | 0,070142 | 0,110744 | 0,526491 | Minerals | 99,95282 |
| Zn | eGFR decline (CKD population) [SH04] | Zinc | Simple mode | 3 | 0,000462 | 0,110319 | 0,997041 | Minerals | 99,95282 |
| Zn | eGFR decline (CKD population) [SH04] | Zinc | Weighted mode | 3 | 0,019932 | 0,094549 | 0,852567 | Minerals | 99,95282 |
| ebi-a-GCST006979-consort | eGFR decline (CKD population) [SH04] | Heel bone mineral density | MR Egger | 483 | 0,013869 | 0,099991 | 0,889746 | Bone disease | 151,4375 |
| ebi-a-GCST006979-consort | eGFR decline (CKD population) [SH04] | Heel bone mineral density | Weighted median | 483 | 0,157954 | 0,079379 | 0,046606 | Bone disease | 151,4375 |
| ebi-a-GCST006979-consort | eGFR decline (CKD population) [SH04] | Heel bone mineral density | Inverse variance weighted | 483 | 0,08916 | 0,051642 | 0,084255 | Bone disease | 151,4375 |
| ebi-a-GCST006979-consort | eGFR decline (CKD population) [SH04] | Heel bone mineral density | Simple mode | 483 | 0,185338 | 0,195609 | 0,343863 | Bone disease | 151,4375 |
| ebi-a-GCST006979-consort | eGFR decline (CKD population) [SH04] | Heel bone mineral density | Weighted mode | 483 | 0,130333 | 0,119089 | 0,274321 | Bone disease | 151,4375 |
| ebi-a-GCST90002227 | eGFR decline (CKD population) [SH04] | Two-hour glucose | MR Egger | 14 | -0,13442 | 0,326808 | 0,688094 | Diabetes | 56,47851 |
| ebi-a-GCST90002227 | eGFR decline (CKD population) [SH04] | Two-hour glucose | Weighted median | 14 | 0,241755 | 0,16463 | 0,141975 | Diabetes | 56,47851 |
| ebi-a-GCST90002227 | eGFR decline (CKD population) [SH04] | Two-hour glucose | Inverse variance weighted | 14 | 0,153782 | 0,12005 | 0,2002 | Diabetes | 56,47851 |
| ebi-a-GCST90002227 | eGFR decline (CKD population) [SH04] | Two-hour glucose | Simple mode | 14 | 0,266873 | 0,285083 | 0,366274 | Diabetes | 56,47851 |
| ebi-a-GCST90002227 | eGFR decline (CKD population) [SH04] | Two-hour glucose | Weighted mode | 14 | 0,304931 | 0,252518 | 0,248727 | Diabetes | 56,47851 |
| ebi-a-GCST90002232 | eGFR decline (CKD population) [SH04] | Fasting glucose | MR Egger | 68 | 0,592179 | 0,318404 | 0,067366 | Diabetes | 124,2211 |
| ebi-a-GCST90002232 | eGFR decline (CKD population) [SH04] | Fasting glucose | Weighted median | 68 | 0,681551 | 0,251309 | 0,006688 | Diabetes | 124,2211 |
| ebi-a-GCST90002232 | eGFR decline (CKD population) [SH04] | Fasting glucose | Inverse variance weighted | 68 | 0,217421 | 0,168394 | 0,196653 | Diabetes | 124,2211 |
| ebi-a-GCST90002232 | eGFR decline (CKD population) [SH04] | Fasting glucose | Simple mode | 68 | 0,653536 | 0,486358 | 0,183567 | Diabetes | 124,2211 |
| ebi-a-GCST90002232 | eGFR decline (CKD population) [SH04] | Fasting glucose | Weighted mode | 68 | 0,681697 | 0,24991 | 0,008134 | Diabetes | 124,2211 |
| ebi-a-GCST90002238 | eGFR decline (CKD population) [SH04] | Fasting insulin | MR Egger | 37 | 0,407626 | 1,183326 | 0,732551 | Diabetes | 52,15552 |
| ebi-a-GCST90002238 | eGFR decline (CKD population) [SH04] | Fasting insulin | Weighted median | 37 | 0,385451 | 0,447825 | 0,389393 | Diabetes | 52,15552 |
| ebi-a-GCST90002238 | eGFR decline (CKD population) [SH04] | Fasting insulin | Inverse variance weighted | 37 | 0,063135 | 0,362478 | 0,861728 | Diabetes | 52,15552 |
| ebi-a-GCST90002238 | eGFR decline (CKD population) [SH04] | Fasting insulin | Simple mode | 37 | 0,41249 | 0,768318 | 0,594658 | Diabetes | 52,15552 |
| ebi-a-GCST90002238 | eGFR decline (CKD population) [SH04] | Fasting insulin | Weighted mode | 37 | 0,377502 | 0,696297 | 0,591051 | Diabetes | 52,15552 |
| ebi-a-GCST90007310 | eGFR decline (CKD population) [SH04] | circulating leptin levels | MR Egger | 3 | 3,130363 | 2,307867 | 0,40444 | Obesity | 56,18818 |
| ebi-a-GCST90007310 | eGFR decline (CKD population) [SH04] | circulating leptin levels | Weighted median | 3 | -0,23889 | 0,343954 | 0,487352 | Obesity | 56,18818 |
| ebi-a-GCST90007310 | eGFR decline (CKD population) [SH04] | circulating leptin levels | Inverse variance weighted | 3 | -0,40597 | 0,322509 | 0,208104 | Obesity | 56,18818 |

| | | | | | | | | | |
|---|---|---|---|---|---|---|---|---|---|
| ebi-a-GCST90007310 | eGFR decline (CKD population) [SH04] | circulating leptin levels | Simple mode | 3 | -0,17116 | 0,42365 | 0,725314 | Obesity | 56,18818 |
| ebi-a-GCST90007310 | eGFR decline (CKD population) [SH04] | circulating leptin levels | Weighted mode | 3 | -0,17116 | 0,419273 | 0,722666 | Obesity | 56,18818 |
| ebi-a-GCST90011998 | eGFR decline (CKD population) [SH04] | Growth differentiation factor-15 levels | MR Egger | 3 | -0,1976 | 0,13766 | 0,387366 | Biomarkers | 294,0397 |
| ebi-a-GCST90011998 | eGFR decline (CKD population) [SH04] | Growth differentiation factor-15 levels | Weighted median | 3 | -0,15374 | 0,095207 | 0,106352 | Biomarkers | 294,0397 |
| ebi-a-GCST90011998 | eGFR decline (CKD population) [SH04] | Growth differentiation factor-15 levels | Inverse variance weighted | 3 | -0,13766 | 0,094151 | 0,143711 | Biomarkers | 294,0397 |
| ebi-a-GCST90011998 | eGFR decline (CKD population) [SH04] | Growth differentiation factor-15 levels | Simple mode | 3 | -0,13595 | 0,179376 | 0,527625 | Biomarkers | 294,0397 |
| ebi-a-GCST90011998 | eGFR decline (CKD population) [SH04] | Growth differentiation factor-15 levels | Weighted mode | 3 | -0,15665 | 0,093215 | 0,234867 | Biomarkers | 294,0397 |
| ebi-a-GCST90012022 | eGFR decline (CKD population) [SH04] | Fibroblast growth factor 23 levels | MR Egger | 8 | 0,188308 | 0,470047 | 0,702579 | Biomarkers | 43,42098 |
| ebi-a-GCST90012022 | eGFR decline (CKD population) [SH04] | Fibroblast growth factor 23 levels | Weighted median | 8 | 0,15445 | 0,167884 | 0,357583 | Biomarkers | 43,42098 |
| ebi-a-GCST90012022 | eGFR decline (CKD population) [SH04] | Fibroblast growth factor 23 levels | Inverse variance weighted | 8 | 0,028927 | 0,127293 | 0,820234 | Biomarkers | 43,42098 |
| ebi-a-GCST90012022 | eGFR decline (CKD population) [SH04] | Fibroblast growth factor 23 levels | Simple mode | 8 | 0,225441 | 0,245657 | 0,389298 | Biomarkers | 43,42098 |
| ebi-a-GCST90012022 | eGFR decline (CKD population) [SH04] | Fibroblast growth factor 23 levels | Weighted mode | 8 | 0,236338 | 0,269336 | 0,409329 | Biomarkers | 43,42098 |
| ebi-a-GCST90012041 | eGFR decline (CKD population) [SH04] | Kidney injury molecule 1 levels | MR Egger | 9 | -0,05217 | 0,115493 | 0,665155 | Biomarkers | 190,9807 |
| ebi-a-GCST90012041 | eGFR decline (CKD population) [SH04] | Kidney injury molecule 1 levels | Weighted median | 9 | -0,04861 | 0,062224 | 0,434695 | Biomarkers | 190,9807 |
| ebi-a-GCST90012041 | eGFR decline (CKD population) [SH04] | Kidney injury molecule 1 levels | Inverse variance weighted | 9 | -0,04514 | 0,069027 | 0,513114 | Biomarkers | 190,9807 |
| ebi-a-GCST90012041 | eGFR decline (CKD population) [SH04] | Kidney injury molecule 1 levels | Simple mode | 9 | -0,01501 | 0,139989 | 0,917234 | Biomarkers | 190,9807 |
| ebi-a-GCST90012041 | eGFR decline (CKD population) [SH04] | Kidney injury molecule 1 levels | Weighted mode | 9 | -0,04715 | 0,062795 | 0,474228 | Biomarkers | 190,9807 |
| ebi-a-GCST90013864 | eGFR decline (CKD population) [SH04] | Coronary artery disease (Firth correction) | MR Egger | 37 | -0,08324 | 0,135178 | 0,542026 | Cardiovascular disease | 59,8949 |
| ebi-a-GCST90013864 | eGFR decline (CKD population) [SH04] | Coronary artery disease (Firth correction) | Weighted median | 37 | -0,0448 | 0,08573 | 0,601281 | Cardiovascular disease | 59,8949 |
| ebi-a-GCST90013864 | eGFR decline (CKD population) [SH04] | Coronary artery disease (Firth correction) | Inverse variance weighted | 37 | -0,02183 | 0,060816 | 0,719686 | Cardiovascular disease | 59,8949 |
| ebi-a-GCST90013864 | eGFR decline (CKD population) [SH04] | Coronary artery disease (Firth correction) | Simple mode | 37 | -0,08451 | 0,174022 | 0,630154 | Cardiovascular disease | 59,8949 |
| ebi-a-GCST90013864 | eGFR decline (CKD population) [SH04] | Coronary artery disease (Firth correction) | Weighted mode | 37 | -0,10313 | 0,107871 | 0,345425 | Cardiovascular disease | 59,8949 |
| ebi-a-GCST90013975 | eGFR decline (CKD population) [SH04] | Body fat percentage | MR Egger | 316 | 0,419407 | 0,397061 | 0,291653 | Obesity | 56,17886 |
| ebi-a-GCST90013975 | eGFR decline (CKD population) [SH04] | Body fat percentage | Weighted median | 316 | 0,012974 | 0,186304 | 0,94448 | Obesity | 56,17886 |
| ebi-a-GCST90013975 | eGFR decline (CKD population) [SH04] | Body fat percentage | Inverse variance weighted | 316 | 0,097163 | 0,123202 | 0,430319 | Obesity | 56,17886 |
| ebi-a-GCST90013975 | eGFR decline (CKD population) [SH04] | Body fat percentage | Simple mode | 316 | 0,376819 | 0,561067 | 0,502324 | Obesity | 56,17886 |
| ebi-a-GCST90013975 | eGFR decline (CKD population) [SH04] | Body fat percentage | Weighted mode | 316 | -0,25925 | 0,424073 | 0,541422 | Obesity | 56,17886 |
| ebi-a-GCST90013989 | eGFR decline (CKD population) [SH04] | Sodium in urine | MR Egger | 41 | 1,196471 | 1,099029 | 0,282985 | Minerals | 43,81211 |
| ebi-a-GCST90013989 | eGFR decline (CKD population) [SH04] | Sodium in urine | Weighted median | 41 | -0,45115 | 0,41808 | 0,280543 | Minerals | 43,81211 |
| ebi-a-GCST90013989 | eGFR decline (CKD population) [SH04] | Sodium in urine | Inverse variance weighted | 41 | -0,18269 | 0,282126 | 0,517285 | Minerals | 43,81211 |
| ebi-a-GCST90013989 | eGFR decline (CKD population) [SH04] | Sodium in urine | Simple mode | 41 | -0,36795 | 0,920067 | 0,691344 | Minerals | 43,81211 |
| ebi-a-GCST90013989 | eGFR decline (CKD population) [SH04] | Sodium in urine | Weighted mode | 41 | -0,72595 | 0,813951 | 0,37779 | Minerals | 43,81211 |
| ebi-a-GCST90014006 | eGFR decline (CKD population) [SH04] | Glycated haemoglobin HbA1c levels | MR Egger | 302 | 0,24771 | 0,122384 | 0,043853 | Diabetes | 128,2905 |
| ebi-a-GCST90014006 | eGFR decline (CKD population) [SH04] | Glycated haemoglobin HbA1c levels | Weighted median | 302 | 0,140769 | 0,109102 | 0,196962 | Diabetes | 128,2905 |
| ebi-a-GCST90014006 | eGFR decline (CKD population) [SH04] | Glycated haemoglobin HbA1c levels | Inverse variance weighted | 302 | 0,060585 | 0,064491 | 0,34751 | Diabetes | 128,2905 |
| ebi-a-GCST90014006 | eGFR decline (CKD population) [SH04] | Glycated haemoglobin HbA1c levels | Simple mode | 302 | -0,01368 | 0,245072 | 0,955535 | Diabetes | 128,2905 |
| ebi-a-GCST90014006 | eGFR decline (CKD population) [SH04] | Glycated haemoglobin HbA1c levels | Weighted mode | 302 | 0,111663 | 0,147306 | 0,449023 | Diabetes | 128,2905 |
| ebi-a-GCST90014010 | eGFR decline (CKD population) [SH04] | Phosphate levels | MR Egger | 160 | 0,175671 | 0,169755 | 0,302322 | Minerals | 87,71201 |
| ebi-a-GCST90014010 | eGFR decline (CKD population) [SH04] | Phosphate levels | Weighted median | 160 | -0,06054 | 0,171835 | 0,724588 | Minerals | 87,71201 |
| ebi-a-GCST90014010 | eGFR decline (CKD population) [SH04] | Phosphate levels | Inverse variance weighted | 160 | -0,04518 | 0,095166 | 0,634954 | Minerals | 87,71201 |
| ebi-a-GCST90014010 | eGFR decline (CKD population) [SH04] | Phosphate levels | Simple mode | 160 | -0,03636 | 0,353809 | 0,918275 | Minerals | 87,71201 |
| ebi-a-GCST90014010 | eGFR decline (CKD population) [SH04] | Phosphate levels | Weighted mode | 160 | 0,098767 | 0,219304 | 0,653061 | Minerals | 87,71201 |
| ebi-a-GCST90014022 | eGFR decline (CKD population) [SH04] | Bone mineral density | MR Egger | 415 | 0,0552 | 0,110269 | 0,616925 | Bone disease | 120,0409 |
| ebi-a-GCST90014022 | eGFR decline (CKD population) [SH04] | Bone mineral density | Weighted median | 415 | 0,106608 | 0,090752 | 0,240108 | Bone disease | 120,0409 |
| ebi-a-GCST90014022 | eGFR decline (CKD population) [SH04] | Bone mineral density | Inverse variance weighted | 415 | 0,082439 | 0,055621 | 0,138299 | Bone disease | 120,0409 |
| ebi-a-GCST90014022 | eGFR decline (CKD population) [SH04] | Bone mineral density | Simple mode | 415 | 0,084697 | 0,220969 | 0,701696 | Bone disease | 120,0409 |
| ebi-a-GCST90014022 | eGFR decline (CKD population) [SH04] | Bone mineral density | Weighted mode | 415 | 0,054048 | 0,131724 | 0,681791 | Bone disease | 120,0409 |
| ebi-a-GCST90014023 | eGFR decline (CKD population) [SH04] | Type 1 diabetes | MR Egger | 76 | -0,02012 | 0,028888 | 0,48831 | Diabetes | 131,7111 |
| ebi-a-GCST90014023 | eGFR decline (CKD population) [SH04] | Type 1 diabetes | Weighted median | 76 | -0,02018 | 0,026988 | 0,454629 | Diabetes | 131,7111 |
| ebi-a-GCST90014023 | eGFR decline (CKD population) [SH04] | Type 1 diabetes | Inverse variance weighted | 76 | -0,01608 | 0,017611 | 0,361185 | Diabetes | 131,7111 |
| ebi-a-GCST90014023 | eGFR decline (CKD population) [SH04] | Type 1 diabetes | Simple mode | 76 | -0,04613 | 0,053595 | 0,392109 | Diabetes | 131,7111 |
| ebi-a-GCST90014023 | eGFR decline (CKD population) [SH04] | Type 1 diabetes | Weighted mode | 76 | -0,03406 | 0,024808 | 0,173917 | Diabetes | 131,7111 |
| ebi-a-GCST90016674 | eGFR decline (CKD population) [SH04] | Liver iron content | MR Egger | 5 | -0,09105 | 0,173754 | 0,636513 | Minerals | 257,3642 |
| ebi-a-GCST90016674 | eGFR decline (CKD population) [SH04] | Liver iron content | Weighted median | 5 | -0,09036 | 0,106942 | 0,398126 | Minerals | 257,3642 |
| ebi-a-GCST90016674 | eGFR decline (CKD population) [SH04] | Liver iron content | Inverse variance weighted | 5 | -0,0847 | 0,101189 | 0,402582 | Minerals | 257,3642 |
| ebi-a-GCST90016674 | eGFR decline (CKD population) [SH04] | Liver iron content | Simple mode | 5 | -0,14732 | 0,192346 | 0,486432 | Minerals | 257,3642 |
| ebi-a-GCST90016674 | eGFR decline (CKD population) [SH04] | Liver iron content | Weighted mode | 5 | -0,09451 | 0,11142 | 0,444108 | Minerals | 257,3642 |
| ebi-a-GCST90016676 | eGFR decline (CKD population) [SH04] | Pancreas iron content | Wald ratio | 1 | -0,26833 | 0,54544 | 0,622754 | Minerals | 30,56027 |
| ebi-a-GCST90018806 | eGFR decline (CKD population) [SH04] | Chronic heart failure | MR Egger | 5 | 0,888623 | 1,303272 | 0,544263 | Cardiovascular disease | 41,81649 |
| ebi-a-GCST90018806 | eGFR decline (CKD population) [SH04] | Chronic heart failure | Weighted median | 5 | 0,175912 | 0,230102 | 0,444573 | Cardiovascular disease | 41,81649 |
| ebi-a-GCST90018806 | eGFR decline (CKD population) [SH04] | Chronic heart failure | Inverse variance weighted | 5 | 0,12837 | 0,187959 | 0,494627 | Cardiovascular disease | 41,81649 |
| ebi-a-GCST90018806 | eGFR decline (CKD population) [SH04] | Chronic heart failure | Simple mode | 5 | 0,265019 | 0,30262 | 0,430605 | Cardiovascular disease | 41,81649 |
| ebi-a-GCST90018806 | eGFR decline (CKD population) [SH04] | Chronic heart failure | Weighted mode | 5 | 0,23984 | 0,302851 | 0,472712 | Cardiovascular disease | 41,81649 |
| ebi-a-GCST90018860 | eGFR decline (CKD population) [SH04] | Hyperthyroidism | MR Egger | 8 | -0,13917 | 0,272448 | 0,627728 | Thyroid disease | 53,27573 |
| ebi-a-GCST90018860 | eGFR decline (CKD population) [SH04] | Hyperthyroidism | Weighted median | 8 | 0,140959 | 0,082706 | 0,088319 | Thyroid disease | 53,27573 |
| ebi-a-GCST90018860 | eGFR decline (CKD population) [SH04] | Hyperthyroidism | Inverse variance weighted | 8 | 0,082605 | 0,076682 | 0,281373 | Thyroid disease | 53,27573 |
| ebi-a-GCST90018860 | eGFR decline (CKD population) [SH04] | Hyperthyroidism | Simple mode | 8 | 0,196359 | 0,152833 | 0,239743 | Thyroid disease | 53,27573 |
| ebi-a-GCST90018860 | eGFR decline (CKD population) [SH04] | Hyperthyroidism | Weighted mode | 8 | 0,176548 | 0,13977 | 0,246985 | Thyroid disease | 53,27573 |
| ebi-a-GCST90018862 | eGFR decline (CKD population) [SH04] | Hypothyroidism | MR Egger | 64 | -0,03359 | 0,088713 | 0,706247 | Thyroid disease | 82,13671 |
| ebi-a-GCST90018862 | eGFR decline (CKD population) [SH04] | Hypothyroidism | Weighted median | 64 | 0,025077 | 0,058649 | 0,668954 | Thyroid disease | 82,13671 |
| ebi-a-GCST90018862 | eGFR decline (CKD population) [SH04] | Hypothyroidism | Inverse variance weighted | 64 | 0,048219 | 0,03822 | 0,207082 | Thyroid disease | 82,13671 |
| ebi-a-GCST90018862 | eGFR decline (CKD population) [SH04] | Hypothyroidism | Simple mode | 64 | 0,044783 | 0,120534 | 0,711482 | Thyroid disease | 82,13671 |
| ebi-a-GCST90018862 | eGFR decline (CKD population) [SH04] | Hypothyroidism | Weighted mode | 64 | -0,01907 | 0,07983 | 0,811939 | Thyroid disease | 82,13671 |
| ebi-a-GCST90018872 | eGFR decline (CKD population) [SH04] | Iron deficiency anemia | Wald ratio | 1 | 0,638406 | 0,400205 | 0,110668 | Minerals | 34,53822 |
| ebi-a-GCST90018877 | eGFR decline (CKD population) [SH04] | Myocardial infarction | MR Egger | 70 | -0,11905 | 0,096553 | 0,22183 | Cardiovascular disease | 66,68595 |
| ebi-a-GCST90018877 | eGFR decline (CKD population) [SH04] | Myocardial infarction | Weighted median | 70 | -0,01858 | 0,062994 | 0,768065 | Cardiovascular disease | 66,68595 |
| ebi-a-GCST90018877 | eGFR decline (CKD population) [SH04] | Myocardial infarction | Inverse variance weighted | 70 | 0,018584 | 0,043176 | 0,666881 | Cardiovascular disease | 66,68595 |
| ebi-a-GCST90018877 | eGFR decline (CKD population) [SH04] | Myocardial infarction | Simple mode | 70 | 0,063626 | 0,12192 | 0,603436 | Cardiovascular disease | 66,68595 |
| ebi-a-GCST90018877 | eGFR decline (CKD population) [SH04] | Myocardial infarction | Weighted mode | 70 | -0,03611 | 0,08276 | 0,664 | Cardiovascular disease | 66,68595 |
| ebi-a-GCST90018916 | eGFR decline (CKD population) [SH04] | Sleep apnea syndrome | Wald ratio | 1 | -0,06204 | 0,241631 | 0,79738 | Sleep disorders | 63,50864 |
| ebi-a-GCST90018977 | eGFR decline (CKD population) [SH04] | Serum uric acid levels | MR Egger | 237 | -0,00526 | 0,15611 | 0,973144 | Uricemia | 110,1094 |
| ebi-a-GCST90018977 | eGFR decline (CKD population) [SH04] | Serum uric acid levels | Weighted median | 237 | 0,111684 | 0,13749 | 0,416615 | Uricemia | 110,1094 |
| ebi-a-GCST90018977 | eGFR decline (CKD population) [SH04] | Serum uric acid levels | Inverse variance weighted | 237 | 0,150646 | 0,086982 | 0,083289 | Uricemia | 110,1094 |
| ebi-a-GCST90018977 | eGFR decline (CKD population) [SH04] | Serum uric acid levels | Simple mode | 237 | 0,319232 | 0,336306 | 0,343474 | Uricemia | 110,1094 |
| ebi-a-GCST90018977 | eGFR decline (CKD population) [SH04] | Serum uric acid levels | Weighted mode | 237 | 0,122479 | 0,148953 | 0,411756 | Uricemia | 110,1094 |
| ebi-a-GCST90019417 | eGFR decline (CKD population) [SH04] | Vitamin K-dependent protein S measurement | MR Egger | 4 | 0,052948 | 0,156194 | 0,766904 | Vitamin | 118,3171 |
| ebi-a-GCST90019417 | eGFR decline (CKD population) [SH04] | Vitamin K-dependent protein S measurement | Weighted median | 4 | 0,039864 | 0,096703 | 0,680171 | Vitamin | 118,3171 |
| ebi-a-GCST90019417 | eGFR decline (CKD population) [SH04] | Vitamin K-dependent protein S measurement | Inverse variance weighted | 4 | 0,024359 | 0,089541 | 0,785586 | Vitamin | 118,3171 |
| ebi-a-GCST90019417 | eGFR decline (CKD population) [SH04] | Vitamin K-dependent protein S measurement | Simple mode | 4 | 0,033559 | 0,124638 | 0,805192 | Vitamin | 118,3171 |
| ebi-a-GCST90019417 | eGFR decline (CKD population) [SH04] | Vitamin K-dependent protein S measurement | Weighted mode | 4 | 0,041132 | 0,115484 | 0,745287 | Vitamin | 118,3171 |
| ebi-a-GCST90019423 | eGFR decline (CKD population) [SH04] | Vitamin K-dependent protein C measurement | MR Egger | 3 | 0,082435 | 0,253411 | 0,799779 | Vitamin | 207,8561 |
| ebi-a-GCST90019423 | eGFR decline (CKD population) [SH04] | Vitamin K-dependent protein C measurement | Weighted median | 3 | 0,071914 | 0,070666 | 0,308839 | Vitamin | 207,8561 |
| ebi-a-GCST90019423 | eGFR decline (CKD population) [SH04] | Vitamin K-dependent protein C measurement | Inverse variance weighted | 3 | 0,041754 | 0,114991 | 0,716527 | Vitamin | 207,8561 |
| ebi-a-GCST90019423 | eGFR decline (CKD population) [SH04] | Vitamin K-dependent protein C measurement | Simple mode | 3 | 0,025774 | 0,178625 | 0,898497 | Vitamin | 207,8561 |
| ebi-a-GCST90019423 | eGFR decline (CKD population) [SH04] | Vitamin K-dependent protein C measurement | Weighted mode | 3 | 0,079151 | 0,071182 | 0,38191 | Vitamin | 207,8561 |
| ebi-a-GCST90025947 | eGFR decline (CKD population) [SH04] | Serum alkaline phosphatase levels | MR Egger | 335 | -0,09182 | 0,101457 | 0,366124 | Minerals | 166,037 |
| ebi-a-GCST90025947 | eGFR decline (CKD population) [SH04] | Serum alkaline phosphatase levels | Weighted median | 335 | 0,036728 | 0,103414 | 0,722477 | Minerals | 166,037 |
| ebi-a-GCST90025947 | eGFR decline (CKD population) [SH04] | Serum alkaline phosphatase levels | Inverse variance weighted | 335 | -0,02155 | 0,059453 | 0,717062 | Minerals | 166,037 |
| ebi-a-GCST90025947 | eGFR decline (CKD population) [SH04] | Serum alkaline phosphatase levels | Simple mode | 335 | 0,102071 | 0,221211 | 0,6448 | Minerals | 166,037 |
| ebi-a-GCST90025947 | eGFR decline (CKD population) [SH04] | Serum alkaline phosphatase levels | Weighted mode | 335 | 0,075623 | 0,119592 | 0,527594 | Minerals | 166,037 |
| ebi-a-GCST90025967 | eGFR decline (CKD population) [SH04] | Vitamin D levels | MR Egger | 81 | -0,05604 | 0,189637 | 0,768388 | Vitamin | 134,1402 |
| ebi-a-GCST90025967 | eGFR decline (CKD population) [SH04] | Vitamin D levels | Weighted median | 81 | -0,22852 | 0,197827 | 0,248038 | Vitamin | 134,1402 |
| ebi-a-GCST90025967 | eGFR decline (CKD population) [SH04] | Vitamin D levels | Inverse variance weighted | 81 | -0,06667 | 0,116295 | 0,566477 | Vitamin | 134,1402 |
| ebi-a-GCST90025967 | eGFR decline (CKD population) [SH04] | Vitamin D levels | Simple mode | 81 | -0,20076 | 0,361763 | 0,580478 | Vitamin | 134,1402 |
| ebi-a-GCST90025967 | eGFR decline (CKD population) [SH04] | Vitamin D levels | Weighted mode | 81 | -0,15223 | 0,165918 | 0,361642 | Vitamin | 134,1402 |
| ebi-a-GCST90025969 | eGFR decline (CKD population) [SH04] | Hemoglobin levels | MR Egger | 325 | -0,00096 | 0,138163 | 0,994432 | Organic compound | 104,3556 |
| ebi-a-GCST90025969 | eGFR decline (CKD population) [SH04] | Hemoglobin levels | Weighted median | 325 | -0,08039 | 0,118624 | 0,49798 | Organic compound | 104,3556 |
| ebi-a-GCST90025969 | eGFR decline (CKD population) [SH04] | Hemoglobin levels | Inverse variance weighted | 325 | -0,1139 | 0,07109 | 0,109107 | Organic compound | 104,3556 |
| ebi-a-GCST90025969 | eGFR decline (CKD population) [SH04] | Hemoglobin levels | Simple mode | 325 | 0,136299 | 0,307193 | 0,657561 | Organic compound | 104,3556 |
| ebi-a-GCST90025969 | eGFR decline (CKD population) [SH04] | Hemoglobin levels | Weighted mode | 325 | -0,07678 | 0,138463 | 0,579602 | Organic compound | 104,3556 |
| ebi-a-GCST90025986 | eGFR decline (CKD population) [SH04] | Blood glucose levels | MR Egger | 109 | 0,194236 | 0,18815 | 0,304237 | Diabetes | 118,7554 |
| ebi-a-GCST90025986 | eGFR decline (CKD population) [SH04] | Blood glucose levels | Weighted median | 109 | 0,312907 | 0,171915 | 0,068739 | Diabetes | 118,7554 |
| ebi-a-GCST90025986 | eGFR decline (CKD population) [SH04] | Blood glucose levels | Inverse variance weighted | 109 | 0,013983 | 0,105969 | 0,895019 | Diabetes | 118,7554 |

| | | | | | | | | | |
|---|---|---|---|---|---|---|---|---|---|
| ebi-a-GCST90025986 | eGFR decline (CKD population) [SH04] | Blood glucose levels | Simple mode | 109 | 0,173435 | 0,384012 | 0,652434 | Diabetes | 118,7554 |
| ebi-a-GCST90025986 | eGFR decline (CKD population) [SH04] | Blood glucose levels | Weighted mode | 109 | 0,315214 | 0,169573 | 0,065768 | Diabetes | 118,7554 |
| ebi-a-GCST90025989 | eGFR decline (CKD population) [SH04] | Insulin-like growth factor 1 levels | MR Egger | 373 | -0,19584 | 0,120951 | 0,106257 | Diabetes | 115,9356 |
| ebi-a-GCST90025989 | eGFR decline (CKD population) [SH04] | Insulin-like growth factor 1 levels | Weighted median | 373 | -0,01347 | 0,105126 | 0,898035 | Diabetes | 115,9356 |
| ebi-a-GCST90025989 | eGFR decline (CKD population) [SH04] | Insulin-like growth factor 1 levels | Inverse variance weighted | 373 | 0,006327 | 0,062524 | 0,919401 | Diabetes | 115,9356 |
| ebi-a-GCST90025989 | eGFR decline (CKD population) [SH04] | Insulin-like growth factor 1 levels | Simple mode | 373 | -0,07953 | 0,248074 | 0,748699 | Diabetes | 115,9356 |
| ebi-a-GCST90025989 | eGFR decline (CKD population) [SH04] | Insulin-like growth factor 1 levels | Weighted mode | 373 | -0,14193 | 0,137575 | 0,30291 | Diabetes | 115,9356 |
| ebi-a-GCST90025990 | eGFR decline (CKD population) [SH04] | Calcium levels | MR Egger | 202 | 0,270319 | 0,153702 | 0,080155 | Minerals | 102,0722 |
| ebi-a-GCST90025990 | eGFR decline (CKD population) [SH04] | Calcium levels | Weighted median | 202 | 0,063487 | 0,171554 | 0,711331 | Minerals | 102,0722 |
| ebi-a-GCST90025990 | eGFR decline (CKD population) [SH04] | Calcium levels | Inverse variance weighted | 202 | 0,054671 | 0,082906 | 0,50962 | Minerals | 102,0722 |
| ebi-a-GCST90025990 | eGFR decline (CKD population) [SH04] | Calcium levels | Simple mode | 202 | -0,00967 | 0,327428 | 0,976458 | Minerals | 102,0722 |
| ebi-a-GCST90025990 | eGFR decline (CKD population) [SH04] | Calcium levels | Weighted mode | 202 | 0,101118 | 0,169365 | 0,551151 | Minerals | 102,0722 |
| ebi-a-GCST90029013 | eGFR decline (CKD population) [SH04] | Educational attainment (years of education) | MR Egger | 209 | 0,014219 | 0,114321 | 0,901136 | Social trait | 45,05626 |
| ebi-a-GCST90029013 | eGFR decline (CKD population) [SH04] | Educational attainment (years of education) | Weighted median | 209 | -0,01951 | 0,038125 | 0,60887 | Social trait | 45,05626 |
| ebi-a-GCST90029013 | eGFR decline (CKD population) [SH04] | Educational attainment (years of education) | Inverse variance weighted | 209 | -0,04471 | 0,026178 | 0,08764 | Social trait | 45,05626 |
| ebi-a-GCST90029013 | eGFR decline (CKD population) [SH04] | Educational attainment (years of education) | Simple mode | 209 | -0,02233 | 0,105026 | 0,8318 | Social trait | 45,05626 |
| ebi-a-GCST90029013 | eGFR decline (CKD population) [SH04] | Educational attainment (years of education) | Weighted mode | 209 | 0,029026 | 0,092303 | 0,753481 | Social trait | 45,05626 |
| ebi-a-GCST90029014 | eGFR decline (CKD population) [SH04] | Smoking status | MR Egger | 117 | -0,3422 | 1,09302 | 0,754788 | Smoking | 42,91926 |
| ebi-a-GCST90029014 | eGFR decline (CKD population) [SH04] | Smoking status | Weighted median | 117 | -0,02027 | 0,377548 | 0,957178 | Smoking | 42,91926 |
| ebi-a-GCST90029014 | eGFR decline (CKD population) [SH04] | Smoking status | Inverse variance weighted | 117 | -0,00097 | 0,258901 | 0,997004 | Smoking | 42,91926 |
| ebi-a-GCST90029014 | eGFR decline (CKD population) [SH04] | Smoking status | Simple mode | 117 | -0,45862 | 0,883596 | 0,604721 | Smoking | 42,91926 |
| ebi-a-GCST90029014 | eGFR decline (CKD population) [SH04] | Smoking status | Weighted mode | 117 | -0,27041 | 0,744818 | 0,717227 | Smoking | 42,91926 |
| ebi-a-GCST90029070 | eGFR decline (CKD population) [SH04] | C-reactive protein levels | MR Egger | 214 | 0,25579 | 0,130084 | 0,050565 | Inflammation | 122,6906 |
| ebi-a-GCST90029070 | eGFR decline (CKD population) [SH04] | C-reactive protein levels | Weighted median | 214 | 0,043622 | 0,126505 | 0,730224 | Inflammation | 122,6906 |
| ebi-a-GCST90029070 | eGFR decline (CKD population) [SH04] | C-reactive protein levels | Inverse variance weighted | 214 | 0,089125 | 0,076712 | 0,245315 | Inflammation | 122,6906 |
| ebi-a-GCST90029070 | eGFR decline (CKD population) [SH04] | C-reactive protein levels | Simple mode | 214 | -0,31258 | 0,266703 | 0,242502 | Inflammation | 122,6906 |
| ebi-a-GCST90029070 | eGFR decline (CKD population) [SH04] | C-reactive protein levels | Weighted mode | 214 | 0,151535 | 0,131764 | 0,251414 | Inflammation | 122,6906 |
| ebi-a-GCST90029073 | eGFR decline (CKD population) [SH04] | Liver fat | MR Egger | 8 | 0,298842 | 0,159294 | 0,109756 | Fatty liver | 140,8023 |
| ebi-a-GCST90029073 | eGFR decline (CKD population) [SH04] | Liver fat | Weighted median | 8 | 0,154881 | 0,118647 | 0,19176 | Fatty liver | 140,8023 |
| ebi-a-GCST90029073 | eGFR decline (CKD population) [SH04] | Liver fat | Inverse variance weighted | 8 | 0,184827 | 0,09654 | 0,055556 | Fatty liver | 140,8023 |
| ebi-a-GCST90029073 | eGFR decline (CKD population) [SH04] | Liver fat | Simple mode | 8 | 0,252299 | 0,171225 | 0,184109 | Fatty liver | 140,8023 |
| ebi-a-GCST90029073 | eGFR decline (CKD population) [SH04] | Liver fat | Weighted mode | 8 | 0,236054 | 0,114491 | 0,07816 | Fatty liver | 140,8023 |
| ebi-a-GCST90086057 | eGFR decline (CKD population) [SH04] | Cardiovascular disease | Inverse variance weighted | 2 | 0,149471 | 0,211689 | 0,480133 | Cardiovascular disease | 32,018 |
| finn-b-D3_ANAEMIA_B12_DEF | eGFR decline (CKD population) [SH04] | Vitamin B12 deficiency anaemia | MR Egger | 4 | -0,06315 | 0,159294 | 0,730078 | Vitamin | 52,61875 |
| finn-b-D3_ANAEMIA_B12_DEF | eGFR decline (CKD population) [SH04] | Vitamin B12 deficiency anaemia | Weighted median | 4 | -0,04992 | 0,06585 | 0,44842 | Vitamin | 52,61875 |
| finn-b-D3_ANAEMIA_B12_DEF | eGFR decline (CKD population) [SH04] | Vitamin B12 deficiency anaemia | Inverse variance weighted | 4 | -0,02177 | 0,053957 | 0,686609 | Vitamin | 52,61875 |
| finn-b-D3_ANAEMIA_B12_DEF | eGFR decline (CKD population) [SH04] | Vitamin B12 deficiency anaemia | Simple mode | 4 | -0,06192 | 0,096083 | 0,565156 | Vitamin | 52,61875 |
| finn-b-D3_ANAEMIA_B12_DEF | eGFR decline (CKD population) [SH04] | Vitamin B12 deficiency anaemia | Weighted mode | 4 | -0,06508 | 0,074428 | 0,446269 | Vitamin | 52,61875 |
| finn-b-E4_OBESITY | eGFR decline (CKD population) [SH04] | Obesity | MR Egger | 7 | 0,005269 | 0,28828 | 0,986125 | Obesity | 59,53804 |
| finn-b-E4_OBESITY | eGFR decline (CKD population) [SH04] | Obesity | Weighted median | 7 | -0,0282 | 0,096011 | 0,769004 | Obesity | 59,53804 |
| finn-b-E4_OBESITY | eGFR decline (CKD population) [SH04] | Obesity | Inverse variance weighted | 7 | 0,02747 | 0,078758 | 0,727249 | Obesity | 59,53804 |
| finn-b-E4_OBESITY | eGFR decline (CKD population) [SH04] | Obesity | Simple mode | 7 | -0,01985 | 0,162402 | 0,906732 | Obesity | 59,53804 |
| finn-b-E4_OBESITY | eGFR decline (CKD population) [SH04] | Obesity | Weighted mode | 7 | -0,03818 | 0,100916 | 0,718181 | Obesity | 59,53804 |
| finn-b-FG_PULMHEART | eGFR decline (CKD population) [SH04] | Pulmonary heart disease | MR Egger | 5 | -0,20491 | 0,15245 | 0,271529 | Cardiovascular disease | 89,00197 |
| finn-b-FG_PULMHEART | eGFR decline (CKD population) [SH04] | Pulmonary heart disease | Weighted median | 5 | 0,062983 | 0,065552 | 0,336641 | Cardiovascular disease | 89,00197 |
| finn-b-FG_PULMHEART | eGFR decline (CKD population) [SH04] | Pulmonary heart disease | Inverse variance weighted | 5 | 0,015126 | 0,054157 | 0,780016 | Cardiovascular disease | 89,00197 |
| finn-b-FG_PULMHEART | eGFR decline (CKD population) [SH04] | Pulmonary heart disease | Simple mode | 5 | 0,10296 | 0,100944 | 0,365412 | Cardiovascular disease | 89,00197 |
| finn-b-FG_PULMHEART | eGFR decline (CKD population) [SH04] | Pulmonary heart disease | Weighted mode | 5 | 0,105607 | 0,101919 | 0,358646 | Cardiovascular disease | 89,00197 |
| ieu-b-102 | eGFR decline (CKD population) [SH04] | Major depression | MR Egger | 48 | 0,151195 | 0,818137 | 0,854195 | Mental health | 38,29386 |
| ieu-b-102 | eGFR decline (CKD population) [SH04] | Major depression | Weighted median | 48 | -0,05989 | 0,21025 | 0,775753 | Mental health | 38,29386 |
| ieu-b-102 | eGFR decline (CKD population) [SH04] | Major depression | Inverse variance weighted | 48 | -0,09045 | 0,145764 | 0,534916 | Mental health | 38,29386 |
| ieu-b-102 | eGFR decline (CKD population) [SH04] | Major depression | Simple mode | 48 | 0,019624 | 0,47956 | 0,967533 | Mental health | 38,29386 |
| ieu-b-102 | eGFR decline (CKD population) [SH04] | Major depression | Weighted mode | 48 | -0,01805 | 0,440845 | 0,967522 | Mental health | 38,29386 |
| ieu-b-107 | eGFR decline (CKD population) [SH04] | apolipoprotein A-I | MR Egger | 269 | -0,1117 | 0,112924 | 0,323461 | Dyslipidemias | 139,7111 |
| ieu-b-107 | eGFR decline (CKD population) [SH04] | apolipoprotein A-I | Weighted median | 269 | -0,12511 | 0,11334 | 0,269671 | Dyslipidemias | 139,7111 |
| ieu-b-107 | eGFR decline (CKD population) [SH04] | apolipoprotein A-I | Inverse variance weighted | 269 | -0,05483 | 0,066826 | 0,41196 | Dyslipidemias | 139,7111 |
| ieu-b-107 | eGFR decline (CKD population) [SH04] | apolipoprotein A-I | Simple mode | 269 | -0,11928 | 0,230215 | 0,604807 | Dyslipidemias | 139,7111 |
| ieu-b-107 | eGFR decline (CKD population) [SH04] | apolipoprotein A-I | Weighted mode | 269 | -0,14799 | 0,127034 | 0,245082 | Dyslipidemias | 139,7111 |
| ieu-b-108 | eGFR decline (CKD population) [SH04] | apolipoprotein B | MR Egger | 173 | 0,070445 | 0,122926 | 0,567349 | Dyslipidemias | 162,3296 |
| ieu-b-108 | eGFR decline (CKD population) [SH04] | apolipoprotein B | Weighted median | 173 | 0,203417 | 0,118237 | 0,085356 | Dyslipidemias | 162,3296 |
| ieu-b-108 | eGFR decline (CKD population) [SH04] | apolipoprotein B | Inverse variance weighted | 173 | 0,082703 | 0,075706 | 0,27465 | Dyslipidemias | 162,3296 |
| ieu-b-108 | eGFR decline (CKD population) [SH04] | apolipoprotein B | Simple mode | 173 | 0,13895 | 0,230888 | 0,548095 | Dyslipidemias | 162,3296 |
| ieu-b-108 | eGFR decline (CKD population) [SH04] | apolipoprotein B | Weighted mode | 173 | 0,183398 | 0,131092 | 0,163614 | Dyslipidemias | 162,3296 |
| ieu-b-25 | eGFR decline (CKD population) [SH04] | Cigarettes per Day | MR Egger | 22 | -0,25458 | 0,171937 | 0,154286 | Smoking | 99,80012 |
| ieu-b-25 | eGFR decline (CKD population) [SH04] | Cigarettes per Day | Weighted median | 22 | -0,1697 | 0,138237 | 0,219607 | Smoking | 99,80012 |
| ieu-b-25 | eGFR decline (CKD population) [SH04] | Cigarettes per Day | Inverse variance weighted | 22 | -0,1677 | 0,098933 | 0,090055 | Smoking | 99,80012 |
| ieu-b-25 | eGFR decline (CKD population) [SH04] | Cigarettes per Day | Simple mode | 22 | -0,46859 | 0,28588 | 0,116085 | Smoking | 99,80012 |
| ieu-b-25 | eGFR decline (CKD population) [SH04] | Cigarettes per Day | Weighted mode | 22 | -0,14277 | 0,138794 | 0,315346 | Smoking | 99,80012 |
| ieu-b-4861 | eGFR decline (CKD population) [SH04] | Chronotype | MR Egger | 60 | -0,44334 | 0,371732 | 0,237868 | Sleep disorders | 44,49296 |
| ieu-b-4861 | eGFR decline (CKD population) [SH04] | Chronotype | Weighted median | 60 | -0,08499 | 0,192491 | 0,65884 | Sleep disorders | 44,49296 |
| ieu-b-4861 | eGFR decline (CKD population) [SH04] | Chronotype | Inverse variance weighted | 60 | 0,00697 | 0,134436 | 0,95865 | Sleep disorders | 44,49296 |
| ieu-b-4861 | eGFR decline (CKD population) [SH04] | Chronotype | Simple mode | 60 | -0,17632 | 0,483775 | 0,716807 | Sleep disorders | 44,49296 |
| ieu-b-4861 | eGFR decline (CKD population) [SH04] | Chronotype | Weighted mode | 60 | -0,1629 | 0,418132 | 0,698244 | Sleep disorders | 44,49296 |
| ieu-b-4877 | eGFR decline (CKD population) [SH04] | smoking initiation | MR Egger | 85 | 1,501068 | 0,598267 | 0,014051 | Smoking | 42,2587 |
| ieu-b-4877 | eGFR decline (CKD population) [SH04] | smoking initiation | Weighted median | 85 | 0,156128 | 0,164828 | 0,343529 | Smoking | 42,2587 |
| ieu-b-4877 | eGFR decline (CKD population) [SH04] | smoking initiation | Inverse variance weighted | 85 | 0,174421 | 0,117639 | 0,138159 | Smoking | 42,2587 |
| ieu-b-4877 | eGFR decline (CKD population) [SH04] | smoking initiation | Simple mode | 85 | 0,291092 | 0,411538 | 0,481321 | Smoking | 42,2587 |
| ieu-b-4877 | eGFR decline (CKD population) [SH04] | smoking initiation | Weighted mode | 85 | 0,221523 | 0,34899 | 0,527313 | Smoking | 42,2587 |
| ieu-b-5144 | eGFR decline (CKD population) [SH04] | Hypertension | MR Egger | 187 | -1,33756 | 0,89406 | 0,136344 | Hypertension | 54,118 |
| ieu-b-5144 | eGFR decline (CKD population) [SH04] | Hypertension | Weighted median | 187 | -0,58666 | 0,415528 | 0,157994 | Hypertension | 54,118 |
| ieu-b-5144 | eGFR decline (CKD population) [SH04] | Hypertension | Inverse variance weighted | 187 | -0,30457 | 0,287239 | 0,288995 | Hypertension | 54,118 |
| ieu-b-5144 | eGFR decline (CKD population) [SH04] | Hypertension | Simple mode | 187 | -0,98473 | 1,040393 | 0,345121 | Hypertension | 54,118 |
| ieu-b-5144 | eGFR decline (CKD population) [SH04] | Hypertension | Weighted mode | 187 | -1,42613 | 0,872898 | 0,103995 | Hypertension | 54,118 |
| ieu-b-72 | eGFR decline (CKD population) [SH04] | Urinary sodium-potassium ratio | MR Egger | 24 | 0,98447 | 1,569786 | 0,537025 | Minerals | 39,88328 |
| ieu-b-72 | eGFR decline (CKD population) [SH04] | Urinary sodium-potassium ratio | Weighted median | 24 | 0,24018 | 0,488208 | 0,622746 | Minerals | 39,88328 |
| ieu-b-72 | eGFR decline (CKD population) [SH04] | Urinary sodium-potassium ratio | Inverse variance weighted | 24 | 0,266722 | 0,362241 | 0,461542 | Minerals | 39,88328 |
| ieu-b-72 | eGFR decline (CKD population) [SH04] | Urinary sodium-potassium ratio | Simple mode | 24 | -0,36328 | 0,915069 | 0,695028 | Minerals | 39,88328 |
| ieu-b-72 | eGFR decline (CKD population) [SH04] | Urinary sodium-potassium ratio | Weighted mode | 24 | -0,15673 | 0,893667 | 0,862319 | Minerals | 39,88328 |
| prot-a-3102 | eGFR decline (CKD population) [SH04] | Thyrotropin-releasing hormone | Inverse variance weighted | 2 | 0,143143 | 0,134041 | 0,285565 | Thyroid disease | 36,35446 |
| prot-c-2652_15_1 | eGFR decline (CKD population) [SH04] | suPAR | Wald ratio | 1 | 0,028847 | 0,103892 | 0,78127 | Biomarkers | 33,5125 |
| ukb-b-4424 | eGFR decline (CKD population) [SH04] | Sleep duration | MR Egger | 66 | -0,2027 | 1,139108 | 0,859325 | Sleep disorders | 42,80283 |
| ukb-b-4424 | eGFR decline (CKD population) [SH04] | Sleep duration | Weighted median | 66 | -0,23688 | 0,443993 | 0,593667 | Sleep disorders | 42,80283 |
| ukb-b-4424 | eGFR decline (CKD population) [SH04] | Sleep duration | Inverse variance weighted | 66 | -0,19004 | 0,29867 | 0,52459 | Sleep disorders | 42,80283 |
| ukb-b-4424 | eGFR decline (CKD population) [SH04] | Sleep duration | Simple mode | 66 | -0,53622 | 0,95318 | 0,575668 | Sleep disorders | 42,80283 |
| ukb-b-4424 | eGFR decline (CKD population) [SH04] | Sleep duration | Weighted mode | 66 | -0,41412 | 0,766035 | 0,590628 | Sleep disorders | 42,80283 |
| ukb-b-5779 | eGFR decline (CKD population) [SH04] | Alcohol intake frequency. | MR Egger | 94 | -0,02543 | 0,271494 | 0,925573 | Alcohol consumption | 53,13815 |
| ukb-b-5779 | eGFR decline (CKD population) [SH04] | Alcohol intake frequency. | Weighted median | 94 | 0,070103 | 0,21659 | 0,746192 | Alcohol consumption | 53,13815 |
| ukb-b-5779 | eGFR decline (CKD population) [SH04] | Alcohol intake frequency. | Inverse variance weighted | 94 | 0,02068 | 0,119689 | 0,86282 | Alcohol consumption | 53,13815 |
| ukb-b-5779 | eGFR decline (CKD population) [SH04] | Alcohol intake frequency. | Simple mode | 94 | -0,32745 | 0,411704 | 0,42843 | Alcohol consumption | 53,13815 |
| ukb-b-5779 | eGFR decline (CKD population) [SH04] | Alcohol intake frequency. | Weighted mode | 94 | 0,025431 | 0,253845 | 0,920414 | Alcohol consumption | 53,13815 |
| ukb-b-7864 | eGFR decline (CKD population) [SH04] | Vitamin B6 | Wald ratio | 1 | -0,89546 | 0,836044 | 0,284141 | Vitamin | 32,75897 |
| ukb-d-30790_irnt | eGFR decline (CKD population) [SH04] | Lipoprotein A | MR Egger | 11 | -0,1037 | 0,193119 | 0,604319 | Dyslipidemias | 985,6435 |
| ukb-d-30790_irnt | eGFR decline (CKD population) [SH04] | Lipoprotein A | Weighted median | 11 | -0,12359 | 0,135684 | 0,362371 | Dyslipidemias | 985,6435 |
| ukb-d-30790_irnt | eGFR decline (CKD population) [SH04] | Lipoprotein A | Inverse variance weighted | 11 | -0,06487 | 0,124914 | 0,603532 | Dyslipidemias | 985,6435 |
| ukb-d-30790_irnt | eGFR decline (CKD population) [SH04] | Lipoprotein A | Simple mode | 11 | -0,08718 | 0,194571 | 0,663653 | Dyslipidemias | 985,6435 |
| ukb-d-30790_irnt | eGFR decline (CKD population) [SH04] | Lipoprotein A | Weighted mode | 11 | -0,17726 | 0,174504 | 0,33367 | Dyslipidemias | 985,6435 |

| exposure | outcome | method | Q | Q_df | Q_pval |
|---|---|---|---|---|---|
| Body mass index | Incident CKD | Inverse variance weighted | 702,4831676 | 505 | 1,27178E-08 |
| Heel bone mineral density | Incident CKD | Inverse variance weighted | 738,7904663 | 497 | 9,64785E-12 |
| Two-hour glucose | Incident CKD | Inverse variance weighted | 35,7802392 | 13 | 0,00064177 |
| Fasting glucose | Incident CKD | Inverse variance weighted | 152,6914517 | 68 | 1,88578E-08 |
| Fasting insulin | Incident CKD | Inverse variance weighted | 77,6691778 | 36 | 6,86687E-05 |
| circulating leptin levels | Incident CKD | Inverse variance weighted | 4,866342659 | 2 | 0,087758081 |
| Growth differentiation factor-15 levels | Incident CKD | Inverse variance weighted | 8,076699377 | 2 | 0,017626538 |
| Fibroblast growth factor 23 levels | Incident CKD | Inverse variance weighted | 91,18483622 | 7 | 7,06405E-17 |
| Kidney injury molecule 1 levels | Incident CKD | Inverse variance weighted | 338,4604322 | 8 | 2,62576E-68 |
| Coronary artery disease (Firth correction) | Incident CKD | Inverse variance weighted | 85,60338251 | 38 | 1,5749E-05 |
| Body fat percentage | Incident CKD | Inverse variance weighted | 492,2808834 | 317 | 9,57243E-10 |
| Sodium in urine | Incident CKD | Inverse variance weighted | 95,84004714 | 40 | 1,75312E-06 |
| Glycated haemoglobin HbA1c levels | Incident CKD | Inverse variance weighted | 545,3763434 | 315 | 1,41093E-14 |
| Phosphate levels | Incident CKD | Inverse variance weighted | 463,4799419 | 161 | 4,49463E-31 |
| Bone mineral density | Incident CKD | Inverse variance weighted | 649,9052642 | 423 | 7,46623E-12 |
| Type 1 diabetes | Incident CKD | Inverse variance weighted | 106,638774 | 76 | 0,011747223 |
| Liver iron content | Incident CKD | Inverse variance weighted | 2,513580215 | 4 | 0,642205707 |
| Chronic heart failure | Incident CKD | Inverse variance weighted | 7,220537795 | 5 | 0,204748511 |
| Hyperthyroidism | Incident CKD | Inverse variance weighted | 14,58476819 | 7 | 0,041706111 |
| Hypothyroidism | Incident CKD | Inverse variance weighted | 108,7305475 | 67 | 0,000957415 |
| Myocardial infarction | Incident CKD | Inverse variance weighted | 192,5110848 | 74 | 1,69391E-12 |
| Serum uric acid levels | Incident CKD | Inverse variance weighted | 1364,404189 | 253 | 1,3902E-151 |
| Vitamin K-dependent protein S measurement | Incident CKD | Inverse variance weighted | 1,886895783 | 3 | 0,596210376 |
| Vitamin K-dependent protein C measurement | Incident CKD | Inverse variance weighted | 3,029530676 | 3 | 0,387094499 |
| Serum alkaline phosphatase levels | Incident CKD | Inverse variance weighted | 613,1581335 | 343 | 1,51995E-17 |
| Vitamin D levels | Incident CKD | Inverse variance weighted | 196,8114039 | 83 | 3,08258E-11 |
| Hemoglobin levels | Incident CKD | Inverse variance weighted | 1106,141543 | 329 | 9,81106E-85 |
| Blood glucose levels | Incident CKD | Inverse variance weighted | 200,7883082 | 109 | 2,06829E-07 |
| Insulin-like growth factor 1 levels | Incident CKD | Inverse variance weighted | 686,3725353 | 381 | 8,67573E-20 |
| Calcium levels | Incident CKD | Inverse variance weighted | 611,4628798 | 213 | 3,65141E-40 |
| Educational attainment (years of education) | Incident CKD | Inverse variance weighted | 303,212949 | 209 | 2,1884E-05 |
| Smoking status | Incident CKD | Inverse variance weighted | 125,4260674 | 116 | 0,259029705 |
| C-reactive protein levels | Incident CKD | Inverse variance weighted | 364,4085638 | 227 | 1,85784E-08 |
| Liver fat | Incident CKD | Inverse variance weighted | 31,93071001 | 7 | 4,18349E-05 |
| Cardiovascular disease | Incident CKD | Inverse variance weighted | 5,62297538 | 1 | 0,01772653 |
| Serum iron | Incident CKD | Inverse variance weighted | 0,146066293 | 1 | 0,702323255 |
| Vitamin B12 deficiency anaemia | Incident CKD | Inverse variance weighted | 0,352303046 | 3 | 0,949909336 |
| Obesity | Incident CKD | Inverse variance weighted | 11,35314611 | 7 | 0,123939564 |
| Pulmonary heart disease | Incident CKD | Inverse variance weighted | 10,84569251 | 5 | 0,054526753 |
| Free triiodothyronine | Incident CKD | Inverse variance weighted | 4,350042179 | 5 | 0,50019434 |
| FT3 FT4 ratio | Incident CKD | Inverse variance weighted | 7,499605259 | 12 | 0,822911511 |
| Free thyroxine | Incident CKD | Inverse variance weighted | 140,5320404 | 62 | 4,97727E-08 |
| HDL cholesterol | Incident CKD | Inverse variance weighted | 593,3537561 | 248 | 2,46676E-30 |
| High TSH | Incident CKD | Inverse variance weighted | 11,94739273 | 5 | 0,035515775 |
| Major depression | Incident CKD | Inverse variance weighted | 63,86226962 | 47 | 0,0512164 |
| apolipoprotein A-I | Incident CKD | Inverse variance weighted | 593,624859 | 284 | 5,18881E-24 |
| apolipoprotein B | Incident CKD | Inverse variance weighted | 412,1350996 | 182 | 6,94096E-20 |
| Cigarettes per Day | Incident CKD | Inverse variance weighted | 21,83358465 | 21 | 0,409153605 |
| Chronotype | Incident CKD | Inverse variance weighted | 99,3546444 | 61 | 0,001395405 |
| smoking initiation | Incident CKD | Inverse variance weighted | 99,69259251 | 84 | 0,11637503 |
| Systolic Blood Pressure (SBP) | Incident CKD | Inverse variance weighted | 612,4087073 | 256 | 3,10894E-31 |
| Diastolic Blood Pressure (DBP) | Incident CKD | Inverse variance weighted | 866,0424946 | 258 | 9,54701E-67 |
| Pulse Pressure (PP) | Incident CKD | Inverse variance weighted | 694,6081713 | 309 | 1,05321E-31 |
| Hypertension | Incident CKD | Inverse variance weighted | 792,3011345 | 191 | 3,5078E-74 |
| Urinary sodium-potassium ratio | Incident CKD | Inverse variance weighted | 63,50775038 | 23 | 1,17033E-05 |
| Insomnia | Incident CKD | Inverse variance weighted | 8,784329628 | 12 | 0,721232824 |
| LDL cholesterol | Incident CKD | Inverse variance weighted | 455,2668505 | 227 | 1,99562E-17 |
| Low TSH | Incident CKD | Inverse variance weighted | 133,0948049 | 27 | 5,5388E-16 |
| Thyrotropin-releasing hormone | Incident CKD | Inverse variance weighted | 0,197763148 | 1 | 0,656532459 |
| Compound smoking index | Incident CKD | Inverse variance weighted | 136,1405548 | 120 | 0,148915025 |
| Type 2 diabetes | Incident CKD | Inverse variance weighted | 642,4647068 | 449 | 5,00486E-09 |
| Total cholesterol | Incident CKD | Inverse variance weighted | 555,0593729 | 275 | 4,38433E-21 |
| Triglyceride | Incident CKD | Inverse variance weighted | 464,695966 | 225 | 8,46002E-19 |
| Thyroid Stimulating Hormone | Incident CKD | Inverse variance weighted | 245,2487901 | 163 | 3,26492E-05 |
| Sleep duration | Incident CKD | Inverse variance weighted | 121,4767273 | 65 | 2,74403E-05 |
| Alcohol intake frequency. | Incident CKD | Inverse variance weighted | 176,7079044 | 94 | 5,2897E-07 |
| Lipoprotein A | Incident CKD | Inverse variance weighted | 17,3106191 | 18 | 0,501855358 |
| Waist-to-hip ratio adjusted BMI | Incident CKD | Inverse variance weighted | 550,0399645 | 296 | 1,72926E-17 |
| Zinc | Incident CKD | Inverse variance weighted | 2,208318496 | 2 | 0,331489465 |
| Body mass index | eGFR decline (CKD population) | Inverse variance weighted | 481,2979624 | 503 | 0,749598667 |
| Heel bone mineral density | eGFR decline (CKD population) | Inverse variance weighted | 427,3931139 | 482 | 0,964717865 |
| Two-hour glucose | eGFR decline (CKD population) | Inverse variance weighted | 13,74713399 | 13 | 0,391890076 |
| Fasting glucose | eGFR decline (CKD population) | Inverse variance weighted | 75,24846013 | 67 | 0,228947636 |
| Fasting insulin | eGFR decline (CKD population) | Inverse variance weighted | 55,47465556 | 37 | 0,026021588 |
| circulating leptin levels | eGFR decline (CKD population) | Inverse variance weighted | 2,981214688 | 2 | 0,225235818 |

| | | | | | |
|---|---|---|---|---|---|
| Growth differentiation factor-15 levels | eGFR decline (CKD population) | Inverse variance weighted | 0,695617579 | 2 | 0,706233903 |
| Fibroblast growth factor 23 levels | eGFR decline (CKD population) | Inverse variance weighted | 6,266596632 | 7 | 0,508988002 |
| Kidney injury molecule 1 levels | eGFR decline (CKD population) | Inverse variance weighted | 34,72804607 | 8 | 2,99666E-05 |
| Coronary artery disease (Firth correction) | eGFR decline (CKD population) | Inverse variance weighted | 24,3283987 | 37 | 0,945820259 |
| Body fat percentage | eGFR decline (CKD population) | Inverse variance weighted | 348,6694455 | 320 | 0,12981921 |
| Sodium in urine | eGFR decline (CKD population) | Inverse variance weighted | 30,95474771 | 40 | 0,846930697 |
| Glycated haemoglobin HbA1c levels | eGFR decline (CKD population) | Inverse variance weighted | 290,9199188 | 303 | 0,680901191 |
| Phosphate levels | eGFR decline (CKD population) | Inverse variance weighted | 164,2274011 | 160 | 0,393016016 |
| Bone mineral density | eGFR decline (CKD population) | Inverse variance weighted | 409,2204474 | 417 | 0,597915015 |
| Type 1 diabetes | eGFR decline (CKD population) | Inverse variance weighted | 59,51640672 | 75 | 0,904567766 |
| Liver iron content | eGFR decline (CKD population) | Inverse variance weighted | 3,711936465 | 5 | 0,591588644 |
| Chronic heart failure | eGFR decline (CKD population) | Inverse variance weighted | 1,620473972 | 4 | 0,805107703 |
| Hyperthyroidism | eGFR decline (CKD population) | Inverse variance weighted | 16,2056976 | 9 | 0,062708369 |
| Hypothyroidism | eGFR decline (CKD population) | Inverse variance weighted | 71,47914683 | 63 | 0,216893832 |
| Myocardial infarction | eGFR decline (CKD population) | Inverse variance weighted | 50,25842413 | 69 | 0,956366346 |
| Serum uric acid levels | eGFR decline (CKD population) | Inverse variance weighted | 252,4191639 | 237 | 0,234566932 |
| Vitamin K-dependent protein S measurement | eGFR decline (CKD population) | Inverse variance weighted | 0,105929612 | 3 | 0,991116475 |
| Vitamin K-dependent protein C measurement | eGFR decline (CKD population) | Inverse variance weighted | 6,330814854 | 2 | 0,042196946 |
| Serum alkaline phosphatase levels | eGFR decline (CKD population) | Inverse variance weighted | 352,7158323 | 334 | 0,230647354 |
| Vitamin D levels | eGFR decline (CKD population) | Inverse variance weighted | 91,02589668 | 80 | 0,187613575 |
| Hemoglobin levels | eGFR decline (CKD population) | Inverse variance weighted | 340,9792801 | 324 | 0,247688471 |
| Blood glucose levels | eGFR decline (CKD population) | Inverse variance weighted | 121,3508104 | 108 | 0,179206254 |
| Insulin-like growth factor 1 levels | eGFR decline (CKD population) | Inverse variance weighted | 365,1866603 | 373 | 0,603969212 |
| Calcium levels | eGFR decline (CKD population) | Inverse variance weighted | 212,3699239 | 204 | 0,329339002 |
| Educational attainment (years of education) | eGFR decline (CKD population) | Inverse variance weighted | 175,0460815 | 208 | 0,953204484 |
| Smoking status | eGFR decline (CKD population) | Inverse variance weighted | 118,6613437 | 116 | 0,414060901 |
| C-reactive protein levels | eGFR decline (CKD population) | Inverse variance weighted | 212,5021919 | 214 | 0,51608032 |
| Liver fat | eGFR decline (CKD population) | Inverse variance weighted | 5,204559433 | 7 | 0,635015538 |
| Cardiovascular disease | eGFR decline (CKD population) | Inverse variance weighted | 0,004058172 | 1 | 0,949206091 |
| Serum iron | eGFR decline (CKD population) | Inverse variance weighted | 0,043050794 | 1 | 0,83562966 |
| Vitamin B12 deficiency anaemia | eGFR decline (CKD population) | Inverse variance weighted | 2,382357828 | 3 | 0,496927157 |
| Obesity | eGFR decline (CKD population) | Inverse variance weighted | 6,589233945 | 6 | 0,36050868 |
| Pulmonary heart disease | eGFR decline (CKD population) | Inverse variance weighted | 3,920455746 | 4 | 0,416878018 |
| Free triiodothyronine | eGFR decline (CKD population) | Inverse variance weighted | 3,380325639 | 5 | 0,641568084 |
| FT3 FT4 ratio | eGFR decline (CKD population) | Inverse variance weighted | 8,274043901 | 12 | 0,763364835 |
| Free thyroxine | eGFR decline (CKD population) | Inverse variance weighted | 49,55164846 | 61 | 0,852704474 |
| HDL cholesterol | eGFR decline (CKD population) | Inverse variance weighted | 265,7141753 | 233 | 0,069387436 |
| High TSH | eGFR decline (CKD population) | Inverse variance weighted | 3,250877808 | 5 | 0,661370248 |
| Major depression | eGFR decline (CKD population) | Inverse variance weighted | 53,95267806 | 47 | 0,225861594 |
| apolipoprotein A-I | eGFR decline (CKD population) | Inverse variance weighted | 257,4855127 | 268 | 0,666889327 |
| apolipoprotein B | eGFR decline (CKD population) | Inverse variance weighted | 142,5223727 | 174 | 0,961228015 |
| Cigarettes per Day | eGFR decline (CKD population) | Inverse variance weighted | 16,96057059 | 21 | 0,713493582 |
| Chronotype | eGFR decline (CKD population) | Inverse variance weighted | 53,24969587 | 58 | 0,652221506 |
| smoking initiation | eGFR decline (CKD population) | Inverse variance weighted | 76,06960386 | 84 | 0,719161983 |
| Systolic Blood Pressure (SBP) | eGFR decline (CKD population) | Inverse variance weighted | 209,8618977 | 253 | 0,977728708 |
| Diastolic Blood Pressure (DBP) | eGFR decline (CKD population) | Inverse variance weighted | 276,4658424 | 256 | 0,181223981 |
| Pulse Pressure (PP) | eGFR decline (CKD population) | Inverse variance weighted | 270,5970872 | 303 | 0,909719229 |
| Hypertension | eGFR decline (CKD population) | Inverse variance weighted | 171,2096031 | 191 | 0,845040927 |
| Urinary sodium-potassium ratio | eGFR decline (CKD population) | Inverse variance weighted | 26,83992057 | 23 | 0,262840654 |
| Insomnia | eGFR decline (CKD population) | Inverse variance weighted | 10,72987801 | 12 | 0,552199053 |
| LDL cholesterol | eGFR decline (CKD population) | Inverse variance weighted | 214,5638161 | 217 | 0,534005634 |
| Low TSH | eGFR decline (CKD population) | Inverse variance weighted | 22,83367655 | 25 | 0,587275056 |
| Thyrotropin-releasing hormone | eGFR decline (CKD population) | Inverse variance weighted | 0,003212279 | 1 | 0,954802516 |
| Compound smoking index | eGFR decline (CKD population) | Inverse variance weighted | 114,8200017 | 120 | 0,616421857 |
| Type 2 diabetes | eGFR decline (CKD population) | Inverse variance weighted | 450,6907556 | 440 | 0,351995778 |
| Total cholesterol | eGFR decline (CKD population) | Inverse variance weighted | 220,9285191 | 263 | 0,972127371 |
| Triglyceride | eGFR decline (CKD population) | Inverse variance weighted | 194,8850031 | 219 | 0,878057706 |
| Thyroid Stimulating Hormone | eGFR decline (CKD population) | Inverse variance weighted | 144,8928097 | 158 | 0,764543217 |
| Sleep duration | eGFR decline (CKD population) | Inverse variance weighted | 43,20077398 | 65 | 0,98300915 |
| Alcohol intake frequency. | eGFR decline (CKD population) | Inverse variance weighted | 89,46619127 | 93 | 0,584494212 |
| Lipoprotein A | eGFR decline (CKD population) | Inverse variance weighted | 19,91306816 | 10 | 0,030085722 |
| Waist-to-hip ratio adjusted BMI | eGFR decline (CKD population) | Inverse variance weighted | 300,8430596 | 295 | 0,394969225 |
| Zinc | eGFR decline (CKD population) | Inverse variance weighted | 3,335665656 | 2 | 0,188655472 |
| Body mass index | eGFR decline (General population) | Inverse variance weighted | 514,4301718 | 504 | 0,364203876 |
| Heel bone mineral density | eGFR decline (General population) | Inverse variance weighted | 542,5717291 | 498 | 0,081776755 |
| Two-hour glucose | eGFR decline (General population) | Inverse variance weighted | 9,97713646 | 13 | 0,695802925 |
| Fasting glucose | eGFR decline (General population) | Inverse variance weighted | 77,16509133 | 68 | 0,209049621 |
| Fasting insulin | eGFR decline (General population) | Inverse variance weighted | 51,66620501 | 37 | 0,055246305 |
| circulating leptin levels | eGFR decline (General population) | Inverse variance weighted | 1,2112577 | 2 | 0,545731136 |
| Growth differentiation factor-15 levels | eGFR decline (General population) | Inverse variance weighted | 4,06356783 | 2 | 0,131101439 |
| Fibroblast growth factor 23 levels | eGFR decline (General population) | Inverse variance weighted | 4,839297087 | 7 | 0,679567081 |
| Kidney injury molecule 1 levels | eGFR decline (General population) | Inverse variance weighted | 192,4623325 | 9 | 1,25665E-36 |
| Coronary artery disease (Firth correction) | eGFR decline (General population) | Inverse variance weighted | 41,84289641 | 39 | 0,348445817 |
| Body fat percentage | eGFR decline (General population) | Inverse variance weighted | 358,0540977 | 327 | 0,114344632 |
| Sodium in urine | eGFR decline (General population) | Inverse variance weighted | 59,32159517 | 40 | 0,025101508 |
| Glycated haemoglobin HbA1c levels | eGFR decline (General population) | Inverse variance weighted | 366,1504609 | 324 | 0,05316156 |

| | | | | | |
|---|---|---|---|---|---|
| Phosphate levels | eGFR decline (General population) | Inverse variance weighted | 184,4683941 | 166 | 0,155164403 |
| Bone mineral density | eGFR decline (General population) | Inverse variance weighted | 425,5023818 | 436 | 0,631475492 |
| Type 1 diabetes | eGFR decline (General population) | Inverse variance weighted | 84,42913564 | 77 | 0,26312011 |
| Liver iron content | eGFR decline (General population) | Inverse variance weighted | 4,927848451 | 5 | 0,424749045 |
| Chronic heart failure | eGFR decline (General population) | Inverse variance weighted | 2,199494981 | 5 | 0,820908913 |
| Hyperthyroidism | eGFR decline (General population) | Inverse variance weighted | 10,48634906 | 10 | 0,398908339 |
| Hypothyroidism | eGFR decline (General population) | Inverse variance weighted | 69,52931444 | 67 | 0,39232572 |
| Iron deficiency anemia | eGFR decline (General population) | Inverse variance weighted | 5,420929978 | 1 | 0,019896759 |
| Myocardial infarction | eGFR decline (General population) | Inverse variance weighted | 96,01697784 | 76 | 0,060257705 |
| Serum uric acid levels | eGFR decline (General population) | Inverse variance weighted | 449,316496 | 259 | 2,11134E-12 |
| Vitamin K-dependent protein S measurement | eGFR decline (General population) | Inverse variance weighted | 9,844900685 | 3 | 0,019931606 |
| Vitamin K-dependent protein C measurement | eGFR decline (General population) | Inverse variance weighted | 11,82894392 | 3 | 0,007992698 |
| Serum alkaline phosphatase levels | eGFR decline (General population) | Inverse variance weighted | 332,265704 | 350 | 0,744297731 |
| Vitamin D levels | eGFR decline (General population) | Inverse variance weighted | 85,99867743 | 83 | 0,389118744 |
| Hemoglobin levels | eGFR decline (General population) | Inverse variance weighted | 432,0103019 | 330 | 0,000132222 |
| Blood glucose levels | eGFR decline (General population) | Inverse variance weighted | 105,2656345 | 112 | 0,660752518 |
| Insulin-like growth factor 1 levels | eGFR decline (General population) | Inverse variance weighted | 436,6754418 | 383 | 0,030098368 |
| Calcium levels | eGFR decline (General population) | Inverse variance weighted | 264,7483146 | 216 | 0,013189245 |
| Educational attainment (years of education) | eGFR decline (General population) | Inverse variance weighted | 209,5226242 | 210 | 0,496316874 |
| Smoking status | eGFR decline (General population) | Inverse variance weighted | 123,1313074 | 116 | 0,307659289 |
| C-reactive protein levels | eGFR decline (General population) | Inverse variance weighted | 259,4480678 | 230 | 0,088633588 |
| Liver fat | eGFR decline (General population) | Inverse variance weighted | 8,378835083 | 8 | 0,397365834 |
| Cardiovascular disease | eGFR decline (General population) | Inverse variance weighted | 2,735467803 | 1 | 0,098142759 |
| Serum iron | eGFR decline (General population) | Inverse variance weighted | 0,05732284 | 1 | 0,81077846 |
| Vitamin B12 deficiency anaemia | eGFR decline (General population) | Inverse variance weighted | 0,186506837 | 3 | 0,979737589 |
| Obesity | eGFR decline (General population) | Inverse variance weighted | 19,85866671 | 7 | 0,005883188 |
| Pulmonary heart disease | eGFR decline (General population) | Inverse variance weighted | 6,987248163 | 5 | 0,221590407 |
| Free triiodothyronine | eGFR decline (General population) | Inverse variance weighted | 3,519852323 | 5 | 0,620385871 |
| FT3 FT4 ratio | eGFR decline (General population) | Inverse variance weighted | 30,01534955 | 12 | 0,002777611 |
| Free thyroxine | eGFR decline (General population) | Inverse variance weighted | 81,61078038 | 62 | 0,048249469 |
| HDL cholesterol | eGFR decline (General population) | Inverse variance weighted | 291,8445717 | 250 | 0,035497923 |
| High TSH | eGFR decline (General population) | Inverse variance weighted | 7,197315001 | 5 | 0,206374476 |
| Major depression | eGFR decline (General population) | Inverse variance weighted | 23,7511825 | 47 | 0,998129525 |
| apolipoprotein A-I | eGFR decline (General population) | Inverse variance weighted | 329,1143906 | 287 | 0,044001337 |
| apolipoprotein B | eGFR decline (General population) | Inverse variance weighted | 189,8623187 | 188 | 0,448248539 |
| Cigarettes per Day | eGFR decline (General population) | Inverse variance weighted | 28,87441992 | 21 | 0,117049243 |
| Chronotype | eGFR decline (General population) | Inverse variance weighted | 85,02077628 | 61 | 0,022770278 |
| smoking initiation | eGFR decline (General population) | Inverse variance weighted | 87,44552802 | 84 | 0,376924332 |
| Systolic Blood Pressure (SBP) | eGFR decline (General population) | Inverse variance weighted | 291,5773839 | 265 | 0,125754022 |
| Diastolic Blood Pressure (DBP) | eGFR decline (General population) | Inverse variance weighted | 493,8713966 | 268 | 1,3493E-15 |
| Pulse Pressure (PP) | eGFR decline (General population) | Inverse variance weighted | 332,2549751 | 316 | 0,253910822 |
| Hypertension | eGFR decline (General population) | Inverse variance weighted | 442,1973505 | 202 | 5,31196E-20 |
| Urinary sodium-potassium ratio | eGFR decline (General population) | Inverse variance weighted | 16,30146142 | 23 | 0,842062975 |
| Insomnia | eGFR decline (General population) | Inverse variance weighted | 2,841885699 | 12 | 0,996561086 |
| LDL cholesterol | eGFR decline (General population) | Inverse variance weighted | 248,690323 | 230 | 0,189489272 |
| Low TSH | eGFR decline (General population) | Inverse variance weighted | 32,83806654 | 26 | 0,166848053 |
| Thyrotropin-releasing hormone | eGFR decline (General population) | Inverse variance weighted | 0,26491123 | 1 | 0,606765973 |
| Compound smoking index | eGFR decline (General population) | Inverse variance weighted | 128,1849964 | 119 | 0,266421658 |
| Type 2 diabetes | eGFR decline (General population) | Inverse variance weighted | 481,2940538 | 450 | 0,148708416 |
| Total cholesterol | eGFR decline (General population) | Inverse variance weighted | 266,5280761 | 278 | 0,679034181 |
| Triglyceride | eGFR decline (General population) | Inverse variance weighted | 265,3644267 | 230 | 0,054585634 |
| Thyroid Stimulating Hormone | eGFR decline (General population) | Inverse variance weighted | 165,7427595 | 163 | 0,425459378 |
| Sleep duration | eGFR decline (General population) | Inverse variance weighted | 59,45125291 | 66 | 0,702449641 |
| Alcohol intake frequency. | eGFR decline (General population) | Inverse variance weighted | 99,21165347 | 94 | 0,33661506 |
| Lipoprotein A | eGFR decline (General population) | Inverse variance weighted | 12,8234662 | 19 | 0,847484312 |
| Waist-to-hip ratio adjusted BMI | eGFR decline (General population) | Inverse variance weighted | 349,8076167 | 295 | 0,01549762 |
| Zinc | eGFR decline (General population) | Inverse variance weighted | 1,881576737 | 2 | 0,390319998 |
| Body mass index | eGFR decline (CKD population) [DB01] | Inverse variance weighted | 481,0872959 | 505 | 0,771436426 |
| Heel bone mineral density | eGFR decline (CKD population) [DB01] | Inverse variance weighted | 433,7312151 | 482 | 0,943871415 |
| Two-hour glucose | eGFR decline (CKD population) [DB01] | Inverse variance weighted | 16,66077653 | 13 | 0,215279001 |
| Fasting glucose | eGFR decline (CKD population) [DB01] | Inverse variance weighted | 71,08345509 | 67 | 0,34340768 |
| Fasting insulin | eGFR decline (CKD population) [DB01] | Inverse variance weighted | 49,47476661 | 36 | 0,066699601 |
| circulating leptin levels | eGFR decline (CKD population) [DB01] | Inverse variance weighted | 2,300436088 | 2 | 0,316567736 |
| Growth differentiation factor-15 levels | eGFR decline (CKD population) [DB01] | Inverse variance weighted | 0,852361673 | 2 | 0,652998245 |
| Fibroblast growth factor 23 levels | eGFR decline (CKD population) [DB01] | Inverse variance weighted | 6,458877233 | 7 | 0,487301986 |
| Kidney injury molecule 1 levels | eGFR decline (CKD population) [DB01] | Inverse variance weighted | 9,73385043 | 8 | 0,284205278 |
| Coronary artery disease (Firth correction) | eGFR decline (CKD population) [DB01] | Inverse variance weighted | 23,18355645 | 36 | 0,951374655 |
| Body fat percentage | eGFR decline (CKD population) [DB01] | Inverse variance weighted | 344,8465687 | 315 | 0,119026287 |
| Sodium in urine | eGFR decline (CKD population) [DB01] | Inverse variance weighted | 33,0665887 | 40 | 0,773182792 |
| Glycated haemoglobin HbA1c levels | eGFR decline (CKD population) [DB01] | Inverse variance weighted | 283,6890498 | 301 | 0,755604121 |
| Phosphate levels | eGFR decline (CKD population) [DB01] | Inverse variance weighted | 137,8977742 | 159 | 0,885385462 |
| Bone mineral density | eGFR decline (CKD population) [DB01] | Inverse variance weighted | 385,5368884 | 414 | 0,838785511 |
| Type 1 diabetes | eGFR decline (CKD population) [DB01] | Inverse variance weighted | 61,69661103 | 75 | 0,864924942 |
| Liver iron content | eGFR decline (CKD population) [DB01] | Inverse variance weighted | 3,762554305 | 4 | 0,439093238 |
| Chronic heart failure | eGFR decline (CKD population) [DB01] | Inverse variance weighted | 1,845381568 | 4 | 0,764169903 |
| Hyperthyroidism | eGFR decline (CKD population) [DB01] | Inverse variance weighted | 14,72776862 | 7 | 0,039651724 |

| | | | | | |
|---|---|---|---|---|---|
| Hypothyroidism | eGFR decline (CKD population) [DB01] | Inverse variance weighted | 79,45965544 | 63 | 0,078770474 |
| Myocardial infarction | eGFR decline (CKD population) [DB01] | Inverse variance weighted | 41,33817402 | 69 | 0,996674223 |
| Serum uric acid levels | eGFR decline (CKD population) [DB01] | Inverse variance weighted | 226,0985537 | 236 | 0,666942032 |
| Vitamin K-dependent protein S measurement | eGFR decline (CKD population) [DB01] | Inverse variance weighted | 0,197892725 | 3 | 0,977928765 |
| Vitamin K-dependent protein C measurement | eGFR decline (CKD population) [DB01] | Inverse variance weighted | 4,828012095 | 2 | 0,08945621 |
| Serum alkaline phosphatase levels | eGFR decline (CKD population) [DB01] | Inverse variance weighted | 334,1438069 | 334 | 0,487491162 |
| Vitamin D levels | eGFR decline (CKD population) [DB01] | Inverse variance weighted | 77,20364313 | 80 | 0,567813541 |
| Hemoglobin levels | eGFR decline (CKD population) [DB01] | Inverse variance weighted | 331,6880402 | 324 | 0,37228779 |
| Blood glucose levels | eGFR decline (CKD population) [DB01] | Inverse variance weighted | 118,972485 | 108 | 0,221330075 |
| Insulin-like growth factor 1 levels | eGFR decline (CKD population) [DB01] | Inverse variance weighted | 356,9926484 | 372 | 0,70306855 |
| Calcium levels | eGFR decline (CKD population) [DB01] | Inverse variance weighted | 185,1524737 | 201 | 0,781923111 |
| Educational attainment (years of education) | eGFR decline (CKD population) [DB01] | Inverse variance weighted | 182,8203336 | 208 | 0,895289421 |
| Smoking status | eGFR decline (CKD population) [DB01] | Inverse variance weighted | 114,8143666 | 116 | 0,513674374 |
| C-reactive protein levels | eGFR decline (CKD population) [DB01] | Inverse variance weighted | 208,8342089 | 213 | 0,567811047 |
| Liver fat | eGFR decline (CKD population) [DB01] | Inverse variance weighted | 2,534607311 | 7 | 0,924468632 |
| Cardiovascular disease | eGFR decline (CKD population) [DB01] | Inverse variance weighted | 0,212989685 | 1 | 0,644434144 |
| Serum iron | eGFR decline (CKD population) [DB01] | Inverse variance weighted | 0,081919836 | 1 | 0,774712372 |
| Vitamin B12 deficiency anaemia | eGFR decline (CKD population) [DB01] | Inverse variance weighted | 2,083649949 | 3 | 0,555227565 |
| Obesity | eGFR decline (CKD population) [DB01] | Inverse variance weighted | 6,934076653 | 6 | 0,326986838 |
| Pulmonary heart disease | eGFR decline (CKD population) [DB01] | Inverse variance weighted | 4,885510574 | 4 | 0,299247832 |
| Free triiodothyronine | eGFR decline (CKD population) [DB01] | Inverse variance weighted | 4,040769798 | 5 | 0,543561165 |
| FT3 FT4 ratio | eGFR decline (CKD population) [DB01] | Inverse variance weighted | 9,042934788 | 12 | 0,69925892 |
| Free thyroxine | eGFR decline (CKD population) [DB01] | Inverse variance weighted | 60,75403631 | 61 | 0,484793696 |
| HDL cholesterol | eGFR decline (CKD population) [DB01] | Inverse variance weighted | 252,9297055 | 232 | 0,164880484 |
| High TSH | eGFR decline (CKD population) [DB01] | Inverse variance weighted | 5,466611594 | 5 | 0,361620933 |
| Major depression | eGFR decline (CKD population) [DB01] | Inverse variance weighted | 52,44155479 | 47 | 0,271215651 |
| apolipoprotein A-I | eGFR decline (CKD population) [DB01] | Inverse variance weighted | 243,0861232 | 268 | 0,860454881 |
| apolipoprotein B | eGFR decline (CKD population) [DB01] | Inverse variance weighted | 145,1820319 | 172 | 0,932190807 |
| Cigarettes per Day | eGFR decline (CKD population) [DB01] | Inverse variance weighted | 15,33276624 | 21 | 0,805891854 |
| Chronotype | eGFR decline (CKD population) [DB01] | Inverse variance weighted | 59,11852991 | 59 | 0,471175633 |
| smoking initiation | eGFR decline (CKD population) [DB01] | Inverse variance weighted | 75,16700915 | 84 | 0,743772617 |
| Systolic Blood Pressure (SBP) | eGFR decline (CKD population) [DB01] | Inverse variance weighted | 194,6478476 | 250 | 0,996042532 |
| Diastolic Blood Pressure (DBP) | eGFR decline (CKD population) [DB01] | Inverse variance weighted | 226,4540288 | 250 | 0,854960912 |
| Pulse Pressure (PP) | eGFR decline (CKD population) [DB01] | Inverse variance weighted | 250,3264128 | 301 | 0,984864741 |
| Hypertension | eGFR decline (CKD population) [DB01] | Inverse variance weighted | 133,9746812 | 186 | 0,998484742 |
| Urinary sodium-potassium ratio | eGFR decline (CKD population) [DB01] | Inverse variance weighted | 27,02689684 | 23 | 0,254824656 |
| Insomnia | eGFR decline (CKD population) [DB01] | Inverse variance weighted | 12,72669406 | 12 | 0,389214746 |
| LDL cholesterol | eGFR decline (CKD population) [DB01] | Inverse variance weighted | 220,4210334 | 216 | 0,403886205 |
| Low TSH | eGFR decline (CKD population) [DB01] | Inverse variance weighted | 25,44947427 | 25 | 0,437423604 |
| Thyrotropin-releasing hormone | eGFR decline (CKD population) [DB01] | Inverse variance weighted | 0,001702975 | 1 | 0,967082951 |
| Compound smoking index | eGFR decline (CKD population) [DB01] | Inverse variance weighted | 107,6766451 | 120 | 0,782606608 |
| Type 2 diabetes | eGFR decline (CKD population) [DB01] | Inverse variance weighted | 434,8606236 | 440 | 0,560181653 |
| Total cholesterol | eGFR decline (CKD population) [DB01] | Inverse variance weighted | 229,4218906 | 263 | 0,933462353 |
| Triglyceride | eGFR decline (CKD population) [DB01] | Inverse variance weighted | 201,9159001 | 218 | 0,77576165 |
| Thyroid Stimulating Hormone | eGFR decline (CKD population) [DB01] | Inverse variance weighted | 160,2445615 | 158 | 0,435202692 |
| Sleep duration | eGFR decline (CKD population) [DB01] | Inverse variance weighted | 49,00036354 | 65 | 0,930377493 |
| Alcohol intake frequency. | eGFR decline (CKD population) [DB01] | Inverse variance weighted | 86,62882435 | 93 | 0,666067851 |
| Lipoprotein A | eGFR decline (CKD population) [DB01] | Inverse variance weighted | 16,22758098 | 10 | 0,093300335 |
| Waist-to-hip ratio adjusted BMI | eGFR decline (CKD population) [DB01] | Inverse variance weighted | 290,5555918 | 295 | 0,562147097 |
| Zinc | eGFR decline (CKD population) [DB01] | Inverse variance weighted | 3,669509215 | 2 | 0,159652675 |
| Body mass index | eGFR decline (CKD population) [SH01] | Inverse variance weighted | 482,9855265 | 505 | 0,752373284 |
| Heel bone mineral density | eGFR decline (CKD population) [SH01] | Inverse variance weighted | 438,2692231 | 482 | 0,923904538 |
| Two-hour glucose | eGFR decline (CKD population) [SH01] | Inverse variance weighted | 17,51532695 | 13 | 0,176803969 |
| Fasting glucose | eGFR decline (CKD population) [SH01] | Inverse variance weighted | 71,04011945 | 67 | 0,344731593 |
| Fasting insulin | eGFR decline (CKD population) [SH01] | Inverse variance weighted | 48,44102661 | 36 | 0,080536263 |
| circulating leptin levels | eGFR decline (CKD population) [SH01] | Inverse variance weighted | 2,187376994 | 2 | 0,334978644 |
| Growth differentiation factor-15 levels | eGFR decline (CKD population) [SH01] | Inverse variance weighted | 0,982283402 | 2 | 0,611927357 |
| Fibroblast growth factor 23 levels | eGFR decline (CKD population) [SH01] | Inverse variance weighted | 7,583074345 | 7 | 0,370787961 |
| Kidney injury molecule 1 levels | eGFR decline (CKD population) [SH01] | Inverse variance weighted | 8,666761919 | 8 | 0,371182419 |
| Coronary artery disease (Firth correction) | eGFR decline (CKD population) [SH01] | Inverse variance weighted | 23,73631627 | 36 | 0,941948559 |
| Body fat percentage | eGFR decline (CKD population) [SH01] | Inverse variance weighted | 346,3326977 | 315 | 0,108316091 |
| Sodium in urine | eGFR decline (CKD population) [SH01] | Inverse variance weighted | 34,27535057 | 40 | 0,725031136 |
| Glycated haemoglobin HbA1c levels | eGFR decline (CKD population) [SH01] | Inverse variance weighted | 285,0127136 | 301 | 0,737677846 |
| Phosphate levels | eGFR decline (CKD population) [SH01] | Inverse variance weighted | 136,5069257 | 159 | 0,901200283 |
| Bone mineral density | eGFR decline (CKD population) [SH01] | Inverse variance weighted | 385,2249796 | 414 | 0,841549999 |
| Type 1 diabetes | eGFR decline (CKD population) [SH01] | Inverse variance weighted | 62,63703592 | 75 | 0,845107925 |
| Liver iron content | eGFR decline (CKD population) [SH01] | Inverse variance weighted | 3,803928269 | 4 | 0,433191087 |
| Chronic heart failure | eGFR decline (CKD population) [SH01] | Inverse variance weighted | 1,961067509 | 4 | 0,742919657 |
| Hyperthyroidism | eGFR decline (CKD population) [SH01] | Inverse variance weighted | 14,99097152 | 7 | 0,036115311 |
| Hypothyroidism | eGFR decline (CKD population) [SH01] | Inverse variance weighted | 81,27086927 | 63 | 0,060489229 |
| Myocardial infarction | eGFR decline (CKD population) [SH01] | Inverse variance weighted | 41,32514968 | 69 | 0,996689956 |
| Serum uric acid levels | eGFR decline (CKD population) [SH01] | Inverse variance weighted | 233,9730284 | 236 | 0,52505835 |
| Vitamin K-dependent protein S measurement | eGFR decline (CKD population) [SH01] | Inverse variance weighted | 0,241124796 | 3 | 0,970692211 |
| Vitamin K-dependent protein C measurement | eGFR decline (CKD population) [SH01] | Inverse variance weighted | 4,441288996 | 2 | 0,108539133 |
| Serum alkaline phosphatase levels | eGFR decline (CKD population) [SH01] | Inverse variance weighted | 333,0580889 | 334 | 0,504258257 |
| Vitamin D levels | eGFR decline (CKD population) [SH01] | Inverse variance weighted | 75,4690787 | 80 | 0,622558512 |

| | | | | | |
|---|---|---|---|---|---|
| Hemoglobin levels | eGFR decline (CKD population) [SH01] | Inverse variance weighted | 338,6202441 | 324 | 0,276920256 |
| Blood glucose levels | eGFR decline (CKD population) [SH01] | Inverse variance weighted | 119,0912275 | 108 | 0,219087368 |
| Insulin-like growth factor 1 levels | eGFR decline (CKD population) [SH01] | Inverse variance weighted | 358,7215014 | 372 | 0,680183323 |
| Calcium levels | eGFR decline (CKD population) [SH01] | Inverse variance weighted | 184,8244531 | 201 | 0,786982298 |
| Educational attainment (years of education) | eGFR decline (CKD population) [SH01] | Inverse variance weighted | 185,6220059 | 208 | 0,865678374 |
| Smoking status | eGFR decline (CKD population) [SH01] | Inverse variance weighted | 113,9953592 | 116 | 0,535271053 |
| C-reactive protein levels | eGFR decline (CKD population) [SH01] | Inverse variance weighted | 209,5617678 | 213 | 0,553748126 |
| Liver fat | eGFR decline (CKD population) [SH01] | Inverse variance weighted | 2,344456597 | 7 | 0,938347397 |
| Cardiovascular disease | eGFR decline (CKD population) [SH01] | Inverse variance weighted | 0,331400941 | 1 | 0,564835325 |
| Serum iron | eGFR decline (CKD population) [SH01] | Inverse variance weighted | 0,091144429 | 1 | 0,762727262 |
| Vitamin B12 deficiency anaemia | eGFR decline (CKD population) [SH01] | Inverse variance weighted | 1,993929059 | 3 | 0,573667707 |
| Obesity | eGFR decline (CKD population) [SH01] | Inverse variance weighted | 7,056524658 | 6 | 0,31565147 |
| Pulmonary heart disease | eGFR decline (CKD population) [SH01] | Inverse variance weighted | 5,075886353 | 4 | 0,279599336 |
| Free triiodothyronine | eGFR decline (CKD population) [SH01] | Inverse variance weighted | 4,180989152 | 5 | 0,523663246 |
| FT3 FT4 ratio | eGFR decline (CKD population) [SH01] | Inverse variance weighted | 9,179888381 | 12 | 0,68749362 |
| Free thyroxine | eGFR decline (CKD population) [SH01] | Inverse variance weighted | 63,95969794 | 61 | 0,373003895 |
| HDL cholesterol | eGFR decline (CKD population) [SH01] | Inverse variance weighted | 253,7396422 | 232 | 0,156234919 |
| High TSH | eGFR decline (CKD population) [SH01] | Inverse variance weighted | 5,997857785 | 5 | 0,306427421 |
| Major depression | eGFR decline (CKD population) [SH01] | Inverse variance weighted | 52,18090258 | 47 | 0,279566464 |
| apolipoprotein A-I | eGFR decline (CKD population) [SH01] | Inverse variance weighted | 243,7817949 | 268 | 0,853215923 |
| apolipoprotein B | eGFR decline (CKD population) [SH01] | Inverse variance weighted | 148,8110725 | 172 | 0,898837363 |
| Cigarettes per Day | eGFR decline (CKD population) [SH01] | Inverse variance weighted | 15,04388109 | 21 | 0,820743555 |
| Chronotype | eGFR decline (CKD population) [SH01] | Inverse variance weighted | 59,78888481 | 59 | 0,446837841 |
| smoking initiation | eGFR decline (CKD population) [SH01] | Inverse variance weighted | 75,19313041 | 84 | 0,743073786 |
| Systolic Blood Pressure (SBP) | eGFR decline (CKD population) [SH01] | Inverse variance weighted | 196,7252325 | 250 | 0,994553827 |
| Diastolic Blood Pressure (DBP) | eGFR decline (CKD population) [SH01] | Inverse variance weighted | 224,9662355 | 250 | 0,870589719 |
| Pulse Pressure (PP) | eGFR decline (CKD population) [SH01] | Inverse variance weighted | 251,4597652 | 301 | 0,982756882 |
| Hypertension | eGFR decline (CKD population) [SH01] | Inverse variance weighted | 134,5777757 | 186 | 0,998281399 |
| Urinary sodium-potassium ratio | eGFR decline (CKD population) [SH01] | Inverse variance weighted | 27,51563247 | 23 | 0,234653778 |
| Insomnia | eGFR decline (CKD population) [SH01] | Inverse variance weighted | 13,09239026 | 12 | 0,362361297 |
| LDL cholesterol | eGFR decline (CKD population) [SH01] | Inverse variance weighted | 224,0667177 | 216 | 0,33893742 |
| Low TSH | eGFR decline (CKD population) [SH01] | Inverse variance weighted | 27,04239311 | 25 | 0,353769968 |
| Thyrotropin-releasing hormone | eGFR decline (CKD population) [SH01] | Inverse variance weighted | 0,003974654 | 1 | 0,949730786 |
| Compound smoking index | eGFR decline (CKD population) [SH01] | Inverse variance weighted | 106,3722615 | 120 | 0,808366124 |
| Type 2 diabetes | eGFR decline (CKD population) [SH01] | Inverse variance weighted | 433,2871072 | 440 | 0,581193723 |
| Total cholesterol | eGFR decline (CKD population) [SH01] | Inverse variance weighted | 234,7968425 | 263 | 0,893856946 |
| Triglyceride | eGFR decline (CKD population) [SH01] | Inverse variance weighted | 206,4010641 | 218 | 0,70318367 |
| Thyroid Stimulating Hormone | eGFR decline (CKD population) [SH01] | Inverse variance weighted | 164,176953 | 158 | 0,351806884 |
| Sleep duration | eGFR decline (CKD population) [SH01] | Inverse variance weighted | 51,11924736 | 65 | 0,895653769 |
| Alcohol intake frequency. | eGFR decline (CKD population) [SH01] | Inverse variance weighted | 86,99633306 | 93 | 0,655746233 |
| Lipoprotein A | eGFR decline (CKD population) [SH01] | Inverse variance weighted | 15,33556967 | 10 | 0,120298106 |
| Waist-to-hip ratio adjusted BMI | eGFR decline (CKD population) [SH01] | Inverse variance weighted | 291,2241561 | 295 | 0,551177629 |
| Zinc | eGFR decline (CKD population) [SH01] | Inverse variance weighted | 3,716912174 | 2 | 0,155913161 |
| Body mass index | eGFR decline (CKD population) [SH02] | Inverse variance weighted | 478,3387302 | 505 | 0,797600678 |
| Heel bone mineral density | eGFR decline (CKD population) [SH02] | Inverse variance weighted | 431,1010248 | 482 | 0,953445349 |
| Two-hour glucose | eGFR decline (CKD population) [SH02] | Inverse variance weighted | 16,4502318 | 13 | 0,225669529 |
| Fasting glucose | eGFR decline (CKD population) [SH02] | Inverse variance weighted | 70,40296252 | 67 | 0,364472421 |
| Fasting insulin | eGFR decline (CKD population) [SH02] | Inverse variance weighted | 49,23593447 | 36 | 0,069704334 |
| circulating leptin levels | eGFR decline (CKD population) [SH02] | Inverse variance weighted | 2,303977332 | 2 | 0,31600771 |
| Growth differentiation factor-15 levels | eGFR decline (CKD population) [SH02] | Inverse variance weighted | 0,83790161 | 2 | 0,657736552 |
| Fibroblast growth factor 23 levels | eGFR decline (CKD population) [SH02] | Inverse variance weighted | 6,196648031 | 7 | 0,516984802 |
| Kidney injury molecule 1 levels | eGFR decline (CKD population) [SH02] | Inverse variance weighted | 9,357118278 | 8 | 0,313071088 |
| Coronary artery disease (Firth correction) | eGFR decline (CKD population) [SH02] | Inverse variance weighted | 22,99285312 | 36 | 0,954359064 |
| Body fat percentage | eGFR decline (CKD population) [SH02] | Inverse variance weighted | 342,7757333 | 315 | 0,135222686 |
| Sodium in urine | eGFR decline (CKD population) [SH02] | Inverse variance weighted | 32,65995364 | 40 | 0,78848705 |
| Glycated haemoglobin HbA1c levels | eGFR decline (CKD population) [SH02] | Inverse variance weighted | 282,0997575 | 301 | 0,776293964 |
| Phosphate levels | eGFR decline (CKD population) [SH02] | Inverse variance weighted | 137,2127506 | 159 | 0,893372878 |
| Bone mineral density | eGFR decline (CKD population) [SH02] | Inverse variance weighted | 383,6774735 | 414 | 0,854824239 |
| Type 1 diabetes | eGFR decline (CKD population) [SH02] | Inverse variance weighted | 61,30956718 | 75 | 0,872606969 |
| Liver iron content | eGFR decline (CKD population) [SH02] | Inverse variance weighted | 3,746974768 | 4 | 0,441330622 |
| Chronic heart failure | eGFR decline (CKD population) [SH02] | Inverse variance weighted | 1,826864228 | 4 | 0,767563884 |
| Hyperthyroidism | eGFR decline (CKD population) [SH02] | Inverse variance weighted | 14,64298667 | 7 | 0,040858033 |
| Hypothyroidism | eGFR decline (CKD population) [SH02] | Inverse variance weighted | 78,67823871 | 63 | 0,087943577 |
| Myocardial infarction | eGFR decline (CKD population) [SH02] | Inverse variance weighted | 41,10928869 | 69 | 0,996941612 |
| Serum uric acid levels | eGFR decline (CKD population) [SH02] | Inverse variance weighted | 221,7275665 | 236 | 0,738856294 |
| Vitamin K-dependent protein S measurement | eGFR decline (CKD population) [SH02] | Inverse variance weighted | 0,193721624 | 3 | 0,978596422 |
| Vitamin K-dependent protein C measurement | eGFR decline (CKD population) [SH02] | Inverse variance weighted | 4,83966654 | 2 | 0,088936445 |
| Serum alkaline phosphatase levels | eGFR decline (CKD population) [SH02] | Inverse variance weighted | 332,2344617 | 334 | 0,51699868 |
| Vitamin D levels | eGFR decline (CKD population) [SH02] | Inverse variance weighted | 76,61708246 | 80 | 0,586435431 |
| Hemoglobin levels | eGFR decline (CKD population) [SH02] | Inverse variance weighted | 327,3140672 | 324 | 0,438048677 |
| Blood glucose levels | eGFR decline (CKD population) [SH02] | Inverse variance weighted | 117,887564 | 108 | 0,242492022 |
| Insulin-like growth factor 1 levels | eGFR decline (CKD population) [SH02] | Inverse variance weighted | 354,8381296 | 372 | 0,730587149 |
| Calcium levels | eGFR decline (CKD population) [SH02] | Inverse variance weighted | 183,535027 | 201 | 0,806249728 |
| Educational attainment (years of education) | eGFR decline (CKD population) [SH02] | Inverse variance weighted | 181,2590351 | 208 | 0,909688949 |
| Smoking status | eGFR decline (CKD population) [SH02] | Inverse variance weighted | 114,5096254 | 116 | 0,521706229 |
| C-reactive protein levels | eGFR decline (CKD population) [SH02] | Inverse variance weighted | 207,7664745 | 213 | 0,588348293 |

| | | | | | |
|---|---|---|---|---|---|
| Liver fat | eGFR decline (CKD population) [SH02] | Inverse variance weighted | 2,522421417 | 7 | 0,925399267 |
| Cardiovascular disease | eGFR decline (CKD population) [SH02] | Inverse variance weighted | 0,201486047 | 1 | 0,653524016 |
| Serum iron | eGFR decline (CKD population) [SH02] | Inverse variance weighted | 0,080873802 | 1 | 0,776116735 |
| Vitamin B12 deficiency anaemia | eGFR decline (CKD population) [SH02] | Inverse variance weighted | 2,083928178 | 3 | 0,555171039 |
| Obesity | eGFR decline (CKD population) [SH02] | Inverse variance weighted | 6,896360892 | 6 | 0,330538193 |
| Pulmonary heart disease | eGFR decline (CKD population) [SH02] | Inverse variance weighted | 4,849767968 | 4 | 0,303062451 |
| Free triiodothyronine | eGFR decline (CKD population) [SH02] | Inverse variance weighted | 4,008440925 | 5 | 0,548201304 |
| FT3 FT4 ratio | eGFR decline (CKD population) [SH02] | Inverse variance weighted | 9,000555879 | 12 | 0,702882955 |
| Free thyroxine | eGFR decline (CKD population) [SH02] | Inverse variance weighted | 59,74301936 | 61 | 0,521566238 |
| HDL cholesterol | eGFR decline (CKD population) [SH02] | Inverse variance weighted | 250,7453627 | 232 | 0,189811169 |
| High TSH | eGFR decline (CKD population) [SH02] | Inverse variance weighted | 5,319014945 | 5 | 0,378199727 |
| Major depression | eGFR decline (CKD population) [SH02] | Inverse variance weighted | 52,24414038 | 47 | 0,277526483 |
| apolipoprotein A-I | eGFR decline (CKD population) [SH02] | Inverse variance weighted | 241,1776569 | 268 | 0,879115614 |
| apolipoprotein B | eGFR decline (CKD population) [SH02] | Inverse variance weighted | 143,7629973 | 172 | 0,942751274 |
| Cigarettes per Day | eGFR decline (CKD population) [SH02] | Inverse variance weighted | 15,29093577 | 21 | 0,808075475 |
| Chronotype | eGFR decline (CKD population) [SH02] | Inverse variance weighted | 58,73171155 | 59 | 0,485358665 |
| smoking initiation | eGFR decline (CKD population) [SH02] | Inverse variance weighted | 74,81059178 | 84 | 0,7532241 |
| Systolic Blood Pressure (SBP) | eGFR decline (CKD population) [SH02] | Inverse variance weighted | 192,5691627 | 250 | 0,997163344 |
| Diastolic Blood Pressure (DBP) | eGFR decline (CKD population) [SH02] | Inverse variance weighted | 224,4312124 | 250 | 0,875929527 |
| Pulse Pressure (PP) | eGFR decline (CKD population) [SH02] | Inverse variance weighted | 248,3586776 | 301 | 0,988012537 |
| Hypertension | eGFR decline (CKD population) [SH02] | Inverse variance weighted | 132,321441 | 186 | 0,998937012 |
| Urinary sodium-potassium ratio | eGFR decline (CKD population) [SH02] | Inverse variance weighted | 26,82123386 | 23 | 0,263650852 |
| Insomnia | eGFR decline (CKD population) [SH02] | Inverse variance weighted | 12,65485634 | 12 | 0,3946181 |
| LDL cholesterol | eGFR decline (CKD population) [SH02] | Inverse variance weighted | 218,5690455 | 216 | 0,438344797 |
| Low TSH | eGFR decline (CKD population) [SH02] | Inverse variance weighted | 24,63323161 | 25 | 0,483071777 |
| Thyrotropin-releasing hormone | eGFR decline (CKD population) [SH02] | Inverse variance weighted | 0,001539809 | 1 | 0,968698722 |
| Compound smoking index | eGFR decline (CKD population) [SH02] | Inverse variance weighted | 107,3719447 | 120 | 0,788777257 |
| Type 2 diabetes | eGFR decline (CKD population) [SH02] | Inverse variance weighted | 432,7558631 | 440 | 0,588250236 |
| Total cholesterol | eGFR decline (CKD population) [SH02] | Inverse variance weighted | 227,4825048 | 263 | 0,944644056 |
| Triglyceride | eGFR decline (CKD population) [SH02] | Inverse variance weighted | 200,1088928 | 218 | 0,802142918 |
| Thyroid Stimulating Hormone | eGFR decline (CKD population) [SH02] | Inverse variance weighted | 158,7685807 | 158 | 0,46785408 |
| Sleep duration | eGFR decline (CKD population) [SH02] | Inverse variance weighted | 48,4496864 | 65 | 0,937880792 |
| Alcohol intake frequency. | eGFR decline (CKD population) [SH02] | Inverse variance weighted | 86,09456409 | 93 | 0,680900834 |
| Lipoprotein A | eGFR decline (CKD population) [SH02] | Inverse variance weighted | 16,25722153 | 10 | 0,092502094 |
| Waist-to-hip ratio adjusted BMI | eGFR decline (CKD population) [SH02] | Inverse variance weighted | 288,90664 | 295 | 0,589053061 |
| Zinc | eGFR decline (CKD population) [SH02] | Inverse variance weighted | 3,650805633 | 2 | 0,161152716 |
| Body mass index | eGFR decline (CKD population) [SH03] | Inverse variance weighted | 473,9643697 | 505 | 0,835524301 |
| Heel bone mineral density | eGFR decline (CKD population) [SH03] | Inverse variance weighted | 426,4664156 | 482 | 0,967163007 |
| Two-hour glucose | eGFR decline (CKD population) [SH03] | Inverse variance weighted | 16,00957955 | 13 | 0,248612948 |
| Fasting glucose | eGFR decline (CKD population) [SH03] | Inverse variance weighted | 69,43494781 | 67 | 0,395384531 |
| Fasting insulin | eGFR decline (CKD population) [SH03] | Inverse variance weighted | 49,07793701 | 36 | 0,07175407 |
| circulating leptin levels | eGFR decline (CKD population) [SH03] | Inverse variance weighted | 2,330522251 | 2 | 0,311841222 |
| Growth differentiation factor-15 levels | eGFR decline (CKD population) [SH03] | Inverse variance weighted | 0,799059784 | 2 | 0,670635243 |
| Fibroblast growth factor 23 levels | eGFR decline (CKD population) [SH03] | Inverse variance weighted | 5,698159059 | 7 | 0,575402054 |
| Kidney injury molecule 1 levels | eGFR decline (CKD population) [SH03] | Inverse variance weighted | 9,262031108 | 8 | 0,320675948 |
| Coronary artery disease (Firth correction) | eGFR decline (CKD population) [SH03] | Inverse variance weighted | 22,64514353 | 36 | 0,959459736 |
| Body fat percentage | eGFR decline (CKD population) [SH03] | Inverse variance weighted | 339,4300577 | 315 | 0,164623378 |
| Sodium in urine | eGFR decline (CKD population) [SH03] | Inverse variance weighted | 31,90312081 | 40 | 0,815618999 |
| Glycated haemoglobin HbA1c levels | eGFR decline (CKD population) [SH03] | Inverse variance weighted | 279,6064268 | 301 | 0,806797604 |
| Phosphate levels | eGFR decline (CKD population) [SH03] | Inverse variance weighted | 136,615489 | 159 | 0,900022761 |
| Bone mineral density | eGFR decline (CKD population) [SH03] | Inverse variance weighted | 381,0151592 | 414 | 0,875937476 |
| Type 1 diabetes | eGFR decline (CKD population) [SH03] | Inverse variance weighted | 60,5904735 | 75 | 0,88614041 |
| Liver iron content | eGFR decline (CKD population) [SH03] | Inverse variance weighted | 3,714333463 | 4 | 0,446044636 |
| Chronic heart failure | eGFR decline (CKD population) [SH03] | Inverse variance weighted | 1,782339238 | 4 | 0,775711865 |
| Hyperthyroidism | eGFR decline (CKD population) [SH03] | Inverse variance weighted | 14,46631802 | 7 | 0,04348246 |
| Hypothyroidism | eGFR decline (CKD population) [SH03] | Inverse variance weighted | 77,26539024 | 63 | 0,10669529 |
| Myocardial infarction | eGFR decline (CKD population) [SH03] | Inverse variance weighted | 40,86289975 | 69 | 0,997208699 |
| Serum uric acid levels | eGFR decline (CKD population) [SH03] | Inverse variance weighted | 215,0209942 | 236 | 0,832704192 |
| Vitamin K-dependent protein S measurement | eGFR decline (CKD population) [SH03] | Inverse variance weighted | 0,181147712 | 3 | 0,980573743 |
| Vitamin K-dependent protein C measurement | eGFR decline (CKD population) [SH03] | Inverse variance weighted | 4,920154473 | 2 | 0,085428353 |
| Serum alkaline phosphatase levels | eGFR decline (CKD population) [SH03] | Inverse variance weighted | 329,7052004 | 334 | 0,556089864 |
| Vitamin D levels | eGFR decline (CKD population) [SH03] | Inverse variance weighted | 76,0979743 | 80 | 0,602835169 |
| Hemoglobin levels | eGFR decline (CKD population) [SH03] | Inverse variance weighted | 320,7052111 | 324 | 0,541279469 |
| Blood glucose levels | eGFR decline (CKD population) [SH03] | Inverse variance weighted | 116,3654036 | 108 | 0,274178705 |
| Insulin-like growth factor 1 levels | eGFR decline (CKD population) [SH03] | Inverse variance weighted | 351,4876431 | 372 | 0,770879266 |
| Calcium levels | eGFR decline (CKD population) [SH03] | Inverse variance weighted | 181,3808164 | 201 | 0,836151099 |
| Educational attainment (years of education) | eGFR decline (CKD population) [SH03] | Inverse variance weighted | 178,6492089 | 208 | 0,930538989 |
| Smoking status | eGFR decline (CKD population) [SH03] | Inverse variance weighted | 114,1777103 | 116 | 0,530460374 |
| C-reactive protein levels | eGFR decline (CKD population) [SH03] | Inverse variance weighted | 206,1004572 | 213 | 0,620033112 |
| Liver fat | eGFR decline (CKD population) [SH03] | Inverse variance weighted | 2,55602364 | 7 | 0,922819679 |
| Cardiovascular disease | eGFR decline (CKD population) [SH03] | Inverse variance weighted | 0,167960397 | 1 | 0,681930865 |
| Serum iron | eGFR decline (CKD population) [SH03] | Inverse variance weighted | 0,077693646 | 1 | 0,780447587 |
| Vitamin B12 deficiency anaemia | eGFR decline (CKD population) [SH03] | Inverse variance weighted | 2,098149223 | 3 | 0,552287279 |
| Obesity | eGFR decline (CKD population) [SH03] | Inverse variance weighted | 6,819182452 | 6 | 0,337893418 |
| Pulmonary heart disease | eGFR decline (CKD population) [SH03] | Inverse variance weighted | 4,759984085 | 4 | 0,312822705 |
| Free triiodothyronine | eGFR decline (CKD population) [SH03] | Inverse variance weighted | 3,933423029 | 5 | 0,559040754 |

| | | | | | |
|---|---|---|---|---|---|
| FT3 FT4 ratio | eGFR decline (CKD population) [SH03] | Inverse variance weighted | 8,9086504 | 12 | 0,710712482 |
| Free thyroxine | eGFR decline (CKD population) [SH03] | Inverse variance weighted | 57,8079968 | 61 | 0,592321259 |
| HDL cholesterol | eGFR decline (CKD population) [SH03] | Inverse variance weighted | 247,4712374 | 232 | 0,231581164 |
| High TSH | eGFR decline (CKD population) [SH03] | Inverse variance weighted | 5,014461074 | 5 | 0,414117889 |
| Major depression | eGFR decline (CKD population) [SH03] | Inverse variance weighted | 51,9863556 | 47 | 0,285897987 |
| apolipoprotein A-I | eGFR decline (CKD population) [SH03] | Inverse variance weighted | 238,3902805 | 268 | 0,903244254 |
| apolipoprotein B | eGFR decline (CKD population) [SH03] | Inverse variance weighted | 141,1743431 | 172 | 0,958775699 |
| Cigarettes per Day | eGFR decline (CKD population) [SH03] | Inverse variance weighted | 15,27944749 | 21 | 0,808673253 |
| Chronotype | eGFR decline (CKD population) [SH03] | Inverse variance weighted | 58,05189625 | 59 | 0,510470543 |
| smoking initiation | eGFR decline (CKD population) [SH03] | Inverse variance weighted | 74,2711662 | 84 | 0,767221449 |
| Systolic Blood Pressure (SBP) | eGFR decline (CKD population) [SH03] | Inverse variance weighted | 189,5179901 | 250 | 0,9983032 |
| Diastolic Blood Pressure (DBP) | eGFR decline (CKD population) [SH03] | Inverse variance weighted | 222,2483346 | 250 | 0,896189542 |
| Pulse Pressure (PP) | eGFR decline (CKD population) [SH03] | Inverse variance weighted | 245,5410155 | 301 | 0,991545686 |
| Hypertension | eGFR decline (CKD population) [SH03] | Inverse variance weighted | 130,2731811 | 186 | 0,999328002 |
| Urinary sodium-potassium ratio | eGFR decline (CKD population) [SH03] | Inverse variance weighted | 26,46385244 | 23 | 0,279461248 |
| Insomnia | eGFR decline (CKD population) [SH03] | Inverse variance weighted | 12,47569066 | 12 | 0,40827105 |
| LDL cholesterol | eGFR decline (CKD population) [SH03] | Inverse variance weighted | 215,2992608 | 216 | 0,500662265 |
| Low TSH | eGFR decline (CKD population) [SH03] | Inverse variance weighted | 23,48133503 | 25 | 0,549478548 |
| Thyrotropin-releasing hormone | eGFR decline (CKD population) [SH03] | Inverse variance weighted | 0,001047311 | 1 | 0,974183218 |
| Compound smoking index | eGFR decline (CKD population) [SH03] | Inverse variance weighted | 107,1313801 | 120 | 0,79358389 |
| Type 2 diabetes | eGFR decline (CKD population) [SH03] | Inverse variance weighted | 429,976977 | 440 | 0,624741416 |
| Total cholesterol | eGFR decline (CKD population) [SH03] | Inverse variance weighted | 223,8340007 | 263 | 0,961741497 |
| Triglyceride | eGFR decline (CKD population) [SH03] | Inverse variance weighted | 196,8294977 | 218 | 0,845303699 |
| Thyroid Stimulating Hormone | eGFR decline (CKD population) [SH03] | Inverse variance weighted | 155,9537496 | 158 | 0,531108417 |
| Sleep duration | eGFR decline (CKD population) [SH03] | Inverse variance weighted | 47,30629286 | 65 | 0,951583072 |
| Alcohol intake frequency. | eGFR decline (CKD population) [SH03] | Inverse variance weighted | 85,30308206 | 93 | 0,702456888 |
| Lipoprotein A | eGFR decline (CKD population) [SH03] | Inverse variance weighted | 16,44965174 | 10 | 0,087463861 |
| Waist-to-hip ratio adjusted BMI | eGFR decline (CKD population) [SH03] | Inverse variance weighted | 286,5104395 | 295 | 0,62756188 |
| Zinc | eGFR decline (CKD population) [SH03] | Inverse variance weighted | 3,612340119 | 2 | 0,164282124 |
| Body mass index | eGFR decline (CKD population) [SH04] | Inverse variance weighted | 471,0321906 | 505 | 0,858291986 |
| Heel bone mineral density | eGFR decline (CKD population) [SH04] | Inverse variance weighted | 420,4034902 | 482 | 0,979993147 |
| Two-hour glucose | eGFR decline (CKD population) [SH04] | Inverse variance weighted | 14,82014979 | 13 | 0,318714637 |
| Fasting glucose | eGFR decline (CKD population) [SH04] | Inverse variance weighted | 69,77111716 | 67 | 0,384531105 |
| Fasting insulin | eGFR decline (CKD population) [SH04] | Inverse variance weighted | 50,75270596 | 36 | 0,05242149 |
| circulating leptin levels | eGFR decline (CKD population) [SH04] | Inverse variance weighted | 2,537505977 | 2 | 0,28118204 |
| Growth differentiation factor-15 levels | eGFR decline (CKD population) [SH04] | Inverse variance weighted | 0,683534754 | 2 | 0,710513467 |
| Fibroblast growth factor 23 levels | eGFR decline (CKD population) [SH04] | Inverse variance weighted | 4,852474897 | 7 | 0,677960791 |
| Kidney injury molecule 1 levels | eGFR decline (CKD population) [SH04] | Inverse variance weighted | 11,28946402 | 8 | 0,185830965 |
| Coronary artery disease (Firth correction) | eGFR decline (CKD population) [SH04] | Inverse variance weighted | 22,34097366 | 36 | 0,963571992 |
| Body fat percentage | eGFR decline (CKD population) [SH04] | Inverse variance weighted | 336,7934373 | 315 | 0,190669058 |
| Sodium in urine | eGFR decline (CKD population) [SH04] | Inverse variance weighted | 30,58261529 | 40 | 0,858352162 |
| Glycated haemoglobin HbA1c levels | eGFR decline (CKD population) [SH04] | Inverse variance weighted | 279,0515132 | 301 | 0,813247389 |
| Phosphate levels | eGFR decline (CKD population) [SH04] | Inverse variance weighted | 141,0885932 | 159 | 0,843049272 |
| Bone mineral density | eGFR decline (CKD population) [SH04] | Inverse variance weighted | 382,4538032 | 414 | 0,864799049 |
| Type 1 diabetes | eGFR decline (CKD population) [SH04] | Inverse variance weighted | 59,3485794 | 75 | 0,907255229 |
| Liver iron content | eGFR decline (CKD population) [SH04] | Inverse variance weighted | 3,617778411 | 4 | 0,460197239 |
| Chronic heart failure | eGFR decline (CKD population) [SH04] | Inverse variance weighted | 1,656675525 | 4 | 0,79857169 |
| Hyperthyroidism | eGFR decline (CKD population) [SH04] | Inverse variance weighted | 13,9782121 | 7 | 0,051570232 |
| Hypothyroidism | eGFR decline (CKD population) [SH04] | Inverse variance weighted | 74,23872967 | 63 | 0,157253926 |
| Myocardial infarction | eGFR decline (CKD population) [SH04] | Inverse variance weighted | 42,41870949 | 69 | 0,995126603 |
| Serum uric acid levels | eGFR decline (CKD population) [SH04] | Inverse variance weighted | 209,5509285 | 236 | 0,891609777 |
| Vitamin K-dependent protein S measurement | eGFR decline (CKD population) [SH04] | Inverse variance weighted | 0,134319603 | 3 | 0,987422483 |
| Vitamin K-dependent protein C measurement | eGFR decline (CKD population) [SH04] | Inverse variance weighted | 5,458833139 | 2 | 0,065257352 |
| Serum alkaline phosphatase levels | eGFR decline (CKD population) [SH04] | Inverse variance weighted | 332,4174269 | 334 | 0,514167674 |
| Vitamin D levels | eGFR decline (CKD population) [SH04] | Inverse variance weighted | 79,12965844 | 80 | 0,506492051 |
| Hemoglobin levels | eGFR decline (CKD population) [SH04] | Inverse variance weighted | 316,3090176 | 324 | 0,609612578 |
| Blood glucose levels | eGFR decline (CKD population) [SH04] | Inverse variance weighted | 116,2370589 | 108 | 0,276954009 |
| Insulin-like growth factor 1 levels | eGFR decline (CKD population) [SH04] | Inverse variance weighted | 350,6178219 | 372 | 0,780797449 |
| Calcium levels | eGFR decline (CKD population) [SH04] | Inverse variance weighted | 183,6407598 | 201 | 0,804707632 |
| Educational attainment (years of education) | eGFR decline (CKD population) [SH04] | Inverse variance weighted | 174,6952688 | 208 | 0,955063865 |
| Smoking status | eGFR decline (CKD population) [SH04] | Inverse variance weighted | 115,1776855 | 116 | 0,504110229 |
| C-reactive protein levels | eGFR decline (CKD population) [SH04] | Inverse variance weighted | 205,631885 | 213 | 0,628841409 |
| Liver fat | eGFR decline (CKD population) [SH04] | Inverse variance weighted | 3,175126579 | 7 | 0,868342715 |
| Cardiovascular disease | eGFR decline (CKD population) [SH04] | Inverse variance weighted | 0,051389551 | 1 | 0,820662753 |
| Serum iron | eGFR decline (CKD population) [SH04] | Inverse variance weighted | 0,063394629 | 1 | 0,801208876 |
| Vitamin B12 deficiency anaemia | eGFR decline (CKD population) [SH04] | Inverse variance weighted | 2,210099256 | 3 | 0,529961873 |
| Obesity | eGFR decline (CKD population) [SH04] | Inverse variance weighted | 6,64453752 | 6 | 0,354973883 |
| Pulmonary heart disease | eGFR decline (CKD population) [SH04] | Inverse variance weighted | 4,417891573 | 4 | 0,352394737 |
| Free triiodothyronine | eGFR decline (CKD population) [SH04] | Inverse variance weighted | 3,685357335 | 5 | 0,595544511 |
| FT3 FT4 ratio | eGFR decline (CKD population) [SH04] | Inverse variance weighted | 8,629999747 | 12 | 0,734166136 |
| Free thyroxine | eGFR decline (CKD population) [SH04] | Inverse variance weighted | 53,24062055 | 61 | 0,749731975 |
| HDL cholesterol | eGFR decline (CKD population) [SH04] | Inverse variance weighted | 247,398241 | 232 | 0,232571741 |
| High TSH | eGFR decline (CKD population) [SH04] | Inverse variance weighted | 4,175724876 | 5 | 0,524403392 |
| Major depression | eGFR decline (CKD population) [SH04] | Inverse variance weighted | 52,27390505 | 47 | 0,276569394 |
| apolipoprotein A-I | eGFR decline (CKD population) [SH04] | Inverse variance weighted | 239,1966298 | 268 | 0,896640873 |
| apolipoprotein B | eGFR decline (CKD population) [SH04] | Inverse variance weighted | 137,4818377 | 172 | 0,975349322 |

| | | | | | |
|---|---|---|---|---|---|
| Cigarettes per Day | eGFR decline (CKD population) [SH04] | Inverse variance weighted | 15,74874741 | 21 | 0,783597708 |
| Chronotype | eGFR decline (CKD population) [SH04] | Inverse variance weighted | 57,12405441 | 59 | 0,544974929 |
| smoking initiation | eGFR decline (CKD population) [SH04] | Inverse variance weighted | 74,15721631 | 84 | 0,77012937 |
| Systolic Blood Pressure (SBP) | eGFR decline (CKD population) [SH04] | Inverse variance weighted | 188,8089973 | 250 | 0,998500766 |
| Diastolic Blood Pressure (DBP) | eGFR decline (CKD population) [SH04] | Inverse variance weighted | 226,4253927 | 250 | 0,855272559 |
| Pulse Pressure (PP) | eGFR decline (CKD population) [SH04] | Inverse variance weighted | 246,3860513 | 301 | 0,990594285 |
| Hypertension | eGFR decline (CKD population) [SH04] | Inverse variance weighted | 131,9099712 | 186 | 0,999028874 |
| Urinary sodium-potassium ratio | eGFR decline (CKD population) [SH04] | Inverse variance weighted | 26,06116876 | 23 | 0,297987039 |
| Insomnia | eGFR decline (CKD population) [SH04] | Inverse variance weighted | 11,7854455 | 12 | 0,463061122 |
| LDL cholesterol | eGFR decline (CKD population) [SH04] | Inverse variance weighted | 210,7418503 | 216 | 0,588200211 |
| Low TSH | eGFR decline (CKD population) [SH04] | Inverse variance weighted | 22,18764648 | 25 | 0,624886541 |
| Thyrotropin-releasing hormone | eGFR decline (CKD population) [SH04] | Inverse variance weighted | 4,58169E-06 | 1 | 0,998292139 |
| Compound smoking index | eGFR decline (CKD population) [SH04] | Inverse variance weighted | 109,0577161 | 120 | 0,75352477 |
| Type 2 diabetes | eGFR decline (CKD population) [SH04] | Inverse variance weighted | 432,610355 | 440 | 0,59017916 |
| Total cholesterol | eGFR decline (CKD population) [SH04] | Inverse variance weighted | 217,6346777 | 263 | 0,981018897 |
| Triglyceride | eGFR decline (CKD population) [SH04] | Inverse variance weighted | 191,6455186 | 218 | 0,90054485 |
| Thyroid Stimulating Hormone | eGFR decline (CKD population) [SH04] | Inverse variance weighted | 149,6611269 | 158 | 0,670007154 |
| Sleep duration | eGFR decline (CKD population) [SH04] | Inverse variance weighted | 44,522638 | 65 | 0,975526724 |
| Alcohol intake frequency. | eGFR decline (CKD population) [SH04] | Inverse variance weighted | 85,40429051 | 93 | 0,69973038 |
| Lipoprotein A | eGFR decline (CKD population) [SH04] | Inverse variance weighted | 17,7365753 | 10 | 0,059573182 |
| Waist-to-hip ratio adjusted BMI | eGFR decline (CKD population) [SH04] | Inverse variance weighted | 287,2191644 | 295 | 0,616263117 |
| Zinc | eGFR decline (CKD population) [SH04] | Inverse variance weighted | 3,500730465 | 2 | 0,173710487 |

| exposure | outcome | egger_intercept | egger_se | egger_pval |
|---|---|---|---|---|
| Body mass index | Incident CKD | 0,00081799 | 0,001421045 | 0,565125085 |
| Heel bone mineral density | Incident CKD | 0,002382089 | 0,00117329 | 0,042862775 |
| Two-hour glucose | Incident CKD | -0,010911368 | 0,013306981 | 0,428218343 |
| Fasting glucose | Incident CKD | -0,001316759 | 0,003936721 | 0,739061289 |
| Fasting insulin | Incident CKD | 0,010650553 | 0,008488405 | 0,217895405 |
| circulating leptin levels | Incident CKD | -0,008934536 | 0,092823766 | 0,938911833 |
| Growth differentiation factor-15 levels | Incident CKD | 0,0140143 | 0,023290114 | 0,655150281 |
| Fibroblast growth factor 23 levels | Incident CKD | -0,073236164 | 0,046388702 | 0,165471518 |
| Kidney injury molecule 1 levels | Incident CKD | 0,044550251 | 0,037518392 | 0,273786583 |
| Coronary artery disease (Firth correction) | Incident CKD | -0,003040796 | 0,006251738 | 0,629557839 |
| Body fat percentage | Incident CKD | 0,002584749 | 0,002405551 | 0,283421379 |
| Sodium in urine | Incident CKD | -0,010566017 | 0,010450902 | 0,318241037 |
| Glycated haemoglobin HbA1c levels | Incident CKD | 0,000523734 | 0,001562612 | 0,737723946 |
| Phosphate levels | Incident CKD | -0,00100714 | 0,002770551 | 0,716699287 |
| Bone mineral density | Incident CKD | 0,000723809 | 0,001328816 | 0,586246065 |
| Type 1 diabetes | Incident CKD | -0,001276621 | 0,002515652 | 0,613313122 |
| Liver iron content | Incident CKD | 0,012630951 | 0,012772403 | 0,395607012 |
| Chronic heart failure | Incident CKD | -0,003501944 | 0,022573898 | 0,884230359 |
| Hyperthyroidism | Incident CKD | -0,009936568 | 0,020501843 | 0,645097586 |
| Hypothyroidism | Incident CKD | -0,001176941 | 0,003827747 | 0,759449028 |
| Myocardial infarction | Incident CKD | -0,002165327 | 0,005124897 | 0,67389385 |
| Serum uric acid levels | Incident CKD | 0,009191665 | 0,002601346 | 0,000487832 |
| Vitamin K-dependent protein S measurement | Incident CKD | 0,00845977 | 0,011136474 | 0,526795917 |
| Vitamin K-dependent protein C measurement | Incident CKD | 0,000393536 | 0,017505065 | 0,984105336 |
| Serum alkaline phosphatase levels | Incident CKD | 0,001867047 | 0,001395734 | 0,181889466 |
| Vitamin D levels | Incident CKD | 0,001954329 | 0,003260988 | 0,550621303 |
| Hemoglobin levels | Incident CKD | 0,000777865 | 0,002128406 | 0,714997519 |
| Blood glucose levels | Incident CKD | -0,001339442 | 0,002637735 | 0,612629031 |
| Insulin-like growth factor 1 levels | Incident CKD | -0,000727605 | 0,001461046 | 0,61876992 |
| Calcium levels | Incident CKD | 0,002713006 | 0,002415969 | 0,262729901 |
| Educational attainment (years of education) | Incident CKD | 0,001851177 | 0,003906252 | 0,636067477 |
| Smoking status | Incident CKD | -0,001611304 | 0,004313788 | 0,709446088 |
| C-reactive protein levels | Incident CKD | 0,001642781 | 0,001589117 | 0,30234889 |
| Liver fat | Incident CKD | -0,006880762 | 0,015917439 | 0,680643226 |
| Vitamin B12 deficiency anaemia | Incident CKD | -0,004147518 | 0,018136688 | 0,840371525 |
| Obesity | Incident CKD | -0,029467026 | 0,013681455 | 0,074714081 |
| Pulmonary heart disease | Incident CKD | -0,032354701 | 0,016349136 | 0,118939614 |
| Free triiodothyronine | Incident CKD | 0,013118419 | 0,019052604 | 0,528969145 |
| FT3 FT4 ratio | Incident CKD | 0,004818712 | 0,005700557 | 0,415948389 |
| Free thyroxine | Incident CKD | -0,006882051 | 0,004202334 | 0,106638675 |
| HDL cholesterol | Incident CKD | -0,003959391 | 0,00176059 | 0,025400678 |
| High TSH | Incident CKD | 0,033235947 | 0,024558871 | 0,247379583 |
| Major depression | Incident CKD | 0,018409532 | 0,009464541 | 0,057889546 |
| apolipoprotein A-I | Incident CKD | -0,002116317 | 0,001628007 | 0,194678091 |
| apolipoprotein B | Incident CKD | -0,001264316 | 0,002076544 | 0,543383272 |
| Cigarettes per Day | Incident CKD | 0,000296288 | 0,00430902 | 0,945863526 |
| Chronotype | Incident CKD | 0,001932895 | 0,00517748 | 0,71022066 |
| smoking initiation | Incident CKD | 0,0047903 | 0,006287405 | 0,448286338 |
| Systolic Blood Pressure (SBP) | Incident CKD | 0,002900902 | 0,003210283 | 0,367045541 |
| Diastolic Blood Pressure (DBP) | Incident CKD | 0,00277637 | 0,003918722 | 0,479283972 |
| Pulse Pressure (PP) | Incident CKD | -0,003149197 | 0,002443873 | 0,19850107 |
| Hypertension | Incident CKD | -0,000329128 | 0,005029529 | 0,947893172 |
| Urinary sodium-potassium ratio | Incident CKD | -0,014192224 | 0,016450504 | 0,397597346 |
| Insomnia | Incident CKD | -0,005380827 | 0,009628444 | 0,587462468 |
| LDL cholesterol | Incident CKD | -0,001393914 | 0,001678473 | 0,407152089 |
| Low TSH | Incident CKD | 0,007638708 | 0,012834264 | 0,556867903 |
| Compound smoking index | Incident CKD | -0,00249349 | 0,004073185 | 0,541591789 |
| Type 2 diabetes | Incident CKD | 0,000391403 | 0,001270349 | 0,758144242 |
| Total cholesterol | Incident CKD | -0,002073661 | 0,001536427 | 0,178237591 |
| Triglyceride | Incident CKD | 0,004406748 | 0,001642039 | 0,007824785 |
| Thyroid Stimulating Hormone | Incident CKD | -0,000356222 | 0,001950194 | 0,855293445 |
| Sleep duration | Incident CKD | 0,008799062 | 0,0067838 | 0,199263767 |
| Alcohol intake frequency. | Incident CKD | 0,002393879 | 0,003494945 | 0,495076191 |
| Lipoprotein A | Incident CKD | 0,012006926 | 0,009654239 | 0,230489585 |
| Waist-to-hip ratio adjusted BMI | Incident CKD | -0,000968684 | 0,002011112 | 0,630400581 |
| Zinc | Incident CKD | 0,010586138 | 0,022445419 | 0,719439998 |

| | | | | |
|---|---|---|---|---|
| Body mass index | eGFR decline (CKD population) | 0,00021839 | 0,003204664 | 0,945695355 |
| Heel bone mineral density | eGFR decline (CKD population) | 0,003110316 | 0,002623895 | 0,236451968 |
| Two-hour glucose | eGFR decline (CKD population) | 0,018501777 | 0,022088719 | 0,41861905 |
| Fasting glucose | eGFR decline (CKD population) | -0,010389619 | 0,007216262 | 0,154664464 |
| Fasting insulin | eGFR decline (CKD population) | -0,001831966 | 0,018651096 | 0,922300095 |
| circulating leptin levels | eGFR decline (CKD population) | -0,184293974 | 0,115877782 | 0,357336497 |
| Growth differentiation factor-15 levels | eGFR decline (CKD population) | 0,019923563 | 0,02540299 | 0,576587925 |
| Fibroblast growth factor 23 levels | eGFR decline (CKD population) | -0,036382959 | 0,037199176 | 0,365817619 |
| Kidney injury molecule 1 levels | eGFR decline (CKD population) | 0,022176007 | 0,0336015 | 0,530380795 |
| Coronary artery disease (Firth correction) | eGFR decline (CKD population) | 0,004135795 | 0,011295492 | 0,716399453 |
| Body fat percentage | eGFR decline (CKD population) | -0,00287748 | 0,005399313 | 0,594449977 |
| Sodium in urine | eGFR decline (CKD population) | -0,027075348 | 0,017744173 | 0,135111305 |
| Glycated haemoglobin HbA1c levels | eGFR decline (CKD population) | -0,005769686 | 0,003256875 | 0,077479262 |
| Phosphate levels | eGFR decline (CKD population) | -0,006952167 | 0,004369027 | 0,113541993 |
| Bone mineral density | eGFR decline (CKD population) | 0,000883287 | 0,002924592 | 0,762787175 |
| Type 1 diabetes | eGFR decline (CKD population) | 0,000615638 | 0,005659616 | 0,913673522 |
| Liver iron content | eGFR decline (CKD population) | -0,004236083 | 0,026276433 | 0,879741043 |
| Chronic heart failure | eGFR decline (CKD population) | -0,072901652 | 0,095183629 | 0,499478764 |
| Hyperthyroidism | eGFR decline (CKD population) | 0,056045685 | 0,042315676 | 0,22193328 |
| Hypothyroidism | eGFR decline (CKD population) | 0,008747177 | 0,008646833 | 0,315658767 |
| Myocardial infarction | eGFR decline (CKD population) | 0,01274046 | 0,008521118 | 0,139498545 |
| Serum uric acid levels | eGFR decline (CKD population) | 0,007416189 | 0,003612827 | 0,041200913 |
| Vitamin K-dependent protein S measurement | eGFR decline (CKD population) | -0,00377531 | 0,029230219 | 0,909050165 |
| Vitamin K-dependent protein C measurement | eGFR decline (CKD population) | -0,01867845 | 0,093713642 | 0,874754003 |
| Serum alkaline phosphatase levels | eGFR decline (CKD population) | 0,002892462 | 0,00288353 | 0,316541981 |
| Vitamin D levels | eGFR decline (CKD population) | -0,000277755 | 0,005976223 | 0,963047595 |
| Hemoglobin levels | eGFR decline (CKD population) | -0,002335458 | 0,003222579 | 0,469150395 |
| Blood glucose levels | eGFR decline (CKD population) | -0,006513157 | 0,005436781 | 0,23357155 |
| Insulin-like growth factor 1 levels | eGFR decline (CKD population) | 0,005452893 | 0,002887743 | 0,05976509 |
| Calcium levels | eGFR decline (CKD population) | -0,006038016 | 0,003961711 | 0,129041892 |
| Educational attainment (years of education) | eGFR decline (CKD population) | -0,004124395 | 0,008688629 | 0,635509166 |
| Smoking status | eGFR decline (CKD population) | 0,002906613 | 0,011109228 | 0,794067397 |
| C-reactive protein levels | eGFR decline (CKD population) | -0,004944907 | 0,003417591 | 0,149395402 |
| Liver fat | eGFR decline (CKD population) | -0,019997044 | 0,018604507 | 0,323753737 |
| Vitamin B12 deficiency anaemia | eGFR decline (CKD population) | 0,0128397 | 0,052178383 | 0,828575668 |
| Obesity | eGFR decline (CKD population) | -0,009023383 | 0,04360009 | 0,844207075 |
| Pulmonary heart disease | eGFR decline (CKD population) | 0,053460209 | 0,039235036 | 0,266290388 |
| Free triiodothyronine | eGFR decline (CKD population) | 0,025984202 | 0,050708479 | 0,635352303 |
| FT3 FT4 ratio | eGFR decline (CKD population) | -0,015383027 | 0,015117434 | 0,33074204 |
| Free thyroxine | eGFR decline (CKD population) | -0,0016636 | 0,007557735 | 0,82652586 |
| HDL cholesterol | eGFR decline (CKD population) | -0,006642276 | 0,003294968 | 0,044965069 |
| High TSH | eGFR decline (CKD population) | -0,012204765 | 0,045587933 | 0,802153442 |
| Major depression | eGFR decline (CKD population) | -0,001167201 | 0,024349967 | 0,961976032 |
| apolipoprotein A-I | eGFR decline (CKD population) | 0,001455919 | 0,003065171 | 0,635183979 |
| apolipoprotein B | eGFR decline (CKD population) | -7,09305E-05 | 0,00380355 | 0,985143004 |
| Cigarettes per Day | eGFR decline (CKD population) | 0,006960787 | 0,010914628 | 0,530871865 |
| Chronotype | eGFR decline (CKD population) | 0,017629696 | 0,010980148 | 0,113889045 |
| smoking initiation | eGFR decline (CKD population) | -0,033527209 | 0,015254641 | 0,030746102 |
| Systolic Blood Pressure (SBP) | eGFR decline (CKD population) | 0,005731737 | 0,00562618 | 0,309293142 |
| Diastolic Blood Pressure (DBP) | eGFR decline (CKD population) | 0,001929415 | 0,006144885 | 0,753787545 |
| Pulse Pressure (PP) | eGFR decline (CKD population) | -0,000225823 | 0,00446526 | 0,959699028 |
| Hypertension | eGFR decline (CKD population) | 0,007272535 | 0,006587948 | 0,271026082 |
| Urinary sodium-potassium ratio | eGFR decline (CKD population) | -0,018460105 | 0,028059655 | 0,517435635 |
| Insomnia | eGFR decline (CKD population) | -0,000896763 | 0,026131388 | 0,973238796 |
| LDL cholesterol | eGFR decline (CKD population) | -0,001020214 | 0,003331461 | 0,759719804 |
| Low TSH | eGFR decline (CKD population) | 0,012106792 | 0,01672241 | 0,476072814 |
| Compound smoking index | eGFR decline (CKD population) | -0,007102568 | 0,010254123 | 0,489875709 |
| Type 2 diabetes | eGFR decline (CKD population) | -0,001531389 | 0,002924806 | 0,600830275 |
| Total cholesterol | eGFR decline (CKD population) | 0,001001177 | 0,002972042 | 0,736488307 |
| Triglyceride | eGFR decline (CKD population) | -0,000797106 | 0,003138839 | 0,799774 |
| Thyroid Stimulating Hormone | eGFR decline (CKD population) | -0,006316519 | 0,004235408 | 0,13787347 |
| Sleep duration | eGFR decline (CKD population) | 0,003166795 | 0,013201413 | 0,811187622 |
| Alcohol intake frequency. | eGFR decline (CKD population) | 0,001572605 | 0,006648484 | 0,813542749 |
| Lipoprotein A | eGFR decline (CKD population) | 0,014937878 | 0,038273414 | 0,70539598 |
| Waist-to-hip ratio adjusted BMI | eGFR decline (CKD population) | 0,003736369 | 0,003950512 | 0,3450306 |
| Zinc | eGFR decline (CKD population) | 0,057049564 | 0,056390654 | 0,496302276 |
| Body mass index | eGFR decline (General population) | 0,000582766 | 0,000571277 | 0,308166822 |

| | | | | |
|---|---|---|---|---|
| Heel bone mineral density | eGFR decline (General population) | -0,000142401 | 0,000467729 | 0,760910905 |
| Two-hour glucose | eGFR decline (General population) | -0,001923837 | 0,003758055 | 0,617994359 |
| Fasting glucose | eGFR decline (General population) | 0,001147581 | 0,001286927 | 0,375733261 |
| Fasting insulin | eGFR decline (General population) | 0,000907579 | 0,003105861 | 0,771799652 |
| circulating leptin levels | eGFR decline (General population) | -0,011049664 | 0,020286321 | 0,68248348 |
| Growth differentiation factor-15 levels | eGFR decline (General population) | 0,006664909 | 0,007894736 | 0,553646758 |
| Fibroblast growth factor 23 levels | eGFR decline (General population) | -0,000732211 | 0,006684474 | 0,91634674 |
| Kidney injury molecule 1 levels | eGFR decline (General population) | 0,011879167 | 0,01153751 | 0,333312488 |
| Coronary artery disease (Firth correction) | eGFR decline (General population) | -0,000194887 | 0,0020261 | 0,92387672 |
| Body fat percentage | eGFR decline (General population) | -0,00027011 | 0,000939796 | 0,773978886 |
| Sodium in urine | eGFR decline (General population) | 0,001305456 | 0,003939194 | 0,742115329 |
| Glycated haemoglobin HbA1c levels | eGFR decline (General population) | 0,000436946 | 0,000577737 | 0,450016872 |
| Phosphate levels | eGFR decline (General population) | 0,000400603 | 0,000784941 | 0,610480544 |
| Bone mineral density | eGFR decline (General population) | -9,2586E-05 | 0,00049257 | 0,850991489 |
| Type 1 diabetes | eGFR decline (General population) | -0,000239194 | 0,001014989 | 0,814329463 |
| Liver iron content | eGFR decline (General population) | 0,005049229 | 0,004382608 | 0,31344922 |
| Chronic heart failure | eGFR decline (General population) | -0,012217904 | 0,009617049 | 0,272793458 |
| Hyperthyroidism | eGFR decline (General population) | 0,006404089 | 0,004718303 | 0,207747493 |
| Hypothyroidism | eGFR decline (General population) | 0,000470689 | 0,001486808 | 0,752564071 |
| Myocardial infarction | eGFR decline (General population) | 0,000197232 | 0,001692217 | 0,90752607 |
| Serum uric acid levels | eGFR decline (General population) | 0,001257676 | 0,000699728 | 0,073444906 |
| Vitamin K-dependent protein S measurement | eGFR decline (General population) | -0,013969114 | 0,005834015 | 0,138967317 |
| Vitamin K-dependent protein C measurement | eGFR decline (General population) | -0,019118903 | 0,008953283 | 0,166261294 |
| Serum alkaline phosphatase levels | eGFR decline (General population) | 0,000232885 | 0,000487154 | 0,632912627 |
| Vitamin D levels | eGFR decline (General population) | 0,000579682 | 0,001003784 | 0,565186123 |
| Hemoglobin levels | eGFR decline (General population) | -0,000657263 | 0,000618602 | 0,288789153 |
| Blood glucose levels | eGFR decline (General population) | -0,000281284 | 0,000912618 | 0,758495496 |
| Insulin-like growth factor 1 levels | eGFR decline (General population) | -0,000138033 | 0,000540114 | 0,798425549 |
| Calcium levels | eGFR decline (General population) | -0,000641925 | 0,000749267 | 0,392543958 |
| Educational attainment (years of education) | eGFR decline (General population) | -0,000150733 | 0,00150085 | 0,920097607 |
| Smoking status | eGFR decline (General population) | -0,001807611 | 0,001990293 | 0,36566485 |
| C-reactive protein levels | eGFR decline (General population) | 0,000309092 | 0,00060962 | 0,612625752 |
| Liver fat | eGFR decline (General population) | 0,000487788 | 0,003538077 | 0,894226686 |
| Vitamin B12 deficiency anaemia | eGFR decline (General population) | 0,001669768 | 0,008702396 | 0,865555979 |
| Obesity | eGFR decline (General population) | -0,013978746 | 0,00931271 | 0,184019863 |
| Pulmonary heart disease | eGFR decline (General population) | 0,004583207 | 0,007907242 | 0,593248727 |
| Free triiodothyronine | eGFR decline (General population) | -0,006771982 | 0,009055771 | 0,496143432 |
| FT3 FT4 ratio | eGFR decline (General population) | 0,000245211 | 0,004416873 | 0,956722298 |
| Free thyroxine | eGFR decline (General population) | -0,000231353 | 0,001542628 | 0,881280551 |
| HDL cholesterol | eGFR decline (General population) | -0,000319797 | 0,000575122 | 0,578675387 |
| High TSH | eGFR decline (General population) | -0,007525571 | 0,010255429 | 0,503754439 |
| Major depression | eGFR decline (General population) | 0,000919733 | 0,003814574 | 0,810541061 |
| apolipoprotein A-I | eGFR decline (General population) | -0,000729384 | 0,000555288 | 0,190058987 |
| apolipoprotein B | eGFR decline (General population) | 0,000379626 | 0,000638037 | 0,552568483 |
| Cigarettes per Day | eGFR decline (General population) | -0,002349848 | 0,002235881 | 0,305805403 |
| Chronotype | eGFR decline (General population) | 0,001240445 | 0,002220281 | 0,578453714 |
| smoking initiation | eGFR decline (General population) | -0,001998048 | 0,002754651 | 0,470286036 |
| Systolic Blood Pressure (SBP) | eGFR decline (General population) | 0,001720254 | 0,000995794 | 0,085243838 |
| Diastolic Blood Pressure (DBP) | eGFR decline (General population) | 0,001619363 | 0,001326455 | 0,223230922 |
| Pulse Pressure (PP) | eGFR decline (General population) | 0,000569632 | 0,000766898 | 0,458172143 |
| Hypertension | eGFR decline (General population) | 0,001661851 | 0,00166116 | 0,318311919 |
| Urinary sodium-potassium ratio | eGFR decline (General population) | 0,003386301 | 0,00500965 | 0,506113191 |
| Insomnia | eGFR decline (General population) | -0,001119792 | 0,004706781 | 0,816324067 |
| LDL cholesterol | eGFR decline (General population) | -0,001549421 | 0,00056925 | 0,006990716 |
| Low TSH | eGFR decline (General population) | -0,001901804 | 0,002981067 | 0,529299824 |
| Compound smoking index | eGFR decline (General population) | -0,003767386 | 0,001834135 | 0,042181016 |
| Type 2 diabetes | eGFR decline (General population) | 0,000381079 | 0,000512217 | 0,457278487 |
| Total cholesterol | eGFR decline (General population) | -0,000798383 | 0,000502281 | 0,113084903 |
| Triglyceride | eGFR decline (General population) | 0,000137394 | 0,000575271 | 0,811448586 |
| Thyroid Stimulating Hormone | eGFR decline (General population) | 0,000828977 | 0,000744935 | 0,267435241 |
| Sleep duration | eGFR decline (General population) | -0,001219207 | 0,002344134 | 0,604754111 |
| Alcohol intake frequency. | eGFR decline (General population) | -0,000138263 | 0,001358021 | 0,919124915 |
| Lipoprotein A | eGFR decline (General population) | -0,003547466 | 0,004450967 | 0,435832701 |
| Waist-to-hip ratio adjusted BMI | eGFR decline (General population) | 0,000570727 | 0,00074317 | 0,443125422 |
| Zinc | eGFR decline (General population) | 0,002764939 | 0,010294806 | 0,832961042 |
| Body mass index | eGFR decline (CKD population) [DB01] | -0,000317161 | 0,003279299 | 0,922990354 |
| Heel bone mineral density | eGFR decline (CKD population) [DB01] | 0,001750242 | 0,002689803 | 0,515553862 |

| | | | | |
|---|---|---|---|---|
| Two-hour glucose | eGFR decline (CKD population) [DB01] | 0,025408569 | 0,024561259 | 0,321302005 |
| Fasting glucose | eGFR decline (CKD population) [DB01] | -0,009590149 | 0,007208633 | 0,187975073 |
| Fasting insulin | eGFR decline (CKD population) [DB01] | -0,007878672 | 0,018994215 | 0,68082486 |
| circulating leptin levels | eGFR decline (CKD population) [DB01] | -0,178714773 | 0,118595798 | 0,372982244 |
| Growth differentiation factor-15 levels | eGFR decline (CKD population) [DB01] | 0,011528918 | 0,026037191 | 0,734632021 |
| Fibroblast growth factor 23 levels | eGFR decline (CKD population) [DB01] | 0,006929055 | 0,039478635 | 0,866449238 |
| Kidney injury molecule 1 levels | eGFR decline (CKD population) [DB01] | -0,004049985 | 0,018739233 | 0,835054645 |
| Coronary artery disease (Firth correction) | eGFR decline (CKD population) [DB01] | 0,006786087 | 0,011589332 | 0,561937477 |
| Body fat percentage | eGFR decline (CKD population) [DB01] | -0,005514005 | 0,005567037 | 0,322705178 |
| Sodium in urine | eGFR decline (CKD population) [DB01] | -0,020855498 | 0,018198668 | 0,258782335 |
| Glycated haemoglobin HbA1c levels | eGFR decline (CKD population) [DB01] | -0,00606055 | 0,003338684 | 0,070483714 |
| Phosphate levels | eGFR decline (CKD population) [DB01] | -0,006369996 | 0,00445061 | 0,154329739 |
| Bone mineral density | eGFR decline (CKD population) [DB01] | 0,000713474 | 0,003002495 | 0,812288196 |
| Type 1 diabetes | eGFR decline (CKD population) [DB01] | 0,001338789 | 0,005799507 | 0,818072086 |
| Liver iron content | eGFR decline (CKD population) [DB01] | -0,001634615 | 0,038531782 | 0,968827463 |
| Chronic heart failure | eGFR decline (CKD population) [DB01] | -0,043919467 | 0,097566204 | 0,683141756 |
| Hyperthyroidism | eGFR decline (CKD population) [DB01] | 0,046988786 | 0,053673418 | 0,414992643 |
| Hypothyroidism | eGFR decline (CKD population) [DB01] | 0,009319412 | 0,009341432 | 0,322330474 |
| Myocardial infarction | eGFR decline (CKD population) [DB01] | 0,01433446 | 0,008735973 | 0,105445507 |
| Serum uric acid levels | eGFR decline (CKD population) [DB01] | 0,002266625 | 0,003613431 | 0,531086117 |
| Vitamin K-dependent protein S measurement | eGFR decline (CKD population) [DB01] | -0,008868885 | 0,029977302 | 0,795232886 |
| Vitamin K-dependent protein C measurement | eGFR decline (CKD population) [DB01] | -0,01775275 | 0,08366356 | 0,866888649 |
| Serum alkaline phosphatase levels | eGFR decline (CKD population) [DB01] | 0,001931256 | 0,002878933 | 0,502798034 |
| Vitamin D levels | eGFR decline (CKD population) [DB01] | -0,001297054 | 0,005708875 | 0,820855467 |
| Hemoglobin levels | eGFR decline (CKD population) [DB01] | -0,002699985 | 0,003256669 | 0,407680885 |
| Blood glucose levels | eGFR decline (CKD population) [DB01] | -0,005856048 | 0,005526996 | 0,291741917 |
| Insulin-like growth factor 1 levels | eGFR decline (CKD population) [DB01] | 0,006005375 | 0,003009148 | 0,046696667 |
| Calcium levels | eGFR decline (CKD population) [DB01] | -0,007296566 | 0,004000695 | 0,069671096 |
| Educational attainment (years of education) | eGFR decline (CKD population) [DB01] | -0,005238578 | 0,008907884 | 0,557117397 |
| Smoking status | eGFR decline (CKD population) [DB01] | 0,004034728 | 0,011215349 | 0,719693452 |
| C-reactive protein levels | eGFR decline (CKD population) [DB01] | -0,005955923 | 0,003501978 | 0,090459801 |
| Liver fat | eGFR decline (CKD population) [DB01] | -0,01603638 | 0,019069407 | 0,432607049 |
| Vitamin B12 deficiency anaemia | eGFR decline (CKD population) [DB01] | 0,01528378 | 0,049726514 | 0,787623776 |
| Obesity | eGFR decline (CKD population) [DB01] | 0,01394974 | 0,045631907 | 0,77215017 |
| Pulmonary heart disease | eGFR decline (CKD population) [DB01] | 0,067170389 | 0,04021383 | 0,193446746 |
| Free triiodothyronine | eGFR decline (CKD population) [DB01] | 0,018136248 | 0,051969315 | 0,744700273 |
| FT3 FT4 ratio | eGFR decline (CKD population) [DB01] | -0,018185195 | 0,015496118 | 0,265366235 |
| Free thyroxine | eGFR decline (CKD population) [DB01] | 0,002530401 | 0,007787569 | 0,746365669 |
| HDL cholesterol | eGFR decline (CKD population) [DB01] | -0,00421306 | 0,003329497 | 0,207012094 |
| High TSH | eGFR decline (CKD population) [DB01] | -0,032157795 | 0,052203649 | 0,571216431 |
| Major depression | eGFR decline (CKD population) [DB01] | -0,011638415 | 0,024533623 | 0,637468743 |
| apolipoprotein A-I | eGFR decline (CKD population) [DB01] | 0,002370101 | 0,003142323 | 0,451363131 |
| apolipoprotein B | eGFR decline (CKD population) [DB01] | 0,000845661 | 0,003908199 | 0,828948573 |
| Cigarettes per Day | eGFR decline (CKD population) [DB01] | 0,006786286 | 0,011189297 | 0,551007083 |
| Chronotype | eGFR decline (CKD population) [DB01] | 0,014332303 | 0,01116918 | 0,204525397 |
| smoking initiation | eGFR decline (CKD population) [DB01] | -0,03639695 | 0,015637522 | 0,022371937 |
| Systolic Blood Pressure (SBP) | eGFR decline (CKD population) [DB01] | 0,003347447 | 0,005795148 | 0,564036489 |
| Diastolic Blood Pressure (DBP) | eGFR decline (CKD population) [DB01] | 0,00057688 | 0,006079711 | 0,92448168 |
| Pulse Pressure (PP) | eGFR decline (CKD population) [DB01] | 0,002138373 | 0,004577597 | 0,640739777 |
| Hypertension | eGFR decline (CKD population) [DB01] | 0,007717967 | 0,006783058 | 0,256662912 |
| Urinary sodium-potassium ratio | eGFR decline (CKD population) [DB01] | -0,009599373 | 0,029099018 | 0,744606285 |
| Insomnia | eGFR decline (CKD population) [DB01] | 0,002403262 | 0,028772203 | 0,934933017 |
| LDL cholesterol | eGFR decline (CKD population) [DB01] | 2,9428E-05 | 0,00346677 | 0,993235034 |
| Low TSH | eGFR decline (CKD population) [DB01] | 0,004810678 | 0,017630577 | 0,787294967 |
| Compound smoking index | eGFR decline (CKD population) [DB01] | -0,005669019 | 0,010504819 | 0,590440707 |
| Type 2 diabetes | eGFR decline (CKD population) [DB01] | -0,001802308 | 0,002959883 | 0,542897812 |
| Total cholesterol | eGFR decline (CKD population) [DB01] | 0,002210534 | 0,003046461 | 0,468727385 |
| Triglyceride | eGFR decline (CKD population) [DB01] | -0,003567797 | 0,003228838 | 0,270392444 |
| Thyroid Stimulating Hormone | eGFR decline (CKD population) [DB01] | -0,00597322 | 0,00436038 | 0,172678666 |
| Sleep duration | eGFR decline (CKD population) [DB01] | -0,002190039 | 0,013533821 | 0,871957596 |
| Alcohol intake frequency. | eGFR decline (CKD population) [DB01] | 0,000570393 | 0,006821427 | 0,933541991 |
| Lipoprotein A | eGFR decline (CKD population) [DB01] | 0,006057967 | 0,035686246 | 0,868956239 |
| Waist-to-hip ratio adjusted BMI | eGFR decline (CKD population) [DB01] | 0,004234886 | 0,004009558 | 0,291745039 |
| Zinc | eGFR decline (CKD population) [DB01] | 0,05072553 | 0,069788478 | 0,599873252 |
| Body mass index | eGFR decline (CKD population) [SH01] | -0,00043514 | 0,00332336 | 0,895879858 |
| Heel bone mineral density | eGFR decline (CKD population) [SH01] | 0,001433741 | 0,002725827 | 0,599141861 |
| Two-hour glucose | eGFR decline (CKD population) [SH01] | 0,026978643 | 0,02547336 | 0,310410964 |

| | | | | |
|---|---|---|---|---|
| Fasting glucose | eGFR decline (CKD population) [SH01] | -0,009383786 | 0,007311887 | 0,203853776 |
| Fasting insulin | eGFR decline (CKD population) [SH01] | -0,009371868 | 0,019032861 | 0,625508118 |
| circulating leptin levels | eGFR decline (CKD population) [SH01] | -0,177412079 | 0,120049124 | 0,378721833 |
| Growth differentiation factor-15 levels | eGFR decline (CKD population) [SH01] | 0,009605389 | 0,026404428 | 0,777885025 |
| Fibroblast growth factor 23 levels | eGFR decline (CKD population) [SH01] | 0,016597584 | 0,043055675 | 0,713170018 |
| Kidney injury molecule 1 levels | eGFR decline (CKD population) [SH01] | -0,010381377 | 0,017637575 | 0,57462605 |
| Coronary artery disease (Firth correction) | eGFR decline (CKD population) [SH01] | 0,007375784 | 0,011744903 | 0,534081324 |
| Body fat percentage | eGFR decline (CKD population) [SH01] | -0,005940562 | 0,005652459 | 0,294080764 |
| Sodium in urine | eGFR decline (CKD population) [SH01] | -0,019367951 | 0,018454642 | 0,300413174 |
| Glycated haemoglobin HbA1c levels | eGFR decline (CKD population) [SH01] | -0,006131894 | 0,003383478 | 0,070938692 |
| Phosphate levels | eGFR decline (CKD population) [SH01] | -0,006307794 | 0,004512773 | 0,164143189 |
| Bone mineral density | eGFR decline (CKD population) [SH01] | 0,000620995 | 0,003042625 | 0,838377234 |
| Type 1 diabetes | eGFR decline (CKD population) [SH01] | 0,00150555 | 0,005875953 | 0,798490184 |
| Liver iron content | eGFR decline (CKD population) [SH01] | -0,003375757 | 0,039220594 | 0,936832661 |
| Chronic heart failure | eGFR decline (CKD population) [SH01] | -0,037203255 | 0,098876005 | 0,731756121 |
| Hyperthyroidism | eGFR decline (CKD population) [SH01] | 0,048353111 | 0,054848252 | 0,411924183 |
| Hypothyroidism | eGFR decline (CKD population) [SH01] | 0,009472877 | 0,009573264 | 0,326257923 |
| Myocardial infarction | eGFR decline (CKD population) [SH01] | 0,014713624 | 0,008856985 | 0,101268374 |
| Serum uric acid levels | eGFR decline (CKD population) [SH01] | 0,001050573 | 0,003663573 | 0,774547398 |
| Vitamin K-dependent protein S measurement | eGFR decline (CKD population) [SH01] | -0,010062659 | 0,030378071 | 0,771944961 |
| Vitamin K-dependent protein C measurement | eGFR decline (CKD population) [SH01] | -0,017524543 | 0,08120914 | 0,864695315 |
| Serum alkaline phosphatase levels | eGFR decline (CKD population) [SH01] | 0,001706014 | 0,002914371 | 0,558688848 |
| Vitamin D levels | eGFR decline (CKD population) [SH01] | -0,001491576 | 0,005788086 | 0,797310382 |
| Hemoglobin levels | eGFR decline (CKD population) [SH01] | -0,002824499 | 0,003338392 | 0,398142346 |
| Blood glucose levels | eGFR decline (CKD population) [SH01] | -0,005717316 | 0,005605293 | 0,310036167 |
| Insulin-like growth factor 1 levels | eGFR decline (CKD population) [SH01] | 0,006065692 | 0,003049655 | 0,047437877 |
| Calcium levels | eGFR decline (CKD population) [SH01] | -0,007622243 | 0,004057793 | 0,061778624 |
| Educational attainment (years of education) | eGFR decline (CKD population) [SH01] | -0,005486281 | 0,009026917 | 0,544006858 |
| Smoking status | eGFR decline (CKD population) [SH01] | 0,004294825 | 0,011365919 | 0,706224691 |
| C-reactive protein levels | eGFR decline (CKD population) [SH01] | -0,006195153 | 0,003548687 | 0,08230146 |
| Liver fat | eGFR decline (CKD population) [SH01] | -0,01507376 | 0,019349589 | 0,465572133 |
| Vitamin B12 deficiency anaemia | eGFR decline (CKD population) [SH01] | 0,015878624 | 0,050367939 | 0,782423131 |
| Obesity | eGFR decline (CKD population) [SH01] | 0,019338011 | 0,046298171 | 0,693508488 |
| Pulmonary heart disease | eGFR decline (CKD population) [SH01] | 0,070370583 | 0,040741205 | 0,182573313 |
| Free triiodothyronine | eGFR decline (CKD population) [SH01] | 0,016298442 | 0,053201007 | 0,774617719 |
| FT3 FT4 ratio | eGFR decline (CKD population) [SH01] | -0,018838997 | 0,015700465 | 0,255383344 |
| Free thyroxine | eGFR decline (CKD population) [SH01] | 0,003451773 | 0,008094034 | 0,671299713 |
| HDL cholesterol | eGFR decline (CKD population) [SH01] | -0,003681359 | 0,003383264 | 0,277681098 |
| High TSH | eGFR decline (CKD population) [SH01] | -0,036552674 | 0,055037767 | 0,542928441 |
| Major depression | eGFR decline (CKD population) [SH01] | -0,014068782 | 0,024791761 | 0,573148684 |
| apolipoprotein A-I | eGFR decline (CKD population) [SH01] | 0,002579446 | 0,003184671 | 0,41868621 |
| apolipoprotein B | eGFR decline (CKD population) [SH01] | 0,001056551 | 0,003960378 | 0,789959665 |
| Cigarettes per Day | eGFR decline (CKD population) [SH01] | 0,006741645 | 0,011337987 | 0,558773756 |
| Chronotype | eGFR decline (CKD population) [SH01] | 0,014330966 | 0,011339204 | 0,211342666 |
| smoking initiation | eGFR decline (CKD population) [SH01] | -0,037066586 | 0,015847356 | 0,021740803 |
| Systolic Blood Pressure (SBP) | eGFR decline (CKD population) [SH01] | 0,002915639 | 0,005877574 | 0,620288583 |
| Diastolic Blood Pressure (DBP) | eGFR decline (CKD population) [SH01] | 0,000219595 | 0,00616547 | 0,971616413 |
| Pulse Pressure (PP) | eGFR decline (CKD population) [SH01] | 0,002638126 | 0,004641716 | 0,570221438 |
| Hypertension | eGFR decline (CKD population) [SH01] | 0,007781171 | 0,006884205 | 0,259815964 |
| Urinary sodium-potassium ratio | eGFR decline (CKD population) [SH01] | -0,007509603 | 0,02982946 | 0,803570192 |
| Insomnia | eGFR decline (CKD population) [SH01] | 0,003155777 | 0,029547769 | 0,916868591 |
| LDL cholesterol | eGFR decline (CKD population) [SH01] | 0,000282866 | 0,003542529 | 0,936432008 |
| Low TSH | eGFR decline (CKD population) [SH01] | 0,003632517 | 0,018471852 | 0,845756043 |
| Compound smoking index | eGFR decline (CKD population) [SH01] | -0,005338385 | 0,01064028 | 0,616795179 |
| Type 2 diabetes | eGFR decline (CKD population) [SH01] | -0,0018745 | 0,002999229 | 0,532299875 |
| Total cholesterol | eGFR decline (CKD population) [SH01] | 0,002499625 | 0,00308751 | 0,418909113 |
| Triglyceride | eGFR decline (CKD population) [SH01] | -0,004179342 | 0,003272887 | 0,202981471 |
| Thyroid Stimulating Hormone | eGFR decline (CKD population) [SH01] | -0,005921349 | 0,004475357 | 0,187725575 |
| Sleep duration | eGFR decline (CKD population) [SH01] | -0,003439757 | 0,013714833 | 0,802767886 |
| Alcohol intake frequency. | eGFR decline (CKD population) [SH01] | 0,000377895 | 0,006923446 | 0,956589948 |
| Lipoprotein A | eGFR decline (CKD population) [SH01] | 0,00400918 | 0,035218053 | 0,911864637 |
| Waist-to-hip ratio adjusted BMI | eGFR decline (CKD population) [SH01] | 0,004344381 | 0,004063226 | 0,285858811 |
| Zinc | eGFR decline (CKD population) [SH01] | 0,049297954 | 0,072870453 | 0,621345102 |
| Body mass index | eGFR decline (CKD population) [SH02] | -0,000292113 | 0,003285146 | 0,929181364 |
| Heel bone mineral density | eGFR decline (CKD population) [SH02] | 0,001776387 | 0,002693763 | 0,509926154 |
| Two-hour glucose | eGFR decline (CKD population) [SH02] | 0,025101995 | 0,024486344 | 0,325517896 |
| Fasting glucose | eGFR decline (CKD population) [SH02] | -0,009551671 | 0,007187531 | 0,18844969 |

| | | | | |
|---|---|---|---|---|
| Fasting insulin | eGFR decline (CKD population) [SH02] | -0,007974311 | 0,01900814 | 0,67739946 |
| circulating leptin levels | eGFR decline (CKD population) [SH02] | -0,178814141 | 0,118658696 | 0,372974724 |
| Growth differentiation factor-15 levels | eGFR decline (CKD population) [SH02] | 0,011799565 | 0,026135825 | 0,730024535 |
| Fibroblast growth factor 23 levels | eGFR decline (CKD population) [SH02] | 0,005040967 | 0,039074521 | 0,901566401 |
| Kidney injury molecule 1 levels | eGFR decline (CKD population) [SH02] | -0,00495613 | 0,018596853 | 0,797531133 |
| Coronary artery disease (Firth correction) | eGFR decline (CKD population) [SH02] | 0,00678689 | 0,011614872 | 0,562747652 |
| Body fat percentage | eGFR decline (CKD population) [SH02] | -0,005458426 | 0,005560636 | 0,327043291 |
| Sodium in urine | eGFR decline (CKD population) [SH02] | -0,020920471 | 0,018248692 | 0,258611104 |
| Glycated haemoglobin HbA1c levels | eGFR decline (CKD population) [SH02] | -0,006064541 | 0,003343872 | 0,070734087 |
| Phosphate levels | eGFR decline (CKD population) [SH02] | -0,006482607 | 0,004463586 | 0,148392373 |
| Bone mineral density | eGFR decline (CKD population) [SH02] | 0,000711025 | 0,003006885 | 0,813188695 |
| Type 1 diabetes | eGFR decline (CKD population) [SH02] | 0,001324533 | 0,005807807 | 0,820228061 |
| Liver iron content | eGFR decline (CKD population) [SH02] | -0,001463921 | 0,038474993 | 0,972039252 |
| Chronic heart failure | eGFR decline (CKD population) [SH02] | -0,044602789 | 0,097731411 | 0,679129728 |
| Hyperthyroidism | eGFR decline (CKD population) [SH02] | 0,046815428 | 0,053662455 | 0,416528444 |
| Hypothyroidism | eGFR decline (CKD population) [SH02] | 0,009364232 | 0,009311981 | 0,318511285 |
| Myocardial infarction | eGFR decline (CKD population) [SH02] | 0,0143465 | 0,008759359 | 0,106072987 |
| Serum uric acid levels | eGFR decline (CKD population) [SH02] | 0,002324095 | 0,003625862 | 0,522162132 |
| Vitamin K-dependent protein S measurement | eGFR decline (CKD population) [SH02] | -0,008773582 | 0,030000233 | 0,797490987 |
| Vitamin K-dependent protein C measurement | eGFR decline (CKD population) [SH02] | -0,017760361 | 0,083767174 | 0,866993196 |
| Serum alkaline phosphatase levels | eGFR decline (CKD population) [SH02] | 0,001970683 | 0,002880703 | 0,494389112 |
| Vitamin D levels | eGFR decline (CKD population) [SH02] | -0,001139497 | 0,005723094 | 0,842692049 |
| Hemoglobin levels | eGFR decline (CKD population) [SH02] | -0,002835131 | 0,003251119 | 0,383829968 |
| Blood glucose levels | eGFR decline (CKD population) [SH02] | -0,005917156 | 0,005506257 | 0,284960477 |
| Insulin-like growth factor 1 levels | eGFR decline (CKD population) [SH02] | 0,00598677 | 0,003016118 | 0,04788883 |
| Calcium levels | eGFR decline (CKD population) [SH02] | -0,007263252 | 0,004018424 | 0,072188869 |
| Educational attainment (years of education) | eGFR decline (CKD population) [SH02] | -0,00518107 | 0,008918728 | 0,56192613 |
| Smoking status | eGFR decline (CKD population) [SH02] | 0,004027054 | 0,011231742 | 0,72059495 |
| C-reactive protein levels | eGFR decline (CKD population) [SH02] | -0,005936912 | 0,0035089 | 0,092123171 |
| Liver fat | eGFR decline (CKD population) [SH02] | -0,01599186 | 0,01917332 | 0,436183373 |
| Vitamin B12 deficiency anaemia | eGFR decline (CKD population) [SH02] | 0,015201019 | 0,049755904 | 0,788841556 |
| Obesity | eGFR decline (CKD population) [SH02] | 0,013455361 | 0,045633109 | 0,779951657 |
| Pulmonary heart disease | eGFR decline (CKD population) [SH02] | 0,066880544 | 0,040250018 | 0,195172905 |
| Free triiodothyronine | eGFR decline (CKD population) [SH02] | 0,018297982 | 0,052001156 | 0,742686049 |
| FT3 FT4 ratio | eGFR decline (CKD population) [SH02] | -0,018123204 | 0,015511084 | 0,267343193 |
| Free thyroxine | eGFR decline (CKD population) [SH02] | 0,002289573 | 0,007764739 | 0,769112597 |
| HDL cholesterol | eGFR decline (CKD population) [SH02] | -0,00423757 | 0,003322143 | 0,203394756 |
| High TSH | eGFR decline (CKD population) [SH02] | -0,031041742 | 0,051722677 | 0,58074605 |
| Major depression | eGFR decline (CKD population) [SH02] | -0,011453623 | 0,024600523 | 0,643709835 |
| apolipoprotein A-I | eGFR decline (CKD population) [SH02] | 0,002341068 | 0,003147486 | 0,457657646 |
| apolipoprotein B | eGFR decline (CKD population) [SH02] | 0,000846886 | 0,00391605 | 0,829042722 |
| Cigarettes per Day | eGFR decline (CKD population) [SH02] | 0,006779935 | 0,011201587 | 0,551809385 |
| Chronotype | eGFR decline (CKD population) [SH02] | 0,01434273 | 0,01118517 | 0,204839411 |
| smoking initiation | eGFR decline (CKD population) [SH02] | -0,036314479 | 0,015662154 | 0,022875759 |
| Systolic Blood Pressure (SBP) | eGFR decline (CKD population) [SH02] | 0,003291837 | 0,005817468 | 0,572002723 |
| Diastolic Blood Pressure (DBP) | eGFR decline (CKD population) [SH02] | 0,00046166 | 0,006101139 | 0,939744113 |
| Pulse Pressure (PP) | eGFR decline (CKD population) [SH02] | 0,002010119 | 0,004593937 | 0,662020742 |
| Hypertension | eGFR decline (CKD population) [SH02] | 0,007930561 | 0,00682342 | 0,246628696 |
| Urinary sodium-potassium ratio | eGFR decline (CKD population) [SH02] | -0,009722722 | 0,029123463 | 0,741657816 |
| Insomnia | eGFR decline (CKD population) [SH02] | 0,002309966 | 0,028724449 | 0,937349171 |
| LDL cholesterol | eGFR decline (CKD population) [SH02] | 4,40675E-06 | 0,003458985 | 0,998984676 |
| Low TSH | eGFR decline (CKD population) [SH02] | 0,006175393 | 0,017451795 | 0,726537835 |
| Compound smoking index | eGFR decline (CKD population) [SH02] | -0,005709017 | 0,010515135 | 0,588191225 |
| Type 2 diabetes | eGFR decline (CKD population) [SH02] | -0,001818657 | 0,002963615 | 0,539756687 |
| Total cholesterol | eGFR decline (CKD population) [SH02] | 0,002203736 | 0,003052026 | 0,47090342 |
| Triglyceride | eGFR decline (CKD population) [SH02] | -0,003467635 | 0,003236995 | 0,285246467 |
| Thyroid Stimulating Hormone | eGFR decline (CKD population) [SH02] | -0,006048438 | 0,004350752 | 0,166434342 |
| Sleep duration | eGFR decline (CKD population) [SH02] | -0,002090091 | 0,013551852 | 0,877914351 |
| Alcohol intake frequency. | eGFR decline (CKD population) [SH02] | 0,000701172 | 0,006853979 | 0,918740081 |
| Lipoprotein A | eGFR decline (CKD population) [SH02] | 0,006286319 | 0,035822439 | 0,864582889 |
| Waist-to-hip ratio adjusted BMI | eGFR decline (CKD population) [SH02] | 0,004198143 | 0,004015287 | 0,296632403 |
| Zinc | eGFR decline (CKD population) [SH02] | 0,051000395 | 0,069366407 | 0,596394309 |
| Body mass index | eGFR decline (CKD population) [SH03] | -0,000235974 | 0,00328838 | 0,942821486 |
| Heel bone mineral density | eGFR decline (CKD population) [SH03] | 0,001869145 | 0,002695105 | 0,488309533 |
| Two-hour glucose | eGFR decline (CKD population) [SH03] | 0,024406115 | 0,024241268 | 0,333901349 |
| Fasting glucose | eGFR decline (CKD population) [SH03] | -0,009528853 | 0,007144552 | 0,18687919 |
| Fasting insulin | eGFR decline (CKD population) [SH03] | -0,007867128 | 0,01903718 | 0,681943961 |

| | | | | |
|---|---|---|---|---|
| circulating leptin levels | eGFR decline (CKD population) [SH03] | -0,179179144 | 0,118563085 | 0,372140678 |
| Growth differentiation factor-15 levels | eGFR decline (CKD population) [SH03] | 0,012534458 | 0,026241353 | 0,716311511 |
| Fibroblast growth factor 23 levels | eGFR decline (CKD population) [SH03] | 0,000797776 | 0,038936825 | 0,984317665 |
| Kidney injury molecule 1 levels | eGFR decline (CKD population) [SH03] | -0,004628758 | 0,018758685 | 0,812179627 |
| Coronary artery disease (Firth correction) | eGFR decline (CKD population) [SH03] | 0,00668892 | 0,011634039 | 0,569006943 |
| Body fat percentage | eGFR decline (CKD population) [SH03] | -0,005305551 | 0,005539803 | 0,338943437 |
| Sodium in urine | eGFR decline (CKD population) [SH03] | -0,021281587 | 0,018292649 | 0,251737821 |
| Glycated haemoglobin HbA1c levels | eGFR decline (CKD population) [SH03] | -0,006057462 | 0,003345939 | 0,071235415 |
| Phosphate levels | eGFR decline (CKD population) [SH03] | -0,006651989 | 0,004473854 | 0,139044919 |
| Bone mineral density | eGFR decline (CKD population) [SH03] | 0,000723316 | 0,003008335 | 0,810110431 |
| Type 1 diabetes | eGFR decline (CKD population) [SH03] | 0,001273078 | 0,005810554 | 0,827177221 |
| Liver iron content | eGFR decline (CKD population) [SH03] | -0,000905991 | 0,038277246 | 0,982602837 |
| Chronic heart failure | eGFR decline (CKD population) [SH03] | -0,046778223 | 0,097816355 | 0,665177669 |
| Hyperthyroidism | eGFR decline (CKD population) [SH03] | 0,046331564 | 0,053471721 | 0,419528522 |
| Hypothyroidism | eGFR decline (CKD population) [SH03] | 0,009381308 | 0,009236573 | 0,313733898 |
| Myocardial infarction | eGFR decline (CKD population) [SH03] | 0,014309998 | 0,008779068 | 0,107722607 |
| Serum uric acid levels | eGFR decline (CKD population) [SH03] | 0,002645516 | 0,003636814 | 0,467688616 |
| Vitamin K-dependent protein S measurement | eGFR decline (CKD population) [SH03] | -0,008425795 | 0,029982628 | 0,805097821 |
| Vitamin K-dependent protein C measurement | eGFR decline (CKD population) [SH03] | -0,017809338 | 0,084329545 | 0,867500848 |
| Serum alkaline phosphatase levels | eGFR decline (CKD population) [SH03] | 0,002068924 | 0,00288283 | 0,473463225 |
| Vitamin D levels | eGFR decline (CKD population) [SH03] | -0,000872355 | 0,005734202 | 0,879470744 |
| Hemoglobin levels | eGFR decline (CKD population) [SH03] | -0,002995406 | 0,003248746 | 0,357206839 |
| Blood glucose levels | eGFR decline (CKD population) [SH03] | -0,006021264 | 0,005467432 | 0,273238914 |
| Insulin-like growth factor 1 levels | eGFR decline (CKD population) [SH03] | 0,005948969 | 0,003021432 | 0,049705784 |
| Calcium levels | eGFR decline (CKD population) [SH03] | -0,007144725 | 0,00403757 | 0,078324204 |
| Educational attainment (years of education) | eGFR decline (CKD population) [SH03] | -0,005061809 | 0,008919703 | 0,570998784 |
| Smoking status | eGFR decline (CKD population) [SH03] | 0,003969773 | 0,011237123 | 0,724530469 |
| C-reactive protein levels | eGFR decline (CKD population) [SH03] | -0,00586899 | 0,003513343 | 0,096298953 |
| Liver fat | eGFR decline (CKD population) [SH03] | -0,016059173 | 0,019294549 | 0,437097441 |
| Vitamin B12 deficiency anaemia | eGFR decline (CKD population) [SH03] | 0,014972973 | 0,049808758 | 0,792082444 |
| Obesity | eGFR decline (CKD population) [SH03] | 0,011788987 | 0,045534075 | 0,806029552 |
| Pulmonary heart disease | eGFR decline (CKD population) [SH03] | 0,065893574 | 0,040236532 | 0,200018224 |
| Free triiodothyronine | eGFR decline (CKD population) [SH03] | 0,018858987 | 0,051960825 | 0,735011245 |
| FT3 FT4 ratio | eGFR decline (CKD population) [SH03] | -0,017916853 | 0,015507299 | 0,272414284 |
| Free thyroxine | eGFR decline (CKD population) [SH03] | 0,001796239 | 0,007779456 | 0,818181804 |
| HDL cholesterol | eGFR decline (CKD population) [SH03] | -0,004364779 | 0,003304619 | 0,187871156 |
| High TSH | eGFR decline (CKD population) [SH03] | -0,02862513 | 0,050526397 | 0,601304477 |
| Major depression | eGFR decline (CKD population) [SH03] | -0,010797067 | 0,024667799 | 0,663652974 |
| apolipoprotein A-I | eGFR decline (CKD population) [SH03] | 0,002263267 | 0,003149818 | 0,473053303 |
| apolipoprotein B | eGFR decline (CKD population) [SH03] | 0,000809593 | 0,003921172 | 0,836671719 |
| Cigarettes per Day | eGFR decline (CKD population) [SH03] | 0,006778583 | 0,011201008 | 0,551867599 |
| Chronotype | eGFR decline (CKD population) [SH03] | 0,01435829 | 0,01118973 | 0,204537309 |
| smoking initiation | eGFR decline (CKD population) [SH03] | -0,036079122 | 0,015672506 | 0,023836412 |
| Systolic Blood Pressure (SBP) | eGFR decline (CKD population) [SH03] | 0,003312747 | 0,005839319 | 0,571009658 |
| Diastolic Blood Pressure (DBP) | eGFR decline (CKD population) [SH03] | 0,000355889 | 0,006120466 | 0,953677912 |
| Pulse Pressure (PP) | eGFR decline (CKD population) [SH03] | 0,001756956 | 0,004609488 | 0,703353758 |
| Hypertension | eGFR decline (CKD population) [SH03] | 0,008102204 | 0,006864971 | 0,239426439 |
| Urinary sodium-potassium ratio | eGFR decline (CKD population) [SH03] | -0,010277321 | 0,029074294 | 0,727089883 |
| Insomnia | eGFR decline (CKD population) [SH03] | 0,002035892 | 0,028525236 | 0,944383127 |
| LDL cholesterol | eGFR decline (CKD population) [SH03] | -7,65229E-05 | 0,003437284 | 0,982259121 |
| Low TSH | eGFR decline (CKD population) [SH03] | 0,007970765 | 0,017392949 | 0,650877083 |
| Compound smoking index | eGFR decline (CKD population) [SH03] | -0,005825911 | 0,010513157 | 0,580513153 |
| Type 2 diabetes | eGFR decline (CKD population) [SH03] | -0,001830618 | 0,00296412 | 0,537164147 |
| Total cholesterol | eGFR decline (CKD population) [SH03] | 0,002142787 | 0,003055186 | 0,48370058 |
| Triglyceride | eGFR decline (CKD population) [SH03] | -0,003221282 | 0,003243535 | 0,3217482 |
| Thyroid Stimulating Hormone | eGFR decline (CKD population) [SH03] | -0,006155509 | 0,004356381 | 0,159639037 |
| Sleep duration | eGFR decline (CKD population) [SH03] | -0,001723545 | 0,013555839 | 0,899224981 |
| Alcohol intake frequency. | eGFR decline (CKD population) [SH03] | 0,000922141 | 0,00689082 | 0,893835885 |
| Lipoprotein A | eGFR decline (CKD population) [SH03] | 0,006983346 | 0,036122302 | 0,850996972 |
| Waist-to-hip ratio adjusted BMI | eGFR decline (CKD population) [SH03] | 0,004124867 | 0,004016992 | 0,305332969 |
| Zinc | eGFR decline (CKD population) [SH03] | 0,051666611 | 0,068192953 | 0,587227707 |
| Body mass index | eGFR decline (CKD population) [SH04] | -4,93008E-05 | 0,003253644 | 0,987916513 |
| Heel bone mineral density | eGFR decline (CKD population) [SH04] | 0,002343671 | 0,002665829 | 0,379757409 |
| Two-hour glucose | eGFR decline (CKD population) [SH04] | 0,022001674 | 0,02319478 | 0,361556075 |
| Fasting glucose | eGFR decline (CKD population) [SH04] | -0,009788303 | 0,007077625 | 0,17132821 |
| Fasting insulin | eGFR decline (CKD population) [SH04] | -0,005888682 | 0,019198956 | 0,760874772 |
| circulating leptin levels | eGFR decline (CKD population) [SH04] | -0,181097572 | 0,117283341 | 0,365867873 |

| | | | | |
|---|---|---|---|---|
| Growth differentiation factor-15 levels | eGFR decline (CKD population) [SH04] | 0,015521448 | 0,026004473 | 0,657422581 |
| Fibroblast growth factor 23 levels | eGFR decline (CKD population) [SH04] | -0,013639806 | 0,038699105 | 0,736545189 |
| Kidney injury molecule 1 levels | eGFR decline (CKD population) [SH04] | 0,001596486 | 0,020666916 | 0,940587592 |
| Coronary artery disease (Firth correction) | eGFR decline (CKD population) [SH04] | 0,005860043 | 0,011516606 | 0,614061255 |
| Body fat percentage | eGFR decline (CKD population) [SH04] | -0,004663077 | 0,005462293 | 0,39393035 |
| Sodium in urine | eGFR decline (CKD population) [SH04] | -0,023505053 | 0,018107734 | 0,201888847 |
| Glycated haemoglobin HbA1c levels | eGFR decline (CKD population) [SH04] | -0,005954068 | 0,003309799 | 0,073036121 |
| Phosphate levels | eGFR decline (CKD population) [SH04] | -0,006960597 | 0,004428563 | 0,118008808 |
| Bone mineral density | eGFR decline (CKD population) [SH04] | 0,000850762 | 0,00297569 | 0,775095033 |
| Type 1 diabetes | eGFR decline (CKD population) [SH04] | 0,001013745 | 0,005748362 | 0,860497795 |
| Liver iron content | eGFR decline (CKD population) [SH04] | 0,001775545 | 0,037322434 | 0,965046326 |
| Chronic heart failure | eGFR decline (CKD population) [SH04] | -0,057044895 | 0,096776678 | 0,597007695 |
| Hyperthyroidism | eGFR decline (CKD population) [SH04] | 0,044300446 | 0,05213439 | 0,428070216 |
| Hypothyroidism | eGFR decline (CKD population) [SH04] | 0,009155756 | 0,008959624 | 0,31080354 |
| Myocardial infarction | eGFR decline (CKD population) [SH04] | 0,013850889 | 0,008690807 | 0,115632181 |
| Serum uric acid levels | eGFR decline (CKD population) [SH04] | 0,004330306 | 0,003602234 | 0,230528224 |
| Vitamin K-dependent protein S measurement | eGFR decline (CKD population) [SH04] | -0,006619887 | 0,029635256 | 0,843981623 |
| Vitamin K-dependent protein C measurement | eGFR decline (CKD population) [SH04] | -0,018133125 | 0,087854859 | 0,870422105 |
| Serum alkaline phosphatase levels | eGFR decline (CKD population) [SH04] | 0,002438353 | 0,00285204 | 0,393193398 |
| Vitamin D levels | eGFR decline (CKD population) [SH04] | -0,000397852 | 0,005679679 | 0,944332251 |
| Hemoglobin levels | eGFR decline (CKD population) [SH04] | -0,003073452 | 0,003219567 | 0,340485579 |
| Blood glucose levels | eGFR decline (CKD population) [SH04] | -0,006256987 | 0,005399277 | 0,249094484 |
| Insulin-like growth factor 1 levels | eGFR decline (CKD population) [SH04] | 0,005838996 | 0,002991032 | 0,051670187 |
| Calcium levels | eGFR decline (CKD population) [SH04] | -0,006666595 | 0,004002483 | 0,097355723 |
| Educational attainment (years of education) | eGFR decline (CKD population) [SH04] | -0,004669808 | 0,008820654 | 0,59708325 |
| Smoking status | eGFR decline (CKD population) [SH04] | 0,003573124 | 0,011118191 | 0,74850851 |
| C-reactive protein levels | eGFR decline (CKD population) [SH04] | -0,005515746 | 0,003477173 | 0,114167822 |
| Liver fat | eGFR decline (CKD population) [SH04] | -0,017210051 | 0,019140177 | 0,403208954 |
| Vitamin B12 deficiency anaemia | eGFR decline (CKD population) [SH04] | 0,014057118 | 0,05070485 | 0,807627366 |
| Obesity | eGFR decline (CKD population) [SH04] | 0,00361105 | 0,044745472 | 0,938809628 |
| Pulmonary heart disease | eGFR decline (CKD population) [SH04] | 0,061008542 | 0,039786059 | 0,222714474 |
| Free triiodothyronine | eGFR decline (CKD population) [SH04] | 0,021667306 | 0,051369228 | 0,694856079 |
| FT3 FT4 ratio | eGFR decline (CKD population) [SH04] | -0,016914538 | 0,015333699 | 0,293533648 |
| Free thyroxine | eGFR decline (CKD population) [SH04] | 0,000303537 | 0,007700869 | 0,968689614 |
| HDL cholesterol | eGFR decline (CKD population) [SH04] | -0,005125203 | 0,003264906 | 0,117833638 |
| High TSH | eGFR decline (CKD population) [SH04] | -0,021291117 | 0,046419666 | 0,670289222 |
| Major depression | eGFR decline (CKD population) [SH04] | -0,007377059 | 0,024553444 | 0,765188371 |
| apolipoprotein A-I | eGFR decline (CKD population) [SH04] | 0,001946475 | 0,003115896 | 0,532706789 |
| apolipoprotein B | eGFR decline (CKD population) [SH04] | 0,000491667 | 0,003880875 | 0,899334812 |
| Cigarettes per Day | eGFR decline (CKD population) [SH04] | 0,006841664 | 0,011076132 | 0,543746778 |
| Chronotype | eGFR decline (CKD population) [SH04] | 0,014376331 | 0,01106542 | 0,19901196 |
| smoking initiation | eGFR decline (CKD population) [SH04] | -0,035059166 | 0,015503732 | 0,026353864 |
| Systolic Blood Pressure (SBP) | eGFR decline (CKD population) [SH04] | 0,003888723 | 0,005784615 | 0,502046148 |
| Diastolic Blood Pressure (DBP) | eGFR decline (CKD population) [SH04] | 0,000794574 | 0,006061338 | 0,8958108 |
| Pulse Pressure (PP) | eGFR decline (CKD population) [SH04] | 0,00087679 | 0,004566058 | 0,847853867 |
| Hypertension | eGFR decline (CKD population) [SH04] | 0,008306087 | 0,006808235 | 0,224015564 |
| Urinary sodium-potassium ratio | eGFR decline (CKD population) [SH04] | -0,013418486 | 0,028535782 | 0,642816291 |
| Insomnia | eGFR decline (CKD population) [SH04] | 0,000839236 | 0,02743748 | 0,976146663 |
| LDL cholesterol | eGFR decline (CKD population) [SH04] | -0,000459411 | 0,003398964 | 0,892610181 |
| Low TSH | eGFR decline (CKD population) [SH04] | 0,010198261 | 0,017246725 | 0,559837605 |
| Compound smoking index | eGFR decline (CKD population) [SH04] | -0,006338611 | 0,010398426 | 0,54330615 |
| Type 2 diabetes | eGFR decline (CKD population) [SH04] | -0,001745084 | 0,002931495 | 0,551957581 |
| Total cholesterol | eGFR decline (CKD population) [SH04] | 0,001718294 | 0,00302291 | 0,570234026 |
| Triglyceride | eGFR decline (CKD population) [SH04] | -0,002281953 | 0,003210965 | 0,478048709 |
| Thyroid Stimulating Hormone | eGFR decline (CKD population) [SH04] | -0,006275171 | 0,004310604 | 0,14745785 |
| Sleep duration | eGFR decline (CKD population) [SH04] | 0,000153793 | 0,013406936 | 0,990883222 |
| Alcohol intake frequency. | eGFR decline (CKD population) [SH04] | 0,001295667 | 0,006829931 | 0,849958816 |
| Lipoprotein A | eGFR decline (CKD population) [SH04] | 0,010147951 | 0,037056469 | 0,7903804 |
| Waist-to-hip ratio adjusted BMI | eGFR decline (CKD population) [SH04] | 0,003931684 | 0,003973172 | 0,323204015 |
| Zinc | eGFR decline (CKD population) [SH04] | 0,053997396 | 0,063339896 | 0,550581527 |

| exposure | outcome | isq |
|---|---|---|
| smoking initiation | Incident CKD | 0,97590436 |
| Body mass index | Incident CKD | 0,986649999 |
| Pulse Pressure (PP) | Incident CKD | 0,985060089 |
| Serum uric acid levels | Incident CKD | 0,991980162 |
| Calcium levels | Incident CKD | 0,990260953 |
| Bone mineral density | Incident CKD | 0,991767909 |
| Hemoglobin levels | Incident CKD | 0,990510029 |
| apolipoprotein A-I | Incident CKD | 0,992954368 |
| C-reactive protein levels | Incident CKD | 0,992335287 |
| Vitamin D levels | Incident CKD | 0,992184114 |
| Systolic Blood Pressure (SBP) | Incident CKD | 0,982553257 |
| Hypertension | Incident CKD | 0,981485144 |
| apolipoprotein B | Incident CKD | 0,994086466 |
| Phosphate levels | Incident CKD | 0,98924133 |
| HDL cholesterol | Incident CKD | 0,996324271 |
| Sodium in urine | Incident CKD | 0,977570047 |
| Glycated haemoglobin HbA1c levels | Incident CKD | 0,992606918 |
| Alcohol intake frequency. | Incident CKD | 0,981070055 |
| Smoking status | Incident CKD | 0,976934018 |
| Waist-to-hip ratio adjusted BMI | Incident CKD | 0,988390623 |
| Diastolic Blood Pressure (DBP) | Incident CKD | 0,982081497 |
| Fasting insulin | Incident CKD | 0,980819964 |
| Compound smoking index | Incident CKD | 0,977516398 |
| Educational attainment (years of education) | Incident CKD | 0,977599247 |
| Heel bone mineral density | Incident CKD | 0,993321648 |
| Hypothyroidism | Incident CKD | 0,987671735 |
| Lipoprotein A | Incident CKD | 0,99896214 |
| Type 1 diabetes | Incident CKD | 0,992598627 |
| Serum alkaline phosphatase levels | Incident CKD | 0,993532682 |
| Zinc | Incident CKD | 0,949737095 |
| Type 2 diabetes | Incident CKD | 0,989412653 |
| Insulin-like growth factor 1 levels | Incident CKD | 0,991342779 |
| Thyroid Stimulating Hormone | Incident CKD | 0,993561477 |
| Total cholesterol | Incident CKD | 0,995998065 |
| Body fat percentage | Incident CKD | 0,982496445 |
| Myocardial infarction | Incident CKD | 0,984987022 |
| LDL cholesterol | Incident CKD | 0,995834689 |
| circulating leptin levels | Incident CKD | 0,985988797 |
| Major depression | Incident CKD | 0,973696529 |
| Triglyceride | Incident CKD | 0,995828613 |
| Hyperthyroidism | Incident CKD | 0,982346643 |
| Blood glucose levels | Incident CKD | 0,991614321 |
| Fasting glucose | Incident CKD | 0,991709504 |
| Low TSH | Incident CKD | 0,985688454 |
| Coronary artery disease (Firth correction) | Incident CKD | 0,982032374 |
| Chronotype | Incident CKD | 0,977583424 |
| Sleep duration | Incident CKD | 0,977071141 |
| Free thyroxine | Incident CKD | 0,988347119 |
| Obesity | Incident CKD | 0,978541294 |
| Urinary sodium-potassium ratio | Incident CKD | 0,975652825 |

| | | |
|---|---|---|
| Two-hour glucose | Incident CKD | 0,982960221 |
| Liver fat | Incident CKD | 0,993302321 |
| Liver iron content | Incident CKD | 0,992280951 |
| FT3 FT4 ratio | Incident CKD | 0,993021633 |
| Insomnia | Incident CKD | 0,976984993 |
| Vitamin K-dependent protein S measurement | Incident CKD | 0,993532126 |
| Vitamin K-dependent protein C measurement | Incident CKD | 0,995142523 |
| Cigarettes per Day | Incident CKD | 0,990555292 |
| Fibroblast growth factor 23 levels | Incident CKD | 0,976736505 |
| Chronic heart failure | Incident CKD | 0,789937309 |
| Pulmonary heart disease | Incident CKD | 0,966931621 |
| Growth differentiation factor-15 levels | Incident CKD | 0,997609987 |
| Kidney injury molecule 1 levels | Incident CKD | 0,99413243 |
| High TSH | Incident CKD | 0,980259152 |
| Vitamin B12 deficiency anaemia | Incident CKD | 0,982106131 |
| Free triiodothyronine | Incident CKD | 0,985485541 |
| Cardiovascular disease | Incident CKD | 0,984322895 |
| Thyrotropin-releasing hormone | Incident CKD | 0,985768051 |
| Serum iron | Incident CKD | 0,992411931 |
| smoking initiation | eGFR decline (CKD population) | 0,97590436 |
| Body mass index | eGFR decline (CKD population) | 0,986653818 |
| Pulse Pressure (PP) | eGFR decline (CKD population) | 0,985083935 |
| Serum uric acid levels | eGFR decline (CKD population) | 0,991063494 |
| Calcium levels | eGFR decline (CKD population) | 0,990173609 |
| Bone mineral density | eGFR decline (CKD population) | 0,991635254 |
| Hemoglobin levels | eGFR decline (CKD population) | 0,990438961 |
| apolipoprotein A-I | eGFR decline (CKD population) | 0,992937409 |
| C-reactive protein levels | eGFR decline (CKD population) | 0,99240901 |
| Vitamin D levels | eGFR decline (CKD population) | 0,992375138 |
| Systolic Blood Pressure (SBP) | eGFR decline (CKD population) | 0,982510184 |
| Hypertension | eGFR decline (CKD population) | 0,981591581 |
| apolipoprotein B | eGFR decline (CKD population) | 0,993846234 |
| Phosphate levels | eGFR decline (CKD population) | 0,989129215 |
| HDL cholesterol | eGFR decline (CKD population) | 0,996331955 |
| Sodium in urine | eGFR decline (CKD population) | 0,977570047 |
| Glycated haemoglobin HbA1c levels | eGFR decline (CKD population) | 0,992483409 |
| Smoking status | eGFR decline (CKD population) | 0,976934018 |
| Alcohol intake frequency. | eGFR decline (CKD population) | 0,981168252 |
| Waist-to-hip ratio adjusted BMI | eGFR decline (CKD population) | 0,988403973 |
| Diastolic Blood Pressure (DBP) | eGFR decline (CKD population) | 0,982004748 |
| Fasting insulin | eGFR decline (CKD population) | 0,980739202 |
| Compound smoking index | eGFR decline (CKD population) | 0,977516398 |
| Educational attainment (years of education) | eGFR decline (CKD population) | 0,977629725 |
| Heel bone mineral density | eGFR decline (CKD population) | 0,993260393 |
| Hypothyroidism | eGFR decline (CKD population) | 0,988106127 |
| Lipoprotein A | eGFR decline (CKD population) | 0,999069444 |
| Type 1 diabetes | eGFR decline (CKD population) | 0,992653253 |
| Serum alkaline phosphatase levels | eGFR decline (CKD population) | 0,993529457 |
| Zinc | eGFR decline (CKD population) | 0,949737095 |
| Type 2 diabetes | eGFR decline (CKD population) | 0,989224675 |
| Insulin-like growth factor 1 levels | eGFR decline (CKD population) | 0,991614654 |

| | | |
|---|---|---|
| Thyroid Stimulating Hormone | eGFR decline (CKD population) | 0,993599269 |
| Total cholesterol | eGFR decline (CKD population) | 0,995843698 |
| Body fat percentage | eGFR decline (CKD population) | 0,982525262 |
| Myocardial infarction | eGFR decline (CKD population) | 0,985192678 |
| LDL cholesterol | eGFR decline (CKD population) | 0,995436999 |
| circulating leptin levels | eGFR decline (CKD population) | 0,985988797 |
| Major depression | eGFR decline (CKD population) | 0,973696529 |
| Triglyceride | eGFR decline (CKD population) | 0,995699019 |
| Hyperthyroidism | eGFR decline (CKD population) | 0,982786373 |
| Blood glucose levels | eGFR decline (CKD population) | 0,991655609 |
| Fasting glucose | eGFR decline (CKD population) | 0,991801815 |
| Low TSH | eGFR decline (CKD population) | 0,985198605 |
| Coronary artery disease (Firth correction) | eGFR decline (CKD population) | 0,981944221 |
| Chronotype | eGFR decline (CKD population) | 0,977716181 |
| Sleep duration | eGFR decline (CKD population) | 0,977071141 |
| Free thyroxine | eGFR decline (CKD population) | 0,988452926 |
| Obesity | eGFR decline (CKD population) | 0,980445637 |
| Urinary sodium-potassium ratio | eGFR decline (CKD population) | 0,975652825 |
| Two-hour glucose | eGFR decline (CKD population) | 0,982960221 |
| Liver fat | eGFR decline (CKD population) | 0,993302321 |
| Liver iron content | eGFR decline (CKD population) | 0,995124053 |
| FT3 FT4 ratio | eGFR decline (CKD population) | 0,993021633 |
| Insomnia | eGFR decline (CKD population) | 0,976984993 |
| Vitamin K-dependent protein S measurement | eGFR decline (CKD population) | 0,993532126 |
| Vitamin K-dependent protein C measurement | eGFR decline (CKD population) | 0,996682971 |
| Cigarettes per Day | eGFR decline (CKD population) | 0,990555292 |
| Fibroblast growth factor 23 levels | eGFR decline (CKD population) | 0,976736505 |
| Chronic heart failure | eGFR decline (CKD population) | 0,286303549 |
| Growth differentiation factor-15 levels | eGFR decline (CKD population) | 0,997609987 |
| Kidney injury molecule 1 levels | eGFR decline (CKD population) | 0,99413243 |
| High TSH | eGFR decline (CKD population) | 0,980259152 |
| Vitamin B12 deficiency anaemia | eGFR decline (CKD population) | 0,982106131 |
| Free triiodothyronine | eGFR decline (CKD population) | 0,985485541 |
| Pulmonary heart disease | eGFR decline (CKD population) | 0,97068422 |
| Cardiovascular disease | eGFR decline (CKD population) | 0,984322895 |
| Thyrotropin-releasing hormone | eGFR decline (CKD population) | 0,985768051 |
| Serum iron | eGFR decline (CKD population) | 0,992411931 |
| smoking initiation | eGFR decline (General population) | 0,97590436 |
| Body mass index | eGFR decline (General population) | 0,986665096 |
| Pulse Pressure (PP) | eGFR decline (General population) | 0,985364644 |
| Serum uric acid levels | eGFR decline (General population) | 0,991936059 |
| Calcium levels | eGFR decline (General population) | 0,990208539 |
| Bone mineral density | eGFR decline (General population) | 0,991734543 |
| Hemoglobin levels | eGFR decline (General population) | 0,99051256 |
| apolipoprotein A-I | eGFR decline (General population) | 0,992942297 |
| C-reactive protein levels | eGFR decline (General population) | 0,992271485 |
| Vitamin D levels | eGFR decline (General population) | 0,992184114 |
| Systolic Blood Pressure (SBP) | eGFR decline (General population) | 0,982602834 |
| Hypertension | eGFR decline (General population) | 0,981203165 |
| apolipoprotein B | eGFR decline (General population) | 0,994107195 |
| Phosphate levels | eGFR decline (General population) | 0,989293477 |

| | | |
|---|---|---|
| HDL cholesterol | eGFR decline (General population) | 0,996342145 |
| Sodium in urine | eGFR decline (General population) | 0,977570047 |
| Glycated haemoglobin HbA1c levels | eGFR decline (General population) | 0,992538187 |
| Alcohol intake frequency. | eGFR decline (General population) | 0,981070055 |
| Smoking status | eGFR decline (General population) | 0,976934018 |
| Waist-to-hip ratio adjusted BMI | eGFR decline (General population) | 0,988414187 |
| Diastolic Blood Pressure (DBP) | eGFR decline (General population) | 0,982010731 |
| Fasting insulin | eGFR decline (General population) | 0,980739202 |
| Compound smoking index | eGFR decline (General population) | 0,977524897 |
| Educational attainment (years of education) | eGFR decline (General population) | 0,977547788 |
| Heel bone mineral density | eGFR decline (General population) | 0,993311404 |
| Hypothyroidism | eGFR decline (General population) | 0,987671735 |
| Lipoprotein A | eGFR decline (General population) | 0,998980031 |
| Type 1 diabetes | eGFR decline (General population) | 0,99401648 |
| Serum alkaline phosphatase levels | eGFR decline (General population) | 0,993472755 |
| Zinc | eGFR decline (General population) | 0,949737095 |
| Type 2 diabetes | eGFR decline (General population) | 0,989407054 |
| Insulin-like growth factor 1 levels | eGFR decline (General population) | 0,991570632 |
| Thyroid Stimulating Hormone | eGFR decline (General population) | 0,99354047 |
| Total cholesterol | eGFR decline (General population) | 0,996062412 |
| Body fat percentage | eGFR decline (General population) | 0,982493248 |
| Myocardial infarction | eGFR decline (General population) | 0,984964316 |
| LDL cholesterol | eGFR decline (General population) | 0,995864858 |
| circulating leptin levels | eGFR decline (General population) | 0,985988797 |
| Major depression | eGFR decline (General population) | 0,973696529 |
| Triglyceride | eGFR decline (General population) | 0,995799426 |
| Hyperthyroidism | eGFR decline (General population) | 0,985543257 |
| Blood glucose levels | eGFR decline (General population) | 0,991472606 |
| Fasting glucose | eGFR decline (General population) | 0,991709504 |
| Low TSH | eGFR decline (General population) | 0,985782585 |
| Coronary artery disease (Firth correction) | eGFR decline (General population) | 0,981994145 |
| Chronotype | eGFR decline (General population) | 0,977583424 |
| Sleep duration | eGFR decline (General population) | 0,976993351 |
| Free thyroxine | eGFR decline (General population) | 0,988347119 |
| Obesity | eGFR decline (General population) | 0,978541294 |
| Urinary sodium-potassium ratio | eGFR decline (General population) | 0,975652825 |
| Two-hour glucose | eGFR decline (General population) | 0,982960221 |
| Liver fat | eGFR decline (General population) | 0,992639173 |
| Liver iron content | eGFR decline (General population) | 0,995124053 |
| FT3 FT4 ratio | eGFR decline (General population) | 0,993021633 |
| Insomnia | eGFR decline (General population) | 0,976984993 |
| Vitamin K-dependent protein S measurement | eGFR decline (General population) | 0,993532126 |
| Vitamin K-dependent protein C measurement | eGFR decline (General population) | 0,995142523 |
| Cigarettes per Day | eGFR decline (General population) | 0,990555292 |
| Fibroblast growth factor 23 levels | eGFR decline (General population) | 0,976736505 |
| Chronic heart failure | eGFR decline (General population) | 0,789937309 |
| Pulmonary heart disease | eGFR decline (General population) | 0,966931621 |
| Growth differentiation factor-15 levels | eGFR decline (General population) | 0,997609987 |
| Iron deficiency anemia | eGFR decline (General population) | 0 |
| Kidney injury molecule 1 levels | eGFR decline (General population) | 0,993441898 |
| High TSH | eGFR decline (General population) | 0,980259152 |

| | | |
|---|---|---|
| Vitamin B12 deficiency anaemia | eGFR decline (General population) | 0,982106131 |
| Free triiodothyronine | eGFR decline (General population) | 0,985485541 |
| Cardiovascular disease | eGFR decline (General population) | 0,984322895 |
| Thyrotropin-releasing hormone | eGFR decline (General population) | 0,985768051 |
| Serum iron | eGFR decline (General population) | 0,992411931 |
| smoking initiation | eGFR decline (CKD population) [DB01] | 0,97590436 |
| Body mass index | eGFR decline (CKD population) [DB01] | 0,986649999 |
| Pulse Pressure (PP) | eGFR decline (CKD population) [DB01] | 0,985115643 |
| Serum uric acid levels | eGFR decline (CKD population) [DB01] | 0,991085597 |
| Calcium levels | eGFR decline (CKD population) [DB01] | 0,990227046 |
| Bone mineral density | eGFR decline (CKD population) [DB01] | 0,991633646 |
| Hemoglobin levels | eGFR decline (CKD population) [DB01] | 0,990438961 |
| apolipoprotein A-I | eGFR decline (CKD population) [DB01] | 0,992937409 |
| C-reactive protein levels | eGFR decline (CKD population) [DB01] | 0,99243685 |
| Vitamin D levels | eGFR decline (CKD population) [DB01] | 0,992375138 |
| Systolic Blood Pressure (SBP) | eGFR decline (CKD population) [DB01] | 0,982598328 |
| Hypertension | eGFR decline (CKD population) [DB01] | 0,981674119 |
| apolipoprotein B | eGFR decline (CKD population) [DB01] | 0,993894088 |
| Phosphate levels | eGFR decline (CKD population) [DB01] | 0,989169476 |
| HDL cholesterol | eGFR decline (CKD population) [DB01] | 0,996315384 |
| Sodium in urine | eGFR decline (CKD population) [DB01] | 0,977570047 |
| Glycated haemoglobin HbA1c levels | eGFR decline (CKD population) [DB01] | 0,992504256 |
| Smoking status | eGFR decline (CKD population) [DB01] | 0,976934018 |
| Alcohol intake frequency. | eGFR decline (CKD population) [DB01] | 0,981168252 |
| Waist-to-hip ratio adjusted BMI | eGFR decline (CKD population) [DB01] | 0,988403973 |
| Diastolic Blood Pressure (DBP) | eGFR decline (CKD population) [DB01] | 0,982124633 |
| Fasting insulin | eGFR decline (CKD population) [DB01] | 0,980819964 |
| Compound smoking index | eGFR decline (CKD population) [DB01] | 0,977516398 |
| Educational attainment (years of education) | eGFR decline (CKD population) [DB01] | 0,977629725 |
| Heel bone mineral density | eGFR decline (CKD population) [DB01] | 0,993260393 |
| Hypothyroidism | eGFR decline (CKD population) [DB01] | 0,988106127 |
| Lipoprotein A | eGFR decline (CKD population) [DB01] | 0,999069444 |
| Type 1 diabetes | eGFR decline (CKD population) [DB01] | 0,992653253 |
| Serum alkaline phosphatase levels | eGFR decline (CKD population) [DB01] | 0,993529457 |
| Zinc | eGFR decline (CKD population) [DB01] | 0,949737095 |
| Type 2 diabetes | eGFR decline (CKD population) [DB01] | 0,989224675 |
| Insulin-like growth factor 1 levels | eGFR decline (CKD population) [DB01] | 0,991371195 |
| Thyroid Stimulating Hormone | eGFR decline (CKD population) [DB01] | 0,993599269 |
| Total cholesterol | eGFR decline (CKD population) [DB01] | 0,995843698 |
| Body fat percentage | eGFR decline (CKD population) [DB01] | 0,982526747 |
| Myocardial infarction | eGFR decline (CKD population) [DB01] | 0,985192678 |
| LDL cholesterol | eGFR decline (CKD population) [DB01] | 0,995447351 |
| circulating leptin levels | eGFR decline (CKD population) [DB01] | 0,985988797 |
| Major depression | eGFR decline (CKD population) [DB01] | 0,973696529 |
| Triglyceride | eGFR decline (CKD population) [DB01] | 0,995709603 |
| Hyperthyroidism | eGFR decline (CKD population) [DB01] | 0,982346643 |
| Blood glucose levels | eGFR decline (CKD population) [DB01] | 0,991655609 |
| Fasting glucose | eGFR decline (CKD population) [DB01] | 0,991801815 |
| Low TSH | eGFR decline (CKD population) [DB01] | 0,985198605 |
| Coronary artery disease (Firth correction) | eGFR decline (CKD population) [DB01] | 0,982210061 |
| Chronotype | eGFR decline (CKD population) [DB01] | 0,977715693 |

| | | |
|---|---|---|
| Sleep duration | eGFR decline (CKD population) [DB01] | 0,977071141 |
| Free thyroxine | eGFR decline (CKD population) [DB01] | 0,988452926 |
| Obesity | eGFR decline (CKD population) [DB01] | 0,980445637 |
| Urinary sodium-potassium ratio | eGFR decline (CKD population) [DB01] | 0,975652825 |
| Two-hour glucose | eGFR decline (CKD population) [DB01] | 0,982960221 |
| Liver fat | eGFR decline (CKD population) [DB01] | 0,993302321 |
| Liver iron content | eGFR decline (CKD population) [DB01] | 0,992280951 |
| FT3 FT4 ratio | eGFR decline (CKD population) [DB01] | 0,993021633 |
| Insomnia | eGFR decline (CKD population) [DB01] | 0,976984993 |
| Vitamin K-dependent protein S measurement | eGFR decline (CKD population) [DB01] | 0,993532126 |
| Vitamin K-dependent protein C measurement | eGFR decline (CKD population) [DB01] | 0,996682971 |
| Cigarettes per Day | eGFR decline (CKD population) [DB01] | 0,990555292 |
| Fibroblast growth factor 23 levels | eGFR decline (CKD population) [DB01] | 0,976736505 |
| Chronic heart failure | eGFR decline (CKD population) [DB01] | 0,286303549 |
| Growth differentiation factor-15 levels | eGFR decline (CKD population) [DB01] | 0,997609987 |
| Kidney injury molecule 1 levels | eGFR decline (CKD population) [DB01] | 0,99413243 |
| High TSH | eGFR decline (CKD population) [DB01] | 0,980259152 |
| Vitamin B12 deficiency anaemia | eGFR decline (CKD population) [DB01] | 0,982106131 |
| Free triiodothyronine | eGFR decline (CKD population) [DB01] | 0,985485541 |
| Pulmonary heart disease | eGFR decline (CKD population) [DB01] | 0,97068422 |
| Cardiovascular disease | eGFR decline (CKD population) [DB01] | 0,984322895 |
| Thyrotropin-releasing hormone | eGFR decline (CKD population) [DB01] | 0,985768051 |
| Serum iron | eGFR decline (CKD population) [DB01] | 0,992411931 |
| smoking initiation | eGFR decline (CKD population) [SH01] | 0,97590436 |
| Body mass index | eGFR decline (CKD population) [SH01] | 0,986649999 |
| Pulse Pressure (PP) | eGFR decline (CKD population) [SH01] | 0,985115643 |
| Serum uric acid levels | eGFR decline (CKD population) [SH01] | 0,991085597 |
| Calcium levels | eGFR decline (CKD population) [SH01] | 0,990227046 |
| Bone mineral density | eGFR decline (CKD population) [SH01] | 0,991633646 |
| Hemoglobin levels | eGFR decline (CKD population) [SH01] | 0,990438961 |
| apolipoprotein A-I | eGFR decline (CKD population) [SH01] | 0,992937409 |
| C-reactive protein levels | eGFR decline (CKD population) [SH01] | 0,99243685 |
| Vitamin D levels | eGFR decline (CKD population) [SH01] | 0,992375138 |
| Systolic Blood Pressure (SBP) | eGFR decline (CKD population) [SH01] | 0,982598328 |
| Hypertension | eGFR decline (CKD population) [SH01] | 0,981674119 |
| apolipoprotein B | eGFR decline (CKD population) [SH01] | 0,993894088 |
| Phosphate levels | eGFR decline (CKD population) [SH01] | 0,989169476 |
| HDL cholesterol | eGFR decline (CKD population) [SH01] | 0,996315384 |
| Sodium in urine | eGFR decline (CKD population) [SH01] | 0,977570047 |
| Glycated haemoglobin HbA1c levels | eGFR decline (CKD population) [SH01] | 0,992504256 |
| Smoking status | eGFR decline (CKD population) [SH01] | 0,976934018 |
| Alcohol intake frequency. | eGFR decline (CKD population) [SH01] | 0,981168252 |
| Waist-to-hip ratio adjusted BMI | eGFR decline (CKD population) [SH01] | 0,988403973 |
| Diastolic Blood Pressure (DBP) | eGFR decline (CKD population) [SH01] | 0,982124633 |
| Fasting insulin | eGFR decline (CKD population) [SH01] | 0,980819964 |
| Compound smoking index | eGFR decline (CKD population) [SH01] | 0,977516398 |
| Educational attainment (years of education) | eGFR decline (CKD population) [SH01] | 0,977629725 |
| Heel bone mineral density | eGFR decline (CKD population) [SH01] | 0,993260393 |
| Hypothyroidism | eGFR decline (CKD population) [SH01] | 0,988106127 |
| Lipoprotein A | eGFR decline (CKD population) [SH01] | 0,999069444 |
| Type 1 diabetes | eGFR decline (CKD population) [SH01] | 0,992653253 |

| | | |
|---|---|---|
| Serum alkaline phosphatase levels | eGFR decline (CKD population) [SH01] | 0,993529457 |
| Zinc | eGFR decline (CKD population) [SH01] | 0,949737095 |
| Type 2 diabetes | eGFR decline (CKD population) [SH01] | 0,989224675 |
| Insulin-like growth factor 1 levels | eGFR decline (CKD population) [SH01] | 0,991371195 |
| Thyroid Stimulating Hormone | eGFR decline (CKD population) [SH01] | 0,993599269 |
| Total cholesterol | eGFR decline (CKD population) [SH01] | 0,995843698 |
| Body fat percentage | eGFR decline (CKD population) [SH01] | 0,982526747 |
| Myocardial infarction | eGFR decline (CKD population) [SH01] | 0,985192678 |
| LDL cholesterol | eGFR decline (CKD population) [SH01] | 0,995447351 |
| circulating leptin levels | eGFR decline (CKD population) [SH01] | 0,985988797 |
| Major depression | eGFR decline (CKD population) [SH01] | 0,973696529 |
| Triglyceride | eGFR decline (CKD population) [SH01] | 0,995709603 |
| Hyperthyroidism | eGFR decline (CKD population) [SH01] | 0,982346643 |
| Blood glucose levels | eGFR decline (CKD population) [SH01] | 0,991655609 |
| Fasting glucose | eGFR decline (CKD population) [SH01] | 0,991801815 |
| Low TSH | eGFR decline (CKD population) [SH01] | 0,985198605 |
| Coronary artery disease (Firth correction) | eGFR decline (CKD population) [SH01] | 0,982210061 |
| Chronotype | eGFR decline (CKD population) [SH01] | 0,977715693 |
| Sleep duration | eGFR decline (CKD population) [SH01] | 0,977071141 |
| Free thyroxine | eGFR decline (CKD population) [SH01] | 0,988452926 |
| Obesity | eGFR decline (CKD population) [SH01] | 0,980445637 |
| Urinary sodium-potassium ratio | eGFR decline (CKD population) [SH01] | 0,975652825 |
| Two-hour glucose | eGFR decline (CKD population) [SH01] | 0,982960221 |
| Liver fat | eGFR decline (CKD population) [SH01] | 0,993302321 |
| Liver iron content | eGFR decline (CKD population) [SH01] | 0,992280951 |
| FT3 FT4 ratio | eGFR decline (CKD population) [SH01] | 0,993021633 |
| Insomnia | eGFR decline (CKD population) [SH01] | 0,976984993 |
| Vitamin K-dependent protein S measurement | eGFR decline (CKD population) [SH01] | 0,993532126 |
| Vitamin K-dependent protein C measurement | eGFR decline (CKD population) [SH01] | 0,996682971 |
| Cigarettes per Day | eGFR decline (CKD population) [SH01] | 0,990555292 |
| Fibroblast growth factor 23 levels | eGFR decline (CKD population) [SH01] | 0,976736505 |
| Chronic heart failure | eGFR decline (CKD population) [SH01] | 0,286303549 |
| Growth differentiation factor-15 levels | eGFR decline (CKD population) [SH01] | 0,997609987 |
| Kidney injury molecule 1 levels | eGFR decline (CKD population) [SH01] | 0,99413243 |
| High TSH | eGFR decline (CKD population) [SH01] | 0,980259152 |
| Vitamin B12 deficiency anaemia | eGFR decline (CKD population) [SH01] | 0,982106131 |
| Free triiodothyronine | eGFR decline (CKD population) [SH01] | 0,985485541 |
| Pulmonary heart disease | eGFR decline (CKD population) [SH01] | 0,97068422 |
| Cardiovascular disease | eGFR decline (CKD population) [SH01] | 0,984322895 |
| Thyrotropin-releasing hormone | eGFR decline (CKD population) [SH01] | 0,985768051 |
| Serum iron | eGFR decline (CKD population) [SH01] | 0,992411931 |
| smoking initiation | eGFR decline (CKD population) [SH02] | 0,97590436 |
| Body mass index | eGFR decline (CKD population) [SH02] | 0,986649999 |
| Pulse Pressure (PP) | eGFR decline (CKD population) [SH02] | 0,985115643 |
| Serum uric acid levels | eGFR decline (CKD population) [SH02] | 0,991085597 |
| Calcium levels | eGFR decline (CKD population) [SH02] | 0,990227046 |
| Bone mineral density | eGFR decline (CKD population) [SH02] | 0,991633646 |
| Hemoglobin levels | eGFR decline (CKD population) [SH02] | 0,990438961 |
| apolipoprotein A-I | eGFR decline (CKD population) [SH02] | 0,992937409 |
| C-reactive protein levels | eGFR decline (CKD population) [SH02] | 0,99243685 |
| Vitamin D levels | eGFR decline (CKD population) [SH02] | 0,992375138 |

| | | |
|---|---|---|
| Systolic Blood Pressure (SBP) | eGFR decline (CKD population) [SH02] | 0,982598328 |
| Hypertension | eGFR decline (CKD population) [SH02] | 0,981674119 |
| apolipoprotein B | eGFR decline (CKD population) [SH02] | 0,993894088 |
| Phosphate levels | eGFR decline (CKD population) [SH02] | 0,989169476 |
| HDL cholesterol | eGFR decline (CKD population) [SH02] | 0,996315384 |
| Sodium in urine | eGFR decline (CKD population) [SH02] | 0,977570047 |
| Glycated haemoglobin HbA1c levels | eGFR decline (CKD population) [SH02] | 0,992504256 |
| Smoking status | eGFR decline (CKD population) [SH02] | 0,976934018 |
| Alcohol intake frequency. | eGFR decline (CKD population) [SH02] | 0,981168252 |
| Waist-to-hip ratio adjusted BMI | eGFR decline (CKD population) [SH02] | 0,988403973 |
| Diastolic Blood Pressure (DBP) | eGFR decline (CKD population) [SH02] | 0,982124633 |
| Fasting insulin | eGFR decline (CKD population) [SH02] | 0,980819964 |
| Compound smoking index | eGFR decline (CKD population) [SH02] | 0,977516398 |
| Educational attainment (years of education) | eGFR decline (CKD population) [SH02] | 0,977629725 |
| Heel bone mineral density | eGFR decline (CKD population) [SH02] | 0,993260393 |
| Hypothyroidism | eGFR decline (CKD population) [SH02] | 0,988106127 |
| Lipoprotein A | eGFR decline (CKD population) [SH02] | 0,999069444 |
| Type 1 diabetes | eGFR decline (CKD population) [SH02] | 0,992653253 |
| Serum alkaline phosphatase levels | eGFR decline (CKD population) [SH02] | 0,993529457 |
| Zinc | eGFR decline (CKD population) [SH02] | 0,949737095 |
| Type 2 diabetes | eGFR decline (CKD population) [SH02] | 0,989224675 |
| Insulin-like growth factor 1 levels | eGFR decline (CKD population) [SH02] | 0,991371195 |
| Thyroid Stimulating Hormone | eGFR decline (CKD population) [SH02] | 0,993599269 |
| Total cholesterol | eGFR decline (CKD population) [SH02] | 0,995843698 |
| Body fat percentage | eGFR decline (CKD population) [SH02] | 0,982526747 |
| Myocardial infarction | eGFR decline (CKD population) [SH02] | 0,985192678 |
| LDL cholesterol | eGFR decline (CKD population) [SH02] | 0,995447351 |
| circulating leptin levels | eGFR decline (CKD population) [SH02] | 0,985988797 |
| Major depression | eGFR decline (CKD population) [SH02] | 0,973696529 |
| Triglyceride | eGFR decline (CKD population) [SH02] | 0,995709603 |
| Hyperthyroidism | eGFR decline (CKD population) [SH02] | 0,982346643 |
| Blood glucose levels | eGFR decline (CKD population) [SH02] | 0,991655609 |
| Fasting glucose | eGFR decline (CKD population) [SH02] | 0,991801815 |
| Low TSH | eGFR decline (CKD population) [SH02] | 0,985198605 |
| Coronary artery disease (Firth correction) | eGFR decline (CKD population) [SH02] | 0,982210061 |
| Chronotype | eGFR decline (CKD population) [SH02] | 0,977715693 |
| Sleep duration | eGFR decline (CKD population) [SH02] | 0,977071141 |
| Free thyroxine | eGFR decline (CKD population) [SH02] | 0,988452926 |
| Obesity | eGFR decline (CKD population) [SH02] | 0,980445637 |
| Urinary sodium-potassium ratio | eGFR decline (CKD population) [SH02] | 0,975652825 |
| Two-hour glucose | eGFR decline (CKD population) [SH02] | 0,982960221 |
| Liver fat | eGFR decline (CKD population) [SH02] | 0,993302321 |
| Liver iron content | eGFR decline (CKD population) [SH02] | 0,992280951 |
| FT3 FT4 ratio | eGFR decline (CKD population) [SH02] | 0,993021633 |
| Insomnia | eGFR decline (CKD population) [SH02] | 0,976984993 |
| Vitamin K-dependent protein S measurement | eGFR decline (CKD population) [SH02] | 0,993532126 |
| Vitamin K-dependent protein C measurement | eGFR decline (CKD population) [SH02] | 0,996682971 |
| Cigarettes per Day | eGFR decline (CKD population) [SH02] | 0,990555292 |
| Fibroblast growth factor 23 levels | eGFR decline (CKD population) [SH02] | 0,976736505 |
| Chronic heart failure | eGFR decline (CKD population) [SH02] | 0,286303549 |
| Growth differentiation factor-15 levels | eGFR decline (CKD population) [SH02] | 0,997609987 |

| | | |
|---|---|---|
| Kidney injury molecule 1 levels | eGFR decline (CKD population) [SH02] | 0,99413243 |
| High TSH | eGFR decline (CKD population) [SH02] | 0,980259152 |
| Vitamin B12 deficiency anaemia | eGFR decline (CKD population) [SH02] | 0,982106131 |
| Free triiodothyronine | eGFR decline (CKD population) [SH02] | 0,985485541 |
| Pulmonary heart disease | eGFR decline (CKD population) [SH02] | 0,97068422 |
| Cardiovascular disease | eGFR decline (CKD population) [SH02] | 0,984322895 |
| Thyrotropin-releasing hormone | eGFR decline (CKD population) [SH02] | 0,985768051 |
| Serum iron | eGFR decline (CKD population) [SH02] | 0,992411931 |
| smoking initiation | eGFR decline (CKD population) [SH03] | 0,97590436 |
| Body mass index | eGFR decline (CKD population) [SH03] | 0,986649999 |
| Pulse Pressure (PP) | eGFR decline (CKD population) [SH03] | 0,985115643 |
| Serum uric acid levels | eGFR decline (CKD population) [SH03] | 0,991085597 |
| Calcium levels | eGFR decline (CKD population) [SH03] | 0,990227046 |
| Bone mineral density | eGFR decline (CKD population) [SH03] | 0,991633646 |
| Hemoglobin levels | eGFR decline (CKD population) [SH03] | 0,990438961 |
| apolipoprotein A-I | eGFR decline (CKD population) [SH03] | 0,992937409 |
| C-reactive protein levels | eGFR decline (CKD population) [SH03] | 0,99243685 |
| Vitamin D levels | eGFR decline (CKD population) [SH03] | 0,992375138 |
| Systolic Blood Pressure (SBP) | eGFR decline (CKD population) [SH03] | 0,982598328 |
| Hypertension | eGFR decline (CKD population) [SH03] | 0,981674119 |
| apolipoprotein B | eGFR decline (CKD population) [SH03] | 0,993894088 |
| Phosphate levels | eGFR decline (CKD population) [SH03] | 0,989169476 |
| HDL cholesterol | eGFR decline (CKD population) [SH03] | 0,996315384 |
| Sodium in urine | eGFR decline (CKD population) [SH03] | 0,977570047 |
| Glycated haemoglobin HbA1c levels | eGFR decline (CKD population) [SH03] | 0,992504256 |
| Smoking status | eGFR decline (CKD population) [SH03] | 0,976934018 |
| Alcohol intake frequency. | eGFR decline (CKD population) [SH03] | 0,981168252 |
| Waist-to-hip ratio adjusted BMI | eGFR decline (CKD population) [SH03] | 0,988403973 |
| Diastolic Blood Pressure (DBP) | eGFR decline (CKD population) [SH03] | 0,982124633 |
| Fasting insulin | eGFR decline (CKD population) [SH03] | 0,980819964 |
| Compound smoking index | eGFR decline (CKD population) [SH03] | 0,977516398 |
| Educational attainment (years of education) | eGFR decline (CKD population) [SH03] | 0,977629725 |
| Heel bone mineral density | eGFR decline (CKD population) [SH03] | 0,993260393 |
| Hypothyroidism | eGFR decline (CKD population) [SH03] | 0,988106127 |
| Lipoprotein A | eGFR decline (CKD population) [SH03] | 0,999069444 |
| Type 1 diabetes | eGFR decline (CKD population) [SH03] | 0,992653253 |
| Serum alkaline phosphatase levels | eGFR decline (CKD population) [SH03] | 0,993529457 |
| Zinc | eGFR decline (CKD population) [SH03] | 0,949737095 |
| Type 2 diabetes | eGFR decline (CKD population) [SH03] | 0,989224675 |
| Insulin-like growth factor 1 levels | eGFR decline (CKD population) [SH03] | 0,991371195 |
| Thyroid Stimulating Hormone | eGFR decline (CKD population) [SH03] | 0,993599269 |
| Total cholesterol | eGFR decline (CKD population) [SH03] | 0,995843698 |
| Body fat percentage | eGFR decline (CKD population) [SH03] | 0,982526747 |
| Myocardial infarction | eGFR decline (CKD population) [SH03] | 0,985192678 |
| LDL cholesterol | eGFR decline (CKD population) [SH03] | 0,995447351 |
| circulating leptin levels | eGFR decline (CKD population) [SH03] | 0,985988797 |
| Major depression | eGFR decline (CKD population) [SH03] | 0,973696529 |
| Triglyceride | eGFR decline (CKD population) [SH03] | 0,995709603 |
| Hyperthyroidism | eGFR decline (CKD population) [SH03] | 0,982346643 |
| Blood glucose levels | eGFR decline (CKD population) [SH03] | 0,991655609 |
| Fasting glucose | eGFR decline (CKD population) [SH03] | 0,991801815 |

| | | |
|---|---|---|
| Low TSH | eGFR decline (CKD population) [SH03] | 0,985198605 |
| Coronary artery disease (Firth correction) | eGFR decline (CKD population) [SH03] | 0,982210061 |
| Chronotype | eGFR decline (CKD population) [SH03] | 0,977715693 |
| Sleep duration | eGFR decline (CKD population) [SH03] | 0,977071141 |
| Free thyroxine | eGFR decline (CKD population) [SH03] | 0,988452926 |
| Obesity | eGFR decline (CKD population) [SH03] | 0,980445637 |
| Urinary sodium-potassium ratio | eGFR decline (CKD population) [SH03] | 0,975652825 |
| Two-hour glucose | eGFR decline (CKD population) [SH03] | 0,982960221 |
| Liver fat | eGFR decline (CKD population) [SH03] | 0,993302321 |
| Liver iron content | eGFR decline (CKD population) [SH03] | 0,992280951 |
| FT3 FT4 ratio | eGFR decline (CKD population) [SH03] | 0,993021633 |
| Insomnia | eGFR decline (CKD population) [SH03] | 0,976984993 |
| Vitamin K-dependent protein S measurement | eGFR decline (CKD population) [SH03] | 0,993532126 |
| Vitamin K-dependent protein C measurement | eGFR decline (CKD population) [SH03] | 0,996682971 |
| Cigarettes per Day | eGFR decline (CKD population) [SH03] | 0,990555292 |
| Fibroblast growth factor 23 levels | eGFR decline (CKD population) [SH03] | 0,976736505 |
| Chronic heart failure | eGFR decline (CKD population) [SH03] | 0,286303549 |
| Growth differentiation factor-15 levels | eGFR decline (CKD population) [SH03] | 0,997609987 |
| Kidney injury molecule 1 levels | eGFR decline (CKD population) [SH03] | 0,99413243 |
| High TSH | eGFR decline (CKD population) [SH03] | 0,980259152 |
| Vitamin B12 deficiency anaemia | eGFR decline (CKD population) [SH03] | 0,982106131 |
| Free triiodothyronine | eGFR decline (CKD population) [SH03] | 0,985485541 |
| Pulmonary heart disease | eGFR decline (CKD population) [SH03] | 0,97068422 |
| Cardiovascular disease | eGFR decline (CKD population) [SH03] | 0,984322895 |
| Thyrotropin-releasing hormone | eGFR decline (CKD population) [SH03] | 0,985768051 |
| Serum iron | eGFR decline (CKD population) [SH03] | 0,992411931 |
| smoking initiation | eGFR decline (CKD population) [SH04] | 0,97590436 |
| Body mass index | eGFR decline (CKD population) [SH04] | 0,986649999 |
| Pulse Pressure (PP) | eGFR decline (CKD population) [SH04] | 0,985115643 |
| Serum uric acid levels | eGFR decline (CKD population) [SH04] | 0,991085597 |
| Calcium levels | eGFR decline (CKD population) [SH04] | 0,990227046 |
| Bone mineral density | eGFR decline (CKD population) [SH04] | 0,991633646 |
| Hemoglobin levels | eGFR decline (CKD population) [SH04] | 0,990438961 |
| apolipoprotein A-I | eGFR decline (CKD population) [SH04] | 0,992937409 |
| C-reactive protein levels | eGFR decline (CKD population) [SH04] | 0,99243685 |
| Vitamin D levels | eGFR decline (CKD population) [SH04] | 0,992375138 |
| Systolic Blood Pressure (SBP) | eGFR decline (CKD population) [SH04] | 0,982598328 |
| Hypertension | eGFR decline (CKD population) [SH04] | 0,981674119 |
| apolipoprotein B | eGFR decline (CKD population) [SH04] | 0,993894088 |
| Phosphate levels | eGFR decline (CKD population) [SH04] | 0,989169476 |
| HDL cholesterol | eGFR decline (CKD population) [SH04] | 0,996315384 |
| Sodium in urine | eGFR decline (CKD population) [SH04] | 0,977570047 |
| Glycated haemoglobin HbA1c levels | eGFR decline (CKD population) [SH04] | 0,992504256 |
| Smoking status | eGFR decline (CKD population) [SH04] | 0,976934018 |
| Alcohol intake frequency. | eGFR decline (CKD population) [SH04] | 0,981168252 |
| Waist-to-hip ratio adjusted BMI | eGFR decline (CKD population) [SH04] | 0,988403973 |
| Diastolic Blood Pressure (DBP) | eGFR decline (CKD population) [SH04] | 0,982124633 |
| Fasting insulin | eGFR decline (CKD population) [SH04] | 0,980819964 |
| Compound smoking index | eGFR decline (CKD population) [SH04] | 0,977516398 |
| Educational attainment (years of education) | eGFR decline (CKD population) [SH04] | 0,977629725 |
| Heel bone mineral density | eGFR decline (CKD population) [SH04] | 0,993260393 |

| | | |
|---|---|---|
| Hypothyroidism | eGFR decline (CKD population) [SH04] | 0,988106127 |
| Lipoprotein A | eGFR decline (CKD population) [SH04] | 0,999069444 |
| Type 1 diabetes | eGFR decline (CKD population) [SH04] | 0,992653253 |
| Serum alkaline phosphatase levels | eGFR decline (CKD population) [SH04] | 0,993529457 |
| Zinc | eGFR decline (CKD population) [SH04] | 0,949737095 |
| Type 2 diabetes | eGFR decline (CKD population) [SH04] | 0,989224675 |
| Insulin-like growth factor 1 levels | eGFR decline (CKD population) [SH04] | 0,991371195 |
| Thyroid Stimulating Hormone | eGFR decline (CKD population) [SH04] | 0,993599269 |
| Total cholesterol | eGFR decline (CKD population) [SH04] | 0,995843698 |
| Body fat percentage | eGFR decline (CKD population) [SH04] | 0,982526747 |
| Myocardial infarction | eGFR decline (CKD population) [SH04] | 0,985192678 |
| LDL cholesterol | eGFR decline (CKD population) [SH04] | 0,995447351 |
| circulating leptin levels | eGFR decline (CKD population) [SH04] | 0,985988797 |
| Major depression | eGFR decline (CKD population) [SH04] | 0,973696529 |
| Triglyceride | eGFR decline (CKD population) [SH04] | 0,995709603 |
| Hyperthyroidism | eGFR decline (CKD population) [SH04] | 0,982346643 |
| Blood glucose levels | eGFR decline (CKD population) [SH04] | 0,991655609 |
| Fasting glucose | eGFR decline (CKD population) [SH04] | 0,991801815 |
| Low TSH | eGFR decline (CKD population) [SH04] | 0,985198605 |
| Coronary artery disease (Firth correction) | eGFR decline (CKD population) [SH04] | 0,982210061 |
| Chronotype | eGFR decline (CKD population) [SH04] | 0,977715693 |
| Sleep duration | eGFR decline (CKD population) [SH04] | 0,977071141 |
| Free thyroxine | eGFR decline (CKD population) [SH04] | 0,988452926 |
| Obesity | eGFR decline (CKD population) [SH04] | 0,980445637 |
| Urinary sodium-potassium ratio | eGFR decline (CKD population) [SH04] | 0,975652825 |
| Two-hour glucose | eGFR decline (CKD population) [SH04] | 0,982960221 |
| Liver fat | eGFR decline (CKD population) [SH04] | 0,993302321 |
| Liver iron content | eGFR decline (CKD population) [SH04] | 0,992280951 |
| FT3 FT4 ratio | eGFR decline (CKD population) [SH04] | 0,993021633 |
| Insomnia | eGFR decline (CKD population) [SH04] | 0,976984993 |
| Vitamin K-dependent protein S measurement | eGFR decline (CKD population) [SH04] | 0,993532126 |
| Vitamin K-dependent protein C measurement | eGFR decline (CKD population) [SH04] | 0,996682971 |
| Cigarettes per Day | eGFR decline (CKD population) [SH04] | 0,990555292 |
| Fibroblast growth factor 23 levels | eGFR decline (CKD population) [SH04] | 0,976736505 |
| Chronic heart failure | eGFR decline (CKD population) [SH04] | 0,286303549 |
| Growth differentiation factor-15 levels | eGFR decline (CKD population) [SH04] | 0,997609987 |
| Kidney injury molecule 1 levels | eGFR decline (CKD population) [SH04] | 0,99413243 |
| High TSH | eGFR decline (CKD population) [SH04] | 0,980259152 |
| Vitamin B12 deficiency anaemia | eGFR decline (CKD population) [SH04] | 0,982106131 |
| Free triiodothyronine | eGFR decline (CKD population) [SH04] | 0,985485541 |
| Pulmonary heart disease | eGFR decline (CKD population) [SH04] | 0,97068422 |
| Cardiovascular disease | eGFR decline (CKD population) [SH04] | 0,984322895 |
| Thyrotropin-releasing hormone | eGFR decline (CKD population) [SH04] | 0,985768051 |
| Serum iron | eGFR decline (CKD population) [SH04] | 0,992411931 |

| exposure | outcome | snp_r2.exposure | snp_r2.outcome | correct_causal_direction | steiger_pval |
|---|---|---|---|---|---|
| Body mass index | Incident CKD | 0,048957245 | 0,001723313 | TRUE | 0 |
| Heel bone mineral density | Incident CKD | 0,146845361 | 0,001832966 | TRUE | 0 |
| Two-hour glucose | Incident CKD | 0,013669376 | 8,83E-05 | TRUE | 5,57E-142 |
| Fasting glucose | Incident CKD | 0,039088018 | 0,000376238 | TRUE | 0 |
| Fasting insulin | Incident CKD | 0,013230373 | 0,000196109 | TRUE | 3,00E-251 |
| circulating leptin levels | Incident CKD | 0,003375637 | 1,14E-05 | TRUE | 4,10E-31 |
| Growth differentiation factor-15 levels | Incident CKD | 0,039780996 | 2,27E-05 | TRUE | 3,20E-177 |
| Fibroblast growth factor 23 levels | Incident CKD | 0,015974496 | 0,0002119 | TRUE | 6,88E-59 |
| Kidney injury molecule 1 levels | Incident CKD | 0,071904785 | 0,000911144 | TRUE | 2,50E-271 |
| Coronary artery disease (Firth correction) | Incident CKD | 0,007300356 | 0,00030194 | TRUE | 1,29E-195 |
| Body fat percentage | Incident CKD | 0,052070021 | 0,001402836 | TRUE | 0 |
| Sodium in urine | Incident CKD | 0,004978117 | 0,000240656 | TRUE | 1,02E-134 |
| Glycated haemoglobin HbA1c levels | Incident CKD | 0,119924528 | 0,001665731 | TRUE | 0 |
| Phosphate levels | Incident CKD | 0,04671408 | 0,001327551 | TRUE | 0 |
| Bone mineral density | Incident CKD | 0,163916445 | 0,001838445 | TRUE | 0 |
| Type 1 diabetes | Incident CKD | 0,018891804 | 0,000274036 | TRUE | 0 |
| Liver iron content | Incident CKD | 0,035861903 | 1,09E-05 | TRUE | 1,09E-236 |
| Pancreas iron content | Incident CKD | 0,001193232 | 1,70E-07 | TRUE | 1,17E-07 |
| Chronic heart failure | Incident CKD | 0,000577602 | 2,08E-05 | TRUE | 2,46E-20 |
| Hyperthyroidism | Incident CKD | 0,00104327 | 3,62E-05 | TRUE | 6,30E-35 |
| Hypothyroidism | Incident CKD | 0,013341761 | 0,000308466 | TRUE | 0 |
| Iron deficiency anemia | Incident CKD | 7,23E-05 | 3,24E-06 | TRUE | 0,0013126 |
| Myocardial infarction | Incident CKD | 0,011171417 | 0,00048964 | TRUE | 0 |
| Sleep apnea syndrome | Incident CKD | 0,000132259 | 7,53E-06 | TRUE | 2,65E-05 |
| Serum uric acid levels | Incident CKD | 0,085035586 | 0,003539104 | TRUE | 0 |
| Vitamin K-dependent protein S measurement | Incident CKD | 0,043730561 | 9,75E-06 | TRUE | 2,78E-101 |
| Vitamin K-dependent protein C measurement | Incident CKD | 0,079490235 | 2,44E-05 | TRUE | 3,62E-186 |
| Serum alkaline phosphatase levels | Incident CKD | 0,120919226 | 0,001525383 | TRUE | 0 |
| Vitamin D levels | Incident CKD | 0,022637596 | 0,000484529 | TRUE | 0 |
| Hemoglobin levels | Incident CKD | 0,075600689 | 0,002652363 | TRUE | 0 |
| Blood glucose levels | Incident CKD | 0,029571829 | 0,000478653 | TRUE | 0 |
| Insulin-like growth factor 1 levels | Incident CKD | 0,102476584 | 0,001702498 | TRUE | 0 |
| Calcium levels | Incident CKD | 0,050109893 | 0,001468945 | TRUE | 0 |
| Educational attainment (years of education) | Incident CKD | 0,021051588 | 0,000769204 | TRUE | 0 |
| Smoking status | Incident CKD | 0,011680752 | 0,000322203 | TRUE | 0 |
| C-reactive protein levels | Incident CKD | 0,048821477 | 0,001031929 | TRUE | 0 |
| Liver fat | Incident CKD | 0,033781378 | 7,56E-05 | TRUE | 1,05E-210 |
| Cardiovascular disease | Incident CKD | 0,001126264 | 1,81E-05 | TRUE | 5,68E-11 |
| Serum iron | Incident CKD | 0,005892681 | 1,27E-05 | TRUE | 1,08E-177 |
| Vitamin B12 deficiency anaemia | Incident CKD | 0,00098957 | 6,25E-06 | TRUE | 9,58E-27 |
| Obesity | Incident CKD | 0,002047881 | 2,64E-05 | TRUE | 5,70E-52 |
| Pulmonary heart disease | Incident CKD | 0,002206318 | 2,91E-05 | TRUE | 1,91E-56 |
| Free triiodothyronine | Incident CKD | 0,009044815 | 2,74E-05 | TRUE | 4,83E-87 |
| FT3 FT4 ratio | Incident CKD | 0,035729539 | 2,40E-05 | TRUE | 0 |
| Free thyroxine | Incident CKD | 0,045866992 | 0,000333761 | TRUE | 0 |
| HDL cholesterol | Incident CKD | 0,039798806 | 0,001451278 | TRUE | 0 |
| High TSH | Incident CKD | 0,001950976 | 3,82E-05 | TRUE | 3,26E-37 |
| Major depression | Incident CKD | 0,003833379 | 0,000159537 | TRUE | 7,59E-123 |
| apolipoprotein A-I | Incident CKD | 0,092091565 | 0,001477442 | TRUE | 0 |
| apolipoprotein B | Incident CKD | 0,060426294 | 0,001001371 | TRUE | 0 |
| Cigarettes per Day | Incident CKD | 0,008938938 | 5,76E-05 | TRUE | 2,69E-261 |
| Chronotype | Incident CKD | 0,011579569 | 0,000244929 | TRUE | 5,79E-288 |
| smoking initiation | Incident CKD | 0,006142932 | 0,000245157 | TRUE | 5,11E-221 |
| Systolic Blood Pressure (SBP) | Incident CKD | 0,034550817 | 0,001600148 | TRUE | 0 |
| Diastolic Blood Pressure (DBP) | Incident CKD | 0,033872522 | 0,002275294 | TRUE | 0 |
| Pulse Pressure (PP) | Incident CKD | 0,049998257 | 0,00175091 | TRUE | 0 |
| Hypertension | Incident CKD | 0,022916908 | 0,002162166 | TRUE | 0 |
| Urinary sodium-potassium ratio | Incident CKD | 0,003293316 | 0,000279529 | TRUE | 1,34E-67 |
| Insomnia | Incident CKD | 0,001403853 | 2,53E-05 | TRUE | 1,29E-46 |
| LDL cholesterol | Incident CKD | 0,042145349 | 0,001145842 | TRUE | 0 |
| Low TSH | Incident CKD | 0,014576008 | 0,000328293 | TRUE | 6,98E-245 |
| Thyrotropin-releasing hormone | Incident CKD | 0,021979716 | 6,59E-07 | TRUE | 2,02E-17 |
| suPAR | Incident CKD | 0,032610864 | 1,19E-06 | TRUE | 1,11E-08 |
| Compound smoking index | Incident CKD | 0,012003941 | 0,000331169 | TRUE | 0 |
| Type 2 diabetes | Incident CKD | 0,022582717 | 0,001594638 | TRUE | 0 |
| Total cholesterol | Incident CKD | 0,045556808 | 0,001394305 | TRUE | 0 |
| Triglyceride | Incident CKD | 0,034480652 | 0,0011388 | TRUE | 0 |
| Thyroid Stimulating Hormone | Incident CKD | 0,089207536 | 0,000651689 | TRUE | 0 |
| Sleep duration | Incident CKD | 0,006419005 | 0,000306172 | TRUE | 6,38E-190 |
| Alcohol intake frequency. | Incident CKD | 0,01121043 | 0,000448026 | TRUE | 0 |
| Vitamin B6 | Incident CKD | 0,000505305 | 3,50E-08 | TRUE | 1,40E-07 |
| Lipoprotein A | Incident CKD | 0,046828556 | 4,36E-05 | TRUE | 0 |
| Waist-to-hip ratio adjusted BMI | Incident CKD | 0,039712217 | 0,001375261 | TRUE | 0 |
| Zinc | Incident CKD | 0,103670008 | 1,06E-05 | TRUE | 1,87E-68 |

| | | | | | |
|---|---|---|---|---|---|
| Body mass index | eGFR decline (General population) | 0,048957245 | 0,001611968 | TRUE | 0 |
| Heel bone mineral density | eGFR decline (General population) | 0,146946994 | 0,001791735 | TRUE | 0 |
| Two-hour glucose | eGFR decline (General population) | 0,013669376 | 3,59E-05 | TRUE | 4,64E-145 |
| Fasting glucose | eGFR decline (General population) | 0,039088018 | 0,000265081 | TRUE | 0 |
| Fasting insulin | eGFR decline (General population) | 0,013476709 | 0,000162173 | TRUE | 2,80E-243 |
| circulating leptin levels | eGFR decline (General population) | 0,003375637 | 6,29E-06 | TRUE | 3,65E-31 |
| Growth differentiation factor-15 levels | eGFR decline (General population) | 0,039780996 | 1,75E-05 | TRUE | 2,40E-175 |
| Fibroblast growth factor 23 levels | eGFR decline (General population) | 0,015974496 | 1,87E-05 | TRUE | 9,96E-69 |
| Kidney injury molecule 1 levels | eGFR decline (General population) | 0,071904785 | 0,000635783 | TRUE | 3,43E-278 |
| Coronary artery disease (Firth correction) | eGFR decline (General population) | 0,007727215 | 0,000160811 | TRUE | 4,73E-206 |
| Body fat percentage | eGFR decline (General population) | 0,05366278 | 0,001592259 | TRUE | 0 |
| Sodium in urine | eGFR decline (General population) | 0,004978117 | 0,000209888 | TRUE | 3,53E-121 |
| Glycated haemoglobin HbA1c levels | eGFR decline (General population) | 0,123238829 | 0,001888794 | TRUE | 0 |
| Phosphate levels | eGFR decline (General population) | 0,048247793 | 0,000768597 | TRUE | 0 |
| Bone mineral density | eGFR decline (General population) | 0,168070624 | 0,001972556 | TRUE | 0 |
| Type 1 diabetes | eGFR decline (General population) | 0,020644013 | 0,000283058 | TRUE | 0 |
| Liver iron content | eGFR decline (General population) | 0,037193104 | 2,87E-05 | TRUE | 6,62E-234 |
| Pancreas iron content | eGFR decline (General population) | 0,001193232 | 1,28E-06 | TRUE | 2,76E-07 |
| Chronic heart failure | eGFR decline (General population) | 0,000577602 | 4,70E-05 | TRUE | 1,49E-13 |
| Hyperthyroidism | eGFR decline (General population) | 0,001666552 | 4,27E-05 | TRUE | 2,11E-47 |
| Hypothyroidism | eGFR decline (General population) | 0,013341761 | 0,000221883 | TRUE | 0 |
| Iron deficiency anemia | eGFR decline (General population) | 0,000151095 | 2,66E-05 | TRUE | 0,0071752 |
| Myocardial infarction | eGFR decline (General population) | 0,011456935 | 0,000453593 | TRUE | 3,55E-299 |
| Sleep apnea syndrome | eGFR decline (General population) | 0,000132259 | 2,74E-08 | TRUE | 4,10E-07 |
| Serum uric acid levels | eGFR decline (General population) | 0,086773692 | 0,001688228 | TRUE | 0 |
| Vitamin K-dependent protein S measurement | eGFR decline (General population) | 0,043730561 | 3,88E-05 | TRUE | 1,40E-97 |
| Vitamin K-dependent protein C measurement | eGFR decline (General population) | 0,079490235 | 3,97E-05 | TRUE | 6,87E-183 |
| Serum alkaline phosphatase levels | eGFR decline (General population) | 0,122527313 | 0,001137685 | TRUE | 0 |
| Vitamin D levels | eGFR decline (General population) | 0,022637596 | 0,000284113 | TRUE | 0 |
| Hemoglobin levels | eGFR decline (General population) | 0,075896159 | 0,001485918 | TRUE | 0 |
| Blood glucose levels | eGFR decline (General population) | 0,029929439 | 0,000362703 | TRUE | 0 |
| Insulin-like growth factor 1 levels | eGFR decline (General population) | 0,104789585 | 0,00150875 | TRUE | 0 |
| Calcium levels | eGFR decline (General population) | 0,050525707 | 0,000866614 | TRUE | 0 |
| Educational attainment (years of education) | eGFR decline (General population) | 0,021117603 | 0,00079487 | TRUE | 0 |
| Smoking status | eGFR decline (General population) | 0,011680752 | 0,000416006 | TRUE | 0 |
| C-reactive protein levels | eGFR decline (General population) | 0,049191093 | 0,001375629 | TRUE | 0 |
| Liver fat | eGFR decline (General population) | 0,03552705 | 2,61E-05 | TRUE | 4,07E-224 |
| Cardiovascular disease | eGFR decline (General population) | 0,001126264 | 8,49E-06 | TRUE | 1,66E-11 |
| Serum iron | eGFR decline (General population) | 0,005892681 | 1,86E-05 | TRUE | 2,85E-153 |
| Vitamin B12 deficiency anaemia | eGFR decline (General population) | 0,00098957 | 7,15E-06 | TRUE | 4,86E-24 |
| Obesity | eGFR decline (General population) | 0,002047881 | 7,06E-05 | TRUE | 3,32E-40 |
| Pulmonary heart disease | eGFR decline (General population) | 0,002206318 | 2,83E-05 | TRUE | 5,16E-51 |
| Free triiodothyronine | eGFR decline (General population) | 0,009044815 | 1,40E-05 | TRUE | 4,35E-87 |
| FT3 FT4 ratio | eGFR decline (General population) | 0,035729539 | 9,55E-05 | TRUE | 2,14E-295 |
| Free thyroxine | eGFR decline (General population) | 0,045866992 | 0,000260291 | TRUE | 0 |
| HDL cholesterol | eGFR decline (General population) | 0,040511734 | 0,001028116 | TRUE | 0 |
| High TSH | eGFR decline (General population) | 0,001950976 | 2,19E-05 | TRUE | 2,38E-37 |
| Major depression | eGFR decline (General population) | 0,003833379 | 8,58E-05 | TRUE | 1,74E-120 |
| apolipoprotein A-I | eGFR decline (General population) | 0,093036358 | 0,001287209 | TRUE | 0 |
| apolipoprotein B | eGFR decline (General population) | 0,062989759 | 0,000681189 | TRUE | 0 |
| Cigarettes per Day | eGFR decline (General population) | 0,008938938 | 9,51E-05 | TRUE | 5,72E-223 |
| Chronotype | eGFR decline (General population) | 0,011579569 | 0,000279281 | TRUE | 1,33E-250 |
| smoking initiation | eGFR decline (General population) | 0,006142932 | 0,000305226 | TRUE | 1,79E-175 |
| Systolic Blood Pressure (SBP) | eGFR decline (General population) | 0,035747767 | 0,00109053 | TRUE | 0 |
| Diastolic Blood Pressure (DBP) | eGFR decline (General population) | 0,034834242 | 0,001715719 | TRUE | 0 |
| Pulse Pressure (PP) | eGFR decline (General population) | 0,052255649 | 0,001311201 | TRUE | 0 |
| Hypertension | eGFR decline (General population) | 0,024062899 | 0,001616229 | TRUE | 0 |
| Urinary sodium-potassium ratio | eGFR decline (General population) | 0,003293316 | 5,94E-05 | TRUE | 1,32E-88 |
| Insomnia | eGFR decline (General population) | 0,001403853 | 1,37E-05 | TRUE | 1,20E-43 |
| LDL cholesterol | eGFR decline (General population) | 0,042327712 | 0,000816975 | TRUE | 0 |
| Low TSH | eGFR decline (General population) | 0,014022142 | 0,000131301 | TRUE | 1,83E-248 |
| Thyrotropin-releasing hormone | eGFR decline (General population) | 0,021979716 | 1,69E-06 | TRUE | 2,91E-17 |
| suPAR | eGFR decline (General population) | 0,032610864 | 8,16E-07 | TRUE | 1,09E-08 |
| Compound smoking index | eGFR decline (General population) | 0,012003941 | 0,000419151 | TRUE | 0 |
| Type 2 diabetes | eGFR decline (General population) | 0,022621917 | 0,001581164 | TRUE | 0 |
| Total cholesterol | eGFR decline (General population) | 0,045708434 | 0,000931952 | TRUE | 0 |
| Triglyceride | eGFR decline (General population) | 0,035079353 | 0,000950133 | TRUE | 0 |
| Thyroid Stimulating Hormone | eGFR decline (General population) | 0,08933009 | 0,000604311 | TRUE | 0 |
| Sleep duration | eGFR decline (General population) | 0,006484323 | 0,000200831 | TRUE | 7,28E-182 |
| Alcohol intake frequency. | eGFR decline (General population) | 0,01121043 | 0,00032632 | TRUE | 0 |
| Vitamin B6 | eGFR decline (General population) | 0,000505305 | 3,63E-07 | TRUE | 4,24E-07 |
| Lipoprotein A | eGFR decline (General population) | 0,050159194 | 4,83E-05 | TRUE | 0 |
| Waist-to-hip ratio adjusted BMI | eGFR decline (General population) | 0,039712217 | 0,001138135 | TRUE | 0 |
| Zinc | eGFR decline (General population) | 0,103670008 | 1,53E-05 | TRUE | 4,75E-68 |
| Body mass index | eGFR decline (CKD population) | 0,048957245 | 0,019281689 | TRUE | 4,62E-41 |

| | | | | | |
|---|---|---|---|---|---|
| Heel bone mineral density | eGFR decline (CKD population) | 0,141152303 | 0,017676884 | TRUE | 0 |
| Two-hour glucose | eGFR decline (CKD population) | 0,013669376 | 0,000637106 | TRUE | 3,46E-35 |
| Fasting glucose | eGFR decline (CKD population) | 0,038931571 | 0,003271421 | TRUE | 4,03E-101 |
| Fasting insulin | eGFR decline (CKD population) | 0,013476709 | 0,002270341 | TRUE | 4,93E-24 |
| circulating leptin levels | eGFR decline (CKD population) | 0,003375637 | 0,000185498 | TRUE | 4,53E-09 |
| Growth differentiation factor-15 levels | eGFR decline (CKD population) | 0,039780996 | 0,0001241 | TRUE | 3,48E-95 |
| Fibroblast growth factor 23 levels | eGFR decline (CKD population) | 0,015974496 | 0,000242319 | TRUE | 1,96E-33 |
| Kidney injury molecule 1 levels | eGFR decline (CKD population) | 0,071904785 | 0,001469186 | TRUE | 2,59E-144 |
| Coronary artery disease (Firth correction) | eGFR decline (CKD population) | 0,007245434 | 0,001392585 | TRUE | 3,97E-13 |
| Body fat percentage | eGFR decline (CKD population) | 0,05252193 | 0,017066909 | TRUE | 2,85E-54 |
| Sodium in urine | eGFR decline (CKD population) | 0,004978117 | 0,001685447 | TRUE | 7,82E-06 |
| Glycated haemoglobin HbA1c levels | eGFR decline (CKD population) | 0,115071734 | 0,014055423 | TRUE | 2,02E-277 |
| Phosphate levels | eGFR decline (CKD population) | 0,046036447 | 0,008151884 | TRUE | 1,76E-82 |
| Bone mineral density | eGFR decline (CKD population) | 0,159116302 | 0,020292775 | TRUE | 0 |
| Type 1 diabetes | eGFR decline (CKD population) | 0,018776408 | 0,002693828 | TRUE | 3,23E-40 |
| Liver iron content | eGFR decline (CKD population) | 0,037193104 | 0,000213713 | TRUE | 7,61E-97 |
| Pancreas iron content | eGFR decline (CKD population) | 0,001193232 | 1,16E-05 | TRUE | 0,0005037 |
| Chronic heart failure | eGFR decline (CKD population) | 0,0005157 | 0,000123677 | TRUE | 0,0677441 |
| Hyperthyroidism | eGFR decline (CKD population) | 0,001242769 | 0,000751519 | TRUE | 0,2373345 |
| Hypothyroidism | eGFR decline (CKD population) | 0,013018766 | 0,002886166 | TRUE | 6,25E-21 |
| Iron deficiency anemia | eGFR decline (CKD population) | 7,23E-05 | 8,12E-05 | FALSE | 0,9355141 |
| Myocardial infarction | eGFR decline (CKD population) | 0,010548576 | 0,002185615 | TRUE | 4,69E-18 |
| Sleep apnea syndrome | eGFR decline (CKD population) | 0,000132259 | 5,58E-08 | TRUE | 0,0740352 |
| Serum uric acid levels | eGFR decline (CKD population) | 0,07299336 | 0,010937675 | TRUE | 1,84E-149 |
| Vitamin K-dependent protein S measurement | eGFR decline (CKD population) | 0,043730561 | 1,38E-05 | TRUE | 2,67E-72 |
| Vitamin K-dependent protein C measurement | eGFR decline (CKD population) | 0,075767721 | 0,000327742 | TRUE | 3,09E-116 |
| Serum alkaline phosphatase levels | eGFR decline (CKD population) | 0,117288675 | 0,014170109 | TRUE | 9,37E-294 |
| Vitamin D levels | eGFR decline (CKD population) | 0,022260413 | 0,003605487 | TRUE | 3,38E-44 |
| Hemoglobin levels | eGFR decline (CKD population) | 0,073803144 | 0,013642669 | TRUE | 3,99E-137 |
| Blood glucose levels | eGFR decline (CKD population) | 0,029428967 | 0,004807063 | TRUE | 3,55E-57 |
| Insulin-like growth factor 1 levels | eGFR decline (CKD population) | 0,102380272 | 0,015138697 | TRUE | 7,74E-227 |
| Calcium levels | eGFR decline (CKD population) | 0,047246365 | 0,008620871 | TRUE | 3,59E-86 |
| Educational attainment (years of education) | eGFR decline (CKD population) | 0,020978674 | 0,007500142 | TRUE | 7,87E-20 |
| Smoking status | eGFR decline (CKD population) | 0,011680752 | 0,004907562 | TRUE | 3,24E-09 |
| C-reactive protein levels | eGFR decline (CKD population) | 0,04611941 | 0,009969282 | TRUE | 1,50E-72 |
| Liver fat | eGFR decline (CKD population) | 0,033781378 | 0,000349849 | TRUE | 2,15E-88 |
| Cardiovascular disease | eGFR decline (CKD population) | 0,001126264 | 3,20E-05 | TRUE | 0,0002128 |
| Serum iron | eGFR decline (CKD population) | 0,005892681 | 0,000235563 | TRUE | 4,15E-20 |
| Vitamin B12 deficiency anaemia | eGFR decline (CKD population) | 0,00098957 | 0,000107712 | TRUE | 0,0022854 |
| Obesity | eGFR decline (CKD population) | 0,001905734 | 0,000259091 | TRUE | 3,27E-05 |
| Pulmonary heart disease | eGFR decline (CKD population) | 0,002036033 | 0,000167918 | TRUE | 1,21E-06 |
| Free triiodothyronine | eGFR decline (CKD population) | 0,009044815 | 0,000198052 | TRUE | 5,01E-27 |
| FT3 FT4 ratio | eGFR decline (CKD population) | 0,035729539 | 0,000436189 | TRUE | 3,66E-106 |
| Free thyroxine | eGFR decline (CKD population) | 0,045460474 | 0,002023864 | TRUE | 4,10E-134 |
| HDL cholesterol | eGFR decline (CKD population) | 0,037079429 | 0,010840053 | TRUE | 5,00E-45 |
| High TSH | eGFR decline (CKD population) | 0,001950976 | 0,000422717 | TRUE | 0,0004154 |
| Major depression | eGFR decline (CKD population) | 0,003833379 | 0,002163987 | TRUE | 0,0172722 |
| apolipoprotein A-I | eGFR decline (CKD population) | 0,086899769 | 0,010862557 | TRUE | 2,83E-204 |
| apolipoprotein B | eGFR decline (CKD population) | 0,057018408 | 0,005828256 | TRUE | 7,49E-144 |
| Cigarettes per Day | eGFR decline (CKD population) | 0,008938938 | 0,000821282 | TRUE | 3,42E-23 |
| Chronotype | eGFR decline (CKD population) | 0,011293411 | 0,002446984 | TRUE | 1,18E-17 |
| smoking initiation | eGFR decline (CKD population) | 0,006142932 | 0,003338315 | TRUE | 0,0012299 |
| Systolic Blood Pressure (SBP) | eGFR decline (CKD population) | 0,03399287 | 0,008768083 | TRUE | 1,04E-45 |
| Diastolic Blood Pressure (DBP) | eGFR decline (CKD population) | 0,033475305 | 0,012264425 | TRUE | 3,02E-29 |
| Pulse Pressure (PP) | eGFR decline (CKD population) | 0,049150855 | 0,011573591 | TRUE | 2,49E-72 |
| Hypertension | eGFR decline (CKD population) | 0,023010936 | 0,00716615 | TRUE | 1,90E-25 |
| Urinary sodium-potassium ratio | eGFR decline (CKD population) | 0,003293316 | 0,001281738 | TRUE | 0,0010458 |
| Insomnia | eGFR decline (CKD population) | 0,001403853 | 0,000473232 | TRUE | 0,0192946 |
| LDL cholesterol | eGFR decline (CKD population) | 0,038621875 | 0,00887698 | TRUE | 2,61E-59 |
| Low TSH | eGFR decline (CKD population) | 0,013131082 | 0,000892791 | TRUE | 9,96E-36 |
| Thyrotropin-releasing hormone | eGFR decline (CKD population) | 0,021979716 | 5,96E-05 | TRUE | 4,03E-14 |
| suPAR | eGFR decline (CKD population) | 0,032610864 | 1,42E-06 | TRUE | 2,01E-08 |
| Compound smoking index | eGFR decline (CKD population) | 0,012003941 | 0,004892174 | TRUE | 6,93E-10 |
| Type 2 diabetes | eGFR decline (CKD population) | 0,021766686 | 0,01873529 | TRUE | 0,0866362 |
| Total cholesterol | eGFR decline (CKD population) | 0,043111951 | 0,008976574 | TRUE | 2,23E-72 |
| Triglyceride | eGFR decline (CKD population) | 0,032446029 | 0,008319774 | TRUE | 2,24E-45 |
| Thyroid Stimulating Hormone | eGFR decline (CKD population) | 0,086786255 | 0,006032089 | TRUE | 1,62E-258 |
| Sleep duration | eGFR decline (CKD population) | 0,006419005 | 0,001831553 | TRUE | 8,26E-09 |
| Alcohol intake frequency. | eGFR decline (CKD population) | 0,011144782 | 0,003670658 | TRUE | 3,53E-12 |
| Vitamin B6 | eGFR decline (CKD population) | 0,000505305 | 5,21E-05 | TRUE | 0,0458522 |
| Lipoprotein A | eGFR decline (CKD population) | 0,026405788 | 0,00087769 | TRUE | 2,32E-86 |
| Waist-to-hip ratio adjusted BMI | eGFR decline (CKD population) | 0,039600922 | 0,012066551 | TRUE | 1,68E-46 |
| Zinc | eGFR decline (CKD population) | 0,103670008 | 0,000178948 | TRUE | 1,72E-58 |
